# COMPREHENSIVE GENETIC INVESTIGATION REVEALS HETEROGENEOUS PATHWAYS TO OBSTRUCTIVE SLEEP APNEA

**DOI:** 10.64898/2026.01.08.26343696

**Authors:** Anne E. Justice, Brendan T. Keenan, Geetha Chittoor, Matthew C. Pahl, Hanne Hoskens, Navya Shilpa Josyula, Shreyash Gupta, Soriul Kim, Gudmar Thorleifsson, Elisabetta Manduchi, Alejandro Gutierrez, Fernando Andrade Oliveira, Cheryl L. Ackert-Bicknell, Bryndís Benediktsdóttir, Jaime Boero, Kenneth M. Borthwick, Ning-hung Chen, Peter A. Cistulli, Lacey J. Favazzo, Daniel J. Gottlieb, Hákon Hákonarson, H. Lester Kirchner, Ulysses J. Magalang, Beth A. Malow, Diego R. Mazzotti, Nigel McArdle, Frank D. Mentch, Timothy I. Morgentheler, Thomas Penzel, James A. Pippin, Laura J. Rasmussen-Torvik, Chol Shin, Bhajan Singh, Tamar Sofer, Olivia J. Veatch, David A. Villani, Andrew D. Wells, Marc S. Williams, Phyllis Zee, Michael J. Zuscik, Ingileif Jonsdottir, Thorarinn Gislason, Janet Robishaw, Kári Stefánsson, Struan FA. Grant, Benedikt Hallgrimsson, Allan I. Pack

**Author notes:** CORRESPONDING AUTHOR Anne E. Justice, Department of Population Health Sciences, Geisinger, Danville, PA 17822, USA; email: aejustice1 at Geisinger dot edu. These authors contributed equally to this work (Anne E. Justice, Brendan T. Keenan, Geetha Chittoor, Matthew C. Pahl, Hanne Hoskens). These authors jointly supervised the work (Kári Stefánsson, Struan FA. Grant, Benedikt Hallgrimsson, Allan I. Pack).

## Abstract

Obstructive sleep apnea (OSA) is a common, heritable disorder with diverse clinical presentations and etiologies. We conducted a genome-wide association meta-analysis of 492,107 individuals, including 46,028 OSA cases, identifying 14 genome-wide significant loci, eight of which are novel. Analyses adjusting for body mass index (BMI) revealed three loci independent of obesity, implicating alternative biological pathways contributing to disease. Integrative functional analyses, including chromatin interaction mapping, fine-mapping, and eQTL colocalization, prioritized candidate effector genes. Notably, implicated genes in chondrocytes were associated with craniofacial morphology in both mouse and human multivariate genotype-phenotype mapping, supporting a role for craniofacial structure in OSA risk. These findings highlight key genetic pathways (obesity-related, neurological, and craniofacial) underlying the development of OSA, offering new insights into its complex etiology.

## INTRODUCTION

Obstructive sleep apnea (OSA) is a common and serious medical condition associated with many adverse health consequences^1–6^. OSA aggregates in families ^7–12^, with individuals with a first degree relative with OSA having an approximately two-fold increased risk of presenting with the disorder,^8^ independent of obesity^8,10^. While this familial aggregation has been known for more than two decades, progress in identifying genetic variants has been relatively recent^13–15^.

One particular difficulty in identifying genetic variants is the considerable heterogeneity in OSA presentation; individuals with similar severity of disease may manifest with different symptoms and have developed OSA through different mechanisms^16,17^. Notably, many risk factors for OSA have been shown to be genetically influenced. For instance, obesity and adiposity are major risk factors for developing OSA^18^, and numerous studies have shown important genetic influences on body mass index (BMI)^19–21^ and waist-hip ratio (WHR)^22–24^. Additionally, characteristics of upper airway anatomy such as soft tissue composition^25–27^ and craniofacial shape^28–30^ are important influences on OSA risk^16^ and have been shown to be heritable and to aggregate in families, even after controlling for overall obesity^31–34^. Finally, OSA risk increases in a variety of neurological conditions, suggesting that some genetic influences may act via neurological mechanisms that affect the regulation of breathing^35^. As such, evidence suggests that the principal genetic influences on OSA risk confer their effects through three, potentially overlapping, mechanistic pathways: obesity, neurological factors and craniofacial morphology.

Recent large case-control genome-wide association studies (GWAS) from FinnGen^14,15^ and the Million Veteran’s Project^13,36^ have documented OSA risk variants residing within known obesity risk loci, and fewer signals in non-obesity-associated regions potentially related to OSA more directly. Additionally, quantitative measures of OSA severity^37–39^, most commonly using the apnea-hypopnea index (AHI), have been examined for genetic factors, but rely on data from small population-based cohorts^37–39^. Notably, some data suggest that individuals with OSA identified in the general population differ from patients presenting clinically^40^ and prevalence of sleep-disordered breathing is often milder in population-based studies, resulting in reduced variability. While these population-based studies have identified some notable genetic associations, there has been little replication. The limited success in GWAS of AHI may also reflect the well-established night-to-night variability in the AHI^41^, which increases the challenge of identifying replicable genetic variants.

To address these challenges and further augment the extant literature on genetics of OSA, we conducted a comprehensive genetic study of individuals mainly recruited from institutions with large biobanks linked to electronic health records (EHRs). Analyses focused on both EHR-derived case/control status^42^ among large numbers of individuals from multiple locations and on quantitative traits among those who received an overnight clinical sleep study evaluation. From the collective analyses, we identified genetic associations related to obesity and non-obesity relevant pathways.

We then performed downstream functional analyses to assess causal genes and biological pathways implicated through these genetic associations. We performed variant to gene mapping^43–45^ based on the examination of open chromatin and chromatin-based interactions in cell types relevant to OSA to implicate causative effector genes^46^, as well as other approaches to prioritize putative genes (e.g. eQTL colocalization^47^ and gene-based association analyses^48^). To test whether some of the identified putative effector genes influence craniofacial dimensions, we utilized a novel multivariate genotype-phenotype (MGP) analysis technique based on comparative insights from diversity outbred mice and humans^49,50^. Together, our findings support the hypothesis that a proportion of OSA genetic etiology acts through influences on relevant aspects of craniofacial morphology, while others act via obesity-related or, perhaps, neurological mechanisms.

## RESULTS

We employed a two-staged meta-analysis approach to identify loci associated with OSA (Stage 1 N_TOTAL_= 161,632, N_CASES_= 25,486; Stage 2 N_TOTAL_= 330,475, N_CASES_= 20,542), AHI (Stage 1 N = 9,120; and Stage 2 N = 19,366), and average oxygen saturation (avgSaO_2_; Stage 1 N = 4,758; and Stage 2 N = 2,150), whereby we selected single nucleotide polymorphisms (SNPs) that achieved at least suggestive statistical significance (*P* < 5x10^-5^) in either of the two independent meta-analyses to bring forward to test for genome-wide significance (*P* < 5x10^-8^) in a joint stage 1 plus stage 2 (S1S2) meta-analysis (**Supplementary Figure 1**). Up to 492,107 individuals were included in our final meta-analysis (**Supplementary Tables 1 and 2**).

### Meta-analysis of OSA case-control status in large clinically-ascertained samples identifies novel risk loci

We identified a total of 14 loci (index SNP ± 500 kb), including 11 from the analyses without BMI adjustment and 4 after adjusting for BMI as a covariate; one locus near the *NACA* gene was significant in both analyses (**Tables 1** and **2**, **Supplementary Table 3, Figure 1, Supplementary Figures 2 and 3**). We did not find any loci with strong evidence of independent secondary signals (**Supplementary Table 4**). Unsurprisingly, given the high polygenicity of BMI and the major role of obesity in conferring OSA risk, 10 of our 14 lead variants are within ±500 kb of known BMI risk loci, including our strongest association at the well-established *FTO* locus^51^. For seven loci, the lead variant was within ±500 kb of previously identified OSA risk variants^14,15,36^. Of these seven, our lead SNPs at the *NACA* locus (rs2958127 for OSA and rs4759254 for OSA adjusted for BMI [OSAadjBMI]) were conditionally independent of previously identified OSA risk variants (**Supplementary Table 5**). Thus, our analyses identified eight new genetic associations with OSA risk, including three that remain significant after adjustment for BMI, and confirm six previously-implicated OSA association signals^13–15^.

**Table 1.**
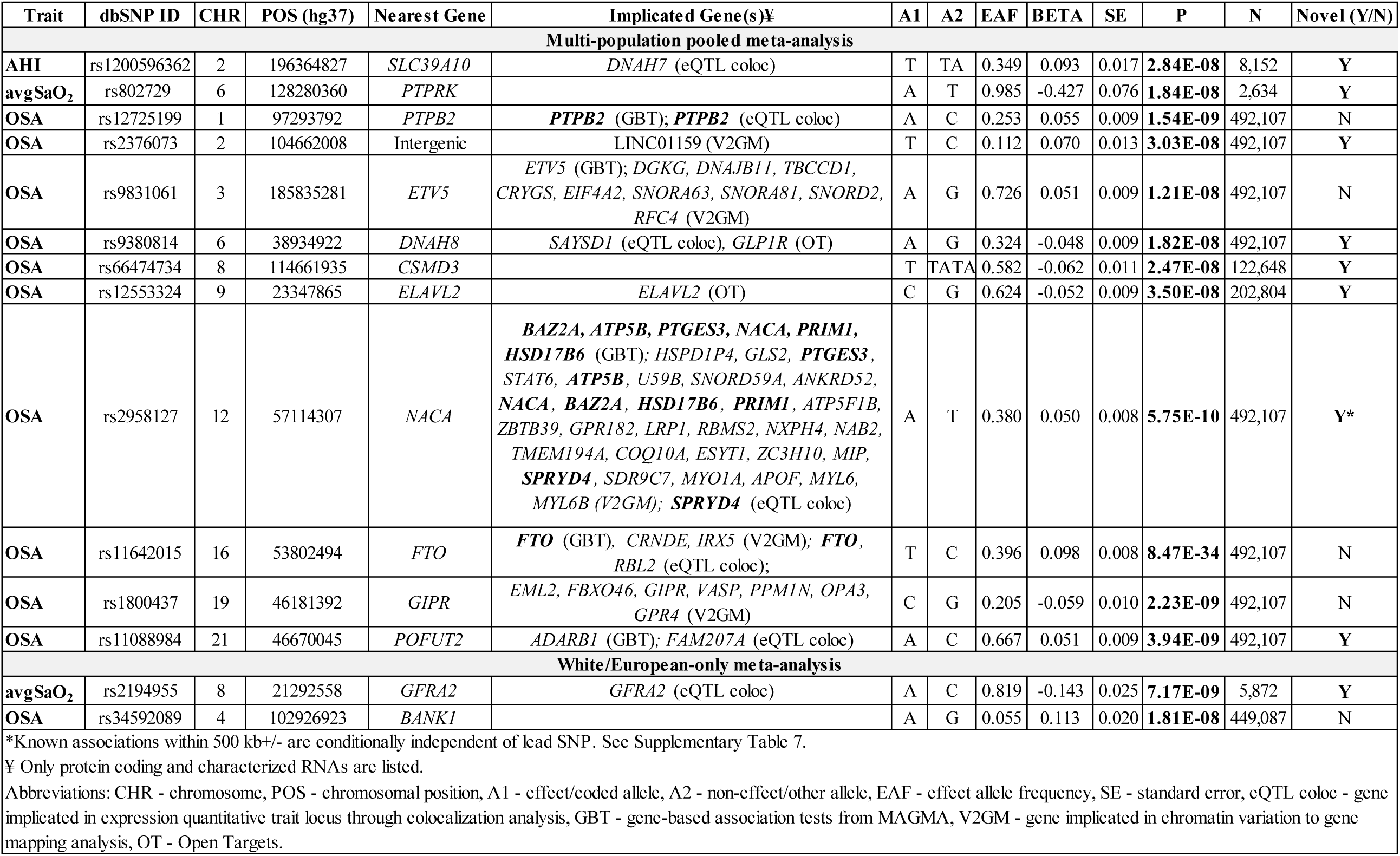
Association results from stage 1 plus stage 2 meta-analysis for the top SNP for each associated locus for model 1 (unadjusted for BMI). Bolded genes are those implicated in more than one follow-up analysis.

**Table 2.** Association results from stage 1 plus stage 2 meta-analysis for the top SNP for each associated locus for model 2 (adjusted for BMI). Bolded genes are those implicated in more than one follow-up analysis.

| Trait | dbSNP ID | CHR | POS | Nearest Gene | Implicated Gene(s)‡ | A1 | A2 | EAF | BETA | SE | P | N | Novel (Y/N) |
| --- | --- | --- | --- | --- | --- | --- | --- | --- | --- | --- | --- | --- | --- |
| **Multi-population pooled meta-analysis** |  |  |  |  |  |  |  |  |  |  |  |  |  |
| **AHIadjBMI** | rs78545995 | 6 | 65804913 | *EYS* |  | A | G | 0.987 | 0.299 | 0.054 | **3.64E-08** | 15,981 | **Y** |
| **avgSaO<sub>2</sub> adjBMI** | rs79914264 | 1 | 29685027 | *PTPRU* |  | A | G | 0.016 | 0.206 | 0.038 | **4.04E-08** | 4,457 | **Y** |
| **OSAadjBMI** | rs2964006 | 5 | 153213880 | *GRIA1* | ***GRIA1*** (V2GM); ***GRIA1***, ***MFAP3*** (eQTL coloc) | T | G | 0.450 | 0.049 | 0.009 | **2.24E-08** | 268,537 | **Y** |
| **OSAadjBMI** | rs4759254 | 12 | 57096674 | *NACA* | *BAZ2A*, *ATP5B*, *PTGES3*, ***NACA***, *PRIM1*, *HSD17B6* (GBT); *MYO1A*, ***NACA***, *ZBTB39*, *SNORD59A*, *ATP5F1B* (V2GM); *SPRYD4* (eQTL coloc) | T | C | 0.644 | -0.055 | 0.009 | **2.99E-09** | 269,620 | **Y\*** |
| **OSAadjBMI** | rs10735928 | 12 | 65769626 | *MSRB3* | ***MSRB3*** (GBT); ***MSRB3*** (OT); *LEMD3*, *WIF1* (eQTL coloc) | A | C | 0.382 | 0.054 | 0.009 | **7.73E-10** | 269,620 | **N** |
| **White/European-only meta-analysis** |  |  |  |  |  |  |  |  |  |  |  |  |  |
| **OSAadjBMI** | rs60791051 | 9 | 71941742 | *FAM189A2* | *FAM189A2*, *TJP2* (V2GM) | A | C | 0.939 | -0.139 | 0.026 | **4.80E-08** | 104,016 | **Y** |

\*Known associations within 500 kb+/- are conditionally independent of lead SNP. See Supplementary Table 5.
‡ Only protein coding and characterized RNAs are listed.
Abbreviations: CHR - chromosome, POS - chromosomal position, A1 - effect/coded allele, A2 - non-effect/other allele, EAF - effect allele frequency, SE - standard error, eQTL coloc - gene implicated in expression quantitative trait locus through colocalization analysis, GBT - gene-based association tests from MAGMA, V2GM - gene implicated in chromatin variation to gene mapping analysis, OT - Open Targets.

To narrow the genetic locus and pinpoint potentially causal variants underlying association signals, we performed fine-mapping analyses. The lead variant from the GWAS had the highest posterior probability (PP) at 12 of the 14 signals. Of the 14, two SNPs displayed a PP of inclusion >0.5 and five loci (four for OSA and one for OSAadjBMI) identified five or fewer SNPs within the 95% credible set (**Supplementary Table 6, Supplementary Figures 4 and 5**). Of note, at the *BANK1* locus, rs34592089 was the only variant in the 95% credible set (PP=0.99). Two additional regions, near *ETV5* and *CSMD3*, were resolved to fewer than 15 variants in the 95% credible set spanning a region <10 kb.

### Meta-analysis of quantitative traits in population-based and clinically ascertained studies identifies genome-wide significant loci

We also conducted a two-stage fixed-effects meta-analysis in up to 28,486 and 6,908 individuals to identify genomic variation associated with the AHI and avgSaO_2_, respectively. Our stage 1 meta-analysis included two studies each with AHI and avgSaO_2_ (N_AHI_ = 9,120, N_avgSaO2_ = 4,758) and stage 2 included five studies with AHI (N_AHI_ = 19,366) and two with avgSaO_2_ (N_avgSaO2_= 2,150), including both population-based and clinically ascertained studies (**Supplementary Tables 1 and 2**). For AHI, two novel loci met our pre-defined significance criteria, one with BMI adjustment and one without (**Tables 1** and **2**, **Supplementary Table 7**, **Figure 1**, **Supplementary Figures 6 and 7**). For avgSaO_2_, we identified three significantly associated loci, including two without and one with BMI adjustment (**Tables 1** and **2**, **Supplementary Table 8**, **Figure 1**, **Supplementary Figures 8 and 9**). We next performed approximate joint conditional analysis on our top variants to identify potential secondary signals, but none were observed (**Supplementary Table 4**). Notably, our top associated variant with avgSaO_2_adjBMI (rs79914264) is nearby (±500 kb) a known BMI association (rs12407884)^21^, but is likely not the same signal due to low linkage disequilibrium (LD) based on the 1000 Genomes reference data (r^2^=0.0052)^52^. Associations with avgSaO_2_ were not within 500kb of index SNPs reported in the Sleep Disorders Knowledge Portal^53^ (which includes suggestive associations with avgSaO_2_), and thus represent the first reported genome-wide significant loci for this trait.

**Figure 1.**
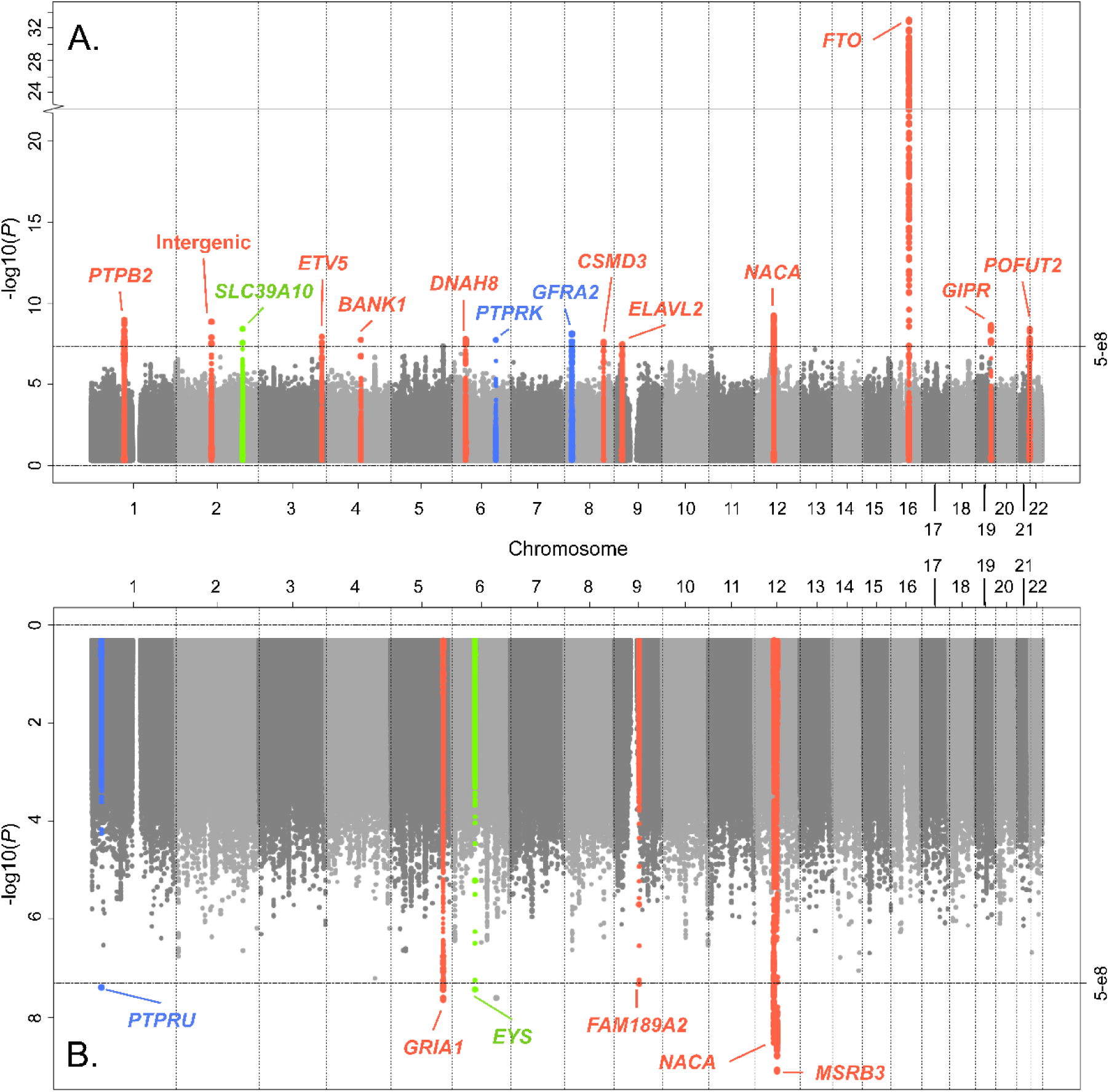
Miami Plot of GWAS results. The plot shows single variant GWAS results for the stage 1 plus stage 2 meta-analysis of OSA, AHI, and avgSaO_2_ without adjustment for BMI (A) and with adjustment for BMI (B). Each significant association signal is annotated with its nearest gene. The horizontal dashed line indicates genome-wide significant threshold P = 5 × 10^-8^.

Fine-mapping of the identified quantitative trait loci was performed using an LD reference based on unrelated individuals of European genetic similarity in the MyCode Study (**Supplementary Table 6**, **Supplementary Figures 10-13**). At all five loci, the lead variant from the GWAS was the variant with the highest PP, including 4 that yielded a PP>0.5. At the *PTPRU* locus for avgSaO_2_adjBMI, rs79914264 was the only variant in the 95% credible set (PP=0.9890). All AHI and avgSaO_2_ loci were resolved to 15 or fewer variants in the 95% credible set.

### Multiple identified loci validate in previously published GWAS data

To determine the generalizability of our results, we compared our top variants to those reported by the Million Veterans Program (MVP)^13^, which included both OSA and OSAadjBMI. We also queried FinnGen Release 11 GWAS results^15^, which were not adjusted for BMI. Of our 14 OSA-associated loci, all were directionally consistent. Additionally, all but three (rs66474734 near *CSMD3*, rs60791051 near *FAM189A2*, and rs11088984 near *POFUT2*) achieved nominal significance (*P*<0.05) and nine achieved Bonferroni-corrected significance in one or both previous GWAS, including three of our novel association signals (**Supplementary Table 9**). Supporting consistent associations across studies, 10 loci were genome-wide significant in a fixed effects meta-analysis between the current study results and lookups. While we found strong replication for association signals from case-control analyses, loci identified in our quantitative trait analyses were not significant in existing GWAS available in the Sleep Disorders Knowledge Portal (SDKP); this may reflect differences in study design, such as the reliance of past studies on population-based samples with less trait variance, covariate adjustments^38,39,54,55^, the inherent night-to-night variability in AHI measurement^41^, and/or low sample size.

### Previously published GWAS loci replicate in our current dataset

To determine whether previously published loci replicated in our dataset, we examined the lead published variants from prior publications (n = 64 OSA SNPs, n = 16 OSAadjBMI SNPs) representing 56 non-overlapping loci (±500 kb)^13–15^ associated with OSA case/control status. We replicated previous findings (*P*<8.93x10^-4^) at 14 loci (**Supplementary Table 10**), including 4 that were robust after adjustment for BMI. An additional 16 loci were nominally associated (*P*<0.05) in our case-control GWAS.

We also investigated previously published GWAS SNPs for quantitative traits reported in the SDKP, including two SNPs for AHI and 49 for avgSaO_2_ in our meta-analyses (**Supplementary Table 11**). Of the 51 lookups, we did not observe any associations meeting a Bonferroni-corrected significance threshold (*P*<0.05/51 = 9.8x10^-4^). However, we replicated four loci for avgSaO_2_ at nominal significance (*P*<0.05), including rs756933193 (*LRP1B*), rs6853180 (*PABPC4L*), rs6948904 (*THSD7A*), and rs565226335 (*ARGLU1*). Of these, all but rs6853180 (*PABPC4L*) were robust following adjustment for BMI. None of these were considered genome-wide significant in the previous publication^54^, yet our results provide further evidence that these loci are valid association signals and warrant future consideration.

### Traits With Shared Genetic Etiologies With OSA

#### Genetic Correlation

Given that we identified nearby associations with other sleep conditions and many known BMI-associated loci, we quantified the shared genetic architecture between OSA traits and others using genetic correlation analyses and previously published GWAS on sleep^56,57^, obesity^20,22,23,58^, and cardiometabolic traits^59–63^ (see **Methods**). Across the 22 comparative traits, the majority (68%) were correlated with our traits at nominal significance or lower (*P*<0.05), including sleep traits, anthropometric traits, obesity traits, coronary artery disease (CAD) and type 2 diabetes (T2D) (**Supplementary Table 12**). All of our OSA-related traits exhibited genetic correlations in the expected direction given phenotypic correlations.

Similar to previous GWAS, we observed significant genetic correlations between OSA and other sleep traits, and cardiometabolic and obesity related traits. OSAadjBMI displayed similar genetic correlations as OSA, although somewhat diminished in magnitude, with the addition of WHRadjBMI meeting Bonferroni-corrected significance for OSAadjBMI. AHI was significantly correlated with BMI (r^2^=45.7%, *P*=8.3E-10), positively associated with sleepiness, and several cardiometabolic and obesity-related traits, and negatively associated with high-density lipoprotein cholesterol [HDL]. After adjusting for BMI, we observed only nominal and positive genetic correlations between AHIadjBMI and OSAadjBMI, waist-to-hip ratio adjusted for BMI [WHRadjBMI], and a nominal and negative correlation with BMI. In addition to OSA, we observed nominal genetic correlations between avgSaO_2_ and several cardiometabolic traits, consistent with expectations (**Supplementary Table 12**).

#### Phenome-wide Association Study (PheWAS)

To further understand potential pleiotropy in our sample and provide initial insights into phenotypic mechanisms or consequences related to the genetic influences on OSA risk, we conducted a PheWAS of our top associated variants in the MyCode study^64^. In total, we identify 142 suggestively significant SNP-PheCode associations (**Supplementary Table 13**, **Figure 2**, *P*<2.9x10^-4^), including 57 that achieved phenome-wide significance (*P*<2.9x10^-5^). Most of these (62% of SNP-PheCode associations) resulted from associations at the *FTO* locus. For our known obesity-associated loci, including *FTO*, we confirmed expected associations with obesity (PheCodes 278.*) and cardiometabolic phenotypes, such as diabetes-related traits (PheCodes 250.*) and hypertension-related traits (PheCodes 401.*), as well as other obesity relevant clinical traits (e.g. PheCodes polycystic ovary disease – 256.4, incisional hernia – 550.6). Among our OSA SNPs without established associations with obesity, rs66474734 near *CSMD3* was associated with pain (PheCodes 338 and 338.2) and intervertebral disc disorders (PheCodes 722 and 722.6). Additionally, rs4759254, which was associated with OSAadjBMI, was associated with atrial fibrillation (PheCodes 427.2 and 427.21), in line with known phenotypic correlations between OSA and atrial fibrillation^65^. We observed suggestive associations between quantitative trait risk loci and cardiometabolic traits; for example, AHIadjBMI SNP rs78545995 was associated with cardiomyopathy (PheCode 425.8) and disorders of pancreatic internal secretion (PheCode 251).

**Figure 2.**
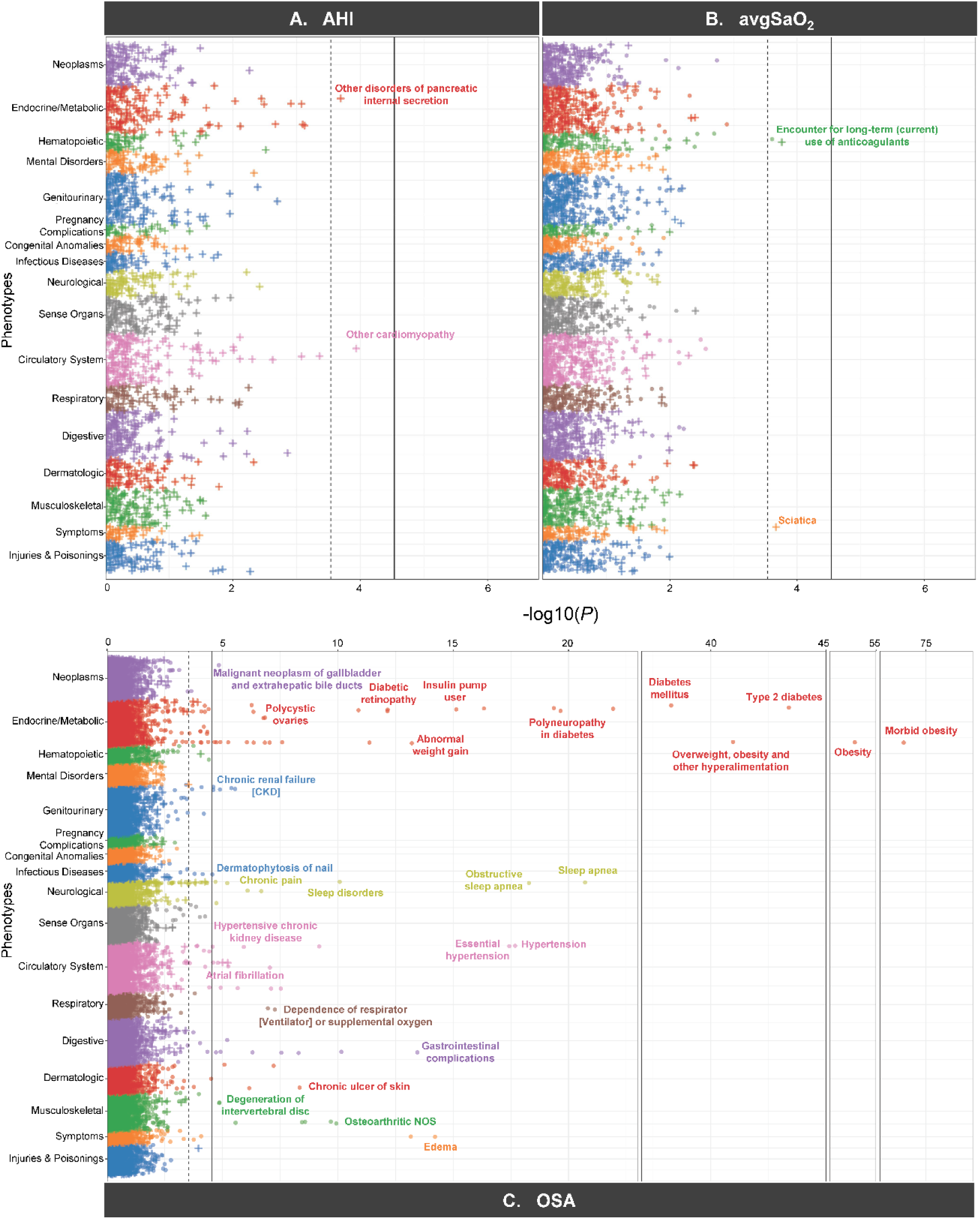
PheWAS Manhattan plots. Manhattan plots of the PheWAS for SNPs associated with A) AHI or AHIadjBMI, B) avgSaO_2_ or avgSaO_2_adjBMI, and C) OSA or OSAadjBMI. The solid line indicates phenome-wide significance threshold (P < 0.05/1,722 PheCodes = 2.9×10^-5^), and the dashed line indicates suggestive significance (P < 2.9x10^-4^). Only a subset of significant PheCodes are annotated with their phenotype, including the ones highlighted in the main text. A full list of suggestively significant associations is available in Supplementary Table 13.

### Identifying Causal Genes and Genomic Pathways to OSA

We next utilized several complementary approaches to prioritize likely causal genes and potential biological mechanisms underlying our association signals, including performing *post-hoc* gene-based analyses (MAGMA^48^ and FUMA^66^); annotation of predicted functional consequences of variants (Variant Effect Predictor [VEP]^67^, Open Targets [OT]^68,69^); integrative analyses that leverage publicly available and newly generated ‘omic resources, including chromatin conformation capture-based variant to gene mapping (V2G) in several relevant tissues (e.g., chondrocytes^43,70^) and eQTL colocalization using publicly available resources (LocusFocus^71^). We hypothesized that noncoding variants would impact enhancers in cells/tissues relevant for OSA, including skeletal, metabolic, and neuronal tissues. Skeletal cell types (e.g., chondrocytes and osteoblasts) are likely to influence craniofacial traits relevant to OSA as these, along with osteoclasts, are involved in craniofacial growth and development^32,72^. Metabolic cell types (e.g., adipocytes and liver) may contribute to OSA through driving increased adiposity (e.g., BMI and/or specific fat depots such as tongue fat^27,73^). Neuronal cell types may influence OSA through circuits important for sleep and breathing^74,75^. Prioritized genes are provided for each association region in **Table 1**.

#### Gene-Based Analyses

In our gene-based analysis^48^ for OSA, we identified 12 significantly associated genes across six regions (**Tables 1** and **2**, **Supplementary Table 14**), including two genes (*TRIM66* and *STK33*) in association regions not identified in our single variant GWAS. Of these 12, only three (*PTBP2*, *ETV5*, and *NACA*) were the nearest gene to our lead SNPs. One region, near *NACA*, identified significant associations with six genes within 200 kb of our lead SNP. For OSAadjBMI, we identified eight significantly associated genes across three regions, including *MED23* (Mediator Complex Subunit 23) on chromosome 6. Notably, the *MED23* region was not represented in our single variant GWAS. While there are no known direct connections between *MED23* and sleep, this gene plays an important role in insulin-mediated adipogenesis^76^ and, like many other sleep-related genes, may affect obesity and metabolism. No additional association regions or genes were identified in our gene-based analysis for quantitative traits. Full FUMA results for all GWAS analyses are publicly available (see **Data Availability Statement**).

#### Functional Consequences of Candidate Variants

To focus on the most likely causal variants underlying association signals, we identified candidate SNPs with the highest PP from our fine-mapping analyses (see **Supplementary Tables 6 and 15**). The majority of the fine-mapped variants were outside of protein coding regions (i.e. intronic, intergenic, within uncharacterized RNA or pseudogenes; **Supplementary Table 15**). Of note, rs1800437 (associated with OSA) is a missense variant in *GIPR* with a moderate impact (based on VEP) and likely damaging based on predictive algorithms (CADD^77^ [combined annotation dependent depletion] = 34 and PolyPhen = 0.999). Our lead SNP within an exon of *NACA*, rs2958127, is also a missense variant, but likely benign. We identified three association regions where all top fine-mapped SNPs exhibited a high Open Targets “locus-to-gene” score (L2G > 0.8): *GLP1R* (rs9380814), *ELALV2* (rs12553324), and *MSRB3* (rs10735928). rs10784447 associated with OSAadjBMI near *MSRB3*, rrs1421085 in *FTO*, rs869399 in the 5’ region of *ETV5*, and rs12740489 in an intergenic region near *PTPB2* exhibit moderate to high CADD scores (>0.15); however, there is no additional evidence that these variants are damaging or regulatory based on evidence from VEP. None of the top fine-mapped variants for quantitative traits were coding variants with likely pathogenic effects or within known regulatory regions based on evidence ascertained from VEP.

#### Physical Variant-to-Gene Mapping for OSA Loci Implicates Effector Genes in OSA-related Cell Types

We next applied our state-of-the-art chromatin conformation capture-based variant-to-gene (V2G) mapping approach to identify candidate effector genes likely to confer associations through specific biological pathways. Given our hypothesis that some OSA risk variants function through genes that determine craniofacial structure, cartilage-producing chondrocytes were of primary interest because chondrocyte differentiation and proliferation within the cranial synchondroses is the major driver of the skeletal shape surrounding the pharynx (e.g., midfacial growth, cranial base length and cranial base flexion^78–82^). As eQTL data in chondrocytes are not publicly available (plus GTEx data are generated from bulk tissue made up of heterogeneous cell types), we utilized epigenetic and chromatin interaction data generated in chondrocytes for 3D genomic variant-to gene-mapping. Accordingly, we generated chromatin accessibility and chromatin contact profiles using ATAC-seq and Hi-C. We defined non-coding cis-regulatory elements harboring associated variants that directly contacted gene promoters and subsequently determined a set of variants in strong LD (r^2^>0.8; proxy variants) with lead variants from the current study, as well as the from a recent meta-analysis for OSA,^13^ that also coincided with open chromatin in chondrocytes (**Figure 3**). In addition, and to contrast our findings with chondrocytes, we queried OSA loci in similar promoter-focused Capture C / Hi-C datasets we have generated from relevant neurological, metabolic, or immune settings that have traditionally been considered to function outside of craniofacial structure (see **Methods** and **Supplementary Table 16** for specific cell types).

**Figure 3.**
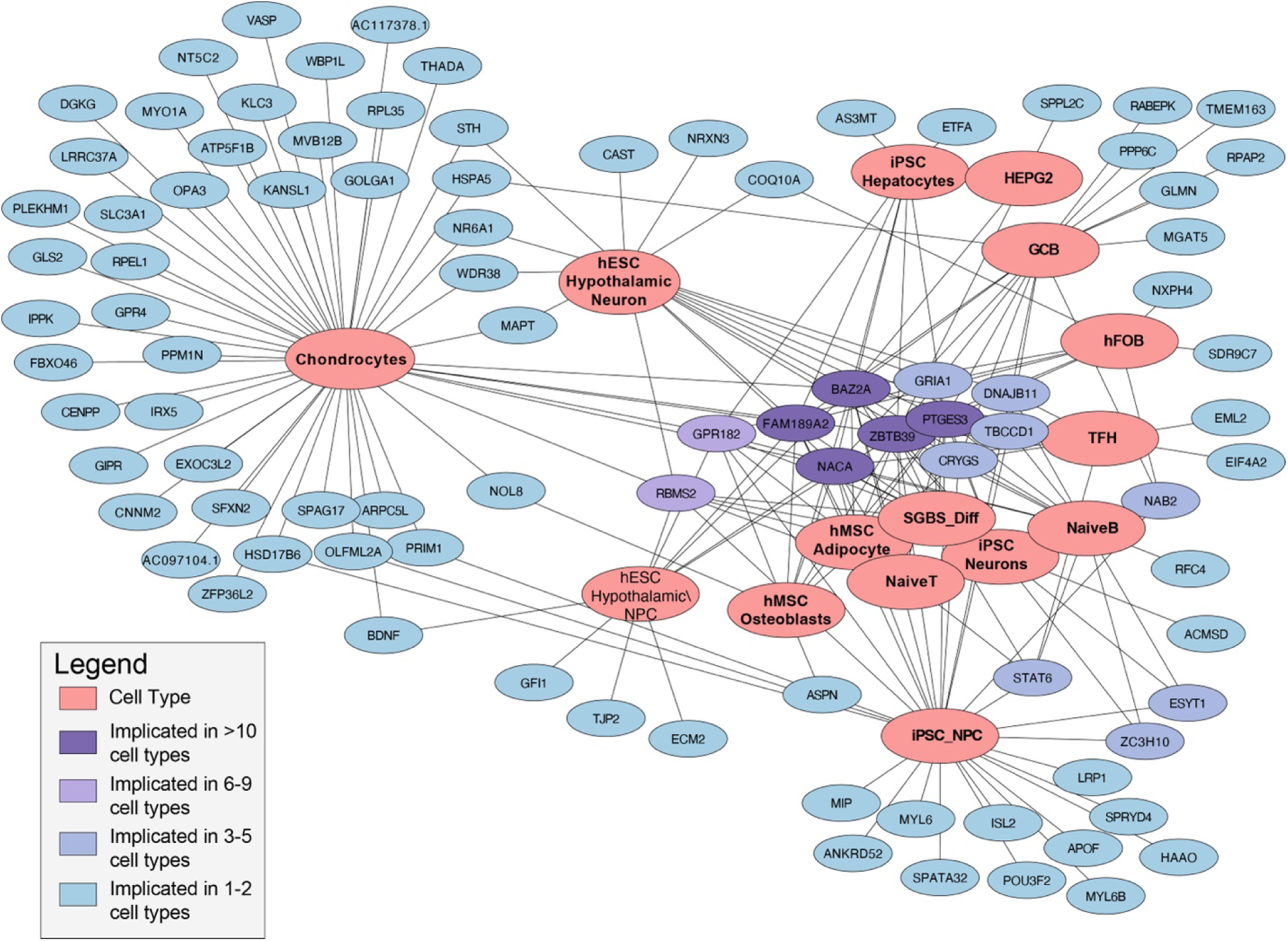
Physical variant-to-gene mapping. Network visualizations of genes nominated by variant-to-gene mapping in each cell type. Nodes represent either a gene (shades of blue) or celltype (pink). Each edge represents that the indicated gene was implicated in the connected cell type.

In chondrocytes, this V2G mapping implicated 47 protein coding genes and 40 non-coding genes, including *THADA, EXOC3L2,* and *NT5C2*. Across all cell types, we observed a total of 184 candidate effector genes whose promoters were contacted by 128 proxy variants in accessible chromatin, corresponding to at least one causal gene being implicated at 34 of the 60 unique OSA-related sentinel variants under investigation (**Supplementary Table 16**). Several genes, such as *ZBTB39*, *BAZ2A*, and *SNORD59A*, displayed contacts across multiple cell types. Other genes were implicated in a more restricted subset of cell types, such as *ASPN* in osteoblasts, *LRP1* and *POU3F2* in neural progenitor cells, and *CAST* and *NRXN3* in embryonic stem cell derived hypothalamic neurons (**Figure 3**). Taken together, these results nominated effector genes for multiple signals, often beyond the nearest gene.

#### Multivariate Genotype Mapping for Mouse and Human Craniofacial Shape

Variation in craniofacial structures is known to influence risk for OSA^28–30,53,83–93^. We used multivariate genotype-phenotype mapping (MGP) to address this mechanism and evaluate whether genes nominated in chondrocytes influenced craniofacial dimensions in both mouse and human data^49,50^. MGP evaluates whether cell type-specific candidate gene sets are jointly associated with variation in craniofacial shape based on 95 3D landmarks quantified on cranial microCT scans of 1266 Diversity Outbred mice (‘mouse MGP’)^49^ and in human samples (‘human MGP’)^50^. These associations allow us to evaluate the hypothesis that genomic variants identified in the human GWAS influence OSA through craniofacial shape. To assess if results were specific to chondrocytes, we similarly examined genes nominated from our V2G mapping analyses in other cell types (see **Supplementary Table 16**).

In the mouse MGP analyses, all gene lists showed statistical significance (P < 0.1/15 = 0.007) in the random gene permutation tests, suggesting enrichment for genes associated with craniofacial shape effects, and were significantly associated with cranial shape when compared to random gene sets of the same length. The chondrocyte specific gene list explained by far the most variation in craniofacial shape when compared to other cell-specific gene lists and was the only list for which the shape permutation test was also significant (*P* < 0.1/15 = 0.007) (**Figure 4A**, **Supplementary Table 17**). Within this list, *Lrrc37a*, *Plekhm1*, *Mapt*, and *Kansl1* had the strongest associations with craniofacial variation along the 1^st^ partial least squares (PLS) axis, which is associated with variation in the relative size of the basicranium, facial prognathism and neurocranial size (**Figure 4A**, **Supplementary Table 18**); at one end of this PLS axis, mice have shorter basicranial, more rounded neurocrania and shorter faces. Two additional gene sets, human mesenchymal stem cell (hMSC) osteoblasts and liver (HEPG2), were nominally significant (*P* < 0.1) for the shape permutation test. While the osteoblasts gene list explained less variation in shape compared to chondrocytes, the 3D morphs and heatmaps show a more pronounced effect on cranial vaulting (**Supplementary Figure 4A**). Given existing data in humans showing that more retrognathic mandibles are a risk factor for OSA^32^, this result is consistent with the hypothesis that putative causal genes differentially expressed in chondrocytes are associated with a phenotypic shape effect that influences OSA risk.

**Figure 4:**
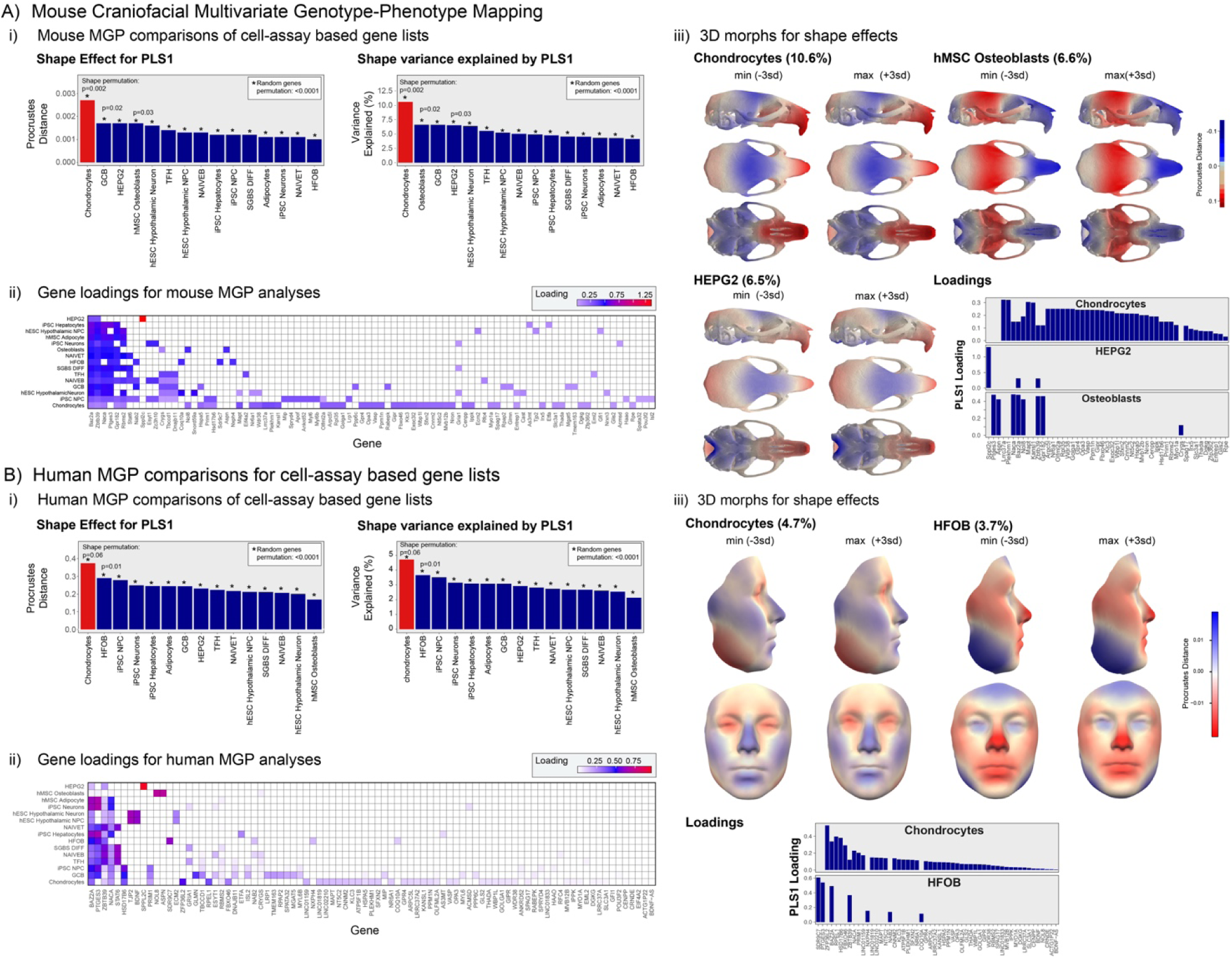
(A) Results of multivariate genotype-phenotype mapping for mouse craniofacial shape using the cell type specific lists from panel A. Ai shows the magnitude of shape effects and shape variance explained by the covariance between markers for each cell type specific list. The chondrocyte list results are indicated in red. Aii shows the gene loadings for all cell type lists. Aiii shows the craniofacial shape effects associated with the three lists that have significant effects as determined by the shape and gene list permutation tests as well as the gene loadings for these three lists on PLS1. These effects represent axes of variation in multivariate shape space associated with the conjoint effects of these genes. (B) shows the analogous multivariate-genotype-phenotype mapping for human craniofacial shape. Again, the chondrocyte list effects are shown in red. Also shown are the 3D morphs and heatmaps that correspond to these the chondrocyte and HFOB lists. Here, the chondrocyte list is the largest but marginally significant in terms of the shape permutation test. For all gene lists, the random gene permutation tests were significant, suggesting enrichment for genes associated with craniofacial shape effects.

The results of the human MGP analyses are generally consistent with the mouse MGP. All gene sets were significantly associated with facial shape when compared to random gene sets of the same length. Similar to the mouse MGP, the chondrocyte gene list explained the greatest proportion of variance in facial shape in height, weight and sex standardized data compared to other gene lists and PLS1 was suggestively associated with variation in craniofacial shape in the permutation test (*P* = 0.06) (**Figure 4B**, **Supplementary Table 19-20**). Additionally, the human fetal osteoblasts (HFOB) gene list was nominally associated with shape in the shape permutation test (*P* = 0.01). Inspection of the 3D morphs and heatmaps for facial shape reveal that this axis in the chondrocyte list is associated with variation in the width and projection of the midface and nose. HFOB is also associated with frontal bossing, which would be broadly homologous to the neurocranial vaulting seen in the mice for hMSC osteoblasts. As in data from mice, these results reveal that the chondrocyte and osteoblast specific gene lists are associated with craniofacial shape variation in humans that is consistent with a morphological role in the etiology of OSA because the direction of MGP effect involves changes in features that are thought to be associated with OSA risk, such as midfacial prognathism and width, as well as nasal projection.

#### eQTL Co-Localization

Genetic alterations may operate on OSA by affecting gene expression. Thus, we also performed colocalization analysis of the top GWAS association regions using the publicly available gene expression quantitative trait loci (eQTL) data from the GTEx project v7 derived from bulk tissue. Unfortunately, data from chondrocytes and other craniofacial-relevant tissues are not available for examination in these resources. Across the 20 regions examined across all traits, we identified significant evidence of colocalization for 14 unique genes within 10 association regions across four tissues relevant to OSA and obesity (**Tables 1** and **2**, **Supplementary Tables 21-22, Supplementary Figure 14**). The strongest colocalization signal observed (-log10*P*=8.35) was with *FTO* in skeletal muscle tissue (**Supplementary Figure 14A**). There was strong evidence for colocalizations for *PTBP2* in skeletal muscle tissue (-log10*P*=7.23), lung (-log10*P*=7.97), and visceral (-log10*P*=7.03) and subcutaneous (-log10*P*=8.00) adipose tissues (**Supplementary Figure 14B**). Also, *GRIA1* showed significant colocalization in lung tissue, with similar strength in association in GWAS and cis-eQTL analyses (**Supplementary Figure 14C**). Two loci showed multiple genes with evidence of eQTL colocalization, including the region around rs11642015 with expression of *FTO* and *RBL2* in skeletal muscle tissue and the region around rs10735928 with *LEMD3* expression in skeletal muscle and subcutaneous adipose tissues and *WIF1* in lung and visceral adipose tissues. Given previously observed enrichment of genes associated with BMI in the hippocampus and hypothalamus^19^, we included both tissues in our colocalization analyses; however, we did not identify any colocalizations for either brain tissue. Additionally, previous studies have noted generalization of eQTLs between disease relevant tissues and whole blood, for example Brotman et al found that ∼30% of adipose eQTL signals were also found in blood^94^; however, we did not find any colocalizations with whole blood.

## DISCUSSION

We conducted a global, multi-institutional GWAS meta-analysis to investigate the genetic basis of OSA, analyzing data from 492,107 individuals, including 46,028 OSA cases. We also examined quantitative sleep traits in up to 28,486 individuals. Given obesity’s strong link to OSA, we performed analyses with and without BMI adjustment^18^. Highlighting obesity-related pathways, our case-control GWAS identified 11 genome-wide significant loci without BMI adjustment, with only one locus remaining significant after adjustment. The strongest signal was at the *FTO* locus. After BMI adjustment, three additional loci emerged, likely reflecting non-obesity mechanisms such as differences in craniofacial structure^28–30^ or neural mechanisms controlling the upper airway and breathing^35^. The quantitative approach identified two associations with AHI, a measure of severity of OSA, and three with avgSaO_2_.

Beyond simply identifying genetic associations, a major goal of our study was to perform functional analyses and determine likely causative genes underlying association signals. For this, we employed several follow-up analyses, including V2G mapping, gene expression colocalization analysis, and MGP mapping, among others. In particular, using results of our V2G mapping, we evaluated genes acting through craniofacial development, particularly via chondrocyte-mediated growth at cranial synchondroses. Genes mapped in chondrocytes and osteoblasts were associated with craniofacial shape variation in both mice and humans, while genes from other cell types showed no robust shape associations, highlighting the importance of craniofacial pathways in risk for developing OSA.

### Overview of Discovery GWAS

Of the 14 loci identified across our analyses, 8 are newly identified associations for OSA, including an independent signal in a known region, near *NACA*. We validated 10 of our association signals using previously published GWAS data, and all 14 were directionally consistent. However, we replicated just over half of the 56 previously reported OSA loci. Differences in study design, population diversity, and phenotype definitions likely explain this, along with statistical phenomena like the Beavis^95^ effect and winner’s curse^96^. For example, the current sample is more diverse in study design and demographics compared to MVP [largely male] and FinnGen [exclusively European and Finnish] and, thus, the underlying genetic architecture of the populations may differ. Notably, of the 56 genome-wide significant hits from previous publications, only six (near *PTBP2*, *GAPVD1*, *PRIM1*, *NRXN3*, *FTO*, and *ZFP64*) were identified in both the MVP and FinnGen studies.

Overall, our results do not replicate previous associations for quantitative traits^37,38,54,55^; we find suggestive evidence supporting four loci for avgSaO_2_, with three robust to BMI adjustment. Lack of replication of newly identified loci in previous GWAS meta-analyses are likely due to smaller sample sizes, phenotype variability, and differences in OSA severity across studies. For example, previous analyses have utilized sleep study data from population-based cohorts with many fewer subjects with moderate/severe OSA^37^ than in the present analysis. Another explanation for lack of replication is that the night-to-night variability in quantitative measures of OSA severity introduces noise, particularly in those with mild disease^97^. There are also data suggesting OSA is a different entity in population samples versus clinical cohorts; in population studies OSA is common but there is a low symptom burden^40^. Finally, genetic associations may differ based on the extensively reported heterogeneity in the causes and consequences of OSA, which are not captured in a single severity measure such as the AHI.

### Potential Pleiotropy

Genetic risk loci for OSA-related traits can recapitulate expected phenotypic associations with comorbid traits. For example, genetic correlations highlight strong and significant shared genetic architecture in expected directions between OSA and other conditions like sleepiness, BMI and cardiometabolic traits. These relationships are mirrored in our PheWAS, which identifies significant associations between specific genetic variants and various clinical traits, confirming known links between OSA and conditions like obesity and atrial fibrillation. These results replicate the major findings of the GWAS study conducted in FinnGen and observed genetic correlations with similar cardiometabolic traits^98^. Our top variant near *GRIA1* has been associated with frequency of tiredness and lethargy among other related traits in the UKBiobank PheWeb, including overall health rating, waist circumference, weight and body fat percentage^99,100^, as well as combined sleep disorders, depression, radiculopathy, and weight in the FinnGen Release 12 PheWAS^101^. The top variant that is intronic to *MSRB3* has also been associated with snoring, height, bone mineral density, and forced expiratory volume (FEV1)^99,102^ in the UK Biobank, and hernia, diverticular disease of intestine, sleep apnea and combined sleep disorders in FinnGen Release 12 PheWAS^101^ . The *PTPB2* variant is also associated with daytime nap and multiple measures of weight and body fat in the UK BioBank^99,103^. These findings reinforce established clinical links between OSA and comorbid traits and while revealing potential new associations, thus highlighting common biological pathways contributing to sleep and related conditions.

### Overlap with Obesity/BMI Loci

The relationship between sleep and obesity is well established, and obesity is a primary risk factor OSA. Obesity can lead to changes in body morphology that cause restriction to air flow ^55,104,105^. Sleep can also increase risk for obesity through reduction in energy output. They both can cause changes in hormonal regulation that can lead to alterations in appetite and satiety and also affect how we process and store energy yielding nutrients^105^. Given this, it may be unsurprising that obesity-risk variants would be identified in our OSA analyses without controlling for BMI.

Largely, BMI risk loci are enriched for genes expressed in neuronal tissue that relate to appetite regulation^19^. As an example, we identify an OSA association signal with lead variant rs11642015 in the well-known *FTO* (Fat Mass and Obesity-Associated Protein) obesity locus. Associations in this region have been observed for many traits for which elevated BMI is a risk factor. There have been multiple independent signals and causal variants identified in this gene, including likely causal variants that operate over long genomic distances on *IRX3* (Iroquois Homeobox 3) and *IRX5* (Iroquois Homeobox 5), which are involved in neuronal development, highly expressed in the hypothalamus, and involved in regulating eating behavior and energy metabolism^106^. Additionally, rs1800437 is in an intron of *GIPR* (Gastric Inhibitory Polypeptide Receptor) and is nearby associations include BMI^107,108^, obesity^109^, heart disease^110,111^, central obesity^23^, T2D^112^ and other cardiometabolic traits^61,113,114^. *GIPR* is essential for glucose homeostasis and a receptor for gastric inhibitory polypeptide (GIP), which is essential for appetite regulation as well^115^. Some rare variants in *GIPR* seem protective against obesity^107^. There is no clear evidence that *DNAH8* (Dynein Axonemal Heavy Chain 8) at the rs9380814 locus is functionally related to sleep or obesity. However, a nearby gene, *GLP1R* (glucagon like peptide 1 receptor; a target for well-known class of obesity and T2D drugs with high efficacy, i.e. semaglutide, tirzepatide, liraglutide)^116^ is prioritized by Open Targets through functionally-informed fine-mapping and evidence that this lead variant is in cis-eQTL for *GLP1R*^68^. Indeed, tirzepatide was recently approved by the Food and Drug Administration (FDA) for treatment of sleep apnea^117^.

However, other BMI-associated regions we find associated with OSA are nearby genes largely operating through non-appetite regulation pathways and may indicate an “OSA-first” relationship to obesity. For example, our lead SNP on chromosome 21, rs11088984 is nearest to *POFUT2*, the next closest gene is *ADARB1* (Adenosine Deaminase RNA Specific B1), which is implicated at this locus by our gene-based analysis. *ADARB1* acts to edit mRNA, including at the translation step. One of its target proteins is GABRA3 (Gamma-Aminobutyric Acid Type A Receptor Subunit Alpha3), which is a target for drugs to treat insomnia. A recent study found that knockdowns of *GABRA3*, while causing a hypnotic state, decreased deep sleep in mice^118^. Additionally, rs9831061 is near *ETV5* (ETS Variant Transcription Factor 5), a well-known BMI and bipolar disorder locus.

A recent study shows that *Drosophila* with knockdown of this gene have high activity, increased startle, and reduced nighttime sleep when knocked down during the developmental stage^119^.

Interestingly, while there have been over a thousand obesity risk loci identified, we find only 10 of these loci are associated with OSA in our well-powered GWAS. This suggests that only certain obesity-related pathways may be relevant for OSA risk (e.g., there is a specific role for fat in the tongue^26,27,31^). As noted above, many of these operate on pathways outside of appetite regulation and seem to have a direct impact on sleep disruption. This is mirrored in a locus not previously associated with BMI; rs2964006, which is associated with OSAadjBMI, is nearest to *GRIA1* (Glutamate Ionotropic Receptor AMPA Type Subunit 1). *GRIA1* was implicated in both our variant to gene mapping analyses and eQTL colocalization. Loss of function mutations in this gene cause rare neurodevelopmental syndromic disorders, in part, characterized by poor sleep^120,121^. Also, mouse knockdowns display sleep pattern disruption and mouse models indicate an important role for this gene in regulating circadian rhythm^122,123^. Other nearby phenotypic associations include schizophrenia^123,124^.

### Gene Prioritization

While GWAS reveals genomic loci associated with OSA related traits, identifying causal variants within these loci is complicated by LD that results in a strong correlation among all genetic variants within a given locus. Adding to the challenge of interpreting GWAS findings, most of the identified variants are found in non-coding regions of the genome that are enriched with regulatory elements, suggesting that these variants likely influence gene expression rather than alter protein structure or function. However, our diverse approaches provide strong support for several genes underlying our association regions. For example, the nearest gene to the top variant in our OSA locus on chromosome 1 is *PTPB2* (polypyrimidine tract-binding protein 2), which is implicated in the gene-based analysis and the colocalization analysis in skeletal muscle, lung, visceral adipose, and subcutaneous adipose tissues. Additionally, *MSRB3* (Methionine Sulfoxide Reductase B3) is the nearest gene to the lead SNP, rs10735928, associated with OSAadjBMI on chromosome 12. Further support for this gene came from our gene-based test and Open Targets. A nearby variant intronic to *MSRB3* in high LD with our top variant and within the top five fine-mapped SNPs, rs10784447, exhibits a high CADD score (CADD phred = 19.64), indicating the likelihood that this variant is deleterious.

### Genetic Risk Operates in Part Through Craniofacial Modifications

Our validation via physical V2G mapping and the subsequent tests for craniofacial shape effects via MGP mapping primarily identified genes implicated in chondrocytes. Smaller but significant effects were seen in genes implicated in the two osteoblast-derived cell lines in both mice and humans and in the human liver cancer cell line in mice. In all cases, the shape axes are similar, although the direction of effect is reversed between chondrocytes and osteoblasts. Cartilage development is pivotal in determining overall craniofacial form, including dimensions related to OSA risk.^82^ Cartilage also affects aspects of craniofacial development via interactions between endochondral elements such as Meckel’s cartilage in the mandible and the dermatocranial structures that form around them^125^. Mouse mutants with reduced proliferation, maturation or altered cell death in synchondroses exhibit a characteristic set of phenotypic effects including a more flexed cranial base, reduced facial length and neurocranial vaulting^78,79,126,127^. It is intriguing, therefore, that the MGP analysis of the OSA risk variants implicated in chondrocytes revealed an axis of craniofacial variation that resembles effects on the pharynx that may relate to OSA risk. In mice this involves projection of the face, basicranial length and flexion, while in humans this involves midfacial prognathism and nasal projection. All of these morphological effects are highly likely to correlate with key dimensions of the nasopharynx. This finding strongly suggests that OSA-risk variants implicated in chondrocytes produce their effects via changes to craniofacial shape mediated by influence on growth at the cranial synchondroses or other cartilaginous components of the developing skull.

The use of MGP mapping as a form of validation for a GWAS study is novel. This approach is ideally suited to studies of highly polygenic traits, as MGP allows for quantitative tests of hypotheses that relate variants in multiple genes to a complex phenotypes. Here, we have shown how the combination of V2G and MGP mappings can facilitate identifying the likely cellular and phenotypic pathways through which variants identified in GWAS convey their effects on disease risk. In this way, the MGP offers an *in-silico* approach to validation that is significantly faster and cheaper than generation of multiple mouse models or even *in situs* that establish plausible localization of effects.

### Limitations and Future Directions

Our study has limitations, most notably with regards to the quantitative association analyses. We did not have access to raw sleep study data for centralized rescoring, and instead relied on sleep study reports from experienced, accredited sites. To mitigate potential biases introduced by differences in sleep study technologies or clinical scoring methods across the included datasets, analyses were either performed separately by site or included study site as a covariate, and rigorous meta-analytic methods were used to combine results, including a two-stage meta-analysis approach. While differences in the definitions of hypopneas could lead to less severe (when using a 4% oxygen desaturation criterion) or more severe (when a 3% desaturation and/or arousal criterion) AHI, we would anticipate that genetic variants relevant for OSA severity would show consistent directions of effect for either definition (particularly when controlling for site). As discussed in this paper, these sources of variance in defining quantitative measures of OSA severity (as well as night-to-night variability within patients) likely impact on the ability to identify reproducible SNP associations with quantitative traits such as AHI.

Further, while we followed a robust two-stage approach to limit spurious associations and provide strong biological support for our findings through downstream analyses, none of our quantitative trait lead variants and three of our case-control lead variants did not generalize on previous GWAS of similar sleep traits. This is not surprising given the differences in study designs between the current GWAS and previous studies, including differences in phenotype definition, ancestral background of participants, race/ethnic diversity of participants, differences in allele frequency, imputation reference panels, etc. Nonetheless, these particular association signals may need to be interpreted with some caution.

Studies that interrogate genetic risk across a range of populations are critical to gain a comprehensive understanding of the genetic architecture of OSA risk. As noted previously, White/European individuals represented 81-92% of our total sample size, and thus this effort focused on pooled, cross-population analyses and White/European individuals only. Our analytical design allowed for heterogenous effects across population groups in pooled analyses to improve power, yet future studies with greater participant diversity and sample size are needed to identify potential population-specific genetic contributors to OSA phenotype heterogeneity and risk.

Lastly, our functional follow-up analyses including V2G mapping and MGP analyses are a strength to the current study; however, they are informed by a limited number of cell types. While we have a strong rationale for the inclusion of cells types (i.e. neuronal, osteoblasts, etc.), additional cell types, such as neural crest cells, could reveal other relevant genes^128^. For example, our focus on chondrocytes and osteoblasts primarily captures post-differentiation gene transcription activity, which does not account for abnormalities in the migration or specification of mesenchymal progenitor cells. Such early developmental processes can affect both cell number and location prior to differentiation and can be early drivers of craniofacial and neuronal determinants of OSA.^129–131^ Indeed, gene activity in these cells may have indirect roles in regulating central respiration. Yet, key neural crest genes, such as *IRX5*, which is associated with Hamamy syndrome and craniofacial anomalies including midface bulging and micrognathia^132,133^, were still represented in our dataset. Future studies incorporating neural crests and other cell types relevant to early developmental stages will be essential to fully characterize gene pathways influencing OSA risk.

### Strengths of This Study

This study has several key strengths. We assembled a large, globally diverse cohort of OSA cases and controls to conduct meta-analyses and comprehensive follow-up investigations of identified OSA risk loci. While smaller than the recent MVP^13^ study, the large cohort we assembled is more diverse. We identified novel genetic associations with OSA and replicated previously reported loci^13–15^. Our study adds further support to the concept that there are both obesity and non-obesity genetic pathways to disease, each with multiple genes involved.

A major strength is our integrative approach to move from association to function. Recognizing that the nearest gene to a variant is not always causal, we used multiple methods to pinpoint likely causal variants and genes, most notably chromatin accessibility and contact data across cell types to map variants to likely effector genes^100,134^. Given the role of craniofacial structure in OSA risk, we specifically explored variants acting in chondrocytes, key cells in craniofacial development, which are absent in publicly available GTEx resources^16^. Using the integration of genome-wide chromatin accessibility/contact and chromatin conformation capture data for V2G mapping and an MGP mapping tool, we found that genes in our identified loci that mapped in chondrocytes likely influence OSA risk through chondrocyte-mediated craniofacial development. To our knowledge, this is the first application of this approach in OSA genetics.

## CONCLUSIONS

This study assembled a diverse cohort from multiple sources worldwide, identified significant genetic loci associated with OSA, validated several previous findings, and highlighted the complexity of genetic determinants influenced by obesity and craniofacial structure. Several OSA-associated loci overlap with BMI-associated regions, indicating shared genetic pathways. Yet, our *in silico* interrogation of significant GWAS loci highlights both obesity and non-obesity genetic pathways to OSA, using multiple strategies to identify causative genes and their mechanisms. We further provide functional evidence for non-obesity-related pathways to OSA, including multiple state-of-the-art analyses showing that some genes related to OSA risk likely convey their effect via chondrocyte-mediated craniofacial development. This approach to assessing craniofacial dimensions via gene mapping is a significant contribution to the understanding of OSA genetics. The findings underscore the importance of considering diverse populations and methodologies in genetic research on OSA. Future work should involve validation of these and previous findings, and leverage newly discovered loci to predict increased risk for development of OSA to inform prevention and treatment strategies.

## MATERIALS AND METHODS

### Study Design

We used a two-staged approach to identify loci associated with OSA (N = 492,107), AHI (N = 28,486), and avgSaO_2_ (N = 6,908), whereby we selected single nucleotide polymorphisms (SNPs) that reached a suggestive statistical significance threshold (*P* < 5x10^-5^) in either stage to bring forward to test for genome-wide significance (*P* < 5x10^-8^) in a joint stage 1 plus stage 2 (S1S2) meta-analysis (**Supplementary Table 1** and **2**, **Supplementary Figure 1**). Joint analyses of two-stage approaches have been shown to maximize power to detect associations while maintaining control of Type I error compared to traditional one-stage or discovery and replication studies^102,103^. As obesity is a major risk factor for OSA^135^, we conducted association analyses without and with adjustment for body mass index (BMI) to identify genetic risk loci independent of BMI. Our stage 1 meta-analysis included up to 161,632 individuals for OSA (case/control), 9,120 for AHI, and 4,758 individuals for avgSaO_2_ from two biobanks linked with electronic health records (EHR), Geisinger’s MyCode Community Health Initiative Study (MyCode) and the Electronic Medical Records and Genomics (eMERGE) Network (**Supplementary Table 1**). Our stage 2 meta-analysis included up to 330,475 individuals for OSA, 19,366 for AHI, and 2,150 for avgSaO_2_ from six data sources, the Penn Medicine Biobank (PMBB), Vanderbilt University’s BioVU (BioVU), Kaiser Permanente Southern California (KPSC), deCODE genetics (deCODE), the Korean Genome and Epidemiology Study (KoGES), and the Sleep Apnea Global Interdisciplinary Consortium (SAGIC). Additionally, our analyses were conducted both pooled across all race and ethnic groups and in White/European individuals only, as this group represented 81-92% of our total sample size, depending on the trait. Results from our final S1S2 meta-analyses are leveraged in all secondary analyses, unless otherwise noted.

### Phenotype Definitions

#### OSA

To identify adult (18 to 89 years) OSA cases and controls, we utilized an EHR-based case identification algorithm previously validated through chart review^42^. Specifically, we required ≥2 instances of unique encounters of OSA-related ICD-9 (327.20, 327.23, 327.29, 780.51, 780.53, 780.57) or ICD-10 (G4730, G4733 or G4739) diagnostic codes for OSA cases and zero instances of any OSA-related diagnosis code for controls. For the MyCode data, we required ≥3 instances of an OSA-related diagnosis code to maintain high accuracy, as described previously^42^.

#### Apnea-Hypopnea Index (AHI)

The apnea-hypopnea index (AHI) was obtained from clinical sleep study reports or through information in the EHR; the first instance of a diagnostic sleep study was used for individuals with multiple studies. Hypopnea scoring was based on either a 4% or on a 3% and arousal hypopnea criterion according to the clinically utilized criteria at the time of the study. To mitigate potential biases introduced by differences in sleep study technologies or clinical scoring methods across the included datasets, analyses were either performed separately by site or included study site as a covariate (depending on the dataset), with meta-analytic approaches used to combine results. Values recorded from full night or unlabeled sleep studies with <120 minutes of total sleep (for in-laboratory studies) or analysis time (for home sleep tests) or from split night studies with <60 minutes of total sleep time were excluded as unreliable.

#### Average Oxygen Saturation (avgSaO_2_)

The average oxygen saturation (avgSaO_2_) value was obtained from clinical sleep study reports or through information available in the EHR. Any values recorded as <50% or >100% were excluded as likely invalid data during the quality assurance process.

### Study-level Genetic Analyses and QC

All study-specific GWAS for OSA and quantitative measures of OSA severity were adjusted for age (age at diagnosis for OSA cases, most recent age for OSA controls, age at sleep study for AHI and avgSaO_2_), sex, principal components (PCs) calculated from genome-wide array data to control for population stratification, study-specific covariates (e.g. clinical trial arm, study center, genotyping array), and race/ethnicity in OSA analyses. For Model 2, analyses were further adjusted for BMI (kg/m^2^) at diagnosis for cases and most recent BMI for controls. Individuals with extreme or unrealistic BMI values were excluded based on study distribution. In the deCode dataset, logistic regression was used to test for association between variants and OSA, assuming a multiplicative model, treating disease status as the response and expected genotype counts from imputation as covariates. The association was adjusted for sex, county of origin, current age or age at death, blood sample availability for the individual, and an indicator function for the overlap of the lifetime of the individual with the time-span of phenotype collection. Testing was performed using the likelihood ratio statistic^136^. A linear mixed model implemented in BOLT-LMM^137^ was used to test for association between sequence variants and AHI, assumed an additive genetic model. The trait values were adjusted for age, sex, year of birth and county of origin, and inverse-normal transformed, prior to the association test. All other studies performed genetic association analyses using SUGEN^138^ assuming an additive genetic model, accounting for relatedness when appropriate, and allowing for heterogeneity of effect by race/ethnicity for quantitative traits. GWAS results underwent centralized quality control filters, including removing invalid values (standard errors [SE]<0, P-values>1), missing values (P, Beta, SE, effect allele frequency [EAF], or sample size [N]), minor allele count (MAC)≤5, duplicate SNPs, and SNPs with N<30. Additionally, studies were examined for signs of systematic errors following standard protocols using the R package EasyQC^139^.

### Meta-analyses

Study-specific GWAS summary statistics were meta-analyzed using a fixed-effect inverse variance weighted meta-analysis in METAL.^140^ All study-specific GWAS were genomic-control (GC)-corrected at the meta-analysis stage. Additionally, S1S2 meta-analyses underwent a second GC-correction for the multi-population meta-analysis for avgSaO_2_ only. Each SNP from the meta-analysis results was filtered for minor allele frequency (MAF) greater than 0.5%.

### Conditional Analysis

To determine if independent association signals were present within our associated loci, we used GCTA^141^ to perform approximate joint conditional analyses on the S1S2 meta-analysis results for each model. Given that a majority of our participants were White/European individuals, we used a subset of unrelated MyCode study participants of genetically-informed European similarity (N=47,177) with genetic data imputed to the 1000 Genomes Phase III global reference panel to calculate linkage disequilibrium (LD). European ancestry was ascertained using genetic data as previously described^142^. We selected all SNPs within ±500 kb from our top associated variant in each locus. We considered significant secondary signals as those with a P_Conditional_<1x10^-5^ and for which β_Conditional_ was not attenuated (decreased by less than 10% from the original β).

Additionally, many of the association signals identified in the current analyses are nearby SNPs previously associated with OSA. For each known locus, GCTA was also used to perform approximate conditional analyses on the OSA S1S2 summary statistics to determine if the loci identified here are independent of the previously identified associations. We considered our signals independent from known signals if the P_Conditional_<1x10^-5^ and β_Conditional_ was not attenuated (decreased by less than 10% from the original β).

### Fine-Mapping

We used FINEMAP^143^ software to identify credible sets of likely causal variant SNPs within our significantly associated regions. We conducted fine-mapping using the shotgun stochastic search (SSS) method. We used summary statistics from our S1S2 meta-analysis results and considered all variants within ±500 kb of our lead variants. We used the same LD reference files as for conditional analyses (unrelated MyCode study participants of genetically-informed European similarity [N=47,177]). Genetic similarity was determined based on k-means clustering of genome-wide PCs with reference populations from the 1000 Genomes population grouped by major geographic regions (i.e. EUR, AFR, EAS, AMR, SAS), as previously described^144^. We assumed a single causal variant for all loci, except one locus with evidence of a secondary signal, for which we assumed two causal SNPs.

### Look-ups of Known OSA loci

To assess generalizability of previously reported associations for OSA, AHI and avgSaO_2_ to our study findings, we conducted lookups of previously published GWAS findings within our S1S2 meta-analyses. We identified a total of 42 loci associated with OSA with or without BMI adjustment reported by Strausz *et al* 2021^15^ or Sofer *et al* 2023^13^, which included both OSA and OSAadjBMI, and 2 AHI-associated loci and 47 avgSaO_2_ reported in the GWAS Catalog available in our current study^38,39,54,145^.

### Look-ups in published sleep-related trait GWAS

We conducted lookups of all of our top associated SNPs in previous GWAS of similar traits. For OSA, we used published GWAS conducted in the Million Veteran’s Program (MVP)^13^ and FinnGen^101^. For quantitative traits, we queried the Sleep Disorder Knowledge Portal, which contains summary data for AHI and avgSaO_2_ GWAS from the Trans-Omics for Precision Medicine (TOPMed) Program^38,39,54,145^.

### Gene-Based Association Analysis

We used FUMAv1.5.2^66^ to implement MAGMA (v1.08) gene-based association analyses. These analyses were performed using the S1S2 meta-analysis summary statistics. Default settings were used in the FUMA analyses. Of note, FUMA performs a series of analyses in addition to gene-based association testing. These full results are available to the public (see Data Availability Statement).

### Functional Mapping and Annotation

Additionally, to gain a better understanding of the potential functional consequence of the genetic variants prioritized in fine-mapping analyses, we used NCBI Ensembl’s Variant Effect Prediction (VEP, v 113.0) tool^67^ to annotate variants with the highest posterior probabilities (PP) of causality. We selected up to five variants within the 95% credible interval with the highest PP. In addition to the default annotation, we included the following plugins available in the public VEP Application Programming Interface (API): CADD^77^ (combined annotation dependent depletion) scores, 1000 Genomes reference population allele frequencies, REVEL (Rare Exome Variant Ensemble Learner) scores^146^, OpenTargets^68^, Gene Ontology (GO)^147,148^, Enformer^149^, AlphaMissense^150^, SpliceAI^151^ scores, ClinPred^152^ , and SIFT^153^ and PolyPhen^154^ predictions.

### Genetic Correlation

To confirm expected direction and strength of shared genetic architecture among OSA, other sleep traits, and obesity, we used LDSC^155^ to estimate genetic correlation across all of the traits included here and with related traits using publicly available GWAS summary data for other sleep traits (insomnia^56^, and sleepiness without and with adjustment for BMI^57^), obesity traits (body mass index [BMI]^20^, visceral adipose tissue [VAT], subcutaneous adipose tissue [SAT], pancreatic volume, pancreatic fat, liver fat^58^, hip circumference adjusted for BMI [HIPadjBMI], waist circumference adjusted for BMI [WCadjBMI]^22^, waist to hip ratio adjusted for BMI [WHRadjBMI]^23^), and cardiometabolic traits (type II diabetes [T2D]^59^, random glucose [RG]^60^, pulse pressure [PuP], systolic blood pressure [SBP], diastolic blood pressure [DBP]^61^, triglycerides [TG], total cholesterol [TC], high-density lipoprotein cholesterol [HDL], low-density lipoprotein cholesterol [LDL]^62^, coronary artery disease [CAD]^63^). Given that our discovery studies and the sample population of our comparative GWAS were primarily White/European descent participants, we used the precomputed publicly available LD scores for European ancestry individuals from the 1000 Genomes Project available at https://alkesgroup.broadinstitute.org/LDSCORE/. The precomputed LD scores cover ∼1.2 million HapMap3 variants that are generally well imputed across studies and populations.

### Phenome-wide Association Study (PheWAS)

To identify underappreciated and known comorbidities with shared genetic underpinnings, we performed a phenome-wide association (PheWAS) for each of our top associated index SNPs. For this analysis, we restricted to an unrelated subset of individuals in the MyCode study population (N = 108,369; Females = 64,568; Males = 43,801), including only one member from each family network based on 2^nd^ degree family networks reconstructed using PRIMUS^156^. Two of our lead variants, rs1200596362 and rs802729, were not available for PheWAS analyses in MyCode. Standardized phenotypes (PheCodes) were derived from ICD-9 and ICD-10 codes based on the PheCode Map v1.2^157^. We restricted to patient visits from all emergency departments, inpatient, outpatient, and tele-medicine appointments. PheCode cases were defined by a minimum of two encounters and PheCode controls were those with zero instances of a relevant PheCode. We excluded PheCodes with less than 20 cases and 20 controls from these analyses. A total of 1,722 PheCodes were included following mapping and exclusions. Association analyses were carried out via the R package PheWAS using logistic regression and adjusting for most recent encounter age, gender (for non-gender specific PheCodes), genotyping array, EHR-reported race/ethnicity, and the first twenty principal components from genome-wide data. Significant PheCode associations are determined based on a P<2.9x10^-5^ (equals 0.05/1,722 PheCodes tested) and suggestive significance as P<2.9x10^-4^.

### Chromatin Accessibility

Chromatin accessibility data were used for prioritizing potential effector genes at OSA-associated loci for variant to gene (V2G) mapping. We used a set of previously generated ATAC-seq and Capture C/Hi-C libraries across multiple cell types, including models representing:

- **Skeletal System:** MSC-derived osteoblasts^158^, a human fetal osteoblast cell line (hFOBs)^159^, and a novel data resource from primary chondrocytes derived from healthy donors (See **Supplementary Note 1** for details on chondrocyte cell preparation, ATAC-seq, etc.);
- **Metabolic tissues**: iPSC Heptocytes^160,161^ and HepG2 (Hepatocarcinoma cell line)^160^, MSC-derived adipocytes^43^, a preadipocytes cell line (SGBS)^158^;
- **Immune cells:** Naive T, T-Follicular Helper Cells (TFH), Naive B cells, and Germinal Center B cells (GCB) derived from Tonsil^162,163^; and
- **Neuronal cells:** ESC derived hypothalamic Progenitors/Neurons^164^, iPSC derived cortical derived progenitors, and neurons^165^.

Details on chromatin accessibility data generation have been previously described^43,158–165^ or are provided in **Supplementary Note 1** for the novel data resource from primary chondrocytes. A brief description of data processing are provided below.

#### ATAC-seq peak processing

Following library preparation, Fastq files of technical replicates were merged and ATAC-seq peaks were called following the ENCODE ATAC-seq pipeline (https://www.encodeproject.org/atac-seq/). Pair-end reads from all replicates for each cell type were aligned to the hg19 or hg38 human genome builds using bowtie2, and duplicate reads were removed from the alignment. Narrow peaks were called independently for pooled replicates for each cell type using macs2 and ENCODE blacklist regions were removed from called peaks. Finally, a consensus of open chromatin regions (OCRs) called in the majority (>50%) of the replicates were obtained by consolidating the peak sets across samples using bedtools intersect (v2.25.0)^76^.

#### Hi-C data processing

Paired-end reads from three biological replicates with two technical replicates each were pre-processed using the HICUP pipeline^166^, with bowtie as aligner and hg38 as the reference genome. The alignment .bam file were parsed to .pairs format using pairtools v0.3.0^167^ and pairix v0.3.7^168^, and eventually converted to pre-binned Hi-C matrix in .cool format by cooler v0.8.10 with multiple resolutions (1kbp, 2kbp, 4kbp ) and normalized with the ICE method^169^. Finally, for each cell type, significant intra-chromosomal interaction loops were determined under multiple resolutions (1kb, 2kb and 4kb) using the Hi-C loop callers mustache v1.0.1 (q-value < 0.1)^170^ and Fit-Hi-C2^171^ v2.0.7 (FDR < 1e-6) on merged replicates matrix. The called loops were merged between both callers to represent cell type loops at each resolution.

#### Variant-to-gene (V2G) mapping

For V2G mapping analyses, we curated lead SNPs from the current study (**Supplementary Tables 3**, **7**, and **8**) and from the recent Million Veterans Program (MVP) OSA GWAS^13^. Proxy variants were then expanded to those in high LD identified from the TOPMed LD reference panel (R^2^ > 0.80)^172^. We identified the subset of variants located in open chromatin and in contact with gene promoters via our ATAC-seq and Capture C/Hi-C data using GenomicRanges (1.50.2) in R (v4.2.3). Resulting gene implications per cell type were visualized using Cytoscape (v.3.10.1)^173^, using complete linkage clustering with the Euclidean distance. The resulting nominated genes per cell type were then selected for additional validation.

### eQTL Co-localization

To narrow in on potential candidate genes and explore the possibility that our GWAS association signals are operating on sleep phenotypes through changes in gene expression, we conducted colocalization analyses. We implemented the simple sums (SS2) colocalization method^174^ using the web-based LocusFocus v.1.6.0 tool^71^ to test for colocalization between GWAS and cis-eQTL signals. We used summary statistics from the S1S2 meta-analysis, including the ±500 kb region surrounding each index SNP. We used the publicly available cis-eQTL data from the GTEx project v7^175^, including expression data from whole blood, skeletal muscle, lung, visceral adipose tissue, subcutaneous adipose tissue, hippocampus, and hypothalamus tissue. We included all protein coding genes within ±500 kb of our leads SNPs for an identified locus. 1000 Genomes EUR samples were used for calculation of LD. We used the default first-stage filter of eQTL results based on a locus-specific Bonferroni-corrected significance threshold (0.05/[number tissues x number of genes]). All gene and tissue combinations with results passing this first stage filter were then used for colocalization. Final determination of significant GWAS eQTL colocalization was determined based on a locus-specific Bonferroni P<0.05/([number of tissues x number of genes]) that passed the first step filter.

### Multivariate Genotype-Phenotype Mapping

As a further validation step, we performed multivariate genotype-phenotype (MGP) mapping for mouse craniofacial and human facial shape^49,50,176^ based on the putative effector genes identified via the cell-type specific assays described above (**see Chromatin Accessibility**). MGP mapping tests for the conjoint effects of a list of genomic variants on a multivariate phenotype^49^. The method uses a two-block partial-least-squares (PLS) ordination to extract latent variables that connect multivariate measures of phenotypic variation to a set of genomic markers as a multivariate dataset (e.g., that explain the covariance between as set of genomic markers and a multivariate phenotype). Each analysis returns a set of PLS axes ordered by the covariance between the two blocks explained, where each PLS axis captures the conjoint effect of multiple genomic variants. These factors represent hypotheses about the combined effects of genes rather than sums of individual gene effects, as in the case of GWAS or polygenic risk scores. Genomic variants with large phenotypic effects will not tend to converge on an MGP effect if their individual effects are in different directions in multivariate phenotype space. However, when genomic variants converge on a common set of directions in multivariate phenotype space, then their conjoint effects will result in a latent variable that is significantly associated with phenotypic variation. The individual effects of genomic variants on PLS axes are quantified as the PLS loadings. The phenotypic effect associated with a PLS axis can be visualized by using the PLS coefficients to project multivariate phenotypic values along each PLS axis. Like any multivariate ordination method, MGP analyses yield multiple loadings ordered by variance explained.

We quantified mouse craniofacial shape from microCT scans of 1266 diversity outbred (DO) mice that subjected to a nonlinear-registration based automated phenotyping pipeline^177^. We used a sparse set of 95 3D landmarks that were obtained via an automated landmarking procedure in which landmarks placed on an atlas are propagated throughout the full set of images with individual landmark placement refined via a machine-learning step based on a large manually landmarked training set^178,179^. Also, we conducted an analysis for the human MGP, whereby we used similar methods as for the established mouse model applied to 3D facial images from GWAS studies of facial shape in North Americans of European descent^180^ (N=2700) and Tanzanian (N=3600) children^181^. Facial shape is quantified from these images via dense representation (5k) landmarking based on the MeshMonk automated registration and phenotyping pipeline used for previous work with these datasets^182,183^. The dense landmarks data are then reduced via principal components analysis. For both datasets, the phenotype matrices consist of the full set of (X, Y, Z) landmark coordinates. We standardized the mouse landmark data for sex and centroid size by mean centering the residuals of a linear regression for centroid size with sex as a factor. We used height, age and sex to standardize the human landmark data in the same way, but used a three term polynomial regression for age due to its nonlinear relationship to facial shape, as in prior work^184^. For human facial data, we also remove the shape variation associated with the first principal component as the variation on this axis is primarily allometric, or associated with size.

To obtain genomic data for the DO mice, we imputed between the Giga16 (177k) and MegaMUGA (77k) arrays using the “calc_genoprob” function in the qtl2 package^185^. This was necessary as some generations had been genotyped with the smaller MUGA array. The genotype block is composed of the full set of DO founder probabilities for each selected marker. A region of chromosome 2 (73.25–124.85 Mb) was removed due to a known meiotic drive effect^186^ resulting in a final number of 120,941 SNPs. . The human genomic data for both datasets is a 12.5M imputed SNP array. We performed a PCA for the SNPs that mapped to each gene and used the PC composite scores to create the genotype matrix.

For both datasets we used PLS to decompose the cross-covariance matrix between the phenotype and genotype matrices. We test for the significance of these PLS regressions in two ways. The shape permutation tests shuffles genotypes with respect to the phenotype matrix. The second permutation test compares the magnitude of the observed phenotypic effect to random gene lists of the same length. These tests establish the significance of the magnitude and direction of phenotypic effect associated with each PLS axis. The phenotypic variation associated with each PLS axis can be visualized by projecting fitted shapes along each axis using the regression coefficients. Notably, the direction along the axis is not meaningful with respect to the GWAS data on which these analyses are based. In other words, the MGP analysis will not indicate which extreme along this axis might be associated with the disease phenotype of interest. However, the shape axis itself is meaningful. These analyses provide a form of validation for the GWAS results as they establish whether the gene lists are associated with a direction and magnitude of phenotypic variation in multivariate phenotype space that is consistent with the biological hypothesis. Here, we specifically test whether any of the cell type specific gene lists are associated with changes in midfacial prognathism and basicranial shape and length that would be consistent with morphological variation that would affect the shape and size of the pharynx as well as the oral and nasal cavities, thereby influencing OSA risk.

## DATA AVAILABILITY

### MyCode Community Health Initiative (MyCode)

MyCode data can be accessed by Geisinger investigators. There are restrictions to the sharing of MyCode DiscovEHR genetic datasets related to agreements between Geisinger and the Regeneron Genetics Center. Requests can be made by contacting the corresponding author.

### Electronic Medical Records and Genomics (eMERGE)

Data for eMERGE III are publicly accessible through the database of Genotypes and Phenotypes (dbGaP), accession numbers phs001584.v2.p2, pht010214.v1.p2, pht009069.v2.p2, pht009070.v2.p2, pht009071.v2.p2, pht009073.v2.p2, and pht010215.v1.p2.

### Amgen deCODE Genetics (deCODE study)

The sequence variants passing GATK filters in our previously described Icelandic population whole-genome sequence data have been deposited at the European Variant Archive under accession code PRJEB15197. Individual level phenotype and genotype data are protected and cannot be shared according to Icelandic law. De-identified GWAS summary statistics included in this study may be made available upon reasonable request. Data access requests should be directed to.

### Penn Medicine BioBank (PMBB)

Individual-level genotype and phenotype data from the PMBB are not publicly available due to privacy concerns. Data and specimens are available to investigators affiliated with Penn Medicine and to external collaborators via scientific collaboration with identified local Penn investigators.^187^

### Korean Genome and Epidemiology Study (KOGES)

Genotyping and cohort data used for this study are available from the National Biobank of Korea (https://biobank.nih.go.kr/eng/) and are managed by the National Institute of Health, Republic of Korea. Data access requests should be directed to. Upon reasonable request, de-identified summary data and results may be shared following approval by the Distribution Review Board in National Institute of Health, Republic of Korea.

### Sleep Apnea Global Interdisciplinary Consortium (SAGIC)

Data access is available to qualified researchers upon request and in accordance with site-specific informed consent. Requests should be directed to Allan Pack and are reviewed for approval by the SAGIC site PIs and Executive Committee.

The following publicly available datasets and pipelines were used, and, when available, web access to results are provided below:

- Functional Mapping and Annotation of Genome-Wide Association Studies (FUMA GWAS) - https://fuma.ctglab.nl/.
- Variant Effect Predictor (VEP) Web Interface - https://useast.ensembl.org/Tools/VEP. Full annotation results are available here

(OSA: https://www.ensembl.org/Multi/Tools/VEP/Ticket?tl=IlOT9OxRF5CxFhRY;

OSAadjBMI: https://www.ensembl.org/Multi/Tools/VEP/Ticket?tl=6sNaSIcx5mDgBm4s;

AHI: https://www.ensembl.org/Multi/Tools/VEP/Ticket?tl=IlOT9OxRF5CxFhRY,

AHIadjBMI: https://www.ensembl.org/Multi/Tools/VEP/Ticket?tl=s2YINiK9dG0W11f9,

avgSaO2: https://www.ensembl.org/Multi/Tools/VEP/Ticket?tl=MdyJP633B4NdLqOb,

avgSaO2adjBMI: https://www.ensembl.org/Multi/Tools/VEP/Ticket?tl=Lg4h2OQ00URxi4cq)

- FinnGen GWAS Release 12- https://r12.finngen.fi/
- LocusFocus - https://locusfocus.research.sickkids.ca/
- UKBiobank PheWeb - https://pheweb.org/UKB-Neale/
- Chromatin conformation and accessibility datasets – Have been deposited at either the Gene Expression Omnibus (https://www.ncbi.nlm.nih.gov/geo) or Array Express

(https://www.ebi.ac.uk/biostudies/arrayexpress). Accession numbers for datasets are hMSC_Osteoblasts (E-MTAB-6835); hMSC_Adipocytes (GSE164912); SFBS_diff (GSE262484); hESC_HypothalamicNPC, hESC_Hypothalamic Neurons (GSE152098); iPSC_Neurons (E-MTAB-9159); Naive T, Naive B, TFH, and GCB (GSE174658); HepG2 (E-MTAB-7144); iPSC-Hepatocytes (GSE189026); Chondrocytes, hFOBs (GSE261284).

- The online multivariate-genotype-phenotype mapping tool is available at (https://hallgrimssonlab.ca/). A free sign up is required for access.

## CODE AVAILABILITY

All protocols and code used for running analyses generated by the authors for use in this study are available upon request following publication.

## ETHICS STATEMENTS

All participants provided written informed consent and each study was approved by their respective institutions, as noted below:

### MyCode

The MyCode Community Initiative parent study was approved by the Geisinger IRB (Study # 2006-0258). The MyCode Governing Board reviews and approves all uses of MyCode samples and data. Additionally, the Geisinger IRB reviewed this study and determined the study did not involve human subjects as defined in 45 CFR 46. 102(f); and therefore was not subject to additional oversight by the IRB (Study #2017-158).

### eMERGE

This parent eMERGE III project at Geisinger was reviewed and approved by the Geisinger IRB (Study #2015-0464). Analyses of this project were conducted at Geisinger. Each parent site within eMERGE III was also approved by their respective sites Institutional Review Boards.

### BioVU

Vanderbilt University Medical Center’s BioVU projects are supported by numerous sources: institutional funding, private agencies, and federal grants. These include NIH funded Shared Instrumentation Grant S10OD017985, S10RR025141, and S10OD025092; CTSA grants UL1TR002243, UL1TR000445, and UL1RR024975. Genomic data are also supported by investigator-led projects that include U01HG004798, R01NS032830, RC2GM092618, P50GM115305, U01HG006378, U19HL065962, R01HD074711; and additional funding sources are detailed here: https://victr.vumc.org/biovu-funding/.

### Amgen deCODE Genetics

The deCODE parent study was reviewed and approved by the National Bioethics Committee in Iceland (Approval no. VSN-02-078, renewed approval no. VSN-19-010), following a review by the Icelandic Data Protection Authority. All participating individuals who donated blood signed informed consent. The personal identities of the participants data and biological samples were encrypted using a third-party system (Idendity Protection System) approved and monitored by the Icelandic Data Protection Authority.

### KOGES

The present study was considered minimal risk to participants and was granted an informed consent waiver by the Institutional Review Board of Korea University Ansan Hospital (IRB no. 2020AS0118) and the Ethics Committee of the Korean Centers for Disease Control and Prevention (no. NBK-2020-093). All procedures were conducted in accordance with relevant guidelines and regulations.

### PMBB

The PMBB is approved under IRB protocol# 813913 and supported by Perelman School of Medicine at University of Pennsylvania, a gift from the Smilow family, and the National Center for Advancing Translational Sciences of the National Institutes of Health under CTSA award number UL1TR001878.

### SAGIC

The SAGIC protocol was approved by the Institutional Review Board (IRB) at the University of Pennsylvania and additional IRB approval was required and obtained at each site. Informed consent was obtained from all participants.

## Data Availability

MyCode Community Health Initiative (MyCode)- MyCode data can be accessed by Geisinger investigators. There are restrictions to the sharing of MyCode DiscovEHR genetic datasets related to agreements between Geisinger and the Regeneron Genetics Center. Requests can be made by contacting the corresponding author.
Electronic Medical Records and Genomics (eMERGE)- Data for eMERGE III are publicly accessible through the database of Genotypes and Phenotypes (dbGaP), accession numbers phs001584.v2.p2, pht010214.v1.p2, pht009069.v2.p2, pht009070.v2.p2, pht009071.v2.p2, pht009073.v2.p2, and pht010215.v1.p2.
Amgen deCODE Genetics (deCODE study)- The sequence variants passing GATK filters in our previously described Icelandic population whole-genome sequence data have been deposited at the European Variant Archive under accession code PRJEB15197. Individual level phenotype and genotype data are protected and cannot be shared according to Icelandic law. De-identified GWAS summary statistics included in this study may be made available upon reasonable request. Data access requests should be directed to.
Penn Medicine BioBank (PMBB)- Individual-level genotype and phenotype data from the PMBB are not publicly available due to privacy concerns. Data and specimens are available to investigators affiliated with Penn Medicine and to external collaborators via scientific collaboration with identified local Penn investigators.187
Korean Genome and Epidemiology Study (KOGES)- Genotyping and cohort data used for this study are available from the National Biobank of Korea (https://biobank.nih.go.kr/eng/) and are managed by the National Institute of Health, Republic of Korea. Data access requests should be directed to. Upon reasonable request, de-identified summary data and results may be shared following approval by the Distribution Review Board in National Institute of Health, Republic of Korea.
Sleep Apnea Global Interdisciplinary Consortium (SAGIC)- Data access is available to qualified researchers upon request and in accordance with site-specific informed consent. Requests should be directed to Allan Pack and are reviewed for approval by the SAGIC site PIs and Executive Committee.

## ACKNOWLEDGEMENTS

We used data from the publicly available FinnGen GWAS Release 12 data. We want to acknowledge the participants and investigators of the FinnGen study for making these data public. Anne E. Justice (AEJ), Geetha Chittoor (GC), Shreyash Gupta (SG), Allan I. Pack (AIP), Brendan Keenan (BK), Struan FA. Grant (SFAG); Benedikt Hallgrimsson (BH) were in part funded by the National Institutes of Health (NIH), National Heart Lung and Blood Institute (NHLBI) grant P01 HL160471. AEJ, GC, Navya Shilpa Josyula (NSJ), H. Lester Kirchner (HLK), AIP, BK, and Janet Robishaw (JB) were in part funded by NIH NHLBI grant R01 HL134015. Peter A Cistulli (PAC) was funded by NHMRC Australia (2008157). Daniel J. Gottlieb (DJB) was funded by I01 BX004821 (VA BLR&D). Ulysses J. Magalang (UJM) was funded by P01 HL160471 and R01HL175579. Marc S. Williams (MSW) was funded by R01 HL134015.

## Geisinger’s MyCode Community Health Initiative Study (MyCode)

We thank all the participants of the MyCode Study. We thank the members of the Geisinger-Regeneron DiscovEHR Collaboration who have been critical in the generation of the genetic data used in this study.

## Electronic Medical Records and Genomics (eMERGE) Network (Phase III)

This phase of the eMERGE Network was initiated and funded by the NHGRI through the following grants: U01HG008657 (Group Health Cooperative/University of Washington); U01HG008685 (Brigham and Women’s Hospital); U01HG008672 (Vanderbilt University Medical Center); U01HG008666 (Cincinnati Children’s Hospital Medical Center); U01HG006379 (Mayo Clinic); U01HG008679 (Geisinger Clinic); U01HG008680 (Columbia University Health Sciences); U01HG008684 (Children’s Hospital of Philadelphia); U01HG008673 (Northwestern University); U01HG008701 (Vanderbilt University Medical Center serving as the Coordinating Center); U01HG008676 (Partners Healthcare/Broad Institute); U01HG008664 (Baylor College of Medicine); and U54MD007593 (Meharry Medical College).

## BioVU

Vanderbilt University Medical Center’s BioVU projects are supported by numerous sources: institutional funding, private agencies, and federal grants. These include NIH funded Shared Instrumentation Grant S10OD017985, S10RR025141, and S10OD025092; CTSA grants UL1TR002243, UL1TR000445, and UL1RR024975. Genomic data are also supported by investigator-led projects that include U01HG004798, R01NS032830, RC2GM092618, P50GM115305, U01HG006378, U19HL065962, R01HD074711; and additional funding sources can be found at https://victr.vumc.org/biovu-funding/.

## Amgen deCODE Genetics

We thank all study participants and the staff at the Icelandic Patient Recruitment Center and the Amgen deCODE Genetics core facilities. We also thank collaborators at Amgen deCODE Genetics who have been critical in the generation, processing and analysis of the genetic data used in this study.

## KOGES

This study was supported by grants from the Korea Disease Control and Prevention Agency (KDCA; Nos. 2001-347-6111-221 and 2002-347-6111-221 to C.S.), the Basic Science Research Program through the National Research Foundation of Korea (NRF) funded by the Ministry of Education (NRF-2022R1I1A1A01065700 [RS-2022-NR075013] to S.K.), and the Korea Health Technology R&D Project through the Korea Health Industry Development Institute (KHIDI) funded by the Ministry of Health and Welfare (HI20C0469 [RS-2020KH084762] to S.K.).

## PMBB

The authors would like to thank the Penn Medicine Biobank and Regeneron Genetics Center for providing genetic data used in this analysis. Great thanks also goes to the patient-participants of Penn Medicine who consented to participate in this research program.

## AUTHOR CONTRIBUTIONS

**Conceived of Study Design:** AEJ, AIP, BTK; **Drafted Manuscript:** AEJ, AIP, BTK, MCP, BH, SFAG; **Acquisition of Data:** AEJ, AIP, BTK, IJ, KS; JAP, CLAB, MJZ, ADW, SFAG, PAC; **Data Preparation:** AEJ, BTK, NSJ, GC, GT, EM, MCP,KB; **Performed Statistical Analyses:** AEJ, NSJ, BTK, GC, BH, HH, AG, SG, SRK, GT, EM, MCP; **Prepared Figures and/or Tables:** AEJ, NSJ, GC, SG, BH, HH, AG, MCP; **Supervised the work:** AIP, AEJ, SFAG, BH, IJ. **Reviewed and approved the manuscript:** AEJ, AIP, BTK, MCP, SFAG, MSW, SRK, DJG, TIM, JDR, PAC, MJZ, GC, BS, SG, DRM, NSJ, EM, AG, CLA, JB, NC, LJF, MG, HH, HLK, UJM, BM, NM, TP, CS, TS, OJV, ADW, PZ, TG, BB, BH; **All authors agree to be held accountable for the content**: AEJ, BTK, GC, MCP, HH, NSJ, SG, SRK, GT, EM, AG, FAO, CLAB, BB, JB, KB, NHC, PAC, PAC, LJF, DJG, HH, HLK, UJM, BAM, DRM, NM, FDM, TIM, TP, JAP, LJRT, CS, BS, TS, OJV, DAV, ADW, MSW, PZ, MJZ, IJ, TG, JR, KS, SFAG, BH, AIP.

## DISCLOSURES/COMPETING INTERESTS

Dr. Timothy I. Morgenthaler (TIM) has served in a consulting capacity on an advisory committee for Apnimed.

## Notes

### Funding Statement

This study was funded by grants and contracts from the National Institutes of Health (P01 HL160471 - HL134015 - U01HG008657 - U01HG008685 - U01HG008672 - U01HG008666 - U01HG006379 - U01HG008679 -U01HG008680 - U01HG008684 - U01HG008673 - U01HG008701- U01HG008676- U01HG008664 - U54MD007593 - R01HL175579 - S10OD017985 -S10RR025141 - S10OD025092 - UL1TR002243 - UL1TR000445 - UL1RR024975 -U01HG004798 - R01NS032830 - RC2GM092618 - P50GM115305 - U01HG006378 - U19HL065962 - R01HD074711) and NHMRC Australia (2008157) and Veterans Affairs (I01 BX004821 VA BLRD) and Korea Disease Control and Prevention Agency (2001-347-6111-221 and 2002-347-6111-221) and the Basic Science Research Program through the National Research Foundation of Korea (NRF) funded by the Ministry of Education (NRF-2022R1I1A1A01065700 - RS-2022-NR075013), and the Korea Health Technology RD Project through the Korea Health Industry Development Institute (KHIDI) funded by the Ministry of Health and Welfare (HI20C0469 - RS-2020KH084762).

### Author Declarations

All participants provided written informed consent and each study was approved by their respective institutions. The Geisinger Institutional Review Board (IRB) reviewed this study and determined the study did not involve human subjects as defined in 45 CFR 46. 102(f), and therefore was not subject to additional oversight by the IRB -Study 2017-158. This parent eMERGE III project at Geisinger was reviewed and approved by the Geisinger IRB -Study - 2015-0464. Analyses for eMERGE III were conducted at Geisinger. The deCODE parent study was reviewed and approved by the National Bioethics Committee in Iceland - Approval no. VSN-02-078, renewed approval no. VSN-19-010 - following a review by the Icelandic Data Protection Authority. All participating individuals who donated blood signed informed consent. The personal identities of the participants data and biological samples were encrypted using a third-party system approved and monitored by the Icelandic Data Protection Authority. The present study was considered minimal risk to participants and was granted an informed consent waiver by the Institutional Review Board of Korea University Ansan Hospital - IRB no. 2020AS0118 - and the Ethics Committee of the Korean Centers for Disease Control and Prevention - no. NBK-2020-093. All procedures were conducted in accordance with relevant guidelines and regulations. The PMBB is approved under IRB protocol - 813913 by the IRB at the Perelman School of Medicine at University of Pennsylvania. The SAGIC and BioVU analyses were also carried out at University of Pennsylvania. The protocol was approved by the Institutional Review Board (IRB) at the University of Pennsylvania and additional IRB approval was required and obtained at each site.

