## Supplemental Materials for "COMPREHENSIVE GENETIC INVESTIGATION REVEALS HETEROGENEOUS PATHWAYS TO OBSTRUCTIVE SLEEP APNEA"

### Index of Supplemental Materials


| Item | Title | Page(s) |
|---|---|---|
| **Supplementary Notes (SN)** | | |
| SN 1 | Detailed methods for chromatin accessibility specifically related to new data in chondrocytes | 2 - 5 |
| **Supplementary Figures (SF)** | | |
| SF 1 | Overview if GWAS study design | 6 |
| SF 2 | Locuszoom regional association plots for OSA case-control association signals | 7 - 12 |
| SF 3 | Locuszoom regional association plots for OSAadjBMI case-control association signals | 13 - 14 |
| SF 4 | Fine-mapping regional plots for OSA case-control association signals | 15 - 20 |
| SF 5 | Fine-mapping regional plots for OSAadjBMI case-control association signals | 21 - 22 |
| SF 6 | Locuszoom regional association plot for AHI association signal | 23 |
| SF 7 | Locuszoom regional association plot for AHIadjBMI association signal | 24 |
| SF 8 | Locuszoom regional association plots for avgSaO2 association signals | 25 |
| SF 9 | Locuszoom regional association plots for avgSaO2adjBMI association signal | 26 |
| SF 10 | Fine-mapping regional plots for AHI association signals | 27 |
| SF 11 | Fine-mapping regional plots for AHIadjBMI association signals | 28 |
| SF 12 | Fine-mapping regional plots for avgSaO2 association signals | 29 |
| SF 13 | Fine-mapping regional plots for avgSaO2adjBMI association signals | 30 |
| SF 14 | Significant cis-eQTL colocalization results | 31 - 32 |
| **Supplementary Tables (ST)** | | |
| ST 1 | Phenotype descriptive statistics by study and race/ethnicity | 33 |
| ST 2 | Genotype and imputation characteristics by study | 34 - 35 |
| ST 3 | Association results for OSA-associated SNPs across all analyses and stages | 36 - 39 |
| ST 4 | Results for approximate conditional analyses to identify secondary signals in identified association regions | 40 |
| ST 5 | Summary of association results for after approximate conditional analyses on known index variants | 41 |
| ST 6 | Fine-mapping results | 42 - 43 |
| ST 7 | Association results for AHI-associated SNPs across all analyses and stages | 44 - 45 |
| ST 8 | Association results for avgSaO2-associated SNPs across all analyses and stages | 46 - 47 |
| ST 9 | Lookups of top associated SNPs from discovery in existing databases of related traits | 48 - 49 |
| ST 10 | Lookup of previously-published OSA and OSAadjBMI SNPs in discovery analysis | 50 - 52 |
| ST 11 | Lookup of previously-published AHI or avgSAO2 SNPs in discovery analysis | 53 - 54 |
| ST 12 | Results from LD score regression | 55 |
| ST 13 | PheWAS results for suggestively significant SNP-PheCode associations | 56 - 57 |
| ST 14 | Gene-based association results from MAGMA analysis | 58 |
| ST 15 | Variant annotation for fine-mapped variants with highest posterior probability | 59 - 82 |
| ST 16 | Prioritized effector genes from variant to gene (V2G) mapping analysis | 83 - 103 |
| ST 17 | Summary of results for the multivariate genotype-phenotype (MGP) in mice on PLS axis 1 | 104 |
| ST 18 | Gene loadings on PLS1 for mouse multivariate genotype-phenotype (MGP) analyses by cell-type gene list | 105 - 106 |
| ST 19 | Summary of results for the MGP in humans on PLS axis 1 | 107 |
| ST 20 | Gene loadings on PLS1 for mouse MGP analyses by cell-type gene list | 108 - 109 |
| ST 21 | Significant eQTL colocalization results | 110 |
| ST 22 | eQTL colocaization results for all gene - tissue pairs that passed stage one significance criteria | 111 |

**Supplementary Note 1.** Detailed methods for chromatin accessibility specifically related to new data in chondrocytes.

##### *Chondrocytes Cell preparation*

Primary chondrocytes derived from healthy donors were procured via AlloSource (Centennial, CO) from eligible donors. Donor eligibility was determined in accordance with American Association of Tissue Banks (AATB) and Food and Drug Administration (FDA) regulations. Tissue fragments that included subchondral bone and cartilage from both the tibial plateau and femoral condyles were surgically removed from donors (N=3) and immediately placed into pre-chilled (wet ice) wash medium composed of DMEM/F12 medium (Cytiva, #SH30023.01) containing amphotericin (1 ng/ml, Sigma, #50-175-7519), gentamycin (0.05 μg/ml, Gibco, #15750-060), and Pen/Strep (1% v/v, VWR, # K952-100ML) for transport to the laboratory. Articular cartilage was removed from the subchondral bone using a scalpel, minced into 1 mm X 1 mm fragments in a Petrie dish, rinsed three times with phosphate-buffered saline, and placed into a 50 ml conical tube containing 30 ml wash medium supplemented with fetal bovine serum (FBS, 25% v/v, Gibco, #10438-026). Chondrocytes were isolated from the cartilage using a modified version of an established method[1]. Specifically, after 60 minutes of gentle shaking at 37°C, medium was aspirated and replaced with 30 ml digestion medium composed of DMEM/F12, 25% FBS, 0.05 μg/ml gentamycin, 1% v/v Pen/Strep, and ascorbic acid (100 μg/ml, Sigma, #A4544-25G), and supplemented with pronase (0.3 μg/ml, Roche, #10165921001). Cartilage fragments were digested in this medium for 90 minutes at 37°C with gentle shaking, followed by centrifugation at 300 X g at 4°C for 7 minutes to pellet tissues and cells, followed by resuspension in 30 ml digestion medium supplemented with collagenase II (1.2 mg/ml, Worthington, #LS004177). Cartilage was digested for 18 hours at 37°C with gentle shaking and filtered through a 70 μm strainer, with a single rinse of the tube with wash medium to recover all remaining cells and tissue, which was again strained. Strained materials were centrifuged to a pellet at 300 X g for 7 minutes at 4°C, resuspended with fresh wash medium, spun once more at 300 X g for 7 minutes at 4°C to form a pellet, and then finally resuspended in DMEM/F12 containing 20% FBS, 0.05 μg/ml gentamycin, 1% v/v

Pen/Strep, and 100 μg/ml ascorbic acid. Isolated suspensions of chondrocytes were counted using a hemocytometer.

##### *Chondrocyte ATAC-seq library preparation*

Three biological replicates of chondrocytes were harvested using Accutase, followed by a DPBS wash, then counted. 50,000 cells of each sample were spun down at 550 ×g for 5 min at 4°C. The cell pellet was then resuspended in 50 μl cold lysis buffer (10 mM Tris-HCl, pH 7.4, 10 mM NaCl, 3 mM MgCl2, 0.1% IGEPAL CA-630) and spun down immediately at 550 ×g for 10 min, 4°C. The nuclei were resuspended on ice in the transposition reaction mix (2x TD Buffer, 2.5ul Tn5 Transposes and Nuclease Free $H_2O$) (Illumina Cat #FC-121-1030, Nextera) on ice and the transposition reaction was incubated at 37°C for 45 min. The transposed DNA was then purified using a the MinElute Kit (Qiagen) adjusted to 10.5 μl elution buffer. The transposed DNA was converted into libraries using NEBNext High Fidelity 2x PCR Master Mix (NEB) and the Nextera Index Kit (illumina) by PCR amplification for 12 cycles. The PCR reaction was subsequently cleaned up using AMPureXP beads (Agencourt), checked on a Bioanalyzer 2100 (Agilent) high sensitivity DNA Chip (Aglient), and paired-end sequenced on the Illumina NovaSeq 6000 platform (51bp read length) at the Center for Spatial and Functional Genomics at CHOP.

##### *ATAC-seq peak calling*

Fastq files of technical replicates were merged, ATAC-seq peaks were called following the ENCODE ATAC-seq pipeline (https://www.encodeproject.org/atac-seq/). Briefly, pair-end reads from all replicates were aligned to hg38 genome using bowtie2, and duplicate reads were removed from the alignment. Aligned tags were generated by modifying the reads alignment by offsetting +4bp for all the reads aligned to the forward strand, and -5bp for all the reads aligned to the reverse strand. Narrow peaks were called independently for pooled replicates for each cell type using macs2 (-p 0.01 --nomodel --shift -75 --extsize 150 -B --SPMR --keep-dup all --call-summits) and ENCODE blacklist regions (https://www.encodeproject.org/files/ENCFF356LFX/) were removed from called peaks. Finally, a consensus of open chromatin regions (OCRs) called in the majority (>50%) of the replicates were obtained by consolidating the peak sets across samples using bedtools intersect (v2.25.0)[76].

##### *Hi-C library preparation*

Hi-C library preparation on primary chondrocytes derived from healthy donors was performed using the Arima-HiC kit (Arima Genomics Inc; cat# A410030), according to the manufacturer's protocols. Briefly, cells were crosslinked using formaldehyde. Cross-linked cells were then subject to the Arima-HiC protocol, which utilizes multiple restriction enzymes to digest chromatin. Arima-HiC sequencing libraries were prepared by first shearing purified proximally-ligated DNA and then size-selecting 200-600 bp DNA fragments using AmpureXP beads (Beckman Coulter; cat# A63882). The size-selected fragments were then enriched using Enrichment Beads (provided in the Arima-HiC kit), and then converted into Illumina-compatible sequencing libraries with the Swift Accel-NGS 2S Plus DNA Library Kit (Swift, 21024) and Swift 2S Indexing Kit (Swift, 26148). The purified, PCR-amplified DNA underwent standard QC (qPCR, Bioanalyzer, and KAPA Library Quantification [Roche, KK4824]) and was sequenced with unique single indexes on the Illumina NovaSeq 6000 Sequencing System at 2x101 base pairs.

##### *Hi-C data processing*

Paired-end reads from three biological replicates with two technical replicates each were pre-processed using the HICUP pipeline v0.7.4[2], with bowtie as aligner and hg38 as the reference genome. The alignment .bam file were parsed to .pairs format using pairtools v 0.3.0[3] and pairix v0.3.7[4], and eventually converted to pre-binned Hi-C matrix in .cool format by cooler v 0.8.10 with multiple resolutions (1kbp, 2kbp, 4kbp ) and normalized with ICE method[5]. Finally, significant intra-chromosomal interaction loops were determined under multiple resolutions (1kb, 2kb and 4kb) using the Hi-C loop callers mustache v1.0.1 (q-value < 0.1) [6] and Fit-Hi-C2 [7] v2.0.7 (FDR < 1e-6) on merged replicates matrix. The called loops were merged between both callers to represent cell type loops at each resolution.

**Supplementary Figure 1.** Overview if GWAS study design.
Sample sizes provided are maximum sample sizes across all traits and population groups.

Model 1. Sleep trait = SNP + Age + Sex + race/ethnicity* + PCs
Model 2. Sleep trait = SNP + Age + Sex + **BMI** + race/ethnicity* + PCs

*Allowing for heterogeneity by self-identified ancestry in All Ancestry analyses for quantitative traits. Adjusting for race/ethnicity in case/control.

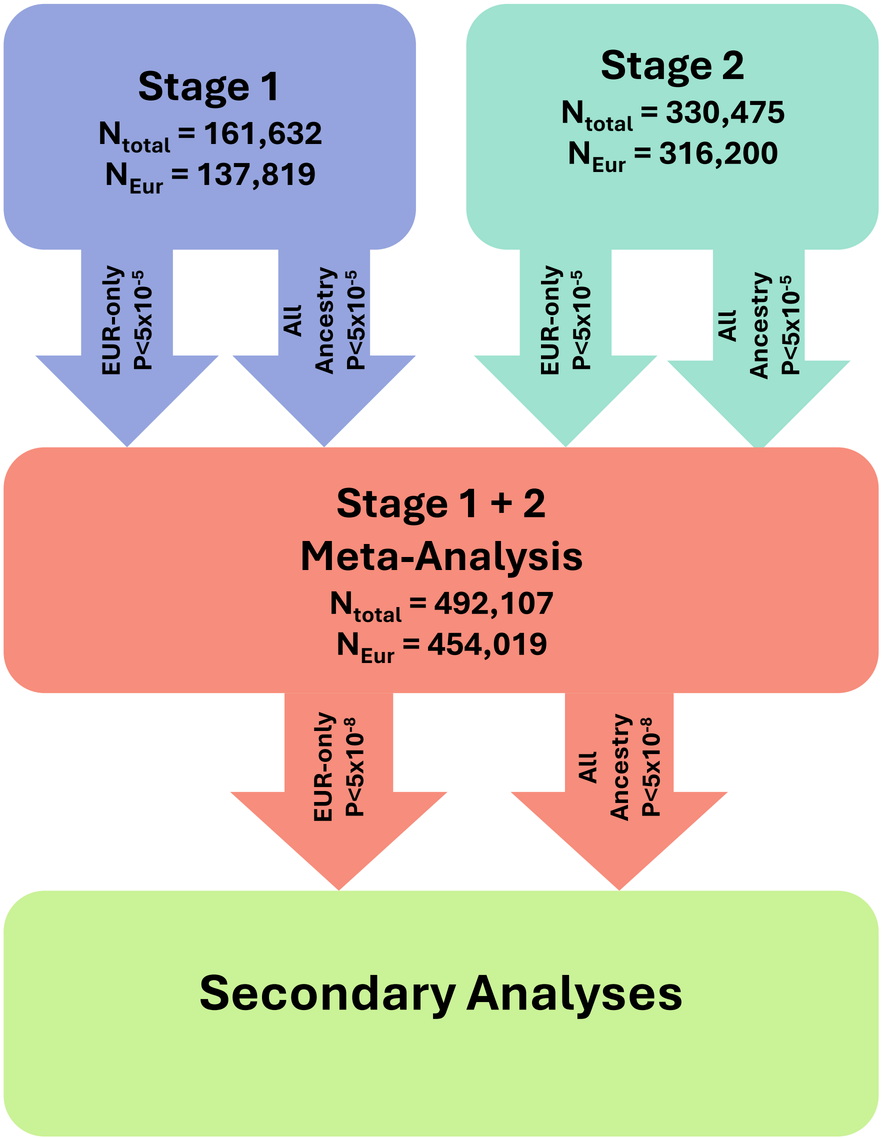

**Supplementary Figure 2**. **Locuszoom regional association plots for OSA case-control association signals.**

Regional association plots of GWAS signals from stage 1 plus stage 2 meta-analyses. The plots appear in order of chromosome and position, strata (pooled vs. European-only), and presentation in Table 1. Unrelated individuals of European genetic similarity from the MyCode study were used to calculate LD displayed in plot. A) rs12725199 – *PTPB2*; B) rs2376073 – intergenic; C) rs9831061 – *ETV5*; D) rs9380814 – *DNAH8*; E) rs66474734 – *CSMD3*; F) rs12553324 – *ELAVL2*; G) rs2958127 – *NACA; H)* rs11642015 – *FTO*; I) rs1800437 – *GIPR*; J) rs11088984 – *POFUT2*; and K) rs34592089 – *BANK1*.

A. rs12725199 – *PTPB2*

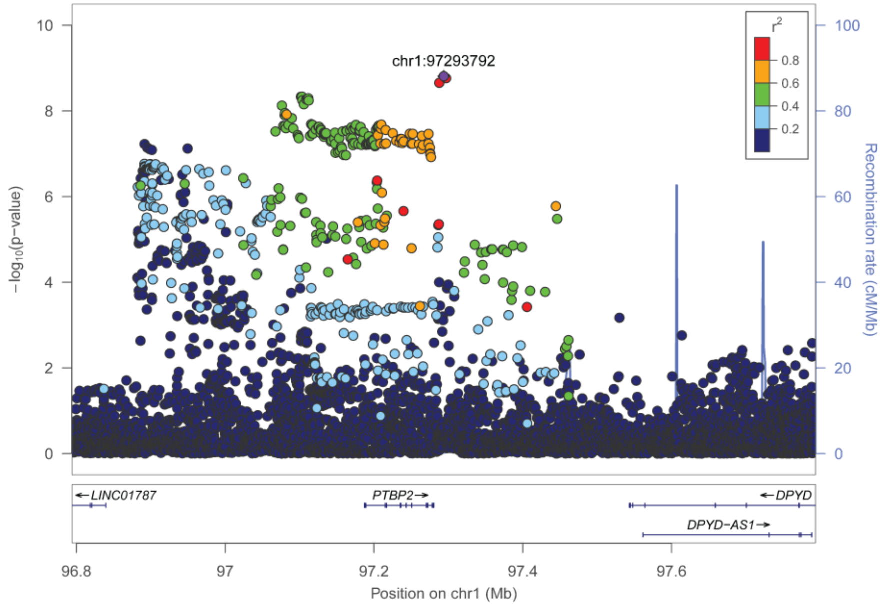


B. rs2376073 – intergenic

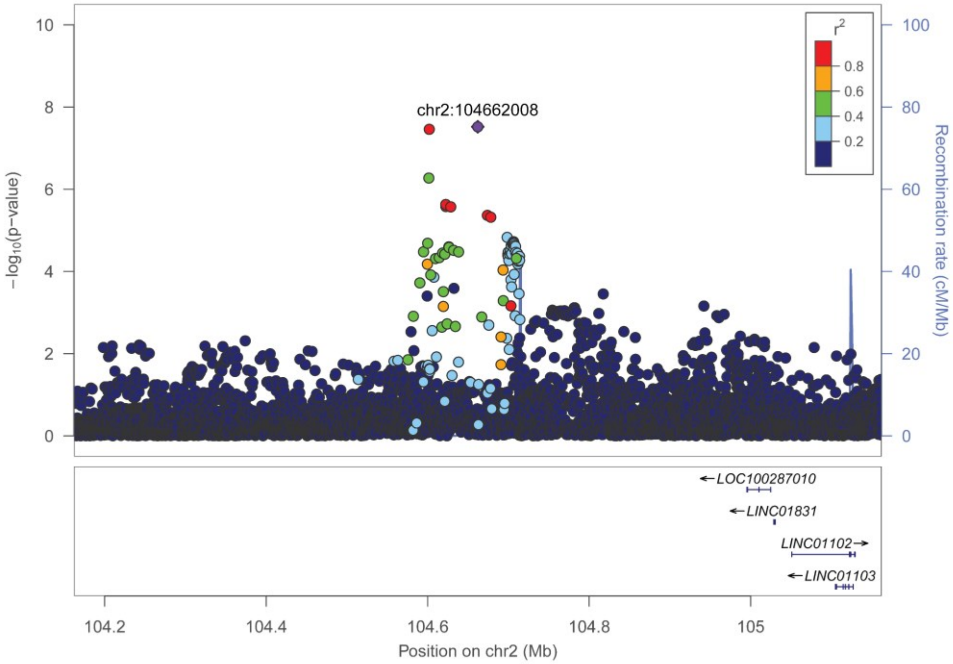

C. rs9831061 – ETV5

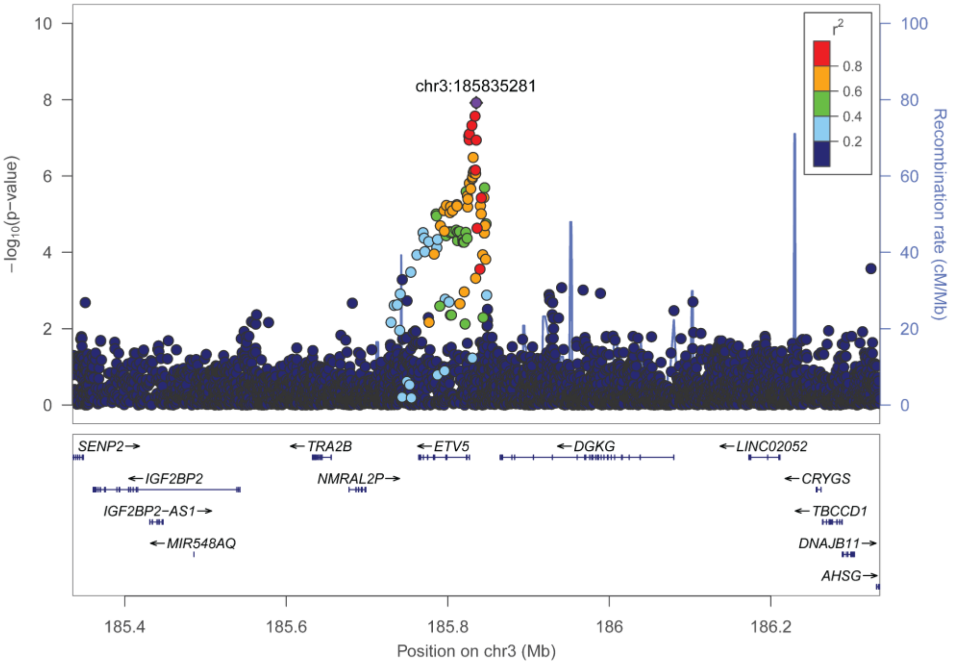


D. rs9380814 – *DNAH8*

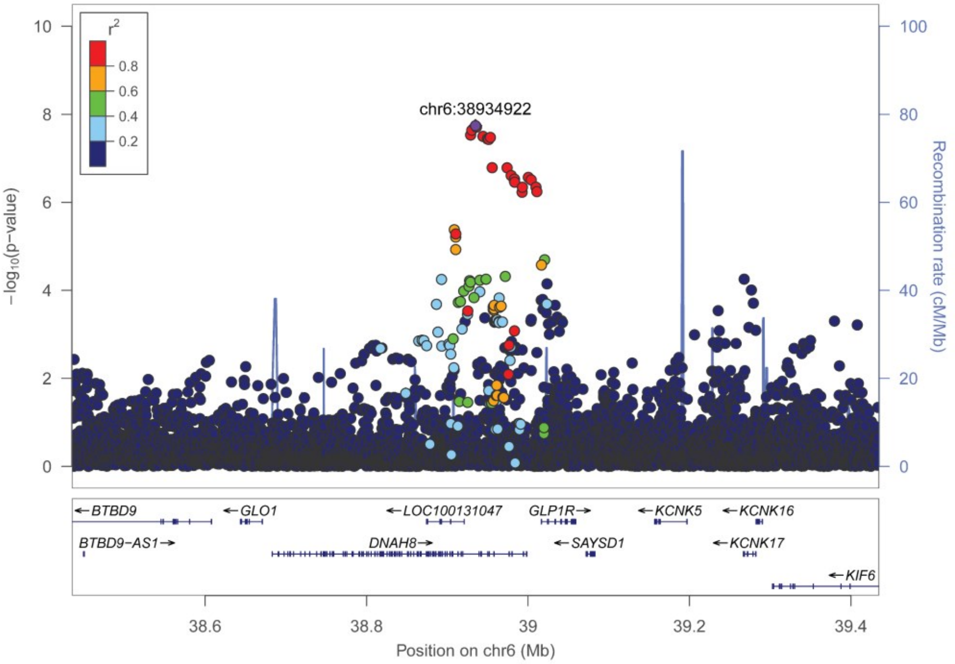

E. rs66474734 – *CSMD3*

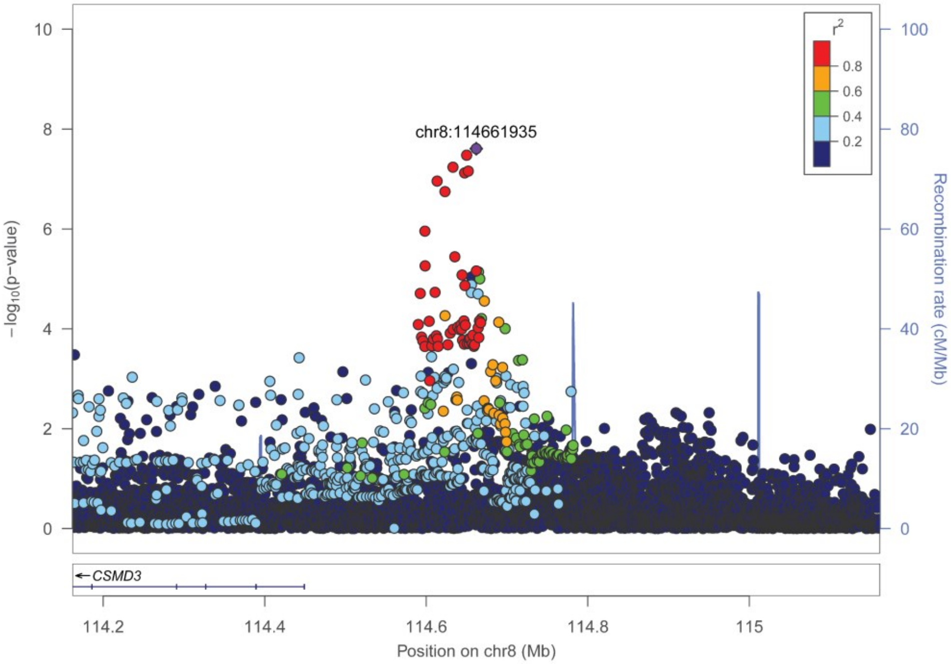


F. rs12553324 – *ELAVL2*

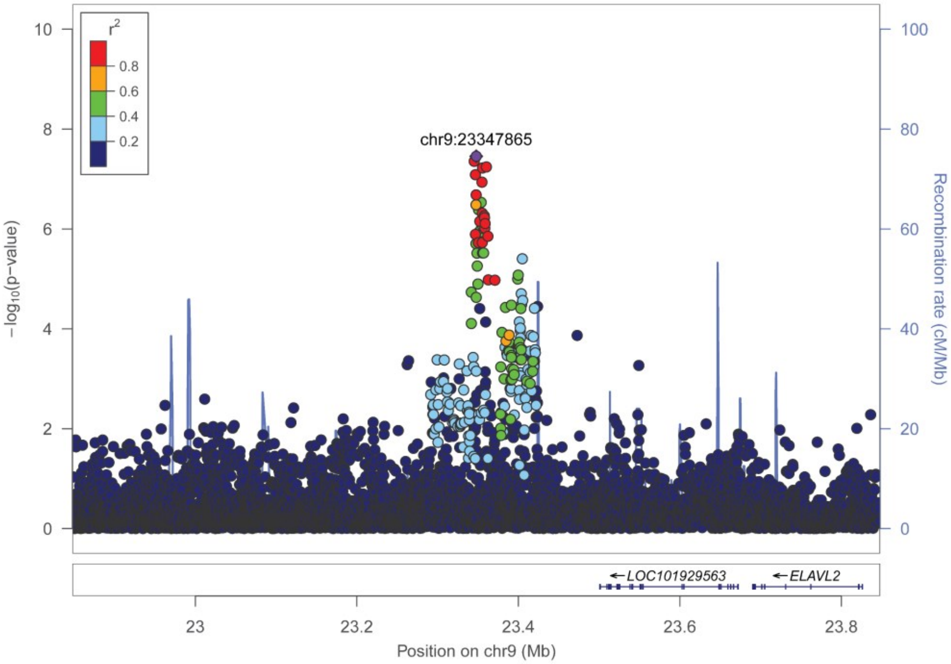

G. rs2958127 – *NACA*

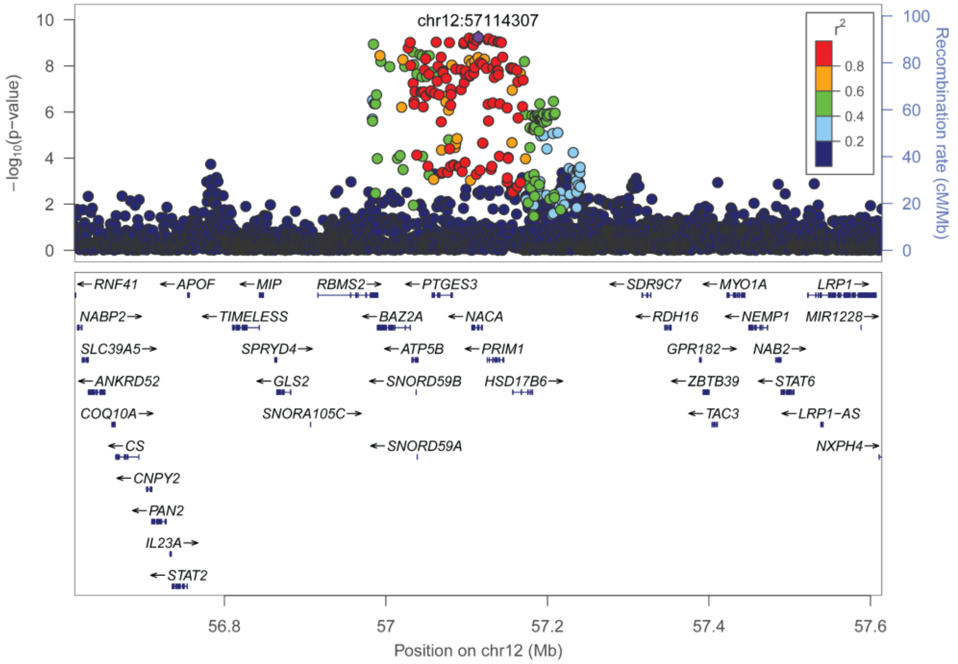


H. rs11642015 – *FTO*

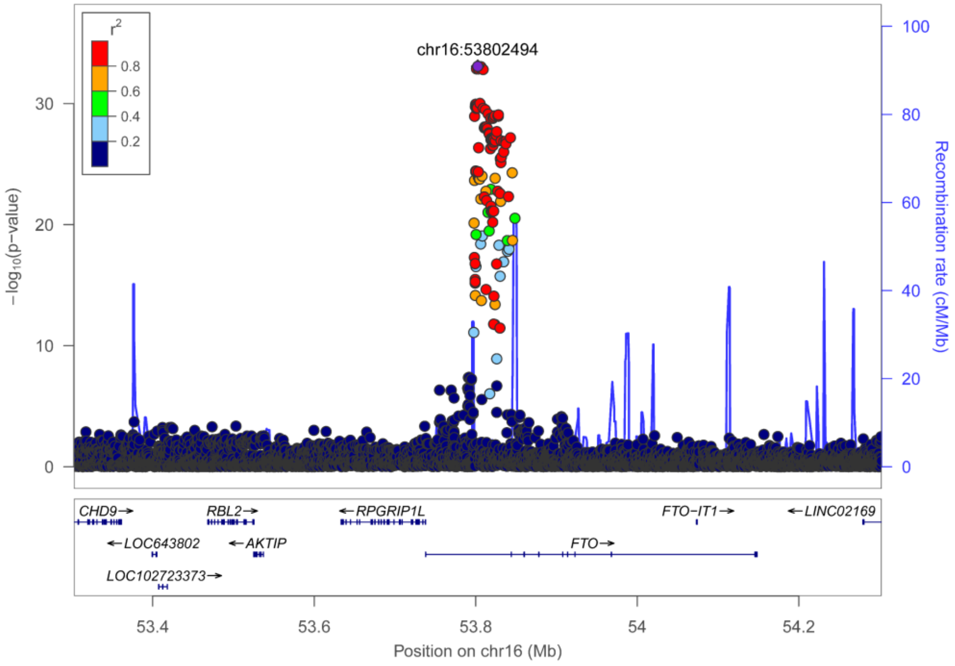

I. rs1800437 – *GIPR*

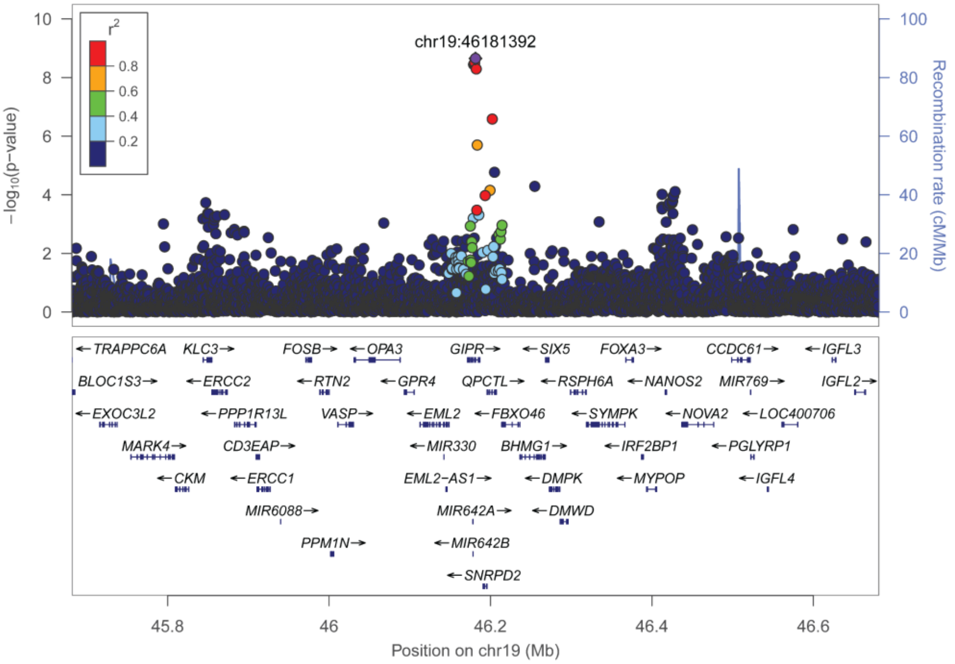


J. rs11088984 – *POFUT2*

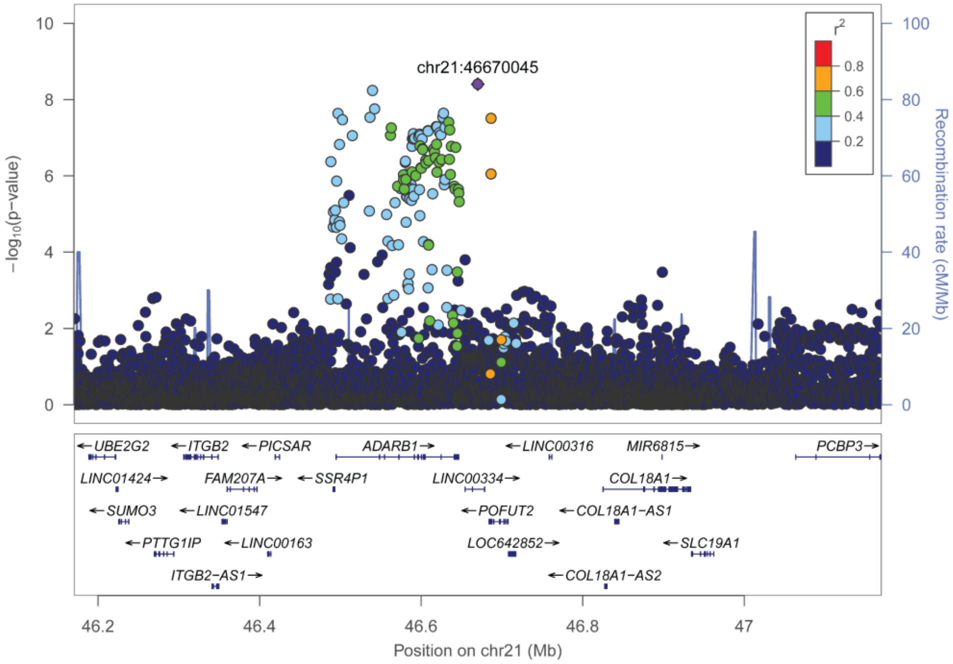

K. rs34592089 – *BANK1*

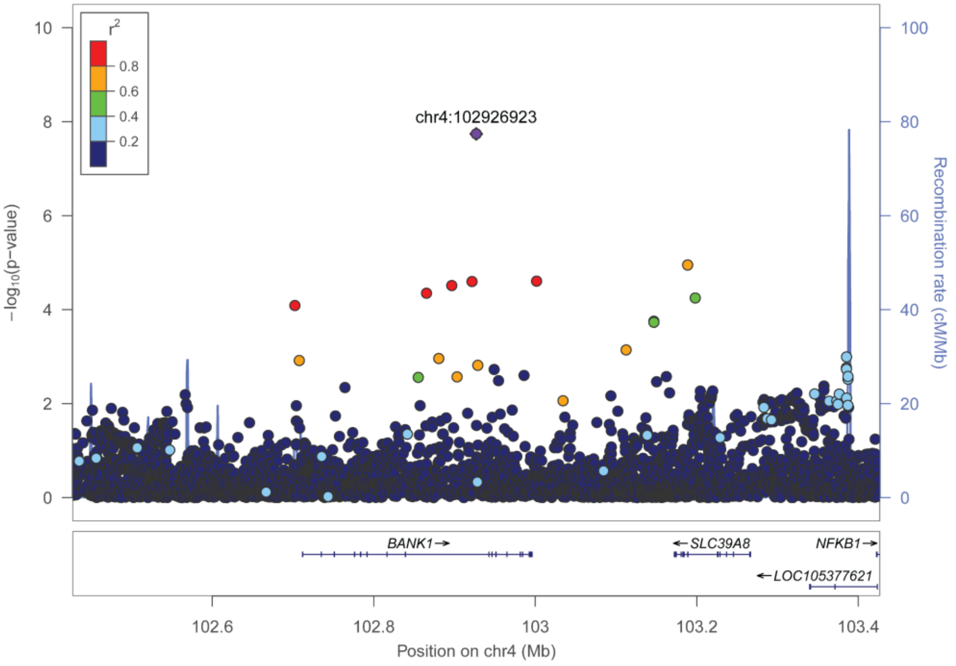

**Supplementary Figure 3**. **Locuszoom regional association plots for OSAadjBMI case-control association signals.**

Regional association plots of GWAS signals from stage 1 plus stage 2 meta-analyses. The plots appear in order of chromosome and position, strata (pooled vs. European-only), and presentation in Table 2. Unrelated individuals of European genetic similarity from the MyCode study were used to calculate LD displayed in plot. A) rs2964006 – *GRIA1*; B) rs4759254 – *NACA*; C) rs10735928 – *MSRB3*; and D) rs60791051 – *FAM189A2*.

A. rs2964006 – *GRIA1*

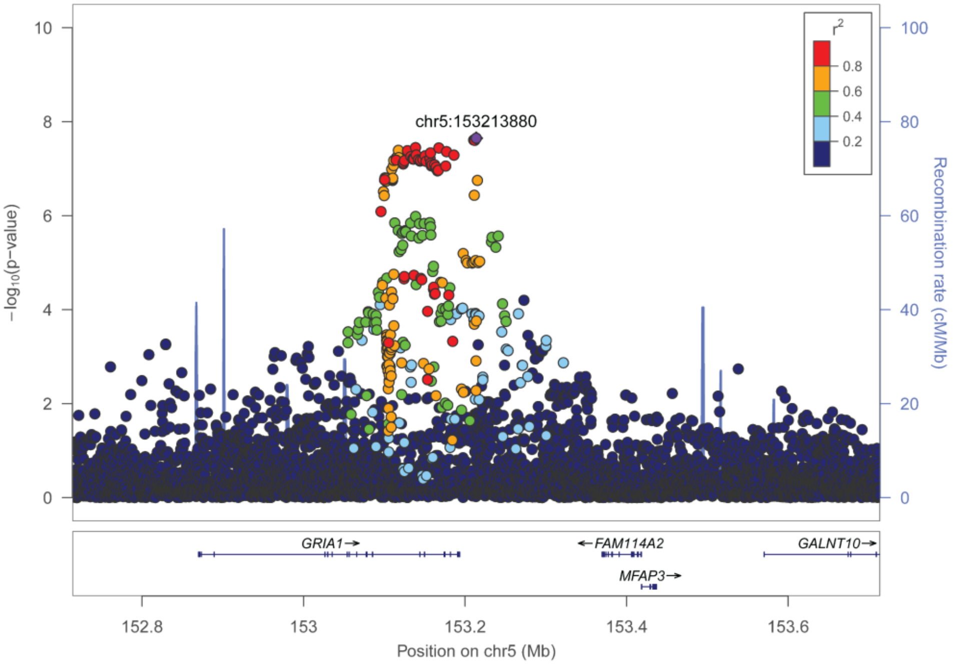


B. rs4759254 – *NACA*

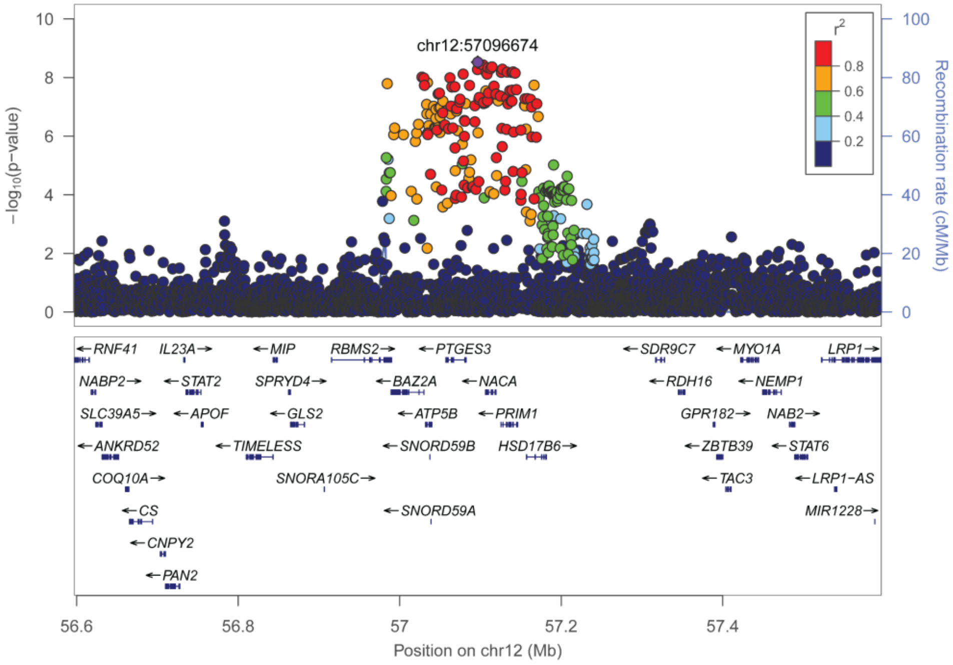

C. rs10735928 – *MSRB3*

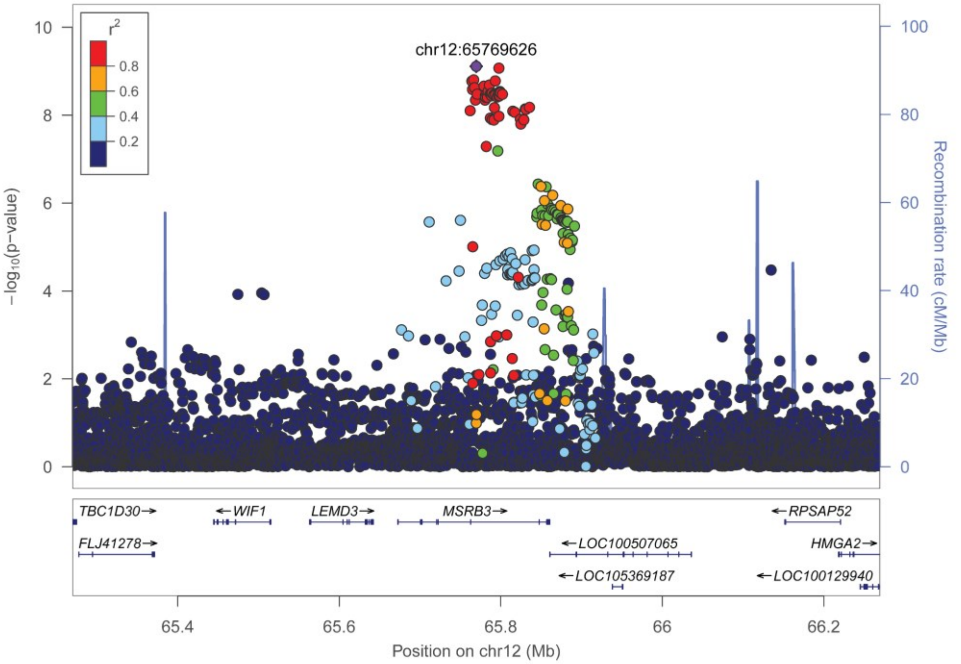


D. rs60791051 – *FAM189A2*

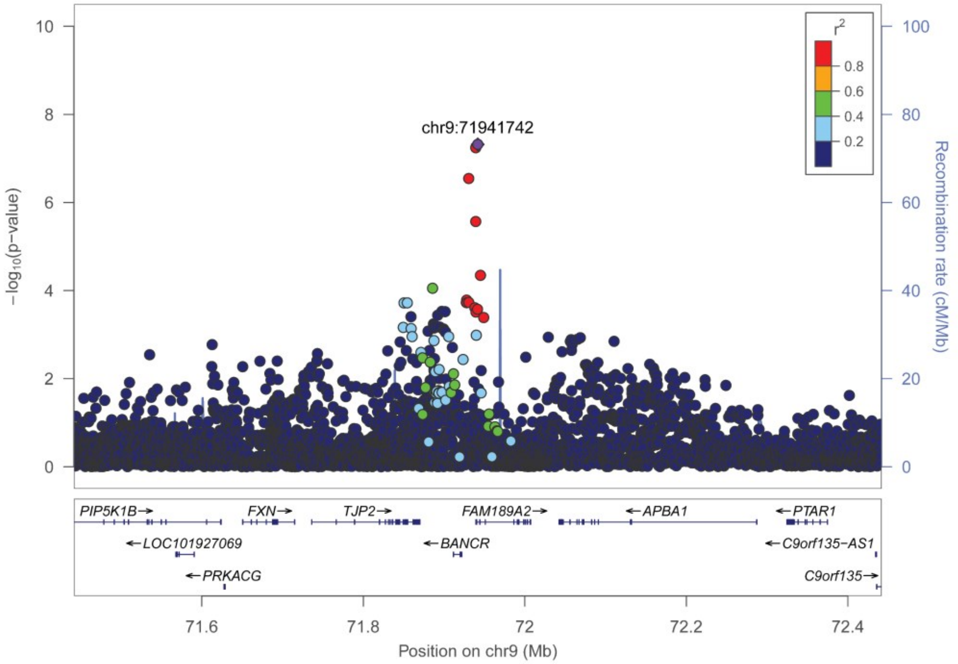

**Supplementary Figure 4**. **Fine-mapping regional plots for OSA case-control association signals.** Regional plots of posterior probability (prob) from fine-mapping analysis conducted in FINEMAP, including all variants ± 500 kb from index variant for each association region. The plots appear in order of chromosome and position, strata (pooled vs. European-only), and presentation in Table 1. Unrelated individuals of European genetic similarity from the MyCode study were used to calculate LD displayed in plot. A) rs12725199 – *PTPB2*; B) rs2376073 – intergenic; C) rs9831061 – *ETV5*; D) rs9380814 – *DNAH8*; E) rs66474734 – *CSMD3*; F) rs12553324 – *ELAVL2*; G) rs2958127 – *NACA; H)* rs11642015 – *FTO*; I) rs1800437 – *GIPR*; J) rs11088984 – *POFUT2*; and K) rs34592089 – *BANK1*.

A. rs12725199 – *PTPB2*

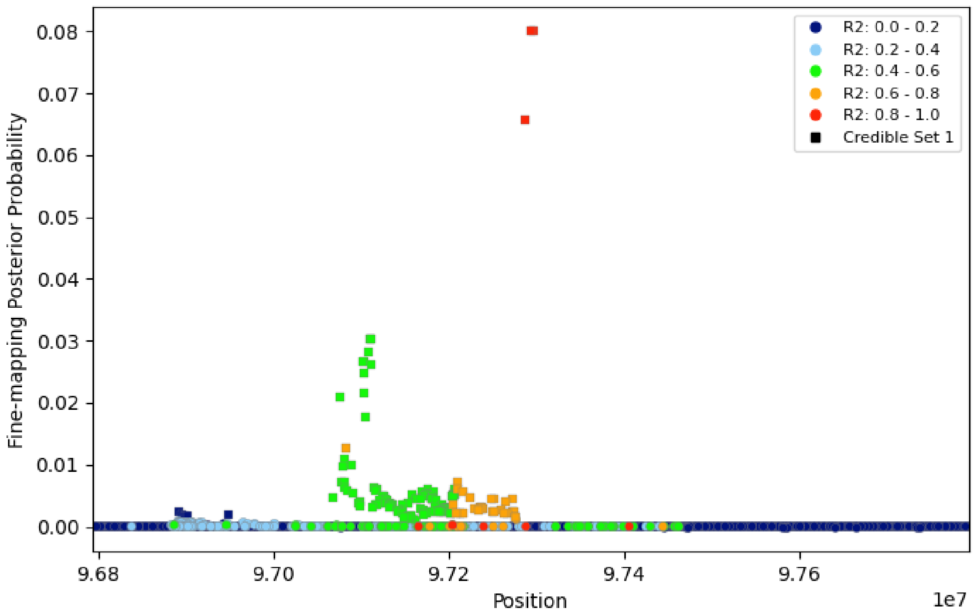


B. rs2376073 – intergenic

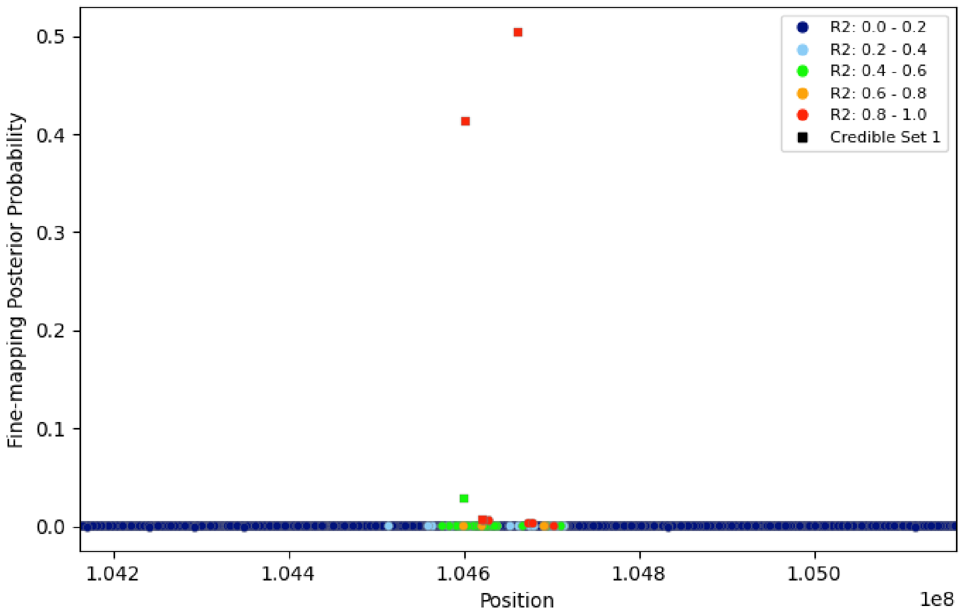

C. rs9831061 – ETV5

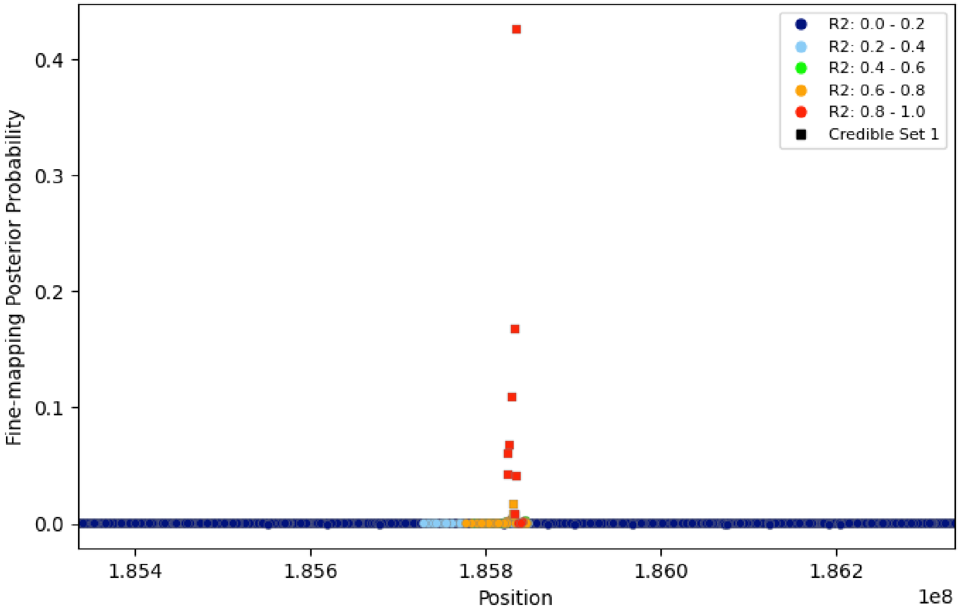


D. rs9380814 – *DNAH8*

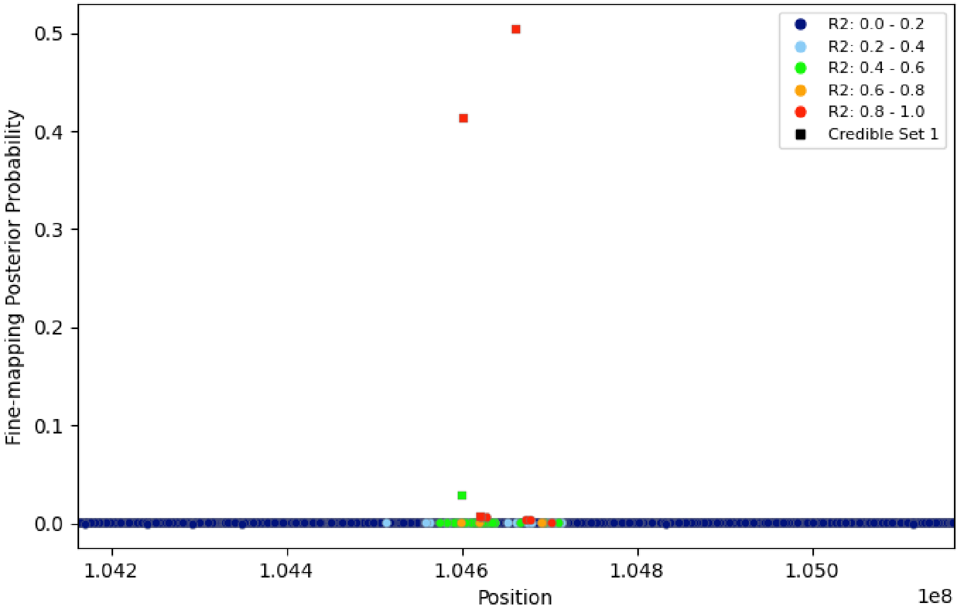

E. rs66474734 – *CSMD3*

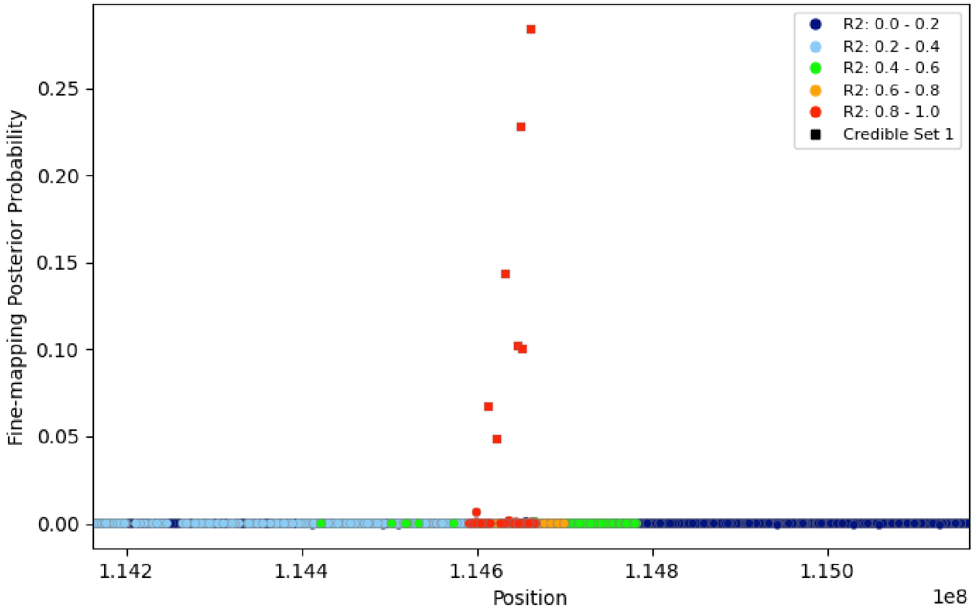


F. rs12553324 – *ELAVL2*

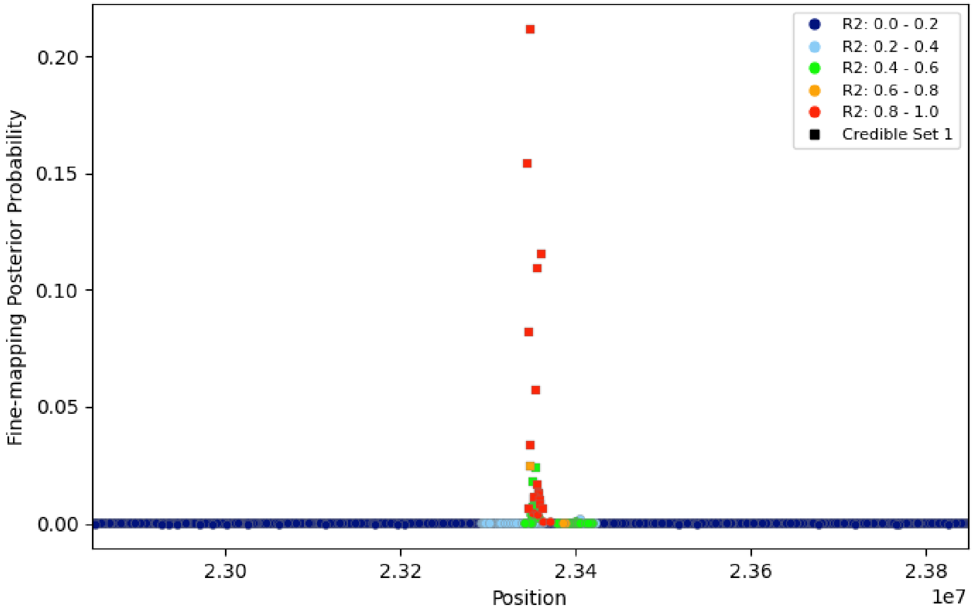

G. rs2958127 – *NACA*

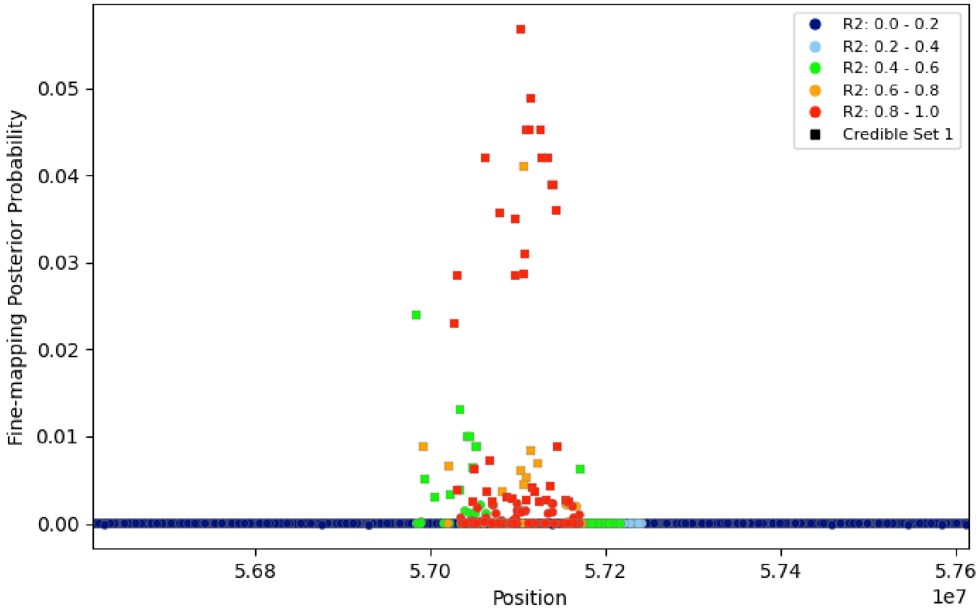


H. rs11642015 – *FTO*

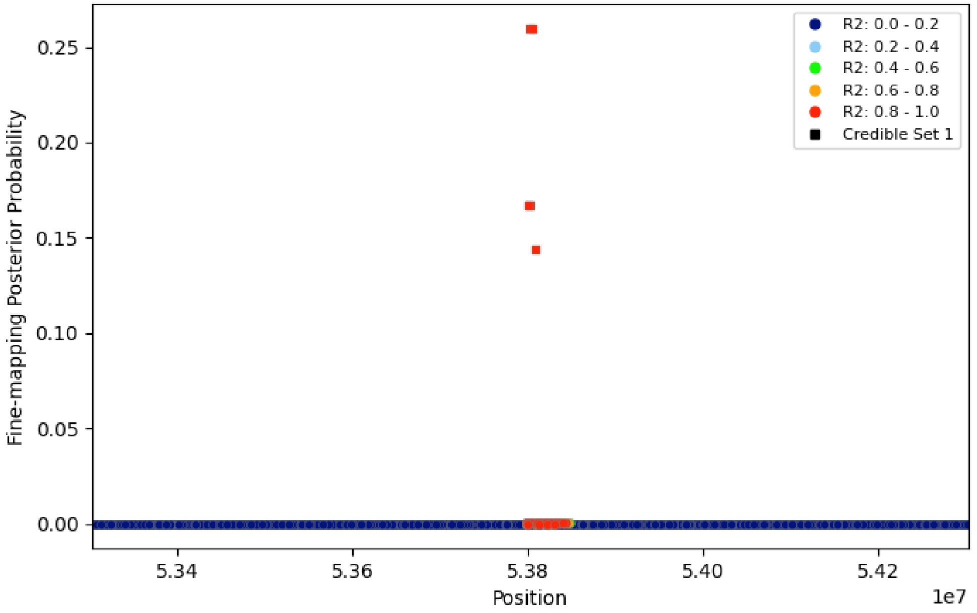

I. rs1800437 – *GIPR*

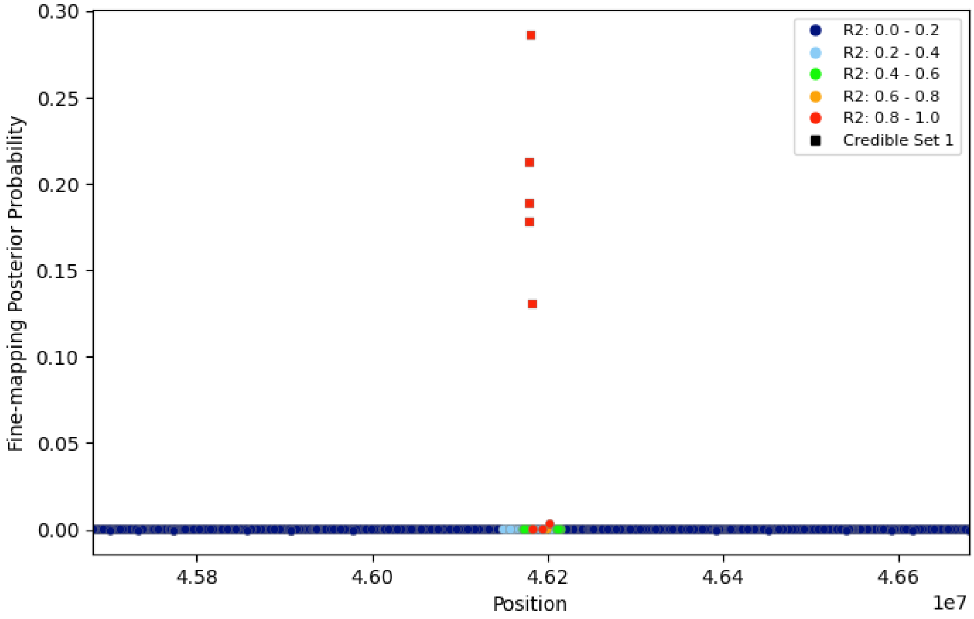


J. rs11088984 – *POFUT2*

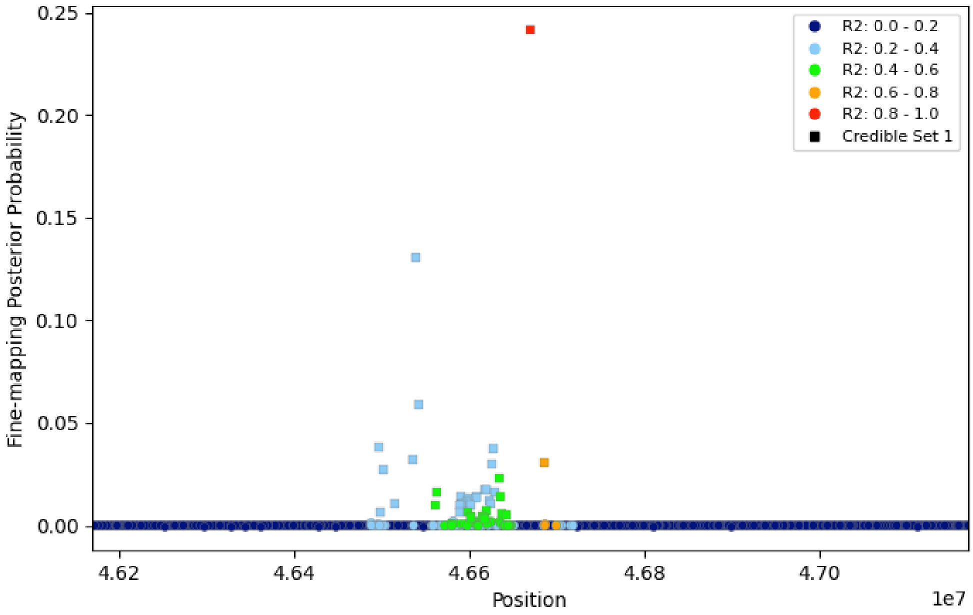

K. rs34592089 – *BANK1*

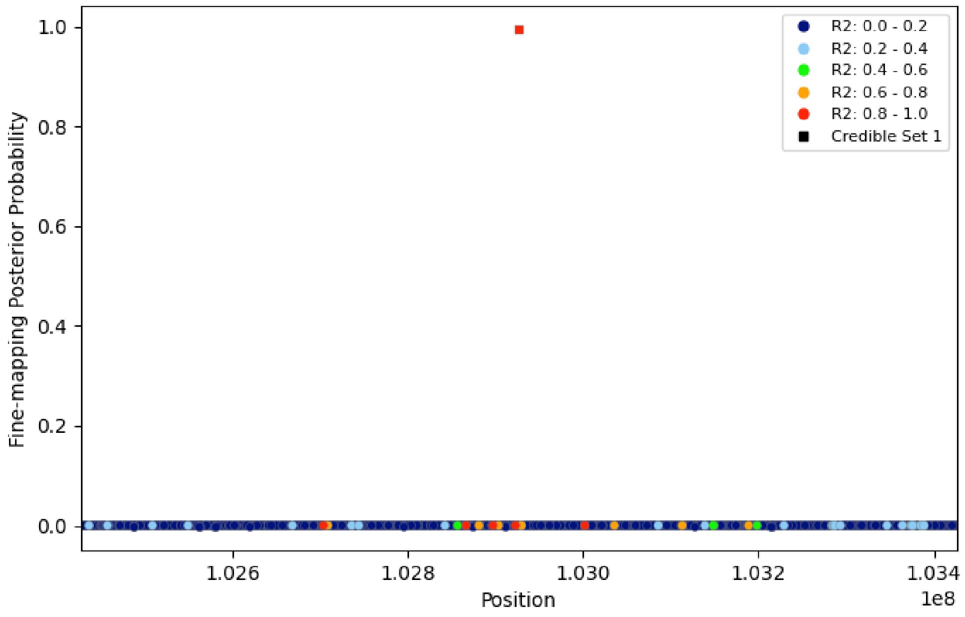

**Supplementary Figure 5**. **Fine-mapping regional plots for OSAadjBMI case-control association signals.** Regional plots of posterior probability (prob) from fine-mapping analysis conducted in FINEMAP, including all variants ± 500 kb from index variant for each association region. The plots appear in order of chromosome and position, strata (pooled vs. European-only), and presentation in Table 2. Unrelated individuals of European genetic similarity from the MyCode study were used to calculate LD displayed in plot. A) rs2964006 – *GRIA1*; B) rs4759254 – *NACA*; C) rs10735928 – *MSRB3*; and D) rs60791051 – *FAM189A2*.

A. rs2964006 – *GRIA1*

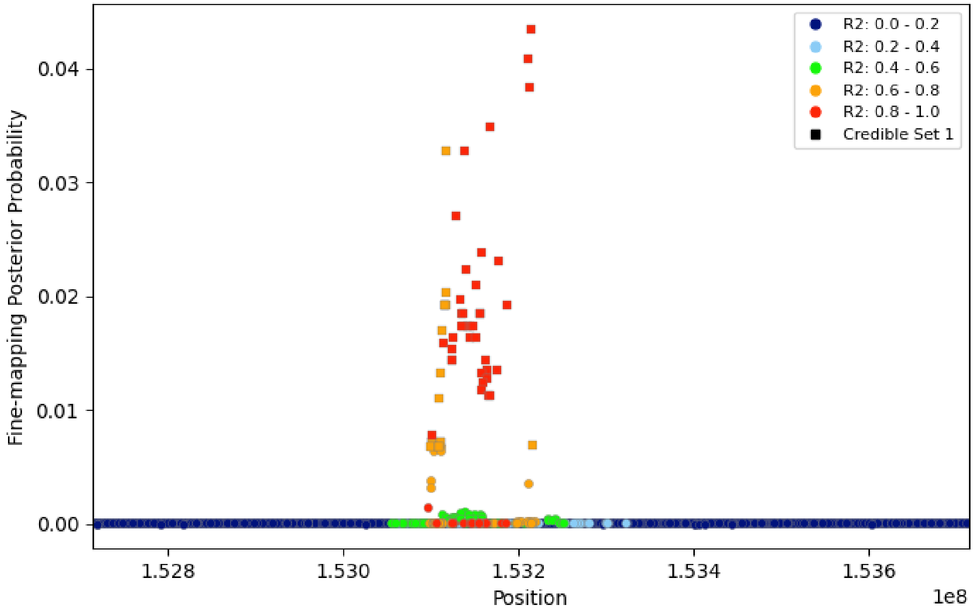


B. rs4759254 – *NACA*

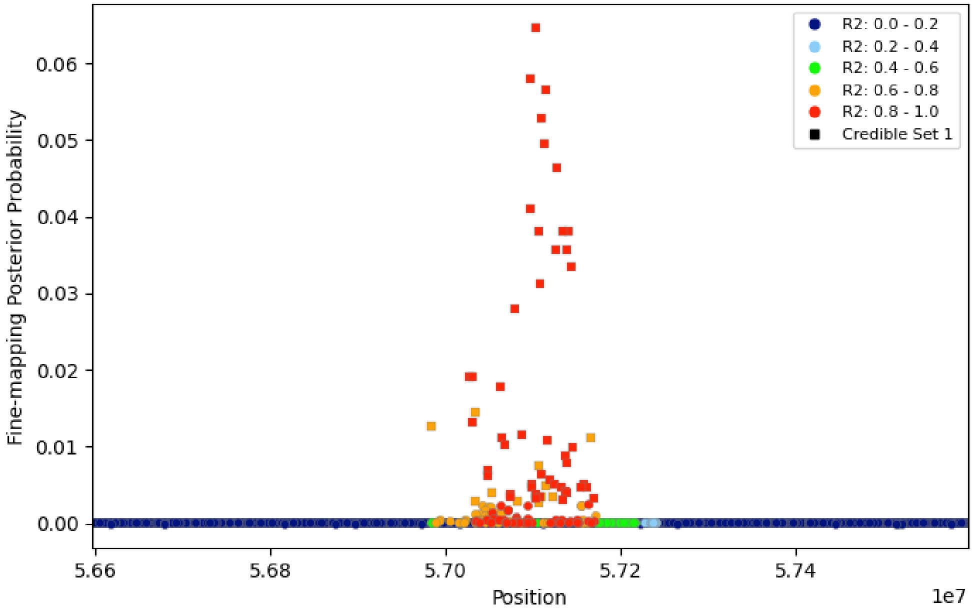

C. rs10735928 – *MSRB3*

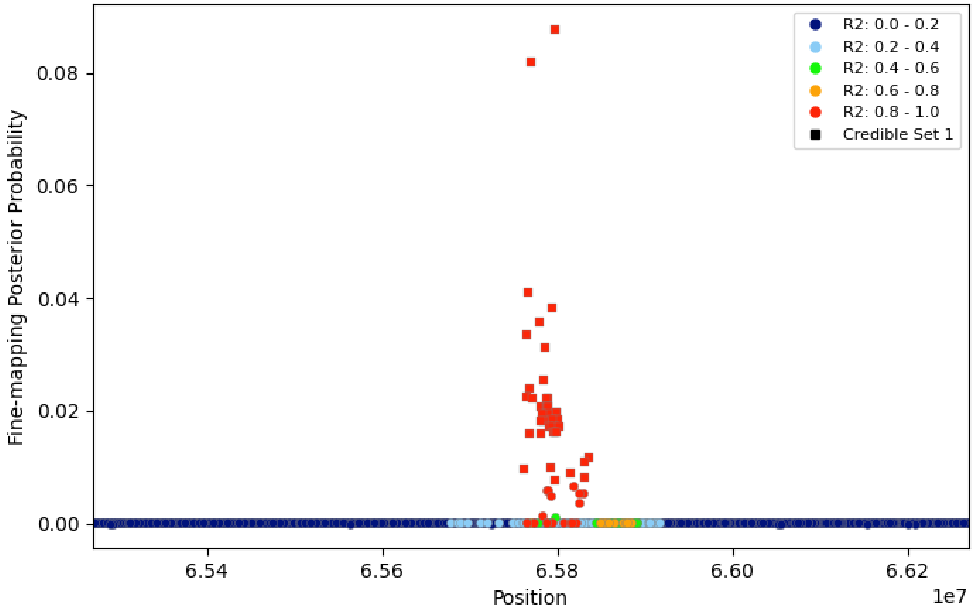


D. rs60791051 – *FAM189A2*

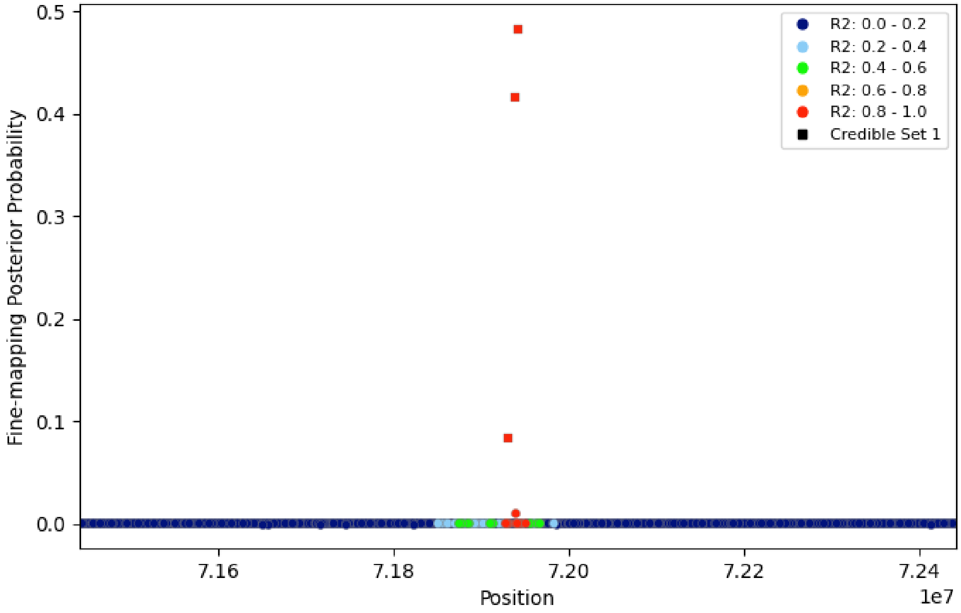

**Supplementary Figure 6. Locuszoom regional association plot for AHI association signal.**
Regional association plots of GWAS signals from stage 1 plus stage 2 meta-analysis. Unrelated individuals of European genetic similarity from the MyCode study were used to calculate LD displayed in plot.

rs1200596362 – *SLC39A10*

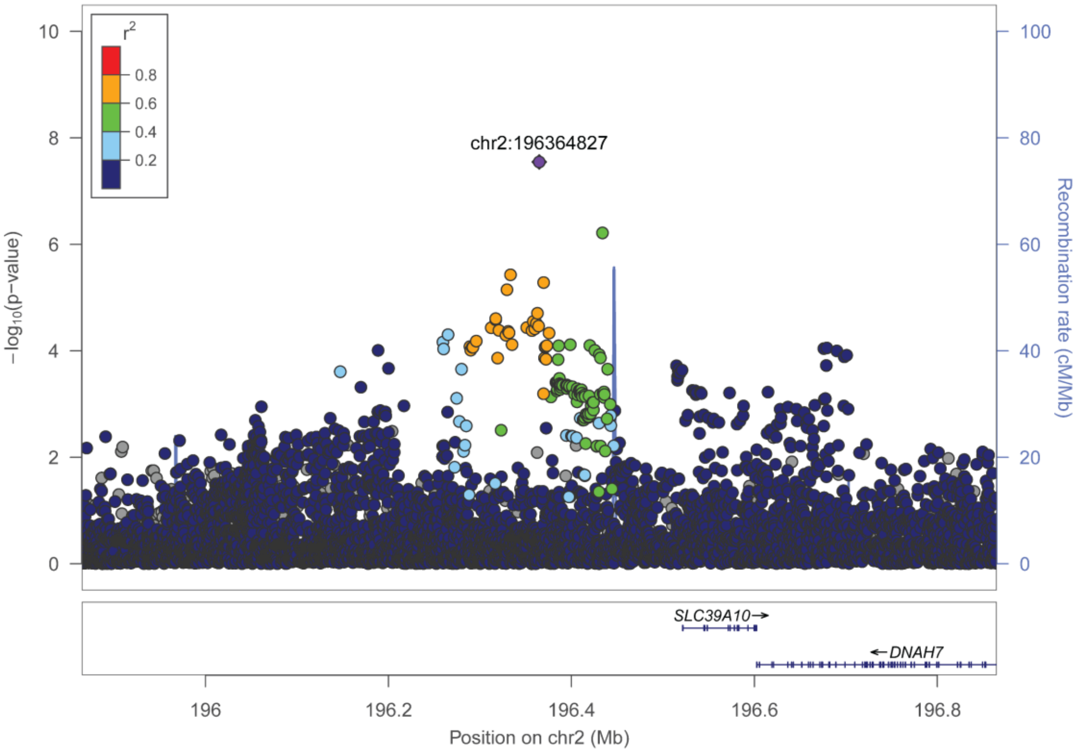

**Supplementary Figure 7. Locuszoom regional association plot for AHIadjBMI association signal.** Regional association plots of GWAS signals from stage 1 plus stage 2 meta-analysis. Unrelated individuals of European genetic similarity from the MyCode study were used to calculate LD displayed in plot.

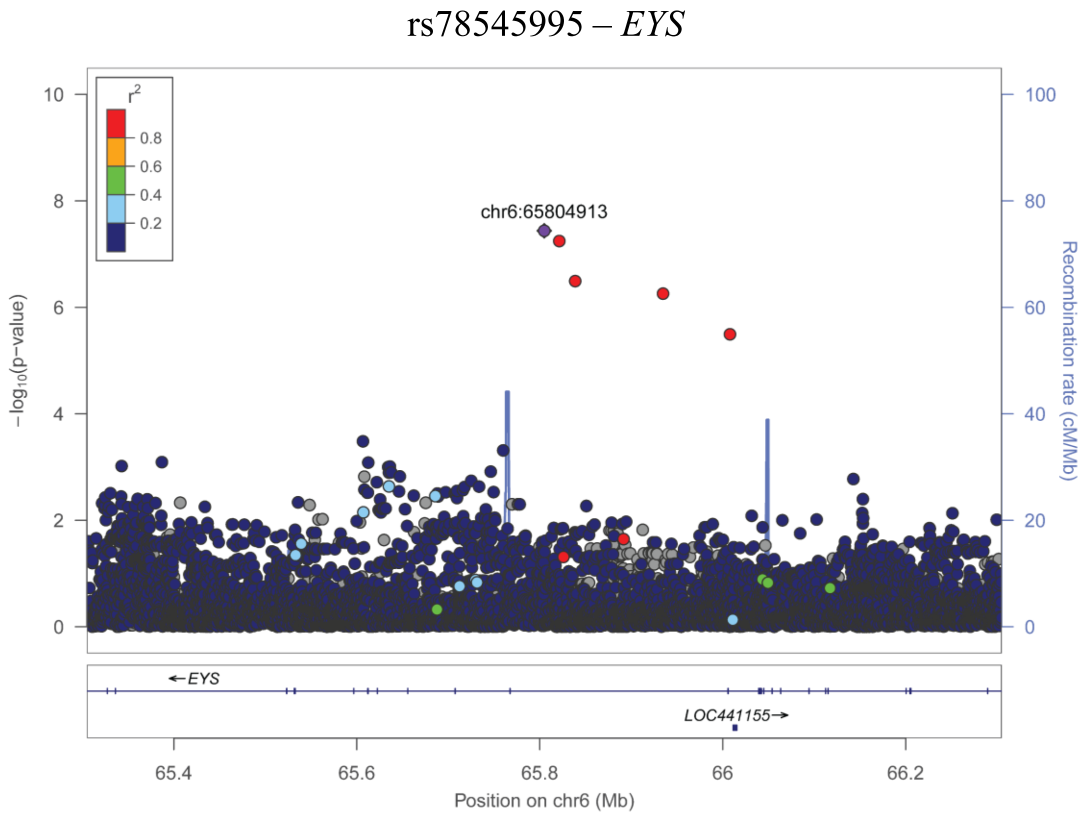

**Supplementary Figure 8**. **Locuszoom regional association plots for avgSaO$_2$ association signals.** Regional association plots of GWAS signals from stage 1 plus stage 2 meta-analyses. The plots appear in order of chromosome and position, strata (pooled vs. European-only), and presentation in Table 1. Unrelated individuals of European genetic similarity from the MyCode study were used to calculate LD displayed in plot. A) rs802729 – *PTPRK*; B) rs2194955 – *GFRA2*.

A. rs802729 – *PTPRK*

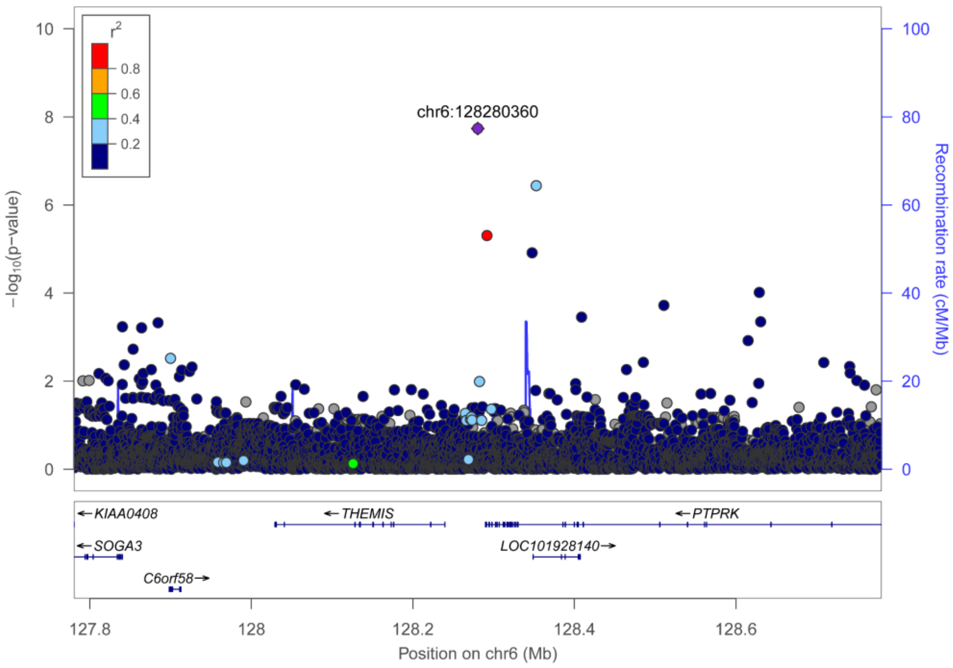


B. rs2194955 – *GFRA2*

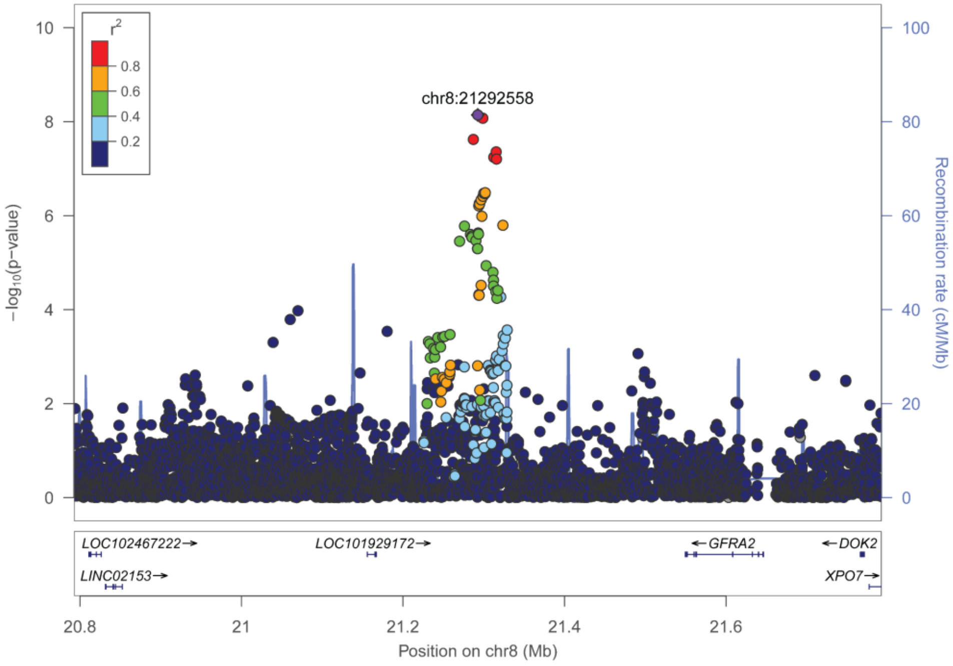

**Supplementary Figure 9**. **Locuszoom regional association plots for avgSaO$_2$adjBMI association signal.** Regional association plots of GWAS signals from stage 1 plus stage 2 meta-analysis. Unrelated individuals of European genetic similarity from the MyCode study were used to calculate LD displayed in plot.

rs79914264 – *PTPRU*

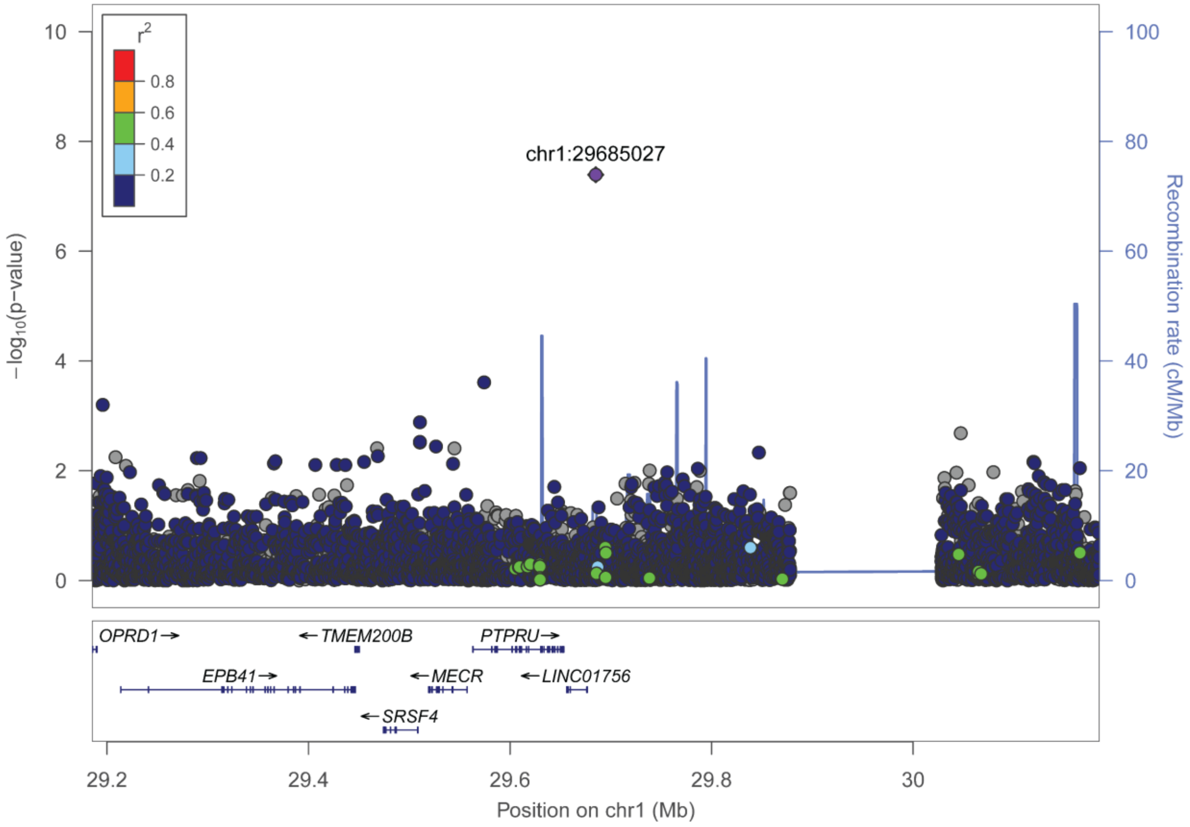

**Supplementary Figure 10**. **Fine-mapping regional plots for AHI association signals.**
Regional plots of posterior probability (prob) from fine-mapping analysis conducted in FINEMAP, including all variants ± 500 kb from index variant for each association region. Unrelated individuals of European genetic similarity from the MyCode study were used to calculate LD displayed in plot.

rs1200596362 – *SLC39A10*

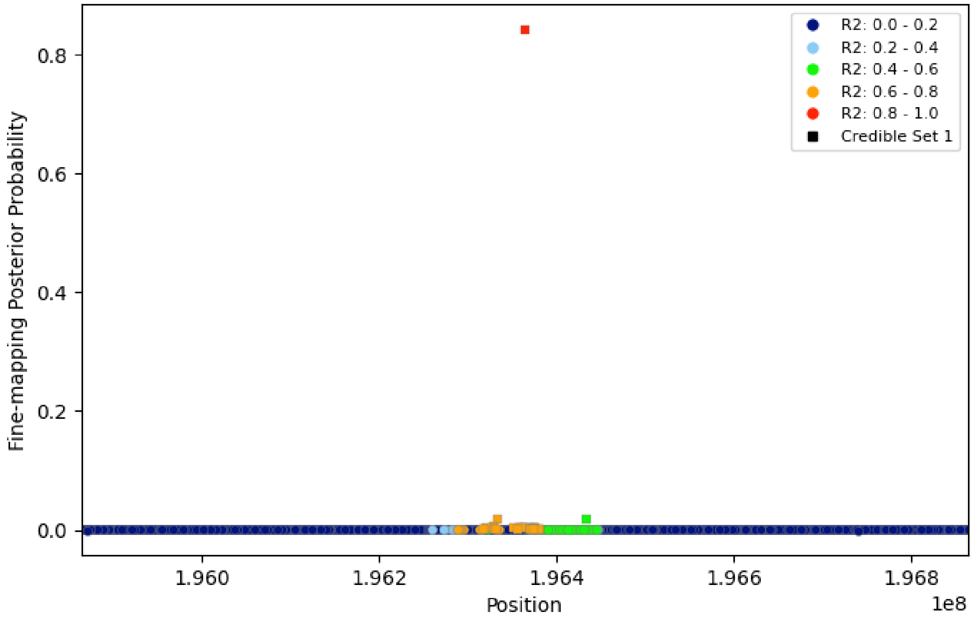

**Supplementary Figure 11**. **Fine-mapping regional plots for AHIadjBMI association signals.** Regional plots of posterior probability (prob) from fine-mapping analysis conducted in FINEMAP, including all variants ± 500 kb from index variant for each association region. Unrelated individuals of European genetic similarity from the MyCode study were used to calculate LD displayed in plot.

rs78545995 – *EYS*

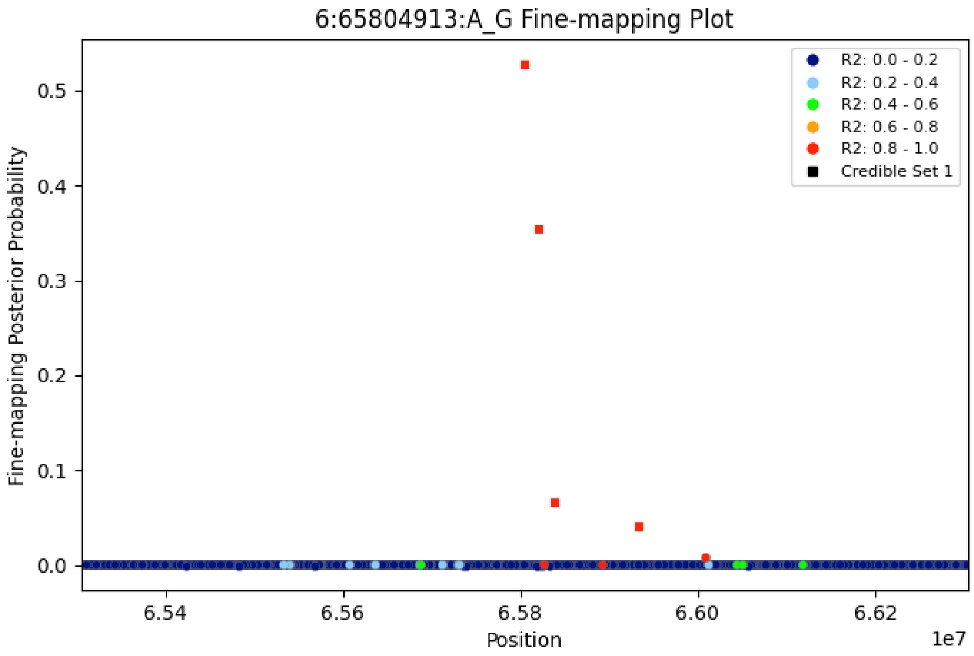

**Supplementary Figure 12**. **Fine-mapping regional plots for avgSaO$_2$ association signals.**
Regional plots of posterior probability (prob) from fine-mapping analysis conducted in FINEMAP, including all variants ± 500 kb from index variant for each association region. Unrelated individuals of European genetic similarity from the MyCode study were used to calculate LD displayed in plot.
The plots appear in order of presentation in Table 1. 1000 Genomes global reference populations were used to calculate LD displayed in plot. A) rs802729 – *PTPRK*; B) rs2194955 – *GFRA2*.

A. rs802729 – *PTPRK*

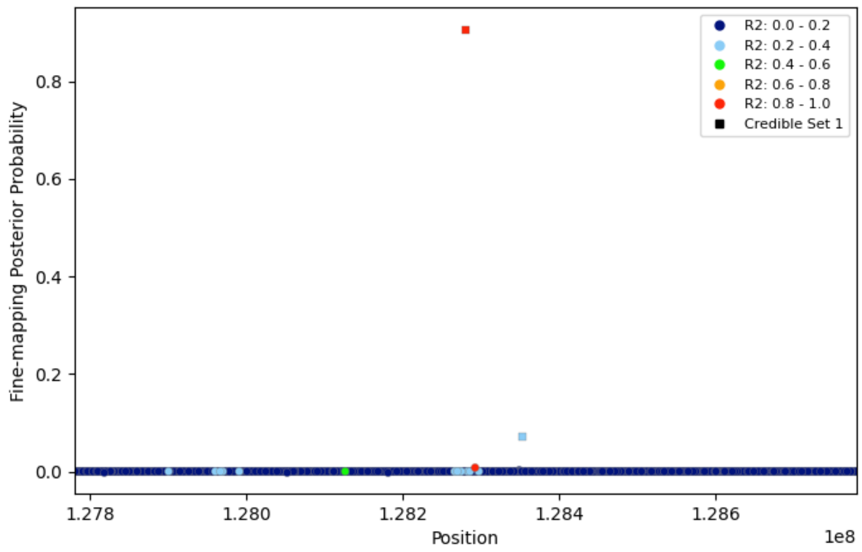


B. rs2194955 – *GFRA2*

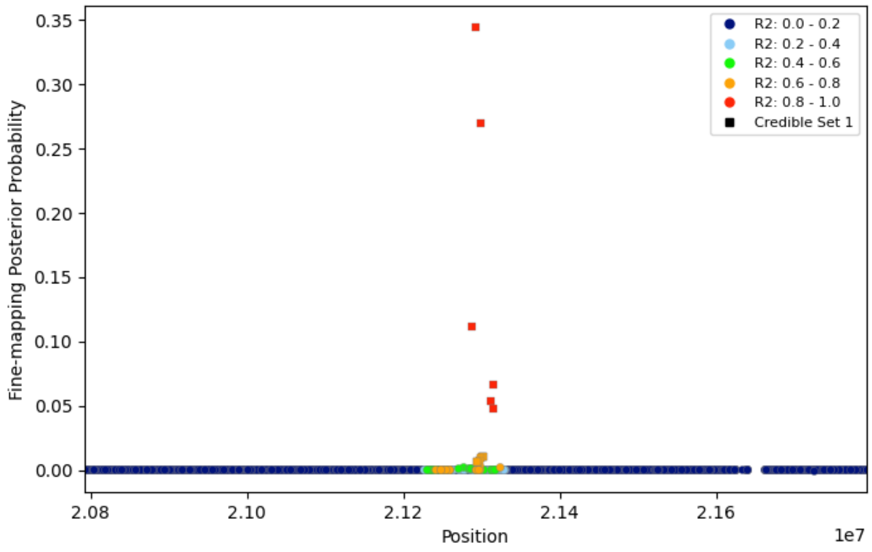

**Supplementary Figure 13**. **Fine-mapping regional plots for avg$SaO_2$adjBMI association signals.** Regional plots of posterior probability (prob) from fine-mapping analysis conducted in FINEMAP, including all variants ± 500 kb from index variant for each association region. Unrelated individuals of European genetic similarity from the MyCode study were used to calculate LD displayed in plot.

rs79914264 – *PTPRU*

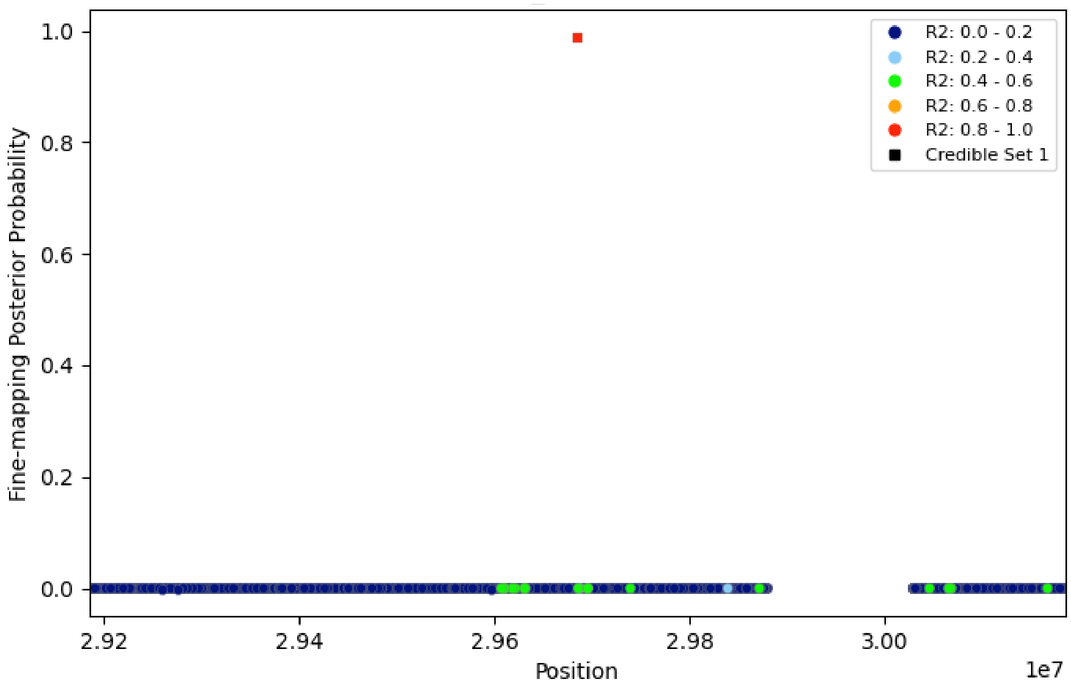

**Supplemental Figure 14.** Significant cis-eQTL colocalization results. A) *FTO*; B) *PTBP2;* C) *GRIA1*. Circles highlight the results of each SNP from the GWAS summary results (colored by LD), while lines highlight the eQTL pattern of association across SNPs (colored by tissue).

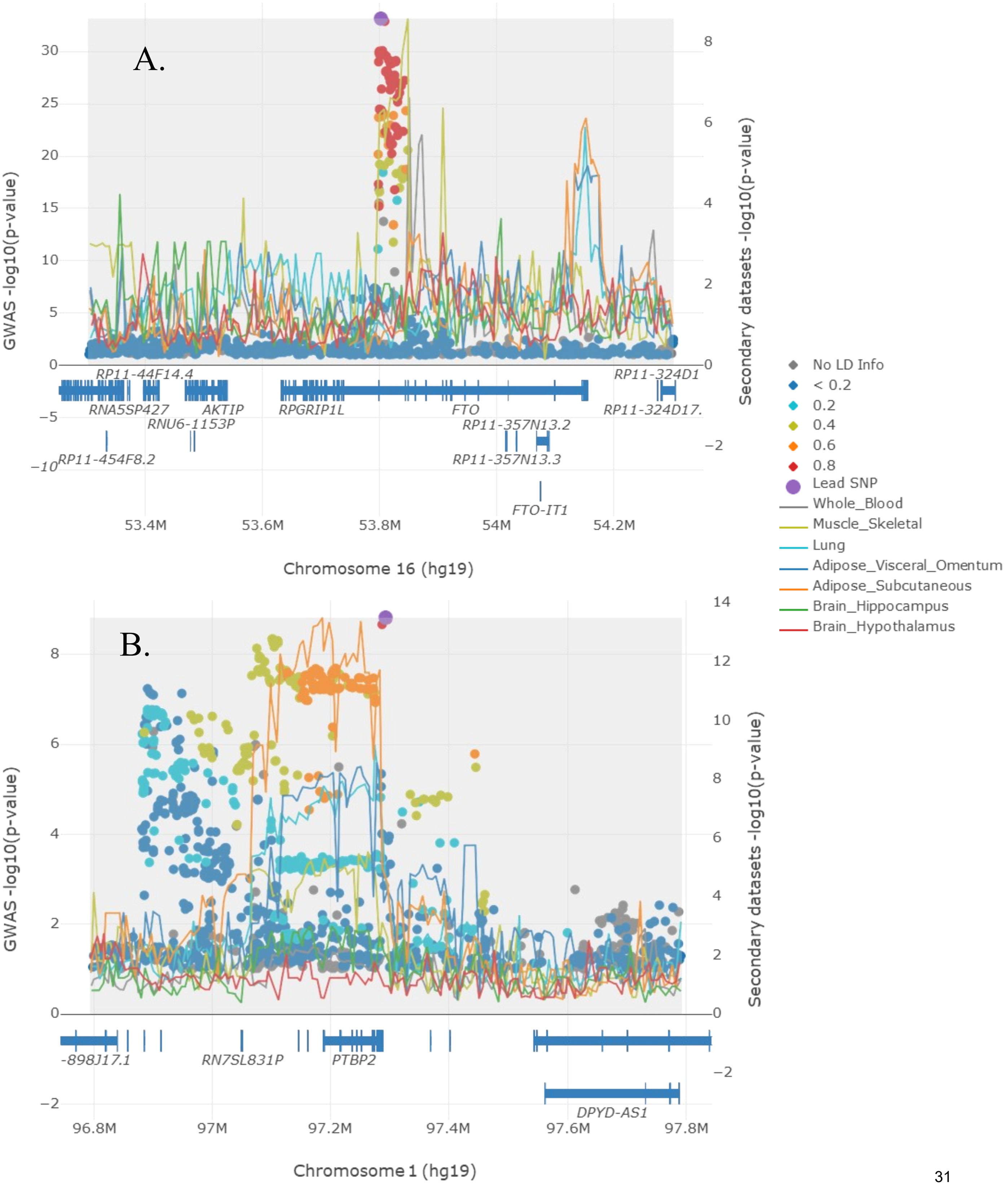

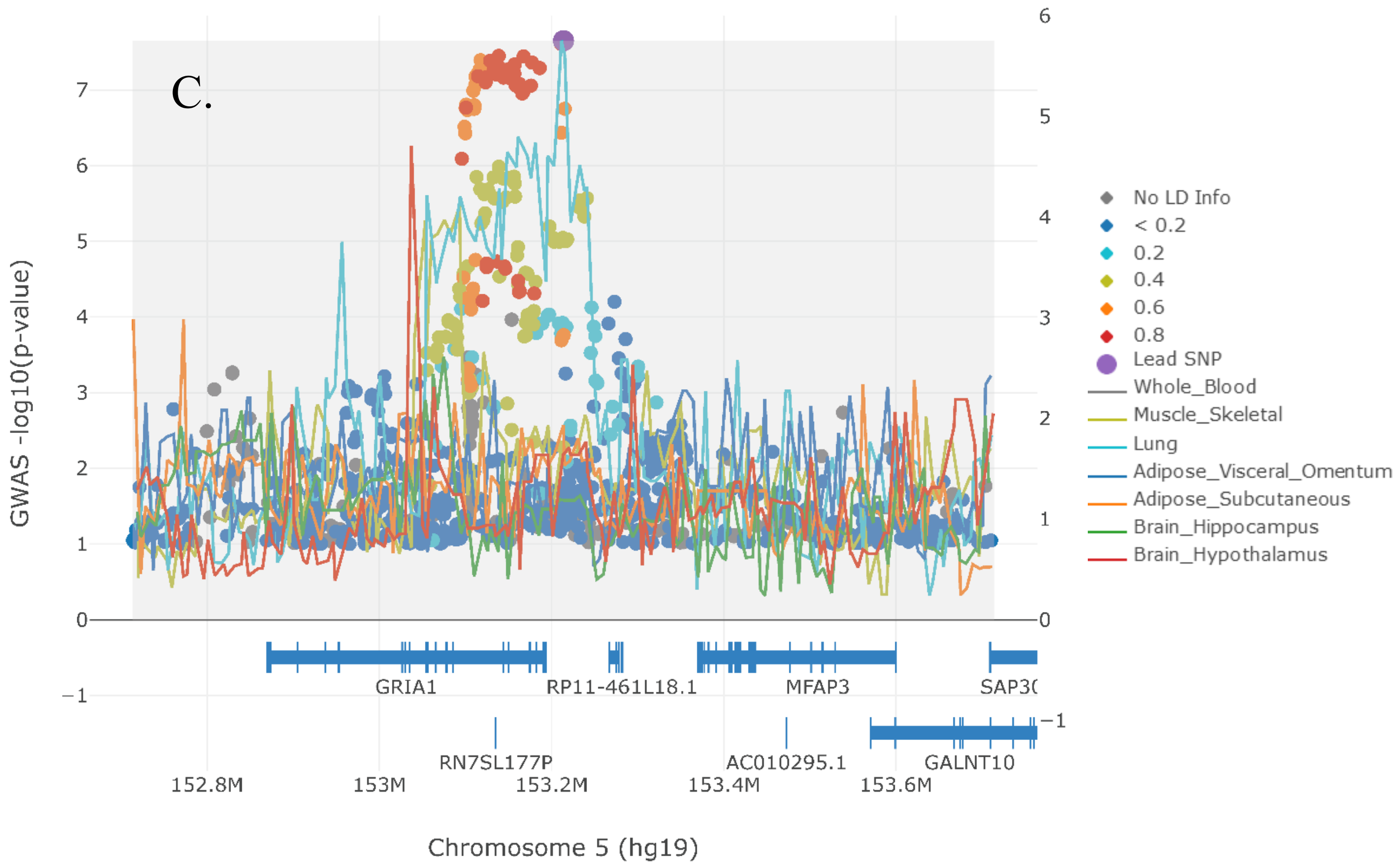
C.
GWAS -log10(p-value)
Chromosome 5 (hg19)
152.8M
153M
153.2M
153.4M
153.6M
GRIA1
RP11-461L18.1
MFAP3
SAP3(
RN7SL177P
AC010295.1
GALNT10
No LD Info
< 0.2
0.2
0.4
0.6
0.8
Lead SNP
Whole_Blood
Muscle_Skeletal
Lung
Adipose_Visceral_Omentum
Adipose_Subcutaneous
Brain_Hippocampus
Brain_Hypothalamus

**Supplementary Table 1.** Phenotype descriptive statistics by study and race/ethnicity.

| Study Abbreviation | Study Full Name | Trait | Race / Ethnicity | N Total | % cases (OSA only) | % Women | Mean BMI (SD) |
|---|---|---|---|---|---|---|---|
| **Stage 1 Studies** | | | | | | | |
| eMERGE | Electronic Medical Records and Genomics Network | AHI | European American / white | 1909 | - | 42.8 | 33.5 (7.58) |
| | | | African American / black | 126 | - | 70.6 | 37.7 (8.64) |
| | | | Hispanic/Latino | 14 | - | 42.9 | 38.4 (7.18) |
| | | | Asian / Asian American | 9 | - | 55.6 | 26.0 (4.91) |
| | | | Other or Mixed | 28 | - | 35.7 | 35.5 (5.38) |
| | | avgSaO$_2$ | European American / white | 1292 | - | 37.6 | 33.0 (7.00) |
| | | | African American / black | 4 | - | 50.0 | 35.0 (4.58) |
| | | | Hispanic/Latino | 7 | - | 14.3 | 34.4 (4.56) |
| | | | Asian / Asian American | 4 | - | 100 | 24.5 (3.64) |
| | | | Other or Mixed | 24 | - | 33.3 | 35.3 (5.40) |
| | | OSA | European American / white | 59411 | 12.1 | 53.9 | 28.4 (6.55) |
| | | | African American / black | 9781 | 8.4 | 66.9 | 30.9 (8.24) |
| | | | Hispanic/Latino | 3375 | 7.9 | 63.5 | 29.6 (6.45) |
| | | | Asian / Asian American | 919 | 4.6 | 58.8 | 25.0 (4.93) |
| | | | Other or Mixed | 6670 | 8.7 | 59.1 | 28.7 (6.87) |
| MyCode | MyCode Community Health Initiative | AHI | European American / white | 6685 | - | 59.7 | 36.7 (8.83) |
| | | | African American / black | 153 | - | 61.4 | 38.6 (9.48) |
| | | | Hispanic/Latino | 165 | - | 70.9 | 39.0 (9.00) |
| | | | Asian / Asian American | 13 | - | 38.5 | 31.1 (5.98) |
| | | | Other or Mixed | 18 | - | 72.2 | 37.5 (7.55) |
| | | avgSaO$_2$ | European American / white | 3250 | - | 58.1 | 36.8 (9.29) |
| | | | African American / black | 70 | - | 57.1 | 39.1 (9.27) |
| | | | Hispanic/Latino | 92 | - | 62 | 39.8 (10.5) |
| | | | Asian / Asian American | 6 | - | 33.3 | 35.4 (3.68) |
| | | | Other or Mixed | 9 | - | 66.7 | 41.0 (8.66) |
| | | OSA | European American / white | 78408 | 20.3 | 61.0 | 31.2 (7.92) |
| | | | African American / black | 1272 | 23.1 | 63.1 | 33.0 (8.77) |
| | | | Hispanic/Latino | 1303 | 21.1 | 70.6 | 32.0 (8.15) |
| | | | Asian / Asian American | 245 | 6.9 | 68.2 | 26.3 (5.21) |
| | | | Other or Mixed | 248 | 18.5 | 64.5 | 30.6 (7.83) |
| **Stage 2 Studies** | | | | | | | |
| BioVU | BioVU | AHI | European American / white | 2184 | - | 49.1 | 33.6 (8.2) |
| | | | African American / black | 427 | - | 66.7 | 38.8 (10.0) |
| | | | Hispanic/Latino | 47 | - | 55.3 | 34.8 (9.3) |
| | | | Asian / Asian American | 18 | - | 50 | 30.1 (8.0) |
| | | | Other or Mixed | 26 | - | 64.2 | 35.3 (8.3) |
| deCODE | deCode Genetics | AHI | European | 12395 | - | 29.8 | 32.7 (6,7) |
| | | OSA | European | 289303 | 4.90% | 49.20 | 26.9 (5.5) |
| KOGES | Korean Genome and Epidemiology Study | AHI | Korean | 2704 | - | 49.8 | 24.6 (2.93) |
| PMBB | Penn Medicine Biobank | AHI | European American / white | 573 | - | 31.2 | 32.6 (7.8) |
| | | | African American / black | 884 | - | 67.9 | 39.3 (10.0) |
| | | | Hispanic/Latino | 41 | - | 46.3 | 36.1 (9.2) |
| | | | Asian / Asian American | 9 | - | 22.2 | 26.4 (3.8) |
| | | | Other or Mixed | 51 | - | 37.3 | 30.3 (6.9) |
| | | avgSaO$_2$ | European American / white | 505 | - | 31.9 | 32.3 (7.7) |
| | | | African American / black | 783 | - | 66.4 | 39.2 (9.9) |
| | | | Hispanic/Latino | 40 | - | 47.5 | 34.8 (8.1) |
| | | | Asian / Asian American | 0 | - | - | - |
| | | | Other or Mixed | 28 | - | 39.3 | 29.8 (6.7) |
| | | OSA | European American / white | 26956 | 13.4 | 46 | 28.4 (6.4) |
| | | | African American / black | 10045 | 21.8 | 63.4 | 32.6 (8.8) |
| | | | Hispanic/Latino | 1111 | 14.8 | 58.7 | 30.2 (7.1) |
| | | | Asian / Asian American | 932 | 4 | 58.7 | 25.3 (4.8) |
| | | | Other or Mixed | 2421 | 9.3 | 43.7 | 28.5 (6.5) |
| SAGIC | Sleep Apnea Global Interdisciplinary Consortium | AHI | European American / white | 1118 | - | 35.7 | 32.4 (7.6) |
| | | | African American / black | 383 | - | 59.3 | 39.5 (10.4) |
| | | | Hispanic/Latino | 52 | - | 63.5 | 35.7 (9.5) |
| | | | Asian / Asian American | 219 | - | 36.1 | 27.9 (5.6) |
| | | | Other or Mixed | 76 | - | 50 | 34.1 (9.1) |
| | | avgSAO$_2$ | European American / white | 847 | - | 37.8 | 32.8 (8.4) |
| | | | African American / black | 292 | - | 59.3 | 40.2 (10.5) |
| | | | Hispanic/Latino | 12 | - | 58.3 | 31.3 (7.5) |
| | | | Asian / Asian American | 182 | - | 33.5 | 27.5 (5.0) |
| | | | Other or Mixed | 40 | - | 45 | 34.0 (10.2) |

**Supplementary Table 2**. Genotype and imputation characteristics by study.

| Study Abbrev | Study Full Name | Study Design | Sample Call Rate | Sample Exclusions | Outcome Ascertainment (measured/sef-report) | References | Genotyping Array |
|---|---|---|---|---|---|---|---|
| **Stage 1 Studies** | | | | | | | |
| eMERGE | Electronic Medical Records and Genomics Network | EHR-linked Biobank | >95% | Sample exclusions variaed by site and array (see PMID: 30298529 for details) | Measured / EHR-derived | PMID: 25414722<br>PMID: 24956226<br>PMID: 3978763/ PMID: 23743551, PMID: 35047833 | Multiple (see PMID: 30298529) |
| MyCode | MyCode Community Health Initiative Study | EHR-linked Biobank | >95% | 1) mismatches between EHR-derived and genetically-determined sex, 2) high rate of heterozygosity (F >0.4), and a low (<95%) call rate. | Measured / EHR-derived | PMIDs: 26866580, 28008009 | Illumina Infinium OMNI v1.2, 1.3, or 1.4; GSA v1 or v2 |
| **Stage 2 Studies** | | | | | | | |
| BioVU | BioVU | EHR-linked Biobank | >95% | 1) mismatches between EHR-derived and genetically-determined sex, 2) unexplained relatedness, and 3) a low (<95%) call rate. | Measured / EHR-derived | | Ilumina MEGA array |
| deCODE | deCode Genetics | EHR-linked Biobank | ≥ 95% | Non-Icelandic individuals | Measured | | Illumina HumanHap and Omni chips |
| KOGES | Korean Genome and Epidemiology Study | population-based | >95% | 1) missing AHI phenotype; 2) low (<95%) call rate | measured | PMIDs: 27439680, 39952983 | Affymetrix Genome Wide Human SNP Array 5.0 |
| SAGIC | Sleep Apnea Global Interdisciplinary Consortium | Patient-sample | >97% | 1) extreme heterozygosity (more than 3SD from mean; 2) sex mismatch; 3) 1st or 2nd relatives | measured | | GSA-24v2-0_A1 and Multi-EthnicGlobal_D1 |
| PMBB | Penn Medicine BioBank | Patient-sample | >90% | 1) sex mismatch; 2) relatives up to 1st cousins | measured | | Illumina Infinium OMNI, GSA_V1, GSA_V2 |

**Supplementary Table 2**. Geno

| Study Abbrev | Software for PCs | Software for Phenotpe Algorithm | SNP exclusions | Phasing | Imputation Software | Imputation Reference Panel |
|---|---|---|---|---|---|---|
| **Stage 1 Studies** | | | | | | |
| eMERGE | Eigenstrat | Data are from the medical record, STATA/SE v15 was used for processing | Missing >2% | BEAGLE | Michaigan Imputation Server | HRC |
| MyCode | Eigenstrat | Data are from the medical record, STATA/SE v15 was used for processing | 1) call rate (<95%), HWE P-val < 1 x 10-15. | Eagle v2.4 | Michigan Imputation Server (Minimac4 1.4.1) | 1KGP3 Global Reference Panel |
| **Stage 2 Studies** | | | | | | |
| BioVU | PLINK | STATA | 1) call rate (<95%), 2) Het Excess < -0.3 or greater than 0.2. | Eagle v2.4 | Michaigan Imputation Server | 1KGP3 Global Reference Panel |
| deCODE | NA | | 1) call rate (<98%), HWE P-val < 1 x 10-6. | deCode long-range phasing software | deCode impudation software | WGS 49,708 Icelandic individuals |
| KOGES | Eigenstrat | SAS ver. 9.4 and R ver. 4.2.3 were used for processing | 1) call rate <95%; 2) MAF<0.01; 5) HWE P-val < 1e-7 | | IMPUTE (v2.644) | 1000 Genomes Phase I (version 3) in NCBI build 37 (hg19) |
| SAGIC | Eigenstrat | Data are from the medical record, STATA/SE v14 was used for processing | 1) indels; 2) call rate <95%; 3) chrM and Y; 4) MAF<0.01; 5) HWE P-val < 1e-6 | Eagle v2.4 | Michigan Imputation Server (Minimac4 1.4.1) | 1KGP3 (Other/Mixed) |
| PMBB | Eigenstrat | Data are from the medical record, STATA/SE v14 was used for processing | 1) call rate < 95%; 2) chrM and Y | EAGLE | TOPMed Imputation Server | TOPMed R2 |

**Supplementary Table 3.** Association results for OSA-associated SNPs across all analyses and stages.

| | | | | | | | | **Pooled Analysis Stage 1 + Stage 2** | | | | | | **EUR Analysis Stage 1 + Stage 2** | | | | | | **Pooled Analysis Stage 1** | | | | | |
|---|---|---|---|---|---|---|---|---|---|---|---|---|---|---|---|---|---|---|---|---|---|---|---|---|---|
| **Locus #** | **Discovery Analysis** | **dbSNP ID** | **CHR** | **POS** | **Nearest Gene** | **A1** | **A2** | **EAF** | **BETA** | **SE** | **P** | **Het $I^2$** | **N** | **EAF** | **BETA** | **SE** | **P** | **Het $I^2$** | **N** | **EAF** | **BETA** | **SE** | **P** | **Het $I^2$** | **N** |
| **Model 1 Results** | | | | | | | | | | | | | | | | | | | | | | | | | |
| 1 | a.m1.s1 | rs12725199 | 1 | 97293792 | *PTPB2* | A | C | 0.253 | 0.055 | 0.009 | **1.54E-09** | 0 | 492,107 | 0.267 | 0.057 | 0.009 | **1.45E-09** | 0 | 451,352 | 0.246 | 0.061 | 0.012 | *3.54E-07* | 44.5 | 161,632 |
| 1 | e.m1.s1 | rs12740489 | 1 | 97296935 | *PTPB2* | A | G | 0.253 | 0.055 | 0.009 | **1.72E-09** | 0 | 492,107 | 0.267 | 0.057 | 0.009 | **1.05E-09** | 0 | 451,402 | 0.246 | 0.060 | 0.012 | *4.26E-07* | 46.2 | 161,632 |
| 2 | e.m1.s1 | rs12467202 | 2 | 104601968 | Intergenic | T | C | 0.893 | -0.070 | 0.013 | **3.49E-08** | 0 | 492,107 | 0.910 | -0.087 | 0.014 | **1.43E-09** | 0 | 451,632 | 0.896 | -0.077 | 0.017 | *6.81E-06* | 72.1 | 161,632 |
| 2 | a.m1.s1 | rs2376073 | 2 | 104662008 | Intergenic | T | C | 0.112 | 0.070 | 0.013 | **3.03E-08** | 0 | 492,107 | 0.091 | 0.081 | 0.014 | **1.31E-08** | 0 | 449,338 | 0.108 | 0.078 | 0.017 | *5.37E-06* | 38.1 | 161,632 |
| 3 | a.m1.s1 | rs9831061 | 3 | 185835281 | *ETV5* | A | G | 0.726 | 0.051 | 0.009 | **1.21E-08** | 0 | 492,107 | 0.752 | 0.050 | 0.010 | 2.63E-07 | 0 | 452,699 | 0.735 | 0.055 | 0.012 | *3.77E-06* | 0 | 161,632 |
| 4 | e.m1.s1 | rs34592089 | 4 | 102926923 | *BANK1* | A | G | 0.055 | 0.097 | 0.019 | 1.87E-07 | 59.5 | 492,107 | 0.055 | 0.113 | 0.020 | **1.81E-08** | 0 | 449,087 | 0.060 | 0.080 | 0.022 | 2.44E-04 | 0 | 161,632 |
| 5 | a.m2.s1 | rs2964006 | 5 | 153213880 | *GRIA1* | T | G | 0.447 | 0.042 | 0.008 | 1.26E-07 | 0 | 492,107 | 0.466 | 0.041 | 0.008 | 1.12E-06 | 0 | 453,015 | 0.448 | 0.043 | 0.010 | *2.25E-05* | 0 | 161,632 |
| 6 | a.m1.s2 | rs9380814 | 6 | 38934922 | *DNAH8* | A | G | 0.324 | -0.048 | 0.009 | **1.82E-08** | 56.2 | 492,107 | 0.344 | -0.049 | 0.009 | **2.24E-08** | 65.5 | 453,433 | 0.317 | -0.038 | 0.011 | 6.34E-04 | 0 | 161,632 |
| 6 | e.m1.s2 | rs1537230 | 6 | 38944196 | *DNAH8* | A | G | 0.324 | -0.047 | 0.009 | **3.18E-08** | 55.2 | 492,107 | 0.345 | -0.050 | 0.009 | **1.63E-08** | 63.6 | 453,528 | 0.317 | -0.037 | 0.011 | 8.71E-04 | 0 | 161,632 |
| 7 | a.m1.s1 | rs66474734 | 8 | 114661935 | *CSMD3* | T | TATA | 0.582 | -0.062 | 0.011 | **2.47E-08** | 0 | 122,648 | 0.612 | -0.061 | 0.012 | 4.35E-07 | 0 | 102,742 | 0.601 | -0.062 | 0.013 | *3.08E-06* | 0 | 81,476 |
| 8 | a.m1.s1 + a.m1.s2 | rs12553324 | 9 | 23347865 | Intergenic | C | G | 0.624 | -0.052 | 0.009 | **3.50E-08** | 74.5 | 202,804 | 0.604 | -0.053 | 0.010 | 1.44E-07 | **82.9** | 164,716 | 0.618 | -0.043 | 0.011 | *4.26E-05* | 45.1 | 161,632 |
| 9 | e.m2.s1 | rs60791051 | 9 | 71941742 | Intergenic | A | C | 0.929 | -0.080 | 0.021 | 1.71E-04 | 51.9 | 122,648 | 0.938 | -0.120 | 0.024 | 6.72E-07 | 0 | 105,305 | 0.940 | -0.104 | 0.027 | 1.25E-04 | 0 | 81,476 |
| 10 | a.m2.s1 | | 12 | 57096674 | *NACA* | T | C | 0.641 | -0.051 | 0.008 | **9.49E-10** | 19.7 | 492,107 | 0.666 | -0.048 | 0.009 | 7.72E-08 | 0 | 454,019 | 0.637 | -0.059 | 0.011 | *6.12E-08* | 0 | 161,632 |
| 10 | e.m1.s1 | rs2926743 | 12 | 57114100 | *NACA* | A | G | 0.246 | 0.054 | 0.009 | **4.30E-09** | 25.5 | 492,107 | 0.258 | 0.054 | 0.010 | **9.14E-09** | 0 | 454,019 | 0.257 | 0.062 | 0.012 | *9.52E-08* | 0 | 161,632 |
| 10 | a.m1.s1 | rs2958127 | 12 | 57114307 | *NACA* | A | T | 0.380 | 0.050 | 0.008 | **5.75E-10** | 0 | 492,107 | 0.363 | 0.044 | 0.009 | 2.79E-07 | 0 | 454,019 | 0.382 | 0.054 | 0.011 | *3.35E-07* | 0 | 161,632 |
| 10 | e.m2.s1 | rs12227117 | 12 | 57166413 | *NACA* | A | G | 0.250 | 0.060 | 0.011 | **2.09E-08** | 0 | 202,804 | 0.268 | 0.062 | 0.011 | **4.42E-08** | 39.8 | 164,716 | 0.256 | 0.059 | 0.012 | *6.84E-07* | 0 | 161,632 |
| 11 | a.m2.s2 | rs10735928 | 12 | 65769626 | *MSRB3* | A | C | 0.382 | 0.038 | 0.008 | 2.58E-06 | 59 | 492,107 | 0.381 | 0.040 | 0.009 | 2.71E-06 | 62.9 | 454,019 | 0.381 | 0.027 | 0.010 | 9.18E-03 | 0.7 | 161,632 |
| 11 | e.m2.s2 | rs10878270 | 12 | 65792175 | *MSRB3* | A | G | 0.605 | -0.037 | 0.008 | 4.79E-06 | 59.4 | 492,107 | 0.618 | -0.041 | 0.009 | 1.67E-06 | 66.8 | 454,019 | 0.607 | -0.026 | 0.011 | 1.29E-02 | 0 | 161,632 |
| 11 | e.m2.s1 | rs4762080 | 12 | 65796310 | *MSRB3* | A | T | 0.208 | 0.046 | 0.010 | 2.68E-06 | 0 | 492,107 | 0.221 | 0.049 | 0.010 | 1.74E-06 | 0 | 454,019 | 0.212 | 0.040 | 0.013 | 1.98E-03 | 0 | 161,632 |
| 12 | a.m1.s1 + a.m1.s2 | rs11642015 | 16 | 53802494 | *FTO* | T | C | 0.396 | 0.098 | 0.008 | **8.47E-34** | 44.3 | 492,107 | 0.420 | 0.100 | 0.008 | **1.82E-32** | 56.6 | 454,019 | 0.399 | 0.107 | 0.010 | **1.39E-24** | 0 | 161,632 |
| 12 | e.m1.s1 + e.m1.s2 | rs56094641 | 16 | 53806453 | *FTO* | A | G | 0.604 | -0.098 | 0.008 | **9.20E-34** | 54.7 | 492,107 | 0.580 | -0.100 | 0.008 | **1.36E-32** | 61.2 | 454,019 | 0.602 | -0.108 | 0.010 | **6.71E-25** | 0 | 161,632 |
| 13 | a.m1.s1 + e.m1.s1 | rs1800437 | 19 | 46181392 | *GIPR* | C | G | 0.205 | -0.059 | 0.010 | **2.23E-09** | 64.3 | 492,107 | 0.213 | -0.058 | 0.010 | **1.92E-08** | 47.4 | 454,019 | 0.202 | -0.073 | 0.013 | **1.55E-08** | 0 | 161,632 |
| 14 | a.m1.s1 + e.m1.s1 | rs11088984 | 21 | 46670045 | *POFUT2* | A | C | 0.667 | 0.051 | 0.009 | **3.94E-09** | 0 | 492,107 | 0.650 | 0.051 | 0.009 | **1.48E-08** | 0 | 454,019 | 0.666 | 0.052 | 0.011 | *4.70E-06* | 0 | 161,632 |
| **Model 2 Results** | | | | | | | | | | | | | | | | | | | | | | | | | |
| 1 | a.m1.s1 | rs12725199 | 1 | 97293792 | *PTPB2* | A | C | 0.251 | 0.038 | 0.010 | 1.54E-04 | 21.4 | 266,865 | 0.266 | 0.042 | 0.010 | 4.74E-05 | 8.1 | 237,586 | 0.244 | 0.047 | 0.013 | 2.38E-04 | 55.1 | 139,337 |
| 1 | e.m1.s1 | rs12740489 | 1 | 97296935 | *PTPB2* | A | G | 0.251 | 0.038 | 0.010 | 1.20E-04 | 19.1 | 266,921 | 0.266 | 0.042 | 0.010 | 3.82E-05 | 5.6 | 237,636 | 0.244 | 0.047 | 0.013 | 2.17E-04 | 58.6 | 139,393 |
| 2 | e.m1.s1 | rs12467202 | 2 | 104601968 | Intergenic | T | C | 0.898 | -0.053 | 0.014 | 2.22E-04 | 0 | 267,073 | 0.911 | -0.071 | 0.016 | 6.87E-06 | 0 | 237,851 | 0.904 | -0.064 | 0.019 | 6.97E-04 | 26.4 | 139,545 |
| 2 | a.m1.s1 | rs2376073 | 2 | 104662008 | Intergenic | T | C | 0.104 | 0.051 | 0.014 | 3.13E-04 | 19.1 | 264,560 | 0.090 | 0.064 | 0.016 | 5.54E-05 | 0 | 235,577 | 0.094 | 0.065 | 0.019 | 6.05E-04 | 0 | 137,032 |
| 3 | a.m1.s1 | rs9831061 | 3 | 185835281 | *ETV5* | A | G | 0.731 | 0.029 | 0.010 | 3.71E-03 | 0 | 268,121 | 0.753 | 0.028 | 0.011 | 7.76E-03 | 0 | 238,912 | 0.742 | 0.035 | 0.013 | 6.36E-03 | 0 | 140,593 |
| 4 | e.m1.s1 | rs34592089 | 4 | 102926923 | *BANK1* | A | G | 0.053 | 0.068 | 0.022 | 2.11E-03 | 0 | 176,681 | 0.059 | 0.071 | 0.023 | 2.11E-03 | 0 | 147,350 | 0.053 | 0.059 | 0.025 | 1.68E-02 | 0 | 137,108 |
| 5 | a.m2.s1 | rs2964006 | 5 | 153213880 | *GRIA1* | T | G | 0.450 | 0.049 | 0.009 | **2.24E-08** | 0 | 268,537 | 0.466 | 0.048 | 0.009 | 8.55E-08 | 0 | 239,234 | 0.453 | 0.047 | 0.011 | *2.00E-05* | 0 | 141,009 |
| 6 | a.m1.s2 | rs9380814 | 6 | 38934922 | *DNAH8* | A | G | 0.326 | -0.035 | 0.009 | 1.52E-04 | 14.5 | 269,014 | 0.344 | -0.036 | 0.010 | 1.74E-04 | 37.5 | 239,648 | 0.322 | -0.028 | 0.012 | 1.92E-02 | 0 | 141,486 |
| 6 | e.m1.s2 | rs1537230 | 6 | 38944196 | *DNAH8* | A | G | 0.326 | -0.035 | 0.009 | 1.70E-04 | 8.5 | 269,094 | 0.344 | -0.037 | 0.010 | 1.35E-04 | 36 | 239,740 | 0.322 | -0.028 | 0.012 | 1.93E-02 | 0 | 141,566 |
| 7 | a.m1.s1 | rs66474734 | 8 | 114661935 | *CSMD3* | T | TATA | 0.587 | -0.057 | 0.012 | 2.55E-06 | 0 | 117,980 | 0.613 | -0.053 | 0.013 | 4.32E-05 | 0 | 101,470 | 0.606 | -0.053 | 0.014 | 1.58E-04 | 0 | 78,407 |
| 8 | a.m1.s1 + a.m1.s2 | rs12553324 | 9 | 23347865 | Intergenic | C | G | 0.620 | -0.049 | 0.010 | 1.55E-06 | **81.7** | 181,665 | 0.604 | -0.047 | 0.011 | 1.16E-05 | **84.2** | 152,274 | 0.614 | -0.038 | 0.011 | 7.50E-04 | 0 | 142,092 |
| 9 | e.m2.s1 | rs60791051 | 9 | 71941742 | Intergenic | A | C | 0.930 | -0.109 | 0.023 | 1.99E-06 | 0 | 120,690 | 0.939 | -0.139 | 0.026 | **4.80E-08** | 0 | 104,016 | 0.940 | -0.122 | 0.029 | *1.80E-05* | 0 | 81,117 |

| | | | | | | | | Pooled Analysis Stage 1 + Stage 2 | | | | | | EUR Analysis Stage 1 + Stage 2 | | | | | | Pooled Analysis Stage 1 | | | | | |
|---|---|---|---|---|---|---|---|---|---|---|---|---|---|---|---|---|---|---|---|---|---|---|---|---|---|
| **Locus #** | **Discovery Analysis** | **dbSNP ID** | **CHR** | **POS** | **Nearest Gene** | **A1** | **A2** | **EAF** | **BETA** | **SE** | **P** | **Het $I^2$** | **N** | **EAF** | **BETA** | **SE** | **P** | **Het $I^2$** | **N** | **EAF** | **BETA** | **SE** | **P** | **Het $I^2$** | **N** |
| 10 | a.m2.s1 | | 12 | 57096674 | *NACA* | T | C | 0.644 | -0.055 | 0.009 | **2.99E-09** | **75.2** | 269,620 | 0.666 | -0.054 | 0.010 | **2.81E-08** | 54.4 | 240,229 | 0.644 | -0.070 | 0.012 | **3.51E-09** | 0 | 142,092 |
| 10 | e.m1.s1 | rs2926743 | 12 | 57114100 | *NACA* | A | G | 0.248 | 0.055 | 0.010 | **3.93E-08** | 72.1 | 269,620 | 0.259 | 0.057 | 0.010 | **3.04E-08** | 51 | 240,229 | 0.260 | 0.069 | 0.012 | **3.36E-08** | 0 | 142,092 |
| 10 | a.m1.s1 | rs2958127 | 12 | 57114307 | *NACA* | A | T | 0.378 | 0.053 | 0.009 | **4.33E-09** | 27.7 | 269,620 | 0.364 | 0.050 | 0.009 | 9.02E-08 | 0 | 240,229 | 0.378 | 0.061 | 0.011 | *8.75E-08* | 0 | 142,092 |
| 10 | e.m2.s1 | rs12227117 | 12 | 57166413 | *NACA* | A | G | 0.253 | 0.065 | 0.012 | **1.83E-08** | 0 | 181,665 | 0.268 | 0.069 | 0.012 | **9.12E-09** | 0 | 152,274 | 0.259 | 0.066 | 0.013 | *1.61E-07* | 0 | 142,092 |
| 11 | a.m2.s2 | rs10735928 | 12 | 65769626 | *MSRB3* | A | C | 0.382 | 0.054 | 0.009 | **7.73E-10** | 57.1 | 269,620 | 0.381 | 0.055 | 0.009 | **4.29E-09** | 39.5 | 240,229 | 0.381 | 0.044 | 0.011 | 1.07E-04 | 0 | 142,092 |
| 11 | e.m2.s2 | rs10878270 | 12 | 65792175 | *MSRB3* | A | G | 0.607 | -0.051 | 0.009 | **6.74E-09** | 49.7 | 269,620 | 0.618 | -0.056 | 0.009 | **2.40E-09** | 46.7 | 240,229 | 0.610 | -0.041 | 0.011 | 2.51E-04 | 0 | 142,092 |
| 11 | e.m2.s1 | rs4762080 | 12 | 65796310 | *MSRB3* | A | T | 0.210 | 0.059 | 0.011 | 6.52E-08 | 0 | 269,620 | 0.221 | 0.064 | 0.011 | **1.11E-08** | 0 | 240,229 | 0.216 | 0.058 | 0.014 | *2.81E-05* | 0 | 142,092 |
| 12 | a.m1.s1 + a.m1.s2 | rs11642015 | 16 | 53802494 | *FTO* | T | C | 0.399 | 0.010 | 0.009 | 2.61E-01 | **89.3** | 269,620 | 0.420 | 0.010 | 0.009 | 2.68E-01 | **90.4** | 240,229 | 0.404 | 0.031 | 0.011 | 5.71E-03 | 0 | 142,092 |
| 12 | e.m1.s1 + e.m1.s2 | rs56094641 | 16 | 53806453 | *FTO* | A | G | 0.601 | -0.010 | 0.009 | 2.86E-01 | **90.1** | 269,620 | 0.580 | -0.010 | 0.009 | 2.78E-01 | **90.9** | 240,229 | 0.596 | -0.031 | 0.011 | 5.26E-03 | 0 | 142,092 |
| 13 | a.m1.s1 + e.m1.s1 | rs1800437 | 19 | 46181392 | *GIPR* | C | G | 0.206 | -0.032 | 0.011 | 2.78E-03 | 51.8 | 269,620 | 0.213 | -0.032 | 0.011 | 4.61E-03 | 22.5 | 240,229 | 0.204 | -0.045 | 0.014 | 1.19E-03 | 40.9 | 142,092 |
| 14 | a.m1.s1 + e.m1.s1 | rs11088984 | 21 | 46670045 | *POFUT2* | A | C | 0.665 | 0.044 | 0.010 | 3.90E-06 | 0 | 269,620 | 0.650 | 0.043 | 0.010 | 1.30E-05 | 0 | 240,229 | 0.662 | 0.047 | 0.012 | 1.23E-04 | 0 | 142,092 |

**Supplementary Table 3.** Associatio

| Locus # | Discovery Analysis | dbSNP ID | EUR Analysis Stage 1 | | | | | | Pooled Analysis Stage 2 | | | | | | EUR Analysis Stage 2 | | | | | |
|---|---|---|---|---|---|---|---|---|---|---|---|---|---|---|---|---|---|---|---|---|
| | | | EAF | BETA | SE | P | Het $I^2$ | N | EAF | BETA | SE | P | Het $I^2$ | N | EAF | BETA | SE | P | Het $I^2$ | N |
| **Model 1 Results** | | | | | | | | | | | | | | | | | | | | |
| 1 | a.m1.s1 | rs12725199 | 0.257 | 0.065 | 0.012 | 1.25E-07 | 64.7 | 135,152 | 0.262 | 0.047 | 0.014 | 8.53E-04 | 25.2 | 330,475 | 0.281 | 0.046 | 0.015 | 1.67E-03 | 49.5 | 316,200 |
| 1 | e.m1.s1 | rs12740489 | 0.257 | 0.065 | 0.012 | 1.01E-07 | 67.2 | 135,202 | 0.262 | 0.048 | 0.014 | 7.65E-04 | 14.7 | 330,475 | 0.281 | 0.046 | 0.015 | 1.44E-03 | 41.7 | 316,200 |
| 2 | e.m1.s1 | rs12467202 | 0.915 | -0.088 | 0.019 | 3.19E-06 | 0 | 135,432 | 0.889 | -0.062 | 0.019 | 1.28E-03 | 0 | 330,475 | 0.905 | -0.085 | 0.022 | 1.21E-04 | 0 | 316,200 |
| 2 | a.m1.s1 | rs2376073 | 0.083 | 0.081 | 0.019 | 2.47E-05 | 0 | 133,138 | 0.116 | 0.061 | 0.019 | 1.29E-03 | 0 | 330,475 | 0.102 | 0.081 | 0.021 | 1.49E-04 | 0 | 316,200 |
| 3 | a.m1.s1 | rs9831061 | 0.758 | 0.057 | 0.013 | 6.94E-06 | 0 | 136,499 | 0.715 | 0.047 | 0.014 | 7.48E-04 | 0 | 330,475 | 0.744 | 0.040 | 0.015 | 8.23E-03 | 55.2 | 316,200 |
| 4 | e.m1.s1 | rs34592089 | 0.057 | 0.102 | 0.024 | *1.55E-05* | 0 | 132,887 | 0.040 | 0.147 | 0.037 | 6.02E-05 | 0 | 330,475 | 0.049 | 0.143 | 0.038 | 1.93E-04 | 0 | 316,200 |
| 5 | a.m2.s1 | rs2964006 | 0.466 | 0.040 | 0.011 | 1.67E-04 | 0 | 136,815 | 0.445 | 0.039 | 0.012 | 1.59E-03 | 0 | 330,475 | 0.466 | 0.041 | 0.013 | 2.12E-03 | 0 | 316,200 |
| 6 | a.m1.s2 | rs9380814 | 0.337 | -0.037 | 0.011 | 1.10E-03 | 0 | 137,233 | 0.333 | -0.064 | 0.014 | *2.12E-06* | 0 | 330,475 | 0.356 | -0.067 | 0.014 | *1.20E-06* | 0 | 316,200 |
| 6 | e.m1.s2 | rs1537230 | 0.337 | -0.038 | 0.011 | 8.65E-04 | 0 | 137,328 | 0.334 | -0.063 | 0.013 | *3.30E-06* | 0 | 330,475 | 0.357 | -0.067 | 0.014 | *1.17E-06* | 0 | 316,200 |
| 7 | a.m1.s1 | rs66474734 | 0.615 | -0.064 | 0.014 | *3.03E-06* | 0 | 75,845 | 0.536 | -0.062 | 0.021 | 2.47E-03 | 0 | 41,172 | 0.603 | -0.051 | 0.026 | 5.19E-02 | 0 | 26,897 |
| 8 | a.m1.s1 + a.m1.s2 | rs12553324 | 0.605 | -0.043 | 0.011 | 9.21E-05 | 68.9 | 137,819 | 0.649 | -0.090 | 0.022 | *2.78E-05* | 0 | 41,172 | 0.598 | -0.112 | 0.026 | *2.02E-05* | 0 | 26,897 |
| 9 | e.m2.s1 | rs60791051 | 0.941 | -0.115 | 0.028 | *3.36E-05* | 0 | 78,408 | 0.913 | -0.041 | 0.034 | 2.28E-01 | 0 | 41,172 | 0.929 | -0.132 | 0.048 | 5.92E-03 | 0 | 26,897 |
| 10 | a.m2.s1 | | 0.657 | -0.053 | 0.012 | *3.72E-06* | 0 | 137,819 | 0.646 | -0.040 | 0.013 | 2.02E-03 | 70.1 | 330,475 | 0.680 | -0.040 | 0.014 | 4.66E-03 | **77.3** | 316,200 |
| 10 | e.m1.s1 | rs2926743 | 0.269 | 0.059 | 0.012 | *7.52E-07* | 0 | 137,819 | 0.227 | 0.040 | 0.015 | 6.33E-03 | 60.2 | 330,475 | 0.240 | 0.046 | 0.015 | 2.69E-03 | **82.3** | 316,200 |
| 10 | a.m1.s1 | rs2958127 | 0.369 | 0.047 | 0.011 | *2.29E-05* | 0 | 137,819 | 0.378 | 0.045 | 0.013 | 4.33E-04 | **78** | 330,475 | 0.356 | 0.040 | 0.014 | 3.33E-03 | **75.5** | 316,200 |
| 10 | e.m2.s1 | rs12227117 | 0.268 | 0.055 | 0.012 | *5.46E-06* | 0 | 137,819 | 0.225 | 0.065 | 0.025 | 8.84E-03 | 0 | 41,172 | 0.270 | 0.096 | 0.029 | 8.65E-04 | 0 | 26,897 |
| 11 | a.m2.s2 | rs10735928 | 0.381 | 0.029 | 0.011 | 9.14E-03 | 32.4 | 137,819 | 0.383 | 0.053 | 0.013 | *2.41E-05* | 40.8 | 330,475 | 0.381 | 0.057 | 0.014 | *2.30E-05* | 0 | 316,200 |
| 11 | e.m2.s2 | rs10878270 | 0.618 | -0.029 | 0.011 | 8.79E-03 | 30.9 | 137,819 | 0.602 | -0.052 | 0.012 | *3.57E-05* | 59.6 | 330,475 | 0.619 | -0.059 | 0.014 | *1.28E-05* | 0 | 316,200 |
| 11 | e.m2.s1 | rs4762080 | 0.225 | 0.043 | 0.013 | 1.12E-03 | 0 | 137,819 | 0.201 | 0.056 | 0.015 | 2.93E-04 | 38.7 | 330,475 | 0.216 | 0.057 | 0.016 | 3.55E-04 | 39 | 316,200 |
| 12 | a.m1.s1 + a.m1.s2 | rs11642015 | 0.420 | 0.110 | 0.011 | **1.91E-24** | 0 | 137,819 | 0.391 | 0.085 | 0.013 | **4.28E-11** | 0 | 330,475 | 0.419 | 0.084 | 0.013 | **3.30E-10** | 0 | 316,200 |
| 12 | e.m1.s1 + e.m1.s2 | rs56094641 | 0.580 | -0.111 | 0.011 | **1.01E-24** | 0 | 137,819 | 0.609 | -0.083 | 0.013 | **9.04E-11** | 0 | 330,475 | 0.581 | -0.083 | 0.013 | **4.57E-10** | 0 | 316,200 |
| 13 | a.m1.s1 + e.m1.s1 | rs1800437 | 0.209 | -0.069 | 0.013 | *2.05E-07* | 0 | 137,819 | 0.209 | -0.039 | 0.015 | 9.96E-03 | 0 | 330,475 | 0.219 | -0.041 | 0.016 | 1.20E-02 | 0 | 316,200 |
| 14 | a.m1.s1 + e.m1.s1 | rs11088984 | 0.650 | 0.055 | 0.012 | *3.62E-06* | 0 | 137,819 | 0.668 | 0.049 | 0.013 | 2.24E-04 | 0 | 330,475 | 0.649 | 0.046 | 0.014 | 1.03E-03 | 0 | 316,200 |
| **Model 2 Results** | | | | | | | | | | | | | | | | | | | | |
| 1 | a.m1.s1 | rs12725199 | 0.257 | 0.050 | 0.013 | 1.17E-04 | 63.7 | 123,726 | 0.261 | 0.024 | 0.016 | 1.40E-01 | 57.9 | 127,528 | 0.281 | 0.028 | 0.017 | 9.18E-02 | 35 | 113,860 |
| 1 | e.m1.s1 | rs12740489 | 0.257 | 0.050 | 0.013 | 1.01E-04 | 65.5 | 123,776 | 0.261 | 0.024 | 0.016 | 1.31E-01 | 55 | 127,528 | 0.281 | 0.029 | 0.017 | 8.39E-02 | 26.8 | 113,860 |
| 2 | e.m1.s1 | rs12467202 | 0.915 | -0.082 | 0.020 | *4.45E-05* | 0 | 123,991 | 0.889 | -0.038 | 0.022 | 8.60E-02 | 0 | 127,528 | 0.905 | -0.053 | 0.025 | 3.74E-02 | 0 | 113,860 |
| 2 | a.m1.s1 | rs2376073 | 0.083 | 0.076 | 0.021 | 2.22E-04 | 0 | 121,717 | 0.116 | 0.033 | 0.022 | 1.22E-01 | 0 | 127,528 | 0.102 | 0.046 | 0.025 | 6.04E-02 | 0 | 113,860 |
| 3 | a.m1.s1 | rs9831061 | 0.758 | 0.036 | 0.014 | 8.50E-03 | 0 | 125,052 | 0.714 | 0.020 | 0.016 | 2.16E-01 | 0 | 127,528 | 0.744 | 0.017 | 0.017 | 3.42E-01 | 51.4 | 113,860 |
| 4 | e.m1.s1 | rs34592089 | 0.056 | 0.064 | 0.025 | 1.17E-02 | 0 | 121,445 | 0.053 | 0.108 | 0.051 | 3.37E-02 | 0 | 39,573 | 0.070 | 0.105 | 0.055 | 5.92E-02 | 0 | 25,905 |
| 5 | a.m2.s1 | rs2964006 | 0.466 | 0.045 | 0.011 | 8.60E-05 | 0 | 125,374 | 0.444 | 0.051 | 0.014 | 2.89E-04 | 52.7 | 127,528 | 0.466 | 0.056 | 0.015 | 2.43E-04 | 32.7 | 113,860 |
| 6 | a.m1.s2 | rs9380814 | 0.337 | -0.027 | 0.012 | 2.65E-02 | 0 | 125,788 | 0.332 | -0.049 | 0.015 | 1.54E-03 | 0 | 127,528 | 0.355 | -0.052 | 0.016 | 1.07E-03 | 0 | 113,860 |
| 6 | e.m1.s2 | rs1537230 | 0.337 | -0.027 | 0.012 | 2.28E-02 | 0 | 125,880 | 0.333 | -0.048 | 0.015 | 1.80E-03 | 0 | 127,528 | 0.356 | -0.052 | 0.016 | 9.87E-04 | 0 | 113,860 |
| 7 | a.m1.s1 | rs66474734 | 0.615 | -0.054 | 0.014 | 1.40E-04 | 0 | 75,565 | 0.535 | -0.066 | 0.023 | 4.60E-03 | 0 | 39,573 | 0.603 | -0.046 | 0.030 | 1.23E-01 | 0 | 25,905 |
| 8 | a.m1.s1 + a.m1.s2 | rs12553324 | 0.605 | -0.037 | 0.012 | 1.58E-03 | 32.4 | 126,369 | 0.649 | -0.101 | 0.024 | *3.65E-05* | 0 | 39,573 | 0.598 | -0.117 | 0.030 | 8.23E-05 | 0 | 25,905 |
| 9 | e.m2.s1 | rs60791051 | 0.941 | -0.135 | 0.029 | *3.47E-06* | 0 | 78,111 | 0.913 | -0.085 | 0.039 | 2.80E-02 | 0 | 39,573 | 0.929 | -0.155 | 0.054 | 3.99E-03 | 0 | 25,905 |

| | | | EUR Analysis Stage 1 | | | | | | Pooled Analysis Stage 2 | | | | | | EUR Analysis Stage 2 | | | | | |
|---|---|---|---|---|---|---|---|---|---|---|---|---|---|---|---|---|---|---|---|---|
| Locus # | Discovery Analysis | dbSNP ID | EAF | BETA | SE | P | Het $I^2$ | N | EAF | BETA | SE | P | Het $I^2$ | N | EAF | BETA | SE | P | Het $I^2$ | N |
| 10 | a.m2.s1 | | 0.658 | -0.065 | 0.012 | *1.09E-07* | 0 | 126,369 | 0.644 | -0.031 | 0.015 | 3.50E-02 | 63.4 | 127,528 | 0.680 | -0.035 | 0.016 | 2.97E-02 | **81.1** | 113,860 |
| 10 | e.m1.s1 | rs2926743 | 0.268 | 0.068 | 0.013 | *9.71E-08* | 0 | 126,369 | 0.227 | 0.029 | 0.017 | 8.03E-02 | 56.4 | 127,528 | 0.240 | 0.037 | 0.018 | 3.56E-02 | **82.1** | 113,860 |
| 10 | a.m1.s1 | rs2958127 | 0.368 | 0.057 | 0.012 | *1.32E-06* | 0 | 126,369 | 0.379 | 0.039 | 0.014 | 6.63E-03 | 65.5 | 127,528 | 0.356 | 0.039 | 0.016 | 1.37E-02 | 73.9 | 113,860 |
| 10 | e.m2.s1 | rs12227117 | 0.267 | 0.065 | 0.013 | *4.57E-07* | 0 | 126,369 | 0.225 | 0.057 | 0.028 | 4.13E-02 | 0 | 39,573 | 0.270 | 0.094 | 0.033 | 3.82E-03 | 0 | 25,905 |
| 11 | a.m2.s2 | rs10735928 | 0.381 | 0.046 | 0.012 | 8.95E-05 | 22.4 | 126,369 | 0.383 | 0.071 | 0.014 | *5.95E-07* | 0 | 127,528 | 0.382 | 0.071 | 0.016 | *5.12E-06* | 0 | 113,860 |
| 11 | e.m2.s2 | rs10878270 | 0.618 | -0.046 | 0.012 | 8.36E-05 | 21.2 | 126,369 | 0.601 | -0.067 | 0.014 | *2.71E-06* | 28.1 | 127,528 | 0.619 | -0.072 | 0.015 | *2.73E-06* | 0 | 113,860 |
| 11 | e.m2.s1 | rs4762080 | 0.225 | 0.064 | 0.014 | *6.28E-06* | 0 | 126,369 | 0.201 | 0.060 | 0.018 | 6.58E-04 | 69.2 | 127,528 | 0.216 | 0.064 | 0.018 | 4.69E-04 | 61.8 | 113,860 |
| 12 | a.m1.s1 + a.m1.s2 | rs11642015 | 0.421 | 0.032 | 0.011 | 4.63E-03 | 0 | 126,369 | 0.390 | -0.025 | 0.015 | 8.41E-02 | 0 | 127,528 | 0.419 | -0.029 | 0.015 | 5.66E-02 | 0 | 113,860 |
| 12 | e.m1.s1 + e.m1.s2 | rs56094641 | 0.579 | -0.033 | 0.011 | 4.35E-03 | 0 | 126,369 | 0.609 | 0.027 | 0.015 | 6.29E-02 | 0 | 127,528 | 0.581 | 0.030 | 0.015 | 4.79E-02 | 0 | 113,860 |
| 13 | a.m1.s1 + e.m1.s1 | rs1800437 | 0.209 | -0.042 | 0.014 | 3.26E-03 | 0 | 126,369 | 0.209 | -0.013 | 0.017 | 4.67E-01 | 0 | 127,528 | 0.218 | -0.015 | 0.018 | 4.07E-01 | 0 | 113,860 |
| 14 | a.m1.s1 + e.m1.s1 | rs11088984 | 0.650 | 0.047 | 0.013 | 1.68E-04 | 0 | 126,369 | 0.669 | 0.039 | 0.015 | 8.91E-03 | 0 | 127,528 | 0.649 | 0.036 | 0.016 | 2.29E-02 | 0 | 113,860 |

**Supplementary Table 4**. Results for approximate conditional analyses to identify secondary signals in identified association regions.

| | | Conditioned on Tag SNP (TS) | | | | | | Most Significant after conditioning | | | | | Before Conditioning | | | After Conditioning | | | |
|---|---|---|---|---|---|---|---|---|---|---|---|---|---|---|---|---|---|---|---|
| **Strata** | **Chr** | **Marker** | **Nearest Gene** | **POS** | **β** | **SE** | **P** | **Marker** | **POS** | **Distance** | **$r^2$** | **EAF** | **β** | **SE** | **P** | **β** | **SE** | **P** | **effN** |
| **OSA** | | | | | | | | | | | | | | | | | | | |
| Multi-population | 1 | rs12725199 | *PTPB2* | 97293792 | 0.055 | 0.009 | **1.54E-09** | 1:96949526:C_T | 96949526 | 344266 | 0.103 | 0.421 | -0.043 | 0.008 | 7.14E-08 | -0.028 | 0.008 | 5.12E-04 | 480,263 |
| Multi-population | 2 | rs2376073 | Intergenic | 104662008 | 0.070 | 0.013 | **3.03E-08** | 2:104967025:C_T | 104967025 | 305017 | 0.013 | 0.045 | -0.116 | 0.035 | 1.09E-03 | -0.123 | 0.035 | 4.87E-04 | 141,517 |
| Multi-population | 3 | rs9831061 | *ETV5* | 185835281 | 0.051 | 0.009 | **1.21E-08** | 3:186324108:A_G | 186324108 | 488827 | 0.000 | 0.006 | 0.237 | 0.065 | 2.65E-04 | 0.240 | 0.065 | 2.14E-04 | 319,166 |
| Multi-population | 6 | rs9380814 | *DNAH8* | 38934922 | -0.048 | 0.009 | **1.82E-08** | 6:39282119:C_G | 39282119 | 347197 | 0.002 | 0.012 | -0.375 | 0.112 | 8.08E-04 | -0.366 | 0.112 | 1.07E-03 | 52,889 |
| Multi-population | 8 | rs66474734 | *CSMD3* | 114661935 | -0.062 | 0.011 | **2.47E-08** | 8:114338684:C_G | 114338684 | 323251 | 0.000 | 0.014 | 0.129 | 0.040 | 1.40E-03 | 0.128 | 0.040 | 1.51E-03 | 333,634 |
| Multi-population | 9 | rs12553324 | *ELAVL2* | 23347865 | -0.052 | 0.009 | **3.50E-08** | 9:23472806:A_G | 23472806 | 124941 | 0.005 | 0.010 | -0.204 | 0.053 | 1.37E-04 | -0.187 | 0.053 | 4.77E-04 | 271,545 |
| Multi-population | 12 | rs2958127 | *NACA* | 57114307 | 0.050 | 0.008 | **5.75E-10** | 12:56783055:A_G | 56783055 | 331252 | 0.001 | 0.091 | -0.092 | 0.025 | 1.81E-04 | -0.093 | 0.025 | 1.47E-04 | 148,730 |
| Multi-population | 16 | rs11642015 | *FTO* | 53802494 | 0.098 | 0.008 | **8.47E-34** | 16:53760912:A_C | 53760912 | 41582 | 0.011 | 0.030 | 0.078 | 0.029 | 6.98E-03 | 0.106 | 0.029 | 2.45E-04 | 310,574 |
| Multi-population | 19 | rs1800437 | *GIPR* | 46181392 | -0.059 | 0.010 | **2.23E-09** | 19:46204938:C_T | 46204938 | 23546 | 0.003 | 0.018 | 0.132 | 0.031 | 1.66E-05 | 0.122 | 0.031 | 7.21E-05 | 456,043 |
| Multi-population | 21 | rs11088984 | *POFUT2* | 46670045 | 0.051 | 0.009 | **3.94E-09** | 21:46898615:C_T | 46898615 | 228570 | 0.002 | 0.030 | 0.146 | 0.041 | 3.39E-04 | 0.141 | 0.041 | 5.47E-04 | 154,559 |
| European-Only | 4 | rs34592089 | *BANK1* | 102926923 | 0.113 | 0.020 | **1.81E-08** | 4:103323995:A_G | 103323995 | 397072 | 0.034 | 0.380 | -0.024 | 0.009 | 5.85E-03 | -0.032 | 0.009 | 2.46E-04 | 455,821 |
| **AHI** | | | | | | | | | | | | | | | | | | | |
| Multi-population | 2 | rs1200596362 | *SLC39A10* | 196364827 | 0.093 | 0.017 | **2.84E-08** | 2:196679162:C_T | 196679162 | 314335 | 0.033 | 0.285 | 0.040 | 0.010 | 8.47E-05 | 0.034 | 0.010 | 7.60E-04 | 31,978 |
| **avgSa$O_2$** | | | | | | | | | | | | | | | | | | | |
| Multi-population | 6 | rs802729 | *PTPRK* | 128280360 | -0.427 | 0.076 | **1.84E-08** | 6:128510821:C_T | 128510821 | 230461 | 0.001 | 0.100 | 0.214 | 0.057 | 1.90E-04 | 0.219 | 0.058 | 1.43E-04 | 1,961 |
| European-Only | 8 | rs2194955 | *GFRA2* | 21292558 | -0.143 | 0.025 | **7.17E-09** | 8:21180360:A_C | 21180360 | 112198 | 0.003 | 0.001 | -1.004 | 0.277 | 2.89E-04 | -1.002 | 0.279 | 3.24E-04 | 1,047 |
| **OSAadjBMI** | | | | | | | | | | | | | | | | | | | |
| Multi-population | 5 | rs2964006 | *GRIA1* | 153213880 | 0.049 | 0.009 | **2.24E-08** | 5:152829369:A_G | 152829369 | 384511 | 0.000 | 0.010 | -0.306 | 0.089 | 5.38E-04 | -0.302 | 0.089 | 6.35E-04 | 67,054 |
| Multi-population | 12 | rs4759254 | *NACA* | 57096674 | -0.055 | 0.009 | **2.99E-09** | 12:56978780:C_T | 56978780 | 117894 | 0.010 | 0.009 | 0.302 | 0.080 | 1.66E-04 | 0.298 | 0.080 | 2.11E-04 | 98,665 |
| Multi-population | 12 | rs10735928 | *MSRB3* | 65769626 | 0.054 | 0.009 | **7.73E-10** | 12:66134754:G_T | 66134754 | 365128 | 0.002 | 0.028 | 0.121 | 0.029 | 3.31E-05 | 0.114 | 0.029 | 1.01E-04 | 263,919 |
| European-Only | 9 | rs60791051 | *FAM189A2* | 71941742 | -0.139 | 0.026 | **4.80E-08** | 9:71859084:C_T | 71859084 | 82658 | 0.001 | 0.037 | 0.088 | 0.025 | 3.97E-04 | 0.092 | 0.025 | 2.31E-04 | 228,821 |
| **AHIadjBMI** | | | | | | | | | | | | | | | | | | | |
| Multi-population | 6 | rs78545995 | *EYS* | 65804913 | 0.299 | 0.054 | **3.64E-08** | 6:65606754:C_G | 65606754 | 198159 | 0.000 | 0.031 | -0.262 | 0.073 | 3.27E-04 | -0.262 | 0.073 | 3.27E-04 | 3,967 |
| **avgSa$O_2$adjBMI** | | | | | | | | | | | | | | | | | | | |
| Multi-population | 1 | rs79914264 | *PTPRU* | 29685027 | 0.206 | 0.038 | **4.04E-08** | 1:29574263:A_G | 29574263 | 110764 | 0.000 | 0.011 | -0.470 | 0.128 | 2.47E-04 | -0.470 | 0.129 | 2.56E-04 | 3,443 |

**Supplementary Table 5**. Summary of association results for after approximate conditional analyses on known index variants within 500 kb +/- of lead variants for each association region.

| Conditioned on Known Signals | | | | | | | | | | | | | | | | | |
|---|---|---|---|---|---|---|---|---|---|---|---|---|---|---|---|---|---|
| | Tag SNP (TS) | | | | | | Known Marker | | | | | | After Conditioning | | | Change | |
| **Chr** | **Marker** | **Nearest Gene** | **POS** | **β** | **SE** | **P** | **Conditioned On** | **Nearest Gene** | **PMID** | **POS** | **Distance** | **$r^2$** | **β** | **SE** | **P** | **β diff** | **β %** |
| **OSA - Multi-Population Meta-Analysis** | | | | | | | | | | | | | | | | | |
| 1 | rs12725199 | *PTPB2* | 97293792 | 0.055 | 0.009 | **1.54E-09** | rs6679458 | *LINC01787* | 36989840 | 96946253 | 347539 | 0.11 | 0.041 | 0.009 | 8.08E-06 | 0.014 | 73.98% |
| | | | | | | | rs17367240 | *LINC01787* | 36989840 | 96960057 | 333735 | 0.06 | 0.046 | 0.009 | 4.87E-07 | 0.046 | 83.40% |
| | | | | | | | rs34013490* | *PTBP2* | 38330144 | 97032649 | 261143 | - | - | - | - | - | - |
| 3 | rs9831061 | *ETV5* | 185835281 | 0.051 | 0.009 | **1.21E-08** | rs869400 | *ETV5* | 36989840 | 185826740 | 8541 | 0.68 | 0.018 | 0.009 | 4.99E-02 | 0.034 | 34.33% |
| **12** | **rs2958127** | ***NACA*** | **57114307** | **0.050** | **0.008** | **5.75E-10** | rs2277339 | *PRIM1* | 36989840 | 57146069 | 31762 | **0.06** | **0.045** | **0.008** | **2.34E-08** | **0.005** | **90.47%** |
| | | | | | | | rs59333125 | *BAZ2A* | 38330144 | 57025908 | 57025908 | **0.01** | **0.048** | **0.008** | **3.56E-09** | **0.048** | **95.64%** |
| 16 | rs11642015 | *FTO* | 53802494 | 0.098 | 0.008 | **8.47E-34** | rs9937053 | *FTO* | 33243845 | 53799507 | 2987 | 0.90 | 0.012 | 0.008 | 1.49E-01 | 0.086 | 11.97% |
| | | | | | | | rs62048402 ¥ | *FTO* | 36989840 | 53803223 | 729 | 1.00 | - | - | - | - | - |
| | | | | | | | rs1558902 ¥ | *FTO* | 36989840 | 53803574 | 1080 | 1.00 | - | - | - | - | - |
| | | | | | | | rs11075985 ¥ | *FTO* | 38330144 | 53805207 | 53805207 | 0.90 | - | - | - | - | - |
| 19 | rs1800437 | *GIPR* | 46181392 | -0.059 | 0.010 | **2.23E-09** | rs10423928 ¥ | *GIPR* | 38330144 | 46182304 | 912 | 1.00 | - | - | - | - | - |
| **OSA - White/European Meta-Analysis** | | | | | | | | | | | | | | | | | |
| 4 | rs34592089 | *BANK1* | 102926923 | 0.113 | 0.020 | **1.81E-08** | rs13107325 | *SLC39A8* | 36989840 | 103188709 | 261786 | 0.61 | 0.051 | 0.020 | 1.12E-02 | 0.062 | 45.05% |
| **OSAadjBMI - Multi-Population Meta-Analysis** | | | | | | | | | | | | | | | | | |
| **12** | **rs4759254** | ***NACA*** | **57096674** | **-0.055** | **0.009** | **2.99E-09** | rs2277339 | *PRIM1* | 36989840 | **57146069** | **49395** | **0.05** | **-0.051** | **0.009** | **3.85E-08** | **0.004** | **93.13%** |
| | | | | | | | rs59333125 | *BAZ2A* | 38330144 | **57025908** | **57025908** | **0.03** | **-0.051** | **0.009** | **4.81E-08** | **0.051** | **92.30%** |
| 12 | rs10735928 | *MSRB3* | 65769626 | 0.054 | 0.009 | **7.73E-10** | rs1826663 ¥ | *MSRB3* | 36989840 | 65797709 | 28083 | 1.00 | - | - | - | - | - |

*Not available in discvoery GWAS summary statisics from current paper.

¥ Did not converge.

**Supplementary Table 6.** Fine-mapping results. Association signals fromt eh stage 1 plus stage 2 meta-analyses were used for fine-mapping. For this analysis, the finemap configuration of 1 causal variant per loci was assumed, as no loci displayed evidence of secondary signals.

| | Locus Tag SNP | | | | | Top FINEMAP Variant | | | | | | 95% Credible Set | | | | | | |
|---|---|---|---|---|---|---|---|---|---|---|---|---|---|---|---|---|---|---|
| **Strata** | **Index variant** | **Chr.** | **POS (hg19)** | **Nearest Gene** | **Index variant available in Reference (Y/N)** | **log10bf (model)** | **log10bf (top variant)** | **Top variant rsID** | **Pos (hg19)** | **PP** | **Is index variant (Y/N)** | **POS-Start** | **POS-End** | **Distance from index variant** | **LD between the finemapped and top variant** | **Range** | **#Variants in Credible Set** | **Top 5 FINEMAP variants within 95% CS** |
| **OSA** | | | | | | | | | | | | | | | | | | |
| Multi-population | rs12725199 | 1 | 97293792 | *PTPB2* | Y | 7.430 | 2.647 | rs12725199 | 97293792 | 0.0800 | Y | 96891671 | 97296935 | 0 | 1 | 405264 | 135 | 1:97293792:A_C (0.080), 1:97296935:A_G (0.080), 1:97287825:G_T (0.0656), 1:97111780:C_T (0.0301), 1:97110637:C_T (0.030) |
| Multi-population | rs2376073 | 2 | 104662008 | Intergenic | Y | 5.555 | 3.698 | rs2376073 | 104662008 | 0.5048 | Y | 104601448 | 104662008 | 0 | 1 | 60560 | 4 | 2:104662008:C_T (0.505), 2:104601968:C_T (0.414), 2:104601448:T_TCACA (0.029), 2:104622478:C_T (0.006) |
| Multi-population | rs9831061 | 3 | 185835281 | *ETV5* | Y | 5.919 | 3.552 | rs9831061 | 185835281 | 0.4263 | Y | 185826028 | 185835281 | 0 | 1 | 9253 | 11 | 3:185835281:A_G (0.426), 3:185833759:A_C (0.167), 3:185829891:A_T (0.109), 3:185826764:C_T (0.067), 3:185826028:A_G (0.060) |
| Multi-population | rs9380814 | 6 | 38934922 | *DNAH8* | Y | 6.158 | 3.103 | rs9380814 | 38934922 | 0.1682 | Y | 38928910 | 39003874 | 0 | 1 | 74964 | 14 | 6:38934922:A_G (0.168), 6:38936096:C_T (0.157), 6:38930402:C_T (0.121), 6:38928910:A_G (0.106), 6:38944196:A_G (0.087) |
| Multi-population | rs66474734 | 8 | 114661935 | *CSMD3* | Y | 6.049 | 3.362 | rs66474734 | 114661935 | 0.2842 | Y | 114613494 | 114661935 | 0 | 1 | 48441 | 7 | 8:114661935:T_TATA (0.284), 8:114650056:G_GA (0.227), 8:114633079:A_AT (0.143), 8:114647559:C_T (0.102), 8:114652051:G_GT (0.100) |
| Multi-population | rs12553324 | 9 | 23347865 | *ELAVL2* | Y | 5.952 | 3.255 | rs12553324 | 23347865 | 0.2121 | Y | 23345347 | 23362311 | 0 | 1 | 16964 | 23 | 9:23347865:C_G (0.212), 9:23345347:A_G (0.154), 9:23360417:C_G (0.116), 9:23355664:A_C (0.109), 9:23346842:C_G (0.082) |
| Multi-population | rs2958127 | 12 | 57114307 | *NACA* | Y | 8.019 | 2.371 | 12:57103162:C_T | 57103162 | 0.0569 | N | 56984606 | 57171516 | 11145 | 1.000 | 186910 | 56 | 12:57103162:C_T (0.057), 12:57114307:A_T (0.049), 12:57109412:C_G (0.045), 12:57112637:A_G (0.045), 12:57125879:A_G (0.045) |
| Multi-population | rs11642015 | 16 | 53802494 | *FTO* | Y | 30.756 | 3.296 | rs11642015 | 53802494 | 0.2597 | Y | 53800954 | 53809123 | 0 | 1 | 8169 | 5 | 16:53802494:C_T (0.260), 16:53806453:A_G (0.260), 16:53800954:C_T (0.166), 16:53803223:A_G (0.166), 16:53809123:C_T (0.143) |
| Multi-population | rs1800437 | 19 | 46181392 | *GIPR* | Y | 6.789 | 3.357 | rs1800437 | 46181392 | 0.2864 | Y | 46179043 | 46182304 | 0 | 1 | 3261 | 5 | 19:46181392:C_G (0.286), 19:46180414:G_T (0.212), 19:46180184:C_T (0.188), 19:46179043:A_G( 0.178), 19:46182304:A_T (0.131) |
| Multi-population | rs11088984 | 21 | 46670045 | *POFUT2* | Y | 6.632 | 3.352 | rs11088984 | 46670045 | 0.2414 | Y | 46496949 | 46686499 | 0 | 1 | 189550 | 40 | 21:46670045:A_C (0.241), 21:46539725:A_G (0.131), 21:46542374:C_T (0.059), 21:46496949:C_T (0.038), 21:46627687:A_G (0.037) |
| European-Only | rs34592089 | 4 | 102926923 | *BANK1* | Y | 5.790 | 5.699 | rs34592089 | 102926923 | 0.9919 | Y | 102926923 | 102926923 | 0 | 1 | 0 | 1 | 4:102926923:A_G (0.992) |
| **AHI** | | | | | | | | | | | | | | | | | | |
| Multi-population | rs1200596362 | 2 | 196364827 | *SLC39A10* | Y | 5.793 | 4.600 | rs1200596362 | 196364827 | 0.8673 | Y | 196317278 | 196433941 | 0 | 1 | 116663 | 9 | 2:196364827:T_TA (0.867), 2:196433941:T_TA (0.053), 2:196333418:A_AAT (0.010), 2:196369621:C_T (0.008), 2:196329529:T_TG (0.006) |
| **avgSaO$_2$** | | | | | | | | | | | | | | | | | | |
| Multi-population | rs802729 | 6 | 128280360 | *PTPRK* | Y | 5.811 | 4.731 | rs802729 | 128280360 | 0.9065 | Y | 128280360 | 128352791 | 0 | 1 | 72431 | 2 | 6:128280360:A_T (0.906), 6:128352791:A_C (0.071) |

| | Locus Tag SNP | | | | | Top FINEMAP Variant | | | | | | 95% Credible Set | | | | | | |
|---|---|---|---|---|---|---|---|---|---|---|---|---|---|---|---|---|---|---|
| **Strata** | **Index variant** | **Chr.** | **POS (hg19)** | **Nearest Gene** | **Index variant available in Reference (Y/N)** | **log10bf (model)** | **log10bf (top variant)** | **Top variant rsID** | **Pos (hg19)** | **PP** | **Is index variant (Y/N)** | **POS-Start** | **POS-End** | **Distance from index variant** | **LD between the finemapped and top variant** | **Range** | **#Variants in Credible Set** | **Top 5 FINEMAP variants within 95% CS** |
| European-Only | rs2194955 | 8 | 21292558 | *GFRA2* | Y | 6.701 | 3.385 | rs2194955 | 21292558 | 0.3441 | Y | 21287105 | 21315809 | 0 | 1 | 28704 | 13 | 8:21292558:A_C (0.344), 8:21298827:A_G (0.270), 8:21287105:A_G (0.112), 8:21315374:A_G (0.066), 8:21312389:A_G (0.053) |
| **OSAadjBMI** | | | | | | | | | | | | | | | | | | |
| Multi-population | rs2964006 | 5 | 153213880 | *GRIA1* | Y | 6.672 | 2.389 | rs2964006 | 153213880 | 0.0435 | Y | 153100273 | 153215342 | 0 | 1 | 115069 | 55 | 5:153213880:G_T (0.0435), 5:153210741:A_C (0.041), 5:153212042:G_T (0.038), 5:153167595:A_G (0.035), 5:153138709:C_T (0.033) |
| Multi-population | rs4759254 | 12 | 57096674 | *NACA* | Y | 7.364 | 2.426 | 12:57103162:C_T | 57103162 | 0.0646 | N | 56984606 | 57169818 | 6488 | 0.872 | 185212 | 58 | 12:57103162:C_T (0.065), 12:57096674:C_T (0.058), 12:57114307:A_T (0.057), 12:57109412:C_G (0.053), 12:57112637:A_G (0.050) |
| Multi-population | rs10735928 | 12 | 65769626 | *MSRB3* | Y | 7.858 | 2.618 | 12:65797709:C_T | 65797709 | 0.0878 | N | 65762058 | 65835548 | 28083 | 0.998 | 73490 | 42 | 12:65797709:C_T (0.0878), 12:65769626:A_C (0.082), 12:65766498:C_T (0.041), 12:65793224:A_G (0.038), 12:65779193:G_T (0.036) |
| European-Only | rs60791051 | 9 | 71941742 | *FAM189A2* | Y | 5.700 | 3.592 | rs60791051 | 71941742 | 0.4830 | Y | 71930561 | 71941742 | 0 | 1 | 11181 | 3 | 9:71941742:A_C (0.483), 9:71939278:A_C (0.416), 9:71930561:T_TA (0.084) |
| **AHIadjBMI** | | | | | | | | | | | | | | | | | | |
| Multi-population | rs78545995 | 6 | 65804913 | *EYS* | Y | 6.004 | 3.909 | rs78545995 | 65804913 | 0.5277 | Y | 65804913 | 65934775 | 0 | 1 | 129862 | 4 | 6:65804913:A_G (0.528), 6:65821376:A_G (0.354), 6:65838774:A_T (0.066), 6:65934775:G_T (0.040) |
| **avgSaO$_2$adjBMI** | | | | | | | | | | | | | | | | | | |
| Multi-population | rs79914264 | 1 | 29685027 | *PTPRU* | Y | 5.905 | 5.493 | rs79914264 | 29685027 | 0.9890 | Y | 29685027 | 29685027 | 0 | 1 | 0 | 1 | 1:29685027:A_G (0.989) |

**Supplementary Table 7.** Association results for AHI-associated SNPs across all analyses and stages.

| | | | | | | | Pooled Analysis Stage 1 + Stage 2 | | | | | | EUR Analysis Stage 1 + Stage 2 | | | | | | Pooled Analysis Stage 1 | | | | | |
|---|---|---|---|---|---|---|---|---|---|---|---|---|---|---|---|---|---|---|---|---|---|---|---|---|
| **Discovery Analysis** | **dbSNP ID** | **CHR** | **POS** | **Nearest Gene** | **A1** | **A2** | **EAF** | **BETA** | **SE** | **P** | **Het $I^2$** | **N** | **EAF** | **BETA** | **SE** | **P** | **Het $I^2$** | **N** | **EAF** | **BETA** | **SE** | **P** | **Het $I^2$** | **N** |
| **Model 1 Results** | | | | | | | | | | | | | | | | | | | | | | | | |
| Pooled Model 1<br>EUR Model 1 | rs1200596362 | 2 | 196364827 | *SLC39A10* | T | TA | 0.349 | 0.093 | 0.017 | **2.84E-08** | 0 | 8,152 | 0.346 | 0.102 | 0.017 | **3.93E-09** | 0 | 7,803 | 0.349 | 0.093 | 0.018 | *2.87E-07* | 0 | 7,034 |
| Pooled Model 2 | rs78545995 | 6 | 65804913 | *EYS* | A | G | 0.987 | 0.234 | 0.050 | 3.44E-06 | 0 | 18,748 | 0.986 | 0.159 | 0.044 | 3.03E-04 | 69.6 | 23,768 | 0.984 | 0.286 | 0.154 | 6.29E-02 | 0 | 2,086 |
| **Model 2 Results** | | | | | | | | | | | | | | | | | | | | | | | | |
| Pooled Model 1<br>EUR Model 1 | rs1200596362 | 2 | 196364827 | *SLC39A10* | T | TA | 0.346 | 0.087 | 0.018 | 2.38E-06 | 0 | 6,945 | 0.344 | 0.097 | 0.019 | 2.27E-07 | 0 | 6,654 | 0.346 | 0.088 | 0.020 | *1.16E-05* | 0 | 5,827 |
| Pooled Model 2 | rs78545995 | 6 | 65804913 | *EYS* | A | G | 0.987 | 0.299 | 0.054 | **3.64E-08** | 0 | 15,981 | 0.986 | 0.217 | 0.048 | 6.09E-06 | 71.5 | 19,888 | 0.984 | 0.369 | 0.153 | 1.56E-02 | 0 | 1,914 |

**Supplementary Table 7.** Association

| Discovery Analysis | dbSNP ID | EUR Analysis Stage 1 | | | | | | Pooled Analysis Stage 2 | | | | | | EUR Analysis Stage 2 | | | | | |
|---|---|---|---|---|---|---|---|---|---|---|---|---|---|---|---|---|---|---|---|
| | | EAF | BETA | SE | P | Het $I^2$ | N | EAF | BETA | SE | P | Het $I^2$ | N | EAF | BETA | SE | P | Het $I^2$ | N |
| **Model 1 Results** | | | | | | | | | | | | | | | | | | | |
| Pooled Model 1<br>EUR Model 1 | rs1200596362 | 0.346 | 0.102 | 0.019 | **3.36E-08** | 0 | 6,685 | 0.349 | 0.098 | 0.047 | 3.70E-02 | 0 | 1,118 | 0.349 | 0.098 | 0.047 | 3.70E-02 | 0 | 1,118 |
| Pooled Model 2 | rs78545995 | 0.983 | 0.046 | 0.077 | 5.51E-01 | 71.2 | 8,594 | 0.988 | 0.228 | 0.053 | *1.93E-05* | 0 | 16,662 | 0.987 | 0.216 | 0.054 | 6.42E-05 | 0 | 15,174 |
| **Model 2 Results** | | | | | | | | | | | | | | | | | | | |
| Pooled Model 1<br>EUR Model 1 | rs1200596362 | 0.343 | 0.101 | 0.021 | *9.95E-07* | 0 | 5,536 | 0.349 | 0.080 | 0.046 | 8.25E-02 | 0 | 1,118 | 0.349 | 0.080 | 0.046 | 8.25E-02 | 0 | 1,118 |
| Pooled Model 2 | rs78545995 | NA | NA | NA | NA | NA | NA | 0.988 | 0.288 | 0.058 | *6.54E-07* | 0 | 14,067 | 0.987 | 0.281 | 0.059 | *1.79E-06* | 0 | 12,606 |

**Supplementary Table 8.** Association results for avg$SaO_2$-associated SNPs across all analyses and stages.

| | | | | | | | Pooled Analysis Stage 1 + Stage 2 | | | | | | EUR Analysis Stage 1 + Stage 2 | | | | | | Pooled Analysis Stage 1 | | | | |
|---|---|---|---|---|---|---|---|---|---|---|---|---|---|---|---|---|---|---|---|---|---|---|---|
| Discovery Analysis | dbSNP ID | CHR | POS | Nearest Gene | A1 | A2 | EAF | BETA | SE | P | Het $I^2$ | N | EAF | BETA | SE | P | Het $I^2$ | N | EAF | BETA | SE | P | Het $I^2$ |
| **Model 1 Results** | | | | | | | | | | | | | | | | | | | | | | | |
| a.m1.s1 | rs802729 | 6 | 128280360 | *PTPRK* | A | T | 0.985 | -0.427 | 0.076 | **1.84E-08** | 0 | 2,634 | 0.987 | -0.135 | 0.093 | 1.48E-01 | 0 | 5,025 | 0.985 | -0.436 | 0.081 | *6.90E-08* | 0 |
| e.m1.s1 | rs2194955 | 8 | 21292558 | *GFRA2* | A | C | 0.785 | -0.116 | 0.022 | *8.35E-08* | 49.1 | 6,908 | 0.819 | -0.143 | 0.025 | **7.17E-09** | 0 | 5,872 | 0.814 | -0.138 | 0.027 | *2.85E-07* | 0 |
| a.m2.s1 | rs79914264 | 1 | 29685027 | *PTPRU* | A | G | 0.020 | 0.118 | 0.077 | 1.25E-01 | 0 | 4,627 | 0.015 | 0.134 | 0.105 | 2.00E-01 | 0 | 3,633 | 0.015 | 0.118 | 0.112 | 2.91E-01 | 0 |
| **Model 2 Results** | | | | | | | | | | | | | | | | | | | | | | | |
| a.m1.s1 | rs802729 | 6 | 128280360 | *PTPRK* | A | T | 0.984 | -0.419 | 0.112 | 1.73E-04 | 0 | 2,473 | 0.987 | -0.124 | 0.095 | 1.89E-01 | 0 | 4,858 | 0.984 | -0.449 | 0.127 | 4.28E-04 | 0 |
| e.m1.s1 | rs2194955 | 8 | 21292558 | *GFRA2* | A | C | 0.785 | -0.107 | 0.022 | 1.25E-06 | 0 | 6,713 | 0.820 | -0.129 | 0.025 | 2.36E-07 | 0 | 5,705 | 0.814 | -0.116 | 0.027 | *2.12E-05* | 0 |
| a.m2.s1 | rs79914264 | 1 | 29685027 | *PTPRU* | A | G | 0.016 | 0.206 | 0.038 | **4.04E-08** | 53.4 | 4,457 | 0.014 | 0.105 | 0.107 | 3.29E-01 | 0 | 3,491 | 0.015 | 0.227 | 0.040 | **1.55E-08** | 65 |

**Supplementary Table 8.** Associatic

| | | | EUR Analysis Stage 1 | | | | | | Pooled Analysis Stage 2 | | | | | | EUR Analysis Stage 2 | | | | | |
|---|---|---|---|---|---|---|---|---|---|---|---|---|---|---|---|---|---|---|---|---|
| **Discovery Analysis** | **dbSNP ID** | **N** | **EAF** | **BETA** | **SE** | **P** | **Het $I^2$** | **N** | **EAF** | **BETA** | **SE** | **P** | **Het $I^2$** | **N** | **EAF** | **BETA** | **SE** | **P** | **Het $I^2$** | **N** |
| **Model 1 Results** | | | | | | | | | | | | | | | | | | | | |
| a.m1.s1 | rs802729 | 1,331 | NA | NA | NA | NA | NA | NA | 0.984 | -0.363 | 0.223 | 1.03E-01 | 0 | 1,303 | 0.977 | -0.355 | 0.279 | 2.04E-01 | 0 | 483 |
| e.m1.s1 | rs2194955 | 4,758 | 0.819 | -0.144 | 0.028 | *2.66E-07* | 0 | 4,542 | 0.731 | -0.075 | 0.036 | 3.81E-02 | 41.7 | 2,150 | 0.821 | -0.138 | 0.052 | 7.77E-03 | 0 | 1,330 |
| a.m2.s1 | rs79914264 | 2,477 | 0.013 | 0.192 | 0.142 | 1.77E-01 | 0 | 2,303 | 0.024 | 0.117 | 0.105 | 2.67E-01 | 0 | 2,150 | 0.017 | 0.066 | 0.155 | 6.72E-01 | 0 | 1,330 |
| **Model 2 Results** | | | | | | | | | | | | | | | | | | | | |
| a.m1.s1 | rs802729 | 1,215 | 0.988 | -0.098 | 0.100 | 3.29E-01 | 25.4 | 4,394 | 0.984 | -0.320 | 0.230 | 1.64E-01 | 0 | 1,258 | 0.977 | -0.348 | 0.290 | 2.31E-01 | 0 | 464 |
| e.m1.s1 | rs2194955 | 4,608 | 0.819 | -0.118 | 0.029 | *3.28E-05* | 0 | 4,394 | NA | NA | NA | NA | NA | NA | 0.820 | -0.166 | 0.052 | 1.44E-03 | 0 | 1,311 |
| a.m2.s1 | rs79914264 | 2,352 | 0.012 | 0.132 | 0.148 | 3.75E-01 | 0 | 2,180 | 0.024 | 0.060 | 0.107 | 5.74E-01 | 0 | 2,105 | 0.017 | 0.075 | 0.155 | 6.29E-01 | 0 | 1,311 |

**Supplementary Table 9.** Lookups of top associated SNPs from discovery in existing databases of related traits.

| | | | | | | | | Discovery + Published Meta-analysis | | | | | MVP - OSA | | | | | MVP - OSAadjBMI | | | | | FinnGen - Slee | | |
|---|---|---|---|---|---|---|---|---|---|---|---|---|---|---|---|---|---|---|---|---|---|---|---|---|---|
| Discovery Trait | Discovery Strata | dbSNP ID | CHR | POS (hg19) | Nearest Gene | A1 | A2 | EAF | BETA | SE | P | N | EAF | BETA | SE | P | N | EAF | BETA | SE | P | N | EAF | BETA | SE |
| **AvgSaO$_2$ adjBMI*** | **multi-pop** | rs79914264 | 1 | 29685027 | *PTPRU* | A | G | NA | 0.1134 | 0.0247 | 4.34E-06 | 25,584 | 0.0207 | 0.0100 | 0.0259 | 7.04E-01 | 568,576 | 0.0207 | -0.0046 | 0.0267 | 8.66E-01 | 559,070 | 0.0027 | -0.0282 | 0.0689 |
| **OSA** | **multi-pop** | rs12725199 | 1 | 97293792 | *PTPB2* | A | C | 0.2160 | 0.0266 | 0.0046 | **5.54E-09** | 1,471,068 | 0.2168 | 0.0199 | 0.0065 | **1.77E-03** | 568,576 | 0.2165 | 0.0031 | 0.0066 | 6.45E-01 | 559,070 | 0.1707 | 0.0115 | 0.0091 |
| **OSA** | **multi-pop** | rs2376073 | 2 | 104662008 | Intergenic | T | C | 0.1176 | 0.0228 | 0.0057 | 5.53E-05 | 1,471,068 | 0.1279 | 0.0030 | 0.0078 | 7.03E-01 | 568,576 | 0.1281 | -0.0020 | 0.0081 | 8.05E-01 | 559,070 | 0.1105 | 0.0262 | 0.0109 |
| **AHI** | **multi-pop** | rs1200596362 | 2 | 196364827 | *SLC39A10* | T | TA | 0.3587 | 0.0596 | 0.0118 | 4.85E-07 | 15,105 | 0.3520 | 0.0091 | 0.0057 | 1.09E-01 | 568,576 | 0.3522 | 0.0068 | 0.0059 | 2.47E-01 | 559,070 | | | |
| **OSA** | **multi-pop** | rs9831061 | 3 | 185835281 | *ETV5* | A | G | 0.7002 | 0.0384 | 0.0041 | **7.54E-21** | 1,471,068 | 0.6736 | 0.0388 | 0.0057 | **6.78E-11** | 568,576 | 0.6730 | 0.0160 | 0.0063 | 9.95E-03 | 559,070 | 0.7055 | 0.0284 | 0.0075 |
| **OSA** | **EUR-only** | rs34592089 | 4 | 102926923 | *BANK1* | A | G | 0.0395 | 0.0812 | 0.0100 | **4.44E-16** | 1,428,048 | 0.0505 | 0.0710 | 0.0121 | **4.26E-10** | 568,576 | 0.0505 | 0.0327 | 0.0121 | 6.01E-03 | 558,257 | 0.0079 | 0.0673 | 0.0385 |
| **OSAadjBMI** | **multi-pop** | rs2964006 | 5 | 153213880 | *GRIA1* | T | G | 0.4349 | 0.0173 | 0.0047 | 2.27E-04 | 827,607 | 0.4280 | 0.0070 | 0.0053 | 1.91E-01 | 568,576 | 0.4277 | 0.0045 | 0.0056 | 4.17E-01 | 559,070 | 0.4375 | 0.0202 | 0.0069 |
| **OSA** | **multi-pop** | rs9380814 | 6 | 38934922 | *DNAH8* | A | G | 0.3048 | -0.0247 | 0.0040 | **1.01E-09** | 1,471,068 | 0.2740 | -0.0167 | 0.0059 | 6.02E-03 | 568,576 | 0.2734 | 0.0015 | 0.0063 | 8.07E-01 | 559,070 | 0.3252 | -0.0197 | 0.0073 |
| **AHI adjBMI*** | **multi-pop** | rs78545995 | 6 | 65804913 | *EYS* | A | G | 0.9857 | 0.1415 | 0.0342 | 3.46E-05 | 29,428 | 0.9876 | -0.0253 | 0.0278 | 3.63E-01 | 568,576 | 0.9875 | 0.0327 | 0.0292 | 2.61E-01 | 552,052 | 0.9946 | 0.0341 | 0.0467 |
| **AvgSaO$_2$** | **multi-pop** | rs802729 | 6 | 128280360 | *PTPRK* | A | T | NA | -0.1906 | 0.0501 | 1.44E-04 | 20,621 | 0.9648 | 0.0287 | 0.0278 | 3.31E-01 | 410,268 | | | | | | | | |
| **AvgSaO$_2$** | **EUR-only** | rs2194955 | 8 | 21292558 | *GFRA2* | A | C | 0.7315 | -0.0181 | 0.0096 | 5.84E-02 | 26,961 | 0.7708 | 0.0079 | 0.0064 | 2.23E-01 | 568,576 | 0.7706 | -0.0009 | 0.0067 | 8.94E-01 | 559,070 | 0.7898 | -0.0041 | 0.0084 |
| **OSA** | **multi-pop** | rs66474734 | 8 | 114661935 | *CSMD3* | T | TATA | 0.5616 | -0.0129 | 0.0048 | 7.17E-03 | 691,224 | 0.5573 | -0.0018 | 0.0053 | 7.38E-01 | 568,576 | 0.5567 | 0.0003 | 0.0056 | 9.60E-01 | 559,070 | | | |
| **OSA** | **multi-pop** | rs12553324 | 9 | 23347865 | *ELAVL2* | C | G | 0.6124 | -0.0295 | 0.0039 | **3.35E-14** | 1,181,765 | 0.6467 | -0.0219 | 0.0055 | **4.57E-05** | 568,576 | 0.6470 | -0.0140 | 0.0057 | 1.27E-02 | 559,070 | 0.5592 | -0.0295 | 0.0069 |
| **OSAadjBMI** | **EUR-only** | rs60791051 | 9 | 71941742 | *FAM189A2* | A | C | 0.9301 | -0.0284 | 0.0101 | 4.89E-03 | 663,086 | 0.9281 | 0.0007 | 0.0103 | 9.43E-01 | 568,576 | 0.9286 | -0.0078 | 0.0110 | 4.77E-01 | 559,070 | 0.9234 | -0.0070 | 0.0128 |
| **OSAadjBMI** | **multi-pop** | rs4759254 | 12 | 57096674 | *NACA* | T | C | 0.6013 | -0.0342 | 0.0049 | **4.12E-12** | 828,690 | 0.5814 | -0.0256 | 0.0056 | **2.67E-06** | 568,576 | 0.5808 | -0.0261 | 0.0058 | **4.75E-06** | 559,070 | 0.6830 | -0.0245 | 0.0073 |
| **OSA** | **multi-pop** | rs2958127 | 12 | 57114307 | *NACA* | A | T | 0.3726 | 0.0313 | 0.0038 | **2.22E-16** | 1,471,068 | 0.4109 | 0.0273 | 0.0053 | **1.12E-07** | 568,576 | 0.4113 | 0.0285 | 0.1513 | **1.17E-07** | 559,070 | 0.3102 | 0.0235 | 0.0074 |
| **OSAadjBMI** | **multi-pop** | rs10735928 | 12 | 65769626 | *MSRB3* | A | C | 0.3834 | 0.0340 | 0.0048 | **1.71E-12** | 828,690 | 0.3840 | 0.0178 | 0.0055 | **9.36E-04** | 568,576 | 0.3841 | 0.0254 | 0.0057 | **6.85E-06** | 559,070 | 0.2922 | 0.0360 | 0.0075 |
| **OSA** | **multi-pop** | rs11642015 | 16 | 53802494 | *FTO* | T | C | 0.3767 | 0.0862 | 0.0039 | **2.99E-108** | 1,471,068 | 0.3305 | 0.0801 | 0.0060 | **3.89E-47** | 568,576 | 0.3301 | 0.0098 | 0.0058 | 9.05E-02 | 559,070 | 0.4183 | 0.0857 | 0.0069 |
| **OSA** | **multi-pop** | rs1800437 | 19 | 46181392 | *GIPR* | C | G | 0.2118 | -0.0416 | 0.0044 | **3.24E-21** | 1,471,068 | 0.1840 | -0.0288 | 0.0063 | **1.12E-05** | 568,576 | 0.1838 | 0.0041 | 0.0068 | 5.53E-01 | 559,070 | 0.2583 | -0.0504 | 0.0078 |
| **OSA** | **multi-pop** | rs11088984 | 21 | 46670045 | *POFUT2* | A | C | 0.6641 | 0.0180 | 0.0041 | 1.45E-05 | 1,471,068 | 0.6980 | 0.0089 | 0.0064 | 1.73E-01 | 568,576 | 0.6984 | 0.0026 | 0.0067 | 7.04E-01 | 559,070 | 0.6139 | 0.0070 | 0.0070 |

*Published GWAS included in meta-analysis was not adjusted for BMI.

**Supplementary Table 9.** Lookups of top associa

| Discovery Trait | Discovery Strata | dbSNP ID | p Apnea | | SDKP - AHI | | | | | SDKP - avgSAO2 (SpO -average) | | | | | Discovery | | | | |
|---|---|---|---|---|---|---|---|---|---|---|---|---|---|---|---|---|---|---|---|
| | | | P | N | EAF | BETA | SE | P | N | EAF | BETA | SE | P | N | EAF | BETA | SE | P | N |
| **$AvgSaO_2$ adjBMI*** | **multi-pop** | rs79914264 | 6.82E-01 | 410,385 | NR | -0.0136 | 0.0285 | 6.34E-01 | 21,244 | | 0.0426 | 0.0328 | 1.93E-01 | 21,127 | 0.016 | 0.206 | 0.038 | **4.04E-08** | 4,457 |
| **OSA** | **multi-pop** | rs12725199 | 2.06E-01 | 410,385 | 0.1910 | -0.0136 | 0.0121 | 2.59E-01 | 19,733 | 0.1910 | -0.0005 | 0.0120 | 9.66E-01 | 20,861 | 0.253 | 0.055 | 0.009 | **1.54E-09** | 492,107 |
| **OSA** | **multi-pop** | rs2376073 | 1.60E-02 | 410,385 | 0.1750 | -0.0018 | 0.0125 | 8.84E-01 | 19,733 | 0.1740 | 0.0087 | 0.0125 | 4.85E-01 | 21,089 | 0.112 | 0.070 | 0.013 | **3.03E-08** | 492,107 |
| **AHI** | **multi-pop** | rs1200596362 | | | 0.3700 | 0.0264 | 0.0167 | 1.13E-01 | 6,953 | 0.3690 | 0.0074 | 0.0158 | 6.42E-01 | 7,810 | 0.349 | 0.093 | 0.017 | **2.84E-08** | 8,152 |
| **OSA** | **multi-pop** | rs9831061 | **1.60E-04** | 410,385 | 0.6330 | 0.0126 | 0.0101 | 2.14E-01 | 19,733 | 0.6330 | 0.0024 | 0.0101 | 8.14E-01 | 21,089 | 0.726 | 0.051 | 0.009 | **1.21E-08** | 492,107 |
| **OSA** | **EUR-only** | rs34592089 | 8.03E-02 | 410,385 | 0.0410 | 0.0239 | 0.0248 | 3.35E-01 | 18,042 | 0.0420 | 0.0088 | 0.0254 | 7.29E-01 | 17,527 | 0.055 | 0.113 | 0.020 | **1.81E-08** | 449,087 |
| **OSAadjBMI** | **multi-pop** | rs2964006 | **3.26E-03** | 410,385 | 0.3650 | -0.0077 | 0.0098 | 4.35E-01 | 19,733 | 0.3650 | 0.0071 | 0.0098 | 4.68E-01 | 21,089 | 0.450 | 0.049 | 0.009 | **2.24E-08** | 268,537 |
| **OSA** | **multi-pop** | rs9380814 | 6.80E-03 | 410,385 | 0.2520 | -0.0056 | 0.0108 | 6.08E-01 | 19,733 | 0.2490 | -0.0155 | 0.0109 | 1.52E-01 | 21,089 | 0.324 | -0.048 | 0.009 | **1.82E-08** | 492,107 |
| **AHI adjBMI*** | **multi-pop** | rs78545995 | 4.66E-01 | 410,385 | 0.9840 | 0.0379 | 0.0440 | 3.89E-01 | 13,447 | 0.9840 | 0.0451 | 0.0467 | 3.34E-01 | 12,825 | 0.987 | 0.299 | 0.054 | **3.64E-08** | 15,981 |
| **$AvgSaO_2$** | **multi-pop** | rs802729 | | | NR | 0.0268 | 0.0507 | 5.98E-01 | 18,073 | | -0.0072 | 0.0668 | 9.14E-01 | 17,987 | 0.985 | -0.427 | 0.076 | **1.84E-08** | 2,634 |
| **$AvgSaO_2$** | **EUR-only** | rs2194955 | 6.22E-01 | 410,385 | 0.7060 | -0.0051 | 0.0104 | 6.27E-01 | 19,732 | 0.7070 | 0.0041 | 0.0104 | 6.94E-01 | 21,089 | 0.819 | -0.143 | 0.025 | **7.17E-09** | 5,872 |
| **OSA** | **multi-pop** | rs66474734 | | | 0.5160 | 0.0143 | 0.0164 | 3.85E-01 | 6,953 | 0.5210 | -0.0173 | 0.0156 | 2.67E-01 | 7,810 | 0.582 | -0.062 | 0.011 | **2.47E-08** | 122,648 |
| **OSA** | **multi-pop** | rs12553324 | **1.73E-05** | 410,385 | 0.6510 | 0.0061 | 0.0097 | 5.28E-01 | 19,733 | 0.6530 | 0.0106 | 0.0097 | 2.76E-01 | 21,089 | 0.624 | -0.052 | 0.009 | **3.50E-08** | 202,804 |
| **OSAadjBMI** | **EUR-only** | rs60791051 | 5.85E-01 | 410,385 | NR | -0.0115 | 0.0187 | 5.38E-01 | 21,244 | | 0.0206 | 0.0235 | 3.82E-01 | 21,127 | 0.939 | -0.139 | 0.026 | **4.80E-08** | 104,016 |
| **OSAadjBMI** | **multi-pop** | rs4759254 | **8.32E-04** | 410,385 | 0.6380 | -0.0184 | 0.0101 | 6.78E-02 | 19,733 | 0.6290 | 0.0042 | 0.0100 | 6.77E-01 | 21,089 | 0.644 | -0.055 | 0.009 | **2.99E-09** | 269,620 |
| **OSA** | **multi-pop** | rs2958127 | **1.39E-03** | 410,385 | 0.3650 | 0.0135 | 0.0098 | 1.70E-01 | 19,733 | 0.3720 | -0.0032 | 0.0098 | 7.42E-01 | 21,089 | 0.380 | 0.050 | 0.008 | **5.75E-10** | 492,107 |
| **OSAadjBMI** | **multi-pop** | rs10735928 | **1.41E-06** | 410,385 | 0.3070 | 0.0306 | 0.0104 | **3.21E-03** | 19,733 | 0.3130 | -0.0057 | 0.0102 | 5.76E-01 | 21,089 | 0.382 | 0.054 | 0.009 | **7.73E-10** | 269,620 |
| **OSA** | **multi-pop** | rs11642015 | **1.06E-35** | 410,385 | 0.2910 | 0.0020 | 0.0106 | 8.53E-01 | 19,733 | 0.2890 | -0.0005 | 0.0105 | 9.66E-01 | 21,089 | 0.396 | 0.098 | 0.008 | **8.47E-34** | 492,107 |
| **OSA** | **multi-pop** | rs1800437 | **1.23E-10** | 410,385 | 0.1500 | 0.0107 | 0.0147 | 4.66E-01 | 15,605 | 0.1520 | 0.0065 | 0.0146 | 6.57E-01 | 16,496 | 0.205 | -0.059 | 0.010 | **2.23E-09** | 492,107 |
| **OSA** | **multi-pop** | rs11088984 | 3.18E-01 | 410,385 | 0.7210 | 0.0066 | 0.0105 | 5.30E-01 | 19,732 | 0.7240 | -0.0062 | 0.0104 | 5.53E-01 | 21,089 | 0.667 | 0.051 | 0.009 | **3.94E-09** | 492,107 |

*Published GWAS included in meta-analysis wa

**Supplementary Table 10.** Lookup of previously-published OSA and OSAadjBMI SNPs in discovery analysis.
All significant associations are highlighted in red, based on Bonferroni-corrected significance for 56 loci (+/- 500 kb). Loci are numbered based on chromosome and position. Each SNP is considered replicated if at least one stratum reached significance in the same trait and model (adjusted for BMI or not adjusted for BMI) and directionally consistent with discovery GWAS. Marginal replications (P<0.05) are noted by N*. Abbreviations: CHR - chromosome, POS - position, A1 - effect allele, A2 - other allele, EAF - effect allele frequency, OR - odds ration, Y - yes, N - No, N* - marginally significant, BETA - effect estimate, SE - standard error, N - sample size, EUR - European, NA - not available.

| | | | | | | | | | | | | | Current Discovery Results - Stage 1 + Stage 2 | | | | | | | | | | | |
|---|---|---|---|---|---|---|---|---|---|---|---|---|---|---|---|---|---|---|---|---|---|---|---|---|
| | | | | | | | Published Results | | | | | Replicated | | | Multi-Population | | | | | EUR only | | | | |
| Published Trait | dbSNP ID | CHR | POS (hg19) | Locus # | Publication | Reported Gene | A1 | A2 | EAF | OR | P | (Y/N*/N) | A1 | A2 | EAF | BETA | SE | P | N | EAF | BETA | SE | P | N |
| OSA | | | | | | | | | | | | | | | | | | | | | | | | |
| OSA - Meta | rs72663521 | 1 | 40005903 | 1 | Sofer et al 2023 | *BMP8A* | A | G | 0.196 | 1.037 | 2.70E-08 | Y | A | G | 0.176 | 0.039 | 0.010 | **1.51E-04** | 492,107 | 0.178 | 0.036 | 0.011 | **8.97E-04** | 449,452 |
| OSA - Meta | rs1087782 | 1 | 75577086 | 2 | Sofer et al 2023 | *AC133865.1* | T | C | 0.408 | 1.030 | 2.80E-09 | N | T | C | 0.442 | 0.018 | 0.011 | 1.10E-01 | 122,648 | 0.468 | 0.020 | 0.012 | 9.09E-02 | 103,660 |
| OSA - Meta | rs10874656 | 1 | 92947024 | 3 | Sofer et al 2023 | *GFI1* | C | T | 0.801 | 0.966 | 2.40E-08 | N* | T | C | 0.186 | 0.020 | 0.010 | **4.98E-02** | 492,107 | 0.198 | 0.018 | 0.010 | 8.55E-02 | 452,820 |
| OSA - Meta | rs6679458 | 1 | 96946253 | 4 | Sofer et al 2023 | *LINC01787* | T | G | 0.544 | 1.034 | 4.90E-12 | Y | T | G | 0.562 | 0.040 | 0.008 | **7.89E-07** | 492,107 | 0.590 | 0.041 | 0.009 | **1.39E-06** | 453,740 |
| OSA - MVP only | rs17367240 | 1 | 96960057 | 4 | Sofer et al 2023 | *LINC01787* | A | G | 0.380 | 0.967 | 2.10E-09 | Y | A | G | 0.342 | -0.036 | 0.009 | **2.15E-05** | 492,107 | 0.310 | -0.036 | 0.009 | **8.99E-05** | 453,195 |
| OSA | rs34013490 | 1 | 97032649 | 4 | Strausz et al 2024 | *PTBP2* | CT | C | 0.660 | 1.051 | 6.33E-09 | - | NA | NA | NA | NA | NA | NA | NA | NA | NA | NA | NA | NA |
| OSA - MVP only | rs11691026 | 2 | 22131865 | 6 | Sofer et al 2023 | *AC018742.1* | A | G | 0.440 | 1.032 | 2.30E-09 | N | A | G | 0.465 | 0.006 | 0.011 | 6.03E-01 | 122,648 | 0.478 | 0.003 | 0.012 | 7.83E-01 | 103,051 |
| OSA - Meta | rs11691026 | 2 | 22131865 | 6 | Sofer et al 2023 | *AC018742.1* | A | G | 0.463 | 1.026 | 3.50E-08 | N | A | G | 0.465 | 0.006 | 0.011 | 6.03E-01 | 122,648 | 0.478 | 0.003 | 0.012 | 7.83E-01 | 103,051 |
| OSA - Meta | rs77881454 | 2 | 43757293 | 7 | Sofer et al 2023 | *THADA* | T | A | 0.132 | 1.050 | 1.20E-10 | N* | A | T | 0.894 | -0.038 | 0.013 | **2.50E-03** | 492,107 | 0.896 | -0.041 | 0.014 | **2.57E-03** | 453,349 |
| OSA - MVP only | rs77881454 | 2 | 43757293 | 7 | Sofer et al 2023 | *THADA* | T | A | 0.110 | 1.054 | 6.70E-11 | N* | A | T | 0.894 | -0.038 | 0.013 | **2.50E-03** | 492,107 | 0.896 | -0.041 | 0.014 | **2.57E-03** | 453,349 |
| OSA - Meta | rs540606 | 2 | 45138507 | 8 | Sofer et al 2023 | *CAMKMT* | G | A | 0.619 | 0.970 | 2.60E-10 | N* | A | G | 0.397 | 0.017 | 0.008 | **3.82E-02** | 492,107 | 0.411 | 0.018 | 0.009 | **3.40E-02** | 449,502 |
| OSA - MVP only | rs540606 | 2 | 45138507 | 8 | Sofer et al 2023 | *CAMKMT* | A | G | 0.390 | 1.035 | 3.70E-10 | N* | A | G | 0.397 | 0.017 | 0.008 | **3.82E-02** | 492,107 | 0.411 | 0.018 | 0.009 | **3.40E-02** | 449,502 |
| OSA - Meta | rs58857776 | 2 | 58890684 | 10 | Sofer et al 2023 | *LINC01122* | T | C | 0.448 | 0.966 | 3.50E-13 | N* | T | C | 0.420 | -0.020 | 0.008 | **1.11E-02** | 492,107 | 0.421 | -0.023 | 0.009 | **6.76E-03** | 452,170 |
| OSA - MVP only | rs58857776 | 2 | 58890684 | 10 | Sofer et al 2023 | *LINC01122* | T | C | 0.410 | 0.963 | 5.10E-13 | N* | T | C | 0.420 | -0.020 | 0.008 | **1.11E-02** | 492,107 | 0.421 | -0.023 | 0.009 | **6.76E-03** | 452,170 |
| OSA | rs76229479 | 2 | 103123912 | 11 | Strausz et al 2024 | *SLC9A4* | C | A | 0.100 | 0.923 | 2.64E-09 | N | A | C | 0.876 | 0.011 | 0.012 | 3.52E-01 | 492,107 | 0.877 | 0.013 | 0.013 | 3.18E-01 | 452,064 |
| OSA - Meta | rs4953923 | 2 | 135445946 | 12 | Sofer et al 2023 | *TMEM163* | T | C | 0.169 | 0.963 | 3.20E-08 | N | T | C | 0.150 | 0.004 | 0.012 | 7.25E-01 | 492,107 | 0.153 | 0.004 | 0.012 | 7.19E-01 | 451,433 |
| OSA - Meta | rs6759321 | 2 | 136322676 | 13 | Sofer et al 2023 | *R3HDM1* | T | G | 0.426 | 0.970 | 2.60E-08 | N | T | G | 0.472 | -0.011 | 0.011 | 3.02E-01 | 202,804 | 0.453 | 0.003 | 0.015 | 8.25E-01 | 119,301 |
| OSA | rs10928560 | 2 | 136991807 | 14 | Strausz et al 2021 | *CXCR4* | T | C | 0.200 | 0.920 | 2.80E-08 | N | T | C | 0.185 | -0.011 | 0.010 | 2.78E-01 | 492,107 | 0.191 | -0.005 | 0.011 | 6.39E-01 | 450,471 |
| OSA - MVP only | rs181355045 | 2 | 156998417 | 15 | Sofer et al 2023 | *LINC01876* | A | C | 0.110 | 1.046 | 2.80E-08 | N* | A | C | 0.117 | 0.039 | 0.017 | **2.56E-02** | 122,648 | 0.115 | 0.051 | 0.018 | **6.12E-03** | 101,976 |
| OSA - Meta | rs181355045 | 2 | 156998417 | 15 | Sofer et al 2023 | *LINC01876* | A | C | 0.149 | 1.042 | 1.20E-08 | N* | A | C | 0.117 | 0.039 | 0.017 | **2.56E-02** | 122,648 | 0.115 | 0.051 | 0.018 | **6.12E-03** | 101,976 |
| OSA - Meta | rs67955391 | 3 | 85640136 | 16 | Sofer et al 2023 | *CADM2* | G | C | 0.597 | 0.973 | 2.80E-08 | N* | C | G | 0.412 | 0.020 | 0.008 | **1.43E-02** | 492,107 | 0.380 | 0.018 | 0.009 | **4.12E-02** | 450,161 |
| OSA | rs114106239 | 3 | 132497894 | 17 | Strausz et al 2024 | *NPHP3-ACAD11* | T | C | 0.040 | 0.887 | 1.19E-08 | N | T | C | 0.016 | 0.029 | 0.039 | 4.53E-01 | 410,631 | 0.018 | 0.032 | 0.041 | 4.31E-01 | 375,611 |
| OSA | rs181010833 | 3 | 168877736 | 19 | Strausz et al 2024 | *MECOM* | T | A | 0.040 | 0.887 | 1.52E-08 | - | NA | NA | NA | NA | NA | NA | NA | NA | NA | NA | NA | NA |
| OSA - MVP only | rs869400 | 3 | 185826740 | 20 | Sofer et al 2023 | *ETV5* | T | G | 0.200 | 0.950 | 2.60E-13 | Y | T | G | 0.190 | -0.048 | 0.010 | **1.55E-06** | 492,107 | 0.187 | -0.054 | 0.011 | **6.67E-07** | 452,733 |
| OSA - Meta | rs869400 | 3 | 185826740 | 20 | Sofer et al 2023 | *ETV5* | T | G | 0.187 | 1.052 | 5.30E-16 | Y | T | G | 0.190 | -0.048 | 0.010 | **1.55E-06** | 492,107 | 0.187 | -0.054 | 0.011 | **6.67E-07** | 452,733 |
| OSA - Meta | rs12507026 | 4 | 45181334 | 21 | Sofer et al 2023 | *AC108467.1* | T | A | 0.423 | 1.031 | 5.60E-11 | Y | A | T | 0.587 | -0.037 | 0.009 | **6.98E-05** | 202,804 | 0.566 | -0.042 | 0.010 | **2.57E-05** | 164,303 |
| OSA - MVP only | rs10938397 | 4 | 45182527 | 21 | Sofer et al 2023 | *AC108467.1* | G | A | 0.390 | 1.032 | 1.40E-09 | Y | A | G | 0.589 | -0.035 | 0.008 | **1.02E-05** | 492,107 | 0.574 | -0.039 | 0.008 | **3.40E-06** | 453,537 |
| OSA | rs34925548 | 4 | 47313520 | 22 | Strausz et al 2024 | *GABRB1* | CA | C | 0.700 | 0.951 | 1.12E-08 | - | NA | NA | NA | NA | NA | NA | NA | NA | NA | NA | NA | NA |
| OSA - MVP only | rs13107325 | 4 | 103188709 | 23 | Sofer et al 2023 | *SLC39A8* | T | C | 0.070 | 1.085 | 3.60E-16 | Y | T | C | 0.070 | 0.076 | 0.016 | **4.36E-06** | 492,107 | 0.076 | 0.075 | 0.017 | **1.12E-05** | 453,360 |
| OSA - Meta | rs13107325 | 4 | 103188709 | 23 | Sofer et al 2023 | *SLC39A8* | T | C | 0.086 | 1.073 | 5.80E-13 | Y | T | C | 0.070 | 0.076 | 0.016 | **4.36E-06** | 492,107 | 0.076 | 0.075 | 0.017 | **1.12E-05** | 453,360 |
| OSA - Meta | rs12514158 | 5 | 95857763 | 24 | Sofer et al 2023 | *LOC101929710* | T | C | 0.520 | 0.973 | 4.30E-09 | N | T | C | 0.580 | -0.001 | 0.008 | 8.75E-01 | 492,107 | 0.588 | -0.002 | 0.009 | 8.16E-01 | 452,585 |
| OSA | rs60700772 | 6 | 7746663 | 25 | Strausz et al 2024 | *BMP6* | C | T | 0.220 | 1.051 | 2.20E-08 | N | T | C | 0.788 | -0.007 | 0.010 | 4.73E-01 | 492,107 | 0.779 | -0.005 | 0.010 | 6.33E-01 | 448,248 |
| OSA - Meta | rs6938973 | 6 | 98421721 | 26 | Sofer et al 2023 | *AL589740.1* | C | T | 0.646 | 1.030 | 1.60E-09 | N | T | C | 0.386 | -0.011 | 0.008 | 1.80E-01 | 492,107 | 0.398 | -0.008 | 0.009 | 3.31E-01 | 453,078 |
| OSA | rs11981973 | 7 | 69445341 | 27 | Strausz et al 2024 | *AUTS2* | G | A | 0.180 | 1.062 | 4.72E-10 | N* | A | G | 0.810 | -0.028 | 0.010 | **5.70E-03** | 492,107 | 0.821 | -0.024 | 0.011 | **2.41E-02** | 453,232 |
| OSA | rs17148727 | 7 | 74444066 | 28 | Strausz et al 2024 | *RCC1L* | C | A | 0.680 | 0.951 | 1.02E-08 | N | A | C | 0.420 | 0.011 | 0.020 | 5.85E-01 | 41,172 | 0.402 | -0.016 | 0.026 | 5.35E-01 | 26,897 |
| OSA | rs679880 | 9 | 79267071 | 30 | Strausz et al 2024 | *PRUNE2* | A | G | 0.750 | 1.051 | 2.53E-08 | N* | A | G | 0.763 | 0.020 | 0.010 | **3.65E-02** | 492,107 | 0.799 | 0.025 | 0.011 | **1.88E-02** | 454,019 |
| OSA - Meta | rs7869742 | 9 | 95256174 | 31 | Sofer et al 2023 | *ECM2* | G | A | 0.407 | 1.028 | 1.10E-08 | N* | A | G | 0.577 | -0.023 | 0.008 | **5.79E-03** | 492,107 | 0.605 | -0.024 | 0.009 | **5.32E-03** | 454,019 |
| OSA - Meta | rs466614 | 9 | 128027707 | 32 | Sofer et al 2023 | *GAPVD1* | T | C | 0.536 | 0.964 | 2.80E-13 | N* | T | C | 0.516 | -0.019 | 0.008 | **1.83E-02** | 492,107 | 0.487 | -0.019 | 0.008 | **2.36E-02** | 454,019 |
| OSA - MVP only | rs5015933 | 9 | 128137418 | 32 | Sofer et al 2023 | *GAPVD1* | C | T | 0.510 | 0.970 | 2.60E-09 | N* | T | C | 0.514 | 0.017 | 0.008 | **3.29E-02** | 492,107 | 0.531 | 0.023 | 0.008 | **6.73E-03** | 454,019 |
| OSA | rs10986730 | 9 | 128137874 | 32 | Strausz et al 2024 | *GAPVD1* | T | C | 0.520 | 0.951 | 1.16E-09 | Y | T | C | 0.455 | -0.023 | 0.008 | **3.90E-03** | 492,107 | 0.444 | -0.028 | 0.008 | **8.68E-04** | 454,019 |
| OSA | rs4837016 | 9 | 128141809 | 32 | Strausz et al 2021 | *GAPVD1* | A | G | 0.470 | 0.930 | 1.50E-08 | N* | A | G | 0.436 | -0.024 | 0.008 | **1.99E-03** | 492,107 | 0.430 | -0.027 | 0.008 | **1.56E-03** | 454,019 |
| OSA | rs185932673 | 10 | 12698439 | 33 | Strausz et al 2021 | *CAMK1D* | T | C | 0.003 | 1.870 | 2.40E-08 | N | T | C | NA | NA | NA | NA | NA | 0.005 | -0.062 | 0.082 | 4.47E-01 | 375,611 |
| OSA | rs113955098 | 10 | 18591226 | 34 | Strausz et al 2024 | *CACNB2* | A | G | 0.070 | 0.905 | 3.38E-10 | N | A | G | 0.015 | -0.004 | 0.037 | 9.22E-01 | 439,042 | 0.018 | -0.030 | 0.035 | 3.83E-01 | 454,019 |
| OSA - Meta | rs2506141 | 10 | 33468014 | 35 | Sofer et al 2023 | *NRP1* | C | T | 0.514 | 0.974 | 4.40E-08 | N | T | C | 0.509 | 0.010 | 0.008 | 2.13E-01 | 492,107 | 0.536 | 0.010 | 0.008 | 2.52E-01 | 454,019 |

| | | | | | | | | | | | | | Current Discovery Results - Stage 1 + Stage 2 | | | | | | | | | | | |
|---|---|---|---|---|---|---|---|---|---|---|---|---|---|---|---|---|---|---|---|---|---|---|---|---|
| | | | | | | | Published Results | | | | | Replicated | | | Multi-Population | | | | | EUR only | | | | |
| Published Trait | dbSNP ID | CHR | POS (hg19) | Locus # | Publication | Reported Gene | A1 | A2 | EAF | OR | P | (Y/N*/N) | A1 | A2 | EAF | BETA | SE | P | N | EAF | BETA | SE | P | N |
| OSA | rs61873510 | 10 | 102626510 | 36 | Strausz et al 2024 | *PAX2* | T | G | 0.300 | 1.051 | 3.28E-08 | N* | T | G | 0.330 | 0.022 | 0.009 | **1.01E-02** | 492,107 | 0.350 | 0.024 | 0.009 | **7.40E-03** | 454,019 |
| OSA - Meta | rs943037 | 10 | 104835919 | 37 | Sofer et al 2023 | *CNNM2* | T | C | 0.120 | 1.047 | 1.90E-08 | N | T | C | 0.084 | 0.002 | 0.014 | 8.68E-01 | 492,107 | 0.086 | -0.003 | 0.015 | 8.25E-01 | 454,019 |
| OSA - MVP only | rs201575845 | 10 | 104861271 | 37 | Sofer et al 2023 | *NT5C2* | A | AT | 0.100 | 1.052 | 2.70E-08 | N | A | AT | 0.689 | 0.005 | 0.015 | 7.24E-01 | 81,476 | 0.688 | 0.003 | 0.015 | 8.65E-01 | 78,408 |
| OSA - Meta | rs34872471 | 10 | 114754071 | 38 | Sofer et al 2023 | *TCF7L2* | C | T | 0.282 | 0.972 | 4.60E-08 | Y | T | C | 0.700 | 0.031 | 0.009 | **3.62E-04** | 492,107 | 0.703 | 0.028 | 0.009 | **2.45E-03** | 454,019 |
| OSA - MVP only | rs34872471 | 10 | 114754071 | 38 | Sofer et al 2023 | *TCF7L2* | C | T | 0.300 | 0.967 | 1.90E-09 | Y | T | C | 0.700 | 0.031 | 0.009 | **3.62E-04** | 492,107 | 0.703 | 0.028 | 0.009 | **2.45E-03** | 454,019 |
| OSA | rs6484367 | 11 | 28387459 | 39 | Strausz et al 2024 | *METTL15* | A | G | 0.500 | 1.051 | 1.13E-09 | N* | A | G | 0.511 | 0.021 | 0.008 | **8.89E-03** | 492,107 | 0.491 | 0.016 | 0.009 | 6.17E-02 | 454,019 |
| OSA - Meta | rs12229673 | 12 | 534957 | 40 | Sofer et al 2023 | *CCDC77* | A | G | 0.405 | 1.029 | 1.30E-08 | N | A | G | 0.421 | 0.015 | 0.008 | 6.21E-02 | 492,107 | 0.447 | 0.014 | 0.009 | 9.43E-02 | 454,019 |
| OSA | rs59333125 | 12 | 57025908 | 41 | Strausz et al 2024 | *BAZ2A* | C | A | 0.080 | 0.923 | 1.32E-08 | N* | A | C | 0.935 | 0.050 | 0.017 | **3.10E-03** | 492,107 | 0.934 | 0.047 | 0.018 | **9.52E-03** | 454,019 |
| OSA - MVP only | rs2277339 | 12 | 57146069 | 41 | Sofer et al 2023 | *PRIM1* | G | T | 0.120 | 0.954 | 9.10E-10 | N* | T | G | 0.889 | 0.030 | 0.013 | **1.71E-02** | 492,107 | 0.898 | 0.028 | 0.014 | **4.13E-02** | 454,019 |
| OSA - Meta | rs2277339 | 12 | 57146069 | 41 | Sofer et al 2023 | *PRIM1* | G | T | 0.159 | 0.954 | 1.20E-11 | N* | T | G | 0.889 | 0.030 | 0.013 | **1.71E-02** | 492,107 | 0.898 | 0.028 | 0.014 | **4.13E-02** | 454,019 |
| OSA | rs10507084 | 12 | 97753152 | 43 | Strausz et al 2021 | *RMST/NEDD1* | T | C | 0.180 | 1.110 | 2.80E-11 | N | T | C | 0.083 | 0.008 | 0.015 | 5.87E-01 | 492,107 | 0.063 | 0.001 | 0.018 | 9.47E-01 | 454,019 |
| OSA | rs10507084 | 12 | 97753152 | 43 | Strausz et al 2024 | *NEDD1* | T | C | 0.180 | 1.062 | 8.23E-11 | N | T | C | 0.083 | 0.008 | 0.015 | 5.87E-01 | 492,107 | 0.063 | 0.001 | 0.018 | 9.47E-01 | 454,019 |
| OSA | rs2016950 | 12 | 107980323 | 44 | Strausz et al 2024 | *BTBD11* | T | C | 0.160 | 0.942 | 4.13E-08 | N | T | C | 0.137 | 0.000 | 0.012 | 9.98E-01 | 492,107 | 0.132 | 0.000 | 0.013 | 9.98E-01 | 454,019 |
| OSA - MVP only | rs201770 | 13 | 51047531 | 45 | Sofer et al 2023 | *DLEU1* | G | A | 0.220 | 0.965 | 1.30E-08 | N | A | G | 0.760 | 0.024 | 0.013 | 6.32E-02 | 122,648 | 0.747 | 0.026 | 0.014 | 5.62E-02 | 105,305 |
| OSA - Meta | rs1373285 | 13 | 54048078 | 46 | Sofer et al 2023 | *AL356295.1* | A | G | 0.311 | 1.033 | 9.00E-10 | Y | A | G | 0.320 | 0.025 | 0.009 | **2.80E-03** | 492,107 | 0.338 | 0.032 | 0.009 | **3.30E-04** | 454,019 |
| OSA | rs2370982 | 14 | 79890677 | 47 | Strausz et al 2024 | *NRXN3* | T | C | 0.240 | 1.051 | 7.97E-09 | N | T | C | 0.208 | 0.014 | 0.010 | 1.60E-01 | 492,107 | 0.207 | 0.014 | 0.010 | 1.81E-01 | 454,019 |
| OSA - Meta | rs10150332 | 14 | 79936964 | 47 | Sofer et al 2023 | *NRXN3* | C | T | 0.268 | 1.031 | 1.30E-08 | N | T | C | 0.774 | -0.018 | 0.009 | 5.42E-02 | 492,107 | 0.788 | -0.016 | 0.010 | 1.22E-01 | 454,019 |
| OSA - MVP only | rs283789 | 15 | 76777577 | 48 | Sofer et al 2023 | *SCAPER* | G | A | 0.360 | 0.970 | 2.60E-08 | N | A | G | 0.663 | 0.007 | 0.008 | 4.21E-01 | 492,107 | 0.673 | 0.006 | 0.009 | 5.07E-01 | 454,019 |
| OSA - Meta | rs283789 | 15 | 76777577 | 48 | Sofer et al 2023 | *SCAPER* | G | A | 0.648 | 1.028 | 1.00E-08 | N | A | G | 0.663 | 0.007 | 0.008 | 4.21E-01 | 492,107 | 0.673 | 0.006 | 0.009 | 5.07E-01 | 454,019 |
| OSA | rs9937053 | 16 | 53799507 | 49 | Strausz et al 2021 | *FTO* | A | G | 0.430 | 1.110 | 4.30E-16 | Y | A | G | 0.437 | 0.090 | 0.008 | **1.78E-30** | 492,107 | 0.440 | 0.097 | 0.008 | **5.00E-31** | 454,019 |
| OSA - Meta | rs62048402 | 16 | 53803223 | 49 | Sofer et al 2023 | *FTO* | A | G | 0.373 | 1.077 | 5.92E-52 | Y | A | G | 0.396 | 0.098 | 0.008 | **1.37E-33** | 492,107 | 0.420 | 0.100 | 0.008 | **1.82E-32** | 454,019 |
| OSA - MVP only | rs1558902 | 16 | 53803574 | 49 | Sofer et al 2023 | *FTO* | A | T | 0.330 | 1.083 | 3.80E-47 | Y | A | T | 0.389 | 0.102 | 0.009 | **4.47E-27** | 202,804 | 0.423 | 0.105 | 0.010 | **5.00E-26** | 164,716 |
| OSA | rs11075985 | 16 | 53805207 | 49 | Strausz et al 2024 | *FTO* | A | C | 0.430 | 1.083 | 2.15E-26 | Y | A | C | 0.438 | 0.090 | 0.008 | **9.70E-31** | 492,107 | 0.441 | 0.097 | 0.008 | **2.61E-31** | 454,019 |
| OSA | rs13333522 | 16 | 73808619 | 50 | Strausz et al 2024 | *PSMD7* | G | C | 0.530 | 1.041 | 4.06E-08 | N | C | G | 0.510 | 0.002 | 0.009 | 8.38E-01 | 202,804 | 0.496 | 0.003 | 0.010 | 7.65E-01 | 164,716 |
| OSA - MVP only | rs58879558 | 17 | 44095467 | 51 | Sofer et al 2023 | *MAPT* | C | T | 0.200 | 1.040 | 8.20E-10 | N* | T | C | 0.791 | -0.030 | 0.010 | **1.90E-03** | 492,107 | 0.780 | -0.034 | 0.010 | **9.14E-04** | 454,019 |
| OSA - Meta | rs199445 | 17 | 44817408 | 52 | Sofer et al 2023 | *NSF* | T | C | 0.175 | 1.035 | 3.60E-08 | N* | T | C | 0.186 | 0.025 | 0.010 | **1.21E-02** | 492,107 | 0.195 | 0.025 | 0.011 | **1.94E-02** | 454,019 |
| OSA - Meta | rs429358 | 19 | 45411941 | 53 | Sofer et al 2023 | *APOE* | C | T | 0.193 | 0.960 | 5.50E-11 | Y | T | C | 0.851 | 0.045 | 0.011 | **5.23E-05** | 492,107 | 0.854 | 0.046 | 0.012 | **1.49E-04** | 454,019 |
| OSA - MVP only | rs429358 | 19 | 45411941 | 53 | Sofer et al 2023 | *APOE* | C | T | 0.150 | 0.955 | 3.40E-11 | Y | T | C | 0.851 | 0.045 | 0.011 | **5.23E-05** | 492,107 | 0.854 | 0.046 | 0.012 | **1.49E-04** | 454,019 |
| OSA | rs10423928 | 19 | 46182304 | 54 | Strausz et al 2024 | *GIPR* | A | T | 0.260 | 0.951 | 7.55E-09 | Y | A | T | 0.208 | -0.057 | 0.010 | **5.10E-09** | 492,107 | 0.213 | -0.057 | 0.010 | **2.28E-08** | 454,019 |
| OSA - Meta | rs13043844 | 20 | 50959453 | 55 | Sofer et al 2023 | *LINC01524* | G | C | 0.178 | 0.962 | 6.60E-09 | N* | C | G | 0.836 | 0.025 | 0.011 | **1.94E-02** | 492,107 | 0.826 | 0.029 | 0.011 | **1.06E-02** | 454,019 |
| OSA | rs4809902 | 20 | 50979653 | 55 | Strausz et al 2024 | *ZFP64* | C | G | 0.230 | 0.942 | 1.51E-09 | N | C | G | 0.297 | -0.014 | 0.009 | 1.04E-01 | 492,107 | 0.282 | -0.016 | 0.009 | 9.55E-02 | 454,019 |
| OSA | rs140896965 | 20 | 57648366 | 56 | Strausz et al 2024 | *PRELID3B* | T | C | 0.050 | 0.896 | 1.11E-09 | N* | T | C | 0.015 | -0.124 | 0.051 | **1.52E-02** | 410,631 | 0.015 | -0.121 | 0.052 | **2.06E-02** | 375,611 |
| **OSAadjBMI** | | | | | | | | | | | | | | | | | | | | | | | | |
| OSAadjBMI - Meta | rs1087782 | 1 | 75577086 | 2 | Sofer et al 2023 | *AC133865.1* | T | C | 0.407 | 1.030 | 3.30E-09 | N | T | C | 0.445 | 0.017 | 0.012 | 1.69E-01 | 118,909 | 0.468 | 0.020 | 0.013 | 1.09E-01 | 102,385 |
| OSAadjBMI - MVP only | rs79950770 | 1 | 119539797 | 5 | Sofer et al 2023 | *TBX15* | T | C | 0.090 | 1.062 | 2.90E-10 | Y | T | C | 0.107 | 0.047 | 0.014 | **9.49E-04** | 267,661 | 0.112 | 0.048 | 0.014 | **8.80E-04** | 238,306 |
| OSAadjBMI - Meta | rs79950770 | 1 | 119539797 | 5 | Sofer et al 2023 | *TBX15* | T | C | 0.144 | 1.046 | 6.10E-09 | Y | T | C | 0.107 | 0.047 | 0.014 | **9.49E-04** | 267,661 | 0.112 | 0.048 | 0.014 | **8.80E-04** | 238,306 |
| OSAadjBMI - MVP only | rs72618639 | 2 | 50795733 | 9 | Sofer et al 2023 | *NRXN1* | G | C | 0.020 | 0.882 | 5.00E-09 | N | C | G | 0.991 | 0.040 | 0.073 | 5.78E-01 | 188,503 | NA | NA | NA | NA | NA |
| OSAadjBMI | rs76229479 | 2 | 103123912 | 11 | Strausz et al 2024 | *SLC9A4* | C | A | 0.100 | NR | 4.97E-10 | N | A | C | 0.879 | 0.017 | 0.013 | 2.08E-01 | 267,543 | 0.878 | 0.022 | 0.014 | 1.27E-01 | 238,280 |
| OSAadjBMI - Meta | rs181355045 | 2 | 156998417 | 15 | Sofer et al 2023 | *LINC01876* | A | C | 0.146 | 1.041 | 4.20E-08 | N* | A | C | 0.109 | 0.037 | 0.019 | **4.93E-02** | 117,268 | 0.114 | 0.044 | 0.020 | **2.60E-02** | 100,712 |
| OSAadjBMI - Meta | rs4681356 | 3 | 147082740 | 18 | Sofer et al 2023 | *AC092957.1* | A | G | 0.313 | 1.029 | 4.30E-08 | N | A | G | 0.312 | 0.000 | 0.009 | 9.95E-01 | 269,597 | 0.320 | 0.006 | 0.010 | 5.43E-01 | 240,206 |
| OSAadjBMI - MVP only | rs34179442 | 8 | 50900444 | 29 | Sofer et al 2023 | *SNTG1* | A | G | 0.470 | 0.966 | 3.70E-08 | N | A | G | 0.553 | -0.017 | 0.009 | 5.32E-02 | 265,819 | 0.582 | -0.017 | 0.009 | 7.41E-02 | 236,601 |
| OSAadjBMI - Meta | rs34179442 | 8 | 50900444 | 29 | Sofer et al 2023 | *SNTG1* | A | G | 0.455 | 1.033 | 1.30E-09 | N | A | G | 0.553 | -0.017 | 0.009 | 5.32E-02 | 265,819 | 0.582 | -0.017 | 0.009 | 7.41E-02 | 236,601 |
| OSAadjBMI - Meta | rs2841329 | 9 | 128134332 | 32 | Sofer et al 2023 | *GAPVD1* | G | A | 0.455 | 1.029 | 2.50E-09 | N | A | G | 0.500 | -0.014 | 0.009 | 1.07E-01 | 269,620 | 0.475 | -0.013 | 0.009 | 1.42E-01 | 240,229 |
| OSAadjBMI - Meta | rs7120537 | 11 | 28399637 | 39 | Sofer et al 2023 | *METTL15* | A | T | 0.489 | 0.970 | 3.20E-10 | N* | A | T | 0.489 | -0.034 | 0.012 | **5.66E-03** | 120,690 | 0.510 | -0.031 | 0.013 | **1.48E-02** | 104,016 |
| OSAadjBMI - MVP only | rs2277339 | 12 | 57146069 | 41 | Sofer et al 2023 | *PRIM1* | G | T | 0.120 | 0.956 | 1.40E-08 | N | T | G | 0.891 | 0.025 | 0.014 | 7.01E-02 | 269,620 | 0.899 | 0.026 | 0.015 | 8.54E-02 | 240,229 |
| OSAadjBMI - Meta | rs2277339 | 12 | 57146069 | 41 | Sofer et al 2023 | *PRIM1* | G | T | 0.156 | 0.954 | 1.30E-11 | N | T | G | 0.891 | 0.025 | 0.014 | 7.01E-02 | 269,620 | 0.899 | 0.026 | 0.015 | 8.54E-02 | 240,229 |
| OSAadjBMI - Meta | rs1826663 | 12 | 65797709 | 42 | Sofer et al 2023 | *MSRB3* | C | T | 0.352 | 1.029 | 6.40E-09 | Y | T | C | 0.618 | -0.054 | 0.009 | **8.51E-10** | 269,620 | 0.619 | -0.055 | 0.009 | **3.98E-09** | 240,229 |
| OSAadjBMI | rs10507084 | 12 | 97753152 | 43 | Strausz et al 2021 | *RMST/NEDD1* | T | C | 0.180 | 1.120 | 9.70E-10 | N | T | C | 0.080 | -0.007 | 0.017 | 6.63E-01 | 269,620 | 0.063 | -0.011 | 0.019 | 5.58E-01 | 240,229 |
| OSAadjBMI - MVP only | rs592333 | 13 | 51340315 | 45 | Sofer et al 2023 | *DLEU1* | G | A | 0.460 | 1.033 | 4.70E-08 | Y | A | G | 0.542 | -0.030 | 0.009 | **6.55E-04** | 269,620 | 0.548 | -0.031 | 0.009 | **9.78E-04** | 240,229 |

| Published Trait | dbSNP ID | CHR | POS (hg19) | Locus # | Publication | Reported Gene | Published Results | | | | | Replicated | Current Discovery Results - Stage 1 + Stage 2 | | | | | | | | | | | |
|---|---|---|---|---|---|---|---|---|---|---|---|---|---|---|---|---|---|---|---|---|---|---|---|---|
| | | | | | | | | | | | | | | | Multi-Population | | | | | EUR only | | | | |
| | | | | | | | A1 | A2 | EAF | OR | P | (Y/N*/N) | A1 | A2 | EAF | BETA | SE | P | N | EAF | BETA | SE | P | N |
| OSAadjBMI - Meta | rs592333 | 13 | 51340315 | 45 | Sofer et al 2023 | *DLEU1* | A | G | 0.513 | 0.970 | 1.20E-09 | Y | A | G | 0.542 | -0.030 | 0.009 | **6.55E-04** | 269,620 | 0.548 | -0.031 | 0.009 | **9.78E-04** | 240,229 |
| OSAadjBMI - Meta | rs1373285 | 13 | 54048078 | 46 | Sofer et al 2023 | *AL356295.1* | A | G | 0.310 | 1.031 | 1.60E-08 | Y | A | G | 0.322 | 0.024 | 0.009 | **1.19E-02** | 269,620 | 0.338 | 0.029 | 0.010 | **2.35E-03** | 240,229 |
| OSAadjBMI - Meta | rs2454451 | 15 | 76861461 | 48 | Sofer et al 2023 | *SCAPER* | T | G | 0.660 | 1.029 | 7.20E-09 | N | T | G | 0.681 | 0.006 | 0.009 | 5.53E-01 | 269,620 | 0.694 | 0.009 | 0.010 | 3.70E-01 | 240,229 |
| OSAadjBMI - MVP only | rs4886823 | 15 | 76965402 | 48 | Sofer et al 2023 | *SCAPER* | G | A | 0.270 | 0.964 | 1.30E-09 | N | A | G | 0.735 | 0.010 | 0.010 | 3.21E-01 | 269,620 | 0.741 | 0.008 | 0.010 | 4.15E-01 | 240,229 |

**Supplementary Table 11.** Lookup of previously-published AHI or avgSAO2 SNPs in discovery analysis.

All marginally significant associations (P<0.05) are highlighted in red.

| | | | | | | | Published Results | | | | | | | | OSA - Pooled Model 1 | | | | | OSA - Pooled Model 2 | | | | | OSA - EUR Model 1 | | | | | OSA - EUR Model 2 | | | | | AHI - Pooled Model 1 | | | |
|---|---|---|---|---|---|---|---|---|---|---|---|---|---|---|---|---|---|---|---|---|---|---|---|---|---|---|---|---|---|---|---|---|---|---|---|---|---|---|
| Published Trait | dbSNP ID | CHR | POS | Locus | cpaid | Reported Gene | A1 | A2 | BETA | SE | P | N | A1 | A2 | EAF | BETA | SE | P | N | EAF | BETA | SE | P | N | EAF | BETA | SE | P | N | EAF | BETA | SE | P | N | EAF | BETA | SE | P |
| AHI | rs711822 | 2 | 176989023 | 1 | 2:176989023:C_T | *HOXD9* | C | T | -0.069 | 0.007 | **3.00E-08** | 47,867 | T | C | 0.679 | -0.008 | 0.010 | 4.06E-01 | 202,804 | 0.678 | -0.008 | 0.011 | 4.44E-01 | 180,529 | 0.668 | -0.008 | 0.011 | 4.47E-01 | 163,647 | 0.668 | -0.008 | 0.011 | 4.87E-01 | 151,211 | 0.641 | 0.003 | 0.012 | 7.88E-01 |
| AHI | rs75546795 | 8 | 31415401 | 2 | 8:31415401:C_T | *NRG1* | C | T | 0.132 | 0.012 | **9.40E-10** | 47,640 | T | C | 0.082 | 0.012 | 0.017 | 4.85E-01 | 202,804 | 0.082 | 0.012 | 0.018 | 5.20E-01 | 180,957 | 0.089 | 0.010 | 0.017 | 5.76E-01 | 164,027 | 0.089 | 0.011 | 0.018 | 5.40E-01 | 151,590 | 0.083 | -0.004 | 0.022 | 8.62E-01 |
| avgSaO$_2$ | rs113419680 | 1 | 5429343 | 3 | 1:5429343:C_G | *KCNAB2* | C | G | 0.236 | 0.045 | 1.90E-07 | 12,522 | C | G | 0.016 | -0.070 | 0.058 | 2.26E-01 | 121,328 | 0.015 | -0.048 | 0.068 | 4.79E-01 | 100,548 | - | - | - | - | - | - | - | - | - | - | - | - | - | - |
| avgSaO$_2$ | rs7537978 | 1 | 65510984 | 4 | 1:65510984:C_T | *JAK1* | C | T | 0.065 | 0.013 | 7.40E-07 | 21,127 | T | C | 0.817 | 0.030 | 0.010 | **4.01E-03** | 492,107 | 0.824 | 0.029 | 0.012 | **1.31E-02** | 264,942 | 0.836 | 0.039 | 0.011 | **6.22E-04** | 449,625 | 0.837 | 0.035 | 0.012 | **4.51E-03** | 235,872 | 0.823 | 0.023 | 0.012 | **4.98E-02** |
| avgSaO$_2$ | rs78728490 | 1 | 84495858 | 5 | 1:84495858:A_G | *TTLL7* | A | G | 0.310 | 0.067 | 3.90E-06 | 11,891 | A | G | 0.011 | -0.008 | 0.066 | 9.07E-01 | 121,328 | 0.011 | -0.019 | 0.079 | 8.08E-01 | 100,548 | - | - | - | - | - | - | - | - | - | - | 0.025 | -0.150 | 0.085 | 7.62E-02 |
| avgSaO$_2$ | rs116133558 | 1 | 203716843 | 6 | 1:203716843:C_T | *LAX1* | T | C | 0.268 | 0.052 | 2.40E-07 | 13,017 | T | C | 0.018 | 0.007 | 0.051 | 8.87E-01 | 121,328 | 0.018 | 0.008 | 0.061 | 9.02E-01 | 100,548 | - | - | - | - | - | - | - | - | - | - | 0.039 | -0.021 | 0.067 | 7.60E-01 |
| avgSaO$_2$ | rs12071201 | 1 | 233950041 | 7 | 1:233950041:C_T | *SLC35F3* | C | T | -0.242 | 0.053 | 4.50E-06 | 16,064 | T | C | 0.985 | -0.030 | 0.054 | 5.78E-01 | 121,328 | 0.986 | -0.026 | 0.063 | 6.76E-01 | 100,548 | - | - | - | - | - | - | - | - | - | - | 0.976 | -0.029 | 0.067 | 6.64E-01 |
| avgSaO$_2$ | rs1208909497 | 1 | 240962587 | 8 | 1:240962587:T_TTGTG | *RGS7* | T | TTGTG | -0.171 | 0.036 | 2.40E-06 | 2,634 | T | TTGTG | - | - | - | - | - | - | - | - | - | - | - | - | - | - | - | - | - | - | - | - | 0.948 | 0.050 | 0.041 | 2.19E-01 |
| avgSaO$_2$ | rs6758412 | 2 | 30163863 | 9 | 2:30163863:A_G | *ALK* | G | A | 0.309 | 0.062 | 6.60E-07 | 1,852 | A | G | 0.987 | -0.119 | 0.084 | 1.55E-01 | 80,156 | 0.991 | -0.032 | 0.107 | 7.67E-01 | 60,975 | - | - | - | - | - | - | - | - | - | - | 0.990 | 0.114 | 0.093 | 2.16E-01 |
| avgSaO$_2$ | rs756933193 | 2 | 142701245 | 10 | 2:142701245:A_AT | *LRP1B* | A | AT | -0.224 | 0.047 | 2.40E-06 | 20,683 | A | AT | 0.021 | -0.073 | 0.067 | 2.77E-01 | 41,172 | 0.021 | -0.098 | 0.077 | 2.07E-01 | 39,573 | 0.010 | -0.290 | 0.142 | **4.06E-02** | 26,897 | 0.010 | -0.351 | 0.160 | **2.85E-02** | 25,905 | 0.037 | -0.055 | 0.098 | 5.72E-01 |
| avgSaO$_2$ | rs116754339 | 3 | 22811163 | 11 | 3:22811163:C_T | *UBE2E2* | T | C | -0.240 | 0.052 | 4.60E-06 | 13,421 | T | C | 0.015 | 0.006 | 0.055 | 9.13E-01 | 121,328 | 0.014 | 0.002 | 0.064 | 9.81E-01 | 100,548 | - | - | - | - | - | - | - | - | - | - | 0.026 | -0.116 | 0.070 | 1.00E-01 |
| avgSaO$_2$ | rs6853180 | 4 | 136754442 | 12 | 4:136754442:A_G | *PABPC4L* | A | G | -0.060 | 0.013 | 4.00E-06 | 21,126 | A | G | 0.739 | 0.005 | 0.009 | 6.24E-01 | 492,107 | 0.745 | 0.002 | 0.010 | 8.70E-01 | 269,610 | 0.778 | 0.000 | 0.010 | 9.94E-01 | 454,012 | 0.779 | -0.003 | 0.011 | 8.11E-01 | 240,222 | 0.693 | 0.019 | 0.010 | 5.42E-02 |
| avgSaO$_2$ | rs28491289 | 4 | 139967889 | 13 | 4:139967889:A_C | *CCRN4L* | C | A | 0.132 | 0.028 | 3.50E-06 | 21,126 | A | C | 0.947 | -0.017 | 0.030 | 5.66E-01 | 121,328 | 0.950 | 0.007 | 0.035 | 8.37E-01 | 100,548 | 0.980 | 0.063 | 0.039 | 1.03E-01 | 158,525 | 0.980 | 0.041 | 0.042 | 3.22E-01 | 146,101 | 0.941 | -0.041 | 0.029 | 1.62E-01 |
| avgSaO$_2$ | rs140597088 | 5 | 88675264 | 14 | 5:88675264:A_G | *MEF2C* | A | G | 1.299 | 0.269 | 1.30E-06 | 3,516 | A | G | - | - | - | - | - | - | - | - | - | - | 0.005 | 0.073 | 0.082 | 3.72E-01 | 375,611 | 0.005 | 0.132 | 0.091 | 1.48E-01 | 162,118 | 0.006 | 0.093 | 0.082 | 2.58E-01 |
| avgSaO$_2$ | rs73718934 | 6 | 3028790 | 15 | 6:3028790:C_T | *NQO2* | T | C | 0.107 | 0.023 | 4.00E-06 | 20,898 | T | C | 0.076 | 0.022 | 0.015 | 1.49E-01 | 492,107 | 0.073 | 0.039 | 0.017 | **1.81E-02** | 268,196 | 0.071 | 0.018 | 0.016 | 2.71E-01 | 452,675 | 0.071 | 0.032 | 0.018 | 6.96E-02 | 238,889 | 0.079 | 0.004 | 0.017 | 8.35E-01 |
| avgSaO$_2$ | rs191894471 | 6 | 13428339 | 16 | 6:13428339:A_G | *GFOD1* | G | A | 1.429 | 0.293 | 1.10E-06 | 3,606 | A | G | - | - | - | - | - | - | - | - | - | - | - | - | - | - | - | - | - | - | - | - | - | - | - | - |
| avgSaO$_2$ | rs3734635 | 6 | 126102486 | 17 | 6:126102486:C_G | *NCOA7* | G | C | 0.130 | 0.028 | 4.50E-06 | 15,801 | C | G | 0.939 | 0.022 | 0.029 | 4.57E-01 | 149,739 | 0.942 | 0.021 | 0.034 | 5.31E-01 | 128,183 | 0.994 | -0.091 | 0.096 | 3.45E-01 | 86,308 | 0.994 | -0.045 | 0.109 | 6.79E-01 | 74,163 | 0.912 | 0.033 | 0.026 | 2.06E-01 |
| avgSaO$_2$ | rs76669606 | 6 | 156889525 | 18 | 6:156889525:G_GC | *ARID1B* | GC | G | 0.046 | 0.010 | 3.30E-06 | 21,089 | G | GC | 0.584 | -0.008 | 0.013 | 5.41E-01 | 81,476 | 0.586 | -0.003 | 0.014 | 8.56E-01 | 76,517 | 0.581 | -0.014 | 0.014 | 3.10E-01 | 74,002 | 0.581 | -0.006 | 0.014 | 6.57E-01 | 73,725 | 0.599 | -0.008 | 0.014 | 5.60E-01 |
| avgSaO$_2$ | rs6948904 | 7 | 11496661 | 19 | 7:11496661:A_G | *THSD7A* | A | G | -0.150 | 0.033 | 4.60E-06 | 14,481 | A | G | 0.027 | -0.045 | 0.043 | 2.98E-01 | 121,328 | 0.026 | -0.056 | 0.052 | 2.80E-01 | 100,548 | - | - | - | - | - | - | - | - | - | - | 0.041 | -0.024 | 0.051 | 6.42E-01 |
| avgSaO$_2$ | rs80108997 | 7 | 32268952 | 20 | 7:32268952:A_C | *PDE1C* | A | C | 0.224 | 0.048 | 2.80E-06 | 13,056 | A | C | 0.012 | 0.016 | 0.061 | 7.99E-01 | 121,328 | 0.011 | 0.056 | 0.072 | 4.42E-01 | 100,548 | - | - | - | - | - | - | - | - | - | - | 0.021 | 0.073 | 0.082 | 3.72E-01 |
| avgSaO$_2$ | rs187178912 | 7 | 33406097 | 21 | 7:33406097:A_G | *BBS9* | A | G | 0.859 | 0.188 | 4.80E-06 | 5,591 | A | G | - | - | - | - | - | - | - | - | - | - | 0.005 | -0.068 | 0.082 | 4.04E-01 | 375,611 | 0.005 | -0.084 | 0.093 | 3.65E-01 | 162,118 | - | - | - | - |
| avgSaO$_2$ | rs112771667 | 8 | 10209611 | 22 | 8:10209611:C_T | *MSRA* | T | C | 0.882 | 0.178 | 7.40E-07 | 8,009 | T | C | - | - | - | - | - | - | - | - | - | - | - | - | - | - | - | - | - | - | - | - | - | - | - | - |
| avgSaO$_2$ | rs11166851 | 8 | 139876084 | 23 | 8:139876084:C_T | *COL22A1* | C | T | -0.063 | 0.014 | 3.90E-06 | 20,130 | T | C | 0.150 | -0.005 | 0.011 | 6.42E-01 | 492,107 | 0.139 | -0.002 | 0.013 | 8.79E-01 | 259,215 | 0.134 | -0.006 | 0.012 | 6.09E-01 | 444,155 | 0.133 | 0.000 | 0.014 | 9.98E-01 | 230,412 | 0.150 | -0.015 | 0.013 | 2.42E-01 |
| avgSaO$_2$ | rs7041166 | 9 | 8392668 | 24 | 9:8392668:A_G | *PTPRD* | G | A | 0.085 | 0.018 | 3.50E-06 | 21,127 | A | G | 0.890 | 0.013 | 0.013 | 3.04E-01 | 492,107 | 0.891 | 0.013 | 0.014 | 3.50E-01 | 269,620 | 0.895 | 0.018 | 0.014 | 1.77E-01 | 454,019 | 0.895 | 0.017 | 0.015 | 2.42E-01 | 240,229 | 0.893 | 0.034 | 0.014 | **1.73E-02** |
| avgSaO$_2$ | rs550460908 | 9 | 90246904 | 25 | 9:90246904:A_G | *CTSL* | G | A | -0.394 | 0.086 | 4.20E-06 | 15,184 | A | G | 0.993 | -0.010 | 0.060 | 8.68E-01 | 410,631 | 0.993 | 0.001 | 0.068 | 9.91E-01 | 188,503 | 0.992 | -0.037 | 0.064 | 5.59E-01 | 375,611 | 0.992 | -0.023 | 0.072 | 7.46E-01 | 162,118 | 0.994 | 0.006 | 0.061 | 9.25E-01 |
| avgSaO$_2$ | rs199819010 | 9 | 113470288 | 26 | 9:113470288:G_GA | *MUSK* | G | GA | -1.287 | 0.281 | 4.60E-06 | 3,818 | G | GA | - | - | - | - | - | - | - | - | - | - | - | - | - | - | - | - | - | - | - | - | - | - | - | - |
| avgSaO$_2$ | rs2812667 | 10 | 34026997 | 27 | 10:34026997:A_G | *NRP1* | A | G | -0.226 | 0.050 | 5.00E-06 | 20,681 | A | G | 0.022 | 0.022 | 0.027 | 4.08E-01 | 492,107 | 0.023 | 0.014 | 0.030 | 6.30E-01 | 269,620 | 0.024 | 0.024 | 0.028 | 3.97E-01 | 454,019 | 0.024 | 0.021 | 0.030 | 4.86E-01 | 240,229 | 0.024 | -0.044 | 0.031 | 1.61E-01 |
| avgSaO$_2$ | rs771251835 | 10 | 71104588 | 28 | 10:71104588:C_CCT | *HK1* | CCT | C | -0.069 | 0.014 | 1.00E-06 | 20,861 | CCT | C | 0.858 | 0.019 | 0.016 | 2.39E-01 | 122,648 | 0.858 | 0.023 | 0.017 | 1.82E-01 | 120,690 | 0.859 | 0.021 | 0.017 | 2.15E-01 | 105,305 | 0.859 | 0.025 | 0.018 | 1.72E-01 | 104,016 | 0.858 | -0.021 | 0.019 | 2.72E-01 |
| avgSaO$_2$ | rs4939452 | 11 | 60537578 | 29 | 11:60537578:C_T | *MS4A15* | C | T | 0.064 | 0.012 | **4.90E-08** | 21,126 | T | C | 0.492 | 0.014 | 0.010 | 1.56E-01 | 410,631 | 0.491 | 0.011 | 0.011 | 3.23E-01 | 188,503 | 0.480 | 0.003 | 0.009 | 6.94E-01 | 454,019 | 0.481 | 0.003 | 0.009 | 7.63E-01 | 240,229 | 0.481 | -0.018 | 0.010 | 6.20E-02 |
| avgSaO$_2$ | rs142696393 | 11 | 71388551 | 30 | 11:71388551:A_G | *AP000867.1* | G | A | 0.258 | 0.055 | 2.80E-06 | 4,500 | A | G | 0.952 | -0.061 | 0.035 | 8.49E-02 | 121,328 | 0.953 | -0.038 | 0.042 | 3.66E-01 | 100,548 | - | - | - | - | - | - | - | - | - | - | 0.901 | -0.002 | 0.044 | 9.64E-01 |
| avgSaO$_2$ | rs114572017 | 11 | 89263553 | 31 | 11:89263553:A_T | *NOX4* | A | T | 0.663 | 0.138 | 1.40E-06 | 4,500 | A | T | 0.006 | 0.002 | 0.088 | 9.79E-01 | 121,328 | 0.006 | 0.043 | 0.105 | 6.80E-01 | 100,548 | - | - | - | - | - | - | - | - | - | - | 0.010 | -0.198 | 0.108 | 6.72E-02 |
| avgSaO$_2$ | rs138247706 | 12 | 17150092 | 32 | 12:17150092:C_T | *LMO3* | T | C | -0.225 | 0.048 | 3.10E-06 | 19,201 | T | C | 0.015 | 0.025 | 0.043 | 5.67E-01 | 410,631 | 0.015 | 0.009 | 0.049 | 8.53E-01 | 188,503 | 0.017 | -0.006 | 0.038 | 8.82E-01 | 454,019 | 0.017 | -0.023 | 0.042 | 5.81E-01 | 240,229 | 0.014 | -0.020 | 0.048 | 6.77E-01 |
| avgSaO$_2$ | rs529918807 | 12 | 44520960 | 33 | 12:44520960:A_T | *TMEM117* | T | A | -0.621 | 0.123 | 4.60E-07 | 7,820 | A | T | 0.991 | 0.089 | 0.115 | 4.37E-01 | 41,172 | 0.991 | 0.058 | 0.127 | 6.50E-01 | 39,573 | 0.989 | -0.013 | 0.123 | 9.17E-01 | 26,897 | 0.989 | -0.035 | 0.138 | 8.02E-01 | 25,905 | 0.993 | -0.044 | 0.127 | 7.33E-01 |
| avgSaO$_2$ | rs2732462 | 12 | 48696384 | 34 | 12:48696384:A_G | *H1FNT* | G | A | 0.069 | 0.013 | 1.70E-07 | 21,127 | A | G | 0.659 | 0.023 | 0.008 | **5.71E-03** | 492,107 | 0.657 | 0.028 | 0.009 | **2.67E-03** | 269,620 | 0.641 | 0.024 | 0.009 | **6.15E-03** | 454,019 | 0.642 | 0.028 | 0.010 | **3.25E-03** | 240,229 | 0.650 | 0.000 | 0.010 | 9.96E-01 |
| avgSaO$_2$ | rs77913955 | 12 | 64425733 | 35 | 12:64425733:G_T | *SRGAP1* | T | G | -0.276 | 0.060 | 4.30E-06 | 12,235 | T | G | 0.013 | -0.012 | 0.060 | 8.47E-01 | 121,328 | 0.013 | -0.053 | 0.073 | 4.72E-01 | 100,548 | NA | NA | NA | NA | NA | NA | NA | NA | NA | NA | 0.025 | -0.111 | 0.075 | 1.41E-01 |
| avgSaO$_2$ | rs117349588 | 13 | 37246102 | 36 | 13:37246102:C_T | *SERTM1* | T | C | -2.128 | 0.447 | 2.00E-06 | 1,614 | T | C | - | - | - | - | - | - | - | - | - | - | - | - | - | - | - | - | - | - | - | - | - | - | - | - |
| avgSaO$_2$ | rs565226335 | 13 | 107701363 | 37 | 13:107701363:A_G | *ARGLU1* | G | A | 0.722 | 0.152 | 2.20E-06 | 7,666 | A | G | 0.988 | 0.060 | 0.051 | 2.39E-01 | 410,631 | 0.988 | 0.097 | 0.058 | 9.65E-02 | 188,503 | 0.987 | 0.042 | 0.052 | 4.21E-01 | 375,611 | 0.987 | 0.078 | 0.059 | 1.87E-01 | 162,118 | 0.985 | 0.004 | 0.055 | 9.38E-01 |
| avgSaO$_2$ | rs79931065 | 14 | 105028164 | 38 | 14:105028164:G_T | *C14orf180* | T | G | 0.181 | 0.040 | 4.70E-06 | 13,017 | T | G | 0.028 | 0.049 | 0.043 | 2.56E-01 | 121,328 | 0.028 | 0.026 | 0.050 | 6.08E-01 | 100,548 | NA | - | - | - | - | - | - | - | - | - | 0.046 | -0.071 | 0.050 | 1.53E-01 |
| avgSaO$_2$ | rs35127183 | 15 | 85215739 | 39 | 15:85215739:A_G | *NMB* | A | G | -0.071 | 0.014 | 1.30E-07 | 21,127 | A | G | 0.326 | -0.005 | 0.012 | 6.77E-01 | 122,648 | 0.327 | 0.000 | 0.013 | 9.79E-01 | 120,690 | 0.344 | -0.012 | 0.012 | 3.56E-01 | 105,305 | 0.344 | -0.004 | 0.013 | 7.80E-01 | 104,016 | 0.318 | -0.004 | 0.014 | 7.49E-01 |
| avgSaO$_2$ | rs75660329 | 16 | 76304656 | 40 | 16:76304656:C_T | *CNTNAP4* | T | C | -0.413 | 0.087 | 2.10E-06 | 9,088 | T | C | 0.005 | 0.069 | 0.093 | 4.59E-01 | 121,328 | 0.005 | 0.130 | 0.109 | 2.33E-01 | 100,548 | - | - | - | - | - | - | - | - | - | - | 0.011 | 0.263 | 0.130 | **4.33E-02** |
| avgSaO$_2$ | rs866759288 | 16 | 87535488 | 41 | 16:87535488:C_T | *ZCCHC14* | C | T | -0.772 | 0.161 | 1.60E-06 | 4,578 | T | C | - | - | - | - | - | - | - | - | - | - | - | - | - | - | - | - | - | - | - | - | - | - | - | - |
| avgSaO$_2$ | rs73939879 | 18 | 9547785 | 42 | 18:9547785:A_T | *PPP4R1* | A | T | -0.746 | 0.146 | 3.10E-07 | 4,717 | A | T | 0.006 | -0.049 | 0.092 | 5.96E-01 | 121,328 | 0.005 | 0.048 | 0.109 | 6.61E-01 | 100,548 | - | - | - | - | - | - | - | - | - | - | 0.013 | 0.071 | 0.119 | 5.50E-01 |
| avgSaO$_2$ | rs187860354 | 18 | 39999243 | 43 | 18:39999243:A_G | *PIK3C3* | G | A | 0.436 | 0.081 | 7.00E-08 | 11,891 | A | G | 0.995 | -0.066 | 0.094 | 4.81E-01 | 121,328 | 0.995 | -0.072 | 0.112 | 5.20E-01 | 100,548 | - | - | - | - | - | - | - | - | - | - | 0.990 | -0.069 | 0.133 | 6.04E-01 |
| avgSaO$_2$ | rs187041178 | 18 | 40320288 | 43 | 18:40320288:C_T | *RIT2* | C | T | 0.379 | 0.079 | 1.60E-06 | 12,235 | T | C | 0.994 | 0.048 | 0.093 | 6.05E-01 | 121,328 | 0.994 | 0.132 | 0.111 | 2.32E-01 | 100,548 | - | - | - | - | - | - | - | - | - | - | 0.991 | 0.044 | 0.121 | 7.18E-01 |
| avgSaO$_2$ | rs868057231 | 18 | 42236396 | 44 | 18:42236396:C_T | *SETBP1* | C | T | 0.533 | 0.109 | 1.10E-06 | 4,500 | T | C | 0.987 | 0.073 | 0.080 | 3.64E-01 | 41,172 | 0.987 | 0.124 | 0.093 | 1.81E-01 | 39,573 | - | - | - | - | - | - | - | - | - | - | 0.974 | 0.301 | 0.114 | **8.04E-03** |
| avgSaO$_2$ | rs16955575 | 18 | 50346481 | 45 | 18:50346481:C_T | *DCC* | T | C | 0.086 | 0.018 | 2.30E-06 | 20,898 | T | C | 0.036 | -0.024 | 0.027 | 3.80E-01 | 410,631 | 0.036 | -0.017 | 0.031 | 5.93E-01 | 188,503 | 0.039 | -0.035 | 0.023 | 1.37E-01 | 454,019 | 0.039 | -0.027 | 0.026 | 2.96E-01 | 240,229 | 0.035 | 0.026 | 0.030 | 3.99E-01 |
| avgSaO$_2$ | rs113197061 | 19 | 7577232 | 46 | 19:7577232:G_T | *ZNF358* | T | G | 0.792 | 0.164 | 1.40E-06 | 4,717 | T | G | 0.005 | -0.238 | 0.130 | 6.63E-02 | 41,172 | 0.005 | -0.134 | 0.148 | 3.65E-01 | 39,573 | - | - | - | - | - | - | - | - | - | - | 0.008 | 0.050 | 0.158 | 7.51E-01 |
| avgSaO$_2$ | rs12609338 | 19 | 57833835 | 47 | 19:57833835:A_G | *ZNF543* | G | A | 0.049 | 0.010 | 4.50E-07 | 21,089 | A | G | 0.679 | -0.003 | 0.008 | 7.40E-01 | 492,107 | 0.678 | 0.007 | 0.009 | 4.78E-01 | 269,620 | 0.674 | -0.001 | 0.009 | 9.07E-01 | 454,019 | 0.674 | 0.008 | 0.010 | 4.33E-01 | 240,229 | 0.666 | -0.009 | 0.009 | 3.13E-01 |
| avgSaO$_2$ | rs76190072 | 19 | 57856774 | 47 | 19:57856774:A_C | *ZNF304* | C | A | 0.391 | 0.081 | 1.50E-06 | 11,891 | A | C | 0.994 | 0.002 | 0.087 | 9.80E-01 | 121,328 | 0.995 | 0.123 | 0.106 | 2.45E-01 | 100,548 | - | - | - | - | - | - | - | - | - | - | 0.987 | 0.165 | 0.115 | 1.52E-01 |
| avgSaO$_2$ | rs233266 | 21 | 46037932 | 48 | 21:46037932:C_G | *KRTAP10-8* | C | G | -0.178 | 0.038 | 3.20E-06 | 20,898 | C | G | 0.042 | -0.015 | 0.028 | 5.93E-01 | 410,631 | 0.042 | -0.013 | 0.031 | 6.80E-01 | 188,503 | 0.045 | -0.016 | 0.029 | 5.74E-01 | 375,611 | 0.045 | -0.016 | 0.032 | 6.14E-01 | 162,118 | 0.047 | -0.083 | 0.033 | **1.31E-02** |
| avgSaO$_2$ | rs57204056 | 22 | 48612376 | 49 | 22:48612376:C_G | *FAM19A5* | C | G | -0.301 | 0.064 | 3.00E-06 | 12,673 | C | G | 0.013 | 0.007 | 0.060 | 9.11E-01 | 121,328 | 0.013 | 0.023 | 0.071 | 7.47E-01 | 100,548 | - | - | - | - | - | - | - | - | - | - | - | - | - | - |

**Supplementary Table 11.** Lookup of pre

All marginally significant associations (F

#### Current Discovery Results - Stage 1 + Stage 2

| | | | AHI - Pooled Model 2 | | | | | AHI - EUR Model 1 | | | | | AHI - EUR Model 2 | | | | | avgSaO2 - Pooled Model 1 | | | | | avgSaO2 - Pooled Model 2 | | | | | avgSaO2 - EUR Model 1 | | | | | avgSaO2 - EUR Model 2 | | | | |
|---|---|---|---|---|---|---|---|---|---|---|---|---|---|---|---|---|---|---|---|---|---|---|---|---|---|---|---|---|---|---|---|---|---|---|---|---|---|
| **Published Trait** | **dbSNP ID** | **N** | **EAF** | **BETA** | **SE** | **P** | **N** | **EAF** | **BETA** | **SE** | **P** | **N** | **EAF** | **BETA** | **SE** | **P** | **N** | **EAF** | **BETA** | **SE** | **P** | **N** | **EAF** | **BETA** | **SE** | **P** | **N** | **EAF** | **BETA** | **SE** | **P** | **N** | **EAF** | **BETA** | **SE** | **P** | **N** |
| AHI | rs711822 | 17,058 | 0.637 | 0.003 | 0.012 | 7.91E-01 | 15,626 | 0.672 | 0.008 | 0.014 | 5.37E-01 | 12,340 | 0.671 | 0.007 | 0.014 | 6.15E-01 | 11,003 | 0.681 | -0.024 | 0.018 | 1.87E-01 | 6,908 | 0.682 | -0.030 | 0.019 | 1.11E-01 | 6,713 | 0.669 | -0.026 | 0.020 | 1.89E-01 | 5,872 | 0.670 | -0.030 | 0.020 | 1.32E-01 | 5,705 |
| AHI | rs75546795 | 14,354 | 0.083 | -0.008 | 0.023 | 7.36E-01 | 12,923 | 0.090 | -0.014 | 0.022 | 5.34E-01 | 12,340 | 0.091 | -0.016 | 0.024 | 4.93E-01 | 11,003 | 0.085 | 0.010 | 0.031 | 7.50E-01 | 6,908 | 0.085 | 0.012 | 0.032 | 6.98E-01 | 6,713 | 0.091 | 0.021 | 0.032 | 5.14E-01 | 5,872 | 0.091 | 0.026 | 0.033 | 4.35E-01 | 5,705 |
| avgSaO$_2$ | rs113419680 | - | 0.028 | -0.041 | 0.076 | 5.90E-01 | 5,978 | - | - | - | - | - | - | - | - | - | - | 0.039 | 0.128 | 0.116 | 2.69E-01 | 1,303 | 0.040 | 0.108 | 0.117 | 3.54E-01 | 1,258 | - | - | - | - | - | - | - | - | - | - |
| avgSaO$_2$ | rs7537978 | 28,486 | 0.824 | 0.024 | 0.013 | 5.22E-02 | 24,511 | 0.831 | 0.019 | 0.013 | 1.38E-01 | 23,768 | 0.832 | 0.022 | 0.014 | 1.16E-01 | 19,888 | 0.812 | -0.031 | 0.023 | 1.66E-01 | 6,908 | 0.813 | -0.017 | 0.023 | 4.67E-01 | 6,713 | 0.839 | -0.039 | 0.026 | 1.32E-01 | 5,872 | 0.840 | -0.020 | 0.026 | 4.47E-01 | 5,705 |
| avgSaO$_2$ | rs78728490 | 7,320 | 0.025 | -0.190 | 0.085 | **2.56E-02** | 7,096 | - | - | - | - | - | - | - | - | - | - | 0.032 | -0.127 | 0.120 | 2.90E-01 | 2,150 | 0.033 | -0.078 | 0.122 | 5.21E-01 | 2,105 | - | - | - | - | - | - | - | - | - | - |
| avgSaO$_2$ | rs116133558 | 7,320 | 0.038 | -0.008 | 0.069 | 9.13E-01 | 7,096 | - | - | - | - | - | - | - | - | - | - | 0.051 | 0.120 | 0.096 | 2.09E-01 | 2,150 | 0.051 | 0.088 | 0.098 | 3.74E-01 | 2,105 | - | - | - | - | - | - | - | - | - | - |
| avgSaO$_2$ | rs12071201 | 7,320 | 0.976 | -0.003 | 0.067 | 9.63E-01 | 7,096 | - | - | - | - | - | - | - | - | - | - | 0.970 | 0.145 | 0.098 | 1.38E-01 | 3,481 | 0.970 | 0.064 | 0.099 | 5.17E-01 | 3,320 | - | - | - | - | - | - | - | - | - | - |
| avgSaO$_2$ | rs1208909497 | 8,625 | 0.947 | 0.046 | 0.041 | 2.69E-01 | 7,875 | - | - | - | - | - | - | - | - | - | - | 0.984 | -0.033 | 0.099 | 7.39E-01 | 2,414 | 0.984 | -0.048 | 0.097 | 6.18E-01 | 2,389 | - | - | - | - | - | - | - | - | - | - |
| avgSaO$_2$ | rs6758412 | 11,661 | - | - | - | - | - | - | - | - | - | - | - | - | - | - | - | - | - | - | - | - | - | - | - | - | - | - | - | - | - | - | - | - | - | - | - |
| avgSaO$_2$ | rs756933193 | 1,557 | 0.038 | 0.017 | 0.099 | 8.67E-01 | 1,505 | 0.005 | 0.190 | 0.480 | 6.92E-01 | 573 | - | - | - | - | - | 0.036 | -0.182 | 0.117 | 1.19E-01 | 1,303 | 0.036 | -0.248 | 0.118 | **3.59E-02** | 1,258 | - | - | - | - | - | - | - | - | - | - |
| avgSaO$_2$ | rs116754339 | 7,320 | 0.026 | -0.123 | 0.071 | 8.26E-02 | 7,096 | NA | - | - | - | - | - | - | - | - | - | 0.035 | 0.070 | 0.105 | 5.07E-01 | 2,634 | 0.038 | 0.109 | 0.110 | 3.22E-01 | 1,258 | - | - | - | - | - | - | - | - | - | - |
| avgSaO$_2$ | rs6853180 | 28,486 | 0.684 | 0.016 | 0.010 | 1.31E-01 | 24,511 | 0.775 | 0.017 | 0.011 | 1.29E-01 | 23,768 | 0.776 | 0.014 | 0.012 | 2.57E-01 | 19,888 | 0.695 | -0.043 | 0.019 | **2.56E-02** | 6,908 | 0.696 | -0.036 | 0.019 | 6.21E-02 | 6,713 | 0.772 | -0.051 | 0.022 | **2.27E-02** | 5,872 | 0.772 | -0.041 | 0.023 | 7.38E-02 | 5,705 |
| avgSaO$_2$ | rs28491289 | 17,053 | 0.941 | -0.051 | 0.030 | 8.67E-02 | 15,624 | 0.971 | -0.048 | 0.047 | 3.15E-01 | 12,340 | 0.972 | -0.098 | 0.050 | **4.98E-02** | 11,003 | 0.927 | 0.022 | 0.046 | 6.28E-01 | 6,906 | 0.928 | 0.030 | 0.047 | 5.16E-01 | 6,711 | 0.970 | -0.073 | 0.070 | 2.95E-01 | 5,872 | 0.970 | -0.074 | 0.071 | 2.93E-01 | 5,705 |
| avgSaO$_2$ | rs140597088 | 18,748 | 0.006 | 0.028 | 0.086 | 7.45E-01 | 15,981 | 0.007 | 0.058 | 0.070 | 4.08E-01 | 23,768 | 0.007 | 0.030 | 0.075 | 6.90E-01 | 19,888 | 0.009 | -0.141 | 0.165 | 3.93E-01 | 3,481 | 0.009 | -0.151 | 0.167 | 3.68E-01 | 3,320 | 0.009 | -0.043 | 0.139 | 7.56E-01 | 5,389 | 0.010 | -0.125 | 0.140 | 3.71E-01 | 5,241 |
| avgSaO$_2$ | rs73718934 | 25,782 | 0.079 | 0.002 | 0.018 | 9.27E-01 | 21,808 | 0.077 | 0.010 | 0.018 | 5.80E-01 | 23,768 | 0.077 | 0.005 | 0.020 | 7.97E-01 | 19,888 | - | - | - | - | - | 0.073 | 0.034 | 0.034 | 3.16E-01 | 6,713 | 0.067 | -0.009 | 0.038 | 8.05E-01 | 5,872 | 0.067 | 0.012 | 0.038 | 7.59E-01 | 5,705 |
| avgSaO$_2$ | rs191894471 | - | - | - | - | - | - | 0.995 | 0.312 | 0.119 | **8.82E-03** | 12,001 | 0.995 | 0.303 | 0.125 | **1.51E-02** | 9,433 | 0.994 | -0.049 | 0.245 | 8.41E-01 | 1,303 | 0.995 | 0.033 | 0.253 | 8.98E-01 | 1,258 | 0.989 | -0.177 | 0.306 | 5.62E-01 | 483 | 0.989 | -0.128 | 0.306 | 6.75E-01 | 464 |
| avgSaO$_2$ | rs3734635 | 17,058 | 0.912 | 0.043 | 0.027 | 1.08E-01 | 15,626 | 0.994 | 0.067 | 0.089 | 4.52E-01 | 12,340 | 0.994 | -0.022 | 0.093 | 8.15E-01 | 11,003 | 0.876 | 0.028 | 0.045 | 5.40E-01 | 6,908 | 0.876 | 0.033 | 0.046 | 4.69E-01 | 6,713 | 0.994 | -0.024 | 0.127 | 8.53E-01 | 5,872 | 0.994 | 0.039 | 0.129 | 7.63E-01 | 5,705 |
| avgSaO$_2$ | rs76669606 | 10,711 | 0.601 | -0.004 | 0.015 | 8.06E-01 | 9,504 | 0.583 | -0.008 | 0.015 | 6.10E-01 | 9,858 | 0.583 | -0.004 | 0.016 | 8.15E-01 | 8,709 | 0.582 | -0.014 | 0.022 | 5.20E-01 | 4,274 | 0.583 | -0.017 | 0.022 | 4.52E-01 | 4,240 | 0.576 | -0.010 | 0.023 | 6.60E-01 | 4,097 | 0.577 | -0.013 | 0.023 | 5.77E-01 | 4,063 |
| avgSaO$_2$ | rs6948904 | 13,231 | 0.041 | -0.044 | 0.052 | 3.96E-01 | 11,803 | - | - | - | - | - | - | - | - | - | - | 0.053 | 0.196 | 0.078 | **1.14E-02** | 4,728 | 0.054 | 0.221 | 0.078 | **4.63E-03** | 4,649 | - | - | - | - | - | - | - | - | - | - |
| avgSaO$_2$ | rs80108997 | 6,202 | 0.021 | 0.100 | 0.082 | 2.22E-01 | 5,978 | - | - | - | - | - | - | - | - | - | - | 0.029 | -0.051 | 0.126 | 6.83E-01 | 1,303 | 0.029 | -0.071 | 0.127 | 5.75E-01 | 1,258 | - | - | - | - | - | - | - | - | - | - |
| avgSaO$_2$ | rs187178912 | - | - | - | - | - | - | 0.005 | 0.057 | 0.079 | 4.66E-01 | 23,768 | NA | - | - | - | - | - | - | - | - | - | - | - | - | - | - | 0.006 | 0.218 | 0.138 | 1.13E-01 | 5,872 | 0.006 | 0.081 | 0.143 | 5.70E-01 | 5,705 |
| avgSaO$_2$ | rs112771667 | - | - | - | - | - | - | - | - | - | - | - | 0.005 | -0.013 | 0.090 | 8.89E-01 | 18,770 | 0.006 | -0.118 | 0.197 | 5.49E-01 | 2,634 | 0.006 | -0.031 | 0.198 | 8.75E-01 | 2,473 | 0.006 | 0.003 | 0.158 | 9.85E-01 | 5,025 | 0.006 | -0.010 | 0.160 | 9.52E-01 | 4,858 |
| avgSaO$_2$ | rs11166851 | 28,486 | 0.150 | -0.014 | 0.014 | 3.28E-01 | 24,511 | 0.146 | -0.009 | 0.014 | 5.34E-01 | 23,768 | 0.146 | -0.008 | 0.015 | 5.83E-01 | 19,888 | 0.161 | -0.032 | 0.025 | 1.96E-01 | 6,908 | 0.161 | -0.030 | 0.026 | 2.34E-01 | 6,713 | 0.149 | -0.047 | 0.028 | 9.22E-02 | 5,872 | 0.149 | -0.040 | 0.028 | 1.56E-01 | 5,705 |
| avgSaO$_2$ | rs7041166 | 28,486 | 0.893 | 0.037 | 0.015 | **1.47E-02** | 24,511 | 0.895 | 0.028 | 0.016 | 7.24E-02 | 23,768 | 0.894 | 0.029 | 0.017 | 8.11E-02 | 19,888 | 0.883 | -0.041 | 0.027 | 1.29E-01 | 6,908 | 0.884 | -0.042 | 0.028 | 1.26E-01 | 6,713 | 0.892 | -0.044 | 0.030 | 1.44E-01 | 5,872 | 0.893 | -0.045 | 0.031 | 1.45E-01 | 5,705 |
| avgSaO$_2$ | rs550460908 | 25,761 | 0.994 | 0.035 | 0.067 | 6.06E-01 | 21,791 | 0.993 | 0.031 | 0.062 | 6.17E-01 | 23,768 | 0.993 | 0.061 | 0.068 | 3.69E-01 | 19,888 | 0.993 | -0.104 | 0.109 | 3.38E-01 | 6,898 | 0.993 | -0.079 | 0.110 | 4.70E-01 | 6,703 | 0.992 | -0.120 | 0.111 | 2.81E-01 | 5,872 | 0.991 | -0.095 | 0.111 | 3.94E-01 | 5,705 |
| avgSaO$_2$ | rs199819010 | - | - | - | - | - | - | 0.007 | 0.021 | 0.101 | 8.34E-01 | 9,858 | 0.007 | -0.121 | 0.109 | 2.64E-01 | 8,709 | 0.005 | 0.417 | 0.260 | 1.09E-01 | 2,150 | 0.005 | 0.469 | 0.262 | 7.31E-02 | 2,105 | 0.008 | 0.179 | 0.143 | 2.09E-01 | 4,580 | 0.008 | 0.236 | 0.144 | 1.01E-01 | 4,527 |
| avgSaO$_2$ | rs2812667 | 25,782 | 0.024 | -0.023 | 0.034 | 5.01E-01 | 21,808 | 0.025 | -0.042 | 0.032 | 1.86E-01 | 23,768 | 0.025 | -0.022 | 0.035 | 5.18E-01 | 19,888 | 0.023 | 0.016 | 0.061 | 7.88E-01 | 6,908 | 0.023 | -0.019 | 0.063 | 7.66E-01 | 6,713 | 0.024 | 0.039 | 0.064 | 5.46E-01 | 5,872 | 0.024 | 0.007 | 0.066 | 9.12E-01 | 5,705 |
| avgSaO$_2$ | rs771251835 | 12,268 | 0.859 | -0.021 | 0.020 | 2.98E-01 | 11,009 | 0.860 | -0.019 | 0.021 | 3.58E-01 | 10,431 | 0.861 | -0.023 | 0.022 | 3.02E-01 | 9,257 | 0.862 | -0.010 | 0.028 | 7.20E-01 | 5,577 | 0.863 | -0.014 | 0.029 | 6.20E-01 | 5,498 | 0.865 | -0.034 | 0.031 | 2.85E-01 | 4,580 | 0.865 | -0.035 | 0.031 | 2.71E-01 | 4,527 |
| avgSaO$_2$ | rs4939452 | 23,691 | 0.484 | -0.019 | 0.011 | 8.40E-02 | 19,892 | 0.467 | -0.021 | 0.010 | **4.36E-02** | 21,859 | 0.469 | -0.019 | 0.011 | 9.55E-02 | 18,142 | 0.522 | 0.020 | 0.021 | 3.50E-01 | 5,575 | 0.522 | 0.019 | 0.021 | 3.74E-01 | 5,496 | 0.497 | 0.016 | 0.023 | 4.88E-01 | 4,580 | 0.498 | 0.016 | 0.023 | 5.03E-01 | 4,527 |
| avgSaO$_2$ | rs142696393 | 7,320 | 0.901 | 0.014 | 0.044 | 7.45E-01 | 7,096 | 0.988 | -1.467 | 0.841 | 8.11E-02 | 1,118 | 0.988 | -1.511 | 0.832 | 6.92E-02 | 1,118 | 0.869 | -0.040 | 0.061 | 5.12E-01 | 2,150 | 0.868 | -0.056 | 0.063 | 3.72E-01 | 2,105 | 0.988 | 2.729 | 1.595 | 8.71E-02 | 847 | 0.988 | 2.706 | 1.593 | 8.94E-02 | 847 |
| avgSaO$_2$ | rs114572017 | 6,202 | 0.010 | -0.067 | 0.109 | 5.36E-01 | 5,978 | - | - | - | - | - | - | - | - | - | - | 0.012 | 0.005 | 0.188 | 9.78E-01 | 1,303 | 0.012 | -0.034 | 0.191 | 8.59E-01 | 1,258 | - | - | - | - | - | - | - | - | - | - |
| avgSaO$_2$ | rs138247706 | 18,748 | 0.014 | -0.049 | 0.052 | 3.50E-01 | 15,981 | 0.016 | 0.011 | 0.043 | 8.03E-01 | 23,768 | 0.016 | 0.021 | 0.047 | 6.61E-01 | 19,888 | 0.017 | 0.060 | 0.104 | 5.61E-01 | 3,481 | 0.017 | 0.120 | 0.109 | 2.70E-01 | 3,320 | 0.020 | 0.036 | 0.085 | 6.70E-01 | 5,872 | 0.020 | 0.058 | 0.086 | 5.04E-01 | 5,705 |
| avgSaO$_2$ | rs529918807 | 5,234 | 0.993 | -0.005 | 0.128 | 9.68E-01 | 5,182 | 0.991 | -0.018 | 0.087 | 8.40E-01 | 10,431 | 0.991 | 0.003 | 0.092 | 9.75E-01 | 9,257 | 0.994 | -0.322 | 0.262 | 2.19E-01 | 2,150 | 0.994 | -0.133 | 0.273 | 6.27E-01 | 2,105 | 0.992 | -0.010 | 0.144 | 9.43E-01 | 4,580 | 0.992 | 0.014 | 0.151 | 9.24E-01 | 4,063 |
| avgSaO$_2$ | rs2732462 | 25,782 | 0.653 | 0.001 | 0.010 | 9.15E-01 | 21,808 | 0.636 | 0.003 | 0.010 | 7.99E-01 | 23,768 | 0.637 | 0.003 | 0.011 | 8.07E-01 | 19,888 | 0.681 | -0.030 | 0.019 | 1.02E-01 | 6,908 | 0.681 | -0.034 | 0.019 | 6.92E-02 | 6,713 | 0.659 | -0.026 | 0.020 | 1.83E-01 | 5,872 | 0.658 | -0.028 | 0.020 | 1.72E-01 | 5,705 |
| avgSaO$_2$ | rs77913955 | 6,202 | 0.026 | -0.104 | 0.075 | 1.66E-01 | 5,978 | - | - | - | - | - | - | - | - | - | - | 0.036 | 0.023 | 0.115 | 8.40E-01 | 1,303 | 0.037 | 0.016 | 0.116 | 8.89E-01 | 1,258 | - | - | - | - | - | - | - | - | - | - |
| avgSaO$_2$ | rs117349588 | - | - | - | - | - | - | 0.005 | 0.132 | 0.130 | 3.07E-01 | 8,740 | - | - | - | - | - | - | - | - | - | - | - | - | - | - | - | 0.006 | 0.070 | 0.210 | 7.40E-01 | 3,250 | 0.006 | 0.015 | 0.208 | 9.42E-01 | 3,216 |
| avgSaO$_2$ | rs565226335 | 15,071 | 0.986 | 0.013 | 0.061 | 8.32E-01 | 12,304 | 0.985 | 0.001 | 0.055 | 9.82E-01 | 13,910 | 0.985 | 0.012 | 0.061 | 8.47E-01 | 11,179 | 0.992 | 0.440 | 0.175 | **1.19E-02** | 2,634 | 0.992 | 0.439 | 0.180 | **1.46E-02** | 2,473 | 0.990 | 0.385 | 0.184 | **3.60E-02** | 1,775 | 0.990 | 0.376 | 0.189 | **4.75E-02** | 1,642 |
| avgSaO$_2$ | rs79931065 | 10,256 | 0.047 | -0.049 | 0.051 | 3.35E-01 | 9,286 | - | - | - | - | - | - | - | - | - | - | 0.064 | 0.091 | 0.076 | 2.33E-01 | 3,273 | 0.065 | 0.092 | 0.077 | 2.31E-01 | 3,210 | - | - | - | - | - | - | - | - | - | - |
| avgSaO$_2$ | rs35127183 | 12,268 | 0.316 | -0.005 | 0.015 | 7.14E-01 | 11,009 | 0.342 | -0.013 | 0.015 | 3.88E-01 | 10,431 | 0.341 | -0.010 | 0.016 | 5.47E-01 | 9,257 | 0.316 | -0.014 | 0.021 | 4.90E-01 | 5,577 | 0.316 | -0.027 | 0.021 | 2.01E-01 | 5,498 | 0.343 | -0.015 | 0.022 | 4.90E-01 | 4,580 | 0.344 | -0.026 | 0.022 | 2.53E-01 | 4,527 |
| avgSaO$_2$ | rs75660329 | 4,116 | 0.011 | 0.189 | 0.131 | 1.47E-01 | 4,064 | - | - | - | - | - | - | - | - | - | - | 0.015 | -0.094 | 0.174 | 5.88E-01 | 1,303 | 0.016 | -0.090 | 0.174 | 6.07E-01 | 1,258 | - | - | - | - | - | - | - | - | - | - |
| avgSaO$_2$ | rs866759288 | - | - | - | - | - | - | - | - | - | - | - | 0.995 | -0.458 | 0.459 | 3.18E-01 | 548 | - | - | - | - | - | - | - | - | - | - | 0.995 | 0.465 | 0.460 | 3.12E-01 | 483 | 0.994 | 0.582 | 0.460 | 2.06E-01 | 464 |
| avgSaO$_2$ | rs73939879 | 6,202 | 0.013 | -0.088 | 0.121 | 4.66E-01 | 5,978 | - | - | - | - | - | - | - | - | - | - | 0.015 | -0.308 | 0.174 | 7.73E-02 | 1,303 | 0.015 | -0.186 | 0.177 | 2.93E-01 | 1,258 | - | - | - | - | - | - | - | - | - | - |
| avgSaO$_2$ | rs187860354 | 4,116 | 0.991 | -0.149 | 0.137 | 2.74E-01 | 4,064 | - | - | - | - | - | - | - | - | - | - | 0.986 | 0.085 | 0.178 | 6.34E-01 | 1,303 | 0.987 | 0.081 | 0.187 | 6.63E-01 | 1,258 | - | - | - | - | - | - | - | - | - | - |
| avgSaO$_2$ | rs187041178 | 4,116 | 0.991 | -0.010 | 0.123 | 9.38E-01 | 4,064 | - | - | - | - | - | - | - | - | - | - | 0.988 | 0.056 | 0.192 | 7.72E-01 | 1,303 | 0.988 | -0.011 | 0.199 | 9.54E-01 | 1,258 | - | - | - | - | - | - | - | - | - | - |
| avgSaO$_2$ | rs868057231 | 1,557 | 0.974 | 0.382 | 0.115 | **9.34E-04** | 1,505 | - | - | - | - | - | - | - | - | - | - | 0.973 | -0.036 | 0.130 | 7.83E-01 | 1,303 | 0.972 | -0.084 | 0.133 | 5.29E-01 | 1,258 | - | - | - | - | - | - | - | - | - | - |
| avgSaO$_2$ | rs16955575 | 18,748 | 0.035 | 0.037 | 0.032 | 2.50E-01 | 15,981 | 0.038 | 0.004 | 0.027 | 8.77E-01 | 23,768 | 0.038 | 0.026 | 0.029 | 3.66E-01 | 19,888 | 0.033 | -0.101 | 0.073 | 1.67E-01 | 3,481 | 0.032 | -0.049 | 0.076 | 5.25E-01 | 3,320 | 0.039 | -0.039 | 0.054 | 4.75E-01 | 5,872 | 0.039 | -0.032 | 0.055 | 5.66E-01 | 5,705 |
| avgSaO$_2$ | rs113197061 | 4,116 | 0.008 | 0.228 | 0.158 | 1.50E-01 | 4,064 | - | - | - | - | - | - | - | - | - | - | 0.010 | -0.171 | 0.229 | 4.57E-01 | 1,303 | 0.010 | -0.242 | 0.230 | 2.93E-01 | 1,258 | - | - | - | - | - | - | - | - | - | - |
| avgSaO$_2$ | rs12609338 | 28,486 | 0.664 | -0.012 | 0.010 | 2.30E-01 | 24,511 | 0.674 | -0.015 | 0.010 | 1.37E-01 | 23,768 | 0.673 | -0.015 | 0.011 | 1.81E-01 | 19,888 | 0.683 | -0.008 | 0.019 | 6.66E-01 | 6,908 | 0.683 | -0.013 | 0.019 | 4.87E-01 | 6,713 | 0.675 | -0.009 | 0.020 | 6.47E-01 | 5,872 | 0.675 | -0.015 | 0.020 | 4.57E-01 | 5,705 |
| avgSaO$_2$ | rs76190072 | 4,116 | 0.986 | 0.142 | 0.115 | 2.18E-01 | 4,064 | - | - | - | - | - | - | - | - | - | - | 0.983 | -0.166 | 0.165 | 3.14E-01 | 1,303 | 0.982 | -0.143 | 0.166 | 3.88E-01 | 1,258 | - | - | - | - | - | - | - | - | - | - |
| avgSaO$_2$ | rs233266 | 12,985 | 0.047 | -0.058 | 0.037 | 1.20E-01 | 10,390 | 0.048 | -0.085 | 0.034 | **1.14E-02** | 12,001 | 0.048 | -0.064 | 0.038 | 8.68E-02 | 9,433 | 0.019 | 0.058 | 0.155 | 7.08E-01 | 1,303 | 0.019 | 0.039 | 0.160 | 8.09E-01 | 1,258 | 0.037 | 0.119 | 0.207 | 5.64E-01 | 483 | 0.036 | 0.108 | 0.217 | 6.17E-01 | 464 |
| avgSaO$_2$ | rs57204056 | - | 0.027 | 0.013 | 0.074 | 8.62E-01 | 7,363 | - | - | - | - | - | - | - | - | - | - | 0.035 | -0.059 | 0.104 | 5.66E-01 | 2,068 | 0.034 | -0.091 | 0.105 | 3.90E-01 | 2,017 | - | - | - | - | - | - | - | - | - | - |

**Supplementary Table 12.** Results from LD score regression using LDSC.

Lower matrix values represent estimated genetic correlation between traits. Upper triangle values are P-values from genetic correlations, with nominally significant associations italicized (P<0.05) and Bonferroni-corrected significant correlations bolded. Abbreviations: AHI - apnea-hypopnea index; BMI - body mas index; adjBMI - adjusted for BMI; avgSaO2 - average oxygen saturation; OSA - obstructive sleep apnea; T2D - type II diabetes; RG - random glucose; VAT - visceral adipose tissue; SAT - subcutaneous adipose tissue; PuP - pulse pressure; SBP - systolic blood pressure; DBP - diastolic blood pressure; TG - triglycerides; TC - total cholesterol; HDL - high-density lipoprotein cholesterol; LDL - low-density lipoprotein cholesterol; HIPadjBMI - hip circumference adjusted for BMI; WCadjBMI - waist circumference adjusted for BMI; WHRadjBMI - waist to hip ratio adjusted for BMI; CAD - coronary artery disease.

| Trait | Lambda | Intercept | Ratio | AHI | AHI adjBMI | avgSaO$_2$ | avgSaO$_2$ adjBMI* | OSA | OSA adjBMI | Insomnia | Sleepiness adjBMI | Sleepiness | T2D | RG | VAT | SAT | Pancreas Volume | Pancreas Fat | Liver Fat | PuP | SBP | DBP | TG | TC | HDL | LDL | HIP adjBMI | WC adjBMI | WHR adjBMI | BMI | CAD |
|---|---|---|---|---|---|---|---|---|---|---|---|---|---|---|---|---|---|---|---|---|---|---|---|---|---|---|---|---|---|---|---|
| **AHI** | 1.0375 | 1.0051 | 0.1453 | | **3.56E-17** | 2.37E-01 | 9.09E-01 | **2.05E-05** | *1.88E-02* | 8.48E-01 | 3.93E-01 | *4.79E-02* | **5.88E-05** | 1.02E-01 | *1.83E-03* | *2.06E-03* | 8.33E-02 | 8.12E-02 | 2.32E-01 | 3.82E-01 | 5.08E-01 | 8.79E-01 | *1.85E-02* | 5.88E-01 | *2.56E-04* | 9.06E-01 | 4.85E-01 | 6.32E-01 | 2.26E-01 | **8.32E-10** | *3.04E-02* |
| **AHI adjBMI** | 1.0436 | 1.0145 | 0.3744 | **0.7466** | | 7.53E-01 | 8.76E-01 | 1.01E-01 | *8.00E-04* | 5.83E-01 | 2.51E-01 | 3.72E-01 | 2.03E-01 | 6.66E-02 | 1.15E-01 | 8.08E-01 | 9.95E-02 | 7.04E-01 | 5.71E-01 | 1.14E-01 | 1.43E-01 | 3.07E-01 | 7.09E-02 | 5.45E-01 | 3.15E-01 | 5.67E-01 | 1.22E-01 | 8.69E-01 | *1.07E-02* | *3.55E-02* | 5.62E-01 |
| **avgSaO$_2$** | 1.0105 | 0.9984 | < 0 | -0.3987 | 0.1329 | | 4.00E-01 | *6.20E-03* | 2.52E-01 | 8.90E-02 | 9.22E-01 | 3.64E-01 | *6.60E-03* | 4.32E-01 | 1.49E-01 | 1.27E-01 | 1.55E-01 | 6.33E-01 | 7.85E-01 | 6.28E-01 | 9.26E-01 | 7.27E-01 | *1.73E-02* | 9.93E-01 | *4.28E-02* | 8.92E-01 | 4.32E-01 | 1.43E-01 | *1.55E-02* | *4.90E-03* | *1.18E-02* |
| **avgSaO$_2$ adjBMI*** | 1.0075 | 1.0002 | 0.0394 | -0.075 | 0.1142 | 0.464 | | 3.31E-01 | 3.22E-01 | 5.30E-01 | 9.65E-01 | 8.16E-01 | 2.86E-01 | 6.30E-01 | 8.35E-01 | 6.34E-01 | 4.28E-01 | 5.52E-01 | 9.69E-01 | 9.47E-01 | 7.32E-01 | 6.47E-01 | 2.91E-01 | 6.72E-01 | 3.63E-01 | 7.57E-01 | NA* | NA* | 3.12E-01 | 3.37E-01 | 3.41E-01 |
| **OSA** | 1.1908 | 0.9704 | < 0 | **0.4435** | 0.1703 | *-0.4748* | -0.3275 | | **0.00E+00** | **8.45E-11** | **7.94E-11** | **7.86E-25** | **2.51E-38** | 2.44E-01 | **6.63E-19** | **2.60E-33** | *1.51E-02* | **6.20E-06** | **6.42E-07** | 5.46E-01 | 6.95E-01 | 7.41E-01 | **1.65E-20** | 5.99E-01 | **1.22E-37** | 3.54E-01 | 3.44E-01 | **1.01E-04** | *4.80E-04* | **3.99E-270** | **5.13E-37** |
| **OSA adjBMI** | 1.105 | 0.9819 | < 0 | *0.2662* | *0.485* | -0.1831 | -0.3928 | **0.8435** | | *2.14E-04* | **5.53E-13** | **3.04E-14** | **3.32E-06** | 4.31E-01 | **1.19E-04** | *3.55E-02* | 6.68E-02 | 2.46E-01 | *6.76E-04* | 9.88E-01 | 4.25E-01 | 1.20E-01 | **9.44E-10** | 2.92E-01 | **1.27E-10** | 1.50E-01 | 4.46E-01 | **5.46E-07** | **4.12E-17** | *5.97E-04* | **2.53E-11** |
| **Insomnia** | 1.3068 | 1.0134 | 0.0369 | -0.016 | -0.0465 | -0.3181 | -0.2634 | **0.2208** | *0.1669* | | **1.58E-11** | **1.11E-16** | **8.97E-18** | *4.10E-03* | **5.63E-08** | **7.89E-06** | 7.78E-01 | *9.25E-04* | **3.32E-05** | *8.13E-03* | **2.08E-05** | *4.88E-04* | **4.85E-13** | *5.26E-03* | **6.00E-08** | *7.35E-03* | *8.06E-03* | 1.27E-01 | **5.81E-06** | **5.04E-19** | **1.09E-23** |
| **Sleepiness adjBMI** | 1.3685 | 1.0163 | 0.0375 | 0.0616 | 0.0945 | 0.0105 | 0.007771 | **0.2014** | **0.2748** | **0.1976** | | **0.00E+00** | 2.57E-01 | 9.79E-02 | 5.53E-01 | 4.86E-01 | 5.49E-01 | 4.87E-01 | 8.26E-01 | 7.19E-01 | 7.09E-01 | 2.74E-01 | *2.85E-02* | 6.97E-01 | *8.30E-03* | 5.50E-01 | 2.68E-01 | 1.61E-01 | *4.60E-02* | 7.97E-02 | *1.60E-03* |
| **Sleepiness** | 1.3685 | 1.0165 | 0.0373 | *0.1424* | 0.0726 | -0.1044 | -0.04232 | **0.3127** | **0.2913** | **0.2342** | **0.9845** | | **2.60E-10** | 4.72E-01 | *1.05E-03* | *8.11E-03* | 6.31E-01 | 2.28E-01 | *4.97E-02* | 8.59E-01 | 6.53E-01 | 3.95E-01 | **6.11E-07** | 6.09E-01 | **8.02E-11** | 8.16E-01 | 3.70E-01 | 2.37E-01 | 7.10E-02 | **8.84E-13** | **2.02E-08** |
| **T2D** | 1.4316 | 0.9355 | < 0 | **0.3083** | 0.1017 | *-0.4952* | -0.4791 | **0.3554** | **0.1419** | **0.1884** | 0.0203 | **0.1112** | | **3.08E-19** | **1.10E-35** | **2.26E-28** | 1.53E-01 | **9.65E-13** | **5.32E-30** | **9.91E-34** | **8.85E-39** | **3.26E-11** | **1.27E-29** | 2.56E-01 | **2.61E-78** | 8.87E-02 | **8.85E-06** | **2.15E-13** | **1.40E-41** | **4.17E-99** | **1.64E-75** |
| **RG** | 1.2597 | 1.0478 | 0.0949 | 0.1179 | 0.1656 | -0.1078 | -0.1553 | 0.0348 | -0.0303 | *0.0888* | -0.0475 | -0.0196 | **0.4383** | | *1.17E-03* | *2.35E-04* | 2.14E-01 | *2.98E-02* | *1.05E-03* | **5.69E-30** | **2.61E-21** | **1.05E-06** | *2.31E-02* | 7.37E-01 | *1.72E-03* | 5.63E-01 | 9.33E-02 | 3.29E-01 | *1.64E-04* | **6.14E-08** | **1.38E-05** |
| **VAT** | 1.1459 | 1.0201 | 0.131 | *0.354* | 0.1924 | -0.2718 | -0.0561 | **0.4203** | **0.2241** | **0.2309** | 0.0262 | *0.1419* | **0.4905** | *0.1339* | | **9.50E-152** | 6.86E-01 | **1.01E-17** | **1.13E-24** | *3.77E-03* | *1.12E-03* | *4.58E-02* | **5.20E-25** | 6.74E-02 | **4.26E-45** | *3.53E-02* | 5.34E-02 | **2.62E-17** | **5.27E-34** | **2.01E-95** | **3.88E-18** |
| **SAT** | 1.1459 | 1.0236 | 0.1515 | *0.3558* | -0.0306 | -0.2994 | 0.1552 | **0.5252** | *0.1275* | **0.2011** | -0.0311 | *0.118* | **0.4783** | *0.1471* | **0.7628** | | 1.45E-01 | **2.03E-07** | **2.11E-19** | 6.08E-02 | 8.81E-02 | 5.54E-01 | **1.19E-17** | 7.02E-01 | **8.13E-30** | 2.36E-01 | **1.49E-10** | **2.17E-09** | *6.75E-03* | **9.97E-160** | **1.32E-14** |
| **Pancreas Volume** | 1.0957 | 1.0098 | 0.0772 | 0.2032 | 0.2252 | 0.3101 | 0.4542 | *0.1124* | 0.1079 | 0.0127 | -0.0231 | **-0.0187** | -0.0551 | -0.0535 | 0.0264 | 0.091 | | **2.46E-08** | 6.58E-01 | 6.09E-02 | *2.61E-02* | 7.28E-02 | 9.70E-01 | 1.30E-01 | 7.20E-02 | 2.14E-01 | *3.11E-04* | *2.84E-04* | 2.58E-01 | 1.87E-01 | 8.51E-02 |
| **Pancreas Fat** | 1.0957 | 1.0188 | 0.1974 | 0.2683 | 0.0623 | -0.106 | 0.2751 | **0.275** | 0.0764 | *0.1875* | -0.0305 | 0.0526 | **0.3412** | *0.1007* | **0.5359** | **0.3491** | **-0.4671** | | *2.19E-04* | *1.34E-02* | *2.51E-02* | 2.85E-01 | **3.68E-09** | 5.17E-02 | **8.89E-10** | *2.45E-02* | 7.88E-01 | *1.35E-04* | **7.13E-10** | **3.85E-26** | **1.67E-13** |
| **Liver Fat** | 1.0957 | 1.0107 | 0.0885 | 0.1484 | 0.0792 | -0.0522 | 0.01174 | **0.2952** | *0.241* | **0.2065** | 0.0098 | *0.088* | **0.5278** | *0.1666* | **0.6746** | **0.5584** | -0.0303 | *0.324* | | **1.01E-04** | *2.68E-04* | 1.63E-01 | **5.25E-06** | 5.69E-01 | **1.08E-15** | 4.89E-01 | *1.42E-03* | **2.93E-05** | **1.20E-18** | **1.77E-17** | **9.79E-07** |
| **PuP** | 2.0513 | 1.0591 | 0.0268 | 0.0475 | 0.1033 | 0.0394 | 0.009181 | -0.0136 | 0.0004 | *0.0493* | -0.0069 | -0.0034 | **0.2017** | **0.2436** | 0.0816 | 0.0521 | -0.0644 | *0.0855* | **0.1307** | | **0.00E+00** | **6.46E-34** | **1.59E-08** | 7.13E-01 | *3.30E-04* | 2.49E-01 | *2.15E-04* | 8.53E-02 | **1.05E-07** | 6.31E-01 | **6.51E-35** |
| **SBP** | 2.2469 | 1.1145 | 0.045 | 0.0362 | 0.1043 | -0.0073 | -0.05289 | -0.0088 | 0.0219 | **0.0839** | 0.0072 | 0.0087 | **0.2055** | **0.2359** | 0.0966 | 0.0501 | *-0.0746* | 0.0788 | *0.1183* | **0.8404** | | **0.00E+00** | **1.88E-11** | 9.23E-01 | **6.31E-08** | 3.28E-01 | **3.07E-05** | *2.78E-02* | **1.11E-09** | 6.44E-01 | **3.71E-57** |
| **DBP** | 2.1633 | 1.1161 | 0.0482 | 0.008 | 0.0717 | -0.0285 | -0.08112 | -0.0069 | 0.0432 | *0.0736* | 0.0215 | 0.0167 | **0.1014** | **0.1278** | 0.064 | 0.0178 | -0.0593 | 0.0383 | 0.0468 | **0.3101** | **0.7752** | | **9.82E-07** | 9.06E-01 | **3.91E-05** | 7.18E-01 | *3.23E-03* | 1.22E-01 | **4.90E-06** | *1.31E-02* | **4.76E-26** |
| **TG** | 2.0302 | 1.2364 | 0.0784 | *0.155* | 0.1279 | *-0.3106* | -0.4027 | **0.2563** | **0.1837** | **0.1373** | *0.0488* | **0.1049** | **0.4438** | *0.0945* | **0.4632** | **0.3458** | 0.0014 | **0.2707** | **0.4766** | **0.1078** | **0.1411** | **0.1024** | | **4.53E-10** | **2.29E-48** | **2.85E-07** | **6.29E-15** | **5.31E-10** | **4.52E-30** | **6.84E-28** | **6.54E-32** |
| **TC** | 1.6908 | 1.4645 | 0.1588 | -0.032 | 0.041 | 0.0008 | -0.06045 | -0.0138 | 0.0293 | *0.0722* | -0.007 | -0.009 | 0.0267 | 0.0103 | 0.0735 | 0.0147 | **-0.0549** | 0.0837 | 0.0458 | -0.0082 | -0.0018 | 0.0022 | **0.403** | | *3.01E-04* | **0.00E+00** | **6.28E-12** | 5.22E-01 | **3.97E-09** | 5.35E-01 | *4.56E-03* |
| **HDL** | 2.2027 | 1.5432 | 0.1624 | *-0.2057* | -0.0583 | *0.241* | 0.2092 | **-0.334** | **-0.19** | **-0.1185** | -0.0467 | **-0.1184** | **-0.4303** | *-0.084* | **-0.4558** | **-0.3958** | -0.0548 | **-0.2577** | **-0.4303** | -0.0552 | **-0.0825** | **-0.0646** | **-0.645** | *0.2682* | | 8.23E-02 | *1.37E-03* | **1.35E-11** | **1.62E-25** | **3.47E-69** | **1.49E-36** |
| **LDL** | 1.5771 | 1.4498 | 0.1971 | 0.008 | 0.0439 | 0.0122 | -0.0446 | 0.0287 | 0.0462 | *0.0761* | -0.0112 | -0.0042 | 0.0416 | 0.0164 | *0.0948* | 0.0512 | -0.0499 | *0.104* | 0.0538 | -0.0309 | -0.022 | -0.0076 | **0.333** | **0.9419** | 0.1099 | | **2.23E-09** | 7.98E-01 | **3.52E-08** | 1.67E-01 | *4.98E-04* |
| **HIP adjBMI** | 1.0466 | 0.8062 | < 0 | -0.0528 | -0.1371 | -0.1238 | NA* | 0.0265 | -0.0286 | *-0.0748* | 0.0276 | 0.0222 | **-0.1022** | -0.0487 | 0.0799 | **0.2368** | *0.1535* | 0.0124 | *-0.1346* | *-0.0854* | **-0.0924** | -0.0601 | **-0.1911** | **-0.1542** | *0.0836* | **-0.1438** | | **3.49E-75** | **3.55E-34** | 6.16E-01 | **4.27E-11** |
| **WC adjBMI** | 1.0466 | 0.8227 | < 0 | -0.0361 | -0.015 | -0.3211 | NA* | **0.1281** | **0.2059** | 0.0445 | 0.0364 | 0.0304 | **0.1863** | 0.0286 | **0.3775** | **0.2514** | *0.1553* | **0.1939** | **0.1868** | 0.037 | *0.0467* | 0.035 | **0.1697** | -0.0154 | **-0.1914** | 0.007 | **0.5297** | | **2.55E-237** | 1.13E-01 | *1.97E-02* |
| **WHR adjBMI** | 1.8044 | 1.1359 | 0.0861 | 0.0811 | *0.1971* | -0.3203 | -0.5716 | *0.0834* | **0.2531** | **0.098** | 0.0416 | 0.0366 | **0.282** | 0.1052 | **0.3904** | 0.0868 | 0.0443 | **0.2734** | **0.3848** | **0.0976** | **0.1214** | **0.0936** | **0.3651** | **0.1283** | **-0.2682** | **0.1348** | **-0.3183** | **0.646** | | *1.92E-02* | **3.49E-22** |
| **BMI** | 2.689 | 1.0458 | 0.0158 | **0.4572** | *-0.1312* | *-0.6503* | -0.2615 | **0.6537** | *0.0963* | **0.1937** | -0.0356 | **0.1399** | **0.5129** | **0.1447** | **0.6608** | **0.8575** | 0.038 | **0.474** | **0.445** | 0.0067 | -0.0072 | *-0.0399* | **0.324** | -0.0131 | **-0.4158** | 0.0366 | -0.0099 | -0.0358 | *-0.0385* | | **4.02E-53** |
| **CAD** | 1.4745 | 1.0407 | 0.0448 | *0.1396* | 0.0415 | *-0.4062* | -0.381 | **0.2793** | **0.1949** | **0.2395** | *0.0662* | **0.12** | **0.3544** | **0.1024** | **0.2942** | **0.2758** | -0.0637 | **0.3154** | **0.2186** | **0.2819** | **0.3317** | **0.2474** | **0.323** | *0.1151* | **-0.2871** | *0.1444* | **-0.1389** | 0.051 | **0.1773** | **0.2897** | |

*Due to the lack of polygenecity in this trait, we could not estimate the genetic correlation between AVGSAO2adjBMI and some traits. These, $R^2$ values are likely biased and should be interpreted with caution.

**Supplementary Table 13.** PheWAS results for suggestively significant SNP-PheCode associations. Abbreviations: SE - standard error, OR - odds ratio, EAF - effect allele frequency.

| Discovery Trait | PheCode | SNP | Effect Allele | Beta | SE | OR | P | N | Ncases | Ncontrols | EAF | Phecode Description | Category |
|---|---|---|---|---|---|---|---|---|---|---|---|---|---|
| AHIadjBMI | 251 | rs78545995 | G | 0.409 | 0.110 | 1.506 | 2.03E-04 | 91,587 | 2,200 | 89,387 | 0.014 | Other disorders of pancreatic internal secretion | endocrine/metabolic |
| AHIadjBMI | 425.8 | rs78545995 | G | 0.993 | 0.258 | 2.700 | 1.16E-04 | 97,556 | 224 | 97,332 | 0.014 | Other cardiomyopathy | circulatory system |
| avgSa$O_2$ | 285.1 | rs2194955 | C | -0.138 | 0.038 | 0.871 | 2.48E-04 | 70,492 | 2,693 | 67,799 | 0.194 | Acute posthemorrhagic anemia | hematopoietic |
| avgSa$O_2$ adjBMI | 286.2 | rs79914264 | A | 0.290 | 0.077 | 1.337 | 1.74E-04 | 93,520 | 6,411 | 87,109 | 0.014 | Encounter for long-term (current) use of anticoagulants | hematopoietic |
| avgSa$O_2$ adjBMI | 764 | rs79914264 | A | -0.246 | 0.066 | 0.782 | 2.15E-04 | 84,317 | 11,980 | 72,337 | 0.014 | Sciatica | symptoms |
| OSA | 278.11 | rs11642015 | T | 0.216 | 0.012 | 1.241 | 9.29E-75 | 64,943 | 25,460 | 39,483 | 0.412 | Morbid obesity | endocrine/metabolic |
| OSA | 278.1 | rs11642015 | T | 0.159 | 0.010 | 1.172 | 8.38E-55 | 82,403 | 42,920 | 39,483 | 0.411 | Obesity | endocrine/metabolic |
| OSA | 250.2 | rs11642015 | T | 0.164 | 0.012 | 1.179 | 4.13E-44 | 85,668 | 24,255 | 61,413 | 0.406 | Type 2 diabetes | endocrine/metabolic |
| OSA | 278 | rs11642015 | T | 0.131 | 0.010 | 1.140 | 1.08E-41 | 92,907 | 53,424 | 39,483 | 0.409 | Overweight, obesity and other hyperalimentation | endocrine/metabolic |
| OSA | 250 | rs11642015 | T | 0.134 | 0.010 | 1.143 | 5.22E-39 | 97,948 | 36,535 | 61,413 | 0.407 | Diabetes mellitus | endocrine/metabolic |
| OSA | 250.24 | rs11642015 | T | 0.190 | 0.019 | 1.210 | 1.16E-22 | 68,211 | 6,798 | 61,413 | 0.400 | Type 2 diabetes with neurological manifestations | endocrine/metabolic |
| OSA | 327.3 | rs11642015 | T | 0.117 | 0.012 | 1.124 | 1.92E-21 | 80,868 | 18,806 | 62,062 | 0.408 | Sleep apnea | neurological |
| OSA | 250.6 | rs11642015 | T | 0.189 | 0.020 | 1.208 | 2.23E-20 | 67,439 | 6,026 | 61,413 | 0.400 | Polyneuropathy in diabetes | endocrine/metabolic |
| OSA | 250.22 | rs11642015 | T | 0.181 | 0.020 | 1.199 | 4.33E-20 | 68,458 | 7,045 | 61,413 | 0.401 | Type 2 diabetes with renal manifestations | endocrine/metabolic |
| OSA | 327.32 | rs11642015 | T | 0.117 | 0.013 | 1.124 | 5.16E-19 | 77,816 | 15,754 | 62,062 | 0.407 | Obstructive sleep apnea | neurological |
| OSA | 401 | rs11642015 | T | 0.098 | 0.011 | 1.102 | 2.06E-18 | 99,691 | 53,959 | 45,732 | 0.408 | Hypertension | circulatory system |
| OSA | 401.1 | rs11642015 | T | 0.097 | 0.011 | 1.102 | 3.70E-18 | 98,954 | 53,222 | 45,732 | 0.407 | Essential hypertension | circulatory system |
| OSA | 250.23 | rs11642015 | T | 0.222 | 0.026 | 1.249 | 4.60E-17 | 64,864 | 3,451 | 61,413 | 0.399 | Type 2 diabetes with ophthalmic manifestations | endocrine/metabolic |
| OSA | 250.3 | rs11642015 | T | 0.166 | 0.021 | 1.181 | 7.33E-16 | 67,160 | 5,747 | 61,413 | 0.399 | Insulin pump user | endocrine/metabolic |
| OSA | 782.3 | rs11642015 | T | 0.111 | 0.014 | 1.118 | 6.10E-15 | 94,009 | 13,020 | 80,989 | 0.407 | Edema | symptoms |
| OSA | 569.2 | rs11642015 | T | 0.190 | 0.025 | 1.209 | 3.57E-14 | 96,202 | 3,511 | 92,691 | 0.407 | Gastrointestinal complications | digestive |
| OSA | 278.4 | rs11642015 | T | 0.161 | 0.021 | 1.175 | 6.20E-14 | 44,949 | 5,466 | 39,483 | 0.397 | Abnormal weight gain | endocrine/metabolic |
| OSA | 782 | rs11642015 | T | 0.104 | 0.014 | 1.110 | 6.89E-14 | 94,851 | 13,862 | 80,989 | 0.407 | Symptoms involving skin and other integumentary tissue | symptoms |
| OSA | 250.4 | rs11642015 | T | 0.092 | 0.013 | 1.096 | 6.93E-13 | 79,393 | 17,980 | 61,413 | 0.401 | Abnormal glucose | endocrine/metabolic |
| OSA | 250.7 | rs11642015 | T | 0.181 | 0.025 | 1.199 | 7.01E-13 | 90,617 | 3,598 | 87,019 | 0.406 | Diabetic retinopathy | endocrine/metabolic |
| OSA | 250.42 | rs11642015 | T | 0.092 | 0.014 | 1.096 | 1.28E-11 | 76,679 | 15,266 | 61,413 | 0.400 | Other abnormal glucose | endocrine/metabolic |
| OSA | 569 | rs11642015 | T | 0.146 | 0.022 | 1.158 | 7.03E-11 | 97,090 | 4,399 | 92,691 | 0.407 | Other disorders of intestine | digestive |
| OSA | 327 | rs11642015 | T | 0.065 | 0.010 | 1.068 | 8.29E-11 | 94,609 | 32,547 | 62,062 | 0.407 | Sleep disorders | neurological |
| OSA | 740.9 | rs11642015 | T | 0.086 | 0.013 | 1.090 | 1.16E-10 | 87,100 | 19,806 | 67,294 | 0.406 | Osteoarthrosis NOS | musculoskeletal |
| OSA | 740 | rs11642015 | T | 0.074 | 0.012 | 1.076 | 2.00E-10 | 95,874 | 28,580 | 67,294 | 0.407 | Osteoarthrosis | musculoskeletal |
| OSA | 401.2 | rs11642015 | T | 0.121 | 0.020 | 1.128 | 6.42E-10 | 58,999 | 13,267 | 45,732 | 0.401 | Hypertensive heart and/or renal disease | circulatory system |
| OSA | 740.1 | rs11642015 | T | 0.074 | 0.012 | 1.077 | 2.63E-09 | 89,668 | 22,374 | 67,294 | 0.406 | Osteoarthritis; localized | musculoskeletal |
| OSA | 740.11 | rs11642015 | T | 0.074 | 0.013 | 1.077 | 3.54E-09 | 89,025 | 21,731 | 67,294 | 0.406 | Osteoarthrosis, localized, primary | musculoskeletal |
| OSA | 707 | rs11642015 | T | 0.132 | 0.023 | 1.141 | 4.60E-09 | 103,926 | 4,438 | 99,488 | 0.407 | Chronic ulcer of skin | dermatologic |
| OSA | 571.5 | rs11642015 | T | 0.100 | 0.017 | 1.105 | 5.05E-09 | 86,380 | 7,926 | 78,454 | 0.407 | Other chronic nonalcoholic liver disease | digestive |
| OSA | 571 | rs11642015 | T | 0.093 | 0.017 | 1.098 | 2.82E-08 | 86,669 | 8,215 | 78,454 | 0.407 | Chronic liver disease and cirrhosis | digestive |
| OSA | 509.8 | rs11642015 | T | 0.122 | 0.022 | 1.129 | 5.37E-08 | 89,182 | 4,469 | 84,713 | 0.406 | Dependence on respirator [Ventilator] or supplemental oxygen | respiratory |
| OSA | 681.5 | rs11642015 | T | 0.106 | 0.020 | 1.112 | 6.07E-08 | 74,555 | 6,100 | 68,455 | 0.405 | Cellulitis and abscess of leg, except foot | dermatologic |
| OSA | 456 | rs11642015 | T | 0.125 | 0.023 | 1.133 | 7.88E-08 | 76,158 | 4,304 | 71,854 | 0.405 | Chronic venous insufficiency [CVI] | circulatory system |
| OSA | 428.4 | rs11642015 | T | 0.121 | 0.023 | 1.129 | 8.76E-08 | 83,538 | 4,926 | 78,612 | 0.405 | Heart failure with preserved EF [Diastolic heart failure] | circulatory system |
| OSA | 509 | rs11642015 | T | 0.092 | 0.017 | 1.096 | 1.08E-07 | 92,659 | 7,946 | 84,713 | 0.407 | Respiratory failure, insufficiency, arrest | respiratory |
| OSA | 256 | rs11642015 | T | 0.187 | 0.036 | 1.206 | 1.46E-07 | 54,966 | 1,766 | 53,200 | 0.405 | Ovarian dysfunction | endocrine/metabolic |
| OSA | 256.4 | rs11642015 | T | 0.189 | 0.036 | 1.208 | 1.69E-07 | 54,912 | 1,712 | 53,200 | 0.405 | Polycystic ovaries | endocrine/metabolic |
| OSA | 251.1 | rs11642015 | T | 0.159 | 0.031 | 1.172 | 4.57E-07 | 56,951 | 2,250 | 54,701 | 0.397 | Hypoglycemia | endocrine/metabolic |
| OSA | 249 | rs11642015 | T | 0.203 | 0.040 | 1.225 | 5.47E-07 | 62,766 | 1,353 | 61,413 | 0.397 | Secondary diabetes mellitus | endocrine/metabolic |
| OSA | 707.2 | rs11642015 | T | 0.127 | 0.026 | 1.136 | 6.94E-07 | 102,839 | 3,351 | 99,488 | 0.406 | Chronic ulcer of leg or foot | dermatologic |
| OSA | 401.22 | rs11642015 | T | 0.114 | 0.023 | 1.120 | 1.19E-06 | 53,802 | 8,070 | 45,732 | 0.399 | Hypertensive chronic kidney disease | circulatory system |
| OSA | 454.11 | rs11642015 | T | 0.135 | 0.029 | 1.145 | 2.20E-06 | 74,547 | 2,693 | 71,854 | 0.404 | Varicose veins of lower extremity, symptomtic | circulatory system |
| OSA | 740.2 | rs11642015 | T | 0.086 | 0.018 | 1.090 | 2.72E-06 | 76,230 | 8,936 | 67,294 | 0.403 | Osteoarthrosis, generalized | musculoskeletal |
| OSA | 585.3 | rs11642015 | T | 0.067 | 0.014 | 1.069 | 2.86E-06 | 87,553 | 18,030 | 69,523 | 0.407 | Chronic renal failure [CKD] | genitourinary |
| OSA | 585 | rs11642015 | T | 0.062 | 0.013 | 1.064 | 4.14E-06 | 90,184 | 20,661 | 69,523 | 0.407 | Renal failure | genitourinary |
| OSA | 428 | rs11642015 | T | 0.077 | 0.017 | 1.080 | 5.61E-06 | 88,480 | 9,868 | 78,612 | 0.406 | Congestive heart failure; nonhypertensive | circulatory system |
| OSA | 539 | rs11642015 | T | 0.051 | 0.011 | 1.052 | 6.25E-06 | 93,472 | 22,236 | 71,236 | 0.407 | Bariatric surgery | digestive |
| OSA | 681 | rs11642015 | T | 0.058 | 0.013 | 1.060 | 8.20E-06 | 83,795 | 15,340 | 68,455 | 0.407 | Superficial cellulitis and abscess | dermatologic |
| OSA | 454.1 | rs11642015 | T | 0.106 | 0.024 | 1.111 | 1.20E-05 | 75,727 | 3,873 | 71,854 | 0.405 | Varicose veins of lower extremity | circulatory system |
| OSA | 585.32 | rs11642015 | T | 0.177 | 0.041 | 1.194 | 1.29E-05 | 70,876 | 1,353 | 69,523 | 0.403 | End stage renal disease | genitourinary |
| OSA | 159.3 | rs11642015 | T | -0.892 | 0.206 | 0.410 | 1.43E-05 | 101,331 | 69 | 101,262 | 0.407 | Malignant neoplasm of gallbladder and extrahepatic bile ducts | neoplasms |
| OSA | 357 | rs11642015 | T | 0.079 | 0.019 | 1.083 | 1.83E-05 | 97,750 | 6,766 | 90,984 | 0.406 | Inflammatory and toxic neuropathy | neurological |
| OSA | 580.3 | rs11642015 | T | 0.167 | 0.040 | 1.182 | 2.49E-05 | 70,934 | 1,411 | 69,523 | 0.403 | Nephritis and nephropathy without mention of glomerulonephritis | genitourinary |
| OSA | 585.33 | rs11642015 | T | 0.064 | 0.015 | 1.066 | 2.51E-05 | 85,557 | 16,034 | 69,523 | 0.406 | Chronic Kidney Disease, Stage III | genitourinary |
| OSA | 401.21 | rs11642015 | T | 0.109 | 0.026 | 1.115 | 2.67E-05 | 50,771 | 5,039 | 45,732 | 0.397 | Hypertensive heart disease | circulatory system |
| OSA | 110.11 | rs11642015 | T | 0.086 | 0.020 | 1.090 | 2.76E-05 | 77,521 | 5,640 | 71,881 | 0.406 | Dermatophytosis of nail | infectious diseases |
| OSA | 700 | rs11642015 | T | 0.112 | 0.027 | 1.119 | 3.11E-05 | 84,148 | 3,097 | 81,051 | 0.405 | Corns and callosities | dermatologic |
| OSA | 415.2 | rs11642015 | T | 0.113 | 0.027 | 1.119 | 3.71E-05 | 98,820 | 2,966 | 95,854 | 0.407 | Chronic pulmonary heart disease | circulatory system |
| OSA | 454 | rs11642015 | T | 0.096 | 0.023 | 1.101 | 3.73E-05 | 75,987 | 4,133 | 71,854 | 0.405 | Varicose veins | circulatory system |
| OSA | 288.2 | rs11642015 | T | 0.139 | 0.034 | 1.149 | 3.91E-05 | 83,621 | 1,851 | 81,770 | 0.408 | Elevated white blood cell count | hematopoietic |
| OSA | 250.25 | rs11642015 | T | 0.142 | 0.035 | 1.153 | 3.97E-05 | 63,324 | 1,911 | 61,413 | 0.397 | Diabetes type 2 with peripheral circulatory disorders | endocrine/metabolic |
| OSA | 272.11 | rs11642015 | T | 0.053 | 0.013 | 1.055 | 4.55E-05 | 80,119 | 37,358 | 42,761 | 0.407 | Hypercholesterolemia | endocrine/metabolic |
| OSA | 274.11 | rs11642015 | T | 0.107 | 0.026 | 1.113 | 4.70E-05 | 102,574 | 3,193 | 99,381 | 0.407 | Gouty arthropathy | endocrine/metabolic |
| OSA | 428.1 | rs11642015 | T | 0.076 | 0.019 | 1.079 | 5.60E-05 | 86,213 | 7,601 | 78,612 | 0.405 | Congestive heart failure (CHF) NOS | circulatory system |
| OSA | 366 | rs11642015 | T | 0.059 | 0.015 | 1.061 | 5.68E-05 | 100,643 | 15,682 | 84,961 | 0.407 | Cataract | sense organs |
| OSA | 250.1 | rs11642015 | T | 0.133 | 0.033 | 1.142 | 5.87E-05 | 63,473 | 2,060 | 61,413 | 0.397 | Type 1 diabetes | endocrine/metabolic |
| OSA | 415 | rs11642015 | T | 0.081 | 0.020 | 1.084 | 6.43E-05 | 101,455 | 5,601 | 95,854 | 0.407 | Pulmonary heart disease | circulatory system |
| OSA | 611.3 | rs11642015 | T | -0.121 | 0.030 | 0.886 | 6.45E-05 | 88,808 | 2,471 | 86,337 | 0.409 | Lump or mass in breast | genitourinary |
| OSA | 269 | rs11642015 | T | 0.109 | 0.027 | 1.115 | 6.46E-05 | 101,055 | 2,972 | 98,083 | 0.407 | Proteinuria | endocrine/metabolic |
| OSA | 110.1 | rs11642015 | T | 0.064 | 0.016 | 1.066 | 7.25E-05 | 81,511 | 9,630 | 71,881 | 0.407 | Dermatophytosis | infectious diseases |
| OSA | 580.31 | rs11642015 | T | 0.171 | 0.043 | 1.187 | 8.16E-05 | 70,696 | 1,173 | 69,523 | 0.403 | Nephritis and nephropathy in diseases classified elsewhere | genitourinary |
| OSA | 272.1 | rs11642015 | T | 0.045 | 0.012 | 1.046 | 8.99E-05 | 98,574 | 55,813 | 42,761 | 0.407 | Hyperlipidemia | endocrine/metabolic |
| OSA | 272 | rs11642015 | T | 0.045 | 0.012 | 1.046 | 9.20E-05 | 98,603 | 55,842 | 42,761 | 0.407 | Disorders of lipoid metabolism | endocrine/metabolic |
| OSA | 550.6 | rs11642015 | T | 0.146 | 0.037 | 1.157 | 9.45E-05 | 90,872 | 1,517 | 89,355 | 0.406 | Incisional hernia | digestive |
| OSA | 366.2 | rs11642015 | T | 0.064 | 0.016 | 1.066 | 9.63E-05 | 96,768 | 11,807 | 84,961 | 0.406 | Senile cataract | sense organs |
| OSA | 710.19 | rs11642015 | T | 0.195 | 0.050 | 1.215 | 1.06E-04 | 82,927 | 833 | 82,094 | 0.404 | Unspecified osteomyelitis | musculoskeletal |
| OSA | 362 | rs11642015 | T | 0.075 | 0.020 | 1.078 | 1.41E-04 | 93,491 | 6,472 | 87,019 | 0.406 | Other retinal disorders | sense organs |
| OSA | 110 | rs11642015 | T | 0.058 | 0.015 | 1.060 | 1.63E-04 | 82,449 | 10,568 | 71,881 | 0.407 | Dermatophytosis / Dermatomycosis | infectious diseases |
| OSA | 790.6 | rs11642015 | T | 0.060 | 0.016 | 1.062 | 1.77E-04 | 91,999 | 9,049 | 82,950 | 0.407 | Other abnormal blood chemistry | symptoms |
| OSA | 280 | rs11642015 | T | 0.061 | 0.016 | 1.063 | 1.83E-04 | 78,519 | 9,483 | 69,036 | 0.407 | Iron deficiency anemias | hematopoietic |
| OSA | 530.15 | rs11642015 | T | -0.378 | 0.101 | 0.685 | 1.84E-04 | 52,561 | 228 | 52,333 | 0.402 | Eosinophilic esophagitis | digestive |
| OSA | 251 | rs11642015 | T | 0.116 | 0.031 | 1.123 | 1.95E-04 | 92,889 | 2,233 | 90,656 | 0.406 | Other disorders of pancreatic internal secretion | endocrine/metabolic |
| OSA | 585.31 | rs11642015 | T | 0.131 | 0.035 | 1.139 | 2.10E-04 | 71,338 | 1,815 | 69,523 | 0.403 | Renal dialysis | genitourinary |
| OSA | 360 | rs11642015 | T | 0.336 | 0.092 | 1.400 | 2.38E-04 | 89,410 | 247 | 89,163 | 0.406 | Disorders of the globe | sense organs |
| OSA | 427.22 | rs11642015 | T | 0.099 | 0.027 | 1.104 | 2.62E-04 | 62,086 | 3,551 | 58,535 | 0.406 | Atrial flutter | circulatory system |
| OSA | 327.1 | rs11642015 | T | 0.092 | 0.025 | 1.096 | 2.67E-04 | 65,594 | 3,532 | 62,062 | 0.402 | Hypersomnia | neurological |
| OSA | 274.1 | rs11642015 | T | 0.081 | 0.022 | 1.084 | 2.69E-04 | 104,089 | 4,708 | 99,381 | 0.407 | Gout | endocrine/metabolic |
| OSA | 278.11 | rs12725199 | A | 0.071 | 0.013 | 1.074 | 9.01E-08 | 64,197 | 25,203 | 38,994 | 0.252 | Morbid obesity | endocrine/metabolic |
| OSA | 569.2 | rs12725199 | A | 0.139 | 0.028 | 1.150 | 5.00E-07 | 95,118 | 3,493 | 91,625 | 0.252 | Gastrointestinal complications | digestive |
| OSA | 278.1 | rs12725199 | A | 0.050 | 0.012 | 1.051 | 1.82E-05 | 81,472 | 42,478 | 38,994 | 0.252 | Obesity | endocrine/metabolic |
| OSA | 278 | rs12725199 | A | 0.047 | 0.011 | 1.048 | 2.21E-05 | 91,871 | 52,877 | 38,994 | 0.251 | Overweight, obesity and other hyperalimentation | endocrine/metabolic |
| OSA | 569 | rs12725199 | A | 0.102 | 0.025 | 1.107 | 4.49E-05 | 95,999 | 4,374 | 91,625 | 0.252 | Other disorders of intestine | digestive |
| OSA | 327.32 | rs12725199 | A | 0.060 | 0.015 | 1.062 | 4.73E-05 | 76,918 | 15,597 | 61,321 | 0.252 | Obstructive sleep apnea | neurological |

**Supplementary Table 13.** PheWAS results for suggestively significant SNP-PheCode associations. Abbreviations: SE - standard error, OR - odds ratio, EAF - effect allele frequency.

| Discovery Trait | PheCode | SNP | Effect Allele | Beta | SE | OR | P | N | Ncases | Ncontrols | EAF | Phecode Description | Category |
|---|---|---|---|---|---|---|---|---|---|---|---|---|---|
| OSA | 327.3 | rs12725199 | A | 0.056 | 0.014 | 1.058 | 5.19E-05 | 79,943 | 18,622 | 61,321 | 0.252 | Sleep apnea | neurological |
| OSA | 251.1 | rs12725199 | A | 0.140 | 0.035 | 1.150 | 5.83E-05 | 56,262 | 2,233 | 54,029 | 0.251 | Hypoglycemia | endocrine/metabolic |
| OSA | 251 | rs12725199 | A | 0.136 | 0.035 | 1.146 | 8.28E-05 | 91,819 | 2,214 | 89,605 | 0.251 | Other disorders of pancreatic internal secretion | endocrine/metabolic |
| OSA | 278.11 | rs1800437 | C | -0.100 | 0.014 | 0.905 | 4.25E-12 | 64,964 | 25,467 | 39,497 | 0.205 | Morbid obesity | endocrine/metabolic |
| OSA | 278.1 | rs1800437 | C | -0.069 | 0.012 | 0.934 | 2.59E-08 | 82,426 | 42,929 | 39,497 | 0.205 | Obesity | endocrine/metabolic |
| OSA | 278 | rs1800437 | C | -0.051 | 0.012 | 0.950 | 1.24E-05 | 92,937 | 53,440 | 39,497 | 0.206 | Overweight, obesity and other hyperalimentation | endocrine/metabolic |
| OSA | 427.21 | rs2958127 | A | 0.080 | 0.018 | 1.083 | 1.59E-05 | 67,177 | 9,837 | 57,340 | 0.374 | Atrial fibrillation | circulatory system |
| OSA | 327 | rs2958127 | A | 0.044 | 0.010 | 1.045 | 2.02E-05 | 92,775 | 31,969 | 60,806 | 0.373 | Sleep disorders | neurological |
| OSA | 427.2 | rs2958127 | A | 0.077 | 0.018 | 1.080 | 2.55E-05 | 67,561 | 10,221 | 57,340 | 0.374 | Atrial fibrillation and flutter | circulatory system |
| OSA | 327.3 | rs2958127 | A | 0.050 | 0.013 | 1.051 | 7.70E-05 | 79,270 | 18,464 | 60,806 | 0.372 | Sleep apnea | neurological |
| OSA | 327.32 | rs2958127 | A | 0.051 | 0.013 | 1.052 | 1.58E-04 | 76,262 | 15,456 | 60,806 | 0.372 | Obstructive sleep apnea | neurological |
| OSA | 456 | rs34592089 | A | 0.253 | 0.046 | 1.288 | 3.00E-08 | 75,262 | 4,235 | 71,027 | 0.057 | Chronic venous insufficiency [CVI] | circulatory system |
| OSA | 278.11 | rs34592089 | A | 0.123 | 0.024 | 1.131 | 4.48E-07 | 64,159 | 25,132 | 39,027 | 0.058 | Morbid obesity | endocrine/metabolic |
| OSA | 278.1 | rs34592089 | A | 0.097 | 0.021 | 1.102 | 5.67E-06 | 81,413 | 42,386 | 39,027 | 0.058 | Obesity | endocrine/metabolic |
| OSA | 278 | rs34592089 | A | 0.083 | 0.020 | 1.087 | 4.17E-05 | 91,796 | 52,769 | 39,027 | 0.058 | Overweight, obesity and other hyperalimentation | endocrine/metabolic |
| OSA | 781.2 | rs34592089 | A | 1.405 | 0.357 | 4.076 | 8.48E-05 | 94,909 | 25 | 94,884 | 0.058 | Abnormal posture | symptoms |
| OSA | 470 | rs34592089 | A | 0.245 | 0.065 | 1.277 | 1.60E-04 | 52,780 | 1,858 | 50,922 | 0.058 | Septal Deviations/Turbinate Hypertrophy | respiratory |
| OSA | 706.3 | rs34592089 | A | 0.670 | 0.179 | 1.954 | 1.75E-04 | 77,653 | 168 | 77,485 | 0.058 | Seborrhea | dermatologic |
| OSA | 338.2 | rs66474734 | TATA | 0.061 | 0.012 | 1.063 | 2.10E-07 | 83,968 | 20,367 | 63,601 | 0.398 | Chronic pain | neurological |
| OSA | 338 | rs66474734 | TATA | 0.055 | 0.011 | 1.057 | 8.24E-07 | 86,918 | 23,317 | 63,601 | 0.398 | Pain | neurological |
| OSA | 327.32 | rs66474734 | TATA | 0.060 | 0.013 | 1.062 | 6.22E-06 | 75,891 | 15,401 | 60,490 | 0.398 | Obstructive sleep apnea | neurological |
| OSA | 722 | rs66474734 | TATA | 0.054 | 0.012 | 1.056 | 1.35E-05 | 86,166 | 18,439 | 67,727 | 0.398 | Intervertebral disc disorders | musculoskeletal |
| OSA | 722.6 | rs66474734 | TATA | 0.064 | 0.015 | 1.066 | 1.39E-05 | 79,659 | 11,932 | 67,727 | 0.398 | Degeneration of intervertebral disc | musculoskeletal |
| OSA | 278.11 | rs66474734 | TATA | 0.047 | 0.012 | 1.048 | 8.21E-05 | 63,300 | 24,852 | 38,448 | 0.397 | Morbid obesity | endocrine/metabolic |
| OSA | 278.1 | rs66474734 | TATA | 0.041 | 0.010 | 1.041 | 8.88E-05 | 80,358 | 41,910 | 38,448 | 0.397 | Obesity | endocrine/metabolic |
| OSA | 327.3 | rs66474734 | TATA | 0.049 | 0.012 | 1.050 | 8.99E-05 | 78,880 | 18,390 | 60,490 | 0.397 | Sleep apnea | neurological |
| OSA | 214 | rs66474734 | TATA | 0.120 | 0.033 | 1.128 | 2.26E-04 | 99,490 | 2,003 | 97,487 | 0.398 | Lipoma | neoplasms |
| OSA | 250 | rs9380814 | A | -0.040 | 0.011 | 0.961 | 1.98E-04 | 97,568 | 36,422 | 61,146 | 0.323 | Diabetes mellitus | endocrine/metabolic |
| OSA | 278.11 | rs9831061 | G | -0.070 | 0.013 | 0.932 | 1.46E-07 | 63,852 | 25,068 | 38,784 | 0.261 | Morbid obesity | endocrine/metabolic |
| OSA | 278.1 | rs9831061 | G | -0.056 | 0.011 | 0.945 | 8.78E-07 | 80,996 | 42,212 | 38,784 | 0.261 | Obesity | endocrine/metabolic |
| OSA | 569 | rs9831061 | G | -0.114 | 0.026 | 0.892 | 1.20E-05 | 95,399 | 4,324 | 91,075 | 0.262 | Other disorders of intestine | digestive |
| OSA | 569.2 | rs9831061 | G | -0.125 | 0.029 | 0.883 | 1.91E-05 | 94,524 | 3,449 | 91,075 | 0.262 | Gastrointestinal complications | digestive |
| OSA | 401 | rs9831061 | G | -0.048 | 0.012 | 0.953 | 1.23E-04 | 97,961 | 53,053 | 44,908 | 0.262 | Hypertension | circulatory system |
| OSA | 401.1 | rs9831061 | G | -0.048 | 0.013 | 0.953 | 1.33E-04 | 97,231 | 52,323 | 44,908 | 0.262 | Essential hypertension | circulatory system |
| OSA | 278 | rs9831061 | G | -0.041 | 0.011 | 0.960 | 1.96E-04 | 91,328 | 52,544 | 38,784 | 0.262 | Overweight, obesity and other hyperalimentation | endocrine/metabolic |
| OSAadjBMI | 809 | rs10735928 | A | 0.075 | 0.019 | 1.077 | 1.10E-04 | 86,170 | 6,159 | 80,011 | 0.382 | Fracture of unspecified bones | injuries & poisonings |
| OSAadjBMI | 427.21 | rs4759254 | C | 0.086 | 0.019 | 1.090 | 5.93E-06 | 65,277 | 9,565 | 55,712 | 0.354 | Atrial fibrillation | circulatory system |
| OSAadjBMI | 427.2 | rs4759254 | C | 0.083 | 0.019 | 1.087 | 9.88E-06 | 65,648 | 9,936 | 55,712 | 0.353 | Atrial fibrillation and flutter | circulatory system |
| OSAadjBMI | 327 | rs4759254 | C | 0.043 | 0.011 | 1.043 | 5.78E-05 | 90,154 | 31,072 | 59,082 | 0.352 | Sleep disorders | neurological |
| OSAadjBMI | 244.2 | rs4759254 | C | 0.076 | 0.020 | 1.079 | 1.21E-04 | 78,097 | 6,378 | 71,719 | 0.353 | Acquired hypothyroidism | endocrine/metabolic |
| OSAadjBMI | 327.3 | rs4759254 | C | 0.049 | 0.013 | 1.050 | 1.49E-04 | 77,031 | 17,949 | 59,082 | 0.352 | Sleep apnea | neurological |
| OSAadjBMI | 244 | rs4759254 | C | 0.048 | 0.013 | 1.049 | 1.92E-04 | 90,338 | 18,619 | 71,719 | 0.352 | Hypothyroidism | endocrine/metabolic |
| OSAadjBMI | 286.13 | rs60791051 | C | 1.340 | 0.342 | 3.819 | 8.84E-05 | 87,438 | 26 | 87,412 | 0.062 | Congenital factor VIII disorder | hematopoietic |

**Supplementary Table 14.** Gene-based association results from MAGMA analysis. NSNPs - number of SNPs in the genomic region; NPARAM - number of SNPs included in the gene-based test.

| Strata | GENE | CHR | START | STOP | NSNPS | NPARAM | N | ZSTAT | P | SYMBOL |
|---|---|---|---|---|---|---|---|---|---|---|
| **OSA** | | | | | | | | | | |
| Multi-Population | ENSG00000117569 | 1 | 97177221 | 97299294 | 563 | 62 | 275,676 | 5.4317 | 2.79E-08 | *PTBP2* |
| European-only | ENSG00000117569 | 1 | 97177221 | 97299294 | 441 | 46 | 293,552 | 5.1634 | 1.21E-07 | *PTBP2* |
| Multi-Population | ENSG00000244405 | 3 | 185754097 | 185838107 | 278 | 46 | 315,329 | 4.927 | 4.17E-07 | *ETV5* |
| European-only | ENSG00000244405 | 3 | 185754097 | 185838107 | 198 | 29 | 369,528 | 4.9452 | 3.80E-07 | *ETV5* |
| European-only | ENSG00000130413 | 11 | 8403418 | 8625836 | 959 | 67 | 352,193 | 4.9377 | 3.95E-07 | *STK33* |
| Multi-Population | ENSG00000166436 | 11 | 8623584 | 8703413 | 417 | 60 | 296,769 | 4.7176 | 1.19E-06 | *TRIM66* |
| European-only | ENSG00000166436 | 11 | 8623584 | 8703413 | 290 | 40 | 357,870 | 5.008 | 2.75E-07 | *TRIM66* |
| Multi-Population | ENSG00000076108 | 12 | 56979380 | 57040600 | 179 | 39 | 286,514 | 4.8695 | 5.59E-07 | *BAZ2A* |
| European-only | ENSG00000076108 | 12 | 56979380 | 57040600 | 125 | 31 | 349,528 | 4.7071 | 1.26E-06 | *BAZ2A* |
| Multi-Population | ENSG00000110955 | 12 | 57021959 | 57049798 | 88 | 22 | 324,184 | 5.9747 | 1.15E-09 | *ATP5B* |
| European-only | ENSG00000110955 | 12 | 57021959 | 57049798 | 72 | 17 | 348,604 | 5.2562 | 7.35E-08 | *ATP5B* |
| Multi-Population | ENSG00000110958 | 12 | 57047127 | 57092159 | 190 | 34 | 297,023 | 5.4742 | 2.20E-08 | *PTGES3* |
| European-only | ENSG00000110958 | 12 | 57047127 | 57092159 | 161 | 27 | 318,641 | 4.8578 | 5.94E-07 | *PTGES3* |
| Multi-Population | ENSG00000196531 | 12 | 57096212 | 57135412 | 126 | 25 | 313,652 | 6.1094 | 5.00E-10 | *NACA* |
| European-only | ENSG00000196531 | 12 | 57096212 | 57135412 | 99 | 17 | 343,451 | 5.2105 | 9.42E-08 | *NACA* |
| Multi-Population | ENSG00000198056 | 12 | 57115380 | 57156157 | 113 | 20 | 285,058 | 5.5983 | 1.08E-08 | *PRIM1* |
| European-only | ENSG00000198056 | 12 | 57115380 | 57156157 | 88 | 14 | 311,235 | 4.9629 | 3.47E-07 | *PRIM1* |
| Multi-Population | ENSG00000025423 | 12 | 57135945 | 57191574 | 205 | 31 | 251,169 | 5.494 | 1.97E-08 | *HSD17B6* |
| European-only | ENSG00000025423 | 12 | 57135945 | 57191574 | 170 | 24 | 268,002 | 4.9075 | 4.61E-07 | *HSD17B6* |
| Multi-Population | ENSG00000140718 | 16 | 53727875 | 54165853 | 2578 | 267 | 309,824 | 4.8303 | 6.82E-07 | *FTO* |
| European-only | ENSG00000140718 | 16 | 53727875 | 54165853 | 1874 | 220 | 359,706 | 5.3678 | 3.98E-08 | *FTO* |
| European-only | ENSG00000197381 | 21 | 46483768 | 46656478 | 696 | 79 | 370,838 | 4.615 | 1.97E-06 | *ADARB1* |
| **OSAadjBMI** | | | | | | | | | | |
| European-only | ENSG00000112282 | 6 | 131885106 | 131959369 | 153 | 36 | 179,626 | 4.969 | 3.37E-07 | *MED23* |
| European-only | ENSG00000076108 | 12 | 56979380 | 57040600 | 125 | 31 | 188,596 | 4.6103 | 2.01E-06 | *BAZ2A* |
| Multi-Population | ENSG00000110955 | 12 | 57021959 | 57049798 | 88 | 22 | 185,315 | 5.5857 | 1.16E-08 | *ATP5B* |
| European-only | ENSG00000110955 | 12 | 57021959 | 57049798 | 72 | 17 | 190,294 | 5.3454 | 4.51E-08 | *ATP5B* |
| Multi-Population | ENSG00000110958 | 12 | 57047127 | 57092159 | 190 | 34 | 170,732 | 5.2137 | 9.26E-08 | *PTGES3* |
| European-only | ENSG00000110958 | 12 | 57047127 | 57092159 | 162 | 28 | 175,267 | 5.0321 | 2.43E-07 | *PTGES3* |
| Multi-Population | ENSG00000196531 | 12 | 57096212 | 57135412 | 124 | 24 | 177,815 | 6.1094 | 5.00E-10 | *NACA* |
| European-only | ENSG00000196531 | 12 | 57096212 | 57135412 | 99 | 17 | 183,703 | 5.4424 | 2.63E-08 | *NACA* |
| Multi-Population | ENSG00000198056 | 12 | 57115380 | 57156157 | 113 | 20 | 168,921 | 5.3778 | 3.77E-08 | *PRIM1* |
| European-only | ENSG00000198056 | 12 | 57115380 | 57156157 | 88 | 14 | 176,882 | 5.2226 | 8.82E-08 | *PRIM1* |
| Multi-Population | ENSG00000025423 | 12 | 57135945 | 57191574 | 205 | 31 | 154,131 | 5.0628 | 2.07E-07 | *HSD17B6* |
| European-only | ENSG00000025423 | 12 | 57135945 | 57191574 | 170 | 24 | 157,855 | 4.8937 | 4.95E-07 | *HSD17B6* |
| Multi-Population | ENSG00000174099 | 12 | 65662423 | 65892024 | 1000 | 101 | 158,305 | 5.463 | 2.34E-08 | *MSRB3* |
| European-only | ENSG00000174099 | 12 | 65662423 | 65892024 | 549 | 58 | 194,891 | 6.1094 | 5.00E-10 | *MSRB3* |

**Supplementary Table 15.** Variant annotation for fine-mapped variants with highest posterior probability.

| Trait | Locus Index Variant | Fine-mapped variant | CHR | POS (hg37) | Location (hg38) | Allele | Consequence | IMPACT | SYMBOL | Gene | Feature_type | Feature | BIOTYPE | EXON | INTRON | HGVSc | HGVSp | cDNA_position | CDS_position | Protein_position | Amino_acids | Codons |
|---|---|---|---|---|---|---|---|---|---|---|---|---|---|---|---|---|---|---|---|---|---|---|
| AHI | rs1200596362 | rs201670852 | 2 | 196329529 | 2:195464805-195464805 | G | intron_variant,non_coding_transcript_variant | MODIFIER | - | ENSG00000271893 | Transcript | ENST00000606986.1 | lncRNA | - | 2/2 | ENST00000606986.1:n.360-12884A>C | - | - | - | - | - | - |
| AHI | rs1200596362 | rs201670852 | 2 | 196329529 | 2:195464805-195464805 | G | intron_variant,non_coding_transcript_variant | MODIFIER | - | ENSG00000224099 | Transcript | ENST00000762253.1 | lncRNA | - | 3/4 | ENST00000762253.1:n.415+6542T>G | - | - | - | - | - | - |
| AHI | rs1200596362 | rs1553539108 | 2 | 196333418 | 2:195468695-195468695 | A | intron_variant,non_coding_transcript_variant | MODIFIER | - | ENSG00000271893 | Transcript | ENST00000606986.1 | lncRNA | - | 2/2 | ENST00000606986.1:n.360-16774dup | - | - | - | - | - | - |
| AHI | rs1200596362 | rs1553539108 | 2 | 196333418 | 2:195468695-195468695 | A | intron_variant,non_coding_transcript_variant | MODIFIER | - | ENSG00000224099 | Transcript | ENST00000762253.1 | lncRNA | - | 3/4 | ENST00000762253.1:n.416-9643dup | - | - | - | - | - | - |
| AHI | rs1200596362 | rs1200596362 | 2 | 196364827 | 2:195500102-195500103 | - | intron_variant,non_coding_transcript_variant | MODIFIER | - | ENSG00000271893 | Transcript | ENST00000606986.1 | lncRNA | - | 1/2 | ENST00000606986.1:n.264-13323del | - | - | - | - | - | - |
| AHI | rs1200596362 | rs13015934 | 2 | 196369621 | 2:195504897-195504897 | T | intron_variant,non_coding_transcript_variant | MODIFIER | - | ENSG00000271893 | Transcript | ENST00000606986.1 | lncRNA | - | 1/2 | ENST00000606986.1:n.263+10540G>A | - | - | - | - | - | - |
| AHI | rs1200596362 | rs903825330 | 2 | 196433941 | 2:195569217-195569217 | A | downstream_gene_variant | MODIFIER | LINC01827 | ENSG00000234919 | Transcript | ENST00000809472.1 | lncRNA | - | - | - | - | - | - | - | - | - |
| AHIadjBMI | rs78545995 | rs78545995 | 6 | 65804913 | 6:65095020-65095020 | G | intron_variant | MODIFIER | EYS | ENSG00000188107 | Transcript | ENST00000503581.6 | protein_coding | - | 12/42 | ENST00000503581.6:c.2024-37293T>C | - | - | - | - | - | - |
| AHIadjBMI | rs78545995 | rs140715014 | 6 | 65821376 | 6:65111483-65111483 | G | intron_variant | MODIFIER | EYS | ENSG00000188107 | Transcript | ENST00000503581.6 | protein_coding | - | 12/42 | ENST00000503581.6:c.2024-53756T>C | - | - | - | - | - | - |
| AHIadjBMI | rs78545995 | rs75433897 | 6 | 65838774 | 6:65128881-65128881 | A | intron_variant | MODIFIER | EYS | ENSG00000188107 | Transcript | ENST00000503581.6 | protein_coding | - | 12/42 | ENST00000503581.6:c.2024-71154A>T | - | - | - | - | - | - |
| AHIadjBMI | rs78545995 | rs149112115 | 6 | 65934775 | 6:65224882-65224882 | T | intron_variant | MODIFIER | EYS | ENSG00000188107 | Transcript | ENST00000503581.6 | protein_coding | - | 12/42 | ENST00000503581.6:c.2023+70981C>A | - | - | - | - | - | - |
| avgSaO2 | rs802729 | rs802729 | 6 | 128280360 | HSCHR6_1_CTG8:304413-304413 | A | intergenic_variant | MODIFIER | - | - | - | - | - | - | - | - | - | - | - | - | - | - |
| avgSaO2 | rs802729 | rs802729 | 6 | 128280360 | 6:127959215-127959215 | A | intergenic_variant | MODIFIER | - | - | - | - | - | - | - | - | - | - | - | - | - | - |
| avgSaO2 | rs802729 | rs117749759 | 6 | 128352791 | HSCHR6_1_CTG8:380980-380980 | C | intron_variant | MODIFIER | PTPRK | ENSG00000273993 | Transcript | ENST00000626594.2 | protein_coding | - | 13/33 | ENST00000626594.2:c.2195-22379T>G | - | - | - | - | - | - |
| avgSaO2 | rs802729 | rs117749759 | 6 | 128352791 | HSCHR6_1_CTG8:380980-380980 | C | intron_variant,non_coding_transcript_variant | MODIFIER | PTPRK-AS1 | ENSG00000280612 | Transcript | ENST00000627470.1 | lncRNA | - | 1/4 | ENST00000627470.1:n.38+3723A>C | - | - | - | - | - | - |
| avgSaO2 | rs802729 | rs117749759 | 6 | 128352791 | 6:128031646-128031646 | C | intron_variant,non_coding_transcript_variant | MODIFIER | PTPRK-AS1 | ENSG00000227945 | Transcript | ENST00000417390.1 | lncRNA | - | 1/4 | ENST00000417390.1:n.38+3723A>C | - | - | - | - | - | - |
| avgSaO2 | rs802729 | rs117749759 | 6 | 128352791 | 6:128031646-128031646 | C | intron_variant | MODIFIER | PTPRK | ENSG00000152894 | Transcript | ENST00000368226.9 | protein_coding | - | 13/29 | ENST00000368226.9:c.2195-22378T>G | - | - | - | - | - | - |
| avgSaO2 | rs2194955 | rs4739235 | 8 | 21287105 | HG2068_PATCH:34085-34085 | A | intergenic_variant | MODIFIER | - | - | - | - | - | - | - | - | - | - | - | - | - | - |
| avgSaO2 | rs2194955 | rs4739235 | 8 | 21287105 | 8:21429594-21429594 | A | intron_variant,non_coding_transcript_variant | MODIFIER | - | ENSG00000305102 | Transcript | ENST00000808781.1 | lncRNA | - | 2/2 | ENST00000808781.1:n.317+18739C>T | - | - | - | - | - | - |
| avgSaO2 | rs2194955 | rs2194955 | 8 | 21292558 | HG2068_PATCH:39538-39538 | C | intergenic_variant | MODIFIER | - | - | - | - | - | - | - | - | - | - | - | - | - | - |
| avgSaO2 | rs2194955 | rs2194955 | 8 | 21292558 | 8:21435047-21435047 | C | intron_variant,non_coding_transcript_variant | MODIFIER | - | ENSG00000305102 | Transcript | ENST00000808781.1 | lncRNA | - | 2/2 | ENST00000808781.1:n.317+13286T>G | - | - | - | - | - | - |
| avgSaO2 | rs2194955 | rs1549471 | 8 | 21298827 | HG2068_PATCH:45807-45807 | A | intergenic_variant | MODIFIER | - | - | - | - | - | - | - | - | - | - | - | - | - | - |
| avgSaO2 | rs2194955 | rs1549471 | 8 | 21298827 | 8:21441316-21441316 | A | intron_variant,non_coding_transcript_variant | MODIFIER | - | ENSG00000305102 | Transcript | ENST00000808781.1 | lncRNA | - | 2/2 | ENST00000808781.1:n.317+7017C>T | - | - | - | - | - | - |
| avgSaO2 | rs2194955 | rs1549471 | 8 | 21298827 | 8:21441316-21441316 | A | upstream_gene_variant | MODIFIER | - | ENSG00000305123 | Transcript | ENST00000808905.1 | lncRNA | - | - | - | - | - | - | - | - | - |
| avgSaO2 | rs2194955 | rs11984977 | 8 | 21312389 | HG2068_PATCH:59369-59369 | G | intergenic_variant | MODIFIER | - | - | - | - | - | - | - | - | - | - | - | - | - | - |
| avgSaO2 | rs2194955 | rs11984977 | 8 | 21312389 | 8:21454878-21454878 | G | upstream_gene_variant | MODIFIER | - | ENSG00000305102 | Transcript | ENST00000808781.1 | lncRNA | - | - | - | - | - | - | - | - | - |
| avgSaO2 | rs2194955 | rs4739247 | 8 | 21315374 | HG2068_PATCH:62354-62354 | G | intergenic_variant | MODIFIER | - | - | - | - | - | - | - | - | - | - | - | - | - | - |
| avgSaO2 | rs2194955 | rs4739247 | 8 | 21315374 | 8:21457863-21457863 | G | intergenic_variant | MODIFIER | - | - | - | - | - | - | - | - | - | - | - | - | - | - |
| avgSaO2adjBMI | rs79914264 | rs79914264 | 1 | 29685027 | 1:29358515-29358515 | A | intergenic_variant | MODIFIER | - | - | - | - | - | - | - | - | - | - | - | - | - | - |
| OSA | rs12725199 | rs55789411 | 1 | 97110637 | 1:96645081-96645081 | T | intergenic_variant | MODIFIER | - | - | - | - | - | - | - | - | - | - | - | - | - | - |
| OSA | rs12725199 | rs12567921 | 1 | 97111780 | 1:96646224-96646224 | T | intergenic_variant | MODIFIER | - | - | - | - | - | - | - | - | - | - | - | - | - | - |
| OSA | rs12725199 | rs35696197 | 1 | 97287825 | 1:96822269-96822269 | T | 3_prime_UTR_variant | MODIFIER | PTBP2 | ENSG00000117569 | Transcript | ENST00000609116.5 | protein_coding | 14/14 | - | ENST00000609116.5:c.*8864G>T | - | 10545 | - | - | - | - |
| OSA | rs12725199 | rs12725199 | 1 | 97293792 | 1:96828236-96828236 | A | downstream_gene_variant | MODIFIER | PTBP2 | ENSG00000117569 | Transcript | ENST00000609116.5 | protein_coding | - | - | - | - | - | - | - | - | - |
| OSA | rs12725199 | rs12740489 | 1 | 97296935 | 1:96831379-96831379 | A | intergenic_variant | MODIFIER | - | - | - | - | - | - | - | - | - | - | - | - | - | - |
| OSA | rs2376073 | rs12467202 | 2 | 104601968 | 2:103985510-103985510 | C | intron_variant,non_coding_transcript_variant | MODIFIER | LINC01965 | ENSG00000256637 | Transcript | ENST00000688553.2 | lncRNA | - | 1/2 | ENST00000688553.2:n.294-67994T>C | - | - | - | - | - | - |
| OSA | rs2376073 | rs35481561 | 2 | 104622478 | 2:104006020-104006020 | T | intron_variant,non_coding_transcript_variant | MODIFIER | LINC01965 | ENSG00000256637 | Transcript | ENST00000688553.2 | lncRNA | - | 1/2 | ENST00000688553.2:n.294-47484C>T | - | - | - | - | - | - |
| OSA | rs2376073 | rs2376073 | 2 | 104662008 | 2:104045550-104045550 | T | intron_variant,non_coding_transcript_variant | MODIFIER | LINC01965 | ENSG00000256637 | Transcript | ENST00000688553.2 | lncRNA | - | 1/2 | ENST00000688553.2:n.294-7954C>T | - | - | - | - | - | - |
| OSA | rs9831061 | rs1410233719 | 3 | 18582602 | 3:18582602-18582602 | C | intron_variant,non_coding_transcript_variant | MODIFIER | SATB1-AS1 | ENSG00000228956 | Transcript | ENST00000683051.1 | lncRNA | - | 1/3 | ENST00000683051.1:n.120+55276A>C | - | - | - | - | - | - |
| OSA | rs9831061 | rs869399 | 3 | 185826764 | 3:186108975-186108975 | T | intron_variant,non_coding_transcript_variant | MODIFIER | DGKG | ENSG00000058866 | Transcript | ENST00000447054.5 | protein_coding_CDS_not_defined | - | 2/3 | ENST00000447054.5:n.132-3033G>A | - | - | - | - | - | - |
| OSA | rs9831061 | rs869399 | 3 | 185826764 | 3:186108975-186108975 | T | regulatory_region_variant | MODIFIER | - | - | RegulatoryFeature | ENSR3_B3CX8H | promoter | - | - | - | - | - | - | - | - | - |
| OSA | rs9831061 | rs869399 | 3 | 185826764 | 3:186108975-186108975 | T | 5_prime_UTR_variant | MODIFIER | ETV5 | ENSG00000244405 | Transcript | ENST00000306376.10 | protein_coding | 1/13 | - | ENST00000306376.10:c.-110G>A | - | 115 | - | - | - | - |
| OSA | rs9831061 | rs1516728 | 3 | 185829891 | 3:186112102-186112102 | A | upstream_gene_variant | MODIFIER | ETV5 | ENSG00000244405 | Transcript | ENST00000306376.10 | protein_coding | - | - | - | - | - | - | - | - | - |
| OSA | rs9831061 | rs1516728 | 3 | 185829891 | 3:186112102-186112102 | A | intron_variant,non_coding_transcript_variant | MODIFIER | DGKG | ENSG00000058866 | Transcript | ENST00000447054.5 | protein_coding_CDS_not_defined | - | 2/3 | ENST00000447054.5:n.132-6160A>T | - | - | - | - | - | - |
| OSA | rs9831061 | rs7635103 | 3 | 185833759 | 3:186115970-186115970 | A | intron_variant,non_coding_transcript_variant | MODIFIER | DGKG | ENSG00000058866 | Transcript | ENST00000447054.5 | protein_coding_CDS_not_defined | - | 2/3 | ENST00000447054.5:n.132-10028G>T | - | - | - | - | - | - |
| OSA | rs9831061 | rs9831061 | 3 | 185835281 | 3:186117492-186117492 | A | intron_variant,non_coding_transcript_variant | MODIFIER | DGKG | ENSG00000058866 | Transcript | ENST00000447054.5 | protein_coding_CDS_not_defined | - | 2/3 | ENST00000447054.5:n.132-11550C>T | - | - | - | - | - | - |
| OSA | rs9831061 | rs9831061 | 3 | 185835281 | 3:186117492-186117492 | A | downstream_gene_variant | MODIFIER | - | ENSG00000305961 | Transcript | ENST00000814422.1 | lncRNA | - | - | - | - | - | - | - | - | - |
| OSA | rs34592089 | rs34592089 | 4 | 102926923 | 4:102005766-102005766 | A | intron_variant | MODIFIER | BANK1 | ENSG00000153064 | Transcript | ENST00000322953.9 | protein_coding | - | 7/16 | ENST00000322953.9:c.1207-15748G>A | - | - | - | - | - | - |

**Supplementary Table 15.** Variant annotation for fine-mapped variants with highest posterior probability.

| Trait | Locus Index Variant | Fine-mapped variant | CHR | POS (hg37) | Location (hg38) | Allele | Consequence | IMPACT | SYMBOL | Gene | Feature_type | Feature | BIOTYPE | EXON | INTRON | HGVSc | HGVSp | cDNA_position | CDS_position | Protein_position | Amino_acids | Codons |
|---|---|---|---|---|---|---|---|---|---|---|---|---|---|---|---|---|---|---|---|---|---|---|
| OSA | rs34592089 | rs34592089 | 4 | 4:102926923 | 4:102005766-102005766 | A | intron_variant,non_coding_transcript_variant | MODIFIER | - | ENSG00000251309 | Transcript | ENST00000505091.1 | lncRNA | - | 2/2 | ENST00000505091.1:n.396+26443C>T | - | - | - | - | - | - |
| OSA | rs9380814 | rs72853773 | 6 | 38928910 | 6:38961134-38961134 | G | intron_variant | MODIFIER | DNAH8 | ENSG00000124721 | Transcript | ENST00000327475.11 | protein_coding | - | 82/92 | ENST00000327475.11:c.12451+9614A>G | - | - | - | - | - | - |
| OSA | rs9380814 | rs9380812 | 6 | 38930402 | 6:38962626-38962626 | T | intron_variant | MODIFIER | DNAH8 | ENSG00000124721 | Transcript | ENST00000327475.11 | protein_coding | - | 82/92 | ENST00000327475.11:c.12452-8966C>T | - | - | - | - | - | - |
| OSA | rs9380814 | rs9380814 | 6 | 38934922 | 6:38967146-38967146 | A | intron_variant | MODIFIER | DNAH8 | ENSG00000124721 | Transcript | ENST00000327475.11 | protein_coding | - | 82/92 | ENST00000327475.11:c.12452-4446G>A | - | - | - | - | - | - |
| OSA | rs9380814 | rs9366987 | 6 | 38936096 | 6:38968320-38968320 | C | intron_variant | MODIFIER | DNAH8 | ENSG00000124721 | Transcript | ENST00000327475.11 | protein_coding | - | 82/92 | ENST00000327475.11:c.12452-3272T>C | - | - | - | - | - | - |
| OSA | rs9380814 | rs1537230 | 6 | 38944196 | 6:38976420-38976420 | A | intron_variant | MODIFIER | DNAH8 | ENSG00000124721 | Transcript | ENST00000327475.11 | protein_coding | - | 85/92 | ENST00000327475.11:c.12834+1891G>A | - | - | - | - | - | - |
| OSA | rs66474734 | rs74818520 | 8 | 114633079 | 8:113620850-113620850 | T | downstream_gene_variant | MODIFIER | - | ENSG00000253207 | Transcript | ENST00000521500.2 | lncRNA | - | - | - | - | - | - | - | - | - |
| OSA | rs66474734 | rs6469468 | 8 | 114647559 | 8:113635330-113635330 | C | intergenic_variant | MODIFIER | - | - | - | - | - | - | - | - | - | - | - | - | - | - |
| OSA | rs66474734 | rs774904698 | 8 | 114650056 | 8:113637826-113637827 | - | intergenic_variant | MODIFIER | - | - | - | - | - | - | - | - | - | - | - | - | - | - |
| OSA | rs66474734 | rs1247136109 | 8 | 114652051 | 8:114652051-114652051 | C | intron_variant,non_coding_transcript_variant | MODIFIER | - | ENSG00000302453 | Transcript | ENST00000786914.1 | lncRNA | - | 2/2 | ENST00000786914.1:n.217+3614T>C | - | - | - | - | - | - |
| OSA | rs66474734 | rs66474734 | 8 | 114661935 | 8:113649707-113649707 | ATA | intergenic_variant | MODIFIER | - | - | - | - | - | - | - | - | - | - | - | - | - | - |
| OSA | rs12553324 | rs11794152 | 9 | 23345347 | 9:23345349-23345349 | G | intron_variant,non_coding_transcript_variant | MODIFIER | - | ENSG00000284418 | Transcript | ENST00000640003.1 | lncRNA | - | 1/9 | ENST00000640003.1:n.140-81012T>C | - | - | - | - | - | - |
| OSA | rs12553324 | rs10429537 | 9 | 23346842 | 9:23346844-23346844 | G | intron_variant,non_coding_transcript_variant | MODIFIER | - | ENSG00000284418 | Transcript | ENST00000640003.1 | lncRNA | - | 1/9 | ENST00000640003.1:n.140-82507G>C | - | - | - | - | - | - |
| OSA | rs12553324 | rs12553324 | 9 | 23347865 | 9:23347867-23347867 | G | intron_variant,non_coding_transcript_variant | MODIFIER | - | ENSG00000284418 | Transcript | ENST00000640003.1 | lncRNA | - | 1/9 | ENST00000640003.1:n.140-83530G>C | - | - | - | - | - | - |
| OSA | rs12553324 | rs4977836 | 9 | 23355664 | 9:23355666-23355666 | A | intron_variant,non_coding_transcript_variant | MODIFIER | - | ENSG00000284418 | Transcript | ENST00000640003.1 | lncRNA | - | 1/9 | ENST00000640003.1:n.139+82896G>T | - | - | - | - | - | - |
| OSA | rs12553324 | rs1590949 | 9 | 23360417 | 9:23360419-23360419 | G | intron_variant,non_coding_transcript_variant | MODIFIER | - | ENSG00000284418 | Transcript | ENST00000640003.1 | lncRNA | - | 1/9 | ENST00000640003.1:n.139+78143G>C | - | - | - | - | - | - |
| OSA | rs2958127 | rs2035081 | 12 | 57125879 | 12:56732095-56732095 | G | intron_variant | MODIFIER | PRIM1 | ENSG00000198056 | Transcript | ENST00000338193.11 | protein_coding | - | 12/12 | ENST00000338193.11:c.1244-361T>C | - | - | - | - | - | - |
| OSA | rs11642015 | rs1421085 | 16 | 53800954 | 16:53767042-53767042 | C | intron_variant | MODIFIER | FTO | ENSG00000140718 | Transcript | ENST00000471389.6 | protein_coding | - | 1/8 | ENST00000471389.6:c.46-43098T>C | - | - | - | - | - | - |
| OSA | rs11642015 | rs11642015 | 16 | 53802494 | 16:53768582-53768582 | T | intron_variant | MODIFIER | FTO | ENSG00000140718 | Transcript | ENST00000471389.6 | protein_coding | - | 1/8 | ENST00000471389.6:c.46-41558C>T | - | - | - | - | - | - |
| OSA | rs11642015 | rs62048402 | 16 | 53803223 | 16:53769311-53769311 | A | intron_variant | MODIFIER | FTO | ENSG00000140718 | Transcript | ENST00000471389.6 | protein_coding | - | 1/8 | ENST00000471389.6:c.46-40829G>A | - | - | - | - | - | - |

**Supplementary Table 15.** Variant annotation for fine-mapped variants with highest posterior probability.

| Trait | Locus Index Variant | Fine-mapped variant | CHR | POS (hg37) | Location (hg38) | Allele | Consequence | IMPACT | SYMBOL | Gene | Feature_type | Feature | BIOTYPE | EXON | INTRON | HGVSc | HGVSp | cDNA_position | CDS_position | Protein_position | Amino_acids | Codons |
|---|---|---|---|---|---|---|---|---|---|---|---|---|---|---|---|---|---|---|---|---|---|---|
| OSA | rs11642015 | rs56094641 | 16 | 53806453 | 16:53772541-53772541 | G | intron_variant | MODIFIER | FTO | ENSG00000140718 | Transcript | ENST00000471389.6 | protein_coding | - | 1/8 | ENST00000471389.6:c.46-37599A>G | - | - | - | - | - | - |
| OSA | rs11642015 | rs55872725 | 16 | 53809123 | 16:53775211-53775211 | T | intron_variant | MODIFIER | FTO | ENSG00000140718 | Transcript | ENST00000471389.6 | protein_coding | - | 1/8 | ENST00000471389.6:c.46-34929C>T | - | - | - | - | - | - |
| OSA | rs1800437 | rs2238691 | 19 | 46179043 | 19:45675785-45675785 | A | downstream_gene_variant | MODIFIER | MIR642A | ENSG00000207773 | Transcript | ENST00000385039.1 | miRNA | - | - | - | - | - | - | - | - | - |
| OSA | rs1800437 | rs2238691 | 19 | 46179043 | 19:45675785-45675785 | A | upstream_gene_variant | MODIFIER | MIR642B | ENSG00000283212 | Transcript | ENST00000637346.1 | miRNA | - | - | - | - | - | - | - | - | - |
| OSA | rs1800437 | rs2238691 | 19 | 46179043 | 19:45675785-45675785 | A | intron_variant | MODIFIER | GIPR | ENSG00000010310 | Transcript | ENST00000590918.6 | protein_coding | - | 7/13 | ENST00000590918.6:c.633+959G>A | - | - | - | - | - | - |
| OSA | rs1800437 | rs11672660 | 19 | 46180184 | 19:45676926-45676926 | T | downstream_gene_variant | MODIFIER | MIR642A | ENSG00000207773 | Transcript | ENST00000385039.1 | miRNA | - | - | - | - | - | - | - | - | - |
| OSA | rs1800437 | rs11672660 | 19 | 46180184 | 19:45676926-45676926 | T | upstream_gene_variant | MODIFIER | MIR642B | ENSG00000283212 | Transcript | ENST00000637346.1 | miRNA | - | - | - | - | - | - | - | - | - |
| OSA | rs1800437 | rs11672660 | 19 | 46180184 | 19:45676926-45676926 | T | intron_variant | MODIFIER | GIPR | ENSG00000010310 | Transcript | ENST00000590918.6 | protein_coding | - | 7/13 | ENST00000590918.6:c.634-23C>T | - | - | - | - | - | - |
| OSA | rs1800437 | rs34783010 | 19 | 46180414 | 19:45677156-45677156 | T | downstream_gene_variant | MODIFIER | MIR642A | ENSG00000207773 | Transcript | ENST00000385039.1 | miRNA | - | - | - | - | - | - | - | - | - |

**Supplementary Table 15.** Variant annotation for fine-mapped variants with highest posterior probability.

| Trait | Locus Index Variant | Fine-mapped variant | CHR | POS (hg37) | Location (hg38) | Allele | Consequence | IMPACT | SYMBOL | Gene | Feature_type | Feature | BIOTYPE | EXON | INTRON | HGVSc | HGVSp | cDNA_position | CDS_position | Protein_position | Amino_acids | Codons |
|---|---|---|---|---|---|---|---|---|---|---|---|---|---|---|---|---|---|---|---|---|---|---|
| OSA | rs1800437 | rs34783010 | 19 | 46180414 | 19:45677156-45677156 | T | upstream_gene_variant | MODIFIER | MIR642B | ENSG00000283212 | Transcript | ENST00000637346.1 | miRNA | - | - | - | - | - | - | - | - | - |
| OSA | rs1800437 | rs34783010 | 19 | 46180414 | 19:45677156-45677156 | T | intron_variant | MODIFIER | GIPR | ENSG00000010310 | Transcript | ENST00000590918.6 | protein_coding | - | 8/13 | ENST00000590918.6:c.793+48G>T | - | - | - | - | - | - |
| OSA | rs1800437 | rs1800437 | 19 | 46181392 | 19:45678134-45678134 | C | downstream_gene_variant | MODIFIER | MIR642A | ENSG00000207773 | Transcript | ENST00000385039.1 | miRNA | - | - | - | - | - | - | - | - | - |
| OSA | rs1800437 | rs1800437 | 19 | 46181392 | 19:45678134-45678134 | C | upstream_gene_variant | MODIFIER | MIR642B | ENSG00000283212 | Transcript | ENST00000637346.1 | miRNA | - | - | - | - | - | - | - | - | - |
| OSA | rs1800437 | rs1800437 | 19 | 46181392 | 19:45678134-45678134 | C | missense_variant | MODERATE | GIPR | ENSG00000010310 | Transcript | ENST00000590918.6 | protein_coding | 12/14 | - | ENST00000590918.6:c.1060G>C | ENSP00000467494.1:p.Glu354Gln | 1182 | 1060 | 354 | E/Q | Gag/Cag |
| OSA | rs1800437 | rs10423928 | 19 | 46182304 | 19:45679046-45679046 | A | downstream_gene_variant | MODIFIER | MIR642A | ENSG00000207773 | Transcript | ENST00000385039.1 | miRNA | - | - | - | - | - | - | - | - | - |
| OSA | rs1800437 | rs10423928 | 19 | 46182304 | 19:45679046-45679046 | A | upstream_gene_variant | MODIFIER | MIR642B | ENSG00000283212 | Transcript | ENST00000637346.1 | miRNA | - | - | - | - | - | - | - | - | - |

**Supplementary Table 15.** Variant annotation for fine-mapped variants with highest posterior probability.

| Trait | Locus Index Variant | Fine-mapped variant | CHR | POS (hg37) | Location (hg38) | Allele | Consequence | IMPACT | SYMBOL | Gene | Feature_type | Feature | BIOTYPE | EXON | INTRON | HGVSc | HGVSp | cDNA_position | CDS_position | Protein_position | Amino_acids | Codons |
|---|---|---|---|---|---|---|---|---|---|---|---|---|---|---|---|---|---|---|---|---|---|---|
| OSA | rs1800437 | rs10423928 | 19 | 46182304 | 19:45679046-45679046 | A | intron_variant | MODIFIER | GIPR | ENSG00000010310 | Transcript | ENST00000590918.6 | protein_coding | - | 12/13 | ENST00000590918.6:c.1152+820T>A | - | - | - | - | - | - |
| OSA | rs11088984 | rs12152096 | 21 | 46496949 | 21:45077034-45077034 | C | upstream_gene_variant | MODIFIER | SSR4P1 | ENSG00000235374 | Transcript | ENST00000415143.1 | transcribed_processed_pseudogene | - | - | - | - | - | - | - | - | - |
| OSA | rs11088984 | rs12152096 | 21 | 46496949 | 21:45077034-45077034 | C | upstream_gene_variant | MODIFIER | SSR4P1 | ENSG00000293210 | Transcript | ENST00000734844.1 | lncRNA | - | - | - | - | - | - | - | - | - |
| OSA | rs11088984 | rs12152096 | 21 | 46496949 | 21:45077034-45077034 | C | intron_variant | MODIFIER | ADARB1 | ENSG00000197381 | Transcript | ENST00000348831.9 | protein_coding | - | 1/10 | ENST00000348831.9:c.-220+2241T>C | - | - | - | - | - | - |
| OSA | rs11088984 | rs2838787 | 21 | 46539725 | 21:45119810-45119810 | A | intron_variant | MODIFIER | ADARB1 | ENSG00000197381 | Transcript | ENST00000348831.9 | protein_coding | - | 1/10 | ENST00000348831.9:c.-219-8592G>A | - | - | - | - | - | - |
| OSA | rs11088984 | rs2838788 | 21 | 46542374 | 21:45122459-45122459 | C | intron_variant | MODIFIER | ADARB1 | ENSG00000197381 | Transcript | ENST00000348831.9 | protein_coding | - | 1/10 | ENST00000348831.9:c.-219-5943T>C | - | - | - | - | - | - |
| OSA | rs11088984 | rs2838813 | 21 | 46627687 | 21:45207772-45207772 | A | intron_variant | MODIFIER | ADARB1 | ENSG00000197381 | Transcript | ENST00000348831.9 | protein_coding | - | 9/10 | ENST00000348831.9:c.1747+3036G>A | - | - | - | - | - | - |
| OSA | rs11088984 | rs11088984 | 21 | 46670045 | 21:45250130-45250130 | A | intron_variant,non_coding_transcript_variant | MODIFIER | LINC00334 | ENSG00000182586 | Transcript | ENST00000660971.1 | lncRNA | - | 1/2 | ENST00000660971.1:n.170-8321C>A | - | - | - | - | - | - |
| OSA | rs11088984 | rs11088984 | 21 | 46670045 | 21:45250130-45250130 | A | downstream_gene_variant | MODIFIER | - | ENSG00000298713 | Transcript | ENST00000757533.1 | lncRNA | - | - | - | - | - | - | - | - | - |
| OSA, OSAadjBMI | rs2958127, rs4759254 | rs12828563 | 12 | 57103162 | 12:56709378-56709378 | C | downstream_gene_variant | MODIFIER | NACA | ENSG00000196531 | Transcript | ENST00000454682.6 | protein_coding | - | - | - | - | - | - | - | - | - |
| OSA, OSAadjBMI | rs2958127, rs4759255 | rs12828563 | 12 | 57103162 | 12:56709378-56709378 | C | intron_variant,non_coding_transcript_variant | MODIFIER | - | ENSG00000306812 | Transcript | ENST00000821264.1 | lncRNA | - | 2/2 | ENST00000821264.1:n.292+225T>C | - | - | - | - | - | - |
| OSA, OSAadjBMI | rs2958127, rs4759255 | rs2926748 | 12 | 57109412 | 12:56715628-56715628 | C | downstream_gene_variant | MODIFIER | - | ENSG00000306812 | Transcript | ENST00000821264.1 | lncRNA | - | - | - | - | - | - | - | - | - |
| OSA, OSAadjBMI | rs2958127, rs4759255 | rs2926748 | 12 | 57109412 | 12:56715628-56715628 | C | intron_variant | MODIFIER | NACA | ENSG00000196531 | Transcript | ENST00000454682.6 | protein_coding | - | 3/8 | ENST00000454682.6:c.5659+243C>G | - | - | - | - | - | - |
| OSA, OSAadjBMI | rs2958127, rs4759255 | rs11171951 | 12 | 57112637 | 12:56718853-56718853 | A | synonymous_variant | LOW | NACA | ENSG00000196531 | Transcript | ENST00000454682.6 | protein_coding | 3/9 | - | ENST00000454682.6:c.2677C>T | ENSP00000403817.1:p.Leu893%3D | 2716 | 2677 | 893 | L | Ctg/Ttg |
| OSA, OSAadjBMI | rs2958127, rs4759255 | rs2958127 | 12 | 57114307 | 12:56720523-56720523 | T | missense_variant | MODERATE | NACA | ENSG00000196531 | Transcript | ENST00000454682.6 | protein_coding | 3/9 | - | ENST00000454682.6:c.1007T>A | ENSP00000403817.1:p.Val336Glu | 1046 | 1007 | 336 | V/E | gTg/gAg |
| OSAadjBMI | rs2964006 | rs7709205 | 5 | 153138709 | 5:153759149-153759149 | C | downstream_gene_variant | MODIFIER | RN7SL177P | ENSG00000242976 | Transcript | ENST00000476596.3 | misc_RNA | - | - | - | - | - | - | - | - | - |
| OSAadjBMI | rs2964006 | rs7709205 | 5 | 153138709 | 5:153759149-153759149 | C | intron_variant | MODIFIER | GRIA1 | ENSG00000155511 | Transcript | ENST00000285900.10 | protein_coding | - | 11/15 | ENST00000285900.10:c.1824-5285T>C | - | - | - | - | - | - |

**Supplementary Table 15.** Variant annotation for fine-mapped variants with highest posterior probability.

| Trait | Locus Index Variant | Fine-mapped variant | CHR | POS (hg37) | Location (hg38) | Allele | Consequence | IMPACT | SYMBOL | Gene | Feature_type | Feature | BIOTYPE | EXON | INTRON | HGVSc | HGVSp | cDNA_position | CDS_position | Protein_position | Amino_acids | Codons |
|---|---|---|---|---|---|---|---|---|---|---|---|---|---|---|---|---|---|---|---|---|---|---|
| OSAadjBMI | rs2964006 | rs2126165 | 5 | 153167595 | 5:153788035-153788035 | G | intron_variant | MODIFIER | GRIA1 | ENSG00000155511 | Transcript | ENST00000285900.10 | protein_coding | - | 13/15 | ENST00000285900.10:c.2271-6586A>G | - | - | - | - | - | - |
| OSAadjBMI | rs2964006 | rs3021428 | 5 | 153210741 | 5:153831181-153831181 | C | intergenic_variant | MODIFIER | - | - | - | - | - | - | - | - | - | - | - | - | - | - |
| OSAadjBMI | rs2964006 | rs2926861 | 5 | 153212042 | 5:153832482-153832482 | T | intergenic_variant | MODIFIER | - | - | - | - | - | - | - | - | - | - | - | - | - | - |
| OSAadjBMI | rs2964006 | rs2964006 | 5 | 153213880 | 5:153834320-153834320 | G | intergenic_variant | MODIFIER | - | - | - | - | - | - | - | - | - | - | - | - | - | - |
| OSAadjBMI | rs60791051 | rs73649650 | 9 | 71939278 | 9:69324362-69324362 | C | upstream_gene_variant | MODIFIER | ENTREP1 | ENSG00000135063 | Transcript | ENST00000303068.14 | protein_coding | - | - | - | - | - | - | - | - | - |
| OSAadjBMI | rs60791051 | rs60791051 | 9 | 71941742 | 9:69326826-69326826 | C | intron_variant | MODIFIER | ENTREP1 | ENSG00000135063 | Transcript | ENST00000303068.14 | protein_coding | - | 1/10 | ENST00000303068.14:c.414+1103A>C | - | - | - | - | - | - |
| OSAadjBMI | rs4759254 | rs4759254 | 12 | 57096674 | 12:56702890-56702890 | T | intron_variant,non_coding_transcript_variant | MODIFIER | - | ENSG00000306812 | Transcript | ENST00000821264.1 | lncRNA | - | 1/2 | ENST00000821264.1:n.195-6166C>T | - | - | - | - | - | - |
| OSAadjBMI | rs10735928 | rs10784447 | 12 | 65766498 | 12:65372718-65372718 | C | intron_variant | MODIFIER | MSRB3 | ENSG00000174099 | Transcript | ENST00000308259.10 | protein_coding | - | 5/6 | ENST00000308259.10:c.292+3692T>C | - | - | - | - | - | - |
| OSAadjBMI | rs10735928 | rs10735928 | 12 | 65769626 | 12:65375846-65375846 | C | intron_variant | MODIFIER | MSRB3 | ENSG00000174099 | Transcript | ENST00000308259.10 | protein_coding | - | 5/6 | ENST00000308259.10:c.292+6820A>C | - | - | - | - | - | - |
| OSAadjBMI | rs10735928 | rs4332569 | 12 | 65793224 | 12:65399444-65399444 | G | intron_variant | MODIFIER | MSRB3 | ENSG00000174099 | Transcript | ENST00000308259.10 | protein_coding | - | 5/6 | ENST00000308259.10:c.292+30418A>G | - | - | - | - | - | - |
| OSAadjBMI | rs10735928 | rs1826663 | 12 | 65797709 | 12:65403929-65403929 | T | intron_variant | MODIFIER | MSRB3 | ENSG00000174099 | Transcript | ENST00000308259.10 | protein_coding | - | 5/6 | ENST00000308259.10:c.292+34903C>T | - | - | - | - | - | - |

**Supplementary Table 15.** Variant annotation for fine-mapped vari

| Trait | Locus Index Variant | Fine-mapped variant | Existing_variation | REF_ALLELE | UPLOADED_ALLELE | DISTANCE | STRAND | SYMBOL_SOURCE | HGNC_ID | CANONICAL | MANE | MANE_SELECT | TSL | APPRIS | ENSP | SIFT | PolyPhen | DOMAINS | AF | AFR_AF | AMR_AF | EAS_AF | EUR_AF |
|---|---|---|---|---|---|---|---|---|---|---|---|---|---|---|---|---|---|---|---|---|---|---|---|
| AHI | rs1200596362 | rs201670852 | rs201670852 | T | T/G | - | -1 | - | - | YES | - | - | 3 | - | - | - | - | | - | - | - | - | - |
| AHI | rs1200596362 | rs201670852 | rs201670852 | T | T/G | - | 1 | - | - | YES | - | - | - | - | - | - | - | | - | - | - | - | - |
| AHI | rs1200596362 | rs1553539108 | - | - | AA/AAA | - | -1 | - | - | YES | - | - | 3 | - | - | - | - | | - | - | - | - | - |
| AHI | rs1200596362 | rs1553539108 | - | - | AA/AAA | - | 1 | - | - | YES | - | - | - | - | - | - | - | | - | - | - | - | - |
| AHI | rs1200596362 | rs1200596362 | rs1200596362 | T | T/- | - | -1 | - | - | YES | - | - | 3 | - | - | - | - | | - | - | - | - | - |
| AHI | rs1200596362 | rs13015934 | rs13015934 | C | C/A/T | - | -1 | - | - | YES | - | - | 3 | - | - | - | - | | 0.1488 | 0.1543 | 0.147 | 0.0446 | 0.2276 |
| AHI | rs1200596362 | rs903825330 | rs903825330 | T | T/A | 414 | -1 | HGNC | HGNC:52633 | YES | - | - | - | - | - | - | - | | - | - | - | - | - |
| AHIadjBMI | rs78545995 | rs78545995 | rs78545995 | A | A/G | - | -1 | HGNC | HGNC:21555 | YES | MANE_Select | NM_001142800.2 | 5 | A2 | ENSP00000424243 | - | - | - | 0.0078 | 0.0008 | 0.0159 | 0 | 0.0189 |
| AHIadjBMI | rs78545995 | rs140715014 | rs140715014 | A | A/G | - | -1 | HGNC | HGNC:21555 | YES | MANE_Select | NM_001142800.2 | 5 | A2 | ENSP00000424243 | - | - | - | 0.0088 | 0.0015 | 0.0173 | 0 | 0.0199 |
| AHIadjBMI | rs78545995 | rs75433897 | rs75433897 | T | T/A | - | -1 | HGNC | HGNC:21555 | YES | MANE_Select | NM_001142800.2 | 5 | A2 | ENSP00000424243 | - | - | - | 0.0078 | 0.0008 | 0.0173 | 0 | 0.0179 |
| AHIadjBMI | rs78545995 | rs149112115 | rs149112115 | G | G/A/C/T | - | -1 | HGNC | HGNC:21555 | YES | MANE_Select | NM_001142800.2 | 5 | A2 | ENSP00000424243 | - | - | - | 0.0078 | 0.0008 | 0.0173 | 0 | 0.0179 |
| avgSaO2 | rs802729 | rs802729 | rs802729 | T | T/A/C/G | - | - | - | - | - | - | - | - | - | - | - | - | - | - | - | - | - | - |
| avgSaO2 | rs802729 | rs802729 | rs802729 | T | T/A/C/G | - | - | - | - | - | - | - | - | - | - | - | - | - | 0.9954 | 1 | 0.9899 | 1 | 0.9841 |
| avgSaO2 | rs802729 | rs117749759 | rs117749759 | A | A/C/T | - | -1 | HGNC | HGNC:9674 | YES | - | - | 5 | - | ENSP00000487005 | - | - | - | - | - | - | - | - |
| avgSaO2 | rs802729 | rs117749759 | rs117749759 | A | A/C/T | - | 1 | HGNC | HGNC:40484 | YES | - | - | 1 | - | - | - | - | - | - | - | - | - | - |
| avgSaO2 | rs802729 | rs117749759 | rs117749759,COSV63893804 | A | A/C/T | - | 1 | HGNC | HGNC:40484 | YES | - | - | 1 | - | - | - | - | - | 0.0036 | 0 | 0.0086 | 0 | 0.0099 |
| avgSaO2 | rs802729 | rs117749759 | rs117749759,COSV63893804 | A | A/C/T | - | -1 | HGNC | HGNC:9674 | YES | MANE_Select | NM_002844.4 | 1 | P4 | ENSP00000357209 | - | - | - | 0.0036 | 0 | 0.0086 | 0 | 0.0099 |
| avgSaO2 | rs2194955 | rs4739235 | rs4739235 | G | G/A/C | - | - | - | - | - | - | - | - | - | - | - | - | - | - | - | - | - | - |
| avgSaO2 | rs2194955 | rs4739235 | rs4739235 | G | G/A/C | - | -1 | - | - | YES | - | - | - | - | - | - | - | - | 0.2069 | 0.0968 | 0.2666 | 0.3849 | 0.1978 |
| avgSaO2 | rs2194955 | rs2194955 | rs2194955 | A | A/C/G/T | - | - | - | - | - | - | - | - | - | - | - | - | - | - | - | - | - | - |
| avgSaO2 | rs2194955 | rs2194955 | rs2194955 | A | A/C/G/T | - | -1 | - | - | YES | - | - | - | - | - | - | - | - | 0.3478 | 0.4145 | 0.3689 | 0.5198 | 0.2147 |
| avgSaO2 | rs2194955 | rs1549471 | rs1549471 | G | G/A | - | - | - | - | - | - | - | - | - | - | - | - | - | - | - | - | - | - |
| avgSaO2 | rs2194955 | rs1549471 | rs1549471 | G | G/A | - | -1 | - | - | YES | - | - | - | - | - | - | - | - | 0.3239 | 0.3472 | 0.4481 | 0.4405 | 0.2107 |
| avgSaO2 | rs2194955 | rs1549471 | rs1549471 | G | G/A | 4551 | 1 | - | - | YES | - | - | - | - | - | - | - | - | 0.3239 | 0.3472 | 0.4481 | 0.4405 | 0.2107 |
| avgSaO2 | rs2194955 | rs11984977 | rs11984977 | A | A/G/T | - | - | - | - | - | - | - | - | - | - | - | - | - | - | - | - | - | - |
| avgSaO2 | rs2194955 | rs11984977 | rs11984977 | A | A/G/T | 4519 | -1 | - | - | YES | - | - | - | - | - | - | - | - | 0.2917 | 0.2708 | 0.4265 | 0.4286 | 0.1849 |
| avgSaO2 | rs2194955 | rs4739247 | rs4739247 | A | A/G | - | - | - | - | - | - | - | - | - | - | - | - | - | - | - | - | - | - |
| avgSaO2 | rs2194955 | rs4739247 | rs4739247 | A | A/G | - | - | - | - | - | - | - | - | - | - | - | - | - | 0.2692 | 0.1929 | 0.415 | 0.4276 | 0.1849 |
| avgSaO2adjBMI | rs79914264 | rs79914264 | rs79914264 | G | G/A | - | - | - | - | - | - | - | - | - | - | - | - | - | 0.0667 | 0.0174 | 0.049 | 0.1944 | 0.0149 |
| OSA | rs12725199 | rs55789411 | rs55789411 | C | C/T | - | - | - | - | - | - | - | - | - | - | - | - | - | 0.2897 | 0.0272 | 0.3098 | 0.4038 | 0.3211 |
| OSA | rs12725199 | rs12567921 | rs12567921 | C | C/T | - | - | - | - | - | - | - | - | - | - | - | - | - | 0.2895 | 0.0272 | 0.3098 | 0.4028 | 0.3211 |
| OSA | rs12725199 | rs35696197 | rs35696197 | G | G/T | - | 1 | HGNC | HGNC:17662 | - | - | - | 1 | A1 | ENSP00000477024 | - | - | - | 0.1020 | 0.0144 | 0.1744 | 0.0129 | 0.2624 |
| OSA | rs12725199 | rs12725199 | rs12725199 | C | C/A/G | 4498 | 1 | HGNC | HGNC:17662 | - | - | - | 1 | A1 | ENSP00000477024 | - | - | - | 0.1038 | 0.0174 | 0.1744 | 0.0119 | 0.2664 |
| OSA | rs12725199 | rs12740489 | rs12740489 | G | G/A/C/T | - | - | - | - | - | - | - | - | - | - | - | - | - | 0.1028 | 0.0136 | 0.1758 | 0.0119 | 0.2664 |
| OSA | rs2376073 | rs12467202 | rs12467202 | T | T/C | - | 1 | HGNC | HGNC:52790 | YES | - | - | - | - | - | - | - | - | 0.2598 | 0.2632 | 0.2378 | 0.3929 | 0.0984 |
| OSA | rs2376073 | rs35481561 | rs35481561 | C | C/A/T | - | 1 | HGNC | HGNC:52790 | YES | - | - | - | - | - | - | - | - | 0.2530 | 0.2564 | 0.2378 | 0.3651 | 0.0994 |
| OSA | rs2376073 | rs2376073 | rs2376073 | C | C/T | - | 1 | HGNC | HGNC:52790 | YES | - | - | - | - | - | - | - | - | 0.2584 | 0.2678 | 0.2392 | 0.3591 | 0.0994 |
| OSA | rs9831061 | rs1410233719 | rs1410233719 | A | A/C | - | 1 | HGNC | HGNC:50687 | YES | - | - | - | - | - | - | - | - | - | - | - | - | - |
| OSA | rs9831061 | rs869399 | rs869399,COSV60503617 | C | C/A/G/T | - | -1 | HGNC | HGNC:2853 | - | - | - | 3 | - | - | - | - | - | 0.5623 | 0.4425 | 0.5663 | 0.4077 | 0.7584 |
| OSA | rs9831061 | rs869399 | rs869399,COSV60503617 | C | C/A/G/T | - | - | - | - | - | - | - | - | - | - | - | - | - | 0.5623 | 0.4425 | 0.5663 | 0.4077 | 0.7584 |
| OSA | rs9831061 | rs869399 | rs869399,COSV60503617 | C | C/A/G/T | - | -1 | HGNC | HGNC:3494 | YES | MANE_Select | NM_004454.3 | 1 | P1 | ENSP00000306894 | - | - | - | 0.5623 | 0.4425 | 0.5663 | 0.4077 | 0.7584 |
| OSA | rs9831061 | rs1516728 | rs1516728 | T | T/A | 3013 | -1 | HGNC | HGNC:3494 | YES | MANE_Select | NM_004454.3 | 1 | P1 | ENSP00000306894 | - | - | - | 0.5623 | 0.4614 | 0.5692 | 0.4028 | 0.7565 |
| OSA | rs9831061 | rs1516728 | rs1516728 | T | T/A | - | -1 | HGNC | HGNC:2853 | - | - | - | 3 | - | - | - | - | - | 0.5623 | 0.4614 | 0.5692 | 0.4028 | 0.7565 |
| OSA | rs9831061 | rs7635103 | rs7635103 | C | C/A/G/T | - | -1 | HGNC | HGNC:2853 | - | - | - | 3 | - | - | - | - | - | 0.5609 | 0.4592 | 0.5677 | 0.4018 | 0.7555 |
| OSA | rs9831061 | rs9831061 | rs9831061 | G | G/A/C/T | - | -1 | HGNC | HGNC:2853 | - | - | - | 3 | - | - | - | - | - | 0.5551 | 0.4448 | 0.5677 | 0.4018 | 0.7505 |
| OSA | rs9831061 | rs9831061 | rs9831061 | G | G/A/C/T | 3861 | -1 | - | - | YES | - | - | - | - | - | - | - | - | 0.5551 | 0.4448 | 0.5677 | 0.4018 | 0.7505 |
| OSA | rs34592089 | rs34592089 | rs34592089 | G | G/A/T | - | 1 | HGNC | HGNC:18233 | YES | MANE_Select | NM_017935.5 | 1 | P1 | ENSP00000320509 | - | - | - | 0.0156 | 0.0015 | 0.0331 | 0 | 0.0527 |

**Supplementary Table 15.** Variant annotation for fine-mapped vari

| Trait | Locus Index Variant | Fine-mapped variant | Existing_variation | REF_ALLELE | UPLOADED_ALLELE | DISTANCE | STRAND | SYMBOL_SOURCE | HGNC_ID | CANONICAL | MANE | MANE_SELECT | TSL | APPRIS | ENSP | SIFT | PolyPhen | DOMAINS | AF | AFR_AF | AMR_AF | EAS_AF | EUR_AF |
|---|---|---|---|---|---|---|---|---|---|---|---|---|---|---|---|---|---|---|---|---|---|---|---|
| OSA | rs34592089 | rs34592089 | rs34592089 | G | G/A/T | - | -1 | - | - | YES | - | - | 3 | - | - | - | - | - | 0.0156 | 0.0015 | 0.0331 | 0 | 0.0527 |
| OSA | rs9380814 | rs72853773 | rs72853773 | A | A/G | - | 1 | HGNC | HGNC:2952 | YES | MANE_Select | NM_001206927.2 | 5 | P2 | ENSP00000333363 | - | - | - | 0.1689 | 0.0174 | 0.219 | 0.1677 | 0.329 |
| OSA | rs9380814 | rs9380812 | rs9380812 | C | C/T | - | 1 | HGNC | HGNC:2952 | YES | MANE_Select | NM_001206927.2 | 5 | P2 | ENSP00000333363 | - | - | - | 0.1733 | 0.0303 | 0.2219 | 0.1706 | 0.329 |
| OSA | rs9380814 | rs9380814 | rs9380814 | G | G/A | - | 1 | HGNC | HGNC:2952 | YES | MANE_Select | NM_001206927.2 | 5 | P2 | ENSP00000333363 | - | - | - | 0.1707 | 0.0227 | 0.2219 | 0.1677 | 0.329 |
| OSA | rs9380814 | rs9366987 | rs9366987 | T | T/A/C | - | 1 | HGNC | HGNC:2952 | YES | MANE_Select | NM_001206927.2 | 5 | P2 | ENSP00000333363 | - | - | - | 0.1705 | 0.0227 | 0.2205 | 0.1687 | 0.33 |
| OSA | rs9380814 | rs1537230 | rs1537230 | G | G/A | - | 1 | HGNC | HGNC:2952 | YES | MANE_Select | NM_001206927.2 | 5 | P2 | ENSP00000333363 | - | - | - | 0.1703 | 0.0227 | 0.2205 | 0.1647 | 0.329 |
| OSA | rs66474734 | rs74818520 | rs74818520 | A | A/T | 3892 | 1 | - | - | YES | - | - | 5 | - | - | - | - | - | - | - | - | - | - |
| OSA | rs66474734 | rs6469468 | rs6469468 | T | T/A/C | - | - | - | - | - | - | - | - | - | - | - | - | - | 0.5703 | 0.5817 | 0.6484 | 0.7113 | 0.3926 |
| OSA | rs66474734 | rs774904698 | rs774904698 | G | G/- | - | - | - | - | - | - | - | - | - | - | - | - | - | - | - | - | - | - |
| OSA | rs66474734 | rs1247136109 | rs1247136109 | T | T/C | - | 1 | - | - | YES | - | - | - | - | - | - | - | - | - | - | - | - | - |
| OSA | rs66474734 | rs66474734 | - | - | TA/TAATA | - | - | - | - | - | - | - | - | - | - | - | - | - | - | - | - | - | - |
| OSA | rs12553324 | rs11794152 | rs11794152 | A | A/G | - | -1 | - | - | YES | - | - | 5 | - | - | - | - | - | 0.3271 | 0.09 | 0.3559 | 0.4325 | 0.4294 |
| OSA | rs12553324 | rs10429537 | rs10429537 | C | C/A/G | - | -1 | - | - | YES | - | - | 5 | - | - | - | - | - | 0.3409 | 0.1097 | 0.3573 | 0.4762 | 0.4284 |
| OSA | rs12553324 | rs12553324 | rs12553324 | C | C/A/G | - | -1 | - | - | YES | - | - | 5 | - | - | - | - | - | 0.3610 | 0.1558 | 0.3631 | 0.4653 | 0.4304 |
| OSA | rs12553324 | rs4977836 | rs4977836 | C | C/A/T | - | -1 | - | - | YES | - | - | 5 | - | - | - | - | - | 0.3401 | 0.1089 | 0.3588 | 0.4663 | 0.4294 |
| OSA | rs12553324 | rs1590949 | rs1590949 | C | C/A/G | - | -1 | - | - | YES | - | - | 5 | - | - | - | - | - | 0.3367 | 0.0961 | 0.3559 | 0.4663 | 0.4324 |
| OSA | rs2958127 | rs2035081 | rs2035081 | A | A/G/T | - | -1 | HGNC | HGNC:9369 | YES | MANE_Select | NM_000946.3 | 1 | P1 | ENSP00000350491 | - | - | - | 0.6076 | 0.3343 | 0.6657 | 0.7401 | 0.6252 |
| OSA | rs11642015 | rs1421085 | rs1421085,CR119358 | T | T/C | - | 1 | HGNC | HGNC:24678 | YES | MANE_Select | NM_001080432.3 | 1 | P1 | ENSP00000418823 | - | - | - | 0.2286 | 0.056 | 0.2392 | 0.1687 | 0.4324 |
| OSA | rs11642015 | rs11642015 | rs11642015 | C | C/G/T | - | 1 | HGNC | HGNC:24678 | YES | MANE_Select | NM_001080432.3 | 1 | P1 | ENSP00000418823 | - | - | - | 0.2284 | 0.056 | 0.2392 | 0.1677 | 0.4324 |
| OSA | rs11642015 | rs62048402 | rs62048402 | G | G/A/T | - | 1 | HGNC | HGNC:24678 | YES | MANE_Select | NM_001080432.3 | 1 | P1 | ENSP00000418823 | - | - | - | 0.2280 | 0.056 | 0.2392 | 0.1657 | 0.4324 |

**Supplementary Table 15.** Variant annotation for fine-mapped vari

| Trait | Locus Index Variant | Fine-mapped variant | Existing_variation | REF_ALLELE | UPLOADED_ALLELE | DISTANCE | STRAND | SYMBOL_SOURCE | HGNC_ID | CANONICAL | MANE | MANE_SELECT | TSL | APPRIS | ENSP | SIFT | PolyPhen | DOMAINS | AF | AFR_AF | AMR_AF | EAS_AF | EUR_AF |
|---|---|---|---|---|---|---|---|---|---|---|---|---|---|---|---|---|---|---|---|---|---|---|---|
| OSA | rs11642015 | rs56094641 | rs56094641 | A | A/G/T | - | 1 | HGNC | HGNC:24678 | YES | MANE_Select | NM_001080432.3 | 1 | P1 | ENSP00000418823 | - | - | - | 0.2288 | 0.0545 | 0.2421 | 0.1677 | 0.4344 |
| OSA | rs11642015 | rs55872725 | rs55872725 | C | C/G/T | - | 1 | HGNC | HGNC:24678 | YES | MANE_Select | NM_001080432.3 | 1 | P1 | ENSP00000418823 | - | - | - | 0.2278 | 0.0537 | 0.2392 | 0.1657 | 0.4324 |
| OSA | rs1800437 | rs2238691 | rs2238691 | G | G/A | 761 | 1 | HGNC | HGNC:32898 | YES | - | - | - | - | - | - | - | - | 0.1603 | 0.1195 | 0.1268 | 0.2083 | 0.2147 |
| OSA | rs1800437 | rs2238691 | rs2238691 | G | G/A | 777 | -1 | HGNC | HGNC:38902 | YES | - | - | - | - | - | - | - | - | 0.1603 | 0.1195 | 0.1268 | 0.2083 | 0.2147 |
| OSA | rs1800437 | rs2238691 | rs2238691 | G | G/A | - | 1 | HGNC | HGNC:4271 | YES | MANE_Select | NM_000164.4 | 1 | P2 | ENSP00000467494 | - | - | - | 0.1603 | 0.1195 | 0.1268 | 0.2083 | 0.2147 |
| OSA | rs1800437 | rs11672660 | rs11672660,COSV54423233 | C | C/T | 1902 | 1 | HGNC | HGNC:32898 | YES | - | - | - | - | - | - | - | - | 0.1595 | 0.1165 | 0.1268 | 0.2083 | 0.2137 |
| OSA | rs1800437 | rs11672660 | rs11672660,COSV54423233 | C | C/T | 1918 | -1 | HGNC | HGNC:38902 | YES | - | - | - | - | - | - | - | - | 0.1595 | 0.1165 | 0.1268 | 0.2083 | 0.2137 |
| OSA | rs1800437 | rs11672660 | rs11672660,COSV54423233 | C | C/T | - | 1 | HGNC | HGNC:4271 | YES | MANE_Select | NM_000164.4 | 1 | P2 | ENSP00000467494 | - | - | - | 0.1595 | 0.1165 | 0.1268 | 0.2083 | 0.2137 |
| OSA | rs1800437 | rs34783010 | rs34783010,COSV54425580 | G | G/T | 2132 | 1 | HGNC | HGNC:32898 | YES | - | - | - | - | - | - | - | - | 0.1595 | 0.1165 | 0.1268 | 0.2083 | 0.2137 |

**Supplementary Table 15.** Variant annotation for fine-mapped vari

| Trait | Locus Index Variant | Fine-mapped variant | Existing_variation | REF_ALLELE | UPLOADED_ALLELE | DISTANCE | STRAND | SYMBOL_SOURCE | HGNC_ID | CANONICAL | MANE | MANE_SELECT | TSL | APPRIS | ENSP | SIFT | PolyPhen | DOMAINS | AF | AFR_AF | AMR_AF | EAS_AF | EUR_AF |
|---|---|---|---|---|---|---|---|---|---|---|---|---|---|---|---|---|---|---|---|---|---|---|---|
| OSA | rs1800437 | rs34783010 | rs34783010,COSV54425580 | G | G/T | 2148 | -1 | HGNC | HGNC:38902 | YES | - | - | - | - | - | - | - | - | 0.1595 | 0.1165 | 0.1268 | 0.2083 | 0.2137 |
| OSA | rs1800437 | rs34783010 | rs34783010,COSV54425580 | G | G/T | - | 1 | HGNC | HGNC:4271 | YES | MANE_Select | NM_000164.4 | 1 | P2 | ENSP00000467494 | - | - | - | 0.1595 | 0.1165 | 0.1268 | 0.2083 | 0.2137 |
| OSA | rs1800437 | rs1800437 | rs1800437,CM960738,COSV54422837,COSV54422925 | G | G/C | 3110 | 1 | HGNC | HGNC:32898 | YES | - | - | - | - | - | - | - | - | 0.1611 | 0.1233 | 0.1268 | 0.2073 | 0.2137 |
| OSA | rs1800437 | rs1800437 | rs1800437,CM960738,COSV54422837,COSV54422925 | G | G/C | 3126 | -1 | HGNC | HGNC:38902 | YES | - | - | - | - | - | - | - | - | 0.1611 | 0.1233 | 0.1268 | 0.2073 | 0.2137 |
| OSA | rs1800437 | rs1800437 | rs1800437,CM960738,COSV54422837,COSV54422925 | G | G/C | - | 1 | HGNC | HGNC:4271 | YES | MANE_Select | NM_000164.4 | 1 | P2 | ENSP00000467494 | tolerated(0.08) | probably_damaging(0.999) | Gene3D:1.20.1070.10,PDB-ENSP_mappings:7dty.R,PDB-ENSP_mappings:7fin.R,PDB-ENSP_mappings:7fiy.R,PDB-ENSP_mappings:7ra3.R,PDB-ENSP_mappings:7rbt.R,PDB-ENSP_mappings:7vab.R,PDB-ENSP_mappings:8itl.R,PDB-ENSP_mappings:8wa3.R,AFDB-ENSP_mappings:AF-P48546-F1,Pfam:PF00002,Prints:PR00249,PROSITE_profiles:PS50261,PANTHER:PTHR45620,Superfamily:SSF81321,Transmembrane_helices:TMhelix,CDD:cd15929 | 0.1611 | 0.1233 | 0.1268 | 0.2073 | 0.2137 |
| OSA | rs1800437 | rs10423928 | rs10423928 | T | T/A | 4022 | 1 | HGNC | HGNC:32898 | YES | - | - | - | - | - | - | - | - | 0.1719 | 0.1596 | 0.1354 | 0.2073 | 0.2137 |
| OSA | rs1800437 | rs10423928 | rs10423928 | T | T/A | 4038 | -1 | HGNC | HGNC:38902 | YES | - | - | - | - | - | - | - | - | 0.1719 | 0.1596 | 0.1354 | 0.2073 | 0.2137 |

**Supplementary Table 15.** Variant annotation for fine-mapped vari

| Trait | Locus Index Variant | Fine-mapped variant | Existing_variation | REF_ALLELE | UPLOADED_ALLELE | DISTANCE | STRAND | SYMBOL_SOURCE | HGNC_ID | CANONICAL | MANE | MANE_SELECT | TSL | APPRIS | ENSP | SIFT | PolyPhen | DOMAINS | AF | AFR_AF | AMR_AF | EAS_AF | EUR_AF |
|---|---|---|---|---|---|---|---|---|---|---|---|---|---|---|---|---|---|---|---|---|---|---|---|
| OSA | rs1800437 | rs10423928 | rs10423928 | T | T/A | - | 1 | HGNC | HGNC:4271 | YES | MANE_Select | NM_000164.4 | 1 | P2 | ENSP00000467494 | - | - | - | 0.1719 | 0.1596 | 0.1354 | 0.2073 | 0.2137 |
| OSA | rs11088984 | rs12152096 | rs12152096 | T | T/A/C | 4795 | -1 | HGNC | HGNC:23131 | YES | - | - | - | - | - | - | - | - | 0.3490 | 0.2935 | 0.2536 | 0.3859 | 0.3847 |
| OSA | rs11088984 | rs12152096 | rs12152096 | T | T/A/C | 3541 | -1 | EntrezGene | HGNC:23131 | YES | - | - | - | - | - | - | - | - | 0.3490 | 0.2935 | 0.2536 | 0.3859 | 0.3847 |
| OSA | rs11088984 | rs12152096 | rs12152096 | T | T/A/C | - | 1 | HGNC | HGNC:226 | YES | MANE_Select | NM_001112.4 | 1 | P1 | ENSP00000015877 | - | - | - | 0.3490 | 0.2935 | 0.2536 | 0.3859 | 0.3847 |
| OSA | rs11088984 | rs2838787 | rs2838787 | G | G/A | - | 1 | HGNC | HGNC:226 | YES | MANE_Select | NM_001112.4 | 1 | P1 | ENSP00000015877 | - | - | - | 0.3554 | 0.3585 | 0.2594 | 0.3423 | 0.3817 |
| OSA | rs11088984 | rs2838788 | rs2838788 | T | T/C | - | 1 | HGNC | HGNC:226 | YES | MANE_Select | NM_001112.4 | 1 | P1 | ENSP00000015877 | - | - | - | 0.3660 | 0.3896 | 0.2622 | 0.3423 | 0.3867 |
| OSA | rs11088984 | rs2838813 | rs2838813 | G | G/A | - | 1 | HGNC | HGNC:226 | YES | MANE_Select | NM_001112.4 | 1 | P1 | ENSP00000015877 | - | - | - | 0.4740 | 0.413 | 0.3876 | 0.4792 | 0.5636 |
| OSA | rs11088984 | rs11088984 | rs11088984 | C | C/A/T | - | 1 | HGNC | HGNC:16425 | YES | - | - | - | - | - | - | - | - | 0.7584 | 0.907 | 0.7622 | 0.7371 | 0.6402 |
| OSA | rs11088984 | rs11088984 | rs11088984 | C | C/A/T | 1534 | 1 | - | - | YES | - | - | - | - | - | - | - | - | 0.7584 | 0.907 | 0.7622 | 0.7371 | 0.6402 |
| OSA, OSAadjBMI | rs2958127, rs4759254 | rs12828563 | rs12828563 | T | T/A/C | 3049 | -1 | HGNC | HGNC:7629 | YES | MANE_Select | NM_001365896.1 | 5 | - | ENSP00000403817 | - | - | - | 0.6076 | 0.3343 | 0.6657 | 0.7401 | 0.6252 |
| OSA, OSAadjBMI | rs2958127, rs4759255 | rs12828563 | rs12828563 | T | T/A/C | - | 1 | - | - | YES | - | - | - | - | - | - | - | - | 0.6076 | 0.3343 | 0.6657 | 0.7401 | 0.6252 |
| OSA, OSAadjBMI | rs2958127, rs4759255 | rs2926748 | rs2926748 | G | G/A/C/T | 2667 | 1 | - | - | YES | - | - | - | - | - | - | - | - | 0.6084 | 0.3374 | 0.6657 | 0.7401 | 0.6252 |
| OSA, OSAadjBMI | rs2958127, rs4759255 | rs2926748 | rs2926748 | G | G/A/C/T | - | -1 | HGNC | HGNC:7629 | YES | MANE_Select | NM_001365896.1 | 5 | - | ENSP00000403817 | - | - | - | 0.6084 | 0.3374 | 0.6657 | 0.7401 | 0.6252 |
| OSA, OSAadjBMI | rs2958127, rs4759255 | rs11171951 | rs11171951,COSV63270734 | G | G/A/T | - | -1 | HGNC | HGNC:7629 | YES | MANE_Select | NM_001365896.1 | 5 | - | ENSP00000403817 | - | - | AFDB-ENSP_mappings:AF-E9PAV3-F1,MobiDB_lite:mobidb-lite | 0.6056 | 0.3343 | 0.6643 | 0.7401 | 0.6203 |
| OSA, OSAadjBMI | rs2958127, rs4759255 | rs2958127 | rs2958127,COSV63272186 | A | A/T | - | -1 | HGNC | HGNC:7629 | YES | MANE_Select | NM_001365896.1 | 5 | - | ENSP00000403817 | tolerated(0.2) | unknown(0) | AFDB-ENSP_mappings:AF-E9PAV3-F1 | 0.6078 | 0.3343 | 0.6657 | 0.7401 | 0.6252 |
| OSAadjBMI | rs2964006 | rs7709205 | rs7709205 | T | T/A/C | 3469 | 1 | HGNC | HGNC:46193 | YES | - | - | - | - | - | - | - | - | 0.7123 | 0.6513 | 0.6369 | 0.9306 | 0.5527 |
| OSAadjBMI | rs2964006 | rs7709205 | rs7709205 | T | T/A/C | - | 1 | HGNC | HGNC:4571 | YES | MANE_Select | NM_000827.4 | 1 | P4 | ENSP00000285900 | - | - | - | 0.7123 | 0.6513 | 0.6369 | 0.9306 | 0.5527 |

**Supplementary Table 15.** Variant annotation for fine-mapped vari

| Trait | Locus Index Variant | Fine-mapped variant | Existing_variation | REF_ALLELE | UPLOADED_ALLELE | DISTANCE | STRAND | SYMBOL_SOURCE | HGNC_ID | CANONICAL | MANE | MANE_SELECT | TSL | APPRIS | ENSP | SIFT | PolyPhen | DOMAINS | AF | AFR_AF | AMR_AF | EAS_AF | EUR_AF |
|---|---|---|---|---|---|---|---|---|---|---|---|---|---|---|---|---|---|---|---|---|---|---|---|
| OSAadjBMI | rs2964006 | rs2126165 | rs2126165,COSV53595837 | A | A/G/T | - | 1 | HGNC | HGNC:4571 | YES | MANE_Select | NM_000827.4 | 1 | P4 | ENSP00000285900 | - | - | - | 0.7340 | 0.7375 | 0.6427 | 0.9325 | 0.5427 |
| OSAadjBMI | rs2964006 | rs3021428 | rs3021428 | A | A/C/G | - | - | - | - | - | - | - | - | - | - | - | - | - | 0.7450 | 0.7595 | 0.6556 | 0.9325 | 0.5577 |
| OSAadjBMI | rs2964006 | rs2926861 | rs2926861 | G | G/A/C/T | - | - | - | - | - | - | - | - | - | - | - | - | - | 0.7450 | 0.7595 | 0.6556 | 0.9325 | 0.5577 |
| OSAadjBMI | rs2964006 | rs2964006 | rs2964006 | T | T/C/G | - | - | - | - | - | - | - | - | - | - | - | - | - | 0.7450 | 0.7595 | 0.6556 | 0.9325 | 0.5577 |
| OSAadjBMI | rs60791051 | rs73649650 | rs73649650 | A | A/C/T | 853 | 1 | HGNC | HGNC:24820 | YES | MANE_Select | NM_001347995.2 | 2 | P1 | ENSP00000304435 | - | - | - | 0.0887 | 0.1581 | 0.0432 | 0.0258 | 0.0736 |
| OSAadjBMI | rs60791051 | rs60791051 | rs60791051 | A | A/C | - | 1 | HGNC | HGNC:24820 | YES | MANE_Select | NM_001347995.2 | 2 | P1 | ENSP00000304435 | - | - | - | 0.0887 | 0.1573 | 0.0447 | 0.0258 | 0.0736 |
| OSAadjBMI | rs4759254 | rs4759254 | rs4759254 | C | C/A/G/T | - | 1 | - | - | YES | - | - | - | - | - | - | - | - | 0.5881 | 0.2436 | 0.6556 | 0.7401 | 0.664 |
| OSAadjBMI | rs10735928 | rs10784447 | rs10784447 | T | T/A/C | - | 1 | HGNC | HGNC:27375 | YES | MANE_Select | NM_001031679.3 | 1 | P1 | ENSP00000312274 | - | - | - | 0.6743 | 0.6747 | 0.7925 | 0.5833 | 0.6342 |
| OSAadjBMI | rs10735928 | rs10735928 | rs10735928 | A | A/C/T | - | 1 | HGNC | HGNC:27375 | YES | MANE_Select | NM_001031679.3 | 1 | P1 | ENSP00000312274 | - | - | - | 0.6410 | 0.5582 | 0.781 | 0.5833 | 0.6292 |
| OSAadjBMI | rs10735928 | rs4332569 | rs4332569 | A | A/G/T | - | 1 | HGNC | HGNC:27375 | YES | MANE_Select | NM_001031679.3 | 1 | P1 | ENSP00000312274 | - | - | - | 0.6741 | 0.6755 | 0.7925 | 0.5833 | 0.6322 |
| OSAadjBMI | rs10735928 | rs1826663 | rs1826663 | C | C/A/G/T | - | 1 | HGNC | HGNC:27375 | YES | MANE_Select | NM_001031679.3 | 1 | P1 | ENSP00000312274 | - | - | - | 0.6416 | 0.5598 | 0.781 | 0.5823 | 0.6302 |

**Supplementary Table 15.** Variant annotation for fine-mapped vari

| Trait | Locus Index Variant | Fine-mapped variant | SAS_AF | CLIN_SIG | SOMATIC | PHENO | PUBMED | SpliceAI_pred_DP_AG | SpliceAI_pred_DP_AL | SpliceAI_pred_DP_DG | SpliceAI_pred_DP_DL | SpliceAI_pred_DS_AG | SpliceAI_pred_DS_AL | SpliceAI_pred_DS_DG | SpliceAI_pred_DS_DL | SpliceAI_pred_SYMBOL | am_class | am_pathogenicity |
|---|---|---|---|---|---|---|---|---|---|---|---|---|---|---|---|---|---|---|
| AHI | rs1200596362 | rs201670852 | - | - | - | - | - | - | - | - | - | - | - | - | - | - | - | - |
| AHI | rs1200596362 | rs201670852 | - | - | - | - | - | - | - | - | - | - | - | - | - | - | - | - |
| AHI | rs1200596362 | rs1553539108 | - | - | - | - | - | - | - | - | - | - | - | - | - | - | - | - |
| AHI | rs1200596362 | rs1553539108 | - | - | - | - | - | - | - | - | - | - | - | - | - | - | - | - |
| AHI | rs1200596362 | rs1200596362 | - | - | - | - | - | - | - | - | - | - | - | - | - | - | - | - |
| AHI | rs1200596362 | rs13015934 | 0.1687 | - | - | - | - | - | - | - | - | - | - | - | - | - | - | - |
| AHI | rs1200596362 | rs903825330 | - | - | - | - | - | - | - | - | - | - | - | - | - | - | - | - |
| AHIadjBMI | rs78545995 | rs78545995 | 0.0082 | - | - | - | - | -3 | 47 | 47 | 27 | 0.00 | 0.00 | 0.00 | 0.00 | EYS | - | - |
| AHIadjBMI | rs78545995 | rs140715014 | 0.0102 | - | - | - | - | -41 | 45 | -2 | 45 | 0.00 | 0.00 | 0.00 | 0.00 | EYS | - | - |
| AHIadjBMI | rs78545995 | rs75433897 | 0.0082 | - | - | - | - | -18 | 24 | 20 | -2 | 0.00 | 0.00 | 0.00 | 0.00 | EYS | - | - |
| AHIadjBMI | rs78545995 | rs149112115 | 0.0082 | - | - | - | - | 44 | 6 | 12 | -31 | 0.00 | 0.00 | 0.01 | 0.00 | EYS | - | - |
| avgSaO2 | rs802729 | rs802729 | - | - | - | - | - | - | - | - | - | - | - | - | - | - | - | - |
| avgSaO2 | rs802729 | rs802729 | 1 | - | - | - | - | - | - | - | - | - | - | - | - | - | - | - |
| avgSaO2 | rs802729 | rs117749759 | - | - | - | - | - | - | - | - | - | - | - | - | - | - | - | - |
| avgSaO2 | rs802729 | rs117749759 | - | - | - | - | - | - | - | - | - | - | - | - | - | - | - | - |
| avgSaO2 | rs802729 | rs117749759 | 0.002 | - | 0,1 | 0,1 | - | - | - | - | - | - | - | - | - | - | - | - |
| avgSaO2 | rs802729 | rs117749759 | 0.002 | - | 0,1 | 0,1 | - | 20 | -48 | -24 | 40 | 0.00 | 0.00 | 0.00 | 0.00 | PTPRK | - | - |
| avgSaO2 | rs2194955 | rs4739235 | - | - | - | 1 | 35361970,30038396 | - | - | - | - | - | - | - | - | - | - | - |
| avgSaO2 | rs2194955 | rs4739235 | 0.1391 | - | - | 1 | 35361970,30038396 | - | - | - | - | - | - | - | - | - | - | - |
| avgSaO2 | rs2194955 | rs2194955 | - | - | - | - | - | - | - | - | - | - | - | - | - | - | - | - |
| avgSaO2 | rs2194955 | rs2194955 | 0.2025 | - | - | - | - | - | - | - | - | - | - | - | - | - | - | - |
| avgSaO2 | rs2194955 | rs1549471 | - | - | - | - | - | - | - | - | - | - | - | - | - | - | - | - |
| avgSaO2 | rs2194955 | rs1549471 | 0.2004 | - | - | - | - | - | - | - | - | - | - | - | - | - | - | - |
| avgSaO2 | rs2194955 | rs1549471 | 0.2004 | - | - | - | - | - | - | - | - | - | - | - | - | - | - | - |
| avgSaO2 | rs2194955 | rs11984977 | - | - | - | - | - | - | - | - | - | - | - | - | - | - | - | - |
| avgSaO2 | rs2194955 | rs11984977 | 0.1933 | - | - | - | - | - | - | - | - | - | - | - | - | - | - | - |
| avgSaO2 | rs2194955 | rs4739247 | - | - | - | - | - | - | - | - | - | - | - | - | - | - | - | - |
| avgSaO2 | rs2194955 | rs4739247 | 0.1922 | - | - | - | - | - | - | - | - | - | - | - | - | - | - | - |
| avgSaO2adjBMI | rs79914264 | rs79914264 | 0.0675 | - | - | 1 | 31598132 | - | - | - | - | - | - | - | - | - | - | - |
| OSA | rs12725199 | rs55789411 | 0.4806 | - | - | - | - | - | - | - | - | - | - | - | - | - | - | - |
| OSA | rs12725199 | rs12567921 | 0.4806 | - | - | - | - | - | - | - | - | - | - | - | - | - | - | - |
| OSA | rs12725199 | rs35696197 | 0.0961 | - | - | - | - | - | - | - | - | - | - | - | - | - | - | - |
| OSA | rs12725199 | rs12725199 | 0.0982 | - | - | - | - | - | - | - | - | - | - | - | - | - | - | - |
| OSA | rs12725199 | rs12740489 | 0.0971 | - | - | - | 19060910 | - | - | - | - | - | - | - | - | - | - | - |
| OSA | rs2376073 | rs12467202 | 0.2996 | - | - | 1 | 29532581 | - | - | - | - | - | - | - | - | - | - | - |
| OSA | rs2376073 | rs35481561 | 0.3016 | - | - | - | - | - | - | - | - | - | - | - | - | - | - | - |
| OSA | rs2376073 | rs2376073 | 0.319 | - | - | - | - | - | - | - | - | - | - | - | - | - | - | - |
| OSA | rs9831061 | rs1410233719 | - | - | - | - | - | - | - | - | - | - | - | - | - | - | - | - |
| OSA | rs9831061 | rs869399 | 0.6789 | - | 0,1 | 0,1 | - | - | - | - | - | - | - | - | - | - | - | - |
| OSA | rs9831061 | rs869399 | 0.6789 | - | 0,1 | 0,1 | - | - | - | - | - | - | - | - | - | - | - | - |
| OSA | rs9831061 | rs869399 | 0.6789 | - | 0,1 | 0,1 | - | -2 | 8 | -35 | 25 | 0.00 | 0.00 | 0.00 | 0.00 | ETV5 | - | - |
| OSA | rs9831061 | rs1516728 | 0.6585 | - | - | 1 | 20502693,23978819,30297969,23701538 | - | - | - | - | - | - | - | - | - | - | - |
| OSA | rs9831061 | rs1516728 | 0.6585 | - | - | 1 | 20502693,23978819,30297969,23701538 | - | - | - | - | - | - | - | - | - | - | - |
| OSA | rs9831061 | rs7635103 | 0.6575 | - | - | - | 31754094,24992585,34761429 | - | - | - | - | - | - | - | - | - | - | - |
| OSA | rs9831061 | rs9831061 | 0.6524 | - | - | - | - | - | - | - | - | - | - | - | - | - | - | - |
| OSA | rs9831061 | rs9831061 | 0.6524 | - | - | - | - | - | - | - | - | - | - | - | - | - | - | - |
| OSA | rs34592089 | rs34592089 | 0 | - | - | 1 | 34170227,33462484,26192919,31879980,35764056,36378351,29326435,29844566,31676860,36893272,35604428,38538606,37247657 | 25 | 0 | -10 | -1 | 0.00 | 0.00 | 0.00 | 0.00 | BANK1 | - | - |

**Supplementary Table 15.** Variant annotation for fine-mapped vari

| Trait | Locus Index Variant | Fine-mapped variant | SAS_AF | CLIN_SIG | SOMATIC | PHENO | PUBMED | SpliceAI_pred_DP_AG | SpliceAI_pred_DP_AL | SpliceAI_pred_DP_DG | SpliceAI_pred_DP_DL | SpliceAI_pred_DS_AG | SpliceAI_pred_DS_AL | SpliceAI_pred_DS_DG | SpliceAI_pred_DS_DL | SpliceAI_pred_SYMBOL | am_class | am_pathogenicity |
|---|---|---|---|---|---|---|---|---|---|---|---|---|---|---|---|---|---|---|
| OSA | rs34592089 | rs34592089 | 0 | - | - | 1 | 34170227,33462484,26192919,31879980,35764056,36378351,29326435,2984456<br>6,31676860,36893272,35604428,38538606,37247657 | - | - | - | - | - | - | - | - | - | - | - |
| OSA | rs9380814 | rs72853773 | 0.1748 | - | - | - | - | -15 | 9 | -1 | 9 | 0.00 | 0.00 | 0.00 | 0.00 | DNAH8 | - | - |
| OSA | rs9380814 | rs9380812 | 0.1748 | - | - | - | - | 16 | -2 | -8 | -3 | 0.00 | 0.00 | 0.00 | 0.00 | DNAH8 | - | - |
| OSA | rs9380814 | rs9380814 | 0.1748 | - | - | - | - | 39 | 47 | 39 | 0 | 0.00 | 0.00 | 0.00 | 0.00 | DNAH8 | - | - |
| OSA | rs9380814 | rs9366987 | 0.1728 | - | - | - | - | -4 | 10 | 37 | -10 | 0.00 | 0.00 | 0.00 | 0.00 | DNAH8 | - | - |
| OSA | rs9380814 | rs1537230 | 0.1769 | - | - | - | - | 14 | 1 | 45 | 28 | 0.00 | 0.00 | 0.00 | 0.00 | DNAH8 | - | - |
| OSA | rs66474734 | rs74818520 | - | - | - | - | - | - | - | - | - | - | - | - | - | - | - | - |
| OSA | rs66474734 | rs6469468 | 0.5368 | - | - | - | - | - | - | - | - | - | - | - | - | - | - | - |
| OSA | rs66474734 | rs774904698 | - | - | - | - | - | - | - | - | - | - | - | - | - | - | - | - |
| OSA | rs66474734 | rs1247136109 | - | - | - | - | - | - | - | - | - | - | - | - | - | - | - | - |
| OSA | rs66474734 | rs66474734 | - | - | - | - | - | - | - | - | - | - | - | - | - | - | - | - |
| OSA | rs12553324 | rs11794152 | 0.4131 | - | - | 1 | 29520040,34855049,29186694 | - | - | - | - | - | - | - | - | - | - | - |
| OSA | rs12553324 | rs10429537 | 0.4121 | - | - | 1 | 29942085,38250258 | - | - | - | - | - | - | - | - | - | - | - |
| OSA | rs12553324 | rs12553324 | 0.4581 | - | - | 1 | 27046643,34613391,29942042,30531795,33097823,27329760,35101157 | - | - | - | - | - | - | - | - | - | - | - |
| OSA | rs12553324 | rs4977836 | 0.4172 | - | - | - | 31222134 | - | - | - | - | - | - | - | - | - | - | - |
| OSA | rs12553324 | rs1590949 | 0.4162 | - | - | 1 | 34211149,38288469 | - | - | - | - | - | - | - | - | - | - | - |
| OSA | rs2958127 | rs2035081 | 0.7812 | - | - | - | 32635923 | -3 | -16 | -41 | 5 | 0.00 | 0.00 | 0.00 | 0.00 | PRIM1 | - | - |
| OSA | rs11642015 | rs1421085 | 0.3067 | risk_factor | 0,1 | 1,1 | 34131278,30584432,31872004,21118518,23799528,34566715,32390949,2825326<br>6,21778217,24992585,30515969,22614171,24879436,27547017,28699988,30607<br>69,35627568,23101478,22513296,26727462,23577239,25477898,19079261,20127<br>379,21976109,19557161,20421936,20616199,21283731,33859285,33966103,2938<br>1148,21124985,27336604,29622662,29652634,27465381,20075150,34556834,276<br>01421,26818947,26833210,30091126,26741700,33891350,22043165,28636007,21<br>150882,32429201,21651756,29220676,27538985,26604143,33222504,30607268,3<br>3729310,32948839,34784930,35450164,22761283,24607704,29410390,21701565,<br>18159244,32271837,29487953,28208657,27182965,34872017,33807560,2208493<br>1,18218107,33888571,20502638,27241637,28954439,20080751,26961502,293418<br>62,27064284,19876010,20309761,21317302,23001569,28464932,24675148,26726<br>774,30929646,17658951,29795460,32651404,22590636,27551991,28445147,2313<br>6261,21573225,19818665,35368675,31311600,32346024,18487448,20442772,193<br>22589,21102551,18379722,18599522,22189543,23236435,24106974,24768648,27<br>478388,29273742,29525920,3082275 | -43 | -44 | -50 | -44 | 0.00 | 0.00 | 0.00 | 0.00 | FTO | - | - |
| OSA | rs11642015 | rs11642015 | 0.3067 | - | - | 1 | 24879436,36088317,23860325,30819183,27820839,35435981,33285050,3153022<br>5,22531089,24385306,25054011,34268397,35715884,35773277,25177340,364775<br>30,32514122,29691431,32491157,34880666,31453325,32147644,26413465,35051<br>171,31659794,30110940,36517512,29455858,35706030,35585065,31409800,2334<br>1774,36966243,30487518,28892062,29403010,30595370,30804565,33980691,345<br>94039,37540242,37745553,37429843,37415668,36709979,38664839,38448586,38<br>604958,38538606,38689001,37280435 | 6 | -9 | -18 | 37 | 0.00 | 0.00 | 0.00 | 0.00 | FTO | - | - |
| OSA | rs11642015 | rs62048402 | 0.3067 | - | - | 1 | 23860325,30099472,33285050,31530225,22531089,24956226,24385306,2505401<br>1,25177340,36477530,36989840,36544708,26413465,29059683,31659794,302547<br>59,36777996,29769355,30456705,28391526,37147289,24094743,34594039,36952<br>292,38584824 | 48 | -19 | -4 | 28 | 0.00 | 0.00 | 0.00 | 0.00 | FTO | - | - |

**Supplementary Table 15.** Variant annotation for fine-mapped vari

| Trait | Locus Index Variant | Fine-mapped variant | SAS_AF | CLIN_SIG | SOMATIC | PHENO | PUBMED | SpliceAI_pred_DP_AG | SpliceAI_pred_DP_AL | SpliceAI_pred_DP_DG | SpliceAI_pred_DP_DL | SpliceAI_pred_DS_AG | SpliceAI_pred_DS_AL | SpliceAI_pred_DS_DG | SpliceAI_pred_DS_DL | SpliceAI_pred_SYMBOL | am_class | am_pathogenicity |
|---|---|---|---|---|---|---|---|---|---|---|---|---|---|---|---|---|---|---|
| OSA | rs11642015 | rs56094641 | 0.3067 | - | - | 1 | 33907307,31568687,33285050,31530225,30566433,35773277,35654975,31919418,37270590,36911739,33928132,31504550,33006084,33972514,37324271,36949158,33326753,34989438,32398868,36376295,36742400,36792603,32910329,35706030,35585065,35459240,35087565,34072866,34912375,35669075,34887591,36856146,32066663,36333784,36297114,33547301,31900758,32376654,31501611,30595370,31589552,34594039,36502284,31015401,38356679,38422093,38628203,38632349,37277458 | 26 | 17 | 7 | 2 | 0.00 | 0.00 | 0.00 | 0.00 | FTO | - | - |
| OSA | rs11642015 | rs55872725 | 0.3088 | benign | - | 1 | 28301549,31998841,30686248,31772290,33285050,31530225,22531089,25054011,29899525,36556195,30388399,33097823,25177340,35551307,35653391,37288296,32393786,35585065,29912962,31046077,28270201,30664634,30857519,32066663,34594039,31015401,37609985,35604428,37282553,38559031,38076869,38683826,36889626,37277458 | 26 | -33 | -31 | -24 | 0.00 | 0.00 | 0.00 | 0.00 | FTO | - | - |
| OSA | rs1800437 | rs2238691 | 0.1339 | - | - | 1 | 32944759,31152163,31453325,37280435 | - | - | - | - | - | - | - | - | - | - | - |
| OSA | rs1800437 | rs2238691 | 0.1339 | - | - | 1 | 32944759,31152163,31453325,37280435 | - | - | - | - | - | - | - | - | - | - | - |
| OSA | rs1800437 | rs2238691 | 0.1339 | - | - | 1 | 32944759,31152163,31453325,37280435 | -10 | 14 | 9 | -39 | 0.00 | 0.00 | 0.00 | 0.00 | GIPR | - | - |
| OSA | rs1800437 | rs11672660 | 0.135 | - | 0,1 | 1,1 | 35124268,22513296,29381148,20081857,29220676,32944759,28843344,23001569,30032228,19254363,34103976,30239722,36140702,26426971,36544708,25673412,35144605,23628854,36329257,33503878,36376304,33459256,28391526,36224396,28448500,35945198,34594039,38529389,38732542 | - | - | - | - | - | - | - | - | - | - | - |
| OSA | rs1800437 | rs11672660 | 0.135 | - | 0,1 | 1,1 | 35124268,22513296,29381148,20081857,29220676,32944759,28843344,23001569,30032228,19254363,34103976,30239722,36140702,26426971,36544708,25673412,35144605,23628854,36329257,33503878,36376304,33459256,28391526,36224396,28448500,35945198,34594039,38529389,38732542 | - | - | - | - | - | - | - | - | - | - | - |
| OSA | rs1800437 | rs11672660 | 0.135 | - | 0,1 | 1,1 | 35124268,22513296,29381148,20081857,29220676,32944759,28843344,23001569,30032228,19254363,34103976,30239722,36140702,26426971,36544708,25673412,35144605,23628854,36329257,33503878,36376304,33459256,28391526,36224396,28448500,35945198,34594039,38529389,38732542 | 23 | 6 | 5 | 22 | 0.00 | 0.00 | 0.00 | 0.00 | GIPR | - | - |
| OSA | rs1800437 | rs34783010 | 0.135 | - | 0,1 | 1,1 | 30910378,33462484,35585065,33459256,31409800,30224653,35213538,36154123,38584824,38459180 | - | - | - | - | - | - | - | - | - | - | - |

**Supplementary Table 15.** Variant annotation for fine-mapped vari

| Trait | Locus Index Variant | Fine-mapped variant | SAS_AF | CLIN_SIG | SOMATIC | PHENO | PUBMED | SpliceAI_pred_DP_AG | SpliceAI_pred_DP_AL | SpliceAI_pred_DP_DG | SpliceAI_pred_DP_DL | SpliceAI_pred_DS_AG | SpliceAI_pred_DS_AL | SpliceAI_pred_DS_DG | SpliceAI_pred_DS_DL | SpliceAI_pred_SYMBOL | am_class | am_pathogenicity |
|---|---|---|---|---|---|---|---|---|---|---|---|---|---|---|---|---|---|---|
| OSA | rs1800437 | rs34783010 | 0.135 | - | 0,1 | 1,1 | 30910378,33462484,35585065,33459256,31409800,30224653,35213538,3615412<br>3,38584824,38459180 | - | - | - | - | - | - | - | - | - | - | - |
| OSA | rs1800437 | rs34783010 | 0.135 | - | 0,1 | 1,1 | 30910378,33462484,35585065,33459256,31409800,30224653,35213538,3615412<br>3,38584824,38459180 | 34 | 35 | -48 | -50 | 0.00 | 0.00 | 0.00 | 0.00 | GIPR | - | - |
| OSA | rs1800437 | rs1800437 | 0.135 | - | 0,1,1,1 | 1,1,1,1 | 17952073,20935630,29303622,19935835,30515958,20081857,31209632,2130084<br>5,30846969,18222353,35754710,28326339,34426508,19254363,20516203,222370<br>64,24446656,28744963,29723152,30784161,31974732,21810601,23349498,35029<br>542,29093273,30239722,34059833,36882778,29632382,25673413,22791748,3145<br>3325,23563607,31646011,37250804,33230300,31905038,27896108,27186349,321<br>93382,29273807,36993363,28774272,34302344,34760890,35366956,33503878,27<br>974210,35810165,28391526,33208488,32888494,25969710,28448500,32376654,3<br>6581621,30593698,37980427,37993469,38137398,37925022 | - | - | - | - | - | - | - | - | - | - | - |
| OSA | rs1800437 | rs1800437 | 0.135 | - | 0,1,1,1 | 1,1,1,1 | 17952073,20935630,29303622,19935835,30515958,20081857,31209632,2130084<br>5,30846969,18222353,35754710,28326339,34426508,19254363,20516203,222370<br>64,24446656,28744963,29723152,30784161,31974732,21810601,23349498,35029<br>542,29093273,30239722,34059833,36882778,29632382,25673413,22791748,3145<br>3325,23563607,31646011,37250804,33230300,31905038,27896108,27186349,321<br>93382,29273807,36993363,28774272,34302344,34760890,35366956,33503878,27<br>974210,35810165,28391526,33208488,32888494,25969710,28448500,32376654,3<br>6581621,30593698,37980427,37993469,38137398,37925022 | - | - | - | - | - | - | - | - | - | - | - |
| OSA | rs1800437 | rs1800437 | 0.135 | - | 0,1,1,1 | 1,1,1,1 | 17952073,20935630,29303622,19935835,30515958,20081857,31209632,2130084<br>5,30846969,18222353,35754710,28326339,34426508,19254363,20516203,222370<br>64,24446656,28744963,29723152,30784161,31974732,21810601,23349498,35029<br>542,29093273,30239722,34059833,36882778,29632382,25673413,22791748,3145<br>3325,23563607,31646011,37250804,33230300,31905038,27896108,27186349,321<br>93382,29273807,36993363,28774272,34302344,34760890,35366956,33503878,27<br>974210,35810165,28391526,33208488,32888494,25969710,28448500,32376654,3<br>6581621,30593698,37980427,37993469,38137398,37925022 | 3 | 25 | 2 | 5 | 0.43 | 0.00 | 0.01 | 0.00 | GIPR | ambiguous | 0.4634 |
| OSA | rs1800437 | rs10423928 | 0.135 | - | - | 1 | 20185807,20886378,21091714,21103350,21421807,23073174,23185617,2577481<br>7,24278702,23236286,30515958,24929251,21445555,20081857,23456907,217892<br>19,31478326,27588103,30089489,31209632,28327672,21300845,35723666,33009<br>421,27160388,34188521,21525464,31860786,28326339,22237064,31974732,2174<br>7410,21810601,22399504,23349498,26395740,31874232,35029542,29093273,323<br>99466,26160912,27864399,27623947,22958899,36140702,34880666,36834796,31<br>669095,30910378,26413465,23563607,31905038,30667137,27896108,29777116,2<br>5825681,37049537,28977447,31592155,22717537,23557707,26963759,22892131,<br>33503878,35585065,26417406,20437211,28592605,30578418,25321336,2857445<br>4,22415877,25969710,29412819,29403010,32376654,31501611,35213538,305953<br>70,36502284,36693378,37540242,37705021,38464367,37795212,38538606,38689<br>001,37280435 | - | - | - | - | - | - | - | - | - | - | - |
| OSA | rs1800437 | rs10423928 | 0.135 | - | - | 1 | 20185807,20886378,21091714,21103350,21421807,23073174,23185617,2577481<br>7,24278702,23236286,30515958,24929251,21445555,20081857,23456907,217892<br>19,31478326,27588103,30089489,31209632,28327672,21300845,35723666,33009<br>421,27160388,34188521,21525464,31860786,28326339,22237064,31974732,2174<br>7410,21810601,22399504,23349498,26395740,31874232,35029542,29093273,323<br>99466,26160912,27864399,27623947,22958899,36140702,34880666,36834796,31<br>669095,30910378,26413465,23563607,31905038,30667137,27896108,29777116,2<br>5825681,37049537,28977447,31592155,22717537,23557707,26963759,22892131,<br>33503878,35585065,26417406,20437211,28592605,30578418,25321336,2857445<br>4,22415877,25969710,29412819,29403010,32376654,31501611,35213538,305953<br>70,36502284,36693378,37540242,37705021,38464367,37795212,38538606,38689<br>001,37280435 | - | - | - | - | - | - | - | - | - | - | - |

**Supplementary Table 15.** Variant annotation for fine-mapped vari

| Trait | Locus Index Variant | Fine-mapped variant | SAS_AF | CLIN_SIG | SOMATIC | PHENO | PUBMED | SpliceAI_pred_DP_AG | SpliceAI_pred_DP_AL | SpliceAI_pred_DP_DG | SpliceAI_pred_DP_DL | SpliceAI_pred_DS_AG | SpliceAI_pred_DS_AL | SpliceAI_pred_DS_DG | SpliceAI_pred_DS_DL | SpliceAI_pred_SYMBOL | am_class | am_pathogenicity |
|---|---|---|---|---|---|---|---|---|---|---|---|---|---|---|---|---|---|---|
| OSA | rs1800437 | rs10423928 | 0.135 | - | - | 1 | 20185807,20886378,21091714,21103350,21421807,23073174,23185617,2577481<br>7,24278702,23236286,30515958,24929251,21445555,20081857,23456907,217892<br>19,31478326,27588103,30089489,31209632,28327672,21300845,35723666,33009<br>421,27160388,34188521,21525464,31860786,28326339,22237064,31974732,2174<br>7410,21810601,22399504,23349498,26395740,31874232,35029542,29093273,323<br>99466,26160912,27864399,27623947,22958899,36140702,34880666,36834796,31<br>669095,30910378,26413465,23563607,31905038,30667137,27896108,29777116,2<br>5825681,37049537,28977447,31592155,22717537,23557707,26963759,22892131,<br>33503878,35585065,26417406,20437211,28592605,30578418,25321336,2857445<br>4,22415877,25969710,29412819,29403010,32376654,31501611,35213538,305953<br>70,36502284,36693378,37540242,37705021,38464367,37795212,38538606,38689<br>001,37280435 | 2 | -31 | -3 | 39 | 0.00 | 0.00 | 0.00 | 0.00 | GIPR | - | - |
| OSA | rs11088984 | rs12152096 | 0.4172 | - | - | - | - | - | - | - | - | - | - | - | - | - | - | - |
| OSA | rs11088984 | rs12152096 | 0.4172 | - | - | - | - | - | - | - | - | - | - | - | - | - | - | - |
| OSA | rs11088984 | rs12152096 | 0.4172 | - | - | - | - | 25 | -3 | 25 | 0 | 0.00 | 0.00 | 0.00 | 0.00 | ADARB1 | - | - |
| OSA | rs11088984 | rs2838787 | 0.4059 | - | - | 1 | 30804565 | 3 | 15 | 2 | 34 | 0.00 | 0.00 | 0.00 | 0.00 | ADARB1 | - | - |
| OSA | rs11088984 | rs2838788 | 0.411 | - | - | 1 | 37464041 | -4 | 49 | 6 | -2 | 0.00 | 0.00 | 0.00 | 0.00 | ADARB1 | - | - |
| OSA | rs11088984 | rs2838813 | 0.5204 | - | - | - | - | 16 | -38 | -39 | 11 | 0.00 | 0.00 | 0.00 | 0.00 | ADARB1 | - | - |
| OSA | rs11088984 | rs11088984 | 0.6984 | - | - | - | - | - | - | - | - | - | - | - | - | - | - | - |
| OSA | rs11088984 | rs11088984 | 0.6984 | - | - | - | - | - | - | - | - | - | - | - | - | - | - | - |
| OSA, OSAadjBMI | rs2958127, rs4759254 | rs12828563 | 0.7812 | - | - | - | - | - | - | - | - | - | - | - | - | - | - | - |
| OSA, OSAadjBMI | rs2958127, rs4759255 | rs12828563 | 0.7812 | - | - | - | - | - | - | - | - | - | - | - | - | - | - | - |
| OSA, OSAadjBMI | rs2958127, rs4759255 | rs2926748 | 0.7812 | - | - | - | - | - | - | - | - | - | - | - | - | - | - | - |
| OSA, OSAadjBMI | rs2958127, rs4759255 | rs2926748 | 0.7812 | - | - | - | - | -27 | 44 | 25 | 12 | 0.00 | 0.00 | 0.00 | 0.00 | NACA | - | - |
| OSA, OSAadjBMI | rs2958127, rs4759255 | rs11171951 | 0.7771 | - | 0,1 | 0,1 | - | -16 | 47 | 7 | -44 | 0.00 | 0.00 | 0.00 | 0.00 | NACA | - | - |
| OSA, OSAadjBMI | rs2958127, rs4759255 | rs2958127 | 0.7822 | - | 0,1 | 1,1 | 38412862 | 36 | -18 | 36 | 28 | 0.00 | 0.00 | 0.00 | 0.00 | NACA | likely_benign | 0.1411 |
| OSAadjBMI | rs2964006 | rs7709205 | 0.7873 | - | - | - | - | - | - | - | - | - | - | - | - | - | - | - |
| OSAadjBMI | rs2964006 | rs7709205 | 0.7873 | - | - | - | - | -34 | 13 | 17 | -26 | 0.00 | 0.00 | 0.00 | 0.00 | GRIA1 | - | - |

**Supplementary Table 15.** Variant annotation for fine-mapped vari

| Trait | Locus Index Variant | Fine-mapped variant | SAS_AF | CLIN_SIG | SOMATIC | PHENO | PUBMED | SpliceAI_pred_DP_AG | SpliceAI_pred_DP_AL | SpliceAI_pred_DP_DG | SpliceAI_pred_DP_DL | SpliceAI_pred_DS_AG | SpliceAI_pred_DS_AL | SpliceAI_pred_DS_DG | SpliceAI_pred_DS_DL | SpliceAI_pred_SYMBOL | am_class | am_pathogenicity |
|---|---|---|---|---|---|---|---|---|---|---|---|---|---|---|---|---|---|---|
| OSAadjBMI | rs2964006 | rs2126165 | 0.7863 | - | 0,1 | 1,1 | 29691431,34880666 | -16 | -28 | 49 | -14 | 0.00 | 0.00 | 0.00 | 0.00 | GRIA1 | - | - |
| OSAadjBMI | rs2964006 | rs3021428 | 0.7883 | - | - | - | - | - | - | - | - | - | - | - | - | - | - | - |
| OSAadjBMI | rs2964006 | rs2926861 | 0.7883 | - | - | - | - | - | - | - | - | - | - | - | - | - | - | - |
| OSAadjBMI | rs2964006 | rs2964006 | 0.7883 | - | - | - | - | - | - | - | - | - | - | - | - | - | - | - |
| OSAadjBMI | rs60791051 | rs73649650 | 0.1074 | - | - | - | - | - | - | - | - | - | - | - | - | - | - | - |
| OSAadjBMI | rs60791051 | rs60791051 | 0.1074 | - | - | - | - | - | - | - | - | - | - | - | - | - | - | - |
| OSAadjBMI | rs4759254 | rs4759254 | 0.771 | - | - | 1 | 36653479 | - | - | - | - | - | - | - | - | - | - | - |
| OSAadjBMI | rs10735928 | rs10784447 | 0.7249 | - | - | 1 | 36800424,31676860 | -22 | 5 | 26 | 5 | 0.00 | 0.00 | 0.00 | 0.00 | MSRB3 | - | - |
| OSAadjBMI | rs10735928 | rs10735928 | 0.7249 | - | - | - | - | 12 | -35 | 23 | -3 | 0.00 | 0.00 | 0.00 | 0.00 | MSRB3 | - | - |
| OSAadjBMI | rs10735928 | rs4332569 | 0.7249 | - | - | - | - | -21 | 2 | 8 | -21 | 0.00 | 0.00 | 0.00 | 0.00 | MSRB3 | - | - |
| OSAadjBMI | rs10735928 | rs1826663 | 0.726 | - | - | 1 | 36989840 | 21 | -36 | -2 | 39 | 0.00 | 0.00 | 0.00 | 0.00 | MSRB3 | - | - |

**Supplementary Table 15.** Variant annotation for fine-mapped vari

| Trait | Locus Index Variant | Fine-mapped variant | PHENOTYPES | ClinPred | CADD_PHRED | CADD_RAW | OpenTargets_geneId | OpenTargets_l2g | Enformer_SAD | Enformer_SAR | GO | REVEL |
|---|---|---|---|---|---|---|---|---|---|---|---|---|
| AHI | rs1200596362 | rs201670852 | - | - | 1.405 | 0.070979 | - | - | - | - | - | - |
| AHI | rs1200596362 | rs201670852 | - | - | 1.405 | 0.070979 | - | - | - | - | - | - |
| AHI | rs1200596362 | rs1553539108 | - | - | - | - | - | - | - | - | - | - |
| AHI | rs1200596362 | rs1553539108 | - | - | - | - | - | - | - | - | - | - |
| AHI | rs1200596362 | rs1200596362 | - | - | 0.006 | -1.403323 | - | - | - | - | - | - |
| AHI | rs1200596362 | rs13015934 | - | - | 6.547 | 0.609688 | - | - | 0.0002494 | 0.0001365 | - | - |
| AHI | rs1200596362 | rs903825330 | - | - | 5.195 | 0.474221 | - | - | - | - | - | - |
| AHIadjBMI | rs78545995 | rs78545995 | Retinitis_pigmentosa+G2P+ENSG00000188107,RETINITIS_PIGMENTOSA_25+MIM_morbid+ENSG00000188107,Retinitis_pigmentosa+Orphanet+ENSG00000188107 | - | 6.925 | 0.650003 | - | - | 7.09E-05 | 4.05E-05 | GO:0001750:photoreceptor_outer_segment,GO:0003674:molecular_function,GO:0005509:calcium_ion_binding,GO:0005576:extracellular_region,GO:0005737:cytoplasm,GO:0005813:centrosome,GO:0005856:cytoskeleton,GO:0005929:cilium,GO:0007601:visual_perception,GO:0033165:interphotoreceptor_matrix,GO:0042995:cell_projection,GO:0043403:skeletal_muscle_tissue_regeneration,GO:0050908:detection_of_light_stimulus_involved_in_visual_perception,GO:0070062:extracellular_exosome | - |
| AHIadjBMI | rs78545995 | rs140715014 | Retinitis_pigmentosa+G2P+ENSG00000188107,RETINITIS_PIGMENTOSA_25+MIM_morbid+ENSG00000188107,Retinitis_pigmentosa+Orphanet+ENSG00000188107 | - | 4.785 | 0.435935 | - | - | -0.0001838 | -0.0001421 | GO:0001750:photoreceptor_outer_segment,GO:0003674:molecular_function,GO:0005509:calcium_ion_binding,GO:0005576:extracellular_region,GO:0005737:cytoplasm,GO:0005813:centrosome,GO:0005856:cytoskeleton,GO:0005929:cilium,GO:0007601:visual_perception,GO:0033165:interphotoreceptor_matrix,GO:0042995:cell_projection,GO:0043403:skeletal_muscle_tissue_regeneration,GO:0050908:detection_of_light_stimulus_involved_in_visual_perception,GO:0070062:extracellular_exosome | - |
| AHIadjBMI | rs78545995 | rs75433897 | Retinitis_pigmentosa+G2P+ENSG00000188107,RETINITIS_PIGMENTOSA_25+MIM_morbid+ENSG00000188107,Retinitis_pigmentosa+Orphanet+ENSG00000188107 | - | 0.832 | -0.065135 | - | - | -0.0001994 | -0.0001776 | GO:0001750:photoreceptor_outer_segment,GO:0003674:molecular_function,GO:0005509:calcium_ion_binding,GO:0005576:extracellular_region,GO:0005737:cytoplasm,GO:0005813:centrosome,GO:0005856:cytoskeleton,GO:0005929:cilium,GO:0007601:visual_perception,GO:0033165:interphotoreceptor_matrix,GO:0042995:cell_projection,GO:0043403:skeletal_muscle_tissue_regeneration,GO:0050908:detection_of_light_stimulus_involved_in_visual_perception,GO:0070062:extracellular_exosome | - |
| AHIadjBMI | rs78545995 | rs149112115 | Retinitis_pigmentosa+G2P+ENSG00000188107,RETINITIS_PIGMENTOSA_25+MIM_morbid+ENSG00000188107,Retinitis_pigmentosa+Orphanet+ENSG00000188107 | - | 0.722 | -0.100941 | - | - | 0.000891 | 0.0005937 | GO:0001750:photoreceptor_outer_segment,GO:0003674:molecular_function,GO:0005509:calcium_ion_binding,GO:0005576:extracellular_region,GO:0005737:cytoplasm,GO:0005813:centrosome,GO:0005856:cytoskeleton,GO:0005929:cilium,GO:0007601:visual_perception,GO:0033165:interphotoreceptor_matrix,GO:0042995:cell_projection,GO:0043403:skeletal_muscle_tissue_regeneration,GO:0050908:detection_of_light_stimulus_involved_in_visual_perception,GO:0070062:extracellular_exosome | - |
| avgSaO2 | rs802729 | rs802729 | - | - | - | - | - | - | - | - | - | - |
| avgSaO2 | rs802729 | rs802729 | - | - | 0.014 | -1.168223 | - | - | - | - | - | - |
| avgSaO2 | rs802729 | rs117749759 | - | - | - | - | - | - | - | - | GO:0004721:phosphoprotein_phosphatase_activity,GO:0004725:protein_tyrosine_phosphatase_activity,GO:0005515:protein_binding,GO:0005886:plasma_membrane,GO:0006470:protein_dephosphorylation,GO:0007155:cell_adhesion,GO:0016020:membrane,GO:0016311:dephosphorylation,GO:0030054:cell_junction,GO:0043231:intracellular_membrane-bounded_organelle | - |
| avgSaO2 | rs802729 | rs117749759 | - | - | - | - | - | - | - | - | - | - |
| avgSaO2 | rs802729 | rs117749759 | - | - | 0.937 | -0.034644 | - | - | -0.0001429 | -0.0001963 | - | - |
| avgSaO2 | rs802729 | rs117749759 | - | - | 0.937 | -0.034644 | - | - | -0.0001429 | -0.0001963 | GO:0004721:phosphoprotein_phosphatase_activity,GO:0004725:protein_tyrosine_phosphatase_activity,GO:0005001:transmembrane_receptor_protein_tyrosine_phosphatase_activity,GO:0005515:protein_binding,GO:0005886:plasma_membrane,GO:0005911:cell-cell_junction,GO:0005912:adherens_junction,GO:0006470:protein_dephosphorylation,GO:0007155:cell_adhesion,GO:0007165:signal_transduction,GO:0007179:transforming_growth_factor_beta_receptor_signaling_pathway,GO:0008013:beta-catenin_binding,GO:0008285:negative_regulation_of_cell_population_proliferation,GO:0009986:cell_surface,GO:0010839:negative_regulation_of_keratinocyte_proliferation,GO:0016020:membrane,GO:0016311:dephosphorylation,GO:0016477:cell_migration,GO:0019901:protein_kinase_binding,GO:0030054:cell_junction,GO:0030336:negative_regulation_of_cell_migration,GO:0031256:leading_edge_membrane,GO:0034394:protein_localization_to_cell_surface,GO:0034614:cellular_response_to_reactive_oxygen_species,GO:0034644:cellular_response_to_UV,GO:0043231:intracellular_membrane-bounded_organelle,GO:0045295:gamma-catenin_binding,GO:0045786:negative_regulation_of_cell_cycle,GO:0045892:negative_regulation_of_DNA-templated_transcription,GO:0048041:focal_adhesion_assembly,GO:0070161:anchoring_junction | - |
| avgSaO2 | rs2194955 | rs4739235 | - | - | - | - | - | - | - | - | - | - |
| avgSaO2 | rs2194955 | rs4739235 | Educational_attainment+NHGRI-EBI_GWAS_catalog+rs4739235,Educational_attainment__years_of_education_+NHGRI-EBI_GWAS_catalog+rs4739235 | - | 7.376 | 0.699254 | ENSG00000168546 | 0.552 | -1.78E-05 | -2.38E-05 | - | - |
| avgSaO2 | rs2194955 | rs2194955 | - | - | - | - | - | - | - | - | - | - |
| avgSaO2 | rs2194955 | rs2194955 | - | - | 0.729 | -0.098756 | ENSG00000168546 | 0.565 | 2.09E-05 | 2.94E-05 | - | - |
| avgSaO2 | rs2194955 | rs1549471 | - | - | - | - | - | - | - | - | - | - |
| avgSaO2 | rs2194955 | rs1549471 | - | - | 5.100 | 0.465265 | ENSG00000168546 | 0.565 | -7.87E-06 | -1.08E-05 | - | - |
| avgSaO2 | rs2194955 | rs1549471 | - | - | 5.100 | 0.465265 | ENSG00000168546 | 0.565 | -7.87E-06 | -1.08E-05 | - | - |
| avgSaO2 | rs2194955 | rs11984977 | - | - | - | - | - | - | - | - | - | - |
| avgSaO2 | rs2194955 | rs11984977 | - | - | 4.893 | 0.445895 | ENSG00000168546 | 0.565 | -5.90E-06 | -8.20E-06 | - | - |
| avgSaO2 | rs2194955 | rs4739247 | - | - | - | - | - | - | - | - | - | - |
| avgSaO2 | rs2194955 | rs4739247 | - | - | 2.026 | 0.167358 | ENSG00000168546 | 0.565 | -3.39E-05 | -4.58E-05 | - | - |
| avgSaO2adjBMI | rs79914264 | rs79914264 | - | - | 6.860 | 0.642926 | - | - | 0.000902 | 0.0006733 | - | - |
| OSA | rs12725199 | rs55789411 | - | - | 3.402 | 0.311872 | ENSG00000117569 | 0.585 | -6.00E-07 | -6.00E-07 | - | - |
| OSA | rs12725199 | rs12567921 | - | - | 7.465 | 0.709121 | ENSG00000117569 | 0.667 | 1.00E-07 | 1.00E-07 | - | - |
| OSA | rs12725199 | rs35696197 | - | - | 0.310 | -0.314038 | ENSG00000117569 | 0.590 | -1.94E-05 | -1.90E-05 | GO:0003676:nucleic_acid_binding,GO:0003723:RNA_binding,GO:0003729:mRNA_binding,GO:0005515:protein_binding,GO:0005634:nucleus,GO:0006397:mRNA_processing,GO:0008380:RNA_splicing,GO:0033119:negative_regulation_of_RNA_splicing,GO:0043484:regulation_of_RNA_splicing | - |
| OSA | rs12725199 | rs12725199 | - | - | 0.986 | -0.021376 | ENSG00000117569 | 0.590 | -1.43E-06 | -1.43E-06 | GO:0003676:nucleic_acid_binding,GO:0003723:RNA_binding,GO:0003729:mRNA_binding,GO:0005515:protein_binding,GO:0005634:nucleus,GO:0006397:mRNA_processing,GO:0008380:RNA_splicing,GO:0033119:negative_regulation_of_RNA_splicing,GO:0043484:regulation_of_RNA_splicing | - |
| OSA | rs12725199 | rs12740489 | - | - | **16.67** | 1.981986 | ENSG00000117569 | 0.590 | 2.47E-05 | 1.93E-05 | - | - |
| OSA | rs2376073 | rs12467202 | Smoking_behaviour__maximum_cigarettes_in_a_24_hour_period_+NHGRI-EBI_GWAS_catalog+rs12467202 | - | 0.204 | -0.421472 | - | - | -0.000509 | -0.00033 | - | - |
| OSA | rs2376073 | rs35481561 | - | - | 6.669 | 0.622541 | - | - | 0.000116 | 8.08E-05 | - | - |
| OSA | rs2376073 | rs2376073 | - | - | 2.725 | 0.247328 | - | - | 8.60E-06 | 5.36E-06 | - | - |
| OSA | rs9831061 | rs1410233719 | - | - | 8.406 | 0.812808 | - | - | - | - | - | - |
| OSA | rs9831061 | rs869399 | - | - | **17.49** | 2.161008 | ENSG00000244405 | 0.664 | - | - | - | - |
| OSA | rs9831061 | rs869399 | - | - | **17.49** | 2.161008 | ENSG00000244405 | 0.664 | - | - | - | - |
| OSA | rs9831061 | rs869399 | - | - | **17.49** | 2.161008 | ENSG00000244405 | 0.664 | - | - | GO:0000122:negative_regulation_of_transcription_by_RNA_polymerase_II,GO:0000785:chromatin,GO:0000976:transcription_cis-regulatory_region_binding,GO:0000977:RNA_polymerase_II_transcription_regulatory_region_sequence-specific_DNA_binding,GO:0000981:DNA-binding_transcription_factor_activity_-_RNA_polymerase_II-specific,GO:0001228:DNA-binding_transcription_activator_activity_-_RNA_polymerase_II-specific,GO:0003677:DNA_binding,GO:0003700:DNA-binding_transcription_factor_activity,GO:0005515:protein_binding,GO:0005634:nucleus,GO:0005654:nucleoplasm,GO:0006355:regulation_of_DNA-templated_transcription,GO:0006357:regulation_of_transcription_by_RNA_polymerase_II,GO:0034599:cellular_response_to_oxidative_stress,GO:0043565:sequence-specific_DNA_binding,GO:0045666:positive_regulation_of_neuron_differentiation,GO:0045944:positive_regulation_of_transcription_by_RNA_polymerase_II,GO:0048133:male_germ-line_stem_cell_asymmetric_division,GO:0060252:positive_regulation_of_glial_cell_proliferation,GO:1990837:sequence-specific_double-stranded_DNA_binding | - |
| OSA | rs9831061 | rs1516728 | Body_Mass_Index+GIANT+rs1516728,Type_2_diabetes+NHGRI-EBI_GWAS_catalog+rs1516728 | - | 14.51 | 1.555257 | ENSG00000244405 | 0.664 | - | - | GO:0000122:negative_regulation_of_transcription_by_RNA_polymerase_II,GO:0000785:chromatin,GO:0000976:transcription_cis-regulatory_region_binding,GO:0000977:RNA_polymerase_II_transcription_regulatory_region_sequence-specific_DNA_binding,GO:0000981:DNA-binding_transcription_factor_activity_-_RNA_polymerase_II-specific,GO:0001228:DNA-binding_transcription_activator_activity_-_RNA_polymerase_II-specific,GO:0003677:DNA_binding,GO:0003700:DNA-binding_transcription_factor_activity,GO:0005515:protein_binding,GO:0005634:nucleus,GO:0005654:nucleoplasm,GO:0006355:regulation_of_DNA-templated_transcription,GO:0006357:regulation_of_transcription_by_RNA_polymerase_II,GO:0034599:cellular_response_to_oxidative_stress,GO:0043565:sequence-specific_DNA_binding,GO:0045666:positive_regulation_of_neuron_differentiation,GO:0045944:positive_regulation_of_transcription_by_RNA_polymerase_II,GO:0048133:male_germ-line_stem_cell_asymmetric_division,GO:0060252:positive_regulation_of_glial_cell_proliferation,GO:1990837:sequence-specific_double-stranded_DNA_binding | - |
| OSA | rs9831061 | rs1516728 | Body_Mass_Index+GIANT+rs1516728,Type_2_diabetes+NHGRI-EBI_GWAS_catalog+rs1516728 | - | 14.51 | 1.555257 | ENSG00000244405 | 0.664 | - | - | - | - |
| OSA | rs9831061 | rs7635103 | Body_Mass_Index+GIANT+rs7635103 | - | 8.756 | 0.848997 | ENSG00000244405 | 0.664 | - | - | - | - |
| OSA | rs9831061 | rs9831061 | - | - | 0.141 | -0.519036 | ENSG00000244405 | 0.619 | - | - | - | - |
| OSA | rs9831061 | rs9831061 | - | - | 0.141 | -0.519036 | ENSG00000244405 | 0.619 | - | - | - | - |
| OSA | rs34592089 | rs34592089 | Systemic_lupus_erythematosus+Orphanet+ENSG00000153064,Apolipoprotein_A1_levels+NHGRI-EBI_GWAS_catalog+rs34592089,Aspartate_aminotransferase_levels+NHGRI-EBI_GWAS_catalog+rs34592089,Attention_deficit_hyperactivity_disorder_or_autism_spectrum_disorder_or_intelligence_pleiotropy+NHGRI-EBI_GWAS_catalog+rs34592089,Body_Mass_Index+NHGRI-EBI_GWAS_catalog+rs34592089,Brain_region_volumes+NHGRI-EBI_GWAS_catalog+rs34592089,Brain_region_volumes+NHGRI-EBI_GWAS_catalog+rs34592089,Calcium_levels+NHGRI-EBI_GWAS_catalog+rs34592089,Crohn's_disease+NHGRI-EBI_GWAS_catalog+rs34592089,General_cognitive_ability+NHGRI-EBI_GWAS_catalog+rs34592089,High_density_lipoprotein_cholesterol_levels+NHGRI-EBI_GWAS_catalog+rs34592089,Hypertension+NHGRI-EBI_GWAS_catalog+rs34592089,Idiopathic_knee_osteoarthritis+NHGRI-EBI_GWAS_catalog+rs34592089,Intelligence__MTAG_+NHGRI-EBI_GWAS_catalog+rs34592089,Intelligence+NHGRI-EBI_GWAS_catalog+rs34592089,Low_density_lipoprotein_cholesterol_levels+NHGRI-EBI_GWAS_catalog+rs34592089,Serum_albumin_levels+NHGRI-EBI_GWAS_catalog+rs34592089,Total_cholesterol_levels+NHGRI-EBI_GWAS_catalog+rs34592089,Urate_levels+NHGRI-EBI_GWAS_catalog+rs34592089,Ventromedial_occipital_thickness_unadjusted_for_global_measures+NHGRI-EBI_GWAS_catalog+rs34592089,Whole_body_fat_mass_UKB_data_field_23100+NHGRI-EBI_GWAS_catalog+rs34592089 | - | 1.519 | 0.091469 | ENSG00000109320,ENSG00000138821,ENSG00000153064 | 0.767<br>0.931<br>0.704 | -1.67E-06 | -2.40E-06 | GO:0000165:MAPK_cascade,GO:0002020:protease_binding,GO:0005102:signaling_receptor_binding,GO:0005515:protein_binding,GO:0007165:signal_transduction,GO:0009617:response_to_bacterium,GO:0032715:negative_regulation_of_interleukin-6_production,GO:0035591:signaling_adaptor_activity,GO:0042113:B_cell_activation,GO:0043274:phospholipase_binding,GO:0043410:positive_regulation_of_MAPK_cascade,GO:0043491:phosphatidylinositol_3-kinase/protein_kinase_B_signal_transduction,GO:0045947:negative_regulation_of_translational_initiation,GO:0048471:perinuclear_region_of_cytoplasm,GO:0050731:positive_regulation_of_peptidyl-tyrosine_phosphorylation,GO:0050853:B_cell_receptor_signaling_pathway,GO:0050869:negative_regulation_of_B_cell_activation,GO:0051898:negative_regulation_of_phosphatidylinositol_3-kinase/protein_kinase_B_signal_transduction,GO:1990782:protein_tyrosine_kinase_binding | - |

**Supplementary Table 15.** Variant annotation for fine-mapped vari

| Trait | Locus Index Variant | Fine-mapped variant | PHENOTYPES | ClinPred | CADD_PHRED | CADD_RAW | OpenTargets_geneId | OpenTargets_l2g | Enformer_SAD | Enformer_SAR | GO | REVEL |
|---|---|---|---|---|---|---|---|---|---|---|---|---|
| OSA | rs34592089 | rs34592089 | Systemic_lupus_erythematosus+Orphanet+ENSG00000153064,Apolipoprotein_A1_levels+NHGRI-EBI_GWAS_catalog+rs34592089,Aspartate_aminotransferase_levels+NHGRI-EBI_GWAS_catalog+rs34592089,Attention_deficit_hyperactivity_disorder_or_autism_spectrum_disorder_or_intelligence_pleiotropy+NHGRI-EBI_GWAS_catalog+rs34592089,Body_Mass_Index+NHGRI-EBI_GWAS_catalog+rs34592089,Brain_region_volumes+NHGRI-EBI_GWAS_catalog+rs34592089,Brain_region_volumes+NHGRI-EBI_GWAS_catalog+rs34592089,Calcium_levels+NHGRI-EBI_GWAS_catalog+rs34592089,Crohn's_disease+NHGRI-EBI_GWAS_catalog+rs34592089,General_cognitive_ability+NHGRI-EBI_GWAS_catalog+rs34592089,High_density_lipoprotein_cholesterol_levels+NHGRI-EBI_GWAS_catalog+rs34592089,Hypertension+NHGRI-EBI_GWAS_catalog+rs34592089,Idiopathic_knee_osteoarthritis+NHGRI-EBI_GWAS_catalog+rs34592089,Intelligence__MTAG_+NHGRI-EBI_GWAS_catalog+rs34592089,Intelligence+NHGRI-EBI_GWAS_catalog+rs34592089,Low_density_lipoprotein_cholesterol_levels+NHGRI-EBI_GWAS_catalog+rs34592089,Serum_albumin_levels+NHGRI-EBI_GWAS_catalog+rs34592089,Total_cholesterol_levels+NHGRI-EBI_GWAS_catalog+rs34592089,Urate_levels+NHGRI-EBI_GWAS_catalog+rs34592089,Ventromedial_occipital_thickness_unadjusted_for_global_measures+NHGRI-EBI_GWAS_catalog+rs34592089,Whole_body_fat_mass_UKB_data_field_23100+NHGRI-EBI_GWAS_catalog+rs34592089 | - | 1.519 | 0.091469 | ENSG00000109320,ENSG00000138821,ENSG00000153064 | 0.767<br>0.931<br>0.704 | -1.67E-06 | -2.40E-06 | - | - |
| OSA | rs9380814 | rs72853773 | SPERMATOGENIC_FAILURE_46+MIM_morbid+ENSG00000124721 | - | 2.279 | 0.198700 | ENSG00000112164 | **0.851** | -0.00062 | -0.0003164 | GO:0005524:ATP_binding,GO:0005737:cytoplasm,GO:0005856:cytoskeleton,GO:0005874:microtubule,GO:0005929:cilium,GO:0005930:axoneme,GO:0007018:microtubule-based_movement,GO:0008569:minus-end-directed_microtubule_motor_activity,GO:0016887:ATP_hydrolysis_activity,GO:0030286:dynein_complex,GO:0045505:dynein_intermediate_chain_binding,GO:0051959:dynein_light_intermediate_chain_binding,GO:0097729:9+2_motile_cilium | - |
| OSA | rs9380814 | rs9380812 | SPERMATOGENIC_FAILURE_46+MIM_morbid+ENSG00000124721 | - | 7.490 | 0.711875 | ENSG00000112164 | **0.851** | -0.0003932 | -0.0002044 | GO:0005524:ATP_binding,GO:0005737:cytoplasm,GO:0005856:cytoskeleton,GO:0005874:microtubule,GO:0005929:cilium,GO:0005930:axoneme,GO:0007018:microtubule-based_movement,GO:0008569:minus-end-directed_microtubule_motor_activity,GO:0016887:ATP_hydrolysis_activity,GO:0030286:dynein_complex,GO:0045505:dynein_intermediate_chain_binding,GO:0051959:dynein_light_intermediate_chain_binding,GO:0097729:9+2_motile_cilium | - |
| OSA | rs9380814 | rs9380814 | SPERMATOGENIC_FAILURE_46+MIM_morbid+ENSG00000124721 | - | 1.340 | 0.058532 | ENSG00000112164 | **0.851** | -0.000696 | -0.000545 | GO:0005524:ATP_binding,GO:0005737:cytoplasm,GO:0005856:cytoskeleton,GO:0005874:microtubule,GO:0005929:cilium,GO:0005930:axoneme,GO:0007018:microtubule-based_movement,GO:0008569:minus-end-directed_microtubule_motor_activity,GO:0016887:ATP_hydrolysis_activity,GO:0030286:dynein_complex,GO:0045505:dynein_intermediate_chain_binding,GO:0051959:dynein_light_intermediate_chain_binding,GO:0097729:9+2_motile_cilium | - |
| OSA | rs9380814 | rs9366987 | SPERMATOGENIC_FAILURE_46+MIM_morbid+ENSG00000124721 | - | 11.82 | 1.151460 | ENSG00000112164 | **0.851** | 0.001945 | 0.0009003 | GO:0005524:ATP_binding,GO:0005737:cytoplasm,GO:0005856:cytoskeleton,GO:0005874:microtubule,GO:0005929:cilium,GO:0005930:axoneme,GO:0007018:microtubule-based_movement,GO:0008569:minus-end-directed_microtubule_motor_activity,GO:0016887:ATP_hydrolysis_activity,GO:0030286:dynein_complex,GO:0045505:dynein_intermediate_chain_binding,GO:0051959:dynein_light_intermediate_chain_binding,GO:0097729:9+2_motile_cilium | - |
| OSA | rs9380814 | rs1537230 | SPERMATOGENIC_FAILURE_46+MIM_morbid+ENSG00000124721 | - | 1.689 | 0.119370 | ENSG00000112164 | **0.851** | 2.04E-05 | 1.35E-05 | GO:0005524:ATP_binding,GO:0005737:cytoplasm,GO:0005856:cytoskeleton,GO:0005874:microtubule,GO:0005929:cilium,GO:0005930:axoneme,GO:0007018:microtubule-based_movement,GO:0008569:minus-end-directed_microtubule_motor_activity,GO:0016887:ATP_hydrolysis_activity,GO:0030286:dynein_complex,GO:0045505:dynein_intermediate_chain_binding,GO:0051959:dynein_light_intermediate_chain_binding,GO:0097729:9+2_motile_cilium | - |
| OSA | rs66474734 | rs74818520 | - | - | 2.355 | 0.207568 | - | - | - | - | - | - |
| OSA | rs66474734 | rs6469468 | - | - | 0.258 | -0.360744 | - | - | -0.001334 | -0.001366 | - | - |
| OSA | rs66474734 | rs774904698 | - | - | - | - | - | - | - | - | - | - |
| OSA | rs66474734 | rs1247136109 | - | - | 4.495 | 0.409501 | - | - | - | - | - | - |
| OSA | rs66474734 | rs66474734 | - | - | - | - | - | - | - | - | - | - |
| OSA | rs12553324 | rs11794152 | Cognitive_ability+NHGRI-EBI_GWAS_catalog+rs11794152,Educational_attainment+NHGRI-EBI_GWAS_catalog+rs11794152,Extremely_high_intelligence+NHGRI-EBI_GWAS_catalog+rs11794152 | - | 2.841 | 0.259008 | ENSG00000107105 | **0.801** | 0.0003078 | 0.0001991 | - | - |
| OSA | rs12553324 | rs10429537 | Depressed_affect+NHGRI-EBI_GWAS_catalog+rs10429537 | - | 1.914 | 0.152303 | ENSG00000107105 | **0.801** | 0.0001808 | 0.0001308 | - | - |
| OSA | rs12553324 | rs12553324 | Appendicular_lean_mass+NHGRI-EBI_GWAS_catalog+rs12553324,Bipolar_disorder+NHGRI-EBI_GWAS_catalog+rs12553324,Educational_attainment+NHGRI-EBI_GWAS_catalog+rs12553324,Occupational_attainment+NHGRI-EBI_GWAS_catalog+rs12553324 | - | 1.944 | 0.156358 | ENSG00000107105 | **0.801** | 0.0001633 | 0.00010526 | - | - |
| OSA | rs12553324 | rs4977836 | - | - | 11.54 | 1.118770 | ENSG00000107105 | **0.801** | 2.86E-06 | 4.10E-06 | - | - |
| OSA | rs12553324 | rs1590949 | Age_at_first_birth+NHGRI-EBI_GWAS_catalog+rs1590949 | - | 0.379 | -0.262441 | ENSG00000107105 | **0.801** | 0.000773 | 0.000455 | - | - |
| OSA | rs2958127 | rs2035081 | PRIM1-related_Primordial_Dwarfism+G2P+ENSG00000198056,PRIMORDIAL_DWARFISM-IMMUNODEFICIENCY-LIPODYSTROPHY_SYNDROME+MIM_morbid+ENSG00000198056 | - | 0.937 | -0.034559 | - | - | - | - | GO:0000287:magnesium_ion_binding,GO:0000428:DNA-directed_RNA_polymerase_complex,GO:0003896:DNA_primase_activity,GO:0005515:protein_binding,GO:0005654:nucleoplasm,GO:0005658:alpha_DNA_polymerase:primase_complex,GO:0006260:DNA_replication,GO:0006269:DNA_replication_-_synthesis_of_primer,GO:0006270:DNA_replication_initiation,GO:0008270:zinc_ion_binding,GO:0016020:membrane,GO:0016779:nucleotidyltransferase_activity,GO:0032553:ribonucleotide_binding,GO:0046872:metal_ion_binding,GO:1990077:primosome_complex | - |
| OSA | rs11642015 | rs1421085 | GROWTH_RETARDATION__DEVELOPMENTAL_DELAY__COARSE_FACIES__AND_EARLY_DEATH+G2P+ENSG00000140718,BODY_MASS_INDEX_QUANTITATIVE_TRAIT_LOCUS_14+MIM_morbid+ENSG00000140718,GROWTH_RETARDATION__DEVELOPMENTAL_DELAY__AND_FACIAL_DYSMORPHISM+MIM_morbid+ENSG00000140718,Lethal_polymalformative_syndrome__Boissel_type+Orphanet+ENSG00000140718,OBESITY__BMIQ14___SUSCEPTIBILITY_TO+ClinVar+rs1421085,Body_Mass_Index+GIANT+rs1421085,Abdominal_adipose_tissue_volumes_to_gluteofemoral_adipose_tissue_volumes_ratio+NHGRI-EBI_GWAS_catalog+rs1421085,Abdominal_fat_ratio+NHGRI-EBI_GWAS_catalog+rs1421085,Abdominal_subcutaneous_adipose_tissue_normalized+NHGRI-EBI_GWAS_catalog+rs1421085,Abdominal_subcutaneous_adipose_tissue+NHGRI-EBI_GWAS_catalog+rs1421085,Abdominal_subcutaneous_adipose_tissue_volume+NHGRI-EBI_GWAS_catalog+rs1421085,Age_at_adiposity_rebound+NHGRI-EBI_GWAS_catalog+rs1421085,Alcohol_consumption+NHGRI-EBI_GWAS_catalog+rs1421085,Alcohol_use_disorder+NHGRI-EBI_GWAS_catalog+rs1421085,Alcohol_use_disorder+NHGRI-EBI_GWAS_catalog+rs1421085,Basal_metabolic_rate_variance+NHGRI-EBI_GWAS_catalog+rs1421085,Basal_metabolic_rate+NHGRI-EBI_GWAS_catalog+rs1421085,Body_fat_percentage+NHGRI-EBI_GWAS_catalog+rs1421085,Body_fat_percentage+NHGRI-EBI_GWAS_catalog+rs1421085,Body_fat_percentage+NHGRI-EBI_GWAS_catalog+rs1421085,Body_fat_percentage+NHGRI-EBI_GWAS_catalog+rs1421085,Body_fat_rate+NHGRI-EBI_GWAS_catalog+rs1421085,Body_mass_index_or_hip_osteoarthritis_pleiotropy+NHGRI-EBI_GWAS_catalog+rs1421085,Body_Mass_Index+NHGRI-EBI_GWAS_catalog+rs1421085,Body_Mass_Index+NHGRI-EBI_GWAS_catalog+rs1421085,Body_Mass_Index+NHGRI- | - | **19.62** | 2.643531 | ENSG00000103479,ENSG00000140718 | 0.707<br>0.751 | 2.07E-05 | 2.27E-05 | GO:0001659:temperature_homeostasis,GO:0005634:nucleus,GO:0005654:nucleoplasm,GO:0005737:cytoplasm,GO:0005829:cytosol,GO:0005886:plasma_membrane,GO:0006307:DNA_alkylation_repair,GO:0008198:ferrous_iron_binding,GO:0010883:regulation_of_lipid_storage,GO:0016180:snRNA_processing,GO:0016607:nuclear_speck,GO:0016706:2-oxoglutarate-dependent_dioxygenase_activity,GO:0016740:transferase_activity,GO:0035515:oxidative_RNA_demethylase_activity,GO:0035516:broad_specificity_oxidative_DNA_demethylase_activity,GO:0040014:regulation_of_multicellular_organism_growth,GO:0042245:RNA_repair,GO:0043231:intracellular_membrane-bounded_organelle,GO:0044065:regulation_of_respiratory_system_process,GO:0046872:metal_ion_binding,GO:0051213:dioxygenase_activity,GO:0060612:adipose_tissue_development,GO:0061157:mRNA_destabilization,GO:0070350:regulation_of_white_fat_cell_proliferation,GO:0090335:regulation_of_brown_fat_cell_differentiation,GO:1990931:mRNA_N6-methyladenosine_dioxygenase_activity,GO:1990984:tRNA_demethylase_activity | - |
| OSA | rs11642015 | rs11642015 | GROWTH_RETARDATION__DEVELOPMENTAL_DELAY__COARSE_FACIES__AND_EARLY_DEATH+G2P+ENSG00000140718,BODY_MASS_INDEX_QUANTITATIVE_TRAIT_LOCUS_14+MIM_morbid+ENSG00000140718,GROWTH_RETARDATION__DEVELOPMENTAL_DELAY__AND_FACIAL_DYSMORPHISM+MIM_morbid+ENSG00000140718,Lethal_polymalformative_syndrome__Boissel_type+Orphanet+ENSG00000140718,Abdominal_adipose_tissue_volumes+NHGRI-EBI_GWAS_catalog+rs11642015,Alanine_aminotransferase_levels+NHGRI-EBI_GWAS_catalog+rs11642015,Aspartate_aminotransferase_levels+NHGRI-EBI_GWAS_catalog+rs11642015,Blood_urea_nitrogen_levels+NHGRI-EBI_GWAS_catalog+rs11642015,Blood_urea_nitrogen_levels+NHGRI-EBI_GWAS_catalog+rs11642015,Body_fat_percentage+NHGRI-EBI_GWAS_catalog+rs11642015,Body_fat_percentage+NHGRI-EBI_GWAS_catalog+rs11642015,Body_mass_index_variance+NHGRI-EBI_GWAS_catalog+rs11642015,Body_Mass_Index+NHGRI-EBI_GWAS_catalog+rs11642015,Body_Mass_Index+NHGRI-EBI_GWAS_catalog+rs11642015,Body_Mass_Index+NHGRI-EBI_GWAS_catalog+rs11642015,Diastolic_blood_pressure+NHGRI-EBI_GWAS_catalog+rs11642015,Diastolic_blood_pressure_x_smoking_status__current_vs_non-current__interaction__2df_test_+NHGRI-EBI_GWAS_catalog+rs11642015,Diastolic_blood_pressure_x_smoking_status__current_vs_non-current__interaction__2df_test_+NHGRI-EBI_GWAS_catalog+rs11642015,Diastolic_blood_pressure_x_smoking_status__ever_vs_never__interaction__2df_test_+NHGRI-EBI_GWAS_catalog+rs11642015,Diastolic_blood_pressure_x_smoking_status__ever_vs_never__interaction__2df_test_+NHGRI-EBI_GWAS_catalog+rs11642015,Drinks_per_week+NHGRI-EBI_GWAS_catalog+rs11642015,Ease_of_getting_up_in_the_morning+NHGRI-EBI_GWAS_catalog+rs11642015,Hand_grip_strength+NHGRI- | - | 9.279 | 0.902093 | ENSG00000140718 | 0.748 | 0.0001688 | 0.00011814 | GO:0001659:temperature_homeostasis,GO:0005634:nucleus,GO:0005654:nucleoplasm,GO:0005737:cytoplasm,GO:0005829:cytosol,GO:0005886:plasma_membrane,GO:0006307:DNA_alkylation_repair,GO:0008198:ferrous_iron_binding,GO:0010883:regulation_of_lipid_storage,GO:0016180:snRNA_processing,GO:0016607:nuclear_speck,GO:0016706:2-oxoglutarate-dependent_dioxygenase_activity,GO:0016740:transferase_activity,GO:0035515:oxidative_RNA_demethylase_activity,GO:0035516:broad_specificity_oxidative_DNA_demethylase_activity,GO:0040014:regulation_of_multicellular_organism_growth,GO:0042245:RNA_repair,GO:0043231:intracellular_membrane-bounded_organelle,GO:0044065:regulation_of_respiratory_system_process,GO:0046872:metal_ion_binding,GO:0051213:dioxygenase_activity,GO:0060612:adipose_tissue_development,GO:0061157:mRNA_destabilization,GO:0070350:regulation_of_white_fat_cell_proliferation,GO:0090335:regulation_of_brown_fat_cell_differentiation,GO:1990931:mRNA_N6-methyladenosine_dioxygenase_activity,GO:1990984:tRNA_demethylase_activity | - |
| OSA | rs11642015 | rs62048402 | GROWTH_RETARDATION__DEVELOPMENTAL_DELAY__COARSE_FACIES__AND_EARLY_DEATH+G2P+ENSG00000140718,BODY_MASS_INDEX_QUANTITATIVE_TRAIT_LOCUS_14+MIM_morbid+ENSG00000140718,GROWTH_RETARDATION__DEVELOPMENTAL_DELAY__AND_FACIAL_DYSMORPHISM+MIM_morbid+ENSG00000140718,Lethal_polymalformative_syndrome__Boissel_type+Orphanet+ENSG00000140718,Breast_Cancer+NHGRI-EBI_GWAS_catalog+rs62048402,Drinks_per_week+NHGRI-EBI_GWAS_catalog+rs62048402,Heart_failure+NHGRI-EBI_GWAS_catalog+rs62048402,Medication_use_diuretics+NHGRI-EBI_GWAS_catalog+rs62048402,Medication_use_diuretics+NHGRI-EBI_GWAS_catalog+rs62048402,Obstructive_sleep_apnea+NHGRI-EBI_GWAS_catalog+rs62048402 | - | 9.296 | 0.903807 | ENSG00000140718 | 0.748 | -0.0007544 | -0.0003288 | GO:0001659:temperature_homeostasis,GO:0005634:nucleus,GO:0005654:nucleoplasm,GO:0005737:cytoplasm,GO:0005829:cytosol,GO:0005886:plasma_membrane,GO:0006307:DNA_alkylation_repair,GO:0008198:ferrous_iron_binding,GO:0010883:regulation_of_lipid_storage,GO:0016180:snRNA_processing,GO:0016607:nuclear_speck,GO:0016706:2-oxoglutarate-dependent_dioxygenase_activity,GO:0016740:transferase_activity,GO:0035515:oxidative_RNA_demethylase_activity,GO:0035516:broad_specificity_oxidative_DNA_demethylase_activity,GO:0040014:regulation_of_multicellular_organism_growth,GO:0042245:RNA_repair,GO:0043231:intracellular_membrane-bounded_organelle,GO:0044065:regulation_of_respiratory_system_process,GO:0046872:metal_ion_binding,GO:0051213:dioxygenase_activity,GO:0060612:adipose_tissue_development,GO:0061157:mRNA_destabilization,GO:0070350:regulation_of_white_fat_cell_proliferation,GO:0090335:regulation_of_brown_fat_cell_differentiation,GO:1990931:mRNA_N6-methyladenosine_dioxygenase_activity,GO:1990984:tRNA_demethylase_activity | - |

**Supplementary Table 15.** Variant annotation for fine-mapped vari

| Trait | Locus Index Variant | Fine-mapped variant | PHENOTYPES | ClinPred | CADD_PHRED | CADD_RAW | OpenTargets_geneId | OpenTargets_l2g | Enformer_SAD | Enformer_SAR | GO | REVEL |
|---|---|---|---|---|---|---|---|---|---|---|---|---|
| OSA | rs11642015 | rs56094641 | GROWTH_RETARDATION__DEVELOPMENTAL_DELAY__COARSE_FACIES__AND_EARLY_DEATH+G2P+ENSG00000140718,BODY_MASS_INDEX_QUANTITATIVE_TRAIT_LOCUS_14+MIM_morbid+ENSG00000140718,GROWTH_RETARDATION__DEVELOPMENTAL_DELAY__AND_FACIAL_DYSMORPHISM+MIM_morbid+ENSG00000140718,Lethal_polymalformative_syndrome__Boissel_type+Orphanet+ENSG00000140718,Abdominal_adipose_tissue_volumes_to_gluteofemoral_adipose_tissue_volumes_ratio+NHGRI-EBI_GWAS_catalog+rs56094641,Abdominal_adipose_tissue_volumes+NHGRI-EBI_GWAS_catalog+rs56094641,Abdominal_adipose_tissue_volumes+NHGRI-EBI_GWAS_catalog+rs56094641,Adult_body_size+NHGRI-EBI_GWAS_catalog+rs56094641,Alanine_aminotransferase_levels+NHGRI-EBI_GWAS_catalog+rs56094641,Appendicular_skeletal_muscle_index_in_chronic_obstructive_pulmonary_disease+NHGRI-EBI_GWAS_catalog+rs56094641,Blood_pressure_pleiotropy_model_1_DBP_adjusted_for_estimated_causal_effects_x_SBP+NHGRI-EBI_GWAS_catalog+rs56094641,Blood_pressure_pleiotropy_model_2_SBP_adjusted_for_estimated_causal_effects_x_DBP+NHGRI-EBI_GWAS_catalog+rs56094641,Body_surface_area+NHGRI-EBI_GWAS_catalog+rs56094641,Childhood_obesity+NHGRI-EBI_GWAS_catalog+rs56094641,Chronic_elevation_of_alanine_aminotransferase_cALT_levels+NHGRI-EBI_GWAS_catalog+rs56094641,C-reactive_protein_levels+NHGRI-EBI_GWAS_catalog+rs56094641,C-reactive_protein_levels+NHGRI-EBI_GWAS_catalog+rs56094641,Dietary_Approaches_to_Stop_Hypertension_DASH_diet+NHGRI-EBI_GWAS_catalog+rs56094641,F-fish_liking_derived_food-liking_factor+NHGRI-EBI_GWAS_catalog+rs56094641,Fish-_and_plant-related_diet+NHGRI-EBI_GWAS_catalog+rs56094641,Fish-_and_plant-related_diet+NHGRI-EBI_GWAS_catalog+rs56094641,F- | - | 0.012 | -1.201580 | ENSG00000140718 | 0.748 | 0.000462 | 0.0002813 | GO:0001659:temperature_homeostasis,GO:0005634:nucleus,GO:0005654:nucleoplasm,GO:0005737:cytoplasm,GO:0005829:cytosol,GO:0005886:plasma_membrane,GO:0006307:DNA_alkylation_repair,GO:0008198:ferrous_iron_binding,GO:0010883:regulation_of_lipid_storage,GO:0016180:snRNA_processing,GO:0016607:nuclear_speck,GO:0016706:2-oxoglutarate-dependent_dioxygenase_activity,GO:0016740:transferase_activity,GO:0035515:oxidative_RNA_demethylase_activity,GO:0035516:broad_specificity_oxidative_DNA_demethylase_activity,GO:0040014:regulation_of_multicellular_organism_growth,GO:0042245:RNA_repair,GO:0043231:intracellular_membrane-bounded_organelle,GO:0044065:regulation_of_respiratory_system_process,GO:0046872:metal_ion_binding,GO:0051213:dioxygenase_activity,GO:0060612:adipose_tissue_development,GO:0061157:mRNA_destabilization,GO:0070350:regulation_of_white_fat_cell_proliferation,GO:0090335:regulation_of_brown_fat_cell_differentiation,GO:1990931:mRNA_N6-methyladenosine_dioxygenase_activity,GO:1990984:tRNA_demethylase_activity | - |
| OSA | rs11642015 | rs55872725 | GROWTH_RETARDATION__DEVELOPMENTAL_DELAY__COARSE_FACIES__AND_EARLY_DEATH+G2P+ENSG00000140718,BODY_MASS_INDEX_QUANTITATIVE_TRAIT_LOCUS_14+MIM_morbid+ENSG00000140718,GROWTH_RETARDATION__DEVELOPMENTAL_DELAY__AND_FACIAL_DYSMORPHISM+MIM_morbid+ENSG00000140718,Lethal_polymalformative_syndrome__Boissel_type+Orphanet+ENSG00000140718,ClinVar:_phenotype_not_specified+ClinVar+rs55872725,Alcohol_consumption__heavy_vs._light/non-drinkers_+NHGRI-EBI_GWAS_catalog+rs55872725,Alcohol_use_disorder__consumption_score_+NHGRI-EBI_GWAS_catalog+rs55872725,Appendicular_lean_mass+NHGRI-EBI_GWAS_catalog+rs55872725,Body_fat_distribution__arm_fat_ratio_+NHGRI-EBI_GWAS_catalog+rs55872725,Body_fat_distribution__arm_fat_ratio_+NHGRI-EBI_GWAS_catalog+rs55872725,Body_fat_distribution__leg_fat_ratio_+NHGRI-EBI_GWAS_catalog+rs55872725,Body_fat_distribution__trunk_fat_ratio_+NHGRI-EBI_GWAS_catalog+rs55872725,Body_fat_distribution__trunk_fat_ratio_+NHGRI-EBI_GWAS_catalog+rs55872725,Body_fat_percentage+NHGRI-EBI_GWAS_catalog+rs55872725,Body_mass_index_or_osteoarthritis_pleiotropy+NHGRI-EBI_GWAS_catalog+rs55872725,Body_Mass_Index+NHGRI-EBI_GWAS_catalog+rs55872725,Butternut_squash_liking+NHGRI-EBI_GWAS_catalog+rs55872725,F-acquired_taste_liking_derived_food-liking_factor+NHGRI-EBI_GWAS_catalog+rs55872725,F-cheese_liking_derived_food-liking_factor+NHGRI-EBI_GWAS_catalog+rs55872725,F-fatty/salty_food_liking_derived_food-liking_factor+NHGRI-EBI_GWAS_catalog+rs55872725,Liver_liking+NHGRI-EBI_GWAS_catalog+rs55872725,Medication_use_agents_acting_on_the_renin-angiotensin_system+NHGRI-EBI_GWAS_catalog+rs55872725,Medication_use_agents_acting_on_the_renin-angiotensin_system+NHGRI-EBI_GWAS_catalog+rs55872725,Multi-trait_sex_score+NHGRI-EBI_GWAS_catalog+rs55872725,Non- | - | 1.138 | 0.015874 | ENSG00000140718 | 0.748 | 0.000348 | 0.0002402 | GO:0001659:temperature_homeostasis,GO:0005634:nucleus,GO:0005654:nucleoplasm,GO:0005737:cytoplasm,GO:0005829:cytosol,GO:0005886:plasma_membrane,GO:0006307:DNA_alkylation_repair,GO:0008198:ferrous_iron_binding,GO:0010883:regulation_of_lipid_storage,GO:0016180:snRNA_processing,GO:0016607:nuclear_speck,GO:0016706:2-oxoglutarate-dependent_dioxygenase_activity,GO:0016740:transferase_activity,GO:0035515:oxidative_RNA_demethylase_activity,GO:0035516:broad_specificity_oxidative_DNA_demethylase_activity,GO:0040014:regulation_of_multicellular_organism_growth,GO:0042245:RNA_repair,GO:0043231:intracellular_membrane-bounded_organelle,GO:0044065:regulation_of_respiratory_system_process,GO:0046872:metal_ion_binding,GO:0051213:dioxygenase_activity,GO:0060612:adipose_tissue_development,GO:0061157:mRNA_destabilization,GO:0070350:regulation_of_white_fat_cell_proliferation,GO:0090335:regulation_of_brown_fat_cell_differentiation,GO:1990931:mRNA_N6-methyladenosine_dioxygenase_activity,GO:1990984:tRNA_demethylase_activity | - |
| OSA | rs1800437 | rs2238691 | Blood_urea_nitrogen_levels+NHGRI-EBI_GWAS_catalog+rs2238691,Body_mass_index_variance+NHGRI-EBI_GWAS_catalog+rs2238691,Body_Mass_Index+NHGRI-EBI_GWAS_catalog+rs2238691,Childhood_body_mass_index+NHGRI-EBI_GWAS_catalog+rs2238691,Hip_circumference_variance+NHGRI-EBI_GWAS_catalog+rs2238691,Hip_circumference+NHGRI-EBI_GWAS_catalog+rs2238691,Pancreatic_beta-cell_glucose_sensitivity+NHGRI-EBI_GWAS_catalog+rs2238691 | - | 3.748 | 0.342890 | ENSG00000010310 | 0.726 | -4.20E-05 | -1.60E-05 | GO:0016442:RISC_complex,GO:0035195:miRNA-mediated_post-transcriptional_gene_silencing | - |
| OSA | rs1800437 | rs2238691 | Blood_urea_nitrogen_levels+NHGRI-EBI_GWAS_catalog+rs2238691,Body_mass_index_variance+NHGRI-EBI_GWAS_catalog+rs2238691,Body_Mass_Index+NHGRI-EBI_GWAS_catalog+rs2238691,Childhood_body_mass_index+NHGRI-EBI_GWAS_catalog+rs2238691,Hip_circumference_variance+NHGRI-EBI_GWAS_catalog+rs2238691,Hip_circumference+NHGRI-EBI_GWAS_catalog+rs2238691,Pancreatic_beta-cell_glucose_sensitivity+NHGRI-EBI_GWAS_catalog+rs2238691 | - | 3.748 | 0.342890 | ENSG00000010310 | 0.726 | -4.20E-05 | -1.60E-05 | GO:0016442:RISC_complex,GO:0035195:miRNA-mediated_post-transcriptional_gene_silencing | - |
| OSA | rs1800437 | rs2238691 | Blood_urea_nitrogen_levels+NHGRI-EBI_GWAS_catalog+rs2238691,Body_mass_index_variance+NHGRI-EBI_GWAS_catalog+rs2238691,Body_Mass_Index+NHGRI-EBI_GWAS_catalog+rs2238691,Childhood_body_mass_index+NHGRI-EBI_GWAS_catalog+rs2238691,Hip_circumference_variance+NHGRI-EBI_GWAS_catalog+rs2238691,Hip_circumference+NHGRI-EBI_GWAS_catalog+rs2238691,Pancreatic_beta-cell_glucose_sensitivity+NHGRI-EBI_GWAS_catalog+rs2238691 | - | 3.748 | 0.342890 | ENSG00000010310 | 0.726 | -4.20E-05 | -1.60E-05 | GO:0002029:desensitization_of_G_protein-coupled_receptor_signaling_pathway,GO:0004888:transmembrane_signaling_receptor_activity,GO:0004930:G_protein-coupled_receptor_activity,GO:0005515:protein_binding,GO:0005886:plasma_membrane,GO:0006091:generation_of_precursor_metabolites_and_energy,GO:0007166:cell_surface_receptor_signaling_pathway,GO:0007186:G_protein-coupled_receptor_signaling_pathway,GO:0007188:adenylate_cyclase-modulating_G_protein-coupled_receptor_signaling_pathway,GO:0007189:adenylate_cyclase-activating_G_protein-coupled_receptor_signaling_pathway,GO:0007190:activation_of_adenylate_cyclase_activity,GO:0007204:positive_regulation_of_cytosolic_calcium_ion_concentration,GO:0007584:response_to_nutrient,GO:0008528:G_protein-coupled_peptide_receptor_activity,GO:0009749:response_to_glucose,GO:0016020:membrane,GO:0016519:gastric_inhibitory_peptide_receptor_activity,GO:0017046:peptide_hormone_binding,GO:0031018:endocrine_pancreas_development,GO:0032024:positive_regulation_of_insulin_secretion,GO:0038192:gastric_inhibitory_peptide_signaling_pathway,GO:0043950:positive_regulation_of_cAMP-mediated_signaling,GO:0048678:response_to_axon_injury,GO:0050796:regulation_of_insulin_secretion,GO:0051592:response_to_calcium_ion,GO:0070542:response_to_fatty_acid,GO:0120022:glucagon_family_peptide_binding | - |
| OSA | rs1800437 | rs11672660 | Blood_urea_nitrogen_levels+NHGRI-EBI_GWAS_catalog+rs11672660,Blood_urea_nitrogen_levels+NHGRI-EBI_GWAS_catalog+rs11672660,Body_mass_index_in_physically_active_individuals+NHGRI-EBI_GWAS_catalog+rs11672660,Body_mass_index_in_physically_active_individuals+NHGRI-EBI_GWAS_catalog+rs11672660,Body_mass_index__joint_analysis_main_effects_and_physical_activity_interaction_+NHGRI-EBI_GWAS_catalog+rs11672660,Body_mass_index_MTAG+NHGRI-EBI_GWAS_catalog+rs11672660,Body_Mass_Index+NHGRI-EBI_GWAS_catalog+rs11672660,Body_Mass_Index+NHGRI-EBI_GWAS_catalog+rs11672660,Body_Mass_Index+NHGRI-EBI_GWAS_catalog+rs11672660,Body_Mass_Index+NHGRI-EBI_GWAS_catalog+rs11672660,Height+NHGRI-EBI_GWAS_catalog+rs11672660,Hip_circumference+NHGRI-EBI_GWAS_catalog+rs11672660 | - | 4.267 | 0.389011 | ENSG00000010310 | 0.731 | 1.60E-05 | 1.09E-05 | GO:0016442:RISC_complex,GO:0035195:miRNA-mediated_post-transcriptional_gene_silencing | - |
| OSA | rs1800437 | rs11672660 | Blood_urea_nitrogen_levels+NHGRI-EBI_GWAS_catalog+rs11672660,Blood_urea_nitrogen_levels+NHGRI-EBI_GWAS_catalog+rs11672660,Body_mass_index_in_physically_active_individuals+NHGRI-EBI_GWAS_catalog+rs11672660,Body_mass_index_in_physically_active_individuals+NHGRI-EBI_GWAS_catalog+rs11672660,Body_mass_index__joint_analysis_main_effects_and_physical_activity_interaction_+NHGRI-EBI_GWAS_catalog+rs11672660,Body_mass_index_MTAG+NHGRI-EBI_GWAS_catalog+rs11672660,Body_Mass_Index+NHGRI-EBI_GWAS_catalog+rs11672660,Body_Mass_Index+NHGRI-EBI_GWAS_catalog+rs11672660,Height+NHGRI-EBI_GWAS_catalog+rs11672660,Hip_circumference+NHGRI-EBI_GWAS_catalog+rs11672660,Hip_circumference+NHGRI-EBI_GWAS_catalog+rs11672660 | - | 4.267 | 0.389011 | ENSG00000010310 | 0.731 | 1.60E-05 | 1.09E-05 | GO:0016442:RISC_complex,GO:0035195:miRNA-mediated_post-transcriptional_gene_silencing | - |
| OSA | rs1800437 | rs11672660 | Blood_urea_nitrogen_levels+NHGRI-EBI_GWAS_catalog+rs11672660,Blood_urea_nitrogen_levels+NHGRI-EBI_GWAS_catalog+rs11672660,Body_mass_index_in_physically_active_individuals+NHGRI-EBI_GWAS_catalog+rs11672660,Body_mass_index_in_physically_active_individuals+NHGRI-EBI_GWAS_catalog+rs11672660,Body_mass_index__joint_analysis_main_effects_and_physical_activity_interaction_+NHGRI-EBI_GWAS_catalog+rs11672660,Body_mass_index__joint_analysis_main_effects_and_physical_activity_interaction_+NHGRI-EBI_GWAS_catalog+rs11672660,Body_mass_index_MTAG+NHGRI-EBI_GWAS_catalog+rs11672660,Body_Mass_Index+NHGRI-EBI_GWAS_catalog+rs11672660,Body_Mass_Index+NHGRI-EBI_GWAS_catalog+rs11672660,Body_Mass_Index+NHGRI-EBI_GWAS_catalog+rs11672660,Body_Mass_Index+NHGRI-EBI_GWAS_catalog+rs11672660,Height+NHGRI-EBI_GWAS_catalog+rs11672660,Hip_circumference+NHGRI-EBI_GWAS_catalog+rs11672660,Hip_circumference+NHGRI-EBI_GWAS_catalog+rs11672660 | - | 4.267 | 0.389011 | ENSG00000010310 | 0.731 | 1.60E-05 | 1.09E-05 | GO:0002029:desensitization_of_G_protein-coupled_receptor_signaling_pathway,GO:0004888:transmembrane_signaling_receptor_activity,GO:0004930:G_protein-coupled_receptor_activity,GO:0005515:protein_binding,GO:0005886:plasma_membrane,GO:0006091:generation_of_precursor_metabolites_and_energy,GO:0007166:cell_surface_receptor_signaling_pathway,GO:0007186:G_protein-coupled_receptor_signaling_pathway,GO:0007188:adenylate_cyclase-modulating_G_protein-coupled_receptor_signaling_pathway,GO:0007189:adenylate_cyclase-activating_G_protein-coupled_receptor_signaling_pathway,GO:0007190:activation_of_adenylate_cyclase_activity,GO:0007204:positive_regulation_of_cytosolic_calcium_ion_concentration,GO:0007584:response_to_nutrient,GO:0008528:G_protein-coupled_peptide_receptor_activity,GO:0009749:response_to_glucose,GO:0016020:membrane,GO:0016519:gastric_inhibitory_peptide_receptor_activity,GO:0017046:peptide_hormone_binding,GO:0031018:endocrine_pancreas_development,GO:0032024:positive_regulation_of_insulin_secretion,GO:0038192:gastric_inhibitory_peptide_signaling_pathway,GO:0043950:positive_regulation_of_cAMP-mediated_signaling,GO:0048678:response_to_axon_injury,GO:0050796:regulation_of_insulin_secretion,GO:0051592:response_to_calcium_ion,GO:0070542:response_to_fatty_acid,GO:0120022:glucagon_family_peptide_binding | - |
| OSA | rs1800437 | rs34783010 | Apolipoprotein_B_levels+NHGRI-EBI_GWAS_catalog+rs34783010,Blood_glucose_levels+NHGRI-EBI_GWAS_catalog+rs34783010,Cheesecake_liking+NHGRI-EBI_GWAS_catalog+rs34783010,Cholesterol_to_total_lipids_ratio_in_small_HDL+NHGRI-EBI_GWAS_catalog+rs34783010,Creatinine_levels+NHGRI-EBI_GWAS_catalog+rs34783010,Estimated_glomerular_filtration_rate+NHGRI-EBI_GWAS_catalog+rs34783010,Free_cholesterol_to_total_lipids_ratio_in_small_LDL+NHGRI-EBI_GWAS_catalog+rs34783010,Low_density_lipoprotein_cholesterol_levels+NHGRI-EBI_GWAS_catalog+rs34783010,Pulse_pressure+NHGRI-EBI_GWAS_catalog+rs34783010,Pulse_pressure+NHGRI-EBI_GWAS_catalog+rs34783010,Total_cholesterol_levels+NHGRI-EBI_GWAS_catalog+rs34783010,Triglycerides_to_total_lipids_ratio_in_small_LDL+NHGRI-EBI_GWAS_catalog+rs34783010,Urea_levels+NHGRI-EBI_GWAS_catalog+rs34783010,Urinary_potassium_to_creatinine_ratio+NHGRI-EBI_GWAS_catalog+rs34783010,Urinary_sodium_excretion+NHGRI-EBI_GWAS_catalog+rs34783010,Urinary_sodium_excretion+NHGRI-EBI_GWAS_catalog+rs34783010 | - | 12.49 | 1.240977 | ENSG00000010310 | 0.731 | 2.65E-05 | 1.78E-05 | GO:0016442:RISC_complex,GO:0035195:miRNA-mediated_post-transcriptional_gene_silencing | - |

**Supplementary Table 15.** Variant annotation for fine-mapped vari

| Trait | Locus Index Variant | Fine-mapped variant | PHENOTYPES | ClinPred | CADD_PHRED | CADD_RAW | OpenTargets_geneId | OpenTargets_l2g | Enformer_SAD | Enformer_SAR | GO | REVEL |
|---|---|---|---|---|---|---|---|---|---|---|---|---|
| OSA | rs1800437 | rs34783010 | Apolipoprotein_B_levels+NHGRI-EBI_GWAS_catalog+rs34783010,Blood_glucose_levels+NHGRI-EBI_GWAS_catalog+rs34783010,Cheesecake_liking+NHGRI-EBI_GWAS_catalog+rs34783010,Cholesterol_to_total_lipids_ratio_in_small_HDL+NHGRI-EBI_GWAS_catalog+rs34783010,Creatinine_levels+NHGRI-EBI_GWAS_catalog+rs34783010,Estimated_glomerular_filtration_rate+NHGRI-EBI_GWAS_catalog+rs34783010,Free_cholesterol_to_total_lipids_ratio_in_small_LDL+NHGRI-EBI_GWAS_catalog+rs34783010,Low_density_lipoprotein_cholesterol_levels+NHGRI-EBI_GWAS_catalog+rs34783010,Pulse_pressure+NHGRI-EBI_GWAS_catalog+rs34783010,Pulse_pressure+NHGRI-EBI_GWAS_catalog+rs34783010,Total_cholesterol_levels+NHGRI-EBI_GWAS_catalog+rs34783010,Triglycerides_to_total_lipids_ratio_in_small_LDL+NHGRI-EBI_GWAS_catalog+rs34783010,Urea_levels+NHGRI-EBI_GWAS_catalog+rs34783010,Urinary_potassium_to_creatinine_ratio+NHGRI-EBI_GWAS_catalog+rs34783010,Urinary_sodium_excretion+NHGRI-EBI_GWAS_catalog+rs34783010,Urinary_sodium_excretion+NHGRI-EBI_GWAS_catalog+rs34783010 | - | 12.49 | 1.240977 | ENSG00000010310 | 0.731 | 2.65E-05 | 1.78E-05 | GO:0016442:RISC_complex,GO:0035195:miRNA-mediated_post-transcriptional_gene_silencing | - |
| OSA | rs1800437 | rs34783010 | Apolipoprotein_B_levels+NHGRI-EBI_GWAS_catalog+rs34783010,Blood_glucose_levels+NHGRI-EBI_GWAS_catalog+rs34783010,Cheesecake_liking+NHGRI-EBI_GWAS_catalog+rs34783010,Cholesterol_to_total_lipids_ratio_in_small_HDL+NHGRI-EBI_GWAS_catalog+rs34783010,Creatinine_levels+NHGRI-EBI_GWAS_catalog+rs34783010,Estimated_glomerular_filtration_rate+NHGRI-EBI_GWAS_catalog+rs34783010,Free_cholesterol_to_total_lipids_ratio_in_small_LDL+NHGRI-EBI_GWAS_catalog+rs34783010,Low_density_lipoprotein_cholesterol_levels+NHGRI-EBI_GWAS_catalog+rs34783010,Pulse_pressure+NHGRI-EBI_GWAS_catalog+rs34783010,Pulse_pressure+NHGRI-EBI_GWAS_catalog+rs34783010,Total_cholesterol_levels+NHGRI-EBI_GWAS_catalog+rs34783010,Triglycerides_to_total_lipids_ratio_in_small_LDL+NHGRI-EBI_GWAS_catalog+rs34783010,Urea_levels+NHGRI-EBI_GWAS_catalog+rs34783010,Urinary_potassium_to_creatinine_ratio+NHGRI-EBI_GWAS_catalog+rs34783010,Urinary_sodium_excretion+NHGRI-EBI_GWAS_catalog+rs34783010,Urinary_sodium_excretion+NHGRI-EBI_GWAS_catalog+rs34783010 | - | 12.49 | 1.240977 | ENSG00000010310 | 0.731 | 2.65E-05 | 1.78E-05 | GO:0002029:desensitization_of_G_protein-coupled_receptor_signaling_pathway,GO:0004888:transmembrane_signaling_receptor_activity,GO:0004930:G_protein-coupled_receptor_activity,GO:0005515:protein_binding,GO:0005886:plasma_membrane,GO:0006091:generation_of_precursor_metabolites_and_energy,GO:0007166:cell_surface_receptor_signaling_pathway,GO:0007186:G_protein-coupled_receptor_signaling_pathway,GO:0007188:adenylate_cyclase-modulating_G_protein-coupled_receptor_signaling_pathway,GO:0007189:adenylate_cyclase-activating_G_protein-coupled_receptor_signaling_pathway,GO:0007190:activation_of_adenylate_cyclase_activity,GO:0007204:positive_regulation_of_cytosolic_calcium_ion_concentration,GO:0007584:response_to_nutrient,GO:0008528:G_protein-coupled_peptide_receptor_activity,GO:0009749:response_to_glucose,GO:0016020:membrane,GO:0016519:gastric_inhibitory_peptide_receptor_activity,GO:0017046:peptide_hormone_binding,GO:0031018:endocrine_pancreas_development,GO:0032024:positive_regulation_of_insulin_secretion,GO:0038192:gastric_inhibitory_peptide_signaling_pathway,GO:0043950:positive_regulation_of_cAMP-mediated_signaling,GO:0048678:response_to_axon_injury,GO:0050796:regulation_of_insulin_secretion,GO:0051592:response_to_calcium_ion,GO:0070542:response_to_fatty_acid,GO:0120022:glucagon_family_peptide_binding | - |
| OSA | rs1800437 | rs1800437 | Body_fat_percentage+NHGRI-EBI_GWAS_catalog+rs1800437,Body_mass_index_in_physically_active_individuals+NHGRI-EBI_GWAS_catalog+rs1800437,Body_mass_index__joint_analysis_main_effects_and_physical_activity_interaction_+NHGRI-EBI_GWAS_catalog+rs1800437,Body_mass_index__joint_analysis_main_effects_and_physical_activity_interaction_+NHGRI-EBI_GWAS_catalog+rs1800437,Body_Mass_Index+NHGRI-EBI_GWAS_catalog+rs1800437,Body_Mass_Index+NHGRI-EBI_GWAS_catalog+rs1800437,Body_Mass_Index+NHGRI-EBI_GWAS_catalog+rs1800437,Body_size_at_age_10+NHGRI-EBI_GWAS_catalog+rs1800437,GIP_levels_in_response_to_oral_glucose_tolerance_test__120_minutes_+NHGRI-EBI_GWAS_catalog+rs1800437,GIP_levels_in_response_to_oral_glucose_tolerance_test__fasting_+NHGRI-EBI_GWAS_catalog+rs1800437,Insulin_levels_in_response_to_oral_glucose_tolerance_test__30_minutes_+NHGRI-EBI_GWAS_catalog+rs1800437,Mean_reticulocyte_volume+NHGRI-EBI_GWAS_catalog+rs1800437,Mean_spheric_corpuscular_volume+NHGRI-EBI_GWAS_catalog+rs1800437,Obesity+NHGRI-EBI_GWAS_catalog+rs1800437,Principal_component-derived_dietary_pattern_29+NHGRI-EBI_GWAS_catalog+rs1800437,Pulse_pressure+NHGRI-EBI_GWAS_catalog+rs1800437,Pulse_pressure+NHGRI-EBI_GWAS_catalog+rs1800437,Random_glucose_levels+NHGRI-EBI_GWAS_catalog+rs1800437,Systolic_blood_pressure+NHGRI-EBI_GWAS_catalog+rs1800437,Two-hour_glucose+NHGRI-EBI_GWAS_catalog+rs1800437,Type_2_diabetes__adjusted_for_BMI_+NHGRI- | - | **34** | 6.054421 | ENSG00000010310 | 0.733 | 3.04E-05 | 2.12E-05 | GO:0016442:RISC_complex,GO:0035195:miRNA-mediated_post-transcriptional_gene_silencing | - |
| OSA | rs1800437 | rs1800437 | Body_fat_percentage+NHGRI-EBI_GWAS_catalog+rs1800437,Body_mass_index_in_physically_active_individuals+NHGRI-EBI_GWAS_catalog+rs1800437,Body_mass_index__joint_analysis_main_effects_and_physical_activity_interaction_+NHGRI-EBI_GWAS_catalog+rs1800437,Body_mass_index__joint_analysis_main_effects_and_physical_activity_interaction_+NHGRI-EBI_GWAS_catalog+rs1800437,Body_Mass_Index+NHGRI-EBI_GWAS_catalog+rs1800437,Body_Mass_Index+NHGRI-EBI_GWAS_catalog+rs1800437,Body_size_at_age_10+NHGRI-EBI_GWAS_catalog+rs1800437,GIP_levels_in_response_to_oral_glucose_tolerance_test__120_minutes_+NHGRI-EBI_GWAS_catalog+rs1800437,GIP_levels_in_response_to_oral_glucose_tolerance_test__fasting_+NHGRI-EBI_GWAS_catalog+rs1800437,Insulin_levels_in_response_to_oral_glucose_tolerance_test__30_minutes_+NHGRI-EBI_GWAS_catalog+rs1800437,Mean_reticulocyte_volume+NHGRI-EBI_GWAS_catalog+rs1800437,Mean_spheric_corpuscular_volume+NHGRI-EBI_GWAS_catalog+rs1800437,Obesity+NHGRI-EBI_GWAS_catalog+rs1800437,Principal_component-derived_dietary_pattern_29+NHGRI-EBI_GWAS_catalog+rs1800437,Pulse_pressure+NHGRI-EBI_GWAS_catalog+rs1800437,Pulse_pressure+NHGRI-EBI_GWAS_catalog+rs1800437,Random_glucose_levels+NHGRI-EBI_GWAS_catalog+rs1800437,Systolic_blood_pressure+NHGRI-EBI_GWAS_catalog+rs1800437,Two-hour_glucose+NHGRI-EBI_GWAS_catalog+rs1800437,Type_2_diabetes__adjusted_for_BMI_+NHGRI-EBI_GWAS_catalog+rs1800437,Type_2_diabetes__adjusted_for_BMI_+NHGRI-EBI_GWAS_catalog+rs1800437,Waist_circumference+NHGRI-EBI_GWAS_catalog+rs1800437,Waist_circumference_variance+NHGRI- | - | **34** | 6.054421 | ENSG00000010310 | 0.733 | 3.04E-05 | 2.12E-05 | GO:0016442:RISC_complex,GO:0035195:miRNA-mediated_post-transcriptional_gene_silencing | - |
| OSA | rs1800437 | rs1800437 | Body_fat_percentage+NHGRI-EBI_GWAS_catalog+rs1800437,Body_fat_percentage+NHGRI-EBI_GWAS_catalog+rs1800437,Body_mass_index_in_physically_active_individuals+NHGRI-EBI_GWAS_catalog+rs1800437,Body_mass_index__joint_analysis_main_effects_and_physical_activity_interaction_+NHGRI-EBI_GWAS_catalog+rs1800437,Body_mass_index__joint_analysis_main_effects_and_physical_activity_interaction_+NHGRI-EBI_GWAS_catalog+rs1800437,Body_Mass_Index+NHGRI-EBI_GWAS_catalog+rs1800437,Body_Mass_Index+NHGRI-EBI_GWAS_catalog+rs1800437,Body_size_at_age_10+NHGRI-EBI_GWAS_catalog+rs1800437,GIP_levels_in_response_to_oral_glucose_tolerance_test__120_minutes_+NHGRI-EBI_GWAS_catalog+rs1800437,GIP_levels_in_response_to_oral_glucose_tolerance_test__fasting_+NHGRI-EBI_GWAS_catalog+rs1800437,Insulin_levels_in_response_to_oral_glucose_tolerance_test__30_minutes_+NHGRI-EBI_GWAS_catalog+rs1800437,Mean_reticulocyte_volume+NHGRI-EBI_GWAS_catalog+rs1800437,Mean_spheric_corpuscular_volume+NHGRI-EBI_GWAS_catalog+rs1800437,Obesity+NHGRI-EBI_GWAS_catalog+rs1800437,Principal_component-derived_dietary_pattern_29+NHGRI-EBI_GWAS_catalog+rs1800437,Pulse_pressure+NHGRI-EBI_GWAS_catalog+rs1800437,Pulse_pressure+NHGRI-EBI_GWAS_catalog+rs1800437,Random_glucose_levels+NHGRI-EBI_GWAS_catalog+rs1800437,Systolic_blood_pressure+NHGRI-EBI_GWAS_catalog+rs1800437,Two-hour_glucose+NHGRI-EBI_GWAS_catalog+rs1800437,Type_2_diabetes__adjusted_for_BMI_+NHGRI- | 0.0232969223930524 | **34** | 6.054421 | ENSG00000010310 | 0.733 | 3.04E-05 | 2.12E-05 | GO:0002029:desensitization_of_G_protein-coupled_receptor_signaling_pathway,GO:0004888:transmembrane_signaling_receptor_activity,GO:0004930:G_protein-coupled_receptor_activity,GO:0005515:protein_binding,GO:0005886:plasma_membrane,GO:0006091:generation_of_precursor_metabolites_and_energy,GO:0007166:cell_surface_receptor_signaling_pathway,GO:0007186:G_protein-coupled_receptor_signaling_pathway,GO:0007188:adenylate_cyclase-modulating_G_protein-coupled_receptor_signaling_pathway,GO:0007189:adenylate_cyclase-activating_G_protein-coupled_receptor_signaling_pathway,GO:0007190:activation_of_adenylate_cyclase_activity,GO:0007204:positive_regulation_of_cytosolic_calcium_ion_concentration,GO:0007584:response_to_nutrient,GO:0008528:G_protein-coupled_peptide_receptor_activity,GO:0009749:response_to_glucose,GO:0016020:membrane,GO:0016519:gastric_inhibitory_peptide_receptor_activity,GO:0017046:peptide_hormone_binding,GO:0031018:endocrine_pancreas_development,GO:0032024:positive_regulation_of_insulin_secretion,GO:0038192:gastric_inhibitory_peptide_signaling_pathway,GO:0043950:positive_regulation_of_cAMP-mediated_signaling,GO:0048678:response_to_axon_injury,GO:0050796:regulation_of_insulin_secretion,GO:0051592:response_to_calcium_ion,GO:0070542:response_to_fatty_acid,GO:0120022:glucagon_family_peptide_binding | - |
| OSA | rs1800437 | rs10423928 | Acetone_levels+NHGRI-EBI_GWAS_catalog+rs10423928,Adult_body_size+NHGRI-EBI_GWAS_catalog+rs10423928,Blood_urea_nitrogen_levels+NHGRI-EBI_GWAS_catalog+rs10423928,Body_Mass_Index+NHGRI-EBI_GWAS_catalog+rs10423928,Body_Mass_Index+NHGRI-EBI_GWAS_catalog+rs10423928,Body_Mass_Index+NHGRI-EBI_GWAS_catalog+rs10423928,Body_surface_area+NHGRI-EBI_GWAS_catalog+rs10423928,Gall_stone_disease_or_coronary_artery_disease+NHGRI-EBI_GWAS_catalog+rs10423928,Obesity+NHGRI-EBI_GWAS_catalog+rs10423928,Oranges_liking+NHGRI-EBI_GWAS_catalog+rs10423928,Predicted_visceral_adipose_tissue+NHGRI-EBI_GWAS_catalog+rs10423928,Pulse_pressure+NHGRI-EBI_GWAS_catalog+rs10423928,Systolic_blood_pressure+NHGRI-EBI_GWAS_catalog+rs10423928,Systolic_blood_pressure+NHGRI-EBI_GWAS_catalog+rs10423928,Two-hour_glucose_challenge+NHGRI-EBI_GWAS_catalog+rs10423928,Whole_body_fat_free_mass_UKB_data_field_23101+NHGRI-EBI_GWAS_catalog+rs10423928,Whole_body_fat_mass_UKB_data_field_23100+NHGRI-EBI_GWAS_catalog+rs10423928 | - | 4.832 | 0.440235 | ENSG00000010310 | 0.733 | 5.17E-05 | 3.18E-05 | GO:0016442:RISC_complex,GO:0035195:miRNA-mediated_post-transcriptional_gene_silencing | - |
| OSA | rs1800437 | rs10423928 | Acetone_levels+NHGRI-EBI_GWAS_catalog+rs10423928,Adult_body_size+NHGRI-EBI_GWAS_catalog+rs10423928,Blood_urea_nitrogen_levels+NHGRI-EBI_GWAS_catalog+rs10423928,Body_Mass_Index+NHGRI-EBI_GWAS_catalog+rs10423928,Body_Mass_Index+NHGRI-EBI_GWAS_catalog+rs10423928,Body_Mass_Index+NHGRI-EBI_GWAS_catalog+rs10423928,Body_surface_area+NHGRI-EBI_GWAS_catalog+rs10423928,Gall_stone_disease_or_coronary_artery_disease+NHGRI-EBI_GWAS_catalog+rs10423928,Obesity+NHGRI-EBI_GWAS_catalog+rs10423928,Oranges_liking+NHGRI-EBI_GWAS_catalog+rs10423928,Predicted_visceral_adipose_tissue+NHGRI-EBI_GWAS_catalog+rs10423928,Pulse_pressure+NHGRI-EBI_GWAS_catalog+rs10423928,Systolic_blood_pressure+NHGRI-EBI_GWAS_catalog+rs10423928,Systolic_blood_pressure+NHGRI-EBI_GWAS_catalog+rs10423928,Two-hour_glucose_challenge+NHGRI-EBI_GWAS_catalog+rs10423928,Whole_body_fat_free_mass_UKB_data_field_23101+NHGRI-EBI_GWAS_catalog+rs10423928,Whole_body_fat_mass_UKB_data_field_23100+NHGRI-EBI_GWAS_catalog+rs10423928 | - | 4.832 | 0.440235 | ENSG00000010310 | 0.733 | 5.17E-05 | 3.18E-05 | GO:0016442:RISC_complex,GO:0035195:miRNA-mediated_post-transcriptional_gene_silencing | - |

**Supplementary Table 15.** Variant annotation for fine-mapped vari

| Trait | Locus Index Variant | Fine-mapped variant | PHENOTYPES | ClinPred | CADD_PHRED | CADD_RAW | OpenTargets_geneId | OpenTargets_l2g | Enformer_SAD | Enformer_SAR | GO | REVEL |
|---|---|---|---|---|---|---|---|---|---|---|---|---|
| OSA | rs1800437 | rs10423928 | Acetone_levels+NHGRI-EBI_GWAS_catalog+rs10423928,Adult_body_size+NHGRI-EBI_GWAS_catalog+rs10423928,Blood_urea_nitrogen_levels+NHGRI-EBI_GWAS_catalog+rs10423928,Body_Mass_Index+NHGRI-EBI_GWAS_catalog+rs10423928,Body_Mass_Index+NHGRI-EBI_GWAS_catalog+rs10423928,Body_Mass_Index+NHGRI-EBI_GWAS_catalog+rs10423928,Body_Mass_Index+NHGRI-EBI_GWAS_catalog+rs10423928,Body_surface_area+NHGRI-EBI_GWAS_catalog+rs10423928,Gall_stone_disease_or_coronary_artery_disease+NHGRI-EBI_GWAS_catalog+rs10423928,Obesity+NHGRI-EBI_GWAS_catalog+rs10423928,Oranges_liking+NHGRI-EBI_GWAS_catalog+rs10423928,Predicted_visceral_adipose_tissue+NHGRI-EBI_GWAS_catalog+rs10423928,Pulse_pressure+NHGRI-EBI_GWAS_catalog+rs10423928,Systolic_blood_pressure+NHGRI-EBI_GWAS_catalog+rs10423928,Systolic_blood_pressure+NHGRI-EBI_GWAS_catalog+rs10423928,Two-hour_glucose_challenge+NHGRI-EBI_GWAS_catalog+rs10423928,Whole_body_fat_free_mass_UKB_data_field_23101+NHGRI-EBI_GWAS_catalog+rs10423928,Whole_body_fat_mass_UKB_data_field_23100+NHGRI-EBI_GWAS_catalog+rs10423928 | - | 4.832 | 0.440235 | ENSG00000010310 | 0.733 | 5.17E-05 | 3.18E-05 | GO:0002029:desensitization_of_G_protein-coupled_receptor_signaling_pathway,GO:0004888:transmembrane_signaling_receptor_activity,GO:0004930:G_protein-coupled_receptor_activity,GO:0005515:protein_binding,GO:0005886:plasma_membrane,GO:0006091:generation_of_precursor_metabolites_and_energy,GO:0007166:cell_surface_receptor_signaling_pathway,GO:0007186:G_protein-coupled_receptor_signaling_pathway,GO:0007188:adenylate_cyclase-modulating_G_protein-coupled_receptor_signaling_pathway,GO:0007189:adenylate_cyclase-activating_G_protein-coupled_receptor_signaling_pathway,GO:0007190:activation_of_adenylate_cyclase_activity,GO:0007204:positive_regulation_of_cytosolic_calcium_ion_concentration,GO:0007584:response_to_nutrient,GO:0008528:G_protein-coupled_peptide_receptor_activity,GO:0009749:response_to_glucose,GO:0016020:membrane,GO:0016519:gastric_inhibitory_peptide_receptor_activity,GO:0017046:peptide_hormone_binding,GO:0031018:endocrine_pancreas_development,GO:0032024:positive_regulation_of_insulin_secretion,GO:0038192:gastric_inhibitory_peptide_signaling_pathway,GO:0043950:positive_regulation_of_cAMP-mediated_signaling,GO:0048678:response_to_axon_injury,GO:0050796:regulation_of_insulin_secretion,GO:0051592:response_to_calcium_ion,GO:0070542:response_to_fatty_acid,GO:0120022:glucagon_family_peptide_binding | - |
| OSA | rs11088984 | rs12152096 | ADARB1-associated_Microcephaly__Intellectual_Disability__and_Seizures+G2P+ENSG00000197381,NEURODEVELOPMENTAL_DISORDER_WITH_HYPOTONIA__MICROCEPHALY__AND_SEIZURES+MIM_morbid+ENSG00000197381 | - | 3.268 | 0.299641 | ENSG00000197381 | 0.588 | 3.46E-06 | 4.70E-06 | - | - |
| OSA | rs11088984 | rs12152096 | ADARB1-associated_Microcephaly__Intellectual_Disability__and_Seizures+G2P+ENSG00000197381,NEURODEVELOPMENTAL_DISORDER_WITH_HYPOTONIA__MICROCEPHALY__AND_SEIZURES+MIM_morbid+ENSG00000197381 | - | 3.268 | 0.299641 | ENSG00000197381 | 0.588 | 3.46E-06 | 4.70E-06 | - | - |
| OSA | rs11088984 | rs12152096 | ADARB1-associated_Microcephaly__Intellectual_Disability__and_Seizures+G2P+ENSG00000197381,NEURODEVELOPMENTAL_DISORDER_WITH_HYPOTONIA__MICROCEPHALY__AND_SEIZURES+MIM_morbid+ENSG00000197381 | - | 3.268 | 0.299641 | ENSG00000197381 | 0.588 | 3.46E-06 | 4.70E-06 | GO:0003723:RNA_binding,GO:0003725:double-stranded_RNA_binding,GO:0003726:double-stranded_RNA_adenosine_deaminase_activity,GO:0003729:mRNA_binding,GO:0004000:adenosine_deaminase_activity,GO:0005515:protein_binding,GO:0005634:nucleus,GO:0005654:nucleoplasm,GO:0005730:nucleolus,GO:0005737:cytoplasm,GO:0005829:cytosol,GO:0006382:adenosine_to_inosine_editing,GO:0006396:RNA_processing,GO:0006397:mRNA_processing,GO:0007274:neuromuscular_synaptic_transmission,GO:0008251:tRNA-specific_adenosine_deaminase_activity,GO:0008285:negative_regulation_of_cell_population_proliferation,GO:0016553:base_conversion_or_substitution_editing,GO:0016787:hydrolase_activity,GO:0021610:facial_nerve_morphogenesis,GO:0021618:hypoglossal_nerve_morphogenesis,GO:0021965:spinal_cord_ventral_commissure_morphogenesis,GO:0030336:negative_regulation_of_cell_migration,GO:0035264:multicellular_organism_growth,GO:0044387:negative_regulation_of_protein_kinase_activity_by_regulation_of_protein_phosphorylation,GO:0045070:positive_regulation_of_viral_genome_replication,GO:0045087:innate_immune_response,GO:0045202:synapse,GO:0046872:metal_ion_binding,GO:0050884:neuromuscular_process_controlling_posture,GO:0051607:defense_response_to_virus,GO:0051726:regulation_of_cell_cycle,GO:0060384:innervation,GO:0060415:muscle_tissue_morphogenesis,GO:0061744:motor_behavior,GO:0097049:motor_neuron_apoptotic_process | - |
| OSA | rs11088984 | rs2838787 | ADARB1-associated_Microcephaly__Intellectual_Disability__and_Seizures+G2P+ENSG00000197381,NEURODEVELOPMENTAL_DISORDER_WITH_HYPOTONIA__MICROCEPHALY__AND_SEIZURES+MIM_morbid+ENSG00000197381,INSOMNIA+NHGRI-EBI_GWAS_catalog+rs2838787 | - | 0.179 | -0.456472 | ENSG00000197381 | 0.588 | -1.40E-06 | -1.97E-06 | GO:0003723:RNA_binding,GO:0003725:double-stranded_RNA_binding,GO:0003726:double-stranded_RNA_adenosine_deaminase_activity,GO:0003729:mRNA_binding,GO:0004000:adenosine_deaminase_activity,GO:0005515:protein_binding,GO:0005634:nucleus,GO:0005654:nucleoplasm,GO:0005730:nucleolus,GO:0005737:cytoplasm,GO:0005829:cytosol,GO:0006382:adenosine_to_inosine_editing,GO:0006396:RNA_processing,GO:0006397:mRNA_processing,GO:0007274:neuromuscular_synaptic_transmission,GO:0008251:tRNA-specific_adenosine_deaminase_activity,GO:0008285:negative_regulation_of_cell_population_proliferation,GO:0016553:base_conversion_or_substitution_editing,GO:0016787:hydrolase_activity,GO:0021610:facial_nerve_morphogenesis,GO:0021618:hypoglossal_nerve_morphogenesis,GO:0021965:spinal_cord_ventral_commissure_morphogenesis,GO:0030336:negative_regulation_of_cell_migration,GO:0035264:multicellular_organism_growth,GO:0044387:negative_regulation_of_protein_kinase_activity_by_regulation_of_protein_phosphorylation,GO:0045070:positive_regulation_of_viral_genome_replication,GO:0045087:innate_immune_response,GO:0045202:synapse,GO:0046872:metal_ion_binding,GO:0050884:neuromuscular_process_controlling_posture,GO:0051607:defense_response_to_virus,GO:0051726:regulation_of_cell_cycle,GO:0060384:innervation,GO:0060415:muscle_tissue_morphogenesis,GO:0061744:motor_behavior,GO:0097049:motor_neuron_apoptotic_process | - |
| OSA | rs11088984 | rs2838788 | ADARB1-associated_Microcephaly__Intellectual_Disability__and_Seizures+G2P+ENSG00000197381,NEURODEVELOPMENTAL_DISORDER_WITH_HYPOTONIA__MICROCEPHALY__AND_SEIZURES+MIM_morbid+ENSG00000197381,Depression+NHGRI-EBI_GWAS_catalog+rs2838788 | - | 0.299 | -0.323167 | ENSG00000197381 | 0.588 | 1.13E-05 | 1.55E-05 | GO:0003723:RNA_binding,GO:0003725:double-stranded_RNA_binding,GO:0003726:double-stranded_RNA_adenosine_deaminase_activity,GO:0003729:mRNA_binding,GO:0004000:adenosine_deaminase_activity,GO:0005515:protein_binding,GO:0005634:nucleus,GO:0005654:nucleoplasm,GO:0005730:nucleolus,GO:0005737:cytoplasm,GO:0005829:cytosol,GO:0006382:adenosine_to_inosine_editing,GO:0006396:RNA_processing,GO:0006397:mRNA_processing,GO:0007274:neuromuscular_synaptic_transmission,GO:0008251:tRNA-specific_adenosine_deaminase_activity,GO:0008285:negative_regulation_of_cell_population_proliferation,GO:0016553:base_conversion_or_substitution_editing,GO:0016787:hydrolase_activity,GO:0021610:facial_nerve_morphogenesis,GO:0021618:hypoglossal_nerve_morphogenesis,GO:0021965:spinal_cord_ventral_commissure_morphogenesis,GO:0030336:negative_regulation_of_cell_migration,GO:0035264:multicellular_organism_growth,GO:0044387:negative_regulation_of_protein_kinase_activity_by_regulation_of_protein_phosphorylation,GO:0045070:positive_regulation_of_viral_genome_replication,GO:0045087:innate_immune_response,GO:0045202:synapse,GO:0046872:metal_ion_binding,GO:0050884:neuromuscular_process_controlling_posture,GO:0051607:defense_response_to_virus,GO:0051726:regulation_of_cell_cycle,GO:0060384:innervation,GO:0060415:muscle_tissue_morphogenesis,GO:0061744:motor_behavior,GO:0097049:motor_neuron_apoptotic_process | - |
| OSA | rs11088984 | rs2838813 | ADARB1-associated_Microcephaly__Intellectual_Disability__and_Seizures+G2P+ENSG00000197381,NEURODEVELOPMENTAL_DISORDER_WITH_HYPOTONIA__MICROCEPHALY__AND_SEIZURES+MIM_morbid+ENSG00000197381 | - | 0.079 | -0.676650 | ENSG00000197381 | 0.697 | - | - | GO:0003723:RNA_binding,GO:0003725:double-stranded_RNA_binding,GO:0003726:double-stranded_RNA_adenosine_deaminase_activity,GO:0003729:mRNA_binding,GO:0004000:adenosine_deaminase_activity,GO:0005515:protein_binding,GO:0005634:nucleus,GO:0005654:nucleoplasm,GO:0005730:nucleolus,GO:0005737:cytoplasm,GO:0005829:cytosol,GO:0006382:adenosine_to_inosine_editing,GO:0006396:RNA_processing,GO:0006397:mRNA_processing,GO:0007274:neuromuscular_synaptic_transmission,GO:0008251:tRNA-specific_adenosine_deaminase_activity,GO:0008285:negative_regulation_of_cell_population_proliferation,GO:0016553:base_conversion_or_substitution_editing,GO:0016787:hydrolase_activity,GO:0021610:facial_nerve_morphogenesis,GO:0021618:hypoglossal_nerve_morphogenesis,GO:0021965:spinal_cord_ventral_commissure_morphogenesis,GO:0030336:negative_regulation_of_cell_migration,GO:0035264:multicellular_organism_growth,GO:0044387:negative_regulation_of_protein_kinase_activity_by_regulation_of_protein_phosphorylation,GO:0045070:positive_regulation_of_viral_genome_replication,GO:0045087:innate_immune_response,GO:0045202:synapse,GO:0046872:metal_ion_binding,GO:0050884:neuromuscular_process_controlling_posture,GO:0051607:defense_response_to_virus,GO:0051726:regulation_of_cell_cycle,GO:0060384:innervation,GO:0060415:muscle_tissue_morphogenesis,GO:0061744:motor_behavior,GO:0097049:motor_neuron_apoptotic_process | - |
| OSA | rs11088984 | rs11088984 | - | - | 0.232 | -0.388179 | - | - | - | - | - | - |
| OSA | rs11088984 | rs11088984 | - | - | 0.232 | -0.388179 | - | - | - | - | - | - |
| OSA, OSAadjBMI | rs2958127, rs4759254 | rs12828563 | - | - | 3.102 | 0.284145 | - | - | - | - | GO:0003677:DNA_binding,GO:0005634:nucleus,GO:0005737:cytoplasm,GO:0005854:nascent_polypeptide-associated_complex,GO:0006612:protein_targeting_to_membrane,GO:0051082:unfolded_protein_binding | - |
| OSA, OSAadjBMI | rs2958127, rs4759255 | rs12828563 | - | - | 3.102 | 0.284145 | - | - | - | - | - | - |
| OSA, OSAadjBMI | rs2958127, rs4759255 | rs2926748 | - | - | 4.759 | 0.433466 | - | - | - | - | - | - |
| OSA, OSAadjBMI | rs2958127, rs4759255 | rs2926748 | - | - | 4.759 | 0.433466 | - | - | - | - | GO:0003677:DNA_binding,GO:0005634:nucleus,GO:0005737:cytoplasm,GO:0005854:nascent_polypeptide-associated_complex,GO:0006612:protein_targeting_to_membrane,GO:0051082:unfolded_protein_binding | - |
| OSA, OSAadjBMI | rs2958127, rs4759255 | rs11171951 | - | - | 0.890 | -0.047938 | ENSG00000196531 | 0.513 | - | - | GO:0003677:DNA_binding,GO:0005634:nucleus,GO:0005737:cytoplasm,GO:0005854:nascent_polypeptide-associated_complex,GO:0006612:protein_targeting_to_membrane,GO:0051082:unfolded_protein_binding | - |
| OSA, OSAadjBMI | rs2958127, rs4759255 | rs2958127 | BACH1/FOXO1_protein_level_ratio+NHGRI-EBI_GWAS_catalog+rs2958127,BACH1/SNAP23_protein_level_ratio+NHGRI-EBI_GWAS_catalog+rs2958127 | 0.0236134595008294 | 3.494 | 0.320202 | ENSG00000196531 | 0.558 | - | - | GO:0003677:DNA_binding,GO:0005634:nucleus,GO:0005737:cytoplasm,GO:0005854:nascent_polypeptide-associated_complex,GO:0006612:protein_targeting_to_membrane,GO:0051082:unfolded_protein_binding | 0.012 |
| OSAadjBMI | rs2964006 | rs7709205 | GRIA1-related_neurodevelopmental_disorder+G2P+ENSG00000155511,Intellectual_developmental_disorder__autosomal_dominant_67+MIM_morbid+ENSG00000155511,INTELLECTUAL_DEVELOPMENTAL_DISORDER__AUTOSOMAL_RECESSIVE_76+MIM_morbid+ENSG00000155511,Autosomal_dominant_non-syndromic_intellectual_disability+Orphanet+ENSG00000155511,Autosomal_recessive_non-syndromic_intellectual_disability+Orphanet+ENSG00000155511 | - | 1.476 | 0.083856 | ENSG00000155511 | 0.683 | - | - | - | - |
| OSAadjBMI | rs2964006 | rs7709205 | GRIA1-related_neurodevelopmental_disorder+G2P+ENSG00000155511,Intellectual_developmental_disorder__autosomal_dominant_67+MIM_morbid+ENSG00000155511,INTELLECTUAL_DEVELOPMENTAL_DISORDER__AUTOSOMAL_RECESSIVE_76+MIM_morbid+ENSG00000155511,Autosomal_dominant_non-syndromic_intellectual_disability+Orphanet+ENSG00000155511,Autosomal_recessive_non-syndromic_intellectual_disability+Orphanet+ENSG00000155511 | - | 1.476 | 0.083856 | ENSG00000155511 | 0.683 | - | - | GO:0001540:amyloid-beta_binding,GO:0001919:regulation_of_receptor_recycling,GO:0001965:G-protein_alpha-subunit_binding,GO:0004970:glutamate-gated_receptor_activity,GO:0004971:AMPA_glutamate_receptor_activity,GO:0005216:monoatomic_ion_channel_activity,GO:0005515:protein_binding,GO:0005768:endosome,GO:0005769:early_endosome,GO:0005783:endoplasmic_reticulum,GO:0005789:endoplasmic_reticulum_membrane,GO:0005829:cytosol,GO:0005886:plasma_membrane,GO:0005911:cell-cell_junction,GO:0006811:monoatomic_ion_transport,GO:0007165:signal_transduction,GO:0007215:glutamate_receptor_signaling_pathway,GO:0007268:chemical_synaptic_transmission,GO:0007416:synapse_assembly,GO:0007616:long-term_memory,GO:0008021:synaptic_vesicle,GO:0008066:glutamate_receptor_activity,GO:0008179:adenylate_cyclase_binding,GO:0008328:ionotropic_glutamate_receptor_complex,GO:0009410:response_to_xenobiotic_stimulus,GO:0009636:response_to_toxic_substance,GO:0009744:response_to_sucrose,GO:0009897:external_side_of_plasma_membrane,GO:0009986:cell_surface,GO:0010226:response_to_lithium_ion,GO:0010628:positive_regulation_of_gene_expression,GO:0012507:ER_to_Golgi_transport_vesicle_membrane,GO:0014069:postsynaptic_density,GO:0014070:response_to_organic_cyclic_compound,GO:0015276:ligand-gated_monoatomic_ion_channel_activity,GO:0016020:membrane,GO:0019228:neuronal_action_potential,GO:0019722:calcium-mediated_signaling,GO:0019865:immunoglobulin_binding,GO:0019901:protein_kinase_binding,GO:0019904:protein_domain_specific_binding,GO:0021510:spinal_cord_development,GO:0021987:cerebral_cortex_development,GO:0022824:transmitter-gated_monoatomic_ion_channel_activity,GO:0022849:glutamate-gated_calcium_ion_channel_activity,GO:0030165:PDZ_domain_binding,GO:0030425:dendrite,GO:0030666:endocytic_vesicle_membrane,GO:0030672:synaptic_vesicle_membrane,GO:0031090:organelle_membrane,GO:0031267:small_GTPase_binding,GO:0031489:myosi | - |

**Supplementary Table 15.** Variant annotation for fine-mapped vari

| Trait | Locus Index Variant | Fine-mapped variant | PHENOTYPES | ClinPred | CADD_PHRED | CADD_RAW | OpenTargets_geneId | OpenTargets_l2g | Enformer_SAD | Enformer_SAR | GO | REVEL |
|---|---|---|---|---|---|---|---|---|---|---|---|---|
| OSAadjBMI | rs2964006 | rs2126165 | GRIA1-related_neurodevelopmental_disorder+G2P+ENSG00000155511,Intellectual_developmental_disorder__autosomal_dominant_67+MIM_morbid+ENSG00000155511,INTELLECTUAL_DEVELOPMENTAL_DISORDER__AUTOSOMAL_RECESSIVE_76+MIM_morbid+ENSG00000155511,Autosomal_dominant_non-syndromic_intellectual_disability+Orphanet+ENSG00000155511,Autosomal_recessive_non-syndromic_intellectual_disability+Orphanet+ENSG00000155511,Hand_grip_strength+NHGRI-EBI_GWAS_catalog+rs2126165 | - | 0.356 | -0.278857 | ENSG00000155511 | 0.683 | - | - | GO:0001540:amyloid-beta_binding,GO:0001919:regulation_of_receptor_recycling,GO:0001965:G-protein_alpha-subunit_binding,GO:0004970:glutamate-gated_receptor_activity,GO:0004971:AMPA_glutamate_receptor_activity,GO:0005216:monoatomic_ion_channel_activity,GO:0005515:protein_binding,GO:0005768:endosome,GO:0005769:early_endosome,GO:0005783:endoplasmic_reticulum,GO:0005789:endoplasmic_reticulum_membrane,GO:0005829:cytosol,GO:0005886:plasma_membrane,GO:0005911:cell-cell_junction,GO:0006811:monoatomic_ion_transport,GO:0007165:signal_transduction,GO:0007215:glutamate_receptor_signaling_pathway,GO:0007268:chemical_synaptic_transmission,GO:0007416:synapse_assembly,GO:0007616:long-term_memory,GO:0008021:synaptic_vesicle,GO:0008066:glutamate_receptor_activity,GO:0008179:adenylate_cyclase_binding,GO:0008328:ionotropic_glutamate_receptor_complex,GO:0009410:response_to_xenobiotic_stimulus,GO:0009636:response_to_toxic_substance,GO:0009744:response_to_sucrose,GO:0009897:external_side_of_plasma_membrane,GO:0009986:cell_surface,GO:0010226:response_to_lithium_ion,GO:0010628:positive_regulation_of_gene_expression,GO:0012507:ER_to_Golgi_transport_vesicle_membrane,GO:0014069:postsynaptic_density,GO:0014070:response_to_organic_cyclic_compound,GO:0015276:ligand-gated_monoatomic_ion_channel_activity,GO:0016020:membrane,GO:0019228:neuronal_action_potential,GO:0019722:calcium-mediated_signaling,GO:0019865:immunoglobulin_binding,GO:0019901:protein_kinase_binding,GO:0019904:protein_domain_specific_binding,GO:0021510:spinal_cord_development,GO:0021987:cerebral_cortex_development,GO:0022824:transmitter-gated_monoatomic_ion_channel_activity,GO:0022849:glutamate-gated_calcium_ion_channel_activity,GO:0030165:PDZ_domain_binding,GO:0030425:dendrite,GO:0030666:endocytic_vesicle_membrane,GO:0030672:synaptic_vesicle_membrane,GO:0031090:organelle_membrane,GO:0031267:small_GTPase_binding,GO:0031489:myosi | - |
| OSAadjBMI | rs2964006 | rs3021428 | - | - | 6.647 | 0.620216 | ENSG00000155511 | 0.683 | - | - | - | - |
| OSAadjBMI | rs2964006 | rs2926861 | - | - | 1.352 | 0.060897 | ENSG00000155511 | 0.683 | - | - | - | - |
| OSAadjBMI | rs2964006 | rs2964006 | - | - | 0.770 | -0.084888 | ENSG00000155511 | 0.683 | - | - | - | - |
| OSAadjBMI | rs60791051 | rs73649650 | - | - | 3.172 | 0.290765 | - | - | 1.74E-05 | 2.33E-05 | GO:0003674:molecular_function,GO:0005515:protein_binding,GO:0005575:cellular_component,GO:0005768:endosome,GO:0005886:plasma_membrane,GO:0008150:biological_process,GO:0016020:membrane,GO:0022401:negative_adaptation_of_signaling_pathway,GO:0031623:receptor_internalization,GO:0031901:early_endosome_membrane,GO:0031902:late_endosome_membrane,GO:0038160:CXCL12-activated_CXCR4_signaling_pathway,GO:0055038:recycling_endosome_membrane,GO:1990757:ubiquitin_ligase_activator_activity | - |
| OSAadjBMI | rs60791051 | rs60791051 | - | - | 4.884 | 0.445010 | - | - | 7.33E-06 | 1.04E-05 | GO:0003674:molecular_function,GO:0005515:protein_binding,GO:0005575:cellular_component,GO:0005768:endosome,GO:0005886:plasma_membrane,GO:0008150:biological_process,GO:0016020:membrane,GO:0022401:negative_adaptation_of_signaling_pathway,GO:0031623:receptor_internalization,GO:0031901:early_endosome_membrane,GO:0031902:late_endosome_membrane,GO:0038160:CXCL12-activated_CXCR4_signaling_pathway,GO:0055038:recycling_endosome_membrane,GO:1990757:ubiquitin_ligase_activator_activity | - |
| OSAadjBMI | rs4759254 | rs4759254 | Medication_use_for_hypertension_number_of_purchases+NHGRI-EBI_GWAS_catalog+rs4759254 | - | 2.602 | 0.234561 | ENSG00000198056 | 0.529 | - | - | - | - |
| OSAadjBMI | rs10735928 | rs10784447 | DEAFNESS__AUTOSOMAL_RECESSIVE_74+MIM_morbid+ENSG00000174099,Rare_autosomal_recessive_non-syndromic_sensorineural_deafness_type_DFNB+Orphanet+ENSG00000174099,Brain_region_volumes+NHGRI-EBI_GWAS_catalog+rs10784447,Edge-level_brain_connectivity_multivariate_analysis+NHGRI-EBI_GWAS_catalog+rs10784447 | - | **19.64** | 2.647949 | ENSG00000174099 | **0.819** | - | - | GO:0005515:protein_binding,GO:0005737:cytoplasm,GO:0005739:mitochondrion,GO:0005783:endoplasmic_reticulum,GO:0005829:cytosol,GO:0006979:response_to_oxidative_stress,GO:0008270:zinc_ion_binding,GO:0016491:oxidoreductase_activity,GO:0016671:oxidoreductase_activity_-_acting_on_a_sulfur_group_of_donors_-_disulfide_as_acceptor,GO:0030091:protein_repair,GO:0033743:peptide-methionine_(R)-S-oxide_reductase_activity,GO:0033745:L-methionine-(R)-S-oxide_reductase_activity,GO:0046872:metal_ion_binding | - |
| OSAadjBMI | rs10735928 | rs10735928 | DEAFNESS__AUTOSOMAL_RECESSIVE_74+MIM_morbid+ENSG00000174099,Rare_autosomal_recessive_non-syndromic_sensorineural_deafness_type_DFNB+Orphanet+ENSG00000174099 | - | 5.167 | 0.471573 | ENSG00000174099 | **0.819** | - | - | GO:0005515:protein_binding,GO:0005737:cytoplasm,GO:0005739:mitochondrion,GO:0005783:endoplasmic_reticulum,GO:0005829:cytosol,GO:0006979:response_to_oxidative_stress,GO:0008270:zinc_ion_binding,GO:0016491:oxidoreductase_activity,GO:0016671:oxidoreductase_activity_-_acting_on_a_sulfur_group_of_donors_-_disulfide_as_acceptor,GO:0030091:protein_repair,GO:0033743:peptide-methionine_(R)-S-oxide_reductase_activity,GO:0033745:L-methionine-(R)-S-oxide_reductase_activity,GO:0046872:metal_ion_binding | - |
| OSAadjBMI | rs10735928 | rs4332569 | DEAFNESS__AUTOSOMAL_RECESSIVE_74+MIM_morbid+ENSG00000174099,Rare_autosomal_recessive_non-syndromic_sensorineural_deafness_type_DFNB+Orphanet+ENSG00000174099 | - | 0.382 | -0.260402 | ENSG00000174099 | **0.819** | - | - | GO:0005515:protein_binding,GO:0005737:cytoplasm,GO:0005739:mitochondrion,GO:0005783:endoplasmic_reticulum,GO:0005829:cytosol,GO:0006979:response_to_oxidative_stress,GO:0008270:zinc_ion_binding,GO:0016491:oxidoreductase_activity,GO:0016671:oxidoreductase_activity_-_acting_on_a_sulfur_group_of_donors_-_disulfide_as_acceptor,GO:0030091:protein_repair,GO:0033743:peptide-methionine_(R)-S-oxide_reductase_activity,GO:0033745:L-methionine-(R)-S-oxide_reductase_activity,GO:0046872:metal_ion_binding | - |
| OSAadjBMI | rs10735928 | rs1826663 | DEAFNESS__AUTOSOMAL_RECESSIVE_74+MIM_morbid+ENSG00000174099,Rare_autosomal_recessive_non-syndromic_sensorineural_deafness_type_DFNB+Orphanet+ENSG00000174099,Obstructive_sleep_apnea_BMI_adjusted+NHGRI-EBI_GWAS_catalog+rs1826663 | - | 4.432 | 0.403746 | ENSG00000174099 | **0.819** | - | - | GO:0005515:protein_binding,GO:0005737:cytoplasm,GO:0005739:mitochondrion,GO:0005783:endoplasmic_reticulum,GO:0005829:cytosol,GO:0006979:response_to_oxidative_stress,GO:0008270:zinc_ion_binding,GO:0016491:oxidoreductase_activity,GO:0016671:oxidoreductase_activity_-_acting_on_a_sulfur_group_of_donors_-_disulfide_as_acceptor,GO:0030091:protein_repair,GO:0033743:peptide-methionine_(R)-S-oxide_reductase_activity,GO:0033745:L-methionine-(R)-S-oxide_reductase_activity,GO:0046872:metal_ion_binding | - |

**Supplementary Table 16.** Prioritized effector genes from variant to gene (V2G) mapping analysis.

| Trait | Discovery Study | Sentinel rsID | Proxy rsID | Gene Symbol | Gene Id | Gene Biotype | Cell | Contact Score (Chicago Score\|-log10(FitHiC2 FDR) |
|---|---|---|---|---|---|---|---|---|
| OSAadjBMI | EUR | rs12227117 | rs2958153 | *ATP5B* | ENSG00000110955.4 | protein_coding | hESC_HypothalamicNeuron | 18.15 |
| OSAadjBMI | EUR | rs12227117 | rs2958153 | *U59B* | U59B | snoRNA | hESC_HypothalamicNeuron | 18.15 |
| OSAadjBMI | EUR | rs12227117 | rs2958153 | *SNORD59A* | ENSG00000207031.1 | snoRNA | hESC_HypothalamicNeuron | 18.15 |
| OSAadjBMI | EUR | rs12227117 | rs941207 | *BAZ2A* | ENSG00000076108.7 | protein_coding | hMSC_Adipocyte | 14.07 |
| OSAadjBMI | EUR | rs12227117 | rs941207 | *BAZ2A* | ENSG00000076108.7 | protein_coding | hESC_HypothalamicNeuron | 13.25 |
| OSAadjBMI | EUR | rs12227117 | rs2958153 | *BAZ2A* | ENSG00000076108.7 | protein_coding | iPSC_Hepatocytes | 13.23 |
| OSAadjBMI | EUR | rs12227117 | rs941207 | *RBMS2* | ENSG00000076067.7 | protein_coding | SGBS_Diff | 12.12 |
| OSAadjBMI | EUR | rs12227117 | rs941207 | *ATP5B* | ENSG00000110955.4 | protein_coding | GCB | 11.88 |
| OSAadjBMI | EUR | rs12227117 | rs941207 | *SNORD59A* | ENSG00000207031.1 | snoRNA | GCB | 11.88 |
| OSAadjBMI | EUR | rs12227117 | rs941207 | *U59B* | U59B | snoRNA | GCB | 11.88 |
| OSAadjBMI | EUR | rs12227117 | rs941207 | *ATP5B* | ENSG00000110955.4 | protein_coding | hESC_HypothalamicNeuron | 11.73 |
| OSAadjBMI | EUR | rs12227117 | rs941207 | *U59B* | U59B | snoRNA | hESC_HypothalamicNeuron | 11.73 |
| OSAadjBMI | EUR | rs12227117 | rs941207 | *SNORD59A* | ENSG00000207031.1 | snoRNA | hESC_HypothalamicNeuron | 11.73 |
| OSAadjBMI | EUR | rs12227117 | rs2958153 | *BAZ2A* | ENSG00000076108.7 | protein_coding | NaiveT | 11.48 |
| OSAadjBMI | EUR | rs12227117 | rs2958153 | *ATP5B* | ENSG00000110955.4 | protein_coding | iPSC_NPC | 11.45 |
| OSAadjBMI | EUR | rs12227117 | rs2958153 | *U59B* | U59B | snoRNA | iPSC_NPC | 11.45 |
| OSAadjBMI | EUR | rs12227117 | rs2958153 | *SNORD59A* | ENSG00000207031.1 | snoRNA | iPSC_NPC | 11.45 |
| OSAadjBMI | EUR | rs12227117 | rs2958153 | *NACA* | ENSG00000196531.6 | protein_coding | iPSC_Hepatocytes | 10.56 |
| OSAadjBMI | EUR | rs12227117 | rs941207 | *RBMS2* | ENSG00000076067.7 | protein_coding | iPSC_NPC | 10.48 |
| OSAadjBMI | EUR | rs12227117 | rs941207 | *BAZ2A* | ENSG00000076108.7 | protein_coding | NaiveB | 10.34 |
| OSAadjBMI | EUR | rs12227117 | rs2958153 | *NACA* | ENSG00000196531.6 | protein_coding | GCB | 10.29 |
| OSAadjBMI | EUR | rs12227117 | rs941207 | *BAZ2A* | ENSG00000076108.7 | protein_coding | GCB | 9.91 |
| OSAadjBMI | EUR | rs12227117 | rs2958153 | *NACA* | ENSG00000196531.6 | protein_coding | TFH | 9.58 |
| OSAadjBMI | EUR | rs12227117 | rs2958153 | *NACA* | ENSG00000196531.6 | protein_coding | hMSC_Adipocyte | 9.57 |
| OSAadjBMI | EUR | rs12227117 | rs941207 | *ATP5B* | ENSG00000110955.4 | protein_coding | TFH | 9.57 |
| OSAadjBMI | EUR | rs12227117 | rs941207 | *SNORD59A* | ENSG00000207031.1 | snoRNA | TFH | 9.57 |
| OSAadjBMI | EUR | rs12227117 | rs941207 | *U59B* | U59B | snoRNA | TFH | 9.57 |
| OSAadjBMI | EUR | rs12227117 | rs2958153 | *ATP5B* | ENSG00000110955.4 | protein_coding | iPSC_Hepatocytes | 9.48 |
| OSAadjBMI | EUR | rs12227117 | rs2958153 | *SNORD59A* | ENSG00000207031.1 | snoRNA | iPSC_Hepatocytes | 9.48 |
| OSAadjBMI | EUR | rs12227117 | rs2958153 | *U59B* | U59B | snoRNA | iPSC_Hepatocytes | 9.48 |
| OSAadjBMI | EUR | rs12227117 | rs941207 | *RBMS2* | ENSG00000076067.7 | protein_coding | hMSC_Adipocyte | 9.43 |
| OSAadjBMI | EUR | rs12227117 | rs941207 | *RBMS2* | ENSG00000076067.7 | protein_coding | hESC_HypothalamicNeuron | 9.42 |
| OSAadjBMI | EUR | rs12227117 | rs941207 | *NACA* | ENSG00000196531.6 | protein_coding | NaiveT | 9.25 |
| OSAadjBMI | EUR | rs12227117 | rs2958153 | *ATP5B* | ENSG00000110955.4 | protein_coding | hMSC_Osteoblasts | 9.21 |
| OSAadjBMI | EUR | rs12227117 | rs2958153 | *SNORD59A* | ENSG00000207031.1 | snoRNA | hMSC_Osteoblasts | 9.21 |
| OSAadjBMI | EUR | rs12227117 | rs2958153 | *U59B* | U59B | snoRNA | hMSC_Osteoblasts | 9.21 |
| OSAadjBMI | EUR | rs12227117 | rs2958153 | *ATP5B* | ENSG00000110955.4 | protein_coding | SGBS_Diff | 9.12 |
| OSAadjBMI | EUR | rs12227117 | rs2958153 | *U59B* | U59B | snoRNA | SGBS_Diff | 9.12 |
| OSAadjBMI | EUR | rs12227117 | rs2958153 | *SNORD59A* | ENSG00000207031.1 | snoRNA | SGBS_Diff | 9.12 |
| OSAadjBMI | EUR | rs12227117 | rs2958153 | *NACA* | ENSG00000196531.6 | protein_coding | NaiveT | 9.01 |
| OSAadjBMI | EUR | rs12227117 | rs2958153 | *NACA* | ENSG00000196531.6 | protein_coding | SGBS_Diff | 8.85 |
| OSAadjBMI | EUR | rs12227117 | rs941207 | *ATP5B* | ENSG00000110955.4 | protein_coding | NaiveB | 8.83 |
| OSAadjBMI | EUR | rs12227117 | rs941207 | *SNORD59A* | ENSG00000207031.1 | snoRNA | NaiveB | 8.83 |
| OSAadjBMI | EUR | rs12227117 | rs941207 | *BAZ2A* | ENSG00000076108.7 | protein_coding | TFH | 8.75 |
| OSAadjBMI | EUR | rs12227117 | rs941207 | *RBMS2* | ENSG00000076067.7 | protein_coding | iPSC_Neurons | 8.74 |
| OSAadjBMI | EUR | rs12227117 | rs2958153 | *NACA* | ENSG00000196531.6 | protein_coding | iPSC_NPC | 8.68 |
| OSAadjBMI | EUR | rs12227117 | rs941207 | *BAZ2A* | ENSG00000076108.7 | protein_coding | iPSC_NPC | 8.63 |
| OSAadjBMI | EUR | rs12227117 | rs941207 | *ZBTB39* | ENSG00000166860.2 | protein_coding | hESC_HypothalamicNeuron | 8.22 |
| OSAadjBMI | EUR | rs12227117 | rs2958153 | *ZBTB39* | ENSG00000166860.2 | protein_coding | iPSC_Hepatocytes | 8.21 |
| OSAadjBMI | EUR | rs12227117 | rs2958153 | *NACA* | ENSG00000196531.6 | protein_coding | hMSC_Osteoblasts | 8.08 |
| OSAadjBMI | EUR | rs12227117 | rs2958153 | *BAZ2A* | ENSG00000076108.7 | protein_coding | hMSC_Osteoblasts | 8 |
| OSAadjBMI | EUR | rs12227117 | rs941207 | *NACA* | ENSG00000196531.6 | protein_coding | TFH | 7.64 |
| OSAadjBMI | EUR | rs12227117 | rs2958153 | *ZBTB39* | ENSG00000166860.2 | protein_coding | iPSC_Neurons | 7.57 |
| OSAadjBMI | EUR | rs12227117 | rs2958153 | *NACA* | ENSG00000196531.6 | protein_coding | hESC_HypothalamicNeuron | 7.5 |
| OSAadjBMI | EUR | rs12227117 | rs941207 | *BAZ2A* | ENSG00000076108.7 | protein_coding | SGBS_Diff | 7.34 |
| OSAadjBMI | EUR | rs12227117 | rs2958153 | *ATP5B* | ENSG00000110955.4 | protein_coding | hESC_HypothalamicNPC | 7.33 |
| OSAadjBMI | EUR | rs12227117 | rs2958153 | *SNORD59A* | ENSG00000207031.1 | snoRNA | hESC_HypothalamicNPC | 7.33 |
| OSAadjBMI | EUR | rs12227117 | rs2958153 | *U59B* | U59B | snoRNA | hESC_HypothalamicNPC | 7.33 |
| OSAadjBMI | EUR | rs12227117 | rs941207 | *NACA* | ENSG00000196531.6 | protein_coding | hMSC_Adipocyte | 7.33 |
| OSAadjBMI | EUR | rs12227117 | rs2958153 | *NACA* | ENSG00000196531.6 | protein_coding | NaiveB | 7.22 |
| OSAadjBMI | EUR | rs12227117 | rs941207 | *ATP5B* | ENSG00000110955.4 | protein_coding | SGBS_Diff | 7.22 |
| OSAadjBMI | EUR | rs12227117 | rs941207 | *SNORD59A* | ENSG00000207031.1 | snoRNA | SGBS_Diff | 7.22 |
| OSAadjBMI | EUR | rs12227117 | rs941207 | *U59B* | U59B | snoRNA | SGBS_Diff | 7.22 |
| OSAadjBMI | EUR | rs12227117 | rs941207 | *ESYT1* | ENSG00000139641.8 | protein_coding | iPSC_NPC | 7.09 |
| OSAadjBMI | EUR | rs12227117 | rs941207 | *ZC3H10* | ENSG00000135482.2 | protein_coding | iPSC_NPC | 7.09 |
| OSAadjBMI | EUR | rs12227117 | rs941207 | *MIP* | ENSG00000135517.6 | protein_coding | iPSC_NPC | 7.05 |
| OSAadjBMI | EUR | rs12227117 | rs941207 | *SPRYD4* | ENSG00000176422.10 | protein_coding | iPSC_NPC | 7.05 |
| OSAadjBMI | EUR | rs12227117 | rs941207 | *NAB2* | ENSG00000166886.8 | protein_coding | iPSC_NPC | 7.02 |
| OSAadjBMI | EUR | rs12227117 | rs941207 | TCONS_00020445 | TCONS_00020445 | lincRNA | iPSC_NPC | 7.02 |
| OSAadjBMI | EUR | rs12227117 | rs941207 | TCONS_00020805 | TCONS_00020805 | lincRNA | iPSC_NPC | 7.02 |
| OSAadjBMI | EUR | rs12227117 | rs941207 | TCONS_00020806 | TCONS_00020806 | lincRNA | iPSC_NPC | 7.02 |
| OSAadjBMI | EUR | rs12227117 | rs941207 | TCONS_00020807 | TCONS_00020807 | lincRNA | iPSC_NPC | 7.02 |
| OSAadjBMI | EUR | rs12227117 | rs941207 | TCONS_00021326 | TCONS_00021326 | lincRNA | iPSC_NPC | 7.02 |
| OSAadjBMI | EUR | rs12227117 | rs941207 | TCONS_00021327 | TCONS_00021327 | lincRNA | iPSC_NPC | 7.02 |
| OSAadjBMI | EUR | rs12227117 | rs941207 | *TMEM194A* | ENSG00000166881.5 | protein_coding | iPSC_NPC | 7.02 |
| OSAadjBMI | EUR | rs12227117 | rs2958153 | *ZBTB39* | ENSG00000166860.2 | protein_coding | GCB | 6.98 |
| OSAadjBMI | EUR | rs12227117 | rs941207 | *COQ10A* | ENSG00000135469.9 | protein_coding | hESC_HypothalamicNeuron | 6.81 |
| OSAadjBMI | EUR | rs12227117 | rs941207 | *ESYT1* | ENSG00000139641.8 | protein_coding | NaiveB | 6.8 |
| OSAadjBMI | EUR | rs12227117 | rs941207 | RP11-603J24.6 | ENSG00000258345.1 | antisense | NaiveB | 6.8 |
| OSAadjBMI | EUR | rs12227117 | rs941207 | *ZC3H10* | ENSG00000135482.2 | protein_coding | NaiveB | 6.8 |
| OSAadjBMI | EUR | rs12227117 | rs2958153 | *ATP5B* | ENSG00000110955.4 | protein_coding | GCB | 6.75 |
| OSAadjBMI | EUR | rs12227117 | rs2958153 | *SNORD59A* | ENSG00000207031.1 | snoRNA | GCB | 6.75 |
| OSAadjBMI | EUR | rs12227117 | rs2958153 | *U59B* | U59B | snoRNA | GCB | 6.75 |
| OSAadjBMI | EUR | rs12227117 | rs941207 | *RBMS2* | ENSG00000076067.7 | protein_coding | hESC_HypothalamicNPC | 6.74 |
| OSAadjBMI | EUR | rs12227117 | rs2958153 | *NACA* | ENSG00000196531.6 | protein_coding | iPSC_Neurons | 6.71 |

| **Supplementary Table 16.** Prioritized effector genes from variant to gene (V2G) mapping analysis. | | | | | | | | |
|---|---|---|---|---|---|---|---|---|
| **Trait** | **Discovery Study** | **Sentinel rsID** | **Proxy rsID** | **Gene Symbol** | **Gene Id** | **Gene Biotype** | **Cell** | **Contact Score (Chicago Score\|-log10(FitHiC2 FDR)** |
| OSAadjBMI | EUR | rs12227117 | rs941207 | *STAT6* | ENSG00000166888.6 | protein_coding | iPSC_NPC | 6.71 |
| OSAadjBMI | EUR | rs12227117 | rs941207 | RP11-603J24.6 | ENSG00000258345.1 | antisense | iPSC_NPC | 6.69 |
| OSAadjBMI | EUR | rs12227117 | rs2958153 | *ZBTB39* | ENSG00000166860.2 | protein_coding | hESC_HypothalamicNeuron | 6.68 |
| OSAadjBMI | EUR | rs12227117 | rs2958153 | *BAZ2A* | ENSG00000076108.7 | protein_coding | iPSC_NPC | 6.65 |
| OSAadjBMI | EUR | rs12227117 | rs941207 | *ATP5B* | ENSG00000110955.4 | protein_coding | NaiveT | 6.4 |
| OSAadjBMI | EUR | rs12227117 | rs941207 | *U59B* | U59B | snoRNA | NaiveT | 6.4 |
| OSAadjBMI | EUR | rs12227117 | rs941207 | *SNORD59A* | ENSG00000207031.1 | snoRNA | NaiveT | 6.4 |
| OSAadjBMI | EUR | rs12227117 | rs2958153 | *BAZ2A* | ENSG00000076108.7 | protein_coding | TFH | 6.25 |
| OSAadjBMI | EUR | rs12227117 | rs2958153 | *ZBTB39* | ENSG00000166860.2 | protein_coding | hMSC_Osteoblasts | 6.17 |
| OSAadjBMI | EUR | rs12227117 | rs2958153 | *ATP5B* | ENSG00000110955.4 | protein_coding | hMSC_Adipocyte | 6.14 |
| OSAadjBMI | EUR | rs12227117 | rs2958153 | *SNORD59A* | ENSG00000207031.1 | snoRNA | hMSC_Adipocyte | 6.14 |
| OSAadjBMI | EUR | rs12227117 | rs2958153 | *U59B* | U59B | snoRNA | hMSC_Adipocyte | 6.14 |
| OSAadjBMI | EUR | rs12227117 | rs2958153 | *ATP5B* | ENSG00000110955.4 | protein_coding | NaiveT | 6.07 |
| OSAadjBMI | EUR | rs12227117 | rs2958153 | *SNORD59A* | ENSG00000207031.1 | snoRNA | NaiveT | 6.07 |
| OSAadjBMI | EUR | rs12227117 | rs2958153 | *U59B* | U59B | snoRNA | NaiveT | 6.07 |
| OSAadjBMI | EUR | rs12227117 | rs941207 | *ATP5B* | ENSG00000110955.4 | protein_coding | hESC_HypothalamicNPC | 5.94 |
| OSAadjBMI | EUR | rs12227117 | rs941207 | *SNORD59A* | ENSG00000207031.1 | snoRNA | hESC_HypothalamicNPC | 5.94 |
| OSAadjBMI | EUR | rs12227117 | rs941207 | *U59B* | U59B | snoRNA | hESC_HypothalamicNPC | 5.94 |
| OSAadjBMI | EUR | rs12227117 | rs2958153 | *BAZ2A* | ENSG00000076108.7 | protein_coding | iPSC_Neurons | 5.94 |
| OSAadjBMI | EUR | rs12227117 | rs941207 | *ATP5B* | ENSG00000110955.4 | protein_coding | hMSC_Adipocyte | 5.89 |
| OSAadjBMI | EUR | rs12227117 | rs941207 | *SNORD59A* | ENSG00000207031.1 | snoRNA | hMSC_Adipocyte | 5.89 |
| OSAadjBMI | EUR | rs12227117 | rs941207 | *U59B* | U59B | snoRNA | hMSC_Adipocyte | 5.89 |
| OSAadjBMI | EUR | rs12227117 | rs941207 | *NACA* | ENSG00000196531.6 | protein_coding | GCB | 5.87 |
| OSAadjBMI | EUR | rs12227117 | rs2958153 | TCONS_00020445 | TCONS_00020445 | lincRNA | iPSC_NPC | 5.8 |
| OSAadjBMI | EUR | rs12227117 | rs2958153 | TCONS_00020807 | TCONS_00020807 | lincRNA | iPSC_NPC | 5.8 |
| OSAadjBMI | EUR | rs12227117 | rs941207 | *BAZ2A* | ENSG00000076108.7 | protein_coding | hESC_HypothalamicNPC | 5.79 |
| OSAadjBMI | EUR | rs12227117 | rs941207 | *BAZ2A* | ENSG00000076108.7 | protein_coding | hMSC_Osteoblasts | 5.77 |
| OSAadjBMI | EUR | rs12227117 | rs2958153 | *BAZ2A* | ENSG00000076108.7 | protein_coding | NaiveB | 5.7 |
| OSAadjBMI | EUR | rs12227117 | rs2958153 | *BAZ2A* | ENSG00000076108.7 | protein_coding | SGBS_Diff | 5.57 |
| OSAadjBMI | EUR | rs12227117 | rs941207 | *NACA* | ENSG00000196531.6 | protein_coding | hESC_HypothalamicNeuron | 5.55 |
| OSAadjBMI | EUR | rs12227117 | rs2958153 | *ESYT1* | ENSG00000139641.8 | protein_coding | iPSC_Neurons | 5.51 |
| OSAadjBMI | EUR | rs12227117 | rs2958153 | *ZC3H10* | ENSG00000135482.2 | protein_coding | iPSC_Neurons | 5.51 |
| OSAadjBMI | EUR | rs12227117 | rs941207 | *BAZ2A* | ENSG00000076108.7 | protein_coding | iPSC_Neurons | 5.39 |
| OSAadjBMI | EUR | rs12227117 | rs2958153 | *ZBTB39* | ENSG00000166860.2 | protein_coding | iPSC_NPC | 5.39 |
| OSAadjBMI | EUR | rs12227117 | rs2958153 | *ATP5B* | ENSG00000110955.4 | protein_coding | TFH | 5.39 |
| OSAadjBMI | EUR | rs12227117 | rs2958153 | *SNORD59A* | ENSG00000207031.1 | snoRNA | TFH | 5.39 |
| OSAadjBMI | EUR | rs12227117 | rs941207 | *RBMS2* | ENSG00000076067.7 | protein_coding | NaiveT | 5.38 |
| OSAadjBMI | EUR | rs12227117 | rs941207 | *U59B* | U59B | snoRNA | NaiveB | 5.28 |
| OSAadjBMI | EUR | rs12227117 | rs2958153 | *ATP5B* | ENSG00000110955.4 | protein_coding | iPSC_Neurons | 5.27 |
| OSAadjBMI | EUR | rs12227117 | rs2958153 | *SNORD59A* | ENSG00000207031.1 | snoRNA | iPSC_Neurons | 5.27 |
| OSAadjBMI | EUR | rs12227117 | rs2958153 | *U59B* | U59B | snoRNA | iPSC_Neurons | 5.27 |
| OSAadjBMI | EUR | rs12227117 | rs941207 | *APOF* | ENSG00000175336.8 | protein_coding | iPSC_NPC | 5.25 |
| OSAadjBMI | EUR | rs12227117 | rs941207 | *NACA* | ENSG00000196531.6 | protein_coding | hMSC_Osteoblasts | 5.18 |
| OSAadjBMI | EUR | rs12227117 | rs941207 | *ATP5B* | ENSG00000110955.4 | protein_coding | iPSC_NPC | 5.12 |
| OSAadjBMI | EUR | rs12227117 | rs941207 | *SNORD59A* | ENSG00000207031.1 | snoRNA | iPSC_NPC | 5.12 |
| OSAadjBMI | EUR | rs12227117 | rs941207 | *U59B* | U59B | snoRNA | iPSC_NPC | 5.12 |
| OSAadjBMI | EUR | rs12227117 | rs941207 | *MYL6* | ENSG00000092841.14 | protein_coding | iPSC_NPC | 5.03 |
| OSAadjBMI | EUR | rs12227117 | rs941207 | *MYL6B* | ENSG00000196465.6 | protein_coding | iPSC_NPC | 5.03 |
| OSAadjBMI | EUR | rs12227117 | rs941207 | RP11-603J24.14 | ENSG00000257809.1 | antisense | iPSC_NPC | 5.03 |
| OSAadjBMI | EUR | rs12227117 | rs2958153 | *ATP5B* | ENSG00000110955.4 | protein_coding | HEPG2 | 5.01 |
| OSAadjBMI | EUR | rs12227117 | rs2958153 | *SNORD59A* | ENSG00000207031.1 | snoRNA | HEPG2 | 5.01 |
| OSAadjBMI | EUR | rs12227117 | rs2958153 | *U59B* | U59B | snoRNA | HEPG2 | 5.01 |
| OSAadjBMI | EUR | rs60791051 | rs73649651 | *FAM189A2* | ENSG00000135063.13 | protein_coding | SGBS_Diff | 9.7 |
| OSAadjBMI | EUR | rs60791051 | rs73649651 | *FAM189A2* | ENSG00000135063.13 | protein_coding | hMSC_Adipocyte | 8.23 |
| OSAadjBMI | EUR | rs60791051 | rs73649651 | *FAM189A2* | ENSG00000135063.13 | protein_coding | hMSC_Osteoblasts | 8.14 |
| OSAadjBMI | EUR | rs60791051 | rs73649651 | *FAM189A2* | ENSG00000135063.13 | protein_coding | hESC_HypothalamicNeuron | 7.98 |
| OSAadjBMI | EUR | rs60791051 | rs77280170 | *FAM189A2* | ENSG00000135063.19 | protein_coding | Chondrocytes | 6.13 |
| OSAadjBMI | EUR | rs60791051 | rs73649651 | *FAM189A2* | ENSG00000135063.13 | protein_coding | hFOB | 5.98 |
| OSAadjBMI | EUR | rs60791051 | rs73649651 | *FAM189A2* | ENSG00000135063.13 | protein_coding | GCB | 5.89 |
| OSAadjBMI | EUR | rs60791051 | rs73649651 | *FAM189A2* | ENSG00000135063.13 | protein_coding | iPSC_Hepatocytes | 5.79 |
| OSAadjBMI | EUR | rs60791051 | rs73649651 | *FAM189A2* | ENSG00000135063.13 | protein_coding | hESC_HypothalamicNPC | 5.6 |
| OSAadjBMI | EUR | rs60791051 | rs73649651 | *TJP2* | ENSG00000119139.12 | protein_coding | hESC_HypothalamicNPC | 5.54 |
| OSAadjBMI | EUR | rs60791051 | rs73649651 | *FAM189A2* | ENSG00000135063.13 | protein_coding | iPSC_NPC | 5.35 |
| OSAadjBMI | EUR | rs60791051 | rs73649651 | TCONS_00016342 | TCONS_00016342 | lincRNA | iPSC_Hepatocytes | 5.3 |
| OSAadjBMI | EUR | rs60791051 | rs73649651 | TCONS_00016343 | TCONS_00016343 | lincRNA | iPSC_Hepatocytes | 5.3 |
| OSAadjBMI | MVP (Sofer et al) | rs2454451 | rs907593 | AC046168.1 | ENSG00000259362.2 | lincRNA | Chondrocytes | 7.98 |
| OSAadjBMI | MVP (Sofer et al) | rs2454451 | rs907594 | AC046168.1 | ENSG00000259362.2 | lincRNA | Chondrocytes | 7.98 |
| OSAadjBMI | MVP (Sofer et al) | rs2841329 | rs12339434 | *NR6A1* | ENSG00000148200.17 | protein_coding | Chondrocytes | 19.78 |
| OSAadjBMI | MVP (Sofer et al) | rs2841329 | rs12339434 | AL354928.1 | ENSG00000236643.1 | antisense | Chondrocytes | 19.78 |
| OSAadjBMI | MVP (Sofer et al) | rs2841329 | rs12339434 | *OLFML2A* | ENSG00000185585.20 | protein_coding | Chondrocytes | 19.71 |
| OSAadjBMI | MVP (Sofer et al) | rs2841329 | rs10760397 | *NR6A1* | ENSG00000148200.17 | protein_coding | Chondrocytes | 15.93 |
| OSAadjBMI | MVP (Sofer et al) | rs2841329 | rs10760397 | AL354928.1 | ENSG00000236643.1 | antisense | Chondrocytes | 15.93 |
| OSAadjBMI | MVP (Sofer et al) | rs2841329 | rs10760397 | *OLFML2A* | ENSG00000185585.20 | protein_coding | Chondrocytes | 12.52 |
| OSAadjBMI | MVP (Sofer et al) | rs2841329 | rs10760397 | *RPL35* | ENSG00000136942.15 | protein_coding | Chondrocytes | 10.99 |
| OSAadjBMI | MVP (Sofer et al) | rs2841329 | rs10760397 | *ARPC5L* | ENSG00000136950.13 | protein_coding | Chondrocytes | 10.99 |
| OSAadjBMI | MVP (Sofer et al) | rs2841329 | rs10760397 | *MVB12B* | ENSG00000196814.15 | protein_coding | Chondrocytes | 10.6 |
| OSAadjBMI | MVP (Sofer et al) | rs2841329 | rs12339434 | *WDR38* | ENSG00000136918.7 | protein_coding | Chondrocytes | 9.64 |
| OSAadjBMI | MVP (Sofer et al) | rs2841329 | rs12339434 | *NRON* | ENSG00000253079.1 | misc_RNA | Chondrocytes | 7.95 |
| OSAadjBMI | MVP (Sofer et al) | rs2841329 | rs556781 | *HSPA5* | ENSG00000044574.8 | protein_coding | Chondrocytes | 7.77 |
| OSAadjBMI | MVP (Sofer et al) | rs2841329 | rs556781 | AL354710.2 | ENSG00000239705.1 | antisense | Chondrocytes | 7.77 |
| OSAadjBMI | MVP (Sofer et al) | rs2841329 | rs10760397 | *WDR38* | ENSG00000136918.7 | protein_coding | Chondrocytes | 7.38 |
| OSAadjBMI | MVP (Sofer et al) | rs2841329 | rs556781 | *ARPC5L* | ENSG00000136950.13 | protein_coding | Chondrocytes | 7 |
| OSAadjBMI | MVP (Sofer et al) | rs2841329 | rs12339434 | *MVB12B* | ENSG00000196814.15 | protein_coding | Chondrocytes | 6.53 |
| OSAadjBMI | MVP (Sofer et al) | rs2841329 | rs10760397 | *GOLGA1* | ENSG00000136935.13 | protein_coding | Chondrocytes | 6.51 |
| OSAadjBMI | MVP (Sofer et al) | rs7120537 | rs2007527 | *BDNF* | ENSG00000176697.19 | protein_coding | Chondrocytes | 11.91 |

**Supplementary Table 16.** Prioritized effector genes from variant to gene (V2G) mapping analysis.

| Trait | Discovery Study | Sentinel rsID | Proxy rsID | Gene Symbol | Gene Id | Gene Biotype | Cell | Contact Score (Chicago Score\|-log10(FitHiC2 FDR) |
|---|---|---|---|---|---|---|---|---|
| OSAadjBMI | MVP (Sofer et al) | rs7120537 | rs11030358 | *BDNF* | ENSG00000176697.19 | protein_coding | Chondrocytes | 11.22 |
| OSAadjBMI | MVP (Sofer et al) | rs7120537 | rs4359193 | *BDNF* | ENSG00000176697.19 | protein_coding | Chondrocytes | 11.2 |
| OSAadjBMI | MVP (Sofer et al) | rs7120537 | rs11030358 | *BDNF-AS* | ENSG00000245573.8 | processed_transcript | Chondrocytes | 8.19 |
| OSAadjBMI | MVP (Sofer et al) | rs7120537 | rs4359193 | *BDNF-AS* | ENSG00000245573.8 | processed_transcript | Chondrocytes | 6.47 |
| OSAadjBMI | MVP (Sofer et al) | rs7120537 | rs11030370 | *BDNF* | ENSG00000176697.14 | protein_coding | hESC_HypothalamicNPC | 5.65 |
| OSAadjBMI | MVP (Sofer et al) | rs79950770 | rs61806235 | *SPAG17* | ENSG00000155761.13 | protein_coding | Chondrocytes | 13.24 |
| OSAadjBMI | MVP (Sofer et al) | rs79950770 | rs61806235 | *RNA5SP56* | ENSG00000222209.1 | rRNA_pseudogene | Chondrocytes | 6.84 |
| OSAadjBMI | Pooled | rs12828563 | rs3214051 | *PTGES3* | ENSG00000110958.11 | protein_coding | TFH | 33.6 |
| OSAadjBMI | Pooled | rs12828563 | rs3214051 | *PTGES3* | ENSG00000110958.11 | protein_coding | hMSC_Adipocyte | 30.51 |
| OSAadjBMI | Pooled | rs12828563 | rs3214051 | *PTGES3* | ENSG00000110958.11 | protein_coding | NaiveT | 28.98 |
| OSAadjBMI | Pooled | rs12828563 | rs3214051 | *PTGES3* | ENSG00000110958.11 | protein_coding | GCB | 28.95 |
| OSAadjBMI | Pooled | rs12828563 | rs3214051 | *PTGES3* | ENSG00000110958.11 | protein_coding | SGBS_Diff | 26.21 |
| OSAadjBMI | Pooled | rs12828563 | rs3214051 | *PTGES3* | ENSG00000110958.11 | protein_coding | hMSC_Osteoblasts | 24.85 |
| OSAadjBMI | Pooled | rs12828563 | rs2255074 | *PTGES3* | ENSG00000110958.11 | protein_coding | iPSC_Hepatocytes | 24.2 |
| OSAadjBMI | Pooled | rs12828563 | rs1107479 | *PTGES3* | ENSG00000110958.11 | protein_coding | iPSC_Hepatocytes | 24.2 |
| OSAadjBMI | Pooled | rs12828563 | rs3214051 | *PTGES3* | ENSG00000110958.11 | protein_coding | NaiveB | 23.5 |
| OSAadjBMI | Pooled | rs12828563 | rs3214051 | *PTGES3* | ENSG00000110958.11 | protein_coding | iPSC_Hepatocytes | 23.2 |
| OSAadjBMI | Pooled | rs12828563 | rs3214051 | *PTGES3* | ENSG00000110958.11 | protein_coding | iPSC_NPC | 22.63 |
| OSAadjBMI | Pooled | rs12828563 | rs3214051 | *PTGES3* | ENSG00000110958.11 | protein_coding | hESC_HypothalamicNeuron | 21.11 |
| OSAadjBMI | Pooled | rs12828563 | rs3214051 | *STAT6* | ENSG00000166888.6 | protein_coding | NaiveT | 21.02 |
| OSAadjBMI | Pooled | rs12828563 | rs3214051 | *PTGES3* | ENSG00000110958.11 | protein_coding | hFOB | 20.67 |
| OSAadjBMI | Pooled | rs12828563 | rs2255074 | *PTGES3* | ENSG00000110958.11 | protein_coding | NaiveT | 19.97 |
| OSAadjBMI | Pooled | rs12828563 | rs1107479 | *PTGES3* | ENSG00000110958.11 | protein_coding | NaiveT | 19.97 |
| OSAadjBMI | Pooled | rs12828563 | rs2255074 | *PTGES3* | ENSG00000110958.11 | protein_coding | hMSC_Osteoblasts | 18.44 |
| OSAadjBMI | Pooled | rs12828563 | rs1107479 | *PTGES3* | ENSG00000110958.11 | protein_coding | hMSC_Osteoblasts | 18.44 |
| OSAadjBMI | Pooled | rs12828563 | rs2958153 | *ATP5B* | ENSG00000110955.4 | protein_coding | hESC_HypothalamicNeuron | 18.15 |
| OSAadjBMI | Pooled | rs12828563 | rs2958153 | *U59B* | U59B | snoRNA | hESC_HypothalamicNeuron | 18.15 |
| OSAadjBMI | Pooled | rs12828563 | rs2958153 | *SNORD59A* | ENSG00000207031.1 | snoRNA | hESC_HypothalamicNeuron | 18.15 |
| OSAadjBMI | Pooled | rs12828563 | rs2255074 | *ANKRD52* | ENSG00000139645.9 | protein_coding | iPSC_NPC | 17.98 |
| OSAadjBMI | Pooled | rs12828563 | rs1107479 | *ANKRD52* | ENSG00000139645.9 | protein_coding | iPSC_NPC | 17.98 |
| OSAadjBMI | Pooled | rs12828563 | rs2255074 | *PTGES3* | ENSG00000110958.11 | protein_coding | TFH | 17.55 |
| OSAadjBMI | Pooled | rs12828563 | rs1107479 | *PTGES3* | ENSG00000110958.11 | protein_coding | TFH | 17.55 |
| OSAadjBMI | Pooled | rs12828563 | rs2255074 | *ATP5B* | ENSG00000110955.4 | protein_coding | hESC_HypothalamicNeuron | 17.42 |
| OSAadjBMI | Pooled | rs12828563 | rs2255074 | *U59B* | U59B | snoRNA | hESC_HypothalamicNeuron | 17.42 |
| OSAadjBMI | Pooled | rs12828563 | rs1107479 | *ATP5B* | ENSG00000110955.4 | protein_coding | hESC_HypothalamicNeuron | 17.42 |
| OSAadjBMI | Pooled | rs12828563 | rs1107479 | *U59B* | U59B | snoRNA | hESC_HypothalamicNeuron | 17.42 |
| OSAadjBMI | Pooled | rs12828563 | rs2255074 | *SNORD59A* | ENSG00000207031.1 | snoRNA | hESC_HypothalamicNeuron | 17.42 |
| OSAadjBMI | Pooled | rs12828563 | rs1107479 | *SNORD59A* | ENSG00000207031.1 | snoRNA | hESC_HypothalamicNeuron | 17.42 |
| OSAadjBMI | Pooled | rs12828563 | rs3214051 | *STAT6* | ENSG00000166888.6 | protein_coding | NaiveB | 17.02 |
| OSAadjBMI | Pooled | rs12828563 | rs3214051 | *ATP5B* | ENSG00000110955.4 | protein_coding | iPSC_Hepatocytes | 16.68 |
| OSAadjBMI | Pooled | rs12828563 | rs3214051 | *SNORD59A* | ENSG00000207031.1 | snoRNA | iPSC_Hepatocytes | 16.68 |
| OSAadjBMI | Pooled | rs12828563 | rs3214051 | *NACA* | ENSG00000196531.6 | protein_coding | iPSC_Hepatocytes | 16.34 |
| OSAadjBMI | Pooled | rs12828563 | rs2255074 | *BAZ2A* | ENSG00000076108.7 | protein_coding | hMSC_Adipocyte | 16.07 |
| OSAadjBMI | Pooled | rs12828563 | rs1107479 | *BAZ2A* | ENSG00000076108.7 | protein_coding | hMSC_Adipocyte | 16.07 |
| OSAadjBMI | Pooled | rs12828563 | rs2255074 | *BAZ2A* | ENSG00000076108.7 | protein_coding | SGBS_Diff | 15.61 |
| OSAadjBMI | Pooled | rs12828563 | rs1107479 | *BAZ2A* | ENSG00000076108.7 | protein_coding | SGBS_Diff | 15.61 |
| OSAadjBMI | Pooled | rs12828563 | rs3214051 | *ATP5B* | ENSG00000110955.4 | protein_coding | hMSC_Osteoblasts | 15.05 |
| OSAadjBMI | Pooled | rs12828563 | rs3214051 | *U59B* | U59B | snoRNA | hMSC_Osteoblasts | 15.05 |
| OSAadjBMI | Pooled | rs12828563 | rs3214051 | *SNORD59A* | ENSG00000207031.1 | snoRNA | hMSC_Osteoblasts | 15.05 |
| OSAadjBMI | Pooled | rs12828563 | rs3214051 | *NACA* | ENSG00000196531.6 | protein_coding | iPSC_Neurons | 14.79 |
| OSAadjBMI | Pooled | rs12828563 | rs3214051 | *NACA* | ENSG00000196531.6 | protein_coding | iPSC_NPC | 14.55 |
| OSAadjBMI | Pooled | rs12828563 | rs2255074 | *BAZ2A* | ENSG00000076108.7 | protein_coding | iPSC_Hepatocytes | 14.54 |
| OSAadjBMI | Pooled | rs12828563 | rs1107479 | *BAZ2A* | ENSG00000076108.7 | protein_coding | iPSC_Hepatocytes | 14.54 |
| OSAadjBMI | Pooled | rs12828563 | rs3214051 | *ATP5B* | ENSG00000110955.4 | protein_coding | hMSC_Adipocyte | 14.42 |
| OSAadjBMI | Pooled | rs12828563 | rs3214051 | *SNORD59A* | ENSG00000207031.1 | snoRNA | hMSC_Adipocyte | 14.42 |
| OSAadjBMI | Pooled | rs12828563 | rs3214051 | *U59B* | U59B | snoRNA | hMSC_Adipocyte | 14.42 |
| OSAadjBMI | Pooled | rs12828563 | rs3214051 | *STAT6* | ENSG00000166888.6 | protein_coding | iPSC_NPC | 14.35 |
| OSAadjBMI | Pooled | rs12828563 | rs3214051 | *HSD17B6* | ENSG00000025423.7 | protein_coding | iPSC_NPC | 14.31 |
| OSAadjBMI | Pooled | rs12828563 | rs3214051 | *PRIM1* | ENSG00000198056.9 | protein_coding | iPSC_NPC | 14.31 |
| OSAadjBMI | Pooled | rs12828563 | rs2255074 | *ATP5B* | ENSG00000110955.4 | protein_coding | TFH | 14.3 |
| OSAadjBMI | Pooled | rs12828563 | rs1107479 | *ATP5B* | ENSG00000110955.4 | protein_coding | TFH | 14.3 |
| OSAadjBMI | Pooled | rs12828563 | rs2255074 | *SNORD59A* | ENSG00000207031.1 | snoRNA | TFH | 14.3 |
| OSAadjBMI | Pooled | rs12828563 | rs1107479 | *SNORD59A* | ENSG00000207031.1 | snoRNA | TFH | 14.3 |
| OSAadjBMI | Pooled | rs12828563 | rs2255074 | *ATP5F1B* | ENSG00000110955.9 | protein_coding | Chondrocytes | 14.19 |
| OSAadjBMI | Pooled | rs12828563 | rs1107479 | *ATP5F1B* | ENSG00000110955.9 | protein_coding | Chondrocytes | 14.19 |
| OSAadjBMI | Pooled | rs12828563 | rs2255074 | *SNORD59A* | ENSG00000207031.1 | snoRNA | Chondrocytes | 14.19 |
| OSAadjBMI | Pooled | rs12828563 | rs1107479 | *SNORD59A* | ENSG00000207031.1 | snoRNA | Chondrocytes | 14.19 |
| OSAadjBMI | Pooled | rs12828563 | rs2255074 | *BAZ2A* | ENSG00000076108.7 | protein_coding | hESC_HypothalamicNeuron | 14.18 |
| OSAadjBMI | Pooled | rs12828563 | rs1107479 | *BAZ2A* | ENSG00000076108.7 | protein_coding | hESC_HypothalamicNeuron | 14.18 |
| OSAadjBMI | Pooled | rs12828563 | rs941207 | *BAZ2A* | ENSG00000076108.7 | protein_coding | hMSC_Adipocyte | 14.07 |
| OSAadjBMI | Pooled | rs12828563 | rs3214051 | *ZBTB39* | ENSG00000166860.2 | protein_coding | hESC_HypothalamicNeuron | 13.76 |
| OSAadjBMI | Pooled | rs12828563 | rs941207 | *BAZ2A* | ENSG00000076108.7 | protein_coding | hESC_HypothalamicNeuron | 13.25 |
| OSAadjBMI | Pooled | rs12828563 | rs2255074 | *ATP5B* | ENSG00000110955.4 | protein_coding | iPSC_Hepatocytes | 13.23 |
| OSAadjBMI | Pooled | rs12828563 | rs2255074 | *SNORD59A* | ENSG00000207031.1 | snoRNA | iPSC_Hepatocytes | 13.23 |
| OSAadjBMI | Pooled | rs12828563 | rs2255074 | *U59B* | U59B | snoRNA | iPSC_Hepatocytes | 13.23 |
| OSAadjBMI | Pooled | rs12828563 | rs1107479 | *ATP5B* | ENSG00000110955.4 | protein_coding | iPSC_Hepatocytes | 13.23 |
| OSAadjBMI | Pooled | rs12828563 | rs1107479 | *SNORD59A* | ENSG00000207031.1 | snoRNA | iPSC_Hepatocytes | 13.23 |
| OSAadjBMI | Pooled | rs12828563 | rs1107479 | *U59B* | U59B | snoRNA | iPSC_Hepatocytes | 13.23 |
| OSAadjBMI | Pooled | rs12828563 | rs2958153 | *BAZ2A* | ENSG00000076108.7 | protein_coding | iPSC_Hepatocytes | 13.23 |
| OSAadjBMI | Pooled | rs12828563 | rs3214051 | *PTGES3* | ENSG00000110958.11 | protein_coding | iPSC_Neurons | 13.17 |
| OSAadjBMI | Pooled | rs12828563 | rs2255074 | *U59B* | U59B | snoRNA | TFH | 13.03 |
| OSAadjBMI | Pooled | rs12828563 | rs1107479 | *U59B* | U59B | snoRNA | TFH | 13.03 |
| OSAadjBMI | Pooled | rs12828563 | rs3214051 | *BAZ2A* | ENSG00000076108.7 | protein_coding | TFH | 12.94 |
| OSAadjBMI | Pooled | rs12828563 | rs3214051 | *GPR182* | ENSG00000166856.1 | protein_coding | hFOB | 12.92 |
| OSAadjBMI | Pooled | rs12828563 | rs3214051 | RP11-474N8.5 | ENSG00000258442.1 | protein_coding | hFOB | 12.92 |

**Supplementary Table 16.** Prioritized effector genes from variant to gene (V2G) mapping analysis.

| Trait | Discovery Study | Sentinel rsID | Proxy rsID | Gene Symbol | Gene Id | Gene Biotype | Cell | Contact Score (Chicago Score\|-log10(FitHiC2 FDR) |
|---|---|---|---|---|---|---|---|---|
| OSAadjBMI | Pooled | rs12828563 | rs3214051 | *BAZ2A* | ENSG00000076108.7 | protein_coding | iPSC_Hepatocytes | 12.79 |
| OSAadjBMI | Pooled | rs12828563 | rs2255074 | *BAZ2A* | ENSG00000076108.7 | protein_coding | NaiveB | 12.45 |
| OSAadjBMI | Pooled | rs12828563 | rs1107479 | *BAZ2A* | ENSG00000076108.7 | protein_coding | NaiveB | 12.45 |
| OSAadjBMI | Pooled | rs12828563 | rs2255074 | *ATP5B* | ENSG00000110955.4 | protein_coding | GCB | 12.44 |
| OSAadjBMI | Pooled | rs12828563 | rs2255074 | *SNORD59A* | ENSG00000207031.1 | snoRNA | GCB | 12.44 |
| OSAadjBMI | Pooled | rs12828563 | rs2255074 | *U59B* | U59B | snoRNA | GCB | 12.44 |
| OSAadjBMI | Pooled | rs12828563 | rs1107479 | *ATP5B* | ENSG00000110955.4 | protein_coding | GCB | 12.44 |
| OSAadjBMI | Pooled | rs12828563 | rs1107479 | *SNORD59A* | ENSG00000207031.1 | snoRNA | GCB | 12.44 |
| OSAadjBMI | Pooled | rs12828563 | rs1107479 | *U59B* | U59B | snoRNA | GCB | 12.44 |
| OSAadjBMI | Pooled | rs12828563 | rs3214051 | *ATP5B* | ENSG00000110955.4 | protein_coding | hFOB | 12.43 |
| OSAadjBMI | Pooled | rs12828563 | rs3214051 | *SNORD59A* | ENSG00000207031.1 | snoRNA | hFOB | 12.43 |
| OSAadjBMI | Pooled | rs12828563 | rs3214051 | *U59B* | U59B | snoRNA | hFOB | 12.43 |
| OSAadjBMI | Pooled | rs12828563 | rs2255074 | *PTGES3* | ENSG00000110958.11 | protein_coding | NaiveB | 12.41 |
| OSAadjBMI | Pooled | rs12828563 | rs1107479 | *PTGES3* | ENSG00000110958.11 | protein_coding | NaiveB | 12.41 |
| OSAadjBMI | Pooled | rs12828563 | rs3214051 | *BAZ2A* | ENSG00000076108.7 | protein_coding | NaiveT | 12.34 |
| OSAadjBMI | Pooled | rs12828563 | rs3214051 | *LRP1* | ENSG00000123384.9 | protein_coding | iPSC_NPC | 12.18 |
| OSAadjBMI | Pooled | rs12828563 | rs3214051 | *GPR182* | ENSG00000166856.1 | protein_coding | hMSC_Adipocyte | 12.17 |
| OSAadjBMI | Pooled | rs12828563 | rs3214051 | RP11-474N8.5 | ENSG00000258442.1 | protein_coding | hMSC_Adipocyte | 12.17 |
| OSAadjBMI | Pooled | rs12828563 | rs3214051 | *U59B* | U59B | snoRNA | iPSC_Hepatocytes | 12.14 |
| OSAadjBMI | Pooled | rs12828563 | rs941207 | *RBMS2* | ENSG00000076067.7 | protein_coding | SGBS_Diff | 12.12 |
| OSAadjBMI | Pooled | rs12828563 | rs3214051 | *BAZ2A* | ENSG00000076108.7 | protein_coding | hMSC_Adipocyte | 12.06 |
| OSAadjBMI | Pooled | rs12828563 | rs3214051 | *ATP5B* | ENSG00000110955.4 | protein_coding | hESC_HypothalamicNeuron | 12.05 |
| OSAadjBMI | Pooled | rs12828563 | rs3214051 | *SNORD59A* | ENSG00000207031.1 | snoRNA | hESC_HypothalamicNeuron | 12.05 |
| OSAadjBMI | Pooled | rs12828563 | rs3214051 | *U59B* | U59B | snoRNA | hESC_HypothalamicNeuron | 12.05 |
| OSAadjBMI | Pooled | rs12828563 | rs2255074 | *ATP5B* | ENSG00000110955.4 | protein_coding | SGBS_Diff | 12.05 |
| OSAadjBMI | Pooled | rs12828563 | rs2255074 | *U59B* | U59B | snoRNA | SGBS_Diff | 12.05 |
| OSAadjBMI | Pooled | rs12828563 | rs2255074 | *SNORD59A* | ENSG00000207031.1 | snoRNA | SGBS_Diff | 12.05 |
| OSAadjBMI | Pooled | rs12828563 | rs1107479 | *ATP5B* | ENSG00000110955.4 | protein_coding | SGBS_Diff | 12.05 |
| OSAadjBMI | Pooled | rs12828563 | rs1107479 | *U59B* | U59B | snoRNA | SGBS_Diff | 12.05 |
| OSAadjBMI | Pooled | rs12828563 | rs1107479 | *SNORD59A* | ENSG00000207031.1 | snoRNA | SGBS_Diff | 12.05 |
| OSAadjBMI | Pooled | rs12828563 | rs941207 | *ATP5B* | ENSG00000110955.4 | protein_coding | GCB | 11.88 |
| OSAadjBMI | Pooled | rs12828563 | rs941207 | *SNORD59A* | ENSG00000207031.1 | snoRNA | GCB | 11.88 |
| OSAadjBMI | Pooled | rs12828563 | rs941207 | *U59B* | U59B | snoRNA | GCB | 11.88 |
| OSAadjBMI | Pooled | rs12828563 | rs2255074 | *BAZ2A* | ENSG00000076108.7 | protein_coding | TFH | 11.81 |
| OSAadjBMI | Pooled | rs12828563 | rs1107479 | *BAZ2A* | ENSG00000076108.7 | protein_coding | TFH | 11.81 |
| OSAadjBMI | Pooled | rs12828563 | rs3214051 | *STAT6* | ENSG00000166888.6 | protein_coding | TFH | 11.8 |
| OSAadjBMI | Pooled | rs12828563 | rs941207 | *ATP5B* | ENSG00000110955.4 | protein_coding | hESC_HypothalamicNeuron | 11.73 |
| OSAadjBMI | Pooled | rs12828563 | rs941207 | *U59B* | U59B | snoRNA | hESC_HypothalamicNeuron | 11.73 |
| OSAadjBMI | Pooled | rs12828563 | rs941207 | *SNORD59A* | ENSG00000207031.1 | snoRNA | hESC_HypothalamicNeuron | 11.73 |
| OSAadjBMI | Pooled | rs12828563 | rs2255074 | *NACA* | ENSG00000196531.6 | protein_coding | iPSC_Hepatocytes | 11.53 |
| OSAadjBMI | Pooled | rs12828563 | rs1107479 | *NACA* | ENSG00000196531.6 | protein_coding | iPSC_Hepatocytes | 11.53 |
| OSAadjBMI | Pooled | rs12828563 | rs2958153 | *BAZ2A* | ENSG00000076108.7 | protein_coding | NaiveT | 11.48 |
| OSAadjBMI | Pooled | rs12828563 | rs2958153 | *ATP5B* | ENSG00000110955.4 | protein_coding | iPSC_NPC | 11.45 |
| OSAadjBMI | Pooled | rs12828563 | rs2958153 | *U59B* | U59B | snoRNA | iPSC_NPC | 11.45 |
| OSAadjBMI | Pooled | rs12828563 | rs2958153 | *SNORD59A* | ENSG00000207031.1 | snoRNA | iPSC_NPC | 11.45 |
| OSAadjBMI | Pooled | rs12828563 | rs3214051 | *ZBTB39* | ENSG00000166860.2 | protein_coding | hMSC_Adipocyte | 11.44 |
| OSAadjBMI | Pooled | rs12828563 | rs3214051 | *ZBTB39* | ENSG00000166860.2 | protein_coding | SGBS_Diff | 11.41 |
| OSAadjBMI | Pooled | rs12828563 | rs3214051 | *ATP5B* | ENSG00000110955.4 | protein_coding | SGBS_Diff | 11.12 |
| OSAadjBMI | Pooled | rs12828563 | rs3214051 | *SNORD59A* | ENSG00000207031.1 | snoRNA | SGBS_Diff | 11.12 |
| OSAadjBMI | Pooled | rs12828563 | rs3214051 | *U59B* | U59B | snoRNA | SGBS_Diff | 11.12 |
| OSAadjBMI | Pooled | rs12828563 | rs3214051 | *ATP5B* | ENSG00000110955.4 | protein_coding | TFH | 11.08 |
| OSAadjBMI | Pooled | rs12828563 | rs3214051 | *SNORD59A* | ENSG00000207031.1 | snoRNA | TFH | 11.08 |
| OSAadjBMI | Pooled | rs12828563 | rs2255074 | *ATP5B* | ENSG00000110955.4 | protein_coding | hMSC_Adipocyte | 10.94 |
| OSAadjBMI | Pooled | rs12828563 | rs2255074 | *U59B* | U59B | snoRNA | hMSC_Adipocyte | 10.94 |
| OSAadjBMI | Pooled | rs12828563 | rs2255074 | *SNORD59A* | ENSG00000207031.1 | snoRNA | hMSC_Adipocyte | 10.94 |
| OSAadjBMI | Pooled | rs12828563 | rs1107479 | *ATP5B* | ENSG00000110955.4 | protein_coding | hMSC_Adipocyte | 10.94 |
| OSAadjBMI | Pooled | rs12828563 | rs1107479 | *U59B* | U59B | snoRNA | hMSC_Adipocyte | 10.94 |
| OSAadjBMI | Pooled | rs12828563 | rs1107479 | *SNORD59A* | ENSG00000207031.1 | snoRNA | hMSC_Adipocyte | 10.94 |
| OSAadjBMI | Pooled | rs12828563 | rs2255074 | *RBMS2* | ENSG00000076067.7 | protein_coding | hMSC_Adipocyte | 10.76 |
| OSAadjBMI | Pooled | rs12828563 | rs1107479 | *RBMS2* | ENSG00000076067.7 | protein_coding | hMSC_Adipocyte | 10.76 |
| OSAadjBMI | Pooled | rs12828563 | rs2255074 | *BAZ2A* | ENSG00000076108.7 | protein_coding | hFOB | 10.65 |
| OSAadjBMI | Pooled | rs12828563 | rs1107479 | *BAZ2A* | ENSG00000076108.7 | protein_coding | hFOB | 10.65 |
| OSAadjBMI | Pooled | rs12828563 | rs2958153 | *NACA* | ENSG00000196531.6 | protein_coding | iPSC_Hepatocytes | 10.56 |
| OSAadjBMI | Pooled | rs12828563 | rs3214051 | *ZBTB39* | ENSG00000166860.2 | protein_coding | NaiveB | 10.5 |
| OSAadjBMI | Pooled | rs12828563 | rs941207 | *RBMS2* | ENSG00000076067.7 | protein_coding | iPSC_NPC | 10.48 |
| OSAadjBMI | Pooled | rs12828563 | rs3214051 | *BAZ2A* | ENSG00000076108.7 | protein_coding | hMSC_Osteoblasts | 10.43 |
| OSAadjBMI | Pooled | rs12828563 | rs2255074 | *ATP5B* | ENSG00000110955.4 | protein_coding | NaiveB | 10.39 |
| OSAadjBMI | Pooled | rs12828563 | rs2255074 | *U59B* | U59B | snoRNA | NaiveB | 10.39 |
| OSAadjBMI | Pooled | rs12828563 | rs1107479 | *ATP5B* | ENSG00000110955.4 | protein_coding | NaiveB | 10.39 |
| OSAadjBMI | Pooled | rs12828563 | rs1107479 | *U59B* | U59B | snoRNA | NaiveB | 10.39 |
| OSAadjBMI | Pooled | rs12828563 | rs941207 | *BAZ2A* | ENSG00000076108.7 | protein_coding | NaiveB | 10.34 |
| OSAadjBMI | Pooled | rs12828563 | rs3214051 | *NACA* | ENSG00000196531.6 | protein_coding | SGBS_Diff | 10.33 |
| OSAadjBMI | Pooled | rs12828563 | rs2958153 | *NACA* | ENSG00000196531.6 | protein_coding | GCB | 10.29 |
| OSAadjBMI | Pooled | rs12828563 | rs2255074 | *ATP5B* | ENSG00000110955.4 | protein_coding | hMSC_Osteoblasts | 10.22 |
| OSAadjBMI | Pooled | rs12828563 | rs2255074 | *U59B* | U59B | snoRNA | hMSC_Osteoblasts | 10.22 |
| OSAadjBMI | Pooled | rs12828563 | rs2255074 | *SNORD59A* | ENSG00000207031.1 | snoRNA | hMSC_Osteoblasts | 10.22 |
| OSAadjBMI | Pooled | rs12828563 | rs1107479 | *ATP5B* | ENSG00000110955.4 | protein_coding | hMSC_Osteoblasts | 10.22 |
| OSAadjBMI | Pooled | rs12828563 | rs1107479 | *U59B* | U59B | snoRNA | hMSC_Osteoblasts | 10.22 |
| OSAadjBMI | Pooled | rs12828563 | rs1107479 | *SNORD59A* | ENSG00000207031.1 | snoRNA | hMSC_Osteoblasts | 10.22 |
| OSAadjBMI | Pooled | rs12828563 | rs3214051 | *GPR182* | ENSG00000166856.1 | protein_coding | iPSC_Hepatocytes | 10.22 |
| OSAadjBMI | Pooled | rs12828563 | rs3214051 | RP11-474N8.5 | ENSG00000258442.1 | protein_coding | iPSC_Hepatocytes | 10.22 |
| OSAadjBMI | Pooled | rs12828563 | rs2255074 | *NACA* | ENSG00000196531.6 | protein_coding | TFH | 10.13 |
| OSAadjBMI | Pooled | rs12828563 | rs1107479 | *NACA* | ENSG00000196531.6 | protein_coding | TFH | 10.13 |
| OSAadjBMI | Pooled | rs12828563 | rs3214051 | *ZBTB39* | ENSG00000166860.2 | protein_coding | iPSC_NPC | 10.09 |
| OSAadjBMI | Pooled | rs12828563 | rs3214051 | *GPR182* | ENSG00000166856.1 | protein_coding | SGBS_Diff | 10.09 |

| Supplementary Table 16. Prioritized effector genes from variant to gene (V2G) mapping analysis. | | | | | | | | |
|---|---|---|---|---|---|---|---|---|
| Trait | Discovery Study | Sentinel rsID | Proxy rsID | Gene Symbol | Gene Id | Gene Biotype | Cell | Contact Score (Chicago Score\|-log10(FitHiC2 FDR) |
| OSAadjBMI | Pooled | rs12828563 | rs3214051 | RP11-474N8.5 | ENSG00000258442.1 | protein_coding | SGBS_Diff | 10.09 |
| OSAadjBMI | Pooled | rs12828563 | rs3214051 | *ZBTB39* | ENSG00000166860.2 | protein_coding | hMSC_Osteoblasts | 10 |
| OSAadjBMI | Pooled | rs12828563 | rs3214051 | TCONS_00020445 | TCONS_00020445 | lincRNA | iPSC_NPC | 9.98 |
| OSAadjBMI | Pooled | rs12828563 | rs3214051 | TCONS_00020807 | TCONS_00020807 | lincRNA | iPSC_NPC | 9.98 |
| OSAadjBMI | Pooled | rs12828563 | rs2255074 | *BAZ2A* | ENSG00000076108.7 | protein_coding | iPSC_NPC | 9.94 |
| OSAadjBMI | Pooled | rs12828563 | rs1107479 | *BAZ2A* | ENSG00000076108.7 | protein_coding | iPSC_NPC | 9.94 |
| OSAadjBMI | Pooled | rs12828563 | rs941207 | *BAZ2A* | ENSG00000076108.7 | protein_coding | GCB | 9.91 |
| OSAadjBMI | Pooled | rs12828563 | rs2255074 | *PTGES3* | ENSG00000110958.11 | protein_coding | iPSC_Neurons | 9.72 |
| OSAadjBMI | Pooled | rs12828563 | rs1107479 | *PTGES3* | ENSG00000110958.11 | protein_coding | iPSC_Neurons | 9.72 |
| OSAadjBMI | Pooled | rs12828563 | rs2255074 | *BAZ2A* | ENSG00000076108.7 | protein_coding | GCB | 9.69 |
| OSAadjBMI | Pooled | rs12828563 | rs1107479 | *BAZ2A* | ENSG00000076108.7 | protein_coding | GCB | 9.69 |
| OSAadjBMI | Pooled | rs12828563 | rs2255074 | *ATP5B* | ENSG00000110955.4 | protein_coding | NaiveT | 9.69 |
| OSAadjBMI | Pooled | rs12828563 | rs2255074 | *SNORD59A* | ENSG00000207031.1 | snoRNA | NaiveT | 9.69 |
| OSAadjBMI | Pooled | rs12828563 | rs2255074 | *U59B* | U59B | snoRNA | NaiveT | 9.69 |
| OSAadjBMI | Pooled | rs12828563 | rs1107479 | *ATP5B* | ENSG00000110955.4 | protein_coding | NaiveT | 9.69 |
| OSAadjBMI | Pooled | rs12828563 | rs1107479 | *SNORD59A* | ENSG00000207031.1 | snoRNA | NaiveT | 9.69 |
| OSAadjBMI | Pooled | rs12828563 | rs1107479 | *U59B* | U59B | snoRNA | NaiveT | 9.69 |
| OSAadjBMI | Pooled | rs12828563 | rs3214051 | TCONS_00020445 | TCONS_00020445 | lincRNA | TFH | 9.62 |
| OSAadjBMI | Pooled | rs12828563 | rs3214051 | TCONS_00020807 | TCONS_00020807 | lincRNA | TFH | 9.62 |
| OSAadjBMI | Pooled | rs12828563 | rs2958153 | *NACA* | ENSG00000196531.6 | protein_coding | TFH | 9.58 |
| OSAadjBMI | Pooled | rs12828563 | rs2958153 | *NACA* | ENSG00000196531.6 | protein_coding | hMSC_Adipocyte | 9.57 |
| OSAadjBMI | Pooled | rs12828563 | rs941207 | *ATP5B* | ENSG00000110955.4 | protein_coding | TFH | 9.57 |
| OSAadjBMI | Pooled | rs12828563 | rs941207 | *SNORD59A* | ENSG00000207031.1 | snoRNA | TFH | 9.57 |
| OSAadjBMI | Pooled | rs12828563 | rs941207 | *U59B* | U59B | snoRNA | TFH | 9.57 |
| OSAadjBMI | Pooled | rs12828563 | rs3214051 | TCONS_00020445 | TCONS_00020445 | lincRNA | GCB | 9.56 |
| OSAadjBMI | Pooled | rs12828563 | rs3214051 | TCONS_00020807 | TCONS_00020807 | lincRNA | GCB | 9.56 |
| OSAadjBMI | Pooled | rs12828563 | rs3214051 | *NXPH4* | ENSG00000182379.9 | protein_coding | hFOB | 9.49 |
| OSAadjBMI | Pooled | rs12828563 | rs2958153 | *ATP5B* | ENSG00000110955.4 | protein_coding | iPSC_Hepatocytes | 9.48 |
| OSAadjBMI | Pooled | rs12828563 | rs2958153 | *SNORD59A* | ENSG00000207031.1 | snoRNA | iPSC_Hepatocytes | 9.48 |
| OSAadjBMI | Pooled | rs12828563 | rs2958153 | *U59B* | U59B | snoRNA | iPSC_Hepatocytes | 9.48 |
| OSAadjBMI | Pooled | rs12828563 | rs3214051 | *GPR182* | ENSG00000166856.1 | protein_coding | hESC_HypothalamicNPC | 9.47 |
| OSAadjBMI | Pooled | rs12828563 | rs3214051 | RP11-474N8.5 | ENSG00000258442.1 | protein_coding | hESC_HypothalamicNPC | 9.47 |
| OSAadjBMI | Pooled | rs12828563 | rs941207 | *RBMS2* | ENSG00000076067.7 | protein_coding | hMSC_Adipocyte | 9.43 |
| OSAadjBMI | Pooled | rs12828563 | rs941207 | *RBMS2* | ENSG00000076067.7 | protein_coding | hESC_HypothalamicNeuron | 9.42 |
| OSAadjBMI | Pooled | rs12828563 | rs3214051 | *STAT6* | ENSG00000166888.6 | protein_coding | SGBS_Diff | 9.42 |
| OSAadjBMI | Pooled | rs12828563 | rs3214051 | TCONS_00020445 | TCONS_00020445 | lincRNA | NaiveB | 9.39 |
| OSAadjBMI | Pooled | rs12828563 | rs3214051 | TCONS_00020807 | TCONS_00020807 | lincRNA | NaiveB | 9.39 |
| OSAadjBMI | Pooled | rs12828563 | rs3214051 | *NACA* | ENSG00000196531.6 | protein_coding | hMSC_Osteoblasts | 9.38 |
| OSAadjBMI | Pooled | rs12828563 | rs2255074 | *BAZ2A* | ENSG00000076108.7 | protein_coding | hMSC_Osteoblasts | 9.36 |
| OSAadjBMI | Pooled | rs12828563 | rs1107479 | *BAZ2A* | ENSG00000076108.7 | protein_coding | hMSC_Osteoblasts | 9.36 |
| OSAadjBMI | Pooled | rs12828563 | rs3214051 | *ZBTB39* | ENSG00000166860.2 | protein_coding | hFOB | 9.3 |
| OSAadjBMI | Pooled | rs12828563 | rs941207 | *NACA* | ENSG00000196531.6 | protein_coding | NaiveT | 9.25 |
| OSAadjBMI | Pooled | rs12828563 | rs2958153 | *ATP5B* | ENSG00000110955.4 | protein_coding | hMSC_Osteoblasts | 9.21 |
| OSAadjBMI | Pooled | rs12828563 | rs2958153 | *SNORD59A* | ENSG00000207031.1 | snoRNA | hMSC_Osteoblasts | 9.21 |
| OSAadjBMI | Pooled | rs12828563 | rs2958153 | *U59B* | U59B | snoRNA | hMSC_Osteoblasts | 9.21 |
| OSAadjBMI | Pooled | rs12828563 | rs2958153 | *ATP5B* | ENSG00000110955.4 | protein_coding | SGBS_Diff | 9.12 |
| OSAadjBMI | Pooled | rs12828563 | rs2958153 | *U59B* | U59B | snoRNA | SGBS_Diff | 9.12 |
| OSAadjBMI | Pooled | rs12828563 | rs2958153 | *SNORD59A* | ENSG00000207031.1 | snoRNA | SGBS_Diff | 9.12 |
| OSAadjBMI | Pooled | rs12828563 | rs2255074 | *SNORD59A* | ENSG00000207031.1 | snoRNA | NaiveB | 9.04 |
| OSAadjBMI | Pooled | rs12828563 | rs1107479 | *SNORD59A* | ENSG00000207031.1 | snoRNA | NaiveB | 9.04 |
| OSAadjBMI | Pooled | rs12828563 | rs2958153 | *NACA* | ENSG00000196531.6 | protein_coding | NaiveT | 9.01 |
| OSAadjBMI | Pooled | rs12828563 | rs2255074 | *NACA* | ENSG00000196531.6 | protein_coding | iPSC_Neurons | 8.96 |
| OSAadjBMI | Pooled | rs12828563 | rs1107479 | *NACA* | ENSG00000196531.6 | protein_coding | iPSC_Neurons | 8.96 |
| OSAadjBMI | Pooled | rs12828563 | rs3214051 | *GPR182* | ENSG00000166856.1 | protein_coding | NaiveB | 8.91 |
| OSAadjBMI | Pooled | rs12828563 | rs3214051 | RP11-474N8.5 | ENSG00000258442.1 | protein_coding | NaiveB | 8.91 |
| OSAadjBMI | Pooled | rs12828563 | rs2958153 | *NACA* | ENSG00000196531.6 | protein_coding | SGBS_Diff | 8.85 |
| OSAadjBMI | Pooled | rs12828563 | rs3214051 | *ATP5B* | ENSG00000110955.4 | protein_coding | GCB | 8.83 |
| OSAadjBMI | Pooled | rs12828563 | rs3214051 | *SNORD59A* | ENSG00000207031.1 | snoRNA | GCB | 8.83 |
| OSAadjBMI | Pooled | rs12828563 | rs3214051 | *U59B* | U59B | snoRNA | GCB | 8.83 |
| OSAadjBMI | Pooled | rs12828563 | rs941207 | *ATP5B* | ENSG00000110955.4 | protein_coding | NaiveB | 8.83 |
| OSAadjBMI | Pooled | rs12828563 | rs941207 | *SNORD59A* | ENSG00000207031.1 | snoRNA | NaiveB | 8.83 |
| OSAadjBMI | Pooled | rs12828563 | rs3214051 | *ZBTB39* | ENSG00000166860.2 | protein_coding | NaiveT | 8.77 |
| OSAadjBMI | Pooled | rs12828563 | rs941207 | *BAZ2A* | ENSG00000076108.7 | protein_coding | TFH | 8.75 |
| OSAadjBMI | Pooled | rs12828563 | rs2255074 | *NACA* | ENSG00000196531.6 | protein_coding | hMSC_Adipocyte | 8.74 |
| OSAadjBMI | Pooled | rs12828563 | rs1107479 | *NACA* | ENSG00000196531.6 | protein_coding | hMSC_Adipocyte | 8.74 |
| OSAadjBMI | Pooled | rs12828563 | rs941207 | *RBMS2* | ENSG00000076067.7 | protein_coding | iPSC_Neurons | 8.74 |
| OSAadjBMI | Pooled | rs12828563 | rs2958153 | *NACA* | ENSG00000196531.6 | protein_coding | iPSC_NPC | 8.68 |
| OSAadjBMI | Pooled | rs12828563 | rs3214051 | *BAZ2A* | ENSG00000076108.7 | protein_coding | SGBS_Diff | 8.66 |
| OSAadjBMI | Pooled | rs12828563 | rs941207 | *BAZ2A* | ENSG00000076108.7 | protein_coding | iPSC_NPC | 8.63 |
| OSAadjBMI | Pooled | rs12828563 | rs2255074 | *ATP5B* | ENSG00000110955.4 | protein_coding | iPSC_NPC | 8.62 |
| OSAadjBMI | Pooled | rs12828563 | rs2255074 | *SNORD59A* | ENSG00000207031.1 | snoRNA | iPSC_NPC | 8.62 |
| OSAadjBMI | Pooled | rs12828563 | rs1107479 | *ATP5B* | ENSG00000110955.4 | protein_coding | iPSC_NPC | 8.62 |
| OSAadjBMI | Pooled | rs12828563 | rs1107479 | *SNORD59A* | ENSG00000207031.1 | snoRNA | iPSC_NPC | 8.62 |
| OSAadjBMI | Pooled | rs12828563 | rs7980317 | *ZBTB39* | ENSG00000166860.2 | protein_coding | hMSC_Adipocyte | 8.6 |
| OSAadjBMI | Pooled | rs12828563 | rs3214051 | *NAB2* | ENSG00000166886.8 | protein_coding | hFOB | 8.54 |
| OSAadjBMI | Pooled | rs12828563 | rs3214051 | TCONS_00020445 | TCONS_00020445 | lincRNA | hFOB | 8.54 |
| OSAadjBMI | Pooled | rs12828563 | rs3214051 | TCONS_00020805 | TCONS_00020805 | lincRNA | hFOB | 8.54 |
| OSAadjBMI | Pooled | rs12828563 | rs3214051 | TCONS_00020806 | TCONS_00020806 | lincRNA | hFOB | 8.54 |
| OSAadjBMI | Pooled | rs12828563 | rs3214051 | TCONS_00020807 | TCONS_00020807 | lincRNA | hFOB | 8.54 |
| OSAadjBMI | Pooled | rs12828563 | rs3214051 | TCONS_00021326 | TCONS_00021326 | lincRNA | hFOB | 8.54 |
| OSAadjBMI | Pooled | rs12828563 | rs3214051 | TCONS_00021327 | TCONS_00021327 | lincRNA | hFOB | 8.54 |
| OSAadjBMI | Pooled | rs12828563 | rs3214051 | *TMEM194A* | ENSG00000166881.5 | protein_coding | hFOB | 8.54 |
| OSAadjBMI | Pooled | rs12828563 | rs3214051 | *BAZ2A* | ENSG00000076108.7 | protein_coding | iPSC_Neurons | 8.46 |
| OSAadjBMI | Pooled | rs12828563 | rs2255074 | *ATP5B* | ENSG00000110955.4 | protein_coding | hFOB | 8.31 |
| OSAadjBMI | Pooled | rs12828563 | rs2255074 | *U59B* | U59B | snoRNA | hFOB | 8.31 |

**Supplementary Table 16.** Prioritized effector genes from variant to gene (V2G) mapping analysis.

| Trait | Discovery Study | Sentinel rsID | Proxy rsID | Gene Symbol | Gene Id | Gene Biotype | Cell | Contact Score (Chicago Score\|-log10(FitHiC2 FDR) |
|---|---|---|---|---|---|---|---|---|
| OSAadjBMI | Pooled | rs12828563 | rs2255074 | *SNORD59A* | ENSG00000207031.1 | snoRNA | hFOB | 8.31 |
| OSAadjBMI | Pooled | rs12828563 | rs1107479 | *ATP5B* | ENSG00000110955.4 | protein_coding | hFOB | 8.31 |
| OSAadjBMI | Pooled | rs12828563 | rs1107479 | *U59B* | U59B | snoRNA | hFOB | 8.31 |
| OSAadjBMI | Pooled | rs12828563 | rs1107479 | *SNORD59A* | ENSG00000207031.1 | snoRNA | hFOB | 8.31 |
| OSAadjBMI | Pooled | rs12828563 | rs2255074 | *NACA* | ENSG00000196531.6 | protein_coding | hMSC_Osteoblasts | 8.3 |
| OSAadjBMI | Pooled | rs12828563 | rs1107479 | *NACA* | ENSG00000196531.6 | protein_coding | hMSC_Osteoblasts | 8.3 |
| OSAadjBMI | Pooled | rs12828563 | rs3214051 | *NACA* | ENSG00000196531.6 | protein_coding | hESC_HypothalamicNeuron | 8.26 |
| OSAadjBMI | Pooled | rs12828563 | rs941207 | *ZBTB39* | ENSG00000166860.2 | protein_coding | hESC_HypothalamicNeuron | 8.22 |
| OSAadjBMI | Pooled | rs12828563 | rs2958153 | *ZBTB39* | ENSG00000166860.2 | protein_coding | iPSC_Hepatocytes | 8.21 |
| OSAadjBMI | Pooled | rs12828563 | rs2255074 | *STAT6* | ENSG00000166888.6 | protein_coding | iPSC_NPC | 8.15 |
| OSAadjBMI | Pooled | rs12828563 | rs1107479 | *STAT6* | ENSG00000166888.6 | protein_coding | iPSC_NPC | 8.15 |
| OSAadjBMI | Pooled | rs12828563 | rs3214051 | *ATP5B* | ENSG00000110955.4 | protein_coding | iPSC_NPC | 8.09 |
| OSAadjBMI | Pooled | rs12828563 | rs3214051 | *SNORD59A* | ENSG00000207031.1 | snoRNA | iPSC_NPC | 8.09 |
| OSAadjBMI | Pooled | rs12828563 | rs3214051 | *U59B* | U59B | snoRNA | iPSC_NPC | 8.09 |
| OSAadjBMI | Pooled | rs12828563 | rs2958153 | *NACA* | ENSG00000196531.6 | protein_coding | hMSC_Osteoblasts | 8.08 |
| OSAadjBMI | Pooled | rs12828563 | rs3214051 | *BAZ2A* | ENSG00000076108.7 | protein_coding | GCB | 8.05 |
| OSAadjBMI | Pooled | rs12828563 | rs3214051 | TCONS_00020445 | TCONS_00020445 | lincRNA | NaiveT | 8.02 |
| OSAadjBMI | Pooled | rs12828563 | rs3214051 | TCONS_00020807 | TCONS_00020807 | lincRNA | NaiveT | 8.02 |
| OSAadjBMI | Pooled | rs12828563 | rs2958153 | *BAZ2A* | ENSG00000076108.7 | protein_coding | hMSC_Osteoblasts | 8 |
| OSAadjBMI | Pooled | rs12828563 | rs2255074 | *NACA* | ENSG00000196531.6 | protein_coding | NaiveT | 7.94 |
| OSAadjBMI | Pooled | rs12828563 | rs1107479 | *NACA* | ENSG00000196531.6 | protein_coding | NaiveT | 7.94 |
| OSAadjBMI | Pooled | rs12828563 | rs3214051 | *NACA* | ENSG00000196531.6 | protein_coding | TFH | 7.9 |
| OSAadjBMI | Pooled | rs12828563 | rs3214051 | *BAZ2A* | ENSG00000076108.7 | protein_coding | NaiveB | 7.76 |
| OSAadjBMI | Pooled | rs12828563 | rs3214051 | *GPR182* | ENSG00000166856.1 | protein_coding | hMSC_Osteoblasts | 7.65 |
| OSAadjBMI | Pooled | rs12828563 | rs3214051 | RP11-474N8.5 | ENSG00000258442.1 | protein_coding | hMSC_Osteoblasts | 7.65 |
| OSAadjBMI | Pooled | rs12828563 | rs941207 | *NACA* | ENSG00000196531.6 | protein_coding | TFH | 7.64 |
| OSAadjBMI | Pooled | rs12828563 | rs2255074 | *COQ10A* | ENSG00000135469.9 | protein_coding | hFOB | 7.57 |
| OSAadjBMI | Pooled | rs12828563 | rs1107479 | *COQ10A* | ENSG00000135469.9 | protein_coding | hFOB | 7.57 |
| OSAadjBMI | Pooled | rs12828563 | rs2958153 | *ZBTB39* | ENSG00000166860.2 | protein_coding | iPSC_Neurons | 7.57 |
| OSAadjBMI | Pooled | rs12828563 | rs3214051 | *ATP5B* | ENSG00000110955.4 | protein_coding | iPSC_Neurons | 7.54 |
| OSAadjBMI | Pooled | rs12828563 | rs3214051 | *SNORD59A* | ENSG00000207031.1 | snoRNA | iPSC_Neurons | 7.54 |
| OSAadjBMI | Pooled | rs12828563 | rs2958153 | *NACA* | ENSG00000196531.6 | protein_coding | hESC_HypothalamicNeuron | 7.5 |
| OSAadjBMI | Pooled | rs12828563 | rs941207 | *BAZ2A* | ENSG00000076108.7 | protein_coding | SGBS_Diff | 7.34 |
| OSAadjBMI | Pooled | rs12828563 | rs2958153 | *ATP5B* | ENSG00000110955.4 | protein_coding | hESC_HypothalamicNPC | 7.33 |
| OSAadjBMI | Pooled | rs12828563 | rs2958153 | *SNORD59A* | ENSG00000207031.1 | snoRNA | hESC_HypothalamicNPC | 7.33 |
| OSAadjBMI | Pooled | rs12828563 | rs2958153 | *U59B* | U59B | snoRNA | hESC_HypothalamicNPC | 7.33 |
| OSAadjBMI | Pooled | rs12828563 | rs941207 | *NACA* | ENSG00000196531.6 | protein_coding | hMSC_Adipocyte | 7.33 |
| OSAadjBMI | Pooled | rs12828563 | rs5798394 | *ZBTB39* | ENSG00000166860.2 | protein_coding | hESC_HypothalamicNeuron | 7.31 |
| OSAadjBMI | Pooled | rs12828563 | rs2255074 | *U59B* | U59B | snoRNA | iPSC_NPC | 7.29 |
| OSAadjBMI | Pooled | rs12828563 | rs1107479 | *U59B* | U59B | snoRNA | iPSC_NPC | 7.29 |
| OSAadjBMI | Pooled | rs12828563 | rs2958153 | *NACA* | ENSG00000196531.6 | protein_coding | NaiveB | 7.22 |
| OSAadjBMI | Pooled | rs12828563 | rs941207 | *ATP5B* | ENSG00000110955.4 | protein_coding | SGBS_Diff | 7.22 |
| OSAadjBMI | Pooled | rs12828563 | rs941207 | *SNORD59A* | ENSG00000207031.1 | snoRNA | SGBS_Diff | 7.22 |
| OSAadjBMI | Pooled | rs12828563 | rs941207 | *U59B* | U59B | snoRNA | SGBS_Diff | 7.22 |
| OSAadjBMI | Pooled | rs12828563 | rs2255074 | *BAZ2A* | ENSG00000076108.7 | protein_coding | HEPG2 | 7.16 |
| OSAadjBMI | Pooled | rs12828563 | rs1107479 | *BAZ2A* | ENSG00000076108.7 | protein_coding | HEPG2 | 7.16 |
| OSAadjBMI | Pooled | rs12828563 | rs3214051 | *BAZ2A* | ENSG00000076108.7 | protein_coding | hESC_HypothalamicNeuron | 7.15 |
| OSAadjBMI | Pooled | rs12828563 | rs3214051 | *GPR182* | ENSG00000166856.1 | protein_coding | iPSC_NPC | 7.15 |
| OSAadjBMI | Pooled | rs12828563 | rs3214051 | RP11-474N8.5 | ENSG00000258442.1 | protein_coding | iPSC_NPC | 7.15 |
| OSAadjBMI | Pooled | rs12828563 | rs3214051 | *ZBTB39* | ENSG00000166860.2 | protein_coding | HEPG2 | 7.14 |
| OSAadjBMI | Pooled | rs12828563 | rs941207 | *ESYT1* | ENSG00000139641.8 | protein_coding | iPSC_NPC | 7.09 |
| OSAadjBMI | Pooled | rs12828563 | rs941207 | *ZC3H10* | ENSG00000135482.2 | protein_coding | iPSC_NPC | 7.09 |
| OSAadjBMI | Pooled | rs12828563 | rs941207 | *MIP* | ENSG00000135517.6 | protein_coding | iPSC_NPC | 7.05 |
| OSAadjBMI | Pooled | rs12828563 | rs941207 | *SPRYD4* | ENSG00000176422.10 | protein_coding | iPSC_NPC | 7.05 |
| OSAadjBMI | Pooled | rs12828563 | rs2255074 | *ZBTB39* | ENSG00000166860.3 | protein_coding | Chondrocytes | 7.02 |
| OSAadjBMI | Pooled | rs12828563 | rs1107479 | *ZBTB39* | ENSG00000166860.3 | protein_coding | Chondrocytes | 7.02 |
| OSAadjBMI | Pooled | rs12828563 | rs941207 | *NAB2* | ENSG00000166886.8 | protein_coding | iPSC_NPC | 7.02 |
| OSAadjBMI | Pooled | rs12828563 | rs941207 | TCONS_00020445 | TCONS_00020445 | lincRNA | iPSC_NPC | 7.02 |
| OSAadjBMI | Pooled | rs12828563 | rs941207 | TCONS_00020805 | TCONS_00020805 | lincRNA | iPSC_NPC | 7.02 |
| OSAadjBMI | Pooled | rs12828563 | rs941207 | TCONS_00020806 | TCONS_00020806 | lincRNA | iPSC_NPC | 7.02 |
| OSAadjBMI | Pooled | rs12828563 | rs941207 | TCONS_00020807 | TCONS_00020807 | lincRNA | iPSC_NPC | 7.02 |
| OSAadjBMI | Pooled | rs12828563 | rs941207 | TCONS_00021326 | TCONS_00021326 | lincRNA | iPSC_NPC | 7.02 |
| OSAadjBMI | Pooled | rs12828563 | rs941207 | TCONS_00021327 | TCONS_00021327 | lincRNA | iPSC_NPC | 7.02 |
| OSAadjBMI | Pooled | rs12828563 | rs941207 | *TMEM194A* | ENSG00000166881.5 | protein_coding | iPSC_NPC | 7.02 |
| OSAadjBMI | Pooled | rs12828563 | rs2255074 | *ESYT1* | ENSG00000139641.8 | protein_coding | iPSC_NPC | 6.99 |
| OSAadjBMI | Pooled | rs12828563 | rs1107479 | *ESYT1* | ENSG00000139641.8 | protein_coding | iPSC_NPC | 6.99 |
| OSAadjBMI | Pooled | rs12828563 | rs2958153 | *ZBTB39* | ENSG00000166860.2 | protein_coding | GCB | 6.98 |
| OSAadjBMI | Pooled | rs12828563 | rs1107479 | *ATP5B* | ENSG00000110955.4 | protein_coding | hESC_HypothalamicNPC | 6.96 |
| OSAadjBMI | Pooled | rs12828563 | rs1107479 | *SNORD59A* | ENSG00000207031.1 | snoRNA | hESC_HypothalamicNPC | 6.96 |
| OSAadjBMI | Pooled | rs12828563 | rs1107479 | *U59B* | U59B | snoRNA | hESC_HypothalamicNPC | 6.96 |
| OSAadjBMI | Pooled | rs12828563 | rs3214051 | *SDR9C7* | ENSG00000170426.1 | protein_coding | hFOB | 6.82 |
| OSAadjBMI | Pooled | rs12828563 | rs941207 | *COQ10A* | ENSG00000135469.9 | protein_coding | hESC_HypothalamicNeuron | 6.81 |
| OSAadjBMI | Pooled | rs12828563 | rs941207 | *ESYT1* | ENSG00000139641.8 | protein_coding | NaiveB | 6.8 |
| OSAadjBMI | Pooled | rs12828563 | rs941207 | RP11-603J24.6 | ENSG00000258345.1 | antisense | NaiveB | 6.8 |
| OSAadjBMI | Pooled | rs12828563 | rs941207 | *ZC3H10* | ENSG00000135482.2 | protein_coding | NaiveB | 6.8 |
| OSAadjBMI | Pooled | rs12828563 | rs2958153 | *ATP5B* | ENSG00000110955.4 | protein_coding | GCB | 6.75 |
| OSAadjBMI | Pooled | rs12828563 | rs2958153 | *SNORD59A* | ENSG00000207031.1 | snoRNA | GCB | 6.75 |
| OSAadjBMI | Pooled | rs12828563 | rs2958153 | *U59B* | U59B | snoRNA | GCB | 6.75 |
| OSAadjBMI | Pooled | rs12828563 | rs941207 | *RBMS2* | ENSG00000076067.7 | protein_coding | hESC_HypothalamicNPC | 6.74 |
| OSAadjBMI | Pooled | rs12828563 | rs2958153 | *NACA* | ENSG00000196531.6 | protein_coding | iPSC_Neurons | 6.71 |
| OSAadjBMI | Pooled | rs12828563 | rs941207 | *STAT6* | ENSG00000166888.6 | protein_coding | iPSC_NPC | 6.71 |
| OSAadjBMI | Pooled | rs12828563 | rs941207 | RP11-603J24.6 | ENSG00000258345.1 | antisense | iPSC_NPC | 6.69 |
| OSAadjBMI | Pooled | rs12828563 | rs2958153 | *ZBTB39* | ENSG00000166860.2 | protein_coding | hESC_HypothalamicNeuron | 6.68 |
| OSAadjBMI | Pooled | rs12828563 | rs3214051 | *NACA* | ENSG00000196531.6 | protein_coding | GCB | 6.67 |
| OSAadjBMI | Pooled | rs12828563 | rs2958153 | *BAZ2A* | ENSG00000076108.7 | protein_coding | iPSC_NPC | 6.65 |

| **Supplementary Table 16.** Prioritized effector genes from variant to gene (V2G) mapping analysis. | | | | | | | | |
|---|---|---|---|---|---|---|---|---|
| **Trait** | **Discovery Study** | **Sentinel rsID** | **Proxy rsID** | **Gene Symbol** | **Gene Id** | **Gene Biotype** | **Cell** | **Contact Score (Chicago Score\|-log10(FitHiC2 FDR)** |
| OSAadjBMI | Pooled | rs12828563 | rs3214051 | *ZBTB39* | ENSG00000166860.2 | protein_coding | hESC_HypothalamicNPC | 6.56 |
| OSAadjBMI | Pooled | rs12828563 | rs2255074 | *ZBTB39* | ENSG00000166860.2 | protein_coding | hESC_HypothalamicNeuron | 6.55 |
| OSAadjBMI | Pooled | rs12828563 | rs1107479 | *ZBTB39* | ENSG00000166860.2 | protein_coding | hESC_HypothalamicNeuron | 6.55 |
| OSAadjBMI | Pooled | rs12828563 | rs1107479 | *BAZ2A* | ENSG00000076108.7 | protein_coding | hESC_HypothalamicNPC | 6.55 |
| OSAadjBMI | Pooled | rs12828563 | rs3214051 | *NACA* | ENSG00000196531.10 | protein_coding | Chondrocytes | 6.54 |
| OSAadjBMI | Pooled | rs12828563 | rs3214051 | *U59B* | U59B | snoRNA | TFH | 6.51 |
| OSAadjBMI | Pooled | rs12828563 | rs3214051 | *MYO1A* | ENSG00000166866.13 | protein_coding | Chondrocytes | 6.49 |
| OSAadjBMI | Pooled | rs12828563 | rs3214051 | *ZBTB39* | ENSG00000166860.2 | protein_coding | iPSC_Hepatocytes | 6.46 |
| OSAadjBMI | Pooled | rs12828563 | rs2255074 | *NACA* | ENSG00000196531.6 | protein_coding | SGBS_Diff | 6.46 |
| OSAadjBMI | Pooled | rs12828563 | rs1107479 | *NACA* | ENSG00000196531.6 | protein_coding | SGBS_Diff | 6.46 |
| OSAadjBMI | Pooled | rs12828563 | rs2255074 | *MIP* | ENSG00000135517.6 | protein_coding | iPSC_NPC | 6.44 |
| OSAadjBMI | Pooled | rs12828563 | rs2255074 | *SPRYD4* | ENSG00000176422.10 | protein_coding | iPSC_NPC | 6.44 |
| OSAadjBMI | Pooled | rs12828563 | rs1107479 | *MIP* | ENSG00000135517.6 | protein_coding | iPSC_NPC | 6.44 |
| OSAadjBMI | Pooled | rs12828563 | rs1107479 | *SPRYD4* | ENSG00000176422.10 | protein_coding | iPSC_NPC | 6.44 |
| OSAadjBMI | Pooled | rs12828563 | rs3214051 | *ATP5B* | ENSG00000110955.4 | protein_coding | NaiveT | 6.43 |
| OSAadjBMI | Pooled | rs12828563 | rs3214051 | *SNORD59A* | ENSG00000207031.1 | snoRNA | NaiveT | 6.43 |
| OSAadjBMI | Pooled | rs12828563 | rs3214051 | *U59B* | U59B | snoRNA | NaiveT | 6.43 |
| OSAadjBMI | Pooled | rs12828563 | rs941207 | *ATP5B* | ENSG00000110955.4 | protein_coding | NaiveT | 6.4 |
| OSAadjBMI | Pooled | rs12828563 | rs941207 | *U59B* | U59B | snoRNA | NaiveT | 6.4 |
| OSAadjBMI | Pooled | rs12828563 | rs941207 | *SNORD59A* | ENSG00000207031.1 | snoRNA | NaiveT | 6.4 |
| OSAadjBMI | Pooled | rs12828563 | rs2958153 | *BAZ2A* | ENSG00000076108.7 | protein_coding | TFH | 6.25 |
| OSAadjBMI | Pooled | rs12828563 | rs2255074 | *BAZ2A* | ENSG00000076108.7 | protein_coding | iPSC_Neurons | 6.23 |
| OSAadjBMI | Pooled | rs12828563 | rs1107479 | *BAZ2A* | ENSG00000076108.7 | protein_coding | iPSC_Neurons | 6.23 |
| OSAadjBMI | Pooled | rs12828563 | rs2958153 | *ZBTB39* | ENSG00000166860.2 | protein_coding | hMSC_Osteoblasts | 6.17 |
| OSAadjBMI | Pooled | rs12828563 | rs2958153 | *ATP5B* | ENSG00000110955.4 | protein_coding | hMSC_Adipocyte | 6.14 |
| OSAadjBMI | Pooled | rs12828563 | rs2958153 | *SNORD59A* | ENSG00000207031.1 | snoRNA | hMSC_Adipocyte | 6.14 |
| OSAadjBMI | Pooled | rs12828563 | rs2958153 | *U59B* | U59B | snoRNA | hMSC_Adipocyte | 6.14 |
| OSAadjBMI | Pooled | rs12828563 | rs2255074 | *BAZ2A* | ENSG00000076108.7 | protein_coding | NaiveT | 6.13 |
| OSAadjBMI | Pooled | rs12828563 | rs1107479 | *BAZ2A* | ENSG00000076108.7 | protein_coding | NaiveT | 6.13 |
| OSAadjBMI | Pooled | rs12828563 | rs3214051 | *BAZ2A* | ENSG00000076108.7 | protein_coding | iPSC_NPC | 6.1 |
| OSAadjBMI | Pooled | rs12828563 | rs2958153 | *ATP5B* | ENSG00000110955.4 | protein_coding | NaiveT | 6.07 |
| OSAadjBMI | Pooled | rs12828563 | rs2958153 | *SNORD59A* | ENSG00000207031.1 | snoRNA | NaiveT | 6.07 |
| OSAadjBMI | Pooled | rs12828563 | rs2958153 | *U59B* | U59B | snoRNA | NaiveT | 6.07 |
| OSAadjBMI | Pooled | rs12828563 | rs2255074 | *NACA* | ENSG00000196531.6 | protein_coding | iPSC_NPC | 6.02 |
| OSAadjBMI | Pooled | rs12828563 | rs1107479 | *NACA* | ENSG00000196531.6 | protein_coding | iPSC_NPC | 6.02 |
| OSAadjBMI | Pooled | rs12828563 | rs10747769 | *ATP5B* | ENSG00000110955.4 | protein_coding | TFH | 5.97 |
| OSAadjBMI | Pooled | rs12828563 | rs10747769 | *SNORD59A* | ENSG00000207031.1 | snoRNA | TFH | 5.97 |
| OSAadjBMI | Pooled | rs12828563 | rs10747769 | *U59B* | U59B | snoRNA | TFH | 5.97 |
| OSAadjBMI | Pooled | rs12828563 | rs2255074 | *ZBTB39* | ENSG00000166860.2 | protein_coding | hMSC_Osteoblasts | 5.95 |
| OSAadjBMI | Pooled | rs12828563 | rs1107479 | *ZBTB39* | ENSG00000166860.2 | protein_coding | hMSC_Osteoblasts | 5.95 |
| OSAadjBMI | Pooled | rs12828563 | rs941207 | *ATP5B* | ENSG00000110955.4 | protein_coding | hESC_HypothalamicNPC | 5.94 |
| OSAadjBMI | Pooled | rs12828563 | rs941207 | *SNORD59A* | ENSG00000207031.1 | snoRNA | hESC_HypothalamicNPC | 5.94 |
| OSAadjBMI | Pooled | rs12828563 | rs941207 | *U59B* | U59B | snoRNA | hESC_HypothalamicNPC | 5.94 |
| OSAadjBMI | Pooled | rs12828563 | rs2958153 | *BAZ2A* | ENSG00000076108.7 | protein_coding | iPSC_Neurons | 5.94 |
| OSAadjBMI | Pooled | rs12828563 | rs3214051 | *ATP5B* | ENSG00000110955.4 | protein_coding | HEPG2 | 5.9 |
| OSAadjBMI | Pooled | rs12828563 | rs3214051 | *SNORD59A* | ENSG00000207031.1 | snoRNA | HEPG2 | 5.9 |
| OSAadjBMI | Pooled | rs12828563 | rs3214051 | *U59B* | U59B | snoRNA | HEPG2 | 5.9 |
| OSAadjBMI | Pooled | rs12828563 | rs941207 | *ATP5B* | ENSG00000110955.4 | protein_coding | hMSC_Adipocyte | 5.89 |
| OSAadjBMI | Pooled | rs12828563 | rs941207 | *SNORD59A* | ENSG00000207031.1 | snoRNA | hMSC_Adipocyte | 5.89 |
| OSAadjBMI | Pooled | rs12828563 | rs941207 | *U59B* | U59B | snoRNA | hMSC_Adipocyte | 5.89 |
| OSAadjBMI | Pooled | rs12828563 | rs941207 | *NACA* | ENSG00000196531.6 | protein_coding | GCB | 5.87 |
| OSAadjBMI | Pooled | rs12828563 | rs2255074 | *NAB2* | ENSG00000166886.8 | protein_coding | NaiveB | 5.87 |
| OSAadjBMI | Pooled | rs12828563 | rs2255074 | TCONS_00020445 | TCONS_00020445 | lincRNA | NaiveB | 5.87 |
| OSAadjBMI | Pooled | rs12828563 | rs2255074 | TCONS_00020805 | TCONS_00020805 | lincRNA | NaiveB | 5.87 |
| OSAadjBMI | Pooled | rs12828563 | rs2255074 | TCONS_00020806 | TCONS_00020806 | lincRNA | NaiveB | 5.87 |
| OSAadjBMI | Pooled | rs12828563 | rs2255074 | TCONS_00020807 | TCONS_00020807 | lincRNA | NaiveB | 5.87 |
| OSAadjBMI | Pooled | rs12828563 | rs2255074 | TCONS_00021326 | TCONS_00021326 | lincRNA | NaiveB | 5.87 |
| OSAadjBMI | Pooled | rs12828563 | rs2255074 | TCONS_00021327 | TCONS_00021327 | lincRNA | NaiveB | 5.87 |
| OSAadjBMI | Pooled | rs12828563 | rs2255074 | *TMEM194A* | ENSG00000166881.5 | protein_coding | NaiveB | 5.87 |
| OSAadjBMI | Pooled | rs12828563 | rs1107479 | *NAB2* | ENSG00000166886.8 | protein_coding | NaiveB | 5.87 |
| OSAadjBMI | Pooled | rs12828563 | rs1107479 | TCONS_00020445 | TCONS_00020445 | lincRNA | NaiveB | 5.87 |
| OSAadjBMI | Pooled | rs12828563 | rs1107479 | TCONS_00020805 | TCONS_00020805 | lincRNA | NaiveB | 5.87 |
| OSAadjBMI | Pooled | rs12828563 | rs1107479 | TCONS_00020806 | TCONS_00020806 | lincRNA | NaiveB | 5.87 |
| OSAadjBMI | Pooled | rs12828563 | rs1107479 | TCONS_00020807 | TCONS_00020807 | lincRNA | NaiveB | 5.87 |
| OSAadjBMI | Pooled | rs12828563 | rs1107479 | TCONS_00021326 | TCONS_00021326 | lincRNA | NaiveB | 5.87 |
| OSAadjBMI | Pooled | rs12828563 | rs1107479 | TCONS_00021327 | TCONS_00021327 | lincRNA | NaiveB | 5.87 |
| OSAadjBMI | Pooled | rs12828563 | rs1107479 | *TMEM194A* | ENSG00000166881.5 | protein_coding | NaiveB | 5.87 |
| OSAadjBMI | Pooled | rs12828563 | rs2958153 | TCONS_00020445 | TCONS_00020445 | lincRNA | iPSC_NPC | 5.8 |
| OSAadjBMI | Pooled | rs12828563 | rs2958153 | TCONS_00020807 | TCONS_00020807 | lincRNA | iPSC_NPC | 5.8 |
| OSAadjBMI | Pooled | rs12828563 | rs941207 | *BAZ2A* | ENSG00000076108.7 | protein_coding | hESC_HypothalamicNPC | 5.79 |
| OSAadjBMI | Pooled | rs12828563 | rs941207 | *BAZ2A* | ENSG00000076108.7 | protein_coding | hMSC_Osteoblasts | 5.77 |
| OSAadjBMI | Pooled | rs12828563 | rs2255074 | *NACA* | ENSG00000196531.6 | protein_coding | GCB | 5.76 |
| OSAadjBMI | Pooled | rs12828563 | rs1107479 | *NACA* | ENSG00000196531.6 | protein_coding | GCB | 5.76 |
| OSAadjBMI | Pooled | rs12828563 | rs2958153 | *BAZ2A* | ENSG00000076108.7 | protein_coding | NaiveB | 5.7 |
| OSAadjBMI | Pooled | rs12828563 | rs2255074 | *RBMS2* | ENSG00000076067.7 | protein_coding | hESC_HypothalamicNeuron | 5.68 |
| OSAadjBMI | Pooled | rs12828563 | rs1107479 | *RBMS2* | ENSG00000076067.7 | protein_coding | hESC_HypothalamicNeuron | 5.68 |
| OSAadjBMI | Pooled | rs12828563 | rs2255074 | *RBMS2* | ENSG00000076067.7 | protein_coding | NaiveT | 5.64 |
| OSAadjBMI | Pooled | rs12828563 | rs1107479 | *RBMS2* | ENSG00000076067.7 | protein_coding | NaiveT | 5.64 |
| OSAadjBMI | Pooled | rs12828563 | rs2255074 | *RBMS2* | ENSG00000076067.7 | protein_coding | SGBS_Diff | 5.63 |
| OSAadjBMI | Pooled | rs12828563 | rs1107479 | *RBMS2* | ENSG00000076067.7 | protein_coding | SGBS_Diff | 5.63 |
| OSAadjBMI | Pooled | rs12828563 | rs2958153 | *BAZ2A* | ENSG00000076108.7 | protein_coding | SGBS_Diff | 5.57 |
| OSAadjBMI | Pooled | rs12828563 | rs941207 | *NACA* | ENSG00000196531.6 | protein_coding | hESC_HypothalamicNeuron | 5.55 |
| OSAadjBMI | Pooled | rs12828563 | rs2958153 | *ESYT1* | ENSG00000139641.8 | protein_coding | iPSC_Neurons | 5.51 |
| OSAadjBMI | Pooled | rs12828563 | rs2958153 | *ZC3H10* | ENSG00000135482.2 | protein_coding | iPSC_Neurons | 5.51 |
| OSAadjBMI | Pooled | rs12828563 | rs3214051 | *NACA* | ENSG00000196531.6 | protein_coding | hESC_HypothalamicNPC | 5.47 |

| **Supplementary Table 16.** Prioritized effector genes from variant to gene (V2G) mapping analysis. | | | | | | | | |
|---|---|---|---|---|---|---|---|---|
| **Trait** | **Discovery Study** | **Sentinel rsID** | **Proxy rsID** | **Gene Symbol** | **Gene Id** | **Gene Biotype** | **Cell** | **Contact Score (Chicago Score\|-log10(FitHiC2 FDR)** |
| OSAadjBMI | Pooled | rs12828563 | rs2255074 | *ATP5B* | ENSG00000110955.4 | protein_coding | iPSC_Neurons | 5.42 |
| OSAadjBMI | Pooled | rs12828563 | rs2255074 | *U59B* | U59B | snoRNA | iPSC_Neurons | 5.42 |
| OSAadjBMI | Pooled | rs12828563 | rs1107479 | *ATP5B* | ENSG00000110955.4 | protein_coding | iPSC_Neurons | 5.42 |
| OSAadjBMI | Pooled | rs12828563 | rs1107479 | *U59B* | U59B | snoRNA | iPSC_Neurons | 5.42 |
| OSAadjBMI | Pooled | rs12828563 | rs2255074 | *SNORD59A* | ENSG00000207031.1 | snoRNA | iPSC_Neurons | 5.42 |
| OSAadjBMI | Pooled | rs12828563 | rs1107479 | *SNORD59A* | ENSG00000207031.1 | snoRNA | iPSC_Neurons | 5.42 |
| OSAadjBMI | Pooled | rs12828563 | rs941207 | *BAZ2A* | ENSG00000076108.7 | protein_coding | iPSC_Neurons | 5.39 |
| OSAadjBMI | Pooled | rs12828563 | rs2958153 | *ZBTB39* | ENSG00000166860.2 | protein_coding | iPSC_NPC | 5.39 |
| OSAadjBMI | Pooled | rs12828563 | rs2958153 | *ATP5B* | ENSG00000110955.4 | protein_coding | TFH | 5.39 |
| OSAadjBMI | Pooled | rs12828563 | rs2958153 | *SNORD59A* | ENSG00000207031.1 | snoRNA | TFH | 5.39 |
| OSAadjBMI | Pooled | rs12828563 | rs941207 | *RBMS2* | ENSG00000076067.7 | protein_coding | NaiveT | 5.38 |
| OSAadjBMI | Pooled | rs12828563 | rs7980835 | *ATP5B* | ENSG00000110955.4 | protein_coding | NaiveT | 5.3 |
| OSAadjBMI | Pooled | rs12828563 | rs7980835 | *SNORD59A* | ENSG00000207031.1 | snoRNA | NaiveT | 5.3 |
| OSAadjBMI | Pooled | rs12828563 | rs7980835 | *U59B* | U59B | snoRNA | NaiveT | 5.3 |
| OSAadjBMI | Pooled | rs12828563 | rs10747769 | *ATP5B* | ENSG00000110955.4 | protein_coding | NaiveT | 5.3 |
| OSAadjBMI | Pooled | rs12828563 | rs10747769 | *SNORD59A* | ENSG00000207031.1 | snoRNA | NaiveT | 5.3 |
| OSAadjBMI | Pooled | rs12828563 | rs10747769 | *U59B* | U59B | snoRNA | NaiveT | 5.3 |
| OSAadjBMI | Pooled | rs12828563 | rs941207 | *U59B* | U59B | snoRNA | NaiveB | 5.28 |
| OSAadjBMI | Pooled | rs12828563 | rs2958153 | *ATP5B* | ENSG00000110955.4 | protein_coding | iPSC_Neurons | 5.27 |
| OSAadjBMI | Pooled | rs12828563 | rs2958153 | *SNORD59A* | ENSG00000207031.1 | snoRNA | iPSC_Neurons | 5.27 |
| OSAadjBMI | Pooled | rs12828563 | rs2958153 | *U59B* | U59B | snoRNA | iPSC_Neurons | 5.27 |
| OSAadjBMI | Pooled | rs12828563 | rs941207 | *APOF* | ENSG00000175336.8 | protein_coding | iPSC_NPC | 5.25 |
| OSAadjBMI | Pooled | rs12828563 | rs10747769 | *NACA* | ENSG00000196531.6 | protein_coding | TFH | 5.25 |
| OSAadjBMI | Pooled | rs12828563 | rs2255074 | *ATP5B* | ENSG00000110955.4 | protein_coding | HEPG2 | 5.2 |
| OSAadjBMI | Pooled | rs12828563 | rs2255074 | *SNORD59A* | ENSG00000207031.1 | snoRNA | HEPG2 | 5.2 |
| OSAadjBMI | Pooled | rs12828563 | rs2255074 | *U59B* | U59B | snoRNA | HEPG2 | 5.2 |
| OSAadjBMI | Pooled | rs12828563 | rs1107479 | *ATP5B* | ENSG00000110955.4 | protein_coding | HEPG2 | 5.2 |
| OSAadjBMI | Pooled | rs12828563 | rs1107479 | *SNORD59A* | ENSG00000207031.1 | snoRNA | HEPG2 | 5.2 |
| OSAadjBMI | Pooled | rs12828563 | rs1107479 | *U59B* | U59B | snoRNA | HEPG2 | 5.2 |
| OSAadjBMI | Pooled | rs12828563 | rs941207 | *NACA* | ENSG00000196531.6 | protein_coding | hMSC_Osteoblasts | 5.18 |
| OSAadjBMI | Pooled | rs12828563 | rs941207 | *ATP5B* | ENSG00000110955.4 | protein_coding | iPSC_NPC | 5.12 |
| OSAadjBMI | Pooled | rs12828563 | rs941207 | *SNORD59A* | ENSG00000207031.1 | snoRNA | iPSC_NPC | 5.12 |
| OSAadjBMI | Pooled | rs12828563 | rs941207 | *U59B* | U59B | snoRNA | iPSC_NPC | 5.12 |
| OSAadjBMI | Pooled | rs12828563 | rs941207 | *MYL6* | ENSG00000092841.14 | protein_coding | iPSC_NPC | 5.03 |
| OSAadjBMI | Pooled | rs12828563 | rs941207 | *MYL6B* | ENSG00000196465.6 | protein_coding | iPSC_NPC | 5.03 |
| OSAadjBMI | Pooled | rs12828563 | rs941207 | RP11-603J24.14 | ENSG00000257809.1 | antisense | iPSC_NPC | 5.03 |
| OSAadjBMI | Pooled | rs12828563 | rs2255074 | *NACA* | ENSG00000196531.6 | protein_coding | NaiveB | 5.02 |
| OSAadjBMI | Pooled | rs12828563 | rs1107479 | *NACA* | ENSG00000196531.6 | protein_coding | NaiveB | 5.02 |
| OSAadjBMI | Pooled | rs12828563 | rs2958153 | *ATP5B* | ENSG00000110955.4 | protein_coding | HEPG2 | 5.01 |
| OSAadjBMI | Pooled | rs12828563 | rs2958153 | *SNORD59A* | ENSG00000207031.1 | snoRNA | HEPG2 | 5.01 |
| OSAadjBMI | Pooled | rs12828563 | rs2958153 | *U59B* | U59B | snoRNA | HEPG2 | 5.01 |
| OSAadjBMI | Pooled | rs2964006 | rs2926854 | *GRIA1* | ENSG00000155511.13 | protein_coding | iPSC_Neurons | 5.72 |
| OSAadjBMI | Pooled | rs2964006 | rs2926852 | *GRIA1* | ENSG00000155511.13 | protein_coding | hESC_HypothalamicNeuron | 5.68 |
| OSAadjBMI | Pooled | rs2964006 | rs2926850 | *GRIA1* | ENSG00000155511.13 | protein_coding | iPSC_Neurons | 5.67 |
| OSAadjBMI | Pooled | rs2964006 | rs2926851 | *GRIA1* | ENSG00000155511.13 | protein_coding | iPSC_Neurons | 5.67 |
| OSAadjBMI | Pooled | rs2964006 | rs2926852 | *GRIA1* | ENSG00000155511.13 | protein_coding | iPSC_Neurons | 5.67 |
| OSAadjBMI | Pooled | rs2964006 | rs2926861 | *GRIA1* | ENSG00000155511.13 | protein_coding | SGBS_Diff | 5.5 |
| OSAadjBMI | Pooled | rs12828563 | rs3214051 | *ZBTB39* | ENSG00000166860.2 | protein_coding | TFH | 5.04 |
| OSA and OSAad | MVP (Sofer et al) | rs2277339 | rs2277339 | *GPR182* | ENSG00000166856.3 | protein_coding | Chondrocytes | 19.47 |
| OSA and OSAad | MVP (Sofer et al) | rs2277339 | rs2277339 | *GPR182* | ENSG00000166856.3 | protein_coding | Chondrocytes | 19.47 |
| OSA and OSAad | MVP (Sofer et al) | rs2277339 | rs2277339 | *ZBTB39* | ENSG00000166860.3 | protein_coding | Chondrocytes | 16.27 |
| OSA and OSAad | MVP (Sofer et al) | rs2277339 | rs2277339 | *ZBTB39* | ENSG00000166860.3 | protein_coding | Chondrocytes | 16.27 |
| OSA and OSAad | MVP (Sofer et al) | rs2277339 | rs2277339 | AC026120.1 | ENSG00000278399.1 | unprocessed_pseudogene | Chondrocytes | 9.35 |
| OSA and OSAad | MVP (Sofer et al) | rs2277339 | rs2277339 | AC026120.1 | ENSG00000278399.1 | unprocessed_pseudogene | Chondrocytes | 9.35 |
| OSA and OSAad | MVP (Sofer et al) | rs2277339 | rs2277339 | *HSD17B6* | ENSG00000025423.11 | protein_coding | Chondrocytes | 6.83 |
| OSA and OSAad | MVP (Sofer et al) | rs2277339 | rs2277339 | *HSD17B6* | ENSG00000025423.11 | protein_coding | Chondrocytes | 6.83 |
| OSA and OSAad | MVP (Sofer et al) | rs2277339 | rs2277339 | *NACA* | ENSG00000196531.6 | protein_coding | iPSC_NPC | 6.27 |
| OSA and OSAad | MVP (Sofer et al) | rs2277339 | rs2277339 | *NAB2* | ENSG00000166886.8 | protein_coding | GCB | 5.6 |
| OSA and OSAad | MVP (Sofer et al) | rs2277339 | rs2277339 | TCONS_00020445 | TCONS_00020445 | lincRNA | GCB | 5.6 |
| OSA and OSAad | MVP (Sofer et al) | rs2277339 | rs2277339 | TCONS_00020805 | TCONS_00020805 | lincRNA | GCB | 5.6 |
| OSA and OSAad | MVP (Sofer et al) | rs2277339 | rs2277339 | TCONS_00020806 | TCONS_00020806 | lincRNA | GCB | 5.6 |
| OSA and OSAad | MVP (Sofer et al) | rs2277339 | rs2277339 | TCONS_00020807 | TCONS_00020807 | lincRNA | GCB | 5.6 |
| OSA and OSAad | MVP (Sofer et al) | rs2277339 | rs2277339 | TCONS_00021326 | TCONS_00021326 | lincRNA | GCB | 5.6 |
| OSA and OSAad | MVP (Sofer et al) | rs2277339 | rs2277339 | TCONS_00021327 | TCONS_00021327 | lincRNA | GCB | 5.6 |
| OSA and OSAad | MVP (Sofer et al) | rs2277339 | rs2277339 | *TMEM194A* | ENSG00000166881.5 | protein_coding | GCB | 5.6 |
| OSA and OSAad | Pooled | rs2958127 | rs941207 | *RBMS2* | ENSG00000076067.13 | protein_coding | Chondrocytes | 15.57 |
| OSA and OSAad | Pooled | rs2958127 | rs1107479 | *RBMS2* | ENSG00000076067.13 | protein_coding | Chondrocytes | 10.55 |
| OSA and OSAad | Pooled | rs2958127 | rs2255074 | *RBMS2* | ENSG00000076067.13 | protein_coding | Chondrocytes | 10.55 |
| OSA and OSAad | Pooled | rs2958127 | rs941207 | *HSPD1P4* | ENSG00000257576.1 | processed_pseudogene | Chondrocytes | 9.65 |
| OSA and OSAad | Pooled | rs2958127 | rs941207 | *GLS2* | ENSG00000135423.13 | protein_coding | Chondrocytes | 6.95 |
| OSA and OSAad | Pooled | rs2958127 | rs941207 | AC097104.1 | ENSG00000285528.1 | protein_coding | Chondrocytes | 6.95 |
| OSA | EUR | rs1800437 | rs11672660 | *EML2* | ENSG00000125746.10 | protein_coding | TFH | 6.47 |
| OSA | EUR | rs1800437 | rs34783010 | *EML2* | ENSG00000125746.10 | protein_coding | TFH | 6.47 |
| OSA | EUR | rs2926743 | rs2958153 | *ATP5B* | ENSG00000110955.4 | protein_coding | hESC_HypothalamicNeuron | 18.15 |
| OSA | EUR | rs2926743 | rs2958153 | *U59B* | U59B | snoRNA | hESC_HypothalamicNeuron | 18.15 |
| OSA | EUR | rs2926743 | rs2958153 | *SNORD59A* | ENSG00000207031.1 | snoRNA | hESC_HypothalamicNeuron | 18.15 |
| OSA | EUR | rs2926743 | rs941207 | *BAZ2A* | ENSG00000076108.7 | protein_coding | hMSC_Adipocyte | 14.07 |
| OSA | EUR | rs2926743 | rs2256341 | *BAZ2A* | ENSG00000076108.11 | protein_coding | Chondrocytes | 14.04 |
| OSA | EUR | rs2926743 | rs941207 | *BAZ2A* | ENSG00000076108.7 | protein_coding | hESC_HypothalamicNeuron | 13.25 |
| OSA | EUR | rs2926743 | rs2958153 | *BAZ2A* | ENSG00000076108.7 | protein_coding | iPSC_Hepatocytes | 13.23 |
| OSA | EUR | rs2926743 | rs941207 | *RBMS2* | ENSG00000076067.7 | protein_coding | SGBS_Diff | 12.12 |
| OSA | EUR | rs2926743 | rs941207 | *ATP5B* | ENSG00000110955.4 | protein_coding | GCB | 11.88 |
| OSA | EUR | rs2926743 | rs941207 | *SNORD59A* | ENSG00000207031.1 | snoRNA | GCB | 11.88 |
| OSA | EUR | rs2926743 | rs941207 | *U59B* | U59B | snoRNA | GCB | 11.88 |
| OSA | EUR | rs2926743 | rs941207 | *ATP5B* | ENSG00000110955.4 | protein_coding | hESC_HypothalamicNeuron | 11.73 |

**Supplementary Table 16.** Prioritized effector genes from variant to gene (V2G) mapping analysis.

| Trait | Discovery Study | Sentinel rsID | Proxy rsID | Gene Symbol | Gene Id | Gene Biotype | Cell | Contact Score (Chicago Score\|-log10(FitHiC2 FDR) |
|---|---|---|---|---|---|---|---|---|
| OSA | EUR | rs2926743 | rs941207 | *U59B* | U59B | snoRNA | hESC_HypothalamicNeuron | 11.73 |
| OSA | EUR | rs2926743 | rs941207 | *SNORD59A* | ENSG00000207031.1 | snoRNA | hESC_HypothalamicNeuron | 11.73 |
| OSA | EUR | rs2926743 | rs2958153 | *BAZ2A* | ENSG00000076108.7 | protein_coding | NaiveT | 11.48 |
| OSA | EUR | rs2926743 | rs2958153 | *ATP5B* | ENSG00000110955.4 | protein_coding | iPSC_NPC | 11.45 |
| OSA | EUR | rs2926743 | rs2958153 | *U59B* | U59B | snoRNA | iPSC_NPC | 11.45 |
| OSA | EUR | rs2926743 | rs2958153 | *SNORD59A* | ENSG00000207031.1 | snoRNA | iPSC_NPC | 11.45 |
| OSA | EUR | rs2926743 | rs2958153 | *NACA* | ENSG00000196531.6 | protein_coding | iPSC_Hepatocytes | 10.56 |
| OSA | EUR | rs2926743 | rs941207 | *RBMS2* | ENSG00000076067.7 | protein_coding | iPSC_NPC | 10.48 |
| OSA | EUR | rs2926743 | rs941207 | *BAZ2A* | ENSG00000076108.7 | protein_coding | NaiveB | 10.34 |
| OSA | EUR | rs2926743 | rs2958153 | *NACA* | ENSG00000196531.6 | protein_coding | GCB | 10.29 |
| OSA | EUR | rs2926743 | rs941207 | *BAZ2A* | ENSG00000076108.7 | protein_coding | GCB | 9.91 |
| OSA | EUR | rs2926743 | rs2958153 | *NACA* | ENSG00000196531.6 | protein_coding | TFH | 9.58 |
| OSA | EUR | rs2926743 | rs2958153 | *NACA* | ENSG00000196531.6 | protein_coding | hMSC_Adipocyte | 9.57 |
| OSA | EUR | rs2926743 | rs941207 | *ATP5B* | ENSG00000110955.4 | protein_coding | TFH | 9.57 |
| OSA | EUR | rs2926743 | rs941207 | *SNORD59A* | ENSG00000207031.1 | snoRNA | TFH | 9.57 |
| OSA | EUR | rs2926743 | rs941207 | *U59B* | U59B | snoRNA | TFH | 9.57 |
| OSA | EUR | rs2926743 | rs2958153 | *ATP5B* | ENSG00000110955.4 | protein_coding | iPSC_Hepatocytes | 9.48 |
| OSA | EUR | rs2926743 | rs2958153 | *SNORD59A* | ENSG00000207031.1 | snoRNA | iPSC_Hepatocytes | 9.48 |
| OSA | EUR | rs2926743 | rs2958153 | *U59B* | U59B | snoRNA | iPSC_Hepatocytes | 9.48 |
| OSA | EUR | rs2926743 | rs941207 | *RBMS2* | ENSG00000076067.7 | protein_coding | hMSC_Adipocyte | 9.43 |
| OSA | EUR | rs2926743 | rs941207 | *RBMS2* | ENSG00000076067.7 | protein_coding | hESC_HypothalamicNeuron | 9.42 |
| OSA | EUR | rs2926743 | rs941207 | *NACA* | ENSG00000196531.6 | protein_coding | NaiveT | 9.25 |
| OSA | EUR | rs2926743 | rs2958153 | *ATP5B* | ENSG00000110955.4 | protein_coding | hMSC_Osteoblasts | 9.21 |
| OSA | EUR | rs2926743 | rs2958153 | *SNORD59A* | ENSG00000207031.1 | snoRNA | hMSC_Osteoblasts | 9.21 |
| OSA | EUR | rs2926743 | rs2958153 | *U59B* | U59B | snoRNA | hMSC_Osteoblasts | 9.21 |
| OSA | EUR | rs2926743 | rs2958153 | *ATP5B* | ENSG00000110955.4 | protein_coding | SGBS_Diff | 9.12 |
| OSA | EUR | rs2926743 | rs2958153 | *U59B* | U59B | snoRNA | SGBS_Diff | 9.12 |
| OSA | EUR | rs2926743 | rs2958153 | *SNORD59A* | ENSG00000207031.1 | snoRNA | SGBS_Diff | 9.12 |
| OSA | EUR | rs2926743 | rs2958153 | *NACA* | ENSG00000196531.6 | protein_coding | NaiveT | 9.01 |
| OSA | EUR | rs2926743 | rs2958153 | *NACA* | ENSG00000196531.6 | protein_coding | SGBS_Diff | 8.85 |
| OSA | EUR | rs2926743 | rs941207 | *ATP5B* | ENSG00000110955.4 | protein_coding | NaiveB | 8.83 |
| OSA | EUR | rs2926743 | rs941207 | *SNORD59A* | ENSG00000207031.1 | snoRNA | NaiveB | 8.83 |
| OSA | EUR | rs2926743 | rs941207 | *BAZ2A* | ENSG00000076108.7 | protein_coding | TFH | 8.75 |
| OSA | EUR | rs2926743 | rs941207 | *RBMS2* | ENSG00000076067.7 | protein_coding | iPSC_Neurons | 8.74 |
| OSA | EUR | rs2926743 | rs2958153 | *NACA* | ENSG00000196531.6 | protein_coding | iPSC_NPC | 8.68 |
| OSA | EUR | rs2926743 | rs941207 | *BAZ2A* | ENSG00000076108.7 | protein_coding | iPSC_NPC | 8.63 |
| OSA | EUR | rs2926743 | rs941207 | *ZBTB39* | ENSG00000166860.2 | protein_coding | hESC_HypothalamicNeuron | 8.22 |
| OSA | EUR | rs2926743 | rs2958153 | *ZBTB39* | ENSG00000166860.2 | protein_coding | iPSC_Hepatocytes | 8.21 |
| OSA | EUR | rs2926743 | rs2958153 | *NACA* | ENSG00000196531.6 | protein_coding | hMSC_Osteoblasts | 8.08 |
| OSA | EUR | rs2926743 | rs2958153 | *BAZ2A* | ENSG00000076108.7 | protein_coding | hMSC_Osteoblasts | 8 |
| OSA | EUR | rs2926743 | rs941207 | *NACA* | ENSG00000196531.6 | protein_coding | TFH | 7.64 |
| OSA | EUR | rs2926743 | rs2958153 | *ZBTB39* | ENSG00000166860.2 | protein_coding | iPSC_Neurons | 7.57 |
| OSA | EUR | rs2926743 | rs2958153 | *NACA* | ENSG00000196531.6 | protein_coding | hESC_HypothalamicNeuron | 7.5 |
| OSA | EUR | rs2926743 | rs941207 | *BAZ2A* | ENSG00000076108.7 | protein_coding | SGBS_Diff | 7.34 |
| OSA | EUR | rs2926743 | rs2958153 | *ATP5B* | ENSG00000110955.4 | protein_coding | hESC_HypothalamicNPC | 7.33 |
| OSA | EUR | rs2926743 | rs2958153 | *SNORD59A* | ENSG00000207031.1 | snoRNA | hESC_HypothalamicNPC | 7.33 |
| OSA | EUR | rs2926743 | rs2958153 | *U59B* | U59B | snoRNA | hESC_HypothalamicNPC | 7.33 |
| OSA | EUR | rs2926743 | rs941207 | *NACA* | ENSG00000196531.6 | protein_coding | hMSC_Adipocyte | 7.33 |
| OSA | EUR | rs2926743 | rs2958153 | *NACA* | ENSG00000196531.6 | protein_coding | NaiveB | 7.22 |
| OSA | EUR | rs2926743 | rs941207 | *ATP5B* | ENSG00000110955.4 | protein_coding | SGBS_Diff | 7.22 |
| OSA | EUR | rs2926743 | rs941207 | *SNORD59A* | ENSG00000207031.1 | snoRNA | SGBS_Diff | 7.22 |
| OSA | EUR | rs2926743 | rs941207 | *U59B* | U59B | snoRNA | SGBS_Diff | 7.22 |
| OSA | EUR | rs2926743 | rs941207 | *ESYT1* | ENSG00000139641.8 | protein_coding | iPSC_NPC | 7.09 |
| OSA | EUR | rs2926743 | rs941207 | *ZC3H10* | ENSG00000135482.2 | protein_coding | iPSC_NPC | 7.09 |
| OSA | EUR | rs2926743 | rs941207 | *MIP* | ENSG00000135517.6 | protein_coding | iPSC_NPC | 7.05 |
| OSA | EUR | rs2926743 | rs941207 | *SPRYD4* | ENSG00000176422.10 | protein_coding | iPSC_NPC | 7.05 |
| OSA | EUR | rs2926743 | rs941207 | *NAB2* | ENSG00000166886.8 | protein_coding | iPSC_NPC | 7.02 |
| OSA | EUR | rs2926743 | rs941207 | TCONS_00020445 | TCONS_00020445 | lincRNA | iPSC_NPC | 7.02 |
| OSA | EUR | rs2926743 | rs941207 | TCONS_00020805 | TCONS_00020805 | lincRNA | iPSC_NPC | 7.02 |
| OSA | EUR | rs2926743 | rs941207 | TCONS_00020806 | TCONS_00020806 | lincRNA | iPSC_NPC | 7.02 |
| OSA | EUR | rs2926743 | rs941207 | TCONS_00020807 | TCONS_00020807 | lincRNA | iPSC_NPC | 7.02 |
| OSA | EUR | rs2926743 | rs941207 | TCONS_00021326 | TCONS_00021326 | lincRNA | iPSC_NPC | 7.02 |
| OSA | EUR | rs2926743 | rs941207 | TCONS_00021327 | TCONS_00021327 | lincRNA | iPSC_NPC | 7.02 |
| OSA | EUR | rs2926743 | rs941207 | *TMEM194A* | ENSG00000166881.5 | protein_coding | iPSC_NPC | 7.02 |
| OSA | EUR | rs2926743 | rs2958153 | *ZBTB39* | ENSG00000166860.2 | protein_coding | GCB | 6.98 |
| OSA | EUR | rs2926743 | rs941207 | *COQ10A* | ENSG00000135469.9 | protein_coding | hESC_HypothalamicNeuron | 6.81 |
| OSA | EUR | rs2926743 | rs941207 | *ESYT1* | ENSG00000139641.8 | protein_coding | NaiveB | 6.8 |
| OSA | EUR | rs2926743 | rs941207 | RP11-603J24.6 | ENSG00000258345.1 | antisense | NaiveB | 6.8 |
| OSA | EUR | rs2926743 | rs941207 | *ZC3H10* | ENSG00000135482.2 | protein_coding | NaiveB | 6.8 |
| OSA | EUR | rs2926743 | rs2958153 | *ATP5B* | ENSG00000110955.4 | protein_coding | GCB | 6.75 |
| OSA | EUR | rs2926743 | rs2958153 | *SNORD59A* | ENSG00000207031.1 | snoRNA | GCB | 6.75 |
| OSA | EUR | rs2926743 | rs2958153 | *U59B* | U59B | snoRNA | GCB | 6.75 |
| OSA | EUR | rs2926743 | rs941207 | *RBMS2* | ENSG00000076067.7 | protein_coding | hESC_HypothalamicNPC | 6.74 |
| OSA | EUR | rs2926743 | rs2958153 | *NACA* | ENSG00000196531.6 | protein_coding | iPSC_Neurons | 6.71 |
| OSA | EUR | rs2926743 | rs941207 | *STAT6* | ENSG00000166888.6 | protein_coding | iPSC_NPC | 6.71 |
| OSA | EUR | rs2926743 | rs941207 | RP11-603J24.6 | ENSG00000258345.1 | antisense | iPSC_NPC | 6.69 |
| OSA | EUR | rs2926743 | rs2958153 | *ZBTB39* | ENSG00000166860.2 | protein_coding | hESC_HypothalamicNeuron | 6.68 |
| OSA | EUR | rs2926743 | rs2958153 | *BAZ2A* | ENSG00000076108.7 | protein_coding | iPSC_NPC | 6.65 |
| OSA | EUR | rs2926743 | rs941207 | *ATP5B* | ENSG00000110955.4 | protein_coding | NaiveT | 6.4 |
| OSA | EUR | rs2926743 | rs941207 | *U59B* | U59B | snoRNA | NaiveT | 6.4 |
| OSA | EUR | rs2926743 | rs941207 | *SNORD59A* | ENSG00000207031.1 | snoRNA | NaiveT | 6.4 |
| OSA | EUR | rs2926743 | rs2958153 | *BAZ2A* | ENSG00000076108.7 | protein_coding | TFH | 6.25 |
| OSA | EUR | rs2926743 | rs2958153 | *ZBTB39* | ENSG00000166860.2 | protein_coding | hMSC_Osteoblasts | 6.17 |
| OSA | EUR | rs2926743 | rs2958153 | *ATP5B* | ENSG00000110955.4 | protein_coding | hMSC_Adipocyte | 6.14 |
| OSA | EUR | rs2926743 | rs2958153 | *SNORD59A* | ENSG00000207031.1 | snoRNA | hMSC_Adipocyte | 6.14 |

**Supplementary Table 16.** Prioritized effector genes from variant to gene (V2G) mapping analysis.

| Trait | Discovery Study | Sentinel rsID | Proxy rsID | Gene Symbol | Gene Id | Gene Biotype | Cell | Contact Score (Chicago Score\|-log10(FitHiC2 FDR) |
|---|---|---|---|---|---|---|---|---|
| OSA | EUR | rs2926743 | rs2958153 | *U59B* | U59B | snoRNA | hMSC_Adipocyte | 6.14 |
| OSA | EUR | rs2926743 | rs2958153 | *ATP5B* | ENSG00000110955.4 | protein_coding | NaiveT | 6.07 |
| OSA | EUR | rs2926743 | rs2958153 | *SNORD59A* | ENSG00000207031.1 | snoRNA | NaiveT | 6.07 |
| OSA | EUR | rs2926743 | rs2958153 | *U59B* | U59B | snoRNA | NaiveT | 6.07 |
| OSA | EUR | rs2926743 | rs941207 | *ATP5B* | ENSG00000110955.4 | protein_coding | hESC_HypothalamicNPC | 5.94 |
| OSA | EUR | rs2926743 | rs941207 | *SNORD59A* | ENSG00000207031.1 | snoRNA | hESC_HypothalamicNPC | 5.94 |
| OSA | EUR | rs2926743 | rs941207 | *U59B* | U59B | snoRNA | hESC_HypothalamicNPC | 5.94 |
| OSA | EUR | rs2926743 | rs2958153 | *BAZ2A* | ENSG00000076108.7 | protein_coding | iPSC_Neurons | 5.94 |
| OSA | EUR | rs2926743 | rs941207 | *ATP5B* | ENSG00000110955.4 | protein_coding | hMSC_Adipocyte | 5.89 |
| OSA | EUR | rs2926743 | rs941207 | *SNORD59A* | ENSG00000207031.1 | snoRNA | hMSC_Adipocyte | 5.89 |
| OSA | EUR | rs2926743 | rs941207 | *U59B* | U59B | snoRNA | hMSC_Adipocyte | 5.89 |
| OSA | EUR | rs2926743 | rs941207 | *NACA* | ENSG00000196531.6 | protein_coding | GCB | 5.87 |
| OSA | EUR | rs2926743 | rs2958153 | TCONS_00020445 | TCONS_00020445 | lincRNA | iPSC_NPC | 5.8 |
| OSA | EUR | rs2926743 | rs2958153 | TCONS_00020807 | TCONS_00020807 | lincRNA | iPSC_NPC | 5.8 |
| OSA | EUR | rs2926743 | rs941207 | *BAZ2A* | ENSG00000076108.7 | protein_coding | hESC_HypothalamicNPC | 5.79 |
| OSA | EUR | rs2926743 | rs941207 | *BAZ2A* | ENSG00000076108.7 | protein_coding | hMSC_Osteoblasts | 5.77 |
| OSA | EUR | rs2926743 | rs2958153 | *BAZ2A* | ENSG00000076108.7 | protein_coding | NaiveB | 5.7 |
| OSA | EUR | rs2926743 | rs2958153 | *BAZ2A* | ENSG00000076108.7 | protein_coding | SGBS_Diff | 5.57 |
| OSA | EUR | rs2926743 | rs941207 | *NACA* | ENSG00000196531.6 | protein_coding | hESC_HypothalamicNeuron | 5.55 |
| OSA | EUR | rs2926743 | rs2958153 | *ESYT1* | ENSG00000139641.8 | protein_coding | iPSC_Neurons | 5.51 |
| OSA | EUR | rs2926743 | rs2958153 | *ZC3H10* | ENSG00000135482.2 | protein_coding | iPSC_Neurons | 5.51 |
| OSA | EUR | rs2926743 | rs941207 | *BAZ2A* | ENSG00000076108.7 | protein_coding | iPSC_Neurons | 5.39 |
| OSA | EUR | rs2926743 | rs2958153 | *ZBTB39* | ENSG00000166860.2 | protein_coding | iPSC_NPC | 5.39 |
| OSA | EUR | rs2926743 | rs2958153 | *ATP5B* | ENSG00000110955.4 | protein_coding | TFH | 5.39 |
| OSA | EUR | rs2926743 | rs2958153 | *SNORD59A* | ENSG00000207031.1 | snoRNA | TFH | 5.39 |
| OSA | EUR | rs2926743 | rs941207 | *RBMS2* | ENSG00000076067.7 | protein_coding | NaiveT | 5.38 |
| OSA | EUR | rs2926743 | rs941207 | *U59B* | U59B | snoRNA | NaiveB | 5.28 |
| OSA | EUR | rs2926743 | rs2958153 | *ATP5B* | ENSG00000110955.4 | protein_coding | iPSC_Neurons | 5.27 |
| OSA | EUR | rs2926743 | rs2958153 | *SNORD59A* | ENSG00000207031.1 | snoRNA | iPSC_Neurons | 5.27 |
| OSA | EUR | rs2926743 | rs2958153 | *U59B* | U59B | snoRNA | iPSC_Neurons | 5.27 |
| OSA | EUR | rs2926743 | rs941207 | *APOF* | ENSG00000175336.8 | protein_coding | iPSC_NPC | 5.25 |
| OSA | EUR | rs2926743 | rs941207 | *NACA* | ENSG00000196531.6 | protein_coding | hMSC_Osteoblasts | 5.18 |
| OSA | EUR | rs2926743 | rs941207 | *ATP5B* | ENSG00000110955.4 | protein_coding | iPSC_NPC | 5.12 |
| OSA | EUR | rs2926743 | rs941207 | *SNORD59A* | ENSG00000207031.1 | snoRNA | iPSC_NPC | 5.12 |
| OSA | EUR | rs2926743 | rs941207 | *U59B* | U59B | snoRNA | iPSC_NPC | 5.12 |
| OSA | EUR | rs2926743 | rs941207 | *MYL6* | ENSG00000092841.14 | protein_coding | iPSC_NPC | 5.03 |
| OSA | EUR | rs2926743 | rs941207 | *MYL6B* | ENSG00000196465.6 | protein_coding | iPSC_NPC | 5.03 |
| OSA | EUR | rs2926743 | rs941207 | RP11-603J24.14 | ENSG00000257809.1 | antisense | iPSC_NPC | 5.03 |
| OSA | EUR | rs2926743 | rs2958153 | *ATP5B* | ENSG00000110955.4 | protein_coding | HEPG2 | 5.01 |
| OSA | EUR | rs2926743 | rs2958153 | *SNORD59A* | ENSG00000207031.1 | snoRNA | HEPG2 | 5.01 |
| OSA | EUR | rs2926743 | rs2958153 | *U59B* | U59B | snoRNA | HEPG2 | 5.01 |
| OSA | EUR | rs56094641 | rs9939973 | *CRNDE* | ENSG00000245694.9 | lincRNA | Chondrocytes | 11.33 |
| OSA | EUR | rs56094641 | rs9940646 | *CRNDE* | ENSG00000245694.9 | lincRNA | Chondrocytes | 11.33 |
| OSA | EUR | rs56094641 | rs9940128 | *CRNDE* | ENSG00000245694.9 | lincRNA | Chondrocytes | 11.33 |
| OSA | EUR | rs56094641 | rs1421085 | *CRNDE* | ENSG00000245694.9 | lincRNA | Chondrocytes | 11.33 |
| OSA | EUR | rs56094641 | rs9939973 | *IRX5* | ENSG00000176842.14 | protein_coding | Chondrocytes | 9.88 |
| OSA | EUR | rs56094641 | rs9940646 | *IRX5* | ENSG00000176842.14 | protein_coding | Chondrocytes | 9.88 |
| OSA | EUR | rs56094641 | rs9940128 | *IRX5* | ENSG00000176842.14 | protein_coding | Chondrocytes | 9.88 |
| OSA | EUR | rs56094641 | rs1421085 | *IRX5* | ENSG00000176842.14 | protein_coding | Chondrocytes | 9.88 |
| OSA | EUR | rs56094641 | rs11642015 | *AC106738.2* | ENSG00000259725.1 | lincRNA | Chondrocytes | 8.5 |
| OSA | EUR | rs56094641 | rs11642015 | *CRNDE* | ENSG00000245694.9 | lincRNA | Chondrocytes | 6.16 |
| OSA | EUR | rs2926743 | rs2256341 | *HSPD1P4* | ENSG00000257576.1 | processed_pseudogene | Chondrocytes | 14.94 |
| OSA | EUR | rs2926743 | rs2256341 | *RBMS2* | ENSG00000076067.13 | protein_coding | Chondrocytes | 11.09 |
| OSA | EUR | rs2926743 | rs2256341 | *RNU6-343P* | ENSG00000201579.1 | snRNA | Chondrocytes | 7.95 |
| OSA | EUR | rs2926743 | rs2256341 | *GLS2* | ENSG00000135423.13 | protein_coding | Chondrocytes | 6.47 |
| OSA | EUR | rs2926743 | rs2256341 | AC097104.1 | ENSG00000285528.1 | protein_coding | Chondrocytes | 6.47 |
| OSA | MVP (Sofer et al) | rs10150332 | rs17109256 | NRXN3 | ENSG00000021645.13 | protein_coding | hESC_HypothalamicNeuron | 5.15 |
| OSA | MVP (Sofer et al) | rs10874656 | rs11164605 | *GFI1* | ENSG00000162676.7 | protein_coding | hESC_HypothalamicNPC | 8.13 |
| OSA | MVP (Sofer et al) | rs10874656 | rs6603975 | *GLMN* | ENSG00000174842.12 | protein_coding | GCB | 5.19 |
| OSA | MVP (Sofer et al) | rs10874656 | rs6603975 | *RPAP2* | ENSG00000122484.8 | protein_coding | GCB | 5.19 |
| OSA | MVP (Sofer et al) | rs12514158 | chr5:96537558:GA:G | *CAST* | ENSG00000153113.19 | protein_coding | hESC_HypothalamicNeuron | 7.15 |
| OSA | MVP (Sofer et al) | rs13043844 | rs71828295 | RP4-715N11.2 | ENSG00000232294.1 | lincRNA | SGBS_Diff | 7.12 |
| OSA | MVP (Sofer et al) | rs13043844 | rs71828295 | TCONS_00028706 | TCONS_00028706 | lincRNA | hMSC_Adipocyte | 5.71 |
| OSA | MVP (Sofer et al) | rs13043844 | rs71828295 | TCONS_00028707 | TCONS_00028707 | lincRNA | hMSC_Adipocyte | 5.71 |
| OSA | MVP (Sofer et al) | rs13043844 | rs71828295 | TCONS_00028708 | TCONS_00028708 | lincRNA | hMSC_Adipocyte | 5.71 |
| OSA | MVP (Sofer et al) | rs13043844 | rs71828295 | TCONS_00028709 | TCONS_00028709 | lincRNA | hMSC_Adipocyte | 5.71 |
| OSA | MVP (Sofer et al) | rs199445 | rs77768866 | *RF00019* | ENSG00000238723.1 | misc_RNA | Chondrocytes | 49.39 |
| OSA | MVP (Sofer et al) | rs199445 | rs17573447 | *RF00019* | ENSG00000238723.1 | misc_RNA | Chondrocytes | 48.27 |
| OSA | MVP (Sofer et al) | rs199445 | rs62064662 | *RF00019* | ENSG00000238723.1 | misc_RNA | Chondrocytes | 48.27 |
| OSA | MVP (Sofer et al) | rs199445 | rs62064663 | *RF00019* | ENSG00000238723.1 | misc_RNA | Chondrocytes | 48.27 |
| OSA | MVP (Sofer et al) | rs199445 | rs4283261 | *RF00019* | ENSG00000238723.1 | misc_RNA | Chondrocytes | 45.94 |
| OSA | MVP (Sofer et al) | rs199445 | rs62062265 | *RF00019* | ENSG00000238723.1 | misc_RNA | Chondrocytes | 45.94 |
| OSA | MVP (Sofer et al) | rs199445 | rs733966 | *RF00019* | ENSG00000238723.1 | misc_RNA | Chondrocytes | 45.94 |
| OSA | MVP (Sofer et al) | rs199445 | rs733967 | *RF00019* | ENSG00000238723.1 | misc_RNA | Chondrocytes | 45.94 |
| OSA | MVP (Sofer et al) | rs199445 | rs733968 | *RF00019* | ENSG00000238723.1 | misc_RNA | Chondrocytes | 45.94 |
| OSA | MVP (Sofer et al) | rs199445 | rs733969 | *RF00019* | ENSG00000238723.1 | misc_RNA | Chondrocytes | 45.94 |
| OSA | MVP (Sofer et al) | rs199445 | rs1052553 | *RF00019* | ENSG00000238723.1 | misc_RNA | Chondrocytes | 41.59 |
| OSA | MVP (Sofer et al) | rs199445 | rs17652121 | *RF00019* | ENSG00000238723.1 | misc_RNA | Chondrocytes | 41.59 |
| OSA | MVP (Sofer et al) | rs199445 | rs55881134 | *RF00019* | ENSG00000238723.1 | misc_RNA | Chondrocytes | 36.02 |
| OSA | MVP (Sofer et al) | rs199445 | rs77768866 | *KANSL1-AS1* | ENSG00000214401.4 | antisense | Chondrocytes | 34.31 |
| OSA | MVP (Sofer et al) | rs199445 | rs77768866 | *KANSL1* | ENSG00000120071.14 | protein_coding | Chondrocytes | 34.31 |
| OSA | MVP (Sofer et al) | rs199445 | rs113417378 | *KANSL1* | ENSG00000120071.14 | protein_coding | Chondrocytes | 34.31 |
| OSA | MVP (Sofer et al) | rs199445 | rs2532236 | *KANSL1* | ENSG00000120071.14 | protein_coding | Chondrocytes | 34.31 |
| OSA | MVP (Sofer et al) | rs199445 | rs2732658 | *KANSL1* | ENSG00000120071.14 | protein_coding | Chondrocytes | 34.31 |
| OSA | MVP (Sofer et al) | rs199445 | rs56266533 | *KANSL1* | ENSG00000120071.14 | protein_coding | Chondrocytes | 34.31 |

| **Supplementary Table 16.** Prioritized effector genes from variant to gene (V2G) mapping analysis. | | | | | | | | |
|---|---|---|---|---|---|---|---|---|
| **Trait** | **Discovery Study** | **Sentinel rsID** | **Proxy rsID** | **Gene Symbol** | **Gene Id** | **Gene Biotype** | **Cell** | **Contact Score (Chicago Score\|-log10(FitHiC2 FDR)** |
| OSA | MVP (Sofer et al) | rs199445 | rs62064665 | *RF00019* | ENSG00000238723.1 | misc_RNA | Chondrocytes | 29.78 |
| OSA | MVP (Sofer et al) | rs199445 | rs55881134 | *KANSL1-AS1* | ENSG00000214401.4 | antisense | Chondrocytes | 24.58 |
| OSA | MVP (Sofer et al) | rs199445 | rs1052594 | *RF00019* | ENSG00000238723.1 | misc_RNA | Chondrocytes | 23.79 |
| OSA | MVP (Sofer et al) | rs199445 | rs16940799 | *RF00019* | ENSG00000238723.1 | misc_RNA | Chondrocytes | 23.79 |
| OSA | MVP (Sofer et al) | rs199445 | rs17574040 | *RF00019* | ENSG00000238723.1 | misc_RNA | Chondrocytes | 23.79 |
| OSA | MVP (Sofer et al) | rs199445 | rs66561280 | *RF00019* | ENSG00000238723.1 | misc_RNA | Chondrocytes | 23.79 |
| OSA | MVP (Sofer et al) | rs199445 | rs62062282 | *RF00019* | ENSG00000238723.1 | misc_RNA | Chondrocytes | 23.45 |
| OSA | MVP (Sofer et al) | rs199445 | rs4283261 | *AC005829.1* | ENSG00000261575.2 | transcribed_unprocessed_pseudogene | Chondrocytes | 21.99 |
| OSA | MVP (Sofer et al) | rs199445 | rs62062265 | *AC005829.1* | ENSG00000261575.2 | transcribed_unprocessed_pseudogene | Chondrocytes | 21.99 |
| OSA | MVP (Sofer et al) | rs199445 | rs733966 | *AC005829.1* | ENSG00000261575.2 | transcribed_unprocessed_pseudogene | Chondrocytes | 21.99 |
| OSA | MVP (Sofer et al) | rs199445 | rs733967 | *AC005829.1* | ENSG00000261575.2 | transcribed_unprocessed_pseudogene | Chondrocytes | 21.99 |
| OSA | MVP (Sofer et al) | rs199445 | rs733968 | *AC005829.1* | ENSG00000261575.2 | transcribed_unprocessed_pseudogene | Chondrocytes | 21.99 |
| OSA | MVP (Sofer et al) | rs199445 | rs733969 | *AC005829.1* | ENSG00000261575.2 | transcribed_unprocessed_pseudogene | Chondrocytes | 21.99 |
| OSA | MVP (Sofer et al) | rs199445 | rs55881134 | *KANSL1* | ENSG00000120071.14 | protein_coding | Chondrocytes | 21.4 |
| OSA | MVP (Sofer et al) | rs199445 | rs113417378 | *CR936218.1* | ENSG00000262372.1 | antisense | Chondrocytes | 21.4 |
| OSA | MVP (Sofer et al) | rs199445 | rs2532236 | *CR936218.1* | ENSG00000262372.1 | antisense | Chondrocytes | 21.4 |
| OSA | MVP (Sofer et al) | rs199445 | rs2732658 | *CR936218.1* | ENSG00000262372.1 | antisense | Chondrocytes | 21.4 |
| OSA | MVP (Sofer et al) | rs199445 | rs56266533 | *CR936218.1* | ENSG00000262372.1 | antisense | Chondrocytes | 21.4 |
| OSA | MVP (Sofer et al) | rs199445 | rs17573447 | *LRRC37A* | ENSG00000176681.14 | protein_coding | Chondrocytes | 21.3 |
| OSA | MVP (Sofer et al) | rs199445 | rs62064662 | *LRRC37A* | ENSG00000176681.14 | protein_coding | Chondrocytes | 21.3 |
| OSA | MVP (Sofer et al) | rs199445 | rs62064663 | *LRRC37A* | ENSG00000176681.14 | protein_coding | Chondrocytes | 21.3 |
| OSA | MVP (Sofer et al) | rs199445 | rs17573447 | *AC005829.2* | ENSG00000262539.1 | processed_pseudogene | Chondrocytes | 20.41 |
| OSA | MVP (Sofer et al) | rs199445 | rs62064662 | *AC005829.2* | ENSG00000262539.1 | processed_pseudogene | Chondrocytes | 20.41 |
| OSA | MVP (Sofer et al) | rs199445 | rs62064663 | *AC005829.2* | ENSG00000262539.1 | processed_pseudogene | Chondrocytes | 20.41 |
| OSA | MVP (Sofer et al) | rs199445 | rs1052553 | *LRRC37A* | ENSG00000176681.14 | protein_coding | Chondrocytes | 20.2 |
| OSA | MVP (Sofer et al) | rs199445 | rs17652121 | *LRRC37A* | ENSG00000176681.14 | protein_coding | Chondrocytes | 20.2 |
| OSA | MVP (Sofer et al) | rs199445 | rs62064665 | *LRRC37A* | ENSG00000176681.14 | protein_coding | Chondrocytes | 18.03 |
| OSA | MVP (Sofer et al) | rs199445 | rs4283261 | *LRRC37A* | ENSG00000176681.14 | protein_coding | Chondrocytes | 16.96 |
| OSA | MVP (Sofer et al) | rs199445 | rs62062265 | *LRRC37A* | ENSG00000176681.14 | protein_coding | Chondrocytes | 16.96 |
| OSA | MVP (Sofer et al) | rs199445 | rs733966 | *LRRC37A* | ENSG00000176681.14 | protein_coding | Chondrocytes | 16.96 |
| OSA | MVP (Sofer et al) | rs199445 | rs733967 | *LRRC37A* | ENSG00000176681.14 | protein_coding | Chondrocytes | 16.96 |
| OSA | MVP (Sofer et al) | rs199445 | rs733968 | *LRRC37A* | ENSG00000176681.14 | protein_coding | Chondrocytes | 16.96 |
| OSA | MVP (Sofer et al) | rs199445 | rs733969 | *LRRC37A* | ENSG00000176681.14 | protein_coding | Chondrocytes | 16.96 |
| OSA | MVP (Sofer et al) | rs199445 | rs17573447 | *AC005829.1* | ENSG00000261575.2 | transcribed_unprocessed_pseudogene | Chondrocytes | 16.37 |
| OSA | MVP (Sofer et al) | rs199445 | rs62064662 | *AC005829.1* | ENSG00000261575.2 | transcribed_unprocessed_pseudogene | Chondrocytes | 16.37 |
| OSA | MVP (Sofer et al) | rs199445 | rs62064663 | *AC005829.1* | ENSG00000261575.2 | transcribed_unprocessed_pseudogene | Chondrocytes | 16.37 |
| OSA | MVP (Sofer et al) | rs199445 | rs4283261 | *AC005829.2* | ENSG00000262539.1 | processed_pseudogene | Chondrocytes | 15.97 |
| OSA | MVP (Sofer et al) | rs199445 | rs62062265 | *AC005829.2* | ENSG00000262539.1 | processed_pseudogene | Chondrocytes | 15.97 |
| OSA | MVP (Sofer et al) | rs199445 | rs733966 | *AC005829.2* | ENSG00000262539.1 | processed_pseudogene | Chondrocytes | 15.97 |
| OSA | MVP (Sofer et al) | rs199445 | rs733967 | *AC005829.2* | ENSG00000262539.1 | processed_pseudogene | Chondrocytes | 15.97 |
| OSA | MVP (Sofer et al) | rs199445 | rs733968 | *AC005829.2* | ENSG00000262539.1 | processed_pseudogene | Chondrocytes | 15.97 |
| OSA | MVP (Sofer et al) | rs199445 | rs733969 | *AC005829.2* | ENSG00000262539.1 | processed_pseudogene | Chondrocytes | 15.97 |
| OSA | MVP (Sofer et al) | rs199445 | rs17651093 | *MAPT* | ENSG00000186868.15 | protein_coding | Chondrocytes | 15.91 |
| OSA | MVP (Sofer et al) | rs199445 | rs17651243 | *MAPT* | ENSG00000186868.15 | protein_coding | Chondrocytes | 15.91 |
| OSA | MVP (Sofer et al) | rs199445 | rs75242405 | *MAPT* | ENSG00000186868.15 | protein_coding | Chondrocytes | 15.91 |
| OSA | MVP (Sofer et al) | rs199445 | rs62063786 | *MAPT* | ENSG00000186868.15 | protein_coding | Chondrocytes | 15.91 |
| OSA | MVP (Sofer et al) | rs199445 | rs63750417 | *MAPT* | ENSG00000186868.15 | protein_coding | Chondrocytes | 15.91 |
| OSA | MVP (Sofer et al) | rs199445 | rs4283261 | *KANSL1-AS1* | ENSG00000214401.4 | antisense | Chondrocytes | 15.81 |
| OSA | MVP (Sofer et al) | rs199445 | rs62062265 | *KANSL1-AS1* | ENSG00000214401.4 | antisense | Chondrocytes | 15.81 |
| OSA | MVP (Sofer et al) | rs199445 | rs733966 | *KANSL1-AS1* | ENSG00000214401.4 | antisense | Chondrocytes | 15.81 |
| OSA | MVP (Sofer et al) | rs199445 | rs733967 | *KANSL1-AS1* | ENSG00000214401.4 | antisense | Chondrocytes | 15.81 |
| OSA | MVP (Sofer et al) | rs199445 | rs733968 | *KANSL1-AS1* | ENSG00000214401.4 | antisense | Chondrocytes | 15.81 |
| OSA | MVP (Sofer et al) | rs199445 | rs733969 | *KANSL1-AS1* | ENSG00000214401.4 | antisense | Chondrocytes | 15.81 |
| OSA | MVP (Sofer et al) | rs199445 | rs55874169 | *RF00017* | ENSG00000274883.1 | misc_RNA | Chondrocytes | 15.61 |
| OSA | MVP (Sofer et al) | rs199445 | rs4283261 | *KANSL1* | ENSG00000120071.14 | protein_coding | Chondrocytes | 15.07 |
| OSA | MVP (Sofer et al) | rs199445 | rs62062265 | *KANSL1* | ENSG00000120071.14 | protein_coding | Chondrocytes | 15.07 |
| OSA | MVP (Sofer et al) | rs199445 | rs733966 | *KANSL1* | ENSG00000120071.14 | protein_coding | Chondrocytes | 15.07 |
| OSA | MVP (Sofer et al) | rs199445 | rs733967 | *KANSL1* | ENSG00000120071.14 | protein_coding | Chondrocytes | 15.07 |
| OSA | MVP (Sofer et al) | rs199445 | rs733968 | *KANSL1* | ENSG00000120071.14 | protein_coding | Chondrocytes | 15.07 |
| OSA | MVP (Sofer et al) | rs199445 | rs733969 | *KANSL1* | ENSG00000120071.14 | protein_coding | Chondrocytes | 15.07 |
| OSA | MVP (Sofer et al) | rs199445 | rs1052594 | *AC005829.1* | ENSG00000261575.2 | transcribed_unprocessed_pseudogene | Chondrocytes | 13.64 |
| OSA | MVP (Sofer et al) | rs199445 | rs16940799 | *AC005829.1* | ENSG00000261575.2 | transcribed_unprocessed_pseudogene | Chondrocytes | 13.64 |
| OSA | MVP (Sofer et al) | rs199445 | rs17574040 | *AC005829.1* | ENSG00000261575.2 | transcribed_unprocessed_pseudogene | Chondrocytes | 13.64 |
| OSA | MVP (Sofer et al) | rs199445 | rs66561280 | *AC005829.1* | ENSG00000261575.2 | transcribed_unprocessed_pseudogene | Chondrocytes | 13.64 |
| OSA | MVP (Sofer et al) | rs199445 | rs62064665 | *KANSL1-AS1* | ENSG00000214401.4 | antisense | Chondrocytes | 13.4 |
| OSA | MVP (Sofer et al) | rs199445 | rs113271363 | *CR936218.2* | ENSG00000262881.1 | antisense | Chondrocytes | 13.37 |
| OSA | MVP (Sofer et al) | rs199445 | rs55760800 | *CR936218.2* | ENSG00000262881.1 | antisense | Chondrocytes | 13.37 |
| OSA | MVP (Sofer et al) | rs199445 | rs56303672 | *CR936218.2* | ENSG00000262881.1 | antisense | Chondrocytes | 13.37 |
| OSA | MVP (Sofer et al) | rs199445 | rs111413387 | *DND1P1* | ENSG00000264070.1 | processed_pseudogene | Chondrocytes | 13.16 |
| OSA | MVP (Sofer et al) | rs199445 | rs112534607 | *DND1P1* | ENSG00000264070.1 | processed_pseudogene | Chondrocytes | 13.16 |
| OSA | MVP (Sofer et al) | rs199445 | rs2532343 | *DND1P1* | ENSG00000264070.1 | processed_pseudogene | Chondrocytes | 13.16 |
| OSA | MVP (Sofer et al) | rs199445 | rs2696690 | *DND1P1* | ENSG00000264070.1 | processed_pseudogene | Chondrocytes | 13.16 |
| OSA | MVP (Sofer et al) | rs199445 | rs2696692 | *DND1P1* | ENSG00000264070.1 | processed_pseudogene | Chondrocytes | 13.16 |
| OSA | MVP (Sofer et al) | rs199445 | rs2732649 | *DND1P1* | ENSG00000264070.1 | processed_pseudogene | Chondrocytes | 13.16 |
| OSA | MVP (Sofer et al) | rs199445 | rs62071575 | *DND1P1* | ENSG00000264070.1 | processed_pseudogene | Chondrocytes | 13.16 |
| OSA | MVP (Sofer et al) | rs199445 | rs111413387 | *AC126544.2* | ENSG00000285668.1 | lincRNA | Chondrocytes | 13.16 |
| OSA | MVP (Sofer et al) | rs199445 | rs112534607 | *AC126544.2* | ENSG00000285668.1 | lincRNA | Chondrocytes | 13.16 |
| OSA | MVP (Sofer et al) | rs199445 | rs2532343 | *AC126544.2* | ENSG00000285668.1 | lincRNA | Chondrocytes | 13.16 |
| OSA | MVP (Sofer et al) | rs199445 | rs2696690 | *AC126544.2* | ENSG00000285668.1 | lincRNA | Chondrocytes | 13.16 |
| OSA | MVP (Sofer et al) | rs199445 | rs2696692 | *AC126544.2* | ENSG00000285668.1 | lincRNA | Chondrocytes | 13.16 |
| OSA | MVP (Sofer et al) | rs199445 | rs2732649 | *AC126544.2* | ENSG00000285668.1 | lincRNA | Chondrocytes | 13.16 |
| OSA | MVP (Sofer et al) | rs199445 | rs62071575 | *AC126544.2* | ENSG00000285668.1 | lincRNA | Chondrocytes | 13.16 |
| OSA | MVP (Sofer et al) | rs199445 | rs62064665 | *AC005829.1* | ENSG00000261575.2 | transcribed_unprocessed_pseudogene | Chondrocytes | 12.89 |
| OSA | MVP (Sofer et al) | rs199445 | rs17573447 | *KANSL1-AS1* | ENSG00000214401.4 | antisense | Chondrocytes | 12.67 |
| OSA | MVP (Sofer et al) | rs199445 | rs62064662 | *KANSL1-AS1* | ENSG00000214401.4 | antisense | Chondrocytes | 12.67 |

| **Supplementary Table 16.** Prioritized effector genes from variant to gene (V2G) mapping analysis. | | | | | | | | |
|---|---|---|---|---|---|---|---|---|
| **Trait** | **Discovery Study** | **Sentinel rsID** | **Proxy rsID** | **Gene Symbol** | **Gene Id** | **Gene Biotype** | **Cell** | **Contact Score (Chicago Score\|-log10(FitHiC2 FDR)** |
| OSA | MVP (Sofer et al) | rs199445 | rs62064663 | *KANSL1-AS1* | ENSG00000214401.4 | antisense | Chondrocytes | 12.67 |
| OSA | MVP (Sofer et al) | rs199445 | rs1052553 | *AC005829.1* | ENSG00000261575.2 | transcribed_unprocessed_pseudogene | Chondrocytes | 12.5 |
| OSA | MVP (Sofer et al) | rs199445 | rs17652121 | *AC005829.1* | ENSG00000261575.2 | transcribed_unprocessed_pseudogene | Chondrocytes | 12.5 |
| OSA | MVP (Sofer et al) | rs199445 | rs111413387 | *STH* | ENSG00000256762.1 | protein_coding | Chondrocytes | 12.5 |
| OSA | MVP (Sofer et al) | rs199445 | rs112534607 | *STH* | ENSG00000256762.1 | protein_coding | Chondrocytes | 12.5 |
| OSA | MVP (Sofer et al) | rs199445 | rs2532343 | *STH* | ENSG00000256762.1 | protein_coding | Chondrocytes | 12.5 |
| OSA | MVP (Sofer et al) | rs199445 | rs2696690 | *STH* | ENSG00000256762.1 | protein_coding | Chondrocytes | 12.5 |
| OSA | MVP (Sofer et al) | rs199445 | rs2696692 | *STH* | ENSG00000256762.1 | protein_coding | Chondrocytes | 12.5 |
| OSA | MVP (Sofer et al) | rs199445 | rs2732649 | *STH* | ENSG00000256762.1 | protein_coding | Chondrocytes | 12.5 |
| OSA | MVP (Sofer et al) | rs199445 | rs62071575 | *STH* | ENSG00000256762.1 | protein_coding | Chondrocytes | 12.5 |
| OSA | MVP (Sofer et al) | rs199445 | rs56398500 | *LINC02210* | ENSG00000204650.14 | transcribed_unitary_pseudogene | Chondrocytes | 12.48 |
| OSA | MVP (Sofer et al) | rs199445 | rs1052594 | *KANSL1-AS1* | ENSG00000214401.4 | antisense | Chondrocytes | 12.06 |
| OSA | MVP (Sofer et al) | rs199445 | rs16940799 | *KANSL1-AS1* | ENSG00000214401.4 | antisense | Chondrocytes | 12.06 |
| OSA | MVP (Sofer et al) | rs199445 | rs17574040 | *KANSL1-AS1* | ENSG00000214401.4 | antisense | Chondrocytes | 12.06 |
| OSA | MVP (Sofer et al) | rs199445 | rs66561280 | *KANSL1-AS1* | ENSG00000214401.4 | antisense | Chondrocytes | 12.06 |
| OSA | MVP (Sofer et al) | rs199445 | rs1052553 | *KANSL1-AS1* | ENSG00000214401.4 | antisense | Chondrocytes | 11.9 |
| OSA | MVP (Sofer et al) | rs199445 | rs17652121 | *KANSL1-AS1* | ENSG00000214401.4 | antisense | Chondrocytes | 11.9 |
| OSA | MVP (Sofer et al) | rs199445 | rs1052553 | *AC005829.2* | ENSG00000262539.1 | processed_pseudogene | Chondrocytes | 11.68 |
| OSA | MVP (Sofer et al) | rs199445 | rs17652121 | *AC005829.2* | ENSG00000262539.1 | processed_pseudogene | Chondrocytes | 11.68 |
| OSA | MVP (Sofer et al) | rs199445 | rs62062282 | *LRRC37A* | ENSG00000176681.14 | protein_coding | Chondrocytes | 11.4 |
| OSA | MVP (Sofer et al) | rs199445 | rs1052594 | *AC005829.2* | ENSG00000262539.1 | processed_pseudogene | Chondrocytes | 10.81 |
| OSA | MVP (Sofer et al) | rs199445 | rs16940799 | *AC005829.2* | ENSG00000262539.1 | processed_pseudogene | Chondrocytes | 10.81 |
| OSA | MVP (Sofer et al) | rs199445 | rs17574040 | *AC005829.2* | ENSG00000262539.1 | processed_pseudogene | Chondrocytes | 10.81 |
| OSA | MVP (Sofer et al) | rs199445 | rs66561280 | *AC005829.2* | ENSG00000262539.1 | processed_pseudogene | Chondrocytes | 10.81 |
| OSA | MVP (Sofer et al) | rs199445 | rs62062770 | *RF00017* | ENSG00000274883.1 | misc_RNA | Chondrocytes | 10.78 |
| OSA | MVP (Sofer et al) | rs199445 | rs55881134 | *LRRC37A* | ENSG00000176681.14 | protein_coding | Chondrocytes | 10.71 |
| OSA | MVP (Sofer et al) | rs199445 | rs62064665 | *AC005829.2* | ENSG00000262539.1 | processed_pseudogene | Chondrocytes | 10.48 |
| OSA | MVP (Sofer et al) | rs199445 | rs62062282 | *AC005829.1* | ENSG00000261575.2 | transcribed_unprocessed_pseudogene | Chondrocytes | 10.29 |
| OSA | MVP (Sofer et al) | rs199445 | rs17571739 | *MAPT* | ENSG00000186868.15 | protein_coding | Chondrocytes | 9.54 |
| OSA | MVP (Sofer et al) | rs199445 | rs78555354 | *MAPT* | ENSG00000186868.15 | protein_coding | Chondrocytes | 9.54 |
| OSA | MVP (Sofer et al) | rs199445 | rs111413387 | *KANSL1* | ENSG00000120071.14 | protein_coding | Chondrocytes | 9.53 |
| OSA | MVP (Sofer et al) | rs199445 | rs112534607 | *KANSL1* | ENSG00000120071.14 | protein_coding | Chondrocytes | 9.53 |
| OSA | MVP (Sofer et al) | rs199445 | rs2532343 | *KANSL1* | ENSG00000120071.14 | protein_coding | Chondrocytes | 9.53 |
| OSA | MVP (Sofer et al) | rs199445 | rs2696690 | *KANSL1* | ENSG00000120071.14 | protein_coding | Chondrocytes | 9.53 |
| OSA | MVP (Sofer et al) | rs199445 | rs2696692 | *KANSL1* | ENSG00000120071.14 | protein_coding | Chondrocytes | 9.53 |
| OSA | MVP (Sofer et al) | rs199445 | rs2732649 | *KANSL1* | ENSG00000120071.14 | protein_coding | Chondrocytes | 9.53 |
| OSA | MVP (Sofer et al) | rs199445 | rs62071575 | *KANSL1* | ENSG00000120071.14 | protein_coding | Chondrocytes | 9.53 |
| OSA | MVP (Sofer et al) | rs199445 | rs1052594 | *KANSL1* | ENSG00000120071.14 | protein_coding | Chondrocytes | 9.22 |
| OSA | MVP (Sofer et al) | rs199445 | rs16940799 | *KANSL1* | ENSG00000120071.14 | protein_coding | Chondrocytes | 9.22 |
| OSA | MVP (Sofer et al) | rs199445 | rs17574040 | *KANSL1* | ENSG00000120071.14 | protein_coding | Chondrocytes | 9.22 |
| OSA | MVP (Sofer et al) | rs199445 | rs66561280 | *KANSL1* | ENSG00000120071.14 | protein_coding | Chondrocytes | 9.22 |
| OSA | MVP (Sofer et al) | rs199445 | rs76594404 | *MAPT-AS1* | ENSG00000264589.1 | antisense | HEPG2 | 8.5 |
| OSA | MVP (Sofer et al) | rs199445 | rs76594404 | *SPPL2C* | ENSG00000185294.5 | protein_coding | HEPG2 | 8.5 |
| OSA | MVP (Sofer et al) | rs199445 | rs55881134 | *AC005829.2* | ENSG00000262539.1 | processed_pseudogene | Chondrocytes | 8.45 |
| OSA | MVP (Sofer et al) | rs199445 | rs1052594 | *LRRC37A* | ENSG00000176681.14 | protein_coding | Chondrocytes | 8.31 |
| OSA | MVP (Sofer et al) | rs199445 | rs16940799 | *LRRC37A* | ENSG00000176681.14 | protein_coding | Chondrocytes | 8.31 |
| OSA | MVP (Sofer et al) | rs199445 | rs17574040 | *LRRC37A* | ENSG00000176681.14 | protein_coding | Chondrocytes | 8.31 |
| OSA | MVP (Sofer et al) | rs199445 | rs17650901 | *MAPT* | ENSG00000186868.15 | protein_coding | Chondrocytes | 8.31 |
| OSA | MVP (Sofer et al) | rs199445 | rs55711941 | *MAPT* | ENSG00000186868.15 | protein_coding | Chondrocytes | 8.31 |
| OSA | MVP (Sofer et al) | rs199445 | rs55881134 | *AC005829.1* | ENSG00000261575.2 | transcribed_unprocessed_pseudogene | Chondrocytes | 8.31 |
| OSA | MVP (Sofer et al) | rs199445 | rs66561280 | *LRRC37A* | ENSG00000176681.14 | protein_coding | Chondrocytes | 8.31 |
| OSA | MVP (Sofer et al) | rs199445 | rs111413387 | *CR936218.1* | ENSG00000262372.1 | antisense | Chondrocytes | 8.31 |
| OSA | MVP (Sofer et al) | rs199445 | rs112534607 | *CR936218.1* | ENSG00000262372.1 | antisense | Chondrocytes | 8.31 |
| OSA | MVP (Sofer et al) | rs199445 | rs2532343 | *CR936218.1* | ENSG00000262372.1 | antisense | Chondrocytes | 8.31 |
| OSA | MVP (Sofer et al) | rs199445 | rs2696690 | *CR936218.1* | ENSG00000262372.1 | antisense | Chondrocytes | 8.31 |
| OSA | MVP (Sofer et al) | rs199445 | rs2696692 | *CR936218.1* | ENSG00000262372.1 | antisense | Chondrocytes | 8.31 |
| OSA | MVP (Sofer et al) | rs199445 | rs2732649 | *CR936218.1* | ENSG00000262372.1 | antisense | Chondrocytes | 8.31 |
| OSA | MVP (Sofer et al) | rs199445 | rs62071575 | *CR936218.1* | ENSG00000262372.1 | antisense | Chondrocytes | 8.31 |
| OSA | MVP (Sofer et al) | rs199445 | rs55975673 | *LINC02210* | ENSG00000204650.14 | transcribed_unitary_pseudogene | Chondrocytes | 8.11 |
| OSA | MVP (Sofer et al) | rs199445 | rs56343551 | *LINC02210* | ENSG00000204650.14 | transcribed_unitary_pseudogene | Chondrocytes | 8.11 |
| OSA | MVP (Sofer et al) | rs199445 | rs56387391 | *LINC02210* | ENSG00000204650.14 | transcribed_unitary_pseudogene | Chondrocytes | 8.11 |
| OSA | MVP (Sofer et al) | rs199445 | rs17573447 | *KANSL1* | ENSG00000120071.14 | protein_coding | Chondrocytes | 7.87 |
| OSA | MVP (Sofer et al) | rs199445 | rs62064662 | *KANSL1* | ENSG00000120071.14 | protein_coding | Chondrocytes | 7.87 |
| OSA | MVP (Sofer et al) | rs199445 | rs62064663 | *KANSL1* | ENSG00000120071.14 | protein_coding | Chondrocytes | 7.87 |
| OSA | MVP (Sofer et al) | rs199445 | rs62056808 | *CR936218.2* | ENSG00000262881.1 | antisense | Chondrocytes | 7.82 |
| OSA | MVP (Sofer et al) | rs199445 | rs62056809 | *CR936218.2* | ENSG00000262881.1 | antisense | Chondrocytes | 7.82 |
| OSA | MVP (Sofer et al) | rs199445 | rs62056810 | *CR936218.2* | ENSG00000262881.1 | antisense | Chondrocytes | 7.82 |
| OSA | MVP (Sofer et al) | rs199445 | rs62062282 | *KANSL1* | ENSG00000120071.14 | protein_coding | Chondrocytes | 7.76 |
| OSA | MVP (Sofer et al) | rs199445 | rs62062282 | *KANSL1-AS1* | ENSG00000214401.4 | antisense | Chondrocytes | 7.76 |
| OSA | MVP (Sofer et al) | rs199445 | rs111413387 | *MAPT* | ENSG00000186868.15 | protein_coding | Chondrocytes | 7.54 |
| OSA | MVP (Sofer et al) | rs199445 | rs112534607 | *MAPT* | ENSG00000186868.15 | protein_coding | Chondrocytes | 7.54 |
| OSA | MVP (Sofer et al) | rs199445 | rs2532343 | *MAPT* | ENSG00000186868.15 | protein_coding | Chondrocytes | 7.54 |
| OSA | MVP (Sofer et al) | rs199445 | rs2696690 | *MAPT* | ENSG00000186868.15 | protein_coding | Chondrocytes | 7.54 |
| OSA | MVP (Sofer et al) | rs199445 | rs2696692 | *MAPT* | ENSG00000186868.15 | protein_coding | Chondrocytes | 7.54 |
| OSA | MVP (Sofer et al) | rs199445 | rs2732649 | *MAPT* | ENSG00000186868.15 | protein_coding | Chondrocytes | 7.54 |
| OSA | MVP (Sofer et al) | rs199445 | rs62071575 | *MAPT* | ENSG00000186868.15 | protein_coding | Chondrocytes | 7.54 |
| OSA | MVP (Sofer et al) | rs199445 | rs55975673 | *PLEKHM1* | ENSG00000225190.11 | protein_coding | Chondrocytes | 7.46 |
| OSA | MVP (Sofer et al) | rs199445 | rs56343551 | *PLEKHM1* | ENSG00000225190.11 | protein_coding | Chondrocytes | 7.46 |
| OSA | MVP (Sofer et al) | rs199445 | rs56387391 | *PLEKHM1* | ENSG00000225190.11 | protein_coding | Chondrocytes | 7.46 |
| OSA | MVP (Sofer et al) | rs199445 | rs78556223 | *MAPT* | ENSG00000186868.11 | protein_coding | hESC_HypothalamicNeuron | 7.31 |
| OSA | MVP (Sofer et al) | rs199445 | rs17651093 | *AC005829.1* | ENSG00000261575.2 | transcribed_unprocessed_pseudogene | Chondrocytes | 7.17 |
| OSA | MVP (Sofer et al) | rs199445 | rs75242405 | *AC005829.1* | ENSG00000261575.2 | transcribed_unprocessed_pseudogene | Chondrocytes | 7.17 |
| OSA | MVP (Sofer et al) | rs199445 | rs77768866 | *LRRC37A* | ENSG00000176681.14 | protein_coding | Chondrocytes | 7.13 |
| OSA | MVP (Sofer et al) | rs199445 | rs77768866 | *AC005829.2* | ENSG00000262539.1 | processed_pseudogene | Chondrocytes | 6.98 |
| OSA | MVP (Sofer et al) | rs199445 | rs17649700 | *CR936218.2* | ENSG00000262881.1 | antisense | Chondrocytes | 6.8 |

**Supplementary Table 16.** Prioritized effector genes from variant to gene (V2G) mapping analysis.

| Trait | Discovery Study | Sentinel rsID | Proxy rsID | Gene Symbol | Gene Id | Gene Biotype | Cell | Contact Score (Chicago Score\|-log10(FitHiC2 FDR) |
|---|---|---|---|---|---|---|---|---|
| OSA | MVP (Sofer et al) | rs199445 | rs56002706 | *CR936218.2* | ENSG00000262881.1 | antisense | Chondrocytes | 6.8 |
| OSA | MVP (Sofer et al) | rs199445 | rs62058962 | *CR936218.2* | ENSG00000262881.1 | antisense | Chondrocytes | 6.8 |
| OSA | MVP (Sofer et al) | rs199445 | rs8078495 | *CR936218.2* | ENSG00000262881.1 | antisense | Chondrocytes | 6.8 |
| OSA | MVP (Sofer et al) | rs199445 | rs77768866 | *AC005829.1* | ENSG00000261575.2 | transcribed_unprocessed_pseudogene | Chondrocytes | 6.72 |
| OSA | MVP (Sofer et al) | rs199445 | rs62062282 | *AC005829.2* | ENSG00000262539.1 | processed_pseudogene | Chondrocytes | 6.28 |
| OSA | MVP (Sofer et al) | rs199445 | rs1052594 | *CR936218.1* | ENSG00000262372.1 | antisense | Chondrocytes | 6.22 |
| OSA | MVP (Sofer et al) | rs199445 | rs16940799 | *CR936218.1* | ENSG00000262372.1 | antisense | Chondrocytes | 6.22 |
| OSA | MVP (Sofer et al) | rs199445 | rs17574040 | *CR936218.1* | ENSG00000262372.1 | antisense | Chondrocytes | 6.22 |
| OSA | MVP (Sofer et al) | rs199445 | rs66561280 | *CR936218.1* | ENSG00000262372.1 | antisense | Chondrocytes | 6.22 |
| OSA | MVP (Sofer et al) | rs199445 | rs11079733 | *STH* | ENSG00000256762.1 | protein_coding | hESC_HypothalamicNeuron | 6.1 |
| OSA | MVP (Sofer et al) | rs199445 | rs17573447 | *MAP3K14-AS1* | ENSG00000267278.1 | antisense | iPSC_NPC | 5.67 |
| OSA | MVP (Sofer et al) | rs199445 | rs17573447 | *SPATA32* | ENSG00000184361.8 | protein_coding | iPSC_NPC | 5.67 |
| OSA | MVP (Sofer et al) | rs199445 | rs199442 | *TCONS_00025674* | TCONS_00025674 | lincRNA | hMSC_Osteoblasts | 5.07 |
| OSA | MVP (Sofer et al) | rs283789 | rs907593 | *AC046168.1* | ENSG00000259362.2 | lincRNA | Chondrocytes | 7.98 |
| OSA | MVP (Sofer et al) | rs283789 | rs907593 | *ETFA* | ENSG00000140374.11 | protein_coding | iPSC_Hepatocytes | 7.47 |
| OSA | MVP (Sofer et al) | rs283789 | rs908817 | *ISL2* | ENSG00000159556.5 | protein_coding | iPSC_NPC | 5.46 |
| OSA | MVP (Sofer et al) | rs429358 | rs429358 | *KLC3* | ENSG00000104892.17 | protein_coding | Chondrocytes | 7.45 |
| OSA | MVP (Sofer et al) | rs429358 | rs429358 | *RPS16P9* | ENSG00000242675.1 | processed_pseudogene | Chondrocytes | 7.45 |
| OSA | MVP (Sofer et al) | rs429358 | rs429358 | *EXOC3L2* | ENSG00000283632.2 | protein_coding | Chondrocytes | 6.81 |
| OSA | MVP (Sofer et al) | rs466614 | rs10760397 | *NR6A1* | ENSG00000148200.17 | protein_coding | Chondrocytes | 15.93 |
| OSA | MVP (Sofer et al) | rs466614 | rs10760397 | *AL354928.1* | ENSG00000236643.1 | antisense | Chondrocytes | 15.93 |
| OSA | MVP (Sofer et al) | rs466614 | rs10760397 | *OLFML2A* | ENSG00000185585.20 | protein_coding | Chondrocytes | 12.52 |
| OSA | MVP (Sofer et al) | rs466614 | rs372225 | *HSPA5* | ENSG00000044574.7 | protein_coding | GCB | 12.01 |
| OSA | MVP (Sofer et al) | rs466614 | rs372225 | *RP11-65N13.8* | ENSG00000239705.1 | antisense | GCB | 12.01 |
| OSA | MVP (Sofer et al) | rs466614 | rs372225 | *TCONS_l2_00029021* | TCONS_l2_00029021 | lincRNA | GCB | 12.01 |
| OSA | MVP (Sofer et al) | rs466614 | rs372225 | *TCONS_l2_00029022* | TCONS_l2_00029022 | lincRNA | GCB | 12.01 |
| OSA | MVP (Sofer et al) | rs466614 | rs10760397 | *RPL35* | ENSG00000136942.15 | protein_coding | Chondrocytes | 10.99 |
| OSA | MVP (Sofer et al) | rs466614 | rs10760397 | *ARPC5L* | ENSG00000136950.13 | protein_coding | Chondrocytes | 10.99 |
| OSA | MVP (Sofer et al) | rs466614 | rs10760397 | *MVB12B* | ENSG00000196814.15 | protein_coding | Chondrocytes | 10.6 |
| OSA | MVP (Sofer et al) | rs466614 | rs372225 | *PPP6C* | ENSG00000119414.7 | protein_coding | GCB | 9.03 |
| OSA | MVP (Sofer et al) | rs466614 | rs372225 | *RABEPK* | ENSG00000136933.12 | protein_coding | GCB | 8.3 |
| OSA | MVP (Sofer et al) | rs466614 | rs372225 | *TCONS_00016868* | TCONS_00016868 | lincRNA | GCB | 8.3 |
| OSA | MVP (Sofer et al) | rs466614 | rs7357647 | *OLFML2A* | ENSG00000185585.15 | protein_coding | iPSC_NPC | 8 |
| OSA | MVP (Sofer et al) | rs466614 | rs556781 | *HSPA5* | ENSG00000044574.8 | protein_coding | Chondrocytes | 7.77 |
| OSA | MVP (Sofer et al) | rs466614 | rs556781 | *AL354710.2* | ENSG00000239705.1 | antisense | Chondrocytes | 7.77 |
| OSA | MVP (Sofer et al) | rs466614 | rs10760397 | *WDR38* | ENSG00000136918.7 | protein_coding | Chondrocytes | 7.38 |
| OSA | MVP (Sofer et al) | rs466614 | rs556781 | *ARPC5L* | ENSG00000136950.13 | protein_coding | Chondrocytes | 7 |
| OSA | MVP (Sofer et al) | rs466614 | rs10760397 | *GOLGA1* | ENSG00000136935.13 | protein_coding | Chondrocytes | 6.51 |
| OSA | MVP (Sofer et al) | rs466614 | rs5015933 | *NR6A1* | ENSG00000148200.12 | protein_coding | hESC_HypothalamicNeuron | 5.26 |
| OSA | MVP (Sofer et al) | rs466614 | rs5015933 | *WDR38* | ENSG00000136918.3 | protein_coding | hESC_HypothalamicNeuron | 5.14 |
| OSA | MVP (Sofer et al) | rs4953923 | rs13018063 | *MGAT5* | ENSG00000152127.4 | protein_coding | GCB | 6.09 |
| OSA | MVP (Sofer et al) | rs4953923 | rs13018063 | *ACMSD* | ENSG00000153086.9 | protein_coding | iPSC_Neurons | 5.72 |
| OSA | MVP (Sofer et al) | rs4953923 | rs13018063 | *MIR3679* | ENSG00000263813.1 | miRNA | NaiveB | 5.72 |
| OSA | MVP (Sofer et al) | rs4953923 | rs71417530 | *TMEM163* | ENSG00000152128.13 | protein_coding | GCB | 5.4 |
| OSA | MVP (Sofer et al) | rs4953923 | rs13018063 | *TCONS_00004454* | TCONS_00004454 | lincRNA | hFOB | 5.14 |
| OSA | MVP (Sofer et al) | rs540606 | rs570305 | *LINC01833* | ENSG00000259439.2 | lincRNA | Chondrocytes | 8.82 |
| OSA | MVP (Sofer et al) | rs540606 | rs570436 | *LINC01833* | ENSG00000259439.2 | lincRNA | Chondrocytes | 8.82 |
| OSA | MVP (Sofer et al) | rs540606 | rs570305 | *SLC3A1* | ENSG00000138079.13 | protein_coding | Chondrocytes | 6.05 |
| OSA | MVP (Sofer et al) | rs540606 | rs570436 | *SLC3A1* | ENSG00000138079.13 | protein_coding | Chondrocytes | 6.05 |
| OSA | MVP (Sofer et al) | rs58857776 | rs6724384 | *ACTG1P22* | ENSG00000271615.1 | transcribed_processed_pseudogene | Chondrocytes | 7.34 |
| OSA | MVP (Sofer et al) | rs62048402 | rs1421085 | *CRNDE* | ENSG00000245694.9 | lincRNA | Chondrocytes | 11.33 |
| OSA | MVP (Sofer et al) | rs62048402 | rs9939973 | *CRNDE* | ENSG00000245694.9 | lincRNA | Chondrocytes | 11.33 |
| OSA | MVP (Sofer et al) | rs62048402 | rs9940128 | *CRNDE* | ENSG00000245694.9 | lincRNA | Chondrocytes | 11.33 |
| OSA | MVP (Sofer et al) | rs62048402 | rs9940646 | *CRNDE* | ENSG00000245694.9 | lincRNA | Chondrocytes | 11.33 |
| OSA | MVP (Sofer et al) | rs62048402 | rs1421085 | *IRX5* | ENSG00000176842.14 | protein_coding | Chondrocytes | 9.88 |
| OSA | MVP (Sofer et al) | rs62048402 | rs9939973 | *IRX5* | ENSG00000176842.14 | protein_coding | Chondrocytes | 9.88 |
| OSA | MVP (Sofer et al) | rs62048402 | rs9940128 | *IRX5* | ENSG00000176842.14 | protein_coding | Chondrocytes | 9.88 |
| OSA | MVP (Sofer et al) | rs62048402 | rs9940646 | *IRX5* | ENSG00000176842.14 | protein_coding | Chondrocytes | 9.88 |
| OSA | MVP (Sofer et al) | rs62048402 | rs11642015 | *AC106738.2* | ENSG00000259725.1 | lincRNA | Chondrocytes | 8.5 |
| OSA | MVP (Sofer et al) | rs62048402 | rs11642015 | *CRNDE* | ENSG00000245694.9 | lincRNA | Chondrocytes | 6.16 |
| OSA | MVP (Sofer et al) | rs6938973 | rs4839715 | *POU3F2* | ENSG00000184486.7 | protein_coding | iPSC_NPC | 6.57 |
| OSA | MVP (Sofer et al) | rs77881454 | rs17030925 | *AC098824.1* | ENSG00000232202.1 | processed_pseudogene | Chondrocytes | 35.12 |
| OSA | MVP (Sofer et al) | rs77881454 | rs17030925 | *ZFP36L2* | ENSG00000152518.8 | protein_coding | Chondrocytes | 32.24 |
| OSA | MVP (Sofer et al) | rs77881454 | rs17030925 | *AC010883.1* | ENSG00000234936.1 | antisense | Chondrocytes | 32.24 |
| OSA | MVP (Sofer et al) | rs77881454 | rs17030925 | *LINC01126* | ENSG00000279873.2 | lincRNA | Chondrocytes | 30.47 |
| OSA | MVP (Sofer et al) | rs77881454 | rs74600494 | *AC098824.1* | ENSG00000232202.1 | processed_pseudogene | Chondrocytes | 20.69 |
| OSA | MVP (Sofer et al) | rs77881454 | rs10169346 | *AC098824.1* | ENSG00000232202.1 | processed_pseudogene | Chondrocytes | 17.05 |
| OSA | MVP (Sofer et al) | rs77881454 | rs17030925 | *THADA* | ENSG00000115970.18 | protein_coding | Chondrocytes | 16.06 |
| OSA | MVP (Sofer et al) | rs77881454 | rs74600494 | *THADA* | ENSG00000115970.18 | protein_coding | Chondrocytes | 12.34 |
| OSA | MVP (Sofer et al) | rs77881454 | rs10169346 | *ZFP36L2* | ENSG00000152518.8 | protein_coding | Chondrocytes | 12.23 |
| OSA | MVP (Sofer et al) | rs77881454 | rs10169346 | *LINC01126* | ENSG00000279873.2 | lincRNA | Chondrocytes | 12.23 |
| OSA | MVP (Sofer et al) | rs77881454 | rs74600494 | *ZFP36L2* | ENSG00000152518.8 | protein_coding | Chondrocytes | 11.1 |
| OSA | MVP (Sofer et al) | rs77881454 | rs74600494 | *LINC01126* | ENSG00000279873.2 | lincRNA | Chondrocytes | 11.1 |
| OSA | MVP (Sofer et al) | rs77881454 | rs74600494 | *AC010883.1* | ENSG00000234936.1 | antisense | Chondrocytes | 10.78 |
| OSA | MVP (Sofer et al) | rs77881454 | rs10169346 | *THADA* | ENSG00000115970.18 | protein_coding | Chondrocytes | 10.23 |
| OSA | MVP (Sofer et al) | rs77881454 | rs74600494 | *AC016735.1* | ENSG00000224739.2 | lincRNA | Chondrocytes | 9.8 |
| OSA | MVP (Sofer et al) | rs77881454 | rs74382177 | *THADA* | ENSG00000115970.18 | protein_coding | Chondrocytes | 8.92 |
| OSA | MVP (Sofer et al) | rs77881454 | rs74382177 | *RN7SL531P* | ENSG00000264071.2 | misc_RNA | Chondrocytes | 8.92 |
| OSA | MVP (Sofer et al) | rs77881454 | rs17030925 | *AC016735.1* | ENSG00000224739.2 | lincRNA | Chondrocytes | 8.8 |
| OSA | MVP (Sofer et al) | rs77881454 | rs78314405 | TCONS_l2_00014535 | TCONS_l2_00014535 | lincRNA | iPSC_NPC | 7.09 |
| OSA | MVP (Sofer et al) | rs77881454 | rs78314405 | TCONS_l2_00014536 | TCONS_l2_00014536 | lincRNA | iPSC_NPC | 7.09 |
| OSA | MVP (Sofer et al) | rs77881454 | rs78886764 | *AC016735.2* | ENSG00000231826.1 | lincRNA | hFOB | 6.62 |
| OSA | MVP (Sofer et al) | rs77881454 | rs78886764 | TCONS_l2_00014537 | TCONS_l2_00014537 | lincRNA | hFOB | 6.62 |
| OSA | MVP (Sofer et al) | rs77881454 | rs17030925 | *FTOP1* | ENSG00000226491.1 | processed_pseudogene | Chondrocytes | 6.61 |
| OSA | MVP (Sofer et al) | rs77881454 | rs17030925 | *LINC01819* | ENSG00000231826.5 | lincRNA | Chondrocytes | 6.51 |

| **Supplementary Table 16.** Prioritized effector genes from variant to gene (V2G) mapping analysis. | | | | | | | | |
|---|---|---|---|---|---|---|---|---|
| **Trait** | **Discovery Study** | **Sentinel rsID** | **Proxy rsID** | **Gene Symbol** | **Gene Id** | **Gene Biotype** | **Cell** | **Contact Score (Chicago Score\|-log10(FitHiC2 FDR)** |
| OSA | MVP (Sofer et al) | rs77881454 | rs17030925 | *LINC02590* | ENSG00000233978.1 | lincRNA | Chondrocytes | 6.51 |
| OSA | MVP (Sofer et al) | rs77881454 | rs17030925 | TCONS_00004223 | TCONS_00004223 | lincRNA | SGBS_Diff | 6.3 |
| OSA | MVP (Sofer et al) | rs77881454 | rs17030925 | TCONS_00004224 | TCONS_00004224 | lincRNA | SGBS_Diff | 6.3 |
| OSA | MVP (Sofer et al) | rs77881454 | rs78314405 | *HAAO* | ENSG00000162882.10 | protein_coding | iPSC_NPC | 5.84 |
| OSA | MVP (Sofer et al) | rs77881454 | rs78886764 | TCONS_00003660 | TCONS_00003660 | lincRNA | hMSC_Adipocyte | 5.66 |
| OSA | MVP (Sofer et al) | rs77881454 | rs78886764 | TCONS_00004230 | TCONS_00004230 | lincRNA | GCB | 5.24 |
| OSA | MVP (Sofer et al) | rs77881454 | rs77527404 | TCONS_00003661 | TCONS_00003661 | lincRNA | HEPG2 | 5.18 |
| OSA | MVP (Sofer et al) | rs77881454 | rs78314405 | *AC093609.1* | ENSG00000230587.1 | lincRNA | iPSC_NPC | 5.1 |
| OSA | MVP (Sofer et al) | rs7869742 | rs4744133 | *AL157827.2* | ENSG00000285650.1 | antisense | Chondrocytes | 24 |
| OSA | MVP (Sofer et al) | rs7869742 | rs4744133 | *IPPK* | ENSG00000127080.10 | protein_coding | Chondrocytes | 21.72 |
| OSA | MVP (Sofer et al) | rs7869742 | rs4744133 | *CENPP* | ENSG00000188312.14 | protein_coding | Chondrocytes | 17.15 |
| OSA | MVP (Sofer et al) | rs7869742 | rs987554 | *MIR4670* | ENSG00000264158.1 | miRNA | SGBS_Diff | 9.7 |
| OSA | MVP (Sofer et al) | rs7869742 | rs4744133 | *ASPN* | ENSG00000106819.7 | protein_coding | hMSC_Osteoblasts | 8.03 |
| OSA | MVP (Sofer et al) | rs7869742 | rs927865 | *ECM2* | ENSG00000106823.8 | protein_coding | hESC_HypothalamicNPC | 6.55 |
| OSA | MVP (Sofer et al) | rs7869742 | rs4744133 | *NOL8* | ENSG00000198000.12 | protein_coding | Chondrocytes | 6.1 |
| OSA | MVP (Sofer et al) | rs7869742 | rs4744133 | *NOL8* | ENSG00000198000.7 | protein_coding | hMSC_Osteoblasts | 5.94 |
| OSA | MVP (Sofer et al) | rs869400 | rs869400 | *DGKG* | ENSG00000058866.15 | protein_coding | Chondrocytes | 17.08 |
| OSA | MVP (Sofer et al) | rs869400 | rs869400 | *DNAJB11* | ENSG00000090520.6 | protein_coding | GCB | 8.91 |
| OSA | MVP (Sofer et al) | rs869400 | rs869400 | *TBCCD1* | ENSG00000113838.8 | protein_coding | GCB | 8.91 |
| OSA | MVP (Sofer et al) | rs869400 | rs869400 | *EIF4A2* | ENSG00000156976.10 | protein_coding | TFH | 6.47 |
| OSA | MVP (Sofer et al) | rs869400 | rs869400 | *hsa-mir-1248* | hsa-mir-1248 | lincRNA | TFH | 6.47 |
| OSA | MVP (Sofer et al) | rs869400 | rs869400 | *RP11-573D15.9* | ENSG00000263826.1 | antisense | TFH | 6.47 |
| OSA | MVP (Sofer et al) | rs869400 | rs869400 | *SNORA63* | ENSG00000200418.1 | snoRNA | TFH | 6.47 |
| OSA | MVP (Sofer et al) | rs869400 | rs869400 | *SNORA81* | ENSG00000221420.2 | snoRNA | TFH | 6.47 |
| OSA | MVP (Sofer et al) | rs869400 | rs869400 | *SNORD2* | ENSG00000238942.1 | snoRNA | TFH | 6.47 |
| OSA | MVP (Sofer et al) | rs869400 | rs869400 | *snR39B* | snR39B | snoRNA | TFH | 6.47 |
| OSA | MVP (Sofer et al) | rs869400 | rs869400 | *RP11-443P15.2* | ENSG00000171658.4 | transcribed_unprocessed_pseudogene | iPSC_Hepatocytes | 5.54 |
| OSA | MVP (Sofer et al) | rs869400 | rs869400 | TCONS_l2_00019043 | TCONS_l2_00019043 | lincRNA | iPSC_Hepatocytes | 5.54 |
| OSA | MVP (Sofer et al) | rs869400 | rs869400 | *RP11-48F14.1* | ENSG00000224406.1 | lincRNA | GCB | 5.18 |
| OSA | MVP (Sofer et al) | rs869400 | rs869400 | TCONS_00005404 | TCONS_00005404 | lincRNA | GCB | 5.18 |
| OSA | MVP (Sofer et al) | rs869400 | rs869400 | TCONS_00006344 | TCONS_00006344 | lincRNA | GCB | 5.18 |
| OSA | MVP (Sofer et al) | rs869400 | rs869400 | TCONS_00006725 | TCONS_00006725 | lincRNA | GCB | 5.18 |
| OSA | MVP (Sofer et al) | rs869400 | rs869400 | *RFC4* | ENSG00000163918.6 | protein_coding | NaiveB | 5.17 |
| OSA | MVP (Sofer et al) | rs869400 | rs869400 | *RP11-573D15.3* | ENSG00000231724.1 | antisense | NaiveB | 5.17 |
| OSA | MVP (Sofer et al) | rs943037 | rs12412038 | *RPEL1* | ENSG00000235376.5 | protein_coding | Chondrocytes | 17.8 |
| OSA | MVP (Sofer et al) | rs943037 | rs12412038 | *RPEL1* | ENSG00000235376.5 | protein_coding | Chondrocytes | 17.8 |
| OSA | MVP (Sofer et al) | rs943037 | rs79780963 | *CNNM2* | ENSG00000148842.18 | protein_coding | Chondrocytes | 12.49 |
| OSA | MVP (Sofer et al) | rs943037 | rs3824754 | *WBP1L* | ENSG00000166272.18 | protein_coding | Chondrocytes | 10.08 |
| OSA | MVP (Sofer et al) | rs943037 | rs12412038 | *MARCKSL1P1* | ENSG00000213277.3 | processed_pseudogene | Chondrocytes | 9.81 |
| OSA | MVP (Sofer et al) | rs943037 | rs12412038 | *NT5C2* | ENSG00000076685.18 | protein_coding | Chondrocytes | 9.81 |
| OSA | MVP (Sofer et al) | rs943037 | rs12412038 | *NT5C2* | ENSG00000076685.18 | protein_coding | Chondrocytes | 9.81 |
| OSA | MVP (Sofer et al) | rs943037 | rs12412038 | *MARCKSL1P1* | ENSG00000213277.3 | processed_pseudogene | Chondrocytes | 9.81 |
| OSA | MVP (Sofer et al) | rs943037 | rs79780963 | *RPS15AP29* | ENSG00000237827.1 | processed_pseudogene | Chondrocytes | 8.94 |
| OSA | MVP (Sofer et al) | rs943037 | rs3824754 | *SFXN2* | ENSG00000156398.13 | protein_coding | Chondrocytes | 8.45 |
| OSA | MVP (Sofer et al) | rs943037 | rs12412038 | *AL356608.1* | ENSG00000272912.1 | lincRNA | Chondrocytes | 6.97 |
| OSA | MVP (Sofer et al) | rs943037 | rs12412038 | *CNNM2* | ENSG00000148842.18 | protein_coding | Chondrocytes | 6.77 |
| OSA | MVP (Sofer et al) | rs943037 | rs12412038 | *SFXN2* | ENSG00000156398.13 | protein_coding | Chondrocytes | 6.56 |
| OSA | MVP (Sofer et al) | rs943037 | rs79780963 | *NT5C2* | ENSG00000076685.18 | protein_coding | Chondrocytes | 6.5 |
| OSA | MVP (Sofer et al) | rs943037 | rs79780963 | *C10orf32* | ENSG00000166275.11 | protein_coding | TFH | 5.72 |
| OSA | MVP (Sofer et al) | rs943037 | rs79780963 | *C10orf32-ASMT* | ENSG00000270316.1 | protein_coding | TFH | 5.72 |
| OSA | MVP (Sofer et al) | rs943037 | rs3824754 | *AS3MT* | ENSG00000214435.3 | protein_coding | iPSC_Hepatocytes | 5.13 |
| OSA | Pooled | rs11642015 | rs9939973 | *CRNDE* | ENSG00000245694.9 | lincRNA | Chondrocytes | 11.33 |
| OSA | Pooled | rs11642015 | rs9940646 | *CRNDE* | ENSG00000245694.9 | lincRNA | Chondrocytes | 11.33 |
| OSA | Pooled | rs11642015 | rs9940128 | *CRNDE* | ENSG00000245694.9 | lincRNA | Chondrocytes | 11.33 |
| OSA | Pooled | rs11642015 | rs1421085 | *CRNDE* | ENSG00000245694.9 | lincRNA | Chondrocytes | 11.33 |
| OSA | Pooled | rs11642015 | rs9939973 | *IRX5* | ENSG00000176842.14 | protein_coding | Chondrocytes | 9.88 |
| OSA | Pooled | rs11642015 | rs9940646 | *IRX5* | ENSG00000176842.14 | protein_coding | Chondrocytes | 9.88 |
| OSA | Pooled | rs11642015 | rs9940128 | *IRX5* | ENSG00000176842.14 | protein_coding | Chondrocytes | 9.88 |
| OSA | Pooled | rs11642015 | rs1421085 | *IRX5* | ENSG00000176842.14 | protein_coding | Chondrocytes | 9.88 |
| OSA | Pooled | rs11642015 | rs11642015 | *AC106738.2* | ENSG00000259725.1 | lincRNA | Chondrocytes | 8.5 |
| OSA | Pooled | rs11642015 | rs11642015 | *CRNDE* | ENSG00000245694.9 | lincRNA | Chondrocytes | 6.16 |
| OSA | Pooled | rs1800437 | rs2287019 | *FBXO46* | ENSG00000177051.6 | protein_coding | Chondrocytes | 15.32 |
| OSA | Pooled | rs1800437 | rs11672660 | *GIPR* | ENSG00000010310.9 | protein_coding | Chondrocytes | 12.34 |
| OSA | Pooled | rs1800437 | rs34783010 | *GIPR* | ENSG00000010310.9 | protein_coding | Chondrocytes | 12.34 |
| OSA | Pooled | rs1800437 | rs11672660 | *VASP* | ENSG00000125753.14 | protein_coding | Chondrocytes | 11.94 |
| OSA | Pooled | rs1800437 | rs34783010 | *VASP* | ENSG00000125753.14 | protein_coding | Chondrocytes | 11.94 |
| OSA | Pooled | rs1800437 | rs11672660 | *MIR642A* | ENSG00000207773.1 | miRNA | Chondrocytes | 11.34 |
| OSA | Pooled | rs1800437 | rs34783010 | *MIR642A* | ENSG00000207773.1 | miRNA | Chondrocytes | 11.34 |
| OSA | Pooled | rs1800437 | rs11672660 | *PPM1N* | ENSG00000213889.10 | protein_coding | Chondrocytes | 10.3 |
| OSA | Pooled | rs1800437 | rs34783010 | *PPM1N* | ENSG00000213889.10 | protein_coding | Chondrocytes | 10.3 |
| OSA | Pooled | rs1800437 | rs2287019 | *VASP* | ENSG00000125753.14 | protein_coding | Chondrocytes | 8.66 |
| OSA | Pooled | rs1800437 | rs11672660 | *EML2* | ENSG00000125746.10 | protein_coding | TFH | 6.47 |
| OSA | Pooled | rs1800437 | rs34783010 | *EML2* | ENSG00000125746.10 | protein_coding | TFH | 6.47 |
| OSA | Pooled | rs1800437 | rs11672660 | *OPA3* | ENSG00000125741.4 | protein_coding | Chondrocytes | 6.32 |
| OSA | Pooled | rs1800437 | rs34783010 | *OPA3* | ENSG00000125741.4 | protein_coding | Chondrocytes | 6.32 |
| OSA | Pooled | rs1800437 | rs11672660 | *GPR4* | ENSG00000177464.5 | protein_coding | Chondrocytes | 6.32 |
| OSA | Pooled | rs1800437 | rs34783010 | *GPR4* | ENSG00000177464.5 | protein_coding | Chondrocytes | 6.32 |
| OSA | Pooled | rs1800437 | rs2287019 | *AC074212.1* | ENSG00000259605.3 | processed_transcript | Chondrocytes | 6.11 |
| OSA | Pooled | rs2376073 | rs79861574 | *LINC01159* | ENSG00000229743.3 | lincRNA | Chondrocytes | 8.1 |
| OSA | Pooled | rs2958127 | rs3214051 | *PTGES3* | ENSG00000110958.11 | protein_coding | TFH | 33.6 |
| OSA | Pooled | rs2958127 | rs3214051 | *PTGES3* | ENSG00000110958.11 | protein_coding | hMSC_Adipocyte | 30.51 |
| OSA | Pooled | rs2958127 | rs3214051 | *PTGES3* | ENSG00000110958.11 | protein_coding | NaiveT | 28.98 |
| OSA | Pooled | rs2958127 | rs3214051 | *PTGES3* | ENSG00000110958.11 | protein_coding | GCB | 28.95 |
| OSA | Pooled | rs2958127 | rs3214051 | *PTGES3* | ENSG00000110958.11 | protein_coding | SGBS_Diff | 26.21 |
| OSA | Pooled | rs2958127 | rs3214051 | *PTGES3* | ENSG00000110958.11 | protein_coding | hMSC_Osteoblasts | 24.85 |

**Supplementary Table 16.** Prioritized effector genes from variant to gene (V2G) mapping analysis.

| Trait | Discovery Study | Sentinel rsID | Proxy rsID | Gene Symbol | Gene Id | Gene Biotype | Cell | Contact Score (Chicago Score\|-log10(FitHiC2 FDR) |
|---|---|---|---|---|---|---|---|---|
| OSA | Pooled | rs2958127 | rs2255074 | *PTGES3* | ENSG00000110958.11 | protein_coding | iPSC_Hepatocytes | 24.2 |
| OSA | Pooled | rs2958127 | rs1107479 | *PTGES3* | ENSG00000110958.11 | protein_coding | iPSC_Hepatocytes | 24.2 |
| OSA | Pooled | rs2958127 | rs3214051 | *PTGES3* | ENSG00000110958.11 | protein_coding | NaiveB | 23.5 |
| OSA | Pooled | rs2958127 | rs3214051 | *PTGES3* | ENSG00000110958.11 | protein_coding | iPSC_Hepatocytes | 23.2 |
| OSA | Pooled | rs2958127 | rs3214051 | *PTGES3* | ENSG00000110958.11 | protein_coding | iPSC_NPC | 22.63 |
| OSA | Pooled | rs2958127 | rs3214051 | *PTGES3* | ENSG00000110958.11 | protein_coding | hESC_HypothalamicNeuron | 21.11 |
| OSA | Pooled | rs2958127 | rs3214051 | *STAT6* | ENSG00000166888.6 | protein_coding | NaiveT | 21.02 |
| OSA | Pooled | rs2958127 | rs3214051 | *PTGES3* | ENSG00000110958.11 | protein_coding | hFOB | 20.67 |
| OSA | Pooled | rs2958127 | rs2255074 | *PTGES3* | ENSG00000110958.11 | protein_coding | NaiveT | 19.97 |
| OSA | Pooled | rs2958127 | rs1107479 | *PTGES3* | ENSG00000110958.11 | protein_coding | NaiveT | 19.97 |
| OSA | Pooled | rs2958127 | rs2255074 | *PTGES3* | ENSG00000110958.11 | protein_coding | hMSC_Osteoblasts | 18.44 |
| OSA | Pooled | rs2958127 | rs1107479 | *PTGES3* | ENSG00000110958.11 | protein_coding | hMSC_Osteoblasts | 18.44 |
| OSA | Pooled | rs2958127 | rs2958153 | *ATP5B* | ENSG00000110955.4 | protein_coding | hESC_HypothalamicNeuron | 18.15 |
| OSA | Pooled | rs2958127 | rs2958153 | *U59B* | U59B | snoRNA | hESC_HypothalamicNeuron | 18.15 |
| OSA | Pooled | rs2958127 | rs2958153 | *SNORD59A* | ENSG00000207031.1 | snoRNA | hESC_HypothalamicNeuron | 18.15 |
| OSA | Pooled | rs2958127 | rs2255074 | *ANKRD52* | ENSG00000139645.9 | protein_coding | iPSC_NPC | 17.98 |
| OSA | Pooled | rs2958127 | rs1107479 | *ANKRD52* | ENSG00000139645.9 | protein_coding | iPSC_NPC | 17.98 |
| OSA | Pooled | rs2958127 | rs2255074 | *PTGES3* | ENSG00000110958.11 | protein_coding | TFH | 17.55 |
| OSA | Pooled | rs2958127 | rs1107479 | *PTGES3* | ENSG00000110958.11 | protein_coding | TFH | 17.55 |
| OSA | Pooled | rs2958127 | rs2255074 | *ATP5B* | ENSG00000110955.4 | protein_coding | hESC_HypothalamicNeuron | 17.42 |
| OSA | Pooled | rs2958127 | rs2255074 | *U59B* | U59B | snoRNA | hESC_HypothalamicNeuron | 17.42 |
| OSA | Pooled | rs2958127 | rs1107479 | *ATP5B* | ENSG00000110955.4 | protein_coding | hESC_HypothalamicNeuron | 17.42 |
| OSA | Pooled | rs2958127 | rs1107479 | *U59B* | U59B | snoRNA | hESC_HypothalamicNeuron | 17.42 |
| OSA | Pooled | rs2958127 | rs2255074 | *SNORD59A* | ENSG00000207031.1 | snoRNA | hESC_HypothalamicNeuron | 17.42 |
| OSA | Pooled | rs2958127 | rs1107479 | *SNORD59A* | ENSG00000207031.1 | snoRNA | hESC_HypothalamicNeuron | 17.42 |
| OSA | Pooled | rs2958127 | rs3214051 | *STAT6* | ENSG00000166888.6 | protein_coding | NaiveB | 17.02 |
| OSA | Pooled | rs2958127 | rs3214051 | *ATP5B* | ENSG00000110955.4 | protein_coding | iPSC_Hepatocytes | 16.68 |
| OSA | Pooled | rs2958127 | rs3214051 | *SNORD59A* | ENSG00000207031.1 | snoRNA | iPSC_Hepatocytes | 16.68 |
| OSA | Pooled | rs2958127 | rs3214051 | *NACA* | ENSG00000196531.6 | protein_coding | iPSC_Hepatocytes | 16.34 |
| OSA | Pooled | rs2958127 | rs2255074 | *BAZ2A* | ENSG00000076108.7 | protein_coding | hMSC_Adipocyte | 16.07 |
| OSA | Pooled | rs2958127 | rs1107479 | *BAZ2A* | ENSG00000076108.7 | protein_coding | hMSC_Adipocyte | 16.07 |
| OSA | Pooled | rs2958127 | rs2255074 | *BAZ2A* | ENSG00000076108.7 | protein_coding | SGBS_Diff | 15.61 |
| OSA | Pooled | rs2958127 | rs1107479 | *BAZ2A* | ENSG00000076108.7 | protein_coding | SGBS_Diff | 15.61 |
| OSA | Pooled | rs2958127 | rs3214051 | *ATP5B* | ENSG00000110955.4 | protein_coding | hMSC_Osteoblasts | 15.05 |
| OSA | Pooled | rs2958127 | rs3214051 | *U59B* | U59B | snoRNA | hMSC_Osteoblasts | 15.05 |
| OSA | Pooled | rs2958127 | rs3214051 | *SNORD59A* | ENSG00000207031.1 | snoRNA | hMSC_Osteoblasts | 15.05 |
| OSA | Pooled | rs2958127 | rs3214051 | *NACA* | ENSG00000196531.6 | protein_coding | iPSC_Neurons | 14.79 |
| OSA | Pooled | rs2958127 | rs3214051 | *NACA* | ENSG00000196531.6 | protein_coding | iPSC_NPC | 14.55 |
| OSA | Pooled | rs2958127 | rs2255074 | *BAZ2A* | ENSG00000076108.7 | protein_coding | iPSC_Hepatocytes | 14.54 |
| OSA | Pooled | rs2958127 | rs1107479 | *BAZ2A* | ENSG00000076108.7 | protein_coding | iPSC_Hepatocytes | 14.54 |
| OSA | Pooled | rs2958127 | rs3214051 | *ATP5B* | ENSG00000110955.4 | protein_coding | hMSC_Adipocyte | 14.42 |
| OSA | Pooled | rs2958127 | rs3214051 | *SNORD59A* | ENSG00000207031.1 | snoRNA | hMSC_Adipocyte | 14.42 |
| OSA | Pooled | rs2958127 | rs3214051 | *U59B* | U59B | snoRNA | hMSC_Adipocyte | 14.42 |
| OSA | Pooled | rs2958127 | rs3214051 | *STAT6* | ENSG00000166888.6 | protein_coding | iPSC_NPC | 14.35 |
| OSA | Pooled | rs2958127 | rs3214051 | *HSD17B6* | ENSG00000025423.7 | protein_coding | iPSC_NPC | 14.31 |
| OSA | Pooled | rs2958127 | rs3214051 | *PRIM1* | ENSG00000198056.9 | protein_coding | iPSC_NPC | 14.31 |
| OSA | Pooled | rs2958127 | rs2255074 | *ATP5B* | ENSG00000110955.4 | protein_coding | TFH | 14.3 |
| OSA | Pooled | rs2958127 | rs1107479 | *ATP5B* | ENSG00000110955.4 | protein_coding | TFH | 14.3 |
| OSA | Pooled | rs2958127 | rs2255074 | *SNORD59A* | ENSG00000207031.1 | snoRNA | TFH | 14.3 |
| OSA | Pooled | rs2958127 | rs1107479 | *SNORD59A* | ENSG00000207031.1 | snoRNA | TFH | 14.3 |
| OSA | Pooled | rs2958127 | rs2255074 | *ATP5F1B* | ENSG00000110955.9 | protein_coding | Chondrocytes | 14.19 |
| OSA | Pooled | rs2958127 | rs1107479 | *ATP5F1B* | ENSG00000110955.9 | protein_coding | Chondrocytes | 14.19 |
| OSA | Pooled | rs2958127 | rs2255074 | *SNORD59A* | ENSG00000207031.1 | snoRNA | Chondrocytes | 14.19 |
| OSA | Pooled | rs2958127 | rs1107479 | *SNORD59A* | ENSG00000207031.1 | snoRNA | Chondrocytes | 14.19 |
| OSA | Pooled | rs2958127 | rs2255074 | *BAZ2A* | ENSG00000076108.7 | protein_coding | hESC_HypothalamicNeuron | 14.18 |
| OSA | Pooled | rs2958127 | rs1107479 | *BAZ2A* | ENSG00000076108.7 | protein_coding | hESC_HypothalamicNeuron | 14.18 |
| OSA | Pooled | rs2958127 | rs941207 | *BAZ2A* | ENSG00000076108.7 | protein_coding | hMSC_Adipocyte | 14.07 |
| OSA | Pooled | rs2958127 | rs3214051 | *ZBTB39* | ENSG00000166860.2 | protein_coding | hESC_HypothalamicNeuron | 13.76 |
| OSA | Pooled | rs2958127 | rs941207 | *BAZ2A* | ENSG00000076108.7 | protein_coding | hESC_HypothalamicNeuron | 13.25 |
| OSA | Pooled | rs2958127 | rs1107479 | *U59B* | U59B | snoRNA | iPSC_Hepatocytes | 13.23 |
| OSA | Pooled | rs2958127 | rs2255074 | *ATP5B* | ENSG00000110955.4 | protein_coding | iPSC_Hepatocytes | 13.23 |
| OSA | Pooled | rs2958127 | rs2255074 | *SNORD59A* | ENSG00000207031.1 | snoRNA | iPSC_Hepatocytes | 13.23 |
| OSA | Pooled | rs2958127 | rs2255074 | *U59B* | U59B | snoRNA | iPSC_Hepatocytes | 13.23 |
| OSA | Pooled | rs2958127 | rs1107479 | *ATP5B* | ENSG00000110955.4 | protein_coding | iPSC_Hepatocytes | 13.23 |
| OSA | Pooled | rs2958127 | rs1107479 | *SNORD59A* | ENSG00000207031.1 | snoRNA | iPSC_Hepatocytes | 13.23 |
| OSA | Pooled | rs2958127 | rs2958153 | *BAZ2A* | ENSG00000076108.7 | protein_coding | iPSC_Hepatocytes | 13.23 |
| OSA | Pooled | rs2958127 | rs3214051 | *PTGES3* | ENSG00000110958.11 | protein_coding | iPSC_Neurons | 13.17 |
| OSA | Pooled | rs2958127 | rs2255074 | *U59B* | U59B | snoRNA | TFH | 13.03 |
| OSA | Pooled | rs2958127 | rs1107479 | *U59B* | U59B | snoRNA | TFH | 13.03 |
| OSA | Pooled | rs2958127 | rs3214051 | *BAZ2A* | ENSG00000076108.7 | protein_coding | TFH | 12.94 |
| OSA | Pooled | rs2958127 | rs3214051 | *GPR182* | ENSG00000166856.1 | protein_coding | hFOB | 12.92 |
| OSA | Pooled | rs2958127 | rs3214051 | *RP11-474N8.5* | ENSG00000258442.1 | protein_coding | hFOB | 12.92 |
| OSA | Pooled | rs2958127 | rs3214051 | *BAZ2A* | ENSG00000076108.7 | protein_coding | iPSC_Hepatocytes | 12.79 |
| OSA | Pooled | rs2958127 | rs2255074 | *BAZ2A* | ENSG00000076108.7 | protein_coding | NaiveB | 12.45 |
| OSA | Pooled | rs2958127 | rs1107479 | *BAZ2A* | ENSG00000076108.7 | protein_coding | NaiveB | 12.45 |
| OSA | Pooled | rs2958127 | rs2255074 | *ATP5B* | ENSG00000110955.4 | protein_coding | GCB | 12.44 |
| OSA | Pooled | rs2958127 | rs2255074 | *SNORD59A* | ENSG00000207031.1 | snoRNA | GCB | 12.44 |
| OSA | Pooled | rs2958127 | rs2255074 | *U59B* | U59B | snoRNA | GCB | 12.44 |
| OSA | Pooled | rs2958127 | rs1107479 | *ATP5B* | ENSG00000110955.4 | protein_coding | GCB | 12.44 |
| OSA | Pooled | rs2958127 | rs1107479 | *SNORD59A* | ENSG00000207031.1 | snoRNA | GCB | 12.44 |
| OSA | Pooled | rs2958127 | rs1107479 | *U59B* | U59B | snoRNA | GCB | 12.44 |
| OSA | Pooled | rs2958127 | rs3214051 | *ATP5B* | ENSG00000110955.4 | protein_coding | hFOB | 12.43 |
| OSA | Pooled | rs2958127 | rs3214051 | *SNORD59A* | ENSG00000207031.1 | snoRNA | hFOB | 12.43 |
| OSA | Pooled | rs2958127 | rs3214051 | *U59B* | U59B | snoRNA | hFOB | 12.43 |
| OSA | Pooled | rs2958127 | rs2255074 | *PTGES3* | ENSG00000110958.11 | protein_coding | NaiveB | 12.41 |

| **Supplementary Table 16.** Prioritized effector genes from variant to gene (V2G) mapping analysis. | | | | | | | | |
|---|---|---|---|---|---|---|---|---|
| **Trait** | **Discovery Study** | **Sentinel rsID** | **Proxy rsID** | **Gene Symbol** | **Gene Id** | **Gene Biotype** | **Cell** | **Contact Score (Chicago Score\|-log10(FitHiC2 FDR)** |
| OSA | Pooled | rs2958127 | rs1107479 | PTGES3 | ENSG00000110958.11 | protein_coding | NaiveB | 12.41 |
| OSA | Pooled | rs2958127 | rs3214051 | BAZ2A | ENSG00000076108.7 | protein_coding | NaiveT | 12.34 |
| OSA | Pooled | rs2958127 | rs3214051 | LRP1 | ENSG00000123384.9 | protein_coding | iPSC_NPC | 12.18 |
| OSA | Pooled | rs2958127 | rs3214051 | GPR182 | ENSG00000166856.1 | protein_coding | hMSC_Adipocyte | 12.17 |
| OSA | Pooled | rs2958127 | rs3214051 | RP11-474N8.5 | ENSG00000258442.1 | protein_coding | hMSC_Adipocyte | 12.17 |
| OSA | Pooled | rs2958127 | rs3214051 | U59B | U59B | snoRNA | iPSC_Hepatocytes | 12.14 |
| OSA | Pooled | rs2958127 | rs941207 | RBMS2 | ENSG00000076067.7 | protein_coding | SGBS_Diff | 12.12 |
| OSA | Pooled | rs2958127 | rs3214051 | BAZ2A | ENSG00000076108.7 | protein_coding | hMSC_Adipocyte | 12.06 |
| OSA | Pooled | rs2958127 | rs3214051 | ATP5B | ENSG00000110955.4 | protein_coding | hESC_HypothalamicNeuron | 12.05 |
| OSA | Pooled | rs2958127 | rs3214051 | SNORD59A | ENSG00000207031.1 | snoRNA | hESC_HypothalamicNeuron | 12.05 |
| OSA | Pooled | rs2958127 | rs3214051 | U59B | U59B | snoRNA | hESC_HypothalamicNeuron | 12.05 |
| OSA | Pooled | rs2958127 | rs2255074 | ATP5B | ENSG00000110955.4 | protein_coding | SGBS_Diff | 12.05 |
| OSA | Pooled | rs2958127 | rs2255074 | U59B | U59B | snoRNA | SGBS_Diff | 12.05 |
| OSA | Pooled | rs2958127 | rs2255074 | SNORD59A | ENSG00000207031.1 | snoRNA | SGBS_Diff | 12.05 |
| OSA | Pooled | rs2958127 | rs1107479 | ATP5B | ENSG00000110955.4 | protein_coding | SGBS_Diff | 12.05 |
| OSA | Pooled | rs2958127 | rs1107479 | U59B | U59B | snoRNA | SGBS_Diff | 12.05 |
| OSA | Pooled | rs2958127 | rs1107479 | SNORD59A | ENSG00000207031.1 | snoRNA | SGBS_Diff | 12.05 |
| OSA | Pooled | rs2958127 | rs941207 | ATP5B | ENSG00000110955.4 | protein_coding | GCB | 11.88 |
| OSA | Pooled | rs2958127 | rs941207 | SNORD59A | ENSG00000207031.1 | snoRNA | GCB | 11.88 |
| OSA | Pooled | rs2958127 | rs941207 | U59B | U59B | snoRNA | GCB | 11.88 |
| OSA | Pooled | rs2958127 | rs2255074 | BAZ2A | ENSG00000076108.7 | protein_coding | TFH | 11.81 |
| OSA | Pooled | rs2958127 | rs1107479 | BAZ2A | ENSG00000076108.7 | protein_coding | TFH | 11.81 |
| OSA | Pooled | rs2958127 | rs3214051 | STAT6 | ENSG00000166888.6 | protein_coding | TFH | 11.8 |
| OSA | Pooled | rs2958127 | rs941207 | ATP5B | ENSG00000110955.4 | protein_coding | hESC_HypothalamicNeuron | 11.73 |
| OSA | Pooled | rs2958127 | rs941207 | U59B | U59B | snoRNA | hESC_HypothalamicNeuron | 11.73 |
| OSA | Pooled | rs2958127 | rs941207 | SNORD59A | ENSG00000207031.1 | snoRNA | hESC_HypothalamicNeuron | 11.73 |
| OSA | Pooled | rs2958127 | rs2255074 | NACA | ENSG00000196531.6 | protein_coding | iPSC_Hepatocytes | 11.53 |
| OSA | Pooled | rs2958127 | rs1107479 | NACA | ENSG00000196531.6 | protein_coding | iPSC_Hepatocytes | 11.53 |
| OSA | Pooled | rs2958127 | rs2958153 | BAZ2A | ENSG00000076108.7 | protein_coding | NaiveT | 11.48 |
| OSA | Pooled | rs2958127 | rs2958153 | ATP5B | ENSG00000110955.4 | protein_coding | iPSC_NPC | 11.45 |
| OSA | Pooled | rs2958127 | rs2958153 | U59B | U59B | snoRNA | iPSC_NPC | 11.45 |
| OSA | Pooled | rs2958127 | rs2958153 | SNORD59A | ENSG00000207031.1 | snoRNA | iPSC_NPC | 11.45 |
| OSA | Pooled | rs2958127 | rs3214051 | ZBTB39 | ENSG00000166860.2 | protein_coding | hMSC_Adipocyte | 11.44 |
| OSA | Pooled | rs2958127 | rs3214051 | ZBTB39 | ENSG00000166860.2 | protein_coding | SGBS_Diff | 11.41 |
| OSA | Pooled | rs2958127 | rs3214051 | ATP5B | ENSG00000110955.4 | protein_coding | SGBS_Diff | 11.12 |
| OSA | Pooled | rs2958127 | rs3214051 | SNORD59A | ENSG00000207031.1 | snoRNA | SGBS_Diff | 11.12 |
| OSA | Pooled | rs2958127 | rs3214051 | U59B | U59B | snoRNA | SGBS_Diff | 11.12 |
| OSA | Pooled | rs2958127 | rs3214051 | ATP5B | ENSG00000110955.4 | protein_coding | TFH | 11.08 |
| OSA | Pooled | rs2958127 | rs3214051 | SNORD59A | ENSG00000207031.1 | snoRNA | TFH | 11.08 |
| OSA | Pooled | rs2958127 | rs2255074 | ATP5B | ENSG00000110955.4 | protein_coding | hMSC_Adipocyte | 10.94 |
| OSA | Pooled | rs2958127 | rs2255074 | U59B | U59B | snoRNA | hMSC_Adipocyte | 10.94 |
| OSA | Pooled | rs2958127 | rs2255074 | SNORD59A | ENSG00000207031.1 | snoRNA | hMSC_Adipocyte | 10.94 |
| OSA | Pooled | rs2958127 | rs1107479 | ATP5B | ENSG00000110955.4 | protein_coding | hMSC_Adipocyte | 10.94 |
| OSA | Pooled | rs2958127 | rs1107479 | U59B | U59B | snoRNA | hMSC_Adipocyte | 10.94 |
| OSA | Pooled | rs2958127 | rs1107479 | SNORD59A | ENSG00000207031.1 | snoRNA | hMSC_Adipocyte | 10.94 |
| OSA | Pooled | rs2958127 | rs2255074 | RBMS2 | ENSG00000076067.7 | protein_coding | hMSC_Adipocyte | 10.76 |
| OSA | Pooled | rs2958127 | rs1107479 | RBMS2 | ENSG00000076067.7 | protein_coding | hMSC_Adipocyte | 10.76 |
| OSA | Pooled | rs2958127 | rs2255074 | BAZ2A | ENSG00000076108.7 | protein_coding | hFOB | 10.65 |
| OSA | Pooled | rs2958127 | rs1107479 | BAZ2A | ENSG00000076108.7 | protein_coding | hFOB | 10.65 |
| OSA | Pooled | rs2958127 | rs2958153 | NACA | ENSG00000196531.6 | protein_coding | iPSC_Hepatocytes | 10.56 |
| OSA | Pooled | rs2958127 | rs3214051 | ZBTB39 | ENSG00000166860.2 | protein_coding | NaiveB | 10.5 |
| OSA | Pooled | rs2958127 | rs941207 | RBMS2 | ENSG00000076067.7 | protein_coding | iPSC_NPC | 10.48 |
| OSA | Pooled | rs2958127 | rs3214051 | BAZ2A | ENSG00000076108.7 | protein_coding | hMSC_Osteoblasts | 10.43 |
| OSA | Pooled | rs2958127 | rs2255074 | ATP5B | ENSG00000110955.4 | protein_coding | NaiveB | 10.39 |
| OSA | Pooled | rs2958127 | rs2255074 | U59B | U59B | snoRNA | NaiveB | 10.39 |
| OSA | Pooled | rs2958127 | rs1107479 | ATP5B | ENSG00000110955.4 | protein_coding | NaiveB | 10.39 |
| OSA | Pooled | rs2958127 | rs1107479 | U59B | U59B | snoRNA | NaiveB | 10.39 |
| OSA | Pooled | rs2958127 | rs941207 | BAZ2A | ENSG00000076108.7 | protein_coding | NaiveB | 10.34 |
| OSA | Pooled | rs2958127 | rs3214051 | NACA | ENSG00000196531.6 | protein_coding | SGBS_Diff | 10.33 |
| OSA | Pooled | rs2958127 | rs2958153 | NACA | ENSG00000196531.6 | protein_coding | GCB | 10.29 |
| OSA | Pooled | rs2958127 | rs2255074 | ATP5B | ENSG00000110955.4 | protein_coding | hMSC_Osteoblasts | 10.22 |
| OSA | Pooled | rs2958127 | rs2255074 | U59B | U59B | snoRNA | hMSC_Osteoblasts | 10.22 |
| OSA | Pooled | rs2958127 | rs2255074 | SNORD59A | ENSG00000207031.1 | snoRNA | hMSC_Osteoblasts | 10.22 |
| OSA | Pooled | rs2958127 | rs1107479 | ATP5B | ENSG00000110955.4 | protein_coding | hMSC_Osteoblasts | 10.22 |
| OSA | Pooled | rs2958127 | rs1107479 | U59B | U59B | snoRNA | hMSC_Osteoblasts | 10.22 |
| OSA | Pooled | rs2958127 | rs1107479 | SNORD59A | ENSG00000207031.1 | snoRNA | hMSC_Osteoblasts | 10.22 |
| OSA | Pooled | rs2958127 | rs3214051 | GPR182 | ENSG00000166856.1 | protein_coding | iPSC_Hepatocytes | 10.22 |
| OSA | Pooled | rs2958127 | rs3214051 | RP11-474N8.5 | ENSG00000258442.1 | protein_coding | iPSC_Hepatocytes | 10.22 |
| OSA | Pooled | rs2958127 | rs2255074 | NACA | ENSG00000196531.6 | protein_coding | TFH | 10.13 |
| OSA | Pooled | rs2958127 | rs1107479 | NACA | ENSG00000196531.6 | protein_coding | TFH | 10.13 |
| OSA | Pooled | rs2958127 | rs3214051 | ZBTB39 | ENSG00000166860.2 | protein_coding | iPSC_NPC | 10.09 |
| OSA | Pooled | rs2958127 | rs3214051 | GPR182 | ENSG00000166856.1 | protein_coding | SGBS_Diff | 10.09 |
| OSA | Pooled | rs2958127 | rs3214051 | RP11-474N8.5 | ENSG00000258442.1 | protein_coding | SGBS_Diff | 10.09 |
| OSA | Pooled | rs2958127 | rs3214051 | ZBTB39 | ENSG00000166860.2 | protein_coding | hMSC_Osteoblasts | 10 |
| OSA | Pooled | rs2958127 | rs3214051 | TCONS_00020445 | TCONS_00020445 | lincRNA | iPSC_NPC | 9.98 |
| OSA | Pooled | rs2958127 | rs3214051 | TCONS_00020807 | TCONS_00020807 | lincRNA | iPSC_NPC | 9.98 |
| OSA | Pooled | rs2958127 | rs2255074 | BAZ2A | ENSG00000076108.7 | protein_coding | iPSC_NPC | 9.94 |
| OSA | Pooled | rs2958127 | rs1107479 | BAZ2A | ENSG00000076108.7 | protein_coding | iPSC_NPC | 9.94 |
| OSA | Pooled | rs2958127 | rs941207 | BAZ2A | ENSG00000076108.7 | protein_coding | GCB | 9.91 |
| OSA | Pooled | rs2958127 | rs2255074 | PTGES3 | ENSG00000110958.11 | protein_coding | iPSC_Neurons | 9.72 |
| OSA | Pooled | rs2958127 | rs1107479 | PTGES3 | ENSG00000110958.11 | protein_coding | iPSC_Neurons | 9.72 |
| OSA | Pooled | rs2958127 | rs2255074 | BAZ2A | ENSG00000076108.7 | protein_coding | GCB | 9.69 |
| OSA | Pooled | rs2958127 | rs1107479 | BAZ2A | ENSG00000076108.7 | protein_coding | GCB | 9.69 |
| OSA | Pooled | rs2958127 | rs2255074 | ATP5B | ENSG00000110955.4 | protein_coding | NaiveT | 9.69 |
| OSA | Pooled | rs2958127 | rs2255074 | SNORD59A | ENSG00000207031.1 | snoRNA | NaiveT | 9.69 |

| **Supplementary Table 16.** Prioritized effector genes from variant to gene (V2G) mapping analysis. | | | | | | | | |
|---|---|---|---|---|---|---|---|---|
| Trait | Discovery Study | Sentinel rsID | Proxy rsID | Gene Symbol | Gene Id | Gene Biotype | Cell | Contact Score (Chicago Score\|-log10(FitHiC2 FDR) |
| OSA | Pooled | rs2958127 | rs2255074 | *U59B* | U59B | snoRNA | NaiveT | 9.69 |
| OSA | Pooled | rs2958127 | rs1107479 | *ATP5B* | ENSG00000110955.4 | protein_coding | NaiveT | 9.69 |
| OSA | Pooled | rs2958127 | rs1107479 | *SNORD59A* | ENSG00000207031.1 | snoRNA | NaiveT | 9.69 |
| OSA | Pooled | rs2958127 | rs1107479 | *U59B* | U59B | snoRNA | NaiveT | 9.69 |
| OSA | Pooled | rs2958127 | rs3214051 | TCONS_00020445 | TCONS_00020445 | lincRNA | TFH | 9.62 |
| OSA | Pooled | rs2958127 | rs3214051 | TCONS_00020807 | TCONS_00020807 | lincRNA | TFH | 9.62 |
| OSA | Pooled | rs2958127 | rs2958153 | *NACA* | ENSG00000196531.6 | protein_coding | TFH | 9.58 |
| OSA | Pooled | rs2958127 | rs2958153 | *NACA* | ENSG00000196531.6 | protein_coding | hMSC_Adipocyte | 9.57 |
| OSA | Pooled | rs2958127 | rs941207 | *ATP5B* | ENSG00000110955.4 | protein_coding | TFH | 9.57 |
| OSA | Pooled | rs2958127 | rs941207 | *SNORD59A* | ENSG00000207031.1 | snoRNA | TFH | 9.57 |
| OSA | Pooled | rs2958127 | rs941207 | *U59B* | U59B | snoRNA | TFH | 9.57 |
| OSA | Pooled | rs2958127 | rs3214051 | TCONS_00020445 | TCONS_00020445 | lincRNA | GCB | 9.56 |
| OSA | Pooled | rs2958127 | rs3214051 | TCONS_00020807 | TCONS_00020807 | lincRNA | GCB | 9.56 |
| OSA | Pooled | rs2958127 | rs3214051 | *NXPH4* | ENSG00000182379.9 | protein_coding | hFOB | 9.49 |
| OSA | Pooled | rs2958127 | rs2958153 | *ATP5B* | ENSG00000110955.4 | protein_coding | iPSC_Hepatocytes | 9.48 |
| OSA | Pooled | rs2958127 | rs2958153 | *SNORD59A* | ENSG00000207031.1 | snoRNA | iPSC_Hepatocytes | 9.48 |
| OSA | Pooled | rs2958127 | rs2958153 | *U59B* | U59B | snoRNA | iPSC_Hepatocytes | 9.48 |
| OSA | Pooled | rs2958127 | rs3214051 | *GPR182* | ENSG00000166856.1 | protein_coding | hESC_HypothalamicNPC | 9.47 |
| OSA | Pooled | rs2958127 | rs3214051 | *RP11-474N8.5* | ENSG00000258442.1 | protein_coding | hESC_HypothalamicNPC | 9.47 |
| OSA | Pooled | rs2958127 | rs941207 | *RBMS2* | ENSG00000076067.7 | protein_coding | hMSC_Adipocyte | 9.43 |
| OSA | Pooled | rs2958127 | rs941207 | *RBMS2* | ENSG00000076067.7 | protein_coding | hESC_HypothalamicNeuron | 9.42 |
| OSA | Pooled | rs2958127 | rs3214051 | *STAT6* | ENSG00000166888.6 | protein_coding | SGBS_Diff | 9.42 |
| OSA | Pooled | rs2958127 | rs3214051 | TCONS_00020445 | TCONS_00020445 | lincRNA | NaiveB | 9.39 |
| OSA | Pooled | rs2958127 | rs3214051 | TCONS_00020807 | TCONS_00020807 | lincRNA | NaiveB | 9.39 |
| OSA | Pooled | rs2958127 | rs3214051 | *NACA* | ENSG00000196531.6 | protein_coding | hMSC_Osteoblasts | 9.38 |
| OSA | Pooled | rs2958127 | rs2255074 | *BAZ2A* | ENSG00000076108.7 | protein_coding | hMSC_Osteoblasts | 9.36 |
| OSA | Pooled | rs2958127 | rs1107479 | *BAZ2A* | ENSG00000076108.7 | protein_coding | hMSC_Osteoblasts | 9.36 |
| OSA | Pooled | rs2958127 | rs3214051 | *ZBTB39* | ENSG00000166860.2 | protein_coding | hFOB | 9.3 |
| OSA | Pooled | rs2958127 | rs941207 | *NACA* | ENSG00000196531.6 | protein_coding | NaiveT | 9.25 |
| OSA | Pooled | rs2958127 | rs2958153 | *ATP5B* | ENSG00000110955.4 | protein_coding | hMSC_Osteoblasts | 9.21 |
| OSA | Pooled | rs2958127 | rs2958153 | *SNORD59A* | ENSG00000207031.1 | snoRNA | hMSC_Osteoblasts | 9.21 |
| OSA | Pooled | rs2958127 | rs2958153 | *U59B* | U59B | snoRNA | hMSC_Osteoblasts | 9.21 |
| OSA | Pooled | rs2958127 | rs2958153 | *ATP5B* | ENSG00000110955.4 | protein_coding | SGBS_Diff | 9.12 |
| OSA | Pooled | rs2958127 | rs2958153 | *U59B* | U59B | snoRNA | SGBS_Diff | 9.12 |
| OSA | Pooled | rs2958127 | rs2958153 | *SNORD59A* | ENSG00000207031.1 | snoRNA | SGBS_Diff | 9.12 |
| OSA | Pooled | rs2958127 | rs2255074 | *SNORD59A* | ENSG00000207031.1 | snoRNA | NaiveB | 9.04 |
| OSA | Pooled | rs2958127 | rs1107479 | *SNORD59A* | ENSG00000207031.1 | snoRNA | NaiveB | 9.04 |
| OSA | Pooled | rs2958127 | rs2958153 | *NACA* | ENSG00000196531.6 | protein_coding | NaiveT | 9.01 |
| OSA | Pooled | rs2958127 | rs2255074 | *NACA* | ENSG00000196531.6 | protein_coding | iPSC_Neurons | 8.96 |
| OSA | Pooled | rs2958127 | rs1107479 | *NACA* | ENSG00000196531.6 | protein_coding | iPSC_Neurons | 8.96 |
| OSA | Pooled | rs2958127 | rs3214051 | *GPR182* | ENSG00000166856.1 | protein_coding | NaiveB | 8.91 |
| OSA | Pooled | rs2958127 | rs3214051 | *RP11-474N8.5* | ENSG00000258442.1 | protein_coding | NaiveB | 8.91 |
| OSA | Pooled | rs2958127 | rs2958153 | *NACA* | ENSG00000196531.6 | protein_coding | SGBS_Diff | 8.85 |
| OSA | Pooled | rs2958127 | rs3214051 | *ATP5B* | ENSG00000110955.4 | protein_coding | GCB | 8.83 |
| OSA | Pooled | rs2958127 | rs3214051 | *SNORD59A* | ENSG00000207031.1 | snoRNA | GCB | 8.83 |
| OSA | Pooled | rs2958127 | rs3214051 | *U59B* | U59B | snoRNA | GCB | 8.83 |
| OSA | Pooled | rs2958127 | rs941207 | *ATP5B* | ENSG00000110955.4 | protein_coding | NaiveB | 8.83 |
| OSA | Pooled | rs2958127 | rs941207 | *SNORD59A* | ENSG00000207031.1 | snoRNA | NaiveB | 8.83 |
| OSA | Pooled | rs2958127 | rs3214051 | *ZBTB39* | ENSG00000166860.2 | protein_coding | NaiveT | 8.77 |
| OSA | Pooled | rs2958127 | rs941207 | *BAZ2A* | ENSG00000076108.7 | protein_coding | TFH | 8.75 |
| OSA | Pooled | rs2958127 | rs2255074 | *NACA* | ENSG00000196531.6 | protein_coding | hMSC_Adipocyte | 8.74 |
| OSA | Pooled | rs2958127 | rs1107479 | *NACA* | ENSG00000196531.6 | protein_coding | hMSC_Adipocyte | 8.74 |
| OSA | Pooled | rs2958127 | rs941207 | *RBMS2* | ENSG00000076067.7 | protein_coding | iPSC_Neurons | 8.74 |
| OSA | Pooled | rs2958127 | rs2958153 | *NACA* | ENSG00000196531.6 | protein_coding | iPSC_NPC | 8.68 |
| OSA | Pooled | rs2958127 | rs3214051 | *BAZ2A* | ENSG00000076108.7 | protein_coding | SGBS_Diff | 8.66 |
| OSA | Pooled | rs2958127 | rs941207 | *BAZ2A* | ENSG00000076108.7 | protein_coding | iPSC_NPC | 8.63 |
| OSA | Pooled | rs2958127 | rs2255074 | *ATP5B* | ENSG00000110955.4 | protein_coding | iPSC_NPC | 8.62 |
| OSA | Pooled | rs2958127 | rs2255074 | *SNORD59A* | ENSG00000207031.1 | snoRNA | iPSC_NPC | 8.62 |
| OSA | Pooled | rs2958127 | rs1107479 | *ATP5B* | ENSG00000110955.4 | protein_coding | iPSC_NPC | 8.62 |
| OSA | Pooled | rs2958127 | rs1107479 | *SNORD59A* | ENSG00000207031.1 | snoRNA | iPSC_NPC | 8.62 |
| OSA | Pooled | rs2958127 | rs7980317 | *ZBTB39* | ENSG00000166860.2 | protein_coding | hMSC_Adipocyte | 8.6 |
| OSA | Pooled | rs2958127 | rs3214051 | *NAB2* | ENSG00000166886.8 | protein_coding | hFOB | 8.54 |
| OSA | Pooled | rs2958127 | rs3214051 | TCONS_00020445 | TCONS_00020445 | lincRNA | hFOB | 8.54 |
| OSA | Pooled | rs2958127 | rs3214051 | TCONS_00020805 | TCONS_00020805 | lincRNA | hFOB | 8.54 |
| OSA | Pooled | rs2958127 | rs3214051 | TCONS_00020806 | TCONS_00020806 | lincRNA | hFOB | 8.54 |
| OSA | Pooled | rs2958127 | rs3214051 | TCONS_00020807 | TCONS_00020807 | lincRNA | hFOB | 8.54 |
| OSA | Pooled | rs2958127 | rs3214051 | TCONS_00021326 | TCONS_00021326 | lincRNA | hFOB | 8.54 |
| OSA | Pooled | rs2958127 | rs3214051 | TCONS_00021327 | TCONS_00021327 | lincRNA | hFOB | 8.54 |
| OSA | Pooled | rs2958127 | rs3214051 | *TMEM194A* | ENSG00000166881.5 | protein_coding | hFOB | 8.54 |
| OSA | Pooled | rs2958127 | rs3214051 | *BAZ2A* | ENSG00000076108.7 | protein_coding | iPSC_Neurons | 8.46 |
| OSA | Pooled | rs2958127 | rs2255074 | *ATP5B* | ENSG00000110955.4 | protein_coding | hFOB | 8.31 |
| OSA | Pooled | rs2958127 | rs2255074 | *U59B* | U59B | snoRNA | hFOB | 8.31 |
| OSA | Pooled | rs2958127 | rs2255074 | *SNORD59A* | ENSG00000207031.1 | snoRNA | hFOB | 8.31 |
| OSA | Pooled | rs2958127 | rs1107479 | *ATP5B* | ENSG00000110955.4 | protein_coding | hFOB | 8.31 |
| OSA | Pooled | rs2958127 | rs1107479 | *U59B* | U59B | snoRNA | hFOB | 8.31 |
| OSA | Pooled | rs2958127 | rs1107479 | *SNORD59A* | ENSG00000207031.1 | snoRNA | hFOB | 8.31 |
| OSA | Pooled | rs2958127 | rs2255074 | *NACA* | ENSG00000196531.6 | protein_coding | hMSC_Osteoblasts | 8.3 |
| OSA | Pooled | rs2958127 | rs1107479 | *NACA* | ENSG00000196531.6 | protein_coding | hMSC_Osteoblasts | 8.3 |
| OSA | Pooled | rs2958127 | rs3214051 | *NACA* | ENSG00000196531.6 | protein_coding | hESC_HypothalamicNeuron | 8.26 |
| OSA | Pooled | rs2958127 | rs941207 | *ZBTB39* | ENSG00000166860.2 | protein_coding | hESC_HypothalamicNeuron | 8.22 |
| OSA | Pooled | rs2958127 | rs2958153 | *ZBTB39* | ENSG00000166860.2 | protein_coding | iPSC_Hepatocytes | 8.21 |
| OSA | Pooled | rs2958127 | rs2255074 | *STAT6* | ENSG00000166888.6 | protein_coding | iPSC_NPC | 8.15 |
| OSA | Pooled | rs2958127 | rs1107479 | *STAT6* | ENSG00000166888.6 | protein_coding | iPSC_NPC | 8.15 |
| OSA | Pooled | rs2958127 | rs3214051 | *ATP5B* | ENSG00000110955.4 | protein_coding | iPSC_NPC | 8.09 |
| OSA | Pooled | rs2958127 | rs3214051 | *SNORD59A* | ENSG00000207031.1 | snoRNA | iPSC_NPC | 8.09 |

| **Supplementary Table 16.** Prioritized effector genes from variant to gene (V2G) mapping analysis. | | | | | | | | |
|---|---|---|---|---|---|---|---|---|
| **Trait** | **Discovery Study** | **Sentinel rsID** | **Proxy rsID** | **Gene Symbol** | **Gene Id** | **Gene Biotype** | **Cell** | **Contact Score (Chicago Score\|-log10(FitHiC2 FDR)** |
| OSA | Pooled | rs2958127 | rs3214051 | *U59B* | U59B | snoRNA | iPSC_NPC | 8.09 |
| OSA | Pooled | rs2958127 | rs2958153 | *NACA* | ENSG00000196531.6 | protein_coding | hMSC_Osteoblasts | 8.08 |
| OSA | Pooled | rs2958127 | rs3214051 | *BAZ2A* | ENSG00000076108.7 | protein_coding | GCB | 8.05 |
| OSA | Pooled | rs2958127 | rs3214051 | TCONS_00020445 | TCONS_00020445 | lincRNA | NaiveT | 8.02 |
| OSA | Pooled | rs2958127 | rs3214051 | TCONS_00020807 | TCONS_00020807 | lincRNA | NaiveT | 8.02 |
| OSA | Pooled | rs2958127 | rs2958153 | *BAZ2A* | ENSG00000076108.7 | protein_coding | hMSC_Osteoblasts | 8 |
| OSA | Pooled | rs2958127 | rs2255074 | *NACA* | ENSG00000196531.6 | protein_coding | NaiveT | 7.94 |
| OSA | Pooled | rs2958127 | rs1107479 | *NACA* | ENSG00000196531.6 | protein_coding | NaiveT | 7.94 |
| OSA | Pooled | rs2958127 | rs3214051 | *NACA* | ENSG00000196531.6 | protein_coding | TFH | 7.9 |
| OSA | Pooled | rs2958127 | rs3214051 | *BAZ2A* | ENSG00000076108.7 | protein_coding | NaiveB | 7.76 |
| OSA | Pooled | rs2958127 | rs3214051 | *GPR182* | ENSG00000166856.1 | protein_coding | hMSC_Osteoblasts | 7.65 |
| OSA | Pooled | rs2958127 | rs3214051 | *RP11-474N8.5* | ENSG00000258442.1 | protein_coding | hMSC_Osteoblasts | 7.65 |
| OSA | Pooled | rs2958127 | rs941207 | *NACA* | ENSG00000196531.6 | protein_coding | TFH | 7.64 |
| OSA | Pooled | rs2958127 | rs2255074 | *COQ10A* | ENSG00000135469.9 | protein_coding | hFOB | 7.57 |
| OSA | Pooled | rs2958127 | rs1107479 | *COQ10A* | ENSG00000135469.9 | protein_coding | hFOB | 7.57 |
| OSA | Pooled | rs2958127 | rs2958153 | *ZBTB39* | ENSG00000166860.2 | protein_coding | iPSC_Neurons | 7.57 |
| OSA | Pooled | rs2958127 | rs3214051 | *ATP5B* | ENSG00000110955.4 | protein_coding | iPSC_Neurons | 7.54 |
| OSA | Pooled | rs2958127 | rs3214051 | *SNORD59A* | ENSG00000207031.1 | snoRNA | iPSC_Neurons | 7.54 |
| OSA | Pooled | rs2958127 | rs2958153 | *NACA* | ENSG00000196531.6 | protein_coding | hESC_HypothalamicNeuron | 7.5 |
| OSA | Pooled | rs2958127 | rs941207 | *BAZ2A* | ENSG00000076108.7 | protein_coding | SGBS_Diff | 7.34 |
| OSA | Pooled | rs2958127 | rs2958153 | *ATP5B* | ENSG00000110955.4 | protein_coding | hESC_HypothalamicNPC | 7.33 |
| OSA | Pooled | rs2958127 | rs2958153 | *SNORD59A* | ENSG00000207031.1 | snoRNA | hESC_HypothalamicNPC | 7.33 |
| OSA | Pooled | rs2958127 | rs2958153 | *U59B* | U59B | snoRNA | hESC_HypothalamicNPC | 7.33 |
| OSA | Pooled | rs2958127 | rs941207 | *NACA* | ENSG00000196531.6 | protein_coding | hMSC_Adipocyte | 7.33 |
| OSA | Pooled | rs2958127 | rs5798394 | *ZBTB39* | ENSG00000166860.2 | protein_coding | hESC_HypothalamicNeuron | 7.31 |
| OSA | Pooled | rs2958127 | rs2255074 | *U59B* | U59B | snoRNA | iPSC_NPC | 7.29 |
| OSA | Pooled | rs2958127 | rs1107479 | *U59B* | U59B | snoRNA | iPSC_NPC | 7.29 |
| OSA | Pooled | rs2958127 | rs2958153 | *NACA* | ENSG00000196531.6 | protein_coding | NaiveB | 7.22 |
| OSA | Pooled | rs2958127 | rs941207 | *ATP5B* | ENSG00000110955.4 | protein_coding | SGBS_Diff | 7.22 |
| OSA | Pooled | rs2958127 | rs941207 | *SNORD59A* | ENSG00000207031.1 | snoRNA | SGBS_Diff | 7.22 |
| OSA | Pooled | rs2958127 | rs941207 | *U59B* | U59B | snoRNA | SGBS_Diff | 7.22 |
| OSA | Pooled | rs2958127 | rs2255074 | *BAZ2A* | ENSG00000076108.7 | protein_coding | HEPG2 | 7.16 |
| OSA | Pooled | rs2958127 | rs1107479 | *BAZ2A* | ENSG00000076108.7 | protein_coding | HEPG2 | 7.16 |
| OSA | Pooled | rs2958127 | rs3214051 | *BAZ2A* | ENSG00000076108.7 | protein_coding | hESC_HypothalamicNeuron | 7.15 |
| OSA | Pooled | rs2958127 | rs3214051 | *GPR182* | ENSG00000166856.1 | protein_coding | iPSC_NPC | 7.15 |
| OSA | Pooled | rs2958127 | rs3214051 | *RP11-474N8.5* | ENSG00000258442.1 | protein_coding | iPSC_NPC | 7.15 |
| OSA | Pooled | rs2958127 | rs3214051 | *ZBTB39* | ENSG00000166860.2 | protein_coding | HEPG2 | 7.14 |
| OSA | Pooled | rs2958127 | rs941207 | *ESYT1* | ENSG00000139641.8 | protein_coding | iPSC_NPC | 7.09 |
| OSA | Pooled | rs2958127 | rs941207 | *ZC3H10* | ENSG00000135482.2 | protein_coding | iPSC_NPC | 7.09 |
| OSA | Pooled | rs2958127 | rs941207 | *MIP* | ENSG00000135517.6 | protein_coding | iPSC_NPC | 7.05 |
| OSA | Pooled | rs2958127 | rs941207 | *SPRYD4* | ENSG00000176422.10 | protein_coding | iPSC_NPC | 7.05 |
| OSA | Pooled | rs2958127 | rs2255074 | *ZBTB39* | ENSG00000166860.3 | protein_coding | Chondrocytes | 7.02 |
| OSA | Pooled | rs2958127 | rs1107479 | *ZBTB39* | ENSG00000166860.3 | protein_coding | Chondrocytes | 7.02 |
| OSA | Pooled | rs2958127 | rs941207 | *NAB2* | ENSG00000166886.8 | protein_coding | iPSC_NPC | 7.02 |
| OSA | Pooled | rs2958127 | rs941207 | TCONS_00020445 | TCONS_00020445 | lincRNA | iPSC_NPC | 7.02 |
| OSA | Pooled | rs2958127 | rs941207 | TCONS_00020805 | TCONS_00020805 | lincRNA | iPSC_NPC | 7.02 |
| OSA | Pooled | rs2958127 | rs941207 | TCONS_00020806 | TCONS_00020806 | lincRNA | iPSC_NPC | 7.02 |
| OSA | Pooled | rs2958127 | rs941207 | TCONS_00020807 | TCONS_00020807 | lincRNA | iPSC_NPC | 7.02 |
| OSA | Pooled | rs2958127 | rs941207 | TCONS_00021326 | TCONS_00021326 | lincRNA | iPSC_NPC | 7.02 |
| OSA | Pooled | rs2958127 | rs941207 | TCONS_00021327 | TCONS_00021327 | lincRNA | iPSC_NPC | 7.02 |
| OSA | Pooled | rs2958127 | rs941207 | *TMEM194A* | ENSG00000166881.5 | protein_coding | iPSC_NPC | 7.02 |
| OSA | Pooled | rs2958127 | rs2255074 | *ESYT1* | ENSG00000139641.8 | protein_coding | iPSC_NPC | 6.99 |
| OSA | Pooled | rs2958127 | rs1107479 | *ESYT1* | ENSG00000139641.8 | protein_coding | iPSC_NPC | 6.99 |
| OSA | Pooled | rs2958127 | rs2958153 | *ZBTB39* | ENSG00000166860.2 | protein_coding | GCB | 6.98 |
| OSA | Pooled | rs2958127 | rs1107479 | *ATP5B* | ENSG00000110955.4 | protein_coding | hESC_HypothalamicNPC | 6.96 |
| OSA | Pooled | rs2958127 | rs1107479 | *SNORD59A* | ENSG00000207031.1 | snoRNA | hESC_HypothalamicNPC | 6.96 |
| OSA | Pooled | rs2958127 | rs1107479 | *U59B* | U59B | snoRNA | hESC_HypothalamicNPC | 6.96 |
| OSA | Pooled | rs2958127 | rs3214051 | *SDR9C7* | ENSG00000170426.1 | protein_coding | hFOB | 6.82 |
| OSA | Pooled | rs2958127 | rs941207 | *COQ10A* | ENSG00000135469.9 | protein_coding | hESC_HypothalamicNeuron | 6.81 |
| OSA | Pooled | rs2958127 | rs941207 | *ESYT1* | ENSG00000139641.8 | protein_coding | NaiveB | 6.8 |
| OSA | Pooled | rs2958127 | rs941207 | *RP11-603J24.6* | ENSG00000258345.1 | antisense | NaiveB | 6.8 |
| OSA | Pooled | rs2958127 | rs941207 | *ZC3H10* | ENSG00000135482.2 | protein_coding | NaiveB | 6.8 |
| OSA | Pooled | rs2958127 | rs2958153 | *ATP5B* | ENSG00000110955.4 | protein_coding | GCB | 6.75 |
| OSA | Pooled | rs2958127 | rs2958153 | *SNORD59A* | ENSG00000207031.1 | snoRNA | GCB | 6.75 |
| OSA | Pooled | rs2958127 | rs2958153 | *U59B* | U59B | snoRNA | GCB | 6.75 |
| OSA | Pooled | rs2958127 | rs941207 | *RBMS2* | ENSG00000076067.7 | protein_coding | hESC_HypothalamicNPC | 6.74 |
| OSA | Pooled | rs2958127 | rs2958153 | *NACA* | ENSG00000196531.6 | protein_coding | iPSC_Neurons | 6.71 |
| OSA | Pooled | rs2958127 | rs941207 | *STAT6* | ENSG00000166888.6 | protein_coding | iPSC_NPC | 6.71 |
| OSA | Pooled | rs2958127 | rs941207 | *RP11-603J24.6* | ENSG00000258345.1 | antisense | iPSC_NPC | 6.69 |
| OSA | Pooled | rs2958127 | rs2958153 | *ZBTB39* | ENSG00000166860.2 | protein_coding | hESC_HypothalamicNeuron | 6.68 |
| OSA | Pooled | rs2958127 | rs3214051 | *NACA* | ENSG00000196531.6 | protein_coding | GCB | 6.67 |
| OSA | Pooled | rs2958127 | rs2958153 | *BAZ2A* | ENSG00000076108.7 | protein_coding | iPSC_NPC | 6.65 |
| OSA | Pooled | rs2958127 | rs3214051 | *ZBTB39* | ENSG00000166860.2 | protein_coding | hESC_HypothalamicNPC | 6.56 |
| OSA | Pooled | rs2958127 | rs2255074 | *ZBTB39* | ENSG00000166860.2 | protein_coding | hESC_HypothalamicNeuron | 6.55 |
| OSA | Pooled | rs2958127 | rs1107479 | *ZBTB39* | ENSG00000166860.2 | protein_coding | hESC_HypothalamicNeuron | 6.55 |
| OSA | Pooled | rs2958127 | rs1107479 | *BAZ2A* | ENSG00000076108.7 | protein_coding | hESC_HypothalamicNPC | 6.55 |
| OSA | Pooled | rs2958127 | rs3214051 | *NACA* | ENSG00000196531.10 | protein_coding | Chondrocytes | 6.54 |
| OSA | Pooled | rs2958127 | rs3214051 | *U59B* | U59B | snoRNA | TFH | 6.51 |
| OSA | Pooled | rs2958127 | rs3214051 | *MYO1A* | ENSG00000166866.13 | protein_coding | Chondrocytes | 6.49 |
| OSA | Pooled | rs2958127 | rs3214051 | *ZBTB39* | ENSG00000166860.2 | protein_coding | iPSC_Hepatocytes | 6.46 |
| OSA | Pooled | rs2958127 | rs2255074 | *NACA* | ENSG00000196531.6 | protein_coding | SGBS_Diff | 6.46 |
| OSA | Pooled | rs2958127 | rs1107479 | *NACA* | ENSG00000196531.6 | protein_coding | SGBS_Diff | 6.46 |
| OSA | Pooled | rs2958127 | rs2255074 | *MIP* | ENSG00000135517.6 | protein_coding | iPSC_NPC | 6.44 |
| OSA | Pooled | rs2958127 | rs2255074 | *SPRYD4* | ENSG00000176422.10 | protein_coding | iPSC_NPC | 6.44 |
| OSA | Pooled | rs2958127 | rs1107479 | *MIP* | ENSG00000135517.6 | protein_coding | iPSC_NPC | 6.44 |

| **Supplementary Table 16.** Prioritized effector genes from variant to gene (V2G) mapping analysis. | | | | | | | | |
|---|---|---|---|---|---|---|---|---|
| **Trait** | **Discovery Study** | **Sentinel rsID** | **Proxy rsID** | **Gene Symbol** | **Gene Id** | **Gene Biotype** | **Cell** | **Contact Score (Chicago Score\|-log10(FitHiC2 FDR)** |
| OSA | Pooled | rs2958127 | rs1107479 | *SPRYD4* | ENSG00000176422.10 | protein_coding | iPSC_NPC | 6.44 |
| OSA | Pooled | rs2958127 | rs3214051 | *ATP5B* | ENSG00000110955.4 | protein_coding | NaiveT | 6.43 |
| OSA | Pooled | rs2958127 | rs3214051 | *SNORD59A* | ENSG00000207031.1 | snoRNA | NaiveT | 6.43 |
| OSA | Pooled | rs2958127 | rs3214051 | *U59B* | U59B | snoRNA | NaiveT | 6.43 |
| OSA | Pooled | rs2958127 | rs941207 | *ATP5B* | ENSG00000110955.4 | protein_coding | NaiveT | 6.4 |
| OSA | Pooled | rs2958127 | rs941207 | *U59B* | U59B | snoRNA | NaiveT | 6.4 |
| OSA | Pooled | rs2958127 | rs941207 | *SNORD59A* | ENSG00000207031.1 | snoRNA | NaiveT | 6.4 |
| OSA | Pooled | rs2958127 | rs2958153 | *BAZ2A* | ENSG00000076108.7 | protein_coding | TFH | 6.25 |
| OSA | Pooled | rs2958127 | rs2255074 | *BAZ2A* | ENSG00000076108.7 | protein_coding | iPSC_Neurons | 6.23 |
| OSA | Pooled | rs2958127 | rs1107479 | *BAZ2A* | ENSG00000076108.7 | protein_coding | iPSC_Neurons | 6.23 |
| OSA | Pooled | rs2958127 | rs2958153 | *ZBTB39* | ENSG00000166860.2 | protein_coding | hMSC_Osteoblasts | 6.17 |
| OSA | Pooled | rs2958127 | rs2958153 | *ATP5B* | ENSG00000110955.4 | protein_coding | hMSC_Adipocyte | 6.14 |
| OSA | Pooled | rs2958127 | rs2958153 | *SNORD59A* | ENSG00000207031.1 | snoRNA | hMSC_Adipocyte | 6.14 |
| OSA | Pooled | rs2958127 | rs2958153 | *U59B* | U59B | snoRNA | hMSC_Adipocyte | 6.14 |
| OSA | Pooled | rs2958127 | rs2255074 | *BAZ2A* | ENSG00000076108.7 | protein_coding | NaiveT | 6.13 |
| OSA | Pooled | rs2958127 | rs1107479 | *BAZ2A* | ENSG00000076108.7 | protein_coding | NaiveT | 6.13 |
| OSA | Pooled | rs2958127 | rs3214051 | *BAZ2A* | ENSG00000076108.7 | protein_coding | iPSC_NPC | 6.1 |
| OSA | Pooled | rs2958127 | rs2958153 | *ATP5B* | ENSG00000110955.4 | protein_coding | NaiveT | 6.07 |
| OSA | Pooled | rs2958127 | rs2958153 | *SNORD59A* | ENSG00000207031.1 | snoRNA | NaiveT | 6.07 |
| OSA | Pooled | rs2958127 | rs2958153 | *U59B* | U59B | snoRNA | NaiveT | 6.07 |
| OSA | Pooled | rs2958127 | rs2255074 | *NACA* | ENSG00000196531.6 | protein_coding | iPSC_NPC | 6.02 |
| OSA | Pooled | rs2958127 | rs1107479 | *NACA* | ENSG00000196531.6 | protein_coding | iPSC_NPC | 6.02 |
| OSA | Pooled | rs2958127 | rs10747769 | *ATP5B* | ENSG00000110955.4 | protein_coding | TFH | 5.97 |
| OSA | Pooled | rs2958127 | rs10747769 | *SNORD59A* | ENSG00000207031.1 | snoRNA | TFH | 5.97 |
| OSA | Pooled | rs2958127 | rs10747769 | *U59B* | U59B | snoRNA | TFH | 5.97 |
| OSA | Pooled | rs2958127 | rs2255074 | *ZBTB39* | ENSG00000166860.2 | protein_coding | hMSC_Osteoblasts | 5.95 |
| OSA | Pooled | rs2958127 | rs1107479 | *ZBTB39* | ENSG00000166860.2 | protein_coding | hMSC_Osteoblasts | 5.95 |
| OSA | Pooled | rs2958127 | rs941207 | *ATP5B* | ENSG00000110955.4 | protein_coding | hESC_HypothalamicNPC | 5.94 |
| OSA | Pooled | rs2958127 | rs941207 | *SNORD59A* | ENSG00000207031.1 | snoRNA | hESC_HypothalamicNPC | 5.94 |
| OSA | Pooled | rs2958127 | rs941207 | *U59B* | U59B | snoRNA | hESC_HypothalamicNPC | 5.94 |
| OSA | Pooled | rs2958127 | rs2958153 | *BAZ2A* | ENSG00000076108.7 | protein_coding | iPSC_Neurons | 5.94 |
| OSA | Pooled | rs2958127 | rs3214051 | *ATP5B* | ENSG00000110955.4 | protein_coding | HEPG2 | 5.9 |
| OSA | Pooled | rs2958127 | rs3214051 | *SNORD59A* | ENSG00000207031.1 | snoRNA | HEPG2 | 5.9 |
| OSA | Pooled | rs2958127 | rs3214051 | *U59B* | U59B | snoRNA | HEPG2 | 5.9 |
| OSA | Pooled | rs2958127 | rs941207 | *ATP5B* | ENSG00000110955.4 | protein_coding | hMSC_Adipocyte | 5.89 |
| OSA | Pooled | rs2958127 | rs941207 | *SNORD59A* | ENSG00000207031.1 | snoRNA | hMSC_Adipocyte | 5.89 |
| OSA | Pooled | rs2958127 | rs941207 | *U59B* | U59B | snoRNA | hMSC_Adipocyte | 5.89 |
| OSA | Pooled | rs2958127 | rs941207 | *NACA* | ENSG00000196531.6 | protein_coding | GCB | 5.87 |
| OSA | Pooled | rs2958127 | rs2255074 | *NAB2* | ENSG00000166886.8 | protein_coding | NaiveB | 5.87 |
| OSA | Pooled | rs2958127 | rs2255074 | TCONS_00020445 | TCONS_00020445 | lincRNA | NaiveB | 5.87 |
| OSA | Pooled | rs2958127 | rs2255074 | TCONS_00020805 | TCONS_00020805 | lincRNA | NaiveB | 5.87 |
| OSA | Pooled | rs2958127 | rs2255074 | TCONS_00020806 | TCONS_00020806 | lincRNA | NaiveB | 5.87 |
| OSA | Pooled | rs2958127 | rs2255074 | TCONS_00020807 | TCONS_00020807 | lincRNA | NaiveB | 5.87 |
| OSA | Pooled | rs2958127 | rs2255074 | TCONS_00021326 | TCONS_00021326 | lincRNA | NaiveB | 5.87 |
| OSA | Pooled | rs2958127 | rs2255074 | TCONS_00021327 | TCONS_00021327 | lincRNA | NaiveB | 5.87 |
| OSA | Pooled | rs2958127 | rs2255074 | *TMEM194A* | ENSG00000166881.5 | protein_coding | NaiveB | 5.87 |
| OSA | Pooled | rs2958127 | rs1107479 | *NAB2* | ENSG00000166886.8 | protein_coding | NaiveB | 5.87 |
| OSA | Pooled | rs2958127 | rs1107479 | TCONS_00020445 | TCONS_00020445 | lincRNA | NaiveB | 5.87 |
| OSA | Pooled | rs2958127 | rs1107479 | TCONS_00020805 | TCONS_00020805 | lincRNA | NaiveB | 5.87 |
| OSA | Pooled | rs2958127 | rs1107479 | TCONS_00020806 | TCONS_00020806 | lincRNA | NaiveB | 5.87 |
| OSA | Pooled | rs2958127 | rs1107479 | TCONS_00020807 | TCONS_00020807 | lincRNA | NaiveB | 5.87 |
| OSA | Pooled | rs2958127 | rs1107479 | TCONS_00021326 | TCONS_00021326 | lincRNA | NaiveB | 5.87 |
| OSA | Pooled | rs2958127 | rs1107479 | TCONS_00021327 | TCONS_00021327 | lincRNA | NaiveB | 5.87 |
| OSA | Pooled | rs2958127 | rs1107479 | *TMEM194A* | ENSG00000166881.5 | protein_coding | NaiveB | 5.87 |
| OSA | Pooled | rs2958127 | rs2958153 | TCONS_00020445 | TCONS_00020445 | lincRNA | iPSC_NPC | 5.8 |
| OSA | Pooled | rs2958127 | rs2958153 | TCONS_00020807 | TCONS_00020807 | lincRNA | iPSC_NPC | 5.8 |
| OSA | Pooled | rs2958127 | rs941207 | *BAZ2A* | ENSG00000076108.7 | protein_coding | hESC_HypothalamicNPC | 5.79 |
| OSA | Pooled | rs2958127 | rs941207 | *BAZ2A* | ENSG00000076108.7 | protein_coding | hMSC_Osteoblasts | 5.77 |
| OSA | Pooled | rs2958127 | rs2255074 | *NACA* | ENSG00000196531.6 | protein_coding | GCB | 5.76 |
| OSA | Pooled | rs2958127 | rs1107479 | *NACA* | ENSG00000196531.6 | protein_coding | GCB | 5.76 |
| OSA | Pooled | rs2958127 | rs2958153 | *BAZ2A* | ENSG00000076108.7 | protein_coding | NaiveB | 5.7 |
| OSA | Pooled | rs2958127 | rs2255074 | *RBMS2* | ENSG00000076067.7 | protein_coding | hESC_HypothalamicNeuron | 5.68 |
| OSA | Pooled | rs2958127 | rs1107479 | *RBMS2* | ENSG00000076067.7 | protein_coding | hESC_HypothalamicNeuron | 5.68 |
| OSA | Pooled | rs2958127 | rs2255074 | *RBMS2* | ENSG00000076067.7 | protein_coding | NaiveT | 5.64 |
| OSA | Pooled | rs2958127 | rs1107479 | *RBMS2* | ENSG00000076067.7 | protein_coding | NaiveT | 5.64 |
| OSA | Pooled | rs2958127 | rs2255074 | *RBMS2* | ENSG00000076067.7 | protein_coding | SGBS_Diff | 5.63 |
| OSA | Pooled | rs2958127 | rs1107479 | *RBMS2* | ENSG00000076067.7 | protein_coding | SGBS_Diff | 5.63 |
| OSA | Pooled | rs2958127 | rs2958153 | *BAZ2A* | ENSG00000076108.7 | protein_coding | SGBS_Diff | 5.57 |
| OSA | Pooled | rs2958127 | rs941207 | *NACA* | ENSG00000196531.6 | protein_coding | hESC_HypothalamicNeuron | 5.55 |
| OSA | Pooled | rs2958127 | rs2958153 | *ESYT1* | ENSG00000139641.8 | protein_coding | iPSC_Neurons | 5.51 |
| OSA | Pooled | rs2958127 | rs2958153 | *ZC3H10* | ENSG00000135482.2 | protein_coding | iPSC_Neurons | 5.51 |
| OSA | Pooled | rs2958127 | rs3214051 | *NACA* | ENSG00000196531.6 | protein_coding | hESC_HypothalamicNPC | 5.47 |
| OSA | Pooled | rs2958127 | rs2255074 | *ATP5B* | ENSG00000110955.4 | protein_coding | iPSC_Neurons | 5.42 |
| OSA | Pooled | rs2958127 | rs2255074 | *U59B* | U59B | snoRNA | iPSC_Neurons | 5.42 |
| OSA | Pooled | rs2958127 | rs1107479 | *ATP5B* | ENSG00000110955.4 | protein_coding | iPSC_Neurons | 5.42 |
| OSA | Pooled | rs2958127 | rs1107479 | *U59B* | U59B | snoRNA | iPSC_Neurons | 5.42 |
| OSA | Pooled | rs2958127 | rs2255074 | *SNORD59A* | ENSG00000207031.1 | snoRNA | iPSC_Neurons | 5.42 |
| OSA | Pooled | rs2958127 | rs1107479 | *SNORD59A* | ENSG00000207031.1 | snoRNA | iPSC_Neurons | 5.42 |
| OSA | Pooled | rs2958127 | rs941207 | *BAZ2A* | ENSG00000076108.7 | protein_coding | iPSC_Neurons | 5.39 |
| OSA | Pooled | rs2958127 | rs2958153 | *ZBTB39* | ENSG00000166860.2 | protein_coding | iPSC_NPC | 5.39 |
| OSA | Pooled | rs2958127 | rs2958153 | *ATP5B* | ENSG00000110955.4 | protein_coding | TFH | 5.39 |
| OSA | Pooled | rs2958127 | rs2958153 | *SNORD59A* | ENSG00000207031.1 | snoRNA | TFH | 5.39 |
| OSA | Pooled | rs2958127 | rs941207 | *RBMS2* | ENSG00000076067.7 | protein_coding | NaiveT | 5.38 |
| OSA | Pooled | rs2958127 | rs7980835 | *ATP5B* | ENSG00000110955.4 | protein_coding | NaiveT | 5.3 |
| OSA | Pooled | rs2958127 | rs7980835 | *SNORD59A* | ENSG00000207031.1 | snoRNA | NaiveT | 5.3 |

| Supplementary Table 16. Prioritized effector genes from variant to gene (V2G) mapping analysis. | | | | | | | | |
|---|---|---|---|---|---|---|---|---|
| Trait | Discovery Study | Sentinel rsID | Proxy rsID | Gene Symbol | Gene Id | Gene Biotype | Cell | Contact Score (Chicago Score\|-log10(FitHiC2 FDR) |
| OSA | Pooled | rs2958127 | rs7980835 | *U59B* | U59B | snoRNA | NaiveT | 5.3 |
| OSA | Pooled | rs2958127 | rs10747769 | *ATP5B* | ENSG00000110955.4 | protein_coding | NaiveT | 5.3 |
| OSA | Pooled | rs2958127 | rs10747769 | *SNORD59A* | ENSG00000207031.1 | snoRNA | NaiveT | 5.3 |
| OSA | Pooled | rs2958127 | rs10747769 | *U59B* | U59B | snoRNA | NaiveT | 5.3 |
| OSA | Pooled | rs2958127 | rs941207 | *U59B* | U59B | snoRNA | NaiveB | 5.28 |
| OSA | Pooled | rs2958127 | rs2958153 | *ATP5B* | ENSG00000110955.4 | protein_coding | iPSC_Neurons | 5.27 |
| OSA | Pooled | rs2958127 | rs2958153 | *SNORD59A* | ENSG00000207031.1 | snoRNA | iPSC_Neurons | 5.27 |
| OSA | Pooled | rs2958127 | rs2958153 | *U59B* | U59B | snoRNA | iPSC_Neurons | 5.27 |
| OSA | Pooled | rs2958127 | rs941207 | *APOF* | ENSG00000175336.8 | protein_coding | iPSC_NPC | 5.25 |
| OSA | Pooled | rs2958127 | rs10747769 | *NACA* | ENSG00000196531.6 | protein_coding | TFH | 5.25 |
| OSA | Pooled | rs2958127 | rs2255074 | *ATP5B* | ENSG00000110955.4 | protein_coding | HEPG2 | 5.2 |
| OSA | Pooled | rs2958127 | rs2255074 | *SNORD59A* | ENSG00000207031.1 | snoRNA | HEPG2 | 5.2 |
| OSA | Pooled | rs2958127 | rs2255074 | *U59B* | U59B | snoRNA | HEPG2 | 5.2 |
| OSA | Pooled | rs2958127 | rs1107479 | *ATP5B* | ENSG00000110955.4 | protein_coding | HEPG2 | 5.2 |
| OSA | Pooled | rs2958127 | rs1107479 | *SNORD59A* | ENSG00000207031.1 | snoRNA | HEPG2 | 5.2 |
| OSA | Pooled | rs2958127 | rs1107479 | *U59B* | U59B | snoRNA | HEPG2 | 5.2 |
| OSA | Pooled | rs2958127 | rs941207 | *NACA* | ENSG00000196531.6 | protein_coding | hMSC_Osteoblasts | 5.18 |
| OSA | Pooled | rs2958127 | rs941207 | *ATP5B* | ENSG00000110955.4 | protein_coding | iPSC_NPC | 5.12 |
| OSA | Pooled | rs2958127 | rs941207 | *SNORD59A* | ENSG00000207031.1 | snoRNA | iPSC_NPC | 5.12 |
| OSA | Pooled | rs2958127 | rs941207 | *U59B* | U59B | snoRNA | iPSC_NPC | 5.12 |
| OSA | Pooled | rs2958127 | rs3214051 | *ZBTB39* | ENSG00000166860.2 | protein_coding | TFH | 5.04 |
| OSA | Pooled | rs2958127 | rs941207 | *MYL6* | ENSG00000092841.14 | protein_coding | iPSC_NPC | 5.03 |
| OSA | Pooled | rs2958127 | rs941207 | *MYL6B* | ENSG00000196465.6 | protein_coding | iPSC_NPC | 5.03 |
| OSA | Pooled | rs2958127 | rs941207 | *RP11-603J24.14* | ENSG00000257809.1 | antisense | iPSC_NPC | 5.03 |
| OSA | Pooled | rs2958127 | rs2255074 | *NACA* | ENSG00000196531.6 | protein_coding | NaiveB | 5.02 |
| OSA | Pooled | rs2958127 | rs1107479 | *NACA* | ENSG00000196531.6 | protein_coding | NaiveB | 5.02 |
| OSA | Pooled | rs2958127 | rs2958153 | *ATP5B* | ENSG00000110955.4 | protein_coding | HEPG2 | 5.01 |
| OSA | Pooled | rs2958127 | rs2958153 | *SNORD59A* | ENSG00000207031.1 | snoRNA | HEPG2 | 5.01 |
| OSA | Pooled | rs2958127 | rs2958153 | *U59B* | U59B | snoRNA | HEPG2 | 5.01 |
| OSA | Pooled | rs9831061 | rs869401 | *DGKG* | ENSG00000058866.15 | protein_coding | Chondrocytes | 17.08 |
| OSA | Pooled | rs9831061 | rs869399 | *DGKG* | ENSG00000058866.15 | protein_coding | Chondrocytes | 17.08 |
| OSA | Pooled | rs9831061 | rs869401 | *DNAJB11* | ENSG00000090520.6 | protein_coding | TFH | 13.41 |
| OSA | Pooled | rs9831061 | rs869401 | *TBCCD1* | ENSG00000113838.8 | protein_coding | TFH | 13.41 |
| OSA | Pooled | rs9831061 | rs869399 | *DNAJB11* | ENSG00000090520.6 | protein_coding | TFH | 13.41 |
| OSA | Pooled | rs9831061 | rs869399 | *TBCCD1* | ENSG00000113838.8 | protein_coding | TFH | 13.41 |
| OSA | Pooled | rs9831061 | rs869401 | *CRYGS* | ENSG00000213139.3 | protein_coding | NaiveB | 10.71 |
| OSA | Pooled | rs9831061 | rs869399 | *CRYGS* | ENSG00000213139.3 | protein_coding | NaiveB | 10.71 |
| OSA | Pooled | rs9831061 | rs869401 | *CRYGS* | ENSG00000213139.3 | protein_coding | GCB | 9.72 |
| OSA | Pooled | rs9831061 | rs869399 | *CRYGS* | ENSG00000213139.3 | protein_coding | GCB | 9.72 |
| OSA | Pooled | rs9831061 | rs869399 | *DNAJB11* | ENSG00000090520.6 | protein_coding | GCB | 8.91 |
| OSA | Pooled | rs9831061 | rs869399 | *TBCCD1* | ENSG00000113838.8 | protein_coding | GCB | 8.91 |
| OSA | Pooled | rs9831061 | rs869401 | *DNAJB11* | ENSG00000090520.6 | protein_coding | GCB | 8.91 |
| OSA | Pooled | rs9831061 | rs869401 | *TBCCD1* | ENSG00000113838.8 | protein_coding | GCB | 8.91 |
| OSA | Pooled | rs9831061 | rs869399 | *CRYGS* | ENSG00000213139.3 | protein_coding | iPSC_NPC | 8.34 |
| OSA | Pooled | rs9831061 | rs869399 | *DNAJB11* | ENSG00000090520.6 | protein_coding | iPSC_NPC | 7.35 |
| OSA | Pooled | rs9831061 | rs869399 | *TBCCD1* | ENSG00000113838.8 | protein_coding | iPSC_NPC | 7.35 |
| OSA | Pooled | rs9831061 | rs869401 | *CRYGS* | ENSG00000213139.3 | protein_coding | hMSC_Osteoblasts | 7.25 |
| OSA | Pooled | rs9831061 | rs869399 | *CRYGS* | ENSG00000213139.3 | protein_coding | hMSC_Osteoblasts | 7.25 |
| OSA | Pooled | rs9831061 | rs869401 | *EIF4A2* | ENSG00000156976.10 | protein_coding | TFH | 6.47 |
| OSA | Pooled | rs9831061 | rs869401 | *hsa-mir-1248* | hsa-mir-1248 | lincRNA | TFH | 6.47 |
| OSA | Pooled | rs9831061 | rs869401 | *RP11-573D15.9* | ENSG00000263826.1 | antisense | TFH | 6.47 |
| OSA | Pooled | rs9831061 | rs869401 | *SNORA63* | ENSG00000200418.1 | snoRNA | TFH | 6.47 |
| OSA | Pooled | rs9831061 | rs869401 | *SNORA81* | ENSG00000221420.2 | snoRNA | TFH | 6.47 |
| OSA | Pooled | rs9831061 | rs869401 | *SNORD2* | ENSG00000238942.1 | snoRNA | TFH | 6.47 |
| OSA | Pooled | rs9831061 | rs869401 | *snR39B* | snR39B | snoRNA | TFH | 6.47 |
| OSA | Pooled | rs9831061 | rs869399 | *EIF4A2* | ENSG00000156976.10 | protein_coding | TFH | 6.47 |
| OSA | Pooled | rs9831061 | rs869399 | *hsa-mir-1248* | hsa-mir-1248 | lincRNA | TFH | 6.47 |
| OSA | Pooled | rs9831061 | rs869399 | *RP11-573D15.9* | ENSG00000263826.1 | antisense | TFH | 6.47 |
| OSA | Pooled | rs9831061 | rs869399 | *SNORA63* | ENSG00000200418.1 | snoRNA | TFH | 6.47 |
| OSA | Pooled | rs9831061 | rs869399 | *SNORA81* | ENSG00000221420.2 | snoRNA | TFH | 6.47 |
| OSA | Pooled | rs9831061 | rs869399 | *SNORD2* | ENSG00000238942.1 | snoRNA | TFH | 6.47 |
| OSA | Pooled | rs9831061 | rs869399 | *snR39B* | snR39B | snoRNA | TFH | 6.47 |
| OSA | Pooled | rs9831061 | rs869401 | *CRYGS* | ENSG00000213139.3 | protein_coding | hESC_HypothalamicNeuron | 6.38 |
| OSA | Pooled | rs9831061 | rs869399 | *CRYGS* | ENSG00000213139.3 | protein_coding | hESC_HypothalamicNeuron | 6.38 |
| OSA | Pooled | rs9831061 | rs869401 | *CRYGS* | ENSG00000213139.3 | protein_coding | TFH | 6.25 |
| OSA | Pooled | rs9831061 | rs869399 | *CRYGS* | ENSG00000213139.3 | protein_coding | TFH | 6.25 |
| OSA | Pooled | rs9831061 | rs869401 | *DNAJB11* | ENSG00000090520.6 | protein_coding | hESC_HypothalamicNeuron | 5.56 |
| OSA | Pooled | rs9831061 | rs869401 | *TBCCD1* | ENSG00000113838.8 | protein_coding | hESC_HypothalamicNeuron | 5.56 |
| OSA | Pooled | rs9831061 | rs869399 | *DNAJB11* | ENSG00000090520.6 | protein_coding | hESC_HypothalamicNeuron | 5.56 |
| OSA | Pooled | rs9831061 | rs869399 | *TBCCD1* | ENSG00000113838.8 | protein_coding | hESC_HypothalamicNeuron | 5.56 |
| OSA | Pooled | rs9831061 | rs869399 | *RP11-443P15.2* | ENSG00000171658.4 | transcribed_unprocessed_pseudogene | iPSC_Hepatocytes | 5.54 |
| OSA | Pooled | rs9831061 | rs869399 | TCONS_l2_00019043 | TCONS_l2_00019043 | lincRNA | iPSC_Hepatocytes | 5.54 |
| OSA | Pooled | rs9831061 | rs869401 | *DNAJB11* | ENSG00000090520.6 | protein_coding | NaiveB | 5.42 |
| OSA | Pooled | rs9831061 | rs869401 | *TBCCD1* | ENSG00000113838.8 | protein_coding | NaiveB | 5.42 |
| OSA | Pooled | rs9831061 | rs869399 | *DNAJB11* | ENSG00000090520.6 | protein_coding | NaiveB | 5.42 |
| OSA | Pooled | rs9831061 | rs869399 | *TBCCD1* | ENSG00000113838.8 | protein_coding | NaiveB | 5.42 |
| OSA | Pooled | rs9831061 | rs869399 | *RP11-48F14.1* | ENSG00000224406.1 | lincRNA | GCB | 5.18 |
| OSA | Pooled | rs9831061 | rs869399 | TCONS_00005404 | TCONS_00005404 | lincRNA | GCB | 5.18 |
| OSA | Pooled | rs9831061 | rs869399 | TCONS_00006344 | TCONS_00006344 | lincRNA | GCB | 5.18 |
| OSA | Pooled | rs9831061 | rs869399 | TCONS_00006725 | TCONS_00006725 | lincRNA | GCB | 5.18 |
| OSA | Pooled | rs9831061 | rs869401 | *RFC4* | ENSG00000163918.6 | protein_coding | NaiveB | 5.17 |
| OSA | Pooled | rs9831061 | rs869401 | *RP11-573D15.3* | ENSG00000231724.1 | antisense | NaiveB | 5.17 |
| OSA | Pooled | rs9831061 | rs869399 | *RFC4* | ENSG00000163918.6 | protein_coding | NaiveB | 5.17 |
| OSA | Pooled | rs9831061 | rs869399 | *RP11-573D15.3* | ENSG00000231724.1 | antisense | NaiveB | 5.17 |
| avgSaO2 | Current | rs4759254 | rs1107479 | *SNORD59A* | ENSG00000207031.1 | snoRNA | Chondrocytes | 14.19 |

**Supplementary Table 16.** Prioritized effector genes from variant to gene (V2G) mapping analysis.

| Trait | Discovery Study | Sentinel rsID | Proxy rsID | Gene Symbol | Gene Id | Gene Biotype | Cell | Contact Score (Chicago Score\|-log10(FitHiC2 FDR) |
|---|---|---|---|---|---|---|---|---|
| avgSaO2 | Current | rs4759254 | rs2255074 | *SNORD59A* | ENSG00000207031.1 | snoRNA | Chondrocytes | 14.19 |
| avgSaO2 | Current | rs4759254 | rs1107479 | *ATP5F1B* | ENSG00000110955.9 | protein_coding | Chondrocytes | 14.19 |
| avgSaO2 | Current | rs4759254 | rs2255074 | *ATP5F1B* | ENSG00000110955.9 | protein_coding | Chondrocytes | 14.19 |
| avgSaO2 | Current | rs4759254 | rs1107479 | *ZBTB39* | ENSG00000166860.3 | protein_coding | Chondrocytes | 7.02 |
| avgSaO2 | Current | rs4759254 | rs2255074 | *ZBTB39* | ENSG00000166860.3 | protein_coding | Chondrocytes | 7.02 |
| avgSaO2 | Current | rs4759254 | rs3214051 | *NACA* | ENSG00000196531.10 | protein_coding | Chondrocytes | 6.54 |
| avgSaO2 | Current | rs4759254 | rs3214051 | *MYO1A* | ENSG00000166866.13 | protein_coding | Chondrocytes | 6.49 |

**Supplementary Table 17.** Summary of results for the multivariate genotype-phenotype (MGP) in mice on PLS axis 1.

| Cell Group | Gene List | PLS Magnitude | Procrustes Distance | Mean Displacement | Maximum Displacement | Variance Explained | P-value (Random Genes test) | P-value (Permutation test) | Effect size (Random Genes Test) | Effect size (Permutation Test) |
|---|---|---|---|---|---|---|---|---|---|---|
| **CHONDROCYTES** | Arpc5l, Baz2a, Cnnm2, Cenpp, Dgkg, Exoc3l2, Entrep1, Fbxo46, Gipr, Glis2, Golga1, Gpr182, Gpr4, Irx5, Ippk, Hspa5, Kansl1, Klc3, Hsd17b6, Mapt, Mvb12b, Lrrc37a, Myo1a, Naca, Nt5c2, Nr6a1, Olfml2a, Nol8, Nron, Prim1, Opa3, Ppm1n, Plekhm1, Rbms2, Rpl35, Sfxn2, Thada, Spag17, Vasp, Rpe, Zbtb39, Wdr38, Zfp36l2, Wbp1l, Slc3a1 | 13.5408 | 0.0027 | 0.074 | 0.1069 | 0.106 | 7.96E-28 | ***0.0026*** | 0.88 | 2.796 |
| HEPG2 | Baz2a, Sppl2c, Zbtb39 | 6.283 | 0.0017 | 0.0438 | 0.1052 | 0.0653 | 6.94E-56 | **0.0189** | 0.37 | 2.076 |
| Osteoblasts | Baz2a, Zbtb39, Naca, Aspn, Gpr182, Ptges3, Nol8, Crygs | 10.8924 | 0.0017 | 0.0437 | 0.0568 | 0.0661 | 8.46E-40 | **0.0276** | 0.63 | 1.917 |
| HESC_HYPOTHALAMICNPC | Gfi1, Gpr182, Naca, Ecm2, Baz2a, Zbtb39, Rbms2, Tjp2 | 12.1952 | 0.0013 | 0.0343 | 0.0484 | 0.0524 | 2.33E-49 | 0.37 | 0.46 | 0.332 |
| hESC_HypothalamicNeuron | Nr6a1, Nrxn3, Ptges3, Naca, Rbms2, Snord59a, Tbccd1, Wdr38, Gria1, Zbtb39, Mapt, Baz2a, Cast, Coq10a, Crygs, Entrep1, Dnajb11 | 13.7783 | 0.0016 | 0.0432 | 0.0572 | 0.064 | 0.0042 | 0.3794 | 0.23 | 0.307 |
| TFH | Baz2a, Eml2, Eif4a2, Dnajb11, Zbtb39, Ptges3, Naca, Stat6, Crygs, Tbccd1 | 13.13 | 0.0014 | 0.0376 | 0.0504 | 0.0554 | 3.69E-53 | 0.4081 | 0.4 | 0.232 |
| IPSC_NEURONS | Naca, Gria1, Esyt1, Baz2a, Acmsd, Ptges3, Rbms2, Zbtb39, Zc3h10 | 15.823 | 0.0011 | 0.0299 | 0.044 | 0.0453 | 5.53E-55 | 0.4142 | 0.42 | 0.217 |
| IPSC_NPC | Myl6, Nab2, Mip, Lrp1, Naca, Myl6b, Ptges3, Prim1, Olfml2a, Esyt1, Crygs, Ankrd52, Baz2a, Dnajb11, Spata32, Pou3f2, Rbms2, Spryd4, Zc3h10, Stat6, Tbccd1, Apof, Gpr182, Zbtb39, Hsd17b6, Isl2, Haao | 24.6934 | 0.0012 | 0.0326 | 0.0447 | 0.0493 | 1.72E-60 | 0.429 | 0.45 | 0.179 |
| NAIVEB | Dnajb11, Esyt1, Crygs, Baz2a, Nab2, Naca, Gpr182, Ptges3, Rfc4, Stat6, Tbccd1, Zc3h10, Zbtb39 | 16.8429 | 0.0013 | 0.0342 | 0.0465 | 0.0504 | 1.26E-59 | 0.4353 | 0.37 | 0.163 |
| SGBS_DIFF | Ptges3, Gria1, Naca, Gpr182, Baz2a, Rbms2, Stat6, Zbtb39 | 15.0828 | 0.0012 | 0.0305 | 0.0457 | 0.0455 | 4.43E-64 | 0.4437 | 0.3 | 0.142 |
| Adipocytes | Naca, Baz2a, Gpr182, Rbms2, Ptges3, Zbtb39 | 14.5473 | 0.0011 | 0.0288 | 0.0426 | 0.0432 | 1.69E-65 | 0.4496 | 0.3 | 0.127 |
| NAIVET | Rbms2, Baz2a, Stat6, Ptges3, Naca, Zbtb39 | 14.5345 | 0.0011 | 0.0285 | 0.0429 | 0.0429 | 2.71E-62 | 0.4694 | 0.31 | 0.077 |
| IPSC_HEPATOCYTES | Ptges3, Naca, Gpr182, As3mt, Etfa, Baz2a, Zbtb39 | 12.885 | 0.0012 | 0.0317 | 0.0473 | 0.0477 | 2.31E-59 | 0.5181 | 0.32 | 0.045 |
| HFOB | Coq10a, Gpr182, Nab2, Ptges3, Nxph4, Sdr9c7, Baz2a, Zbtb39 | 16.6155 | 0.001 | 0.0277 | 0.0411 | 0.0413 | 8.20E-61 | 0.6093 | 0.28 | 0.277 |
| GCB | Hspa5, Baz2a, Glmn, Crygs, Dnajb11, Mgat5, Naca, Ppp6c, Ptges3, Nab2, Rpap2, Rabepk, Zbtb39, Tbccd1, Tmem163 | 11.8551 | 0.0017 | 0.0455 | 0.0676 | 0.066 | 3.94E-73 | 0.6116 | 0.23 | 0.283 |

**Supplementary Table 18. Gene loadings on PLS1 for mouse MGP analyses by cell-type gene list.**

| Gene | Chondrocytes | TFH | GCB | HEPG2 | HESC_ Hypothalamic_ NPC | HFOB | Osteoblasts | IPSC_ Hepatocytes | IPSC_NPC | IPSC_ Neurons | SGBS_ DIFF | NAIVEB | NAIVET | HMSC_ Adipocyte | hESC_ HypothalamicNeuron |
|---|---|---|---|---|---|---|---|---|---|---|---|---|---|---|---|
| Lrrc37a | 0.3232 | NA | NA | NA | NA | NA | NA | NA | NA | NA | NA | NA | NA | NA | NA |
| Plekhm1 | 0.3219 | NA | NA | NA | NA | NA | NA | NA | NA | NA | NA | NA | NA | NA | NA |
| Mapt | 0.3048 | NA | NA | NA | NA | NA | NA | NA | NA | NA | NA | NA | NA | NA | 0.0535 |
| Kansl1 | 0.3001 | NA | NA | NA | NA | NA | NA | NA | NA | NA | NA | NA | NA | NA | NA |
| Arpc5l | 0.2515 | NA | NA | NA | NA | NA | NA | NA | NA | NA | NA | NA | NA | NA | NA |
| Nr6a1 | 0.2515 | NA | NA | NA | NA | NA | NA | NA | NA | NA | NA | NA | NA | NA | 0.0879 |
| Olfml2a | 0.2515 | NA | NA | NA | NA | NA | NA | NA | 0.0148 | NA | NA | NA | NA | NA | NA |
| Rpl35 | 0.2515 | NA | NA | NA | NA | NA | NA | NA | NA | NA | NA | NA | NA | NA | NA |
| Wdr38 | 0.2515 | NA | NA | NA | NA | NA | NA | NA | NA | NA | NA | NA | NA | NA | 0.0879 |
| Golga1 | 0.2513 | NA | NA | NA | NA | NA | NA | NA | NA | NA | NA | NA | NA | NA | NA |
| Gpr4 | 0.2436 | NA | NA | NA | NA | NA | NA | NA | NA | NA | NA | NA | NA | NA | NA |
| Opa3 | 0.2432 | NA | NA | NA | NA | NA | NA | NA | NA | NA | NA | NA | NA | NA | NA |
| Vasp | 0.2432 | NA | NA | NA | NA | NA | NA | NA | NA | NA | NA | NA | NA | NA | NA |
| Ppm1n | 0.243 | NA | NA | NA | NA | NA | NA | NA | NA | NA | NA | NA | NA | NA | NA |
| Gipr | 0.2421 | NA | NA | NA | NA | NA | NA | NA | NA | NA | NA | NA | NA | NA | NA |
| Fbxo46 | 0.2407 | NA | NA | NA | NA | NA | NA | NA | NA | NA | NA | NA | NA | NA | NA |
| Klc3 | 0.2341 | NA | NA | NA | NA | NA | NA | NA | NA | NA | NA | NA | NA | NA | NA |
| Exoc3l2 | 0.2323 | NA | NA | NA | NA | NA | NA | NA | NA | NA | NA | NA | NA | NA | NA |
| Wbp1l | 0.2165 | NA | NA | NA | NA | NA | NA | NA | NA | NA | NA | NA | NA | NA | NA |
| Sfxn2 | 0.2159 | NA | NA | NA | NA | NA | NA | NA | NA | NA | NA | NA | NA | NA | NA |
| Cnnm2 | 0.2141 | NA | NA | NA | NA | NA | NA | NA | NA | NA | NA | NA | NA | NA | NA |
| Nt5c2 | 0.2136 | NA | NA | NA | NA | NA | NA | NA | NA | NA | NA | NA | NA | NA | NA |
| Hspa5 | 0.2118 | NA | 0.243 | NA | NA | NA | NA | NA | NA | NA | NA | NA | NA | NA | NA |
| Mvb12b | 0.2011 | NA | NA | NA | NA | NA | NA | NA | NA | NA | NA | NA | NA | NA | NA |
| Nron | 0.2011 | NA | NA | NA | NA | NA | NA | NA | NA | NA | NA | NA | NA | NA | NA |
| Nol8 | 0.1914 | NA | NA | NA | NA | NA | 0.424 | NA | NA | NA | NA | NA | NA | NA | NA |
| Cenpp | 0.1913 | NA | NA | NA | NA | NA | NA | NA | NA | NA | NA | NA | NA | NA | NA |
| Ippk | 0.1912 | NA | NA | NA | NA | NA | NA | NA | NA | NA | NA | NA | NA | NA | NA |
| Hsd17b6 | 0.1497 | NA | NA | NA | NA | NA | NA | NA | 0.2968 | NA | NA | NA | NA | NA | NA |
| Naca | 0.1495 | 0.468 | 0.5229 | NA | 0.5772 | NA | 0.4827 | 0.5942 | 0.2978 | 0.45 | 0.5202 | 0.4118 | 0.5072 | 0.5455 | 0.5232 |
| Prim1 | 0.1494 | NA | NA | NA | NA | NA | NA | NA | 0.2979 | NA | NA | NA | NA | NA | NA |
| Rbms2 | 0.1494 | NA | NA | NA | 0.5718 | NA | NA | NA | 0.2957 | 0.4484 | 0.5154 | 0.4077 | 0.5028 | 0.5417 | 0.5195 |
| Baz2a | 0.1493 | 0.4628 | 0.517 | 0.3086 | 0.5726 | 0.4353 | 0.4775 | 0.5895 | 0.296 | 0.4488 | 0.5162 | 0.4083 | 0.5035 | 0.5425 | 0.5203 |
| Myo1a | 0.1227 | NA | NA | NA | NA | NA | NA | NA | NA | NA | NA | NA | NA | NA | NA |
| Gpr182 | 0.1204 | NA | NA | NA | 0.5199 | 0.4287 | 0.461 | 0.5347 | 0.2546 | NA | 0.474 | 0.3712 | 0.463 | 0.4775 | NA |
| Zbtb39 | 0.1204 | 0.4148 | 0.4781 | 0.3019 | 0.5199 | 0.4287 | 0.461 | 0.5347 | 0.2546 | 0.3622 | 0.474 | 0.3712 | 0.463 | 0.4775 | 0.4207 |
| Spag17 | 0.1174 | NA | NA | NA | NA | NA | NA | NA | NA | NA | NA | NA | NA | NA | NA |
| Irx5 | 0.0923 | NA | NA | NA | NA | NA | NA | NA | NA | NA | NA | NA | NA | NA | NA |
| Slc3a1 | 0.0836 | NA | NA | NA | NA | NA | NA | NA | NA | NA | NA | NA | NA | NA | NA |
| Thada | 0.0766 | NA | NA | NA | NA | NA | NA | NA | NA | NA | NA | NA | NA | NA | NA |
| Dgkg | 0.0739 | NA | NA | NA | NA | NA | NA | NA | NA | NA | NA | NA | NA | NA | NA |
| Zfp36l2 | 0.0737 | NA | NA | NA | NA | NA | NA | NA | NA | NA | NA | NA | NA | NA | NA |
| Entrep1 | 0.0558 | NA | NA | NA | NA | NA | NA | NA | NA | NA | NA | NA | NA | NA | 0.0563 |
| Glis2 | 0.0522 | NA | NA | NA | NA | NA | NA | NA | NA | NA | NA | NA | NA | NA | NA |
| Rpe | 0.0319 | NA | NA | NA | NA | NA | NA | NA | NA | NA | NA | NA | NA | NA | NA |
| Ptges3 | NA | 0.4645 | 0.5185 | NA | NA | 0.4368 | 0.4785 | 0.591 | 0.2966 | 0.4493 | 0.5176 | 0.4094 | 0.5047 | 0.5434 | 0.5214 |
| Stat6 | NA | 0.4054 | NA | NA | NA | NA | NA | NA | 0.2496 | NA | 0.4657 | 0.3642 | 0.455 | NA | NA |
| Tbccd1 | NA | 0.3507 | 0.3233 | NA | NA | NA | NA | NA | 0.0381 | NA | NA | 0.1288 | NA | NA | 0.1752 |
| Dnajb11 | NA | 0.3497 | 0.3227 | NA | NA | NA | NA | NA | 0.0379 | NA | NA | 0.1285 | NA | NA | 0.1748 |
| Crygs | NA | 0.3481 | 0.3214 | NA | NA | NA | 0.1193 | NA | 0.0366 | NA | NA | 0.127 | NA | NA | 0.1729 |
| Eif4a2 | NA | 0.3432 | NA | NA | NA | NA | NA | NA | NA | NA | NA | NA | NA | NA | NA |
| Eml2 | NA | 0.0644 | NA | NA | NA | NA | NA | NA | NA | NA | NA | NA | NA | NA | NA |
| Nab2 | NA | NA | 0.4718 | NA | NA | 0.4231 | NA | NA | 0.2503 | NA | NA | 0.3653 | NA | NA | NA |
| Ppp6c | NA | NA | 0.2443 | NA | NA | NA | NA | NA | NA | NA | NA | NA | NA | NA | NA |
| Rabepk | NA | NA | 0.243 | NA | NA | NA | NA | NA | NA | NA | NA | NA | NA | NA | NA |
| Rpap2 | NA | NA | 0.115 | NA | NA | NA | NA | NA | NA | NA | NA | NA | NA | NA | NA |
| Glmn | NA | NA | 0.1136 | NA | NA | NA | NA | NA | NA | NA | NA | NA | NA | NA | NA |
| Mgat5 | NA | NA | 0.0759 | NA | NA | NA | NA | NA | NA | NA | NA | NA | NA | NA | NA |
| Tmem163 | NA | NA | 0.0747 | NA | NA | NA | NA | NA | NA | NA | NA | NA | NA | NA | NA |
| Sppl2c | NA | NA | NA | 1.3182 | NA | NA | NA | NA | NA | NA | NA | NA | NA | NA | NA |
| Ecm2 | NA | NA | NA | NA | 0.133 | NA | NA | NA | NA | NA | NA | NA | NA | NA | NA |
| Tjp2 | NA | NA | NA | NA | 0.1023 | NA | NA | NA | NA | NA | NA | NA | NA | NA | NA |
| Gfi1 | NA | NA | NA | NA | 0.0571 | NA | NA | NA | NA | NA | NA | NA | NA | NA | NA |
| Sdr9c7 | NA | NA | NA | NA | NA | 0.4373 | NA | NA | NA | NA | NA | NA | NA | NA | NA |
| Coq10a | NA | NA | NA | NA | NA | 0.4301 | NA | NA | NA | NA | NA | NA | NA | NA | 0.5144 |
| Nxph4 | NA | NA | NA | NA | NA | 0.4193 | NA | NA | NA | NA | NA | NA | NA | NA | NA |
| Aspn | NA | NA | NA | NA | NA | NA | 0.4241 | NA | NA | NA | NA | NA | NA | NA | NA |
| As3mt | NA | NA | NA | NA | NA | NA | NA | 0.1067 | NA | NA | NA | NA | NA | NA | NA |
| Etfa | NA | NA | NA | NA | NA | NA | NA | 0.0875 | NA | NA | NA | NA | NA | NA | NA |
| Mip | NA | NA | NA | NA | NA | NA | NA | NA | 0.2956 | NA | NA | NA | NA | NA | NA |
| Spryd4 | NA | NA | NA | NA | NA | NA | NA | NA | 0.2956 | NA | NA | NA | NA | NA | NA |
| Apof | NA | NA | NA | NA | NA | NA | NA | NA | 0.2953 | NA | NA | NA | NA | NA | NA |
| Ankrd52 | NA | NA | NA | NA | NA | NA | NA | NA | 0.2937 | NA | NA | NA | NA | NA | NA |
| Myl6 | NA | NA | NA | NA | NA | NA | NA | NA | 0.2931 | NA | NA | NA | NA | NA | NA |
| Myl6b | NA | NA | NA | NA | NA | NA | NA | NA | 0.2931 | NA | NA | NA | NA | NA | NA |
| Esyt1 | NA | NA | NA | NA | NA | NA | NA | NA | 0.2923 | 0.4437 | NA | 0.4032 | NA | NA | NA |
| Zc3h10 | NA | NA | NA | NA | NA | NA | NA | NA | 0.292 | 0.4431 | NA | 0.4027 | NA | NA | NA |
| Lrp1 | NA | NA | NA | NA | NA | NA | NA | NA | 0.2474 | NA | NA | NA | NA | NA | NA |
| Haao | NA | NA | NA | NA | NA | NA | NA | NA | 0.0405 | NA | NA | NA | NA | NA | NA |
| Spata32 | NA | NA | NA | NA | NA | NA | NA | NA | 0.0283 | NA | NA | NA | NA | NA | NA |

**Supplementary Table 18. Gene loadings on PLS1 for mouse MGP analyses by cell-type gene list.**

| Gene | Chondrocytes | TFH | GCB | HEPG2 | HESC_ Hypothalamic_ NPC | HFOB | Osteoblasts | IPSC_ Hepatocytes | IPSC_NPC | IPSC_ Neurons | SGBS_ DIFF | NAIVEB | NAIVET | HMSC_ Adipocyte | hESC_ HypothalamicNeuron |
|---|---|---|---|---|---|---|---|---|---|---|---|---|---|---|---|
| **Pou3f2** | NA | NA | NA | NA | NA | NA | NA | NA | 0.0258 | NA | NA | NA | NA | NA | NA |
| **Isl2** | NA | NA | NA | NA | NA | NA | NA | NA | 0.0256 | NA | NA | NA | NA | NA | NA |
| **Gria1** | NA | NA | NA | NA | NA | NA | NA | NA | NA | 0.0544 | 0.0643 | NA | NA | NA | 0.0764 |
| **Acmsd** | NA | NA | NA | NA | NA | NA | NA | NA | NA | 0.0432 | NA | NA | NA | NA | NA |
| **Rfc4** | NA | NA | NA | NA | NA | NA | NA | NA | NA | NA | NA | 0.1245 | NA | NA | NA |
| **Snord59a** | NA | NA | NA | NA | NA | NA | NA | NA | NA | NA | NA | NA | NA | NA | 0.5214 |
| **Cast** | NA | NA | NA | NA | NA | NA | NA | NA | NA | NA | NA | NA | NA | NA | 0.1115 |
| **Nrxn3** | NA | NA | NA | NA | NA | NA | NA | NA | NA | NA | NA | NA | NA | NA | 0.0568 |

**Supplementary Table 19.** Summary of results for the multivariate genotype-phenotype (MGP) in humans on PLS axis 1.

| Cell Group | Gene List | PLS Magnitude | Procrustes Distance | Mean Displacement | Maximum Displacement | Variance Explained | P-value (Random Genes test) | P-value (Permutation test) | Effect size (Random Genes Test) | Effect size (Permutation Test) |
|---|---|---|---|---|---|---|---|---|---|---|
| HFOB | *BAZ2A, COQ10A, NAB2, NXPH4, PTGES3, SDR9C7, ZBTB39* | 7.7118 | 0.2903 | 0.0044 | 0.0072 | 0.0366 | 2.24E-59 | 0.0132 | 0.3 | 2.22 |
| HEPG2 | *BAZ2A, SPPL2C, ZBTB39* | 5.9902 | 0.2313 | 0.0033 | 0.0106 | 0.0292 | 3.87E-53 | 0.0656 | 0.34 | 1.51 |
| Human chondrocyte list | *ACTG1P22, CENPP, BDNF-AS, ARPC5L, BAZ2A, ATP5F1B, BDNF, CNNM2, CRNDE, DGKG, GIPR, GLS2, GOLGA1, GPR4, HSD17B6, FBXO46, HSPA5, IPPK, KLC3, KANSL1, LINC01159, LINC01819, LINC01833, LRRC37A, LINC02210, MAPT, NACA, MVB12B, NOL8, MYO1A, NT5C2, NR6A1, OLFML2A, PPM1N, PLEKHM1, PRIM1, OPA3, RPEL1, RBMS2, SFXN2, SPAG17, VASP, SLC3A1, THADA, WDR38, WBP1L, ZBTB39, ZFP36L2* | 6.3905 | 0.3741 | 0.0058 | 0.0095 | 0.0472 | 5.27E-40 | 0.0668 | 0.44 | 1.5 |
| Adipocytes | *BAZ2A, NACA, PTGES3, RBMS2, ZBTB39* | 8.6738 | 0.2445 | 0.0037 | 0.006 | 0.0308 | 1.03E-63 | 0.0684 | 0.27 | 1.488 |
| IPSC_HEPATOCYTES | *AS3MT, ETFA, NACA, PTGES3, BAZ2A, ZBTB39* | 7.9191 | 0.2451 | 0.0037 | 0.0059 | 0.0309 | 1.33E-62 | 0.0965 | 0.28 | 1.302 |
| IPSC_NEURONS | *NACA, GRIA1, ESYT1, BAZ2A, ACMSD, PTGES3, ZBTB39, RBMS2* | 8.3987 | 0.2496 | 0.0037 | 0.0061 | 0.0315 | 4.69E-60 | 0.1236 | 0.3 | 1.157 |
| NAIVET 0.028 lambda | *BAZ2A, RBMS2, NACA, PTGES3, STAT6, ZBTB39* | 7.6782 | 0.2175 | 0.0033 | 0.0049 | 0.0274 | 2.49E-127 | 0.1922 | 0.27 | 0.87 |
| IPSC_NPC | *AC093609.1, ANKRD52, APOF, ATP5B, BAZ2A, CRYGS, DNAJB11, ESYT1, FAM189A2, GPR182, HAAO, HSD17B6, ISL2, LRP1, MAP3K14-AS1, MIP, MYL6, MYL6B, NAB2, NACA, NACA, OLFML2A, POU3F2, PRIM1, PTGES3, RBMS2, RP11-474N8.5, RP11-603J24.14, RP11-603J24.6, SNORD59A, SPATA32, SPRYD4, STAT6, TBCCD1, TCONS_00020445, TCONS_00020805, TCONS_00020806, TCONS_00020807, TCONS_00021326, TCONS_00021327, TCONS_L2_00014535, TCONS_L2_00014536, TMEM194A, U59B, ZBTB39, ZC3H10* | 9.8678 | 0.2789 | 0.0043 | 0.0072 | 0.0352 | 1.77E-44 | 0.2171 | 0.37 | 0.782 |
| SGBS_DIFF | *BAZ2A, GRIA1, NACA, PTGES3, RBMS2, STAT6, ZBTB39* | 7.2924 | 0.2115 | 0.0032 | 0.0052 | 0.0267 | 0.0989 | 0.3587 | 0.15 | 0.362 |
| HESC_HYPOTHALAMICNPC | *BAZ2A, ECM2, BDNF, RBMS2, GFI1, NACA, TJP2, ZBTB39* | 6.1265 | 0.2119 | 0.0032 | 0.0065 | 0.0267 | 7.08E-67 | 0.4302 | 0.27 | 0.176 |
| TFH | *BAZ2A, GRIA1, NACA, PTGES3, RBMS2, STAT6, ZBTB39, DNAJB11, CRYGS, EML2, EIF4A2, TBCCD1* | 7.3456 | 0.2234 | 0.0034 | 0.0053 | 0.0282 | 4.70E-132 | 0.4328 | 0.27 | 0.169 |
| GCB | *ATP5B, BAZ2A, CRYGS, DNAJB11, DNAJB11, FAM189A2, GLMN, HSPA5, MGAT5, NAB2, NACA, PPP6C, PTGES3, RABEPK, RP11-48F14.1, RP11-48F14.1, RP11-65N13.8, RPAP2, SNORD59A, TBCCD1, TBCCD1, TCONS_00004230, TCONS_00005404, TCONS_00005404, TCONS_00006344, TCONS_00006344, TCONS_00006725, TCONS_00006725, TCONS_00016868, TCONS_00020445, TCONS_00020445, TCONS_00020805, TCONS_00020806, TCONS_00020807, TCONS_00020807, TCONS_00021326, TCONS_00021327, TCONS_L2_00029021, TCONS_L2_00029022, TMEM163, TMEM194A, U59B, ZBTB39* | 6.9563 | 0.2442 | 0.0038 | 0.0055 | 0.0308 | 1.17E-54 | 0.5158 | 0.33 | 0.04 |
| NAIVEB | *ZBTB39, NACA, BAZ2A, RBMS2, PTGES3, ESYT1, STAT6, NAB2, RFC4, TBCCD1, DNAJB11, CRYGS* | 7.5388 | 0.2072 | 0.0032 | 0.0055 | 0.0261 | 0.1442 | 0.5396 | 0.37 | 0.099 |
| HESC_HYPOTHALAMICNEURON | *CAST, COQ10A, CRYGS, GRIA1, MAPT, DNAJB11, NACA, BAZ2A, NR6A1, PTGES3, RBMS2, NRXN3, WDR38, ZBTB39, TBCCD1* | 6.1133 | 0.2011 | 0.0031 | 0.0059 | 0.0254 | 7.63E-64 | 0.6328 | 0.26 | 0.339 |
| HMSC_OSTEOBLASTS | *ASPN, BAZ2A, CRYGS, NOL8, NACA, PTGES3, ZBTB39* | 8.4182 | 0.169 | 0.0026 | 0.0052 | 0.0213 | 5.01E-77 | 0.6418 | 0.19 | 0.363 |

**Supplementary Table 20.** Gene loadings on PLS1 for mouse MGP analyses by cell-type gene list.

| Cell | Chondrocytes | HFOB | HMSC_ Adipocyte | TFH | SGBS_DIFF | NAIVEB | HEPG2 | IPSC_NPC | IPSC_ Neurons | IPSC_ Hepatocytes | HESC_ Hypothalamic_ neuron | HMSC_ Osteoblasts | GCB | HESC_ Hypothalamic_ NPC | NAIVET |
|---|---|---|---|---|---|---|---|---|---|---|---|---|---|---|---|
| ZFP36L2 | 0.5284 | NA | NA | NA | NA | NA | NA | NA | NA | NA | NA | NA | NA | NA | NA |
| RPEL1 | 0.3967 | NA | NA | NA | NA | NA | NA | NA | NA | NA | NA | NA | NA | NA | NA |
| HSD17B6 | 0.3816 | NA | NA | NA | NA | NA | NA | 0.5652 | NA | NA | NA | NA | 0.4871 | NA | NA |
| BAZ2A | 0.3388 | 0.49 | 0.6296 | 0.3653 | 0.3382 | 0.3099 | 0.1541 | 0.5098 | 0.6368 | 0.6167 | 0.27 | 0.0329 | 0.435 | 0.27 | 0.394 |
| FBXO46 | 0.2922 | NA | NA | NA | NA | NA | NA | NA | NA | NA | NA | NA | NA | NA | NA |
| NACA | 0.2284 | NA | 0.4725 | 0.2645 | 0.2442 | 0.2287 | NA | 0.3107 | 0.4798 | 0.472 | 0.2185 | 0.0254 | 0.2727 | 0.2185 | 0.2834 |
| PRIM1 | 0.2031 | NA | NA | NA | NA | NA | NA | 0.3212 | NA | NA | NA | NA | 0.2723 | NA | NA |
| LINC01159 | 0.1751 | NA | NA | NA | NA | NA | NA | NA | NA | NA | NA | NA | NA | NA | NA |
| LINC01819 | 0.1482 | NA | NA | NA | NA | NA | NA | NA | NA | NA | NA | NA | NA | NA | NA |
| LINC02210 | 0.1464 | NA | NA | NA | NA | NA | NA | NA | NA | NA | NA | NA | NA | NA | NA |
| MAPT | 0.1443 | NA | NA | NA | NA | NA | NA | NA | NA | NA | NA | NA | NA | NA | NA |
| NT5C2 | 0.1399 | NA | NA | NA | NA | NA | NA | NA | NA | NA | NA | NA | NA | NA | NA |
| CNNM2 | 0.1376 | NA | NA | NA | NA | NA | NA | NA | NA | NA | NA | NA | NA | NA | NA |
| KLC3 | 0.1257 | NA | NA | NA | NA | NA | NA | NA | NA | NA | NA | NA | NA | NA | NA |
| ATP5F1B | 0.1233 | NA | NA | NA | NA | NA | NA | NA | NA | NA | NA | NA | NA | NA | NA |
| PLEKHM1 | 0.1215 | NA | NA | NA | NA | NA | NA | NA | NA | NA | NA | NA | NA | NA | NA |
| ZBTB39 | 0.121 | 0.3676 | 0.1223 | 0.5498 | 0.5693 | 0.589 | 0.2265 | 0.1181 | 0.1345 | 0.218 | 0.1662 | 0.0103 | 0.0448 | 0.1662 | 0.5527 |
| SFXN2 | 0.1193 | NA | NA | NA | NA | NA | NA | NA | NA | NA | NA | NA | NA | NA | NA |
| NR6A1 | 0.1138 | NA | NA | NA | NA | NA | NA | NA | NA | NA | NA | NA | NA | NA | NA |
| GPR4 | 0.1041 | NA | NA | NA | NA | NA | NA | NA | NA | NA | NA | NA | NA | NA | NA |
| ARPC5L | 0.0977 | NA | NA | NA | NA | NA | NA | NA | NA | NA | NA | NA | NA | NA | NA |
| LRRC37A2 | 0.0926 | NA | NA | NA | NA | NA | NA | NA | NA | NA | NA | NA | NA | NA | NA |
| KANSL1 | 0.0924 | NA | NA | NA | NA | NA | NA | NA | NA | NA | NA | NA | NA | NA | NA |
| HSPA5 | 0.0905 | NA | NA | NA | NA | NA | NA | NA | NA | NA | NA | NA | 0.032 | NA | NA |
| PPM1N | 0.0886 | NA | NA | NA | NA | NA | NA | NA | NA | NA | NA | NA | NA | NA | NA |
| VASP | 0.0843 | NA | NA | NA | NA | NA | NA | NA | NA | NA | NA | NA | NA | NA | NA |
| OPA3 | 0.0796 | NA | NA | NA | NA | NA | NA | NA | NA | NA | NA | NA | NA | NA | NA |
| OLFML2A | 0.0713 | NA | NA | NA | NA | NA | NA | 0.0023 | NA | NA | NA | NA | 0.0147 | NA | NA |
| GLS2 | 0.069 | NA | NA | NA | NA | NA | NA | NA | NA | NA | NA | NA | NA | NA | NA |
| THADA | 0.0689 | NA | NA | NA | NA | NA | NA | NA | NA | NA | NA | NA | NA | NA | NA |
| WBP1L | 0.0677 | NA | NA | NA | NA | NA | NA | NA | NA | NA | NA | NA | NA | NA | NA |
| GOLGA1 | 0.0664 | NA | NA | NA | NA | NA | NA | NA | NA | NA | NA | NA | NA | NA | NA |
| GIPR | 0.0642 | NA | NA | NA | NA | NA | NA | NA | NA | NA | NA | NA | NA | NA | NA |
| WDR38 | 0.0599 | NA | NA | NA | NA | NA | NA | NA | NA | NA | NA | NA | NA | NA | NA |
| RBMS2 | 0.0517 | NA | 0.0347 | 0.0598 | 0.0657 | 0.0129 | NA | 0.0331 | 0.0061 | NA | 0.0033 | NA | 0.0402 | 0.0033 | 0.0108 |
| SPAG17 | 0.0477 | NA | NA | NA | NA | NA | NA | NA | NA | NA | NA | NA | NA | NA | NA |
| LINC01833 | 0.0421 | NA | NA | NA | NA | NA | NA | NA | NA | NA | NA | NA | NA | NA | NA |
| MVB12B | 0.0357 | NA | NA | NA | NA | NA | NA | NA | NA | NA | NA | NA | NA | NA | NA |
| IPPK | 0.0334 | NA | NA | NA | NA | NA | NA | NA | NA | NA | NA | NA | NA | NA | NA |
| MYO1A | 0.0315 | NA | NA | NA | NA | NA | NA | NA | NA | NA | NA | NA | NA | NA | NA |
| DGKG | 0.0295 | NA | NA | NA | NA | NA | NA | NA | NA | NA | NA | NA | NA | NA | NA |
| LRRC37A | 0.0282 | NA | NA | NA | NA | NA | NA | NA | NA | NA | NA | NA | NA | NA | NA |
| SLC3A1 | 0.0225 | NA | NA | NA | NA | NA | NA | NA | NA | NA | NA | NA | NA | NA | NA |
| CENPP | 0.0156 | NA | NA | NA | NA | NA | NA | NA | NA | NA | NA | NA | NA | NA | NA |
| BDNF | 0.0145 | NA | NA | NA | NA | NA | NA | NA | NA | NA | 0.6082 | NA | NA | 0.6082 | NA |
| NOL8 | 0.0133 | NA | NA | NA | NA | NA | NA | NA | NA | NA | NA | 0.7071 | NA | NA | NA |
| CRNDE | 0.009 | NA | NA | NA | NA | NA | NA | NA | NA | NA | NA | NA | NA | NA | NA |
| ACTG1P22 | 0.0078 | NA | NA | NA | NA | NA | NA | NA | NA | NA | NA | NA | NA | NA | NA |
| BDNF-AS | 0.0054 | NA | NA | NA | NA | NA | NA | NA | NA | NA | NA | NA | NA | NA | NA |
| SDR9C7 | NA | 0.608 | NA | NA | NA | NA | NA | NA | NA | NA | NA | NA | NA | NA | NA |
| PTGES3 | NA | 0.5405 | 0.6664 | 0.4135 | 0.385 | 0.363 | NA | 0.551 | 0.672 | 0.657 | NA | 0.0352 | 0.4618 | NA | 0.4418 |
| NXPH4 | NA | 0.153 | NA | NA | NA | NA | NA | NA | NA | NA | NA | NA | NA | NA | NA |
| NAB2 | NA | 0.1385 | NA | NA | NA | 0.0944 | NA | 0.0174 | NA | NA | NA | NA | 0.0063 | NA | NA |
| COQ10A | NA | 0.1133 | NA | NA | NA | NA | NA | NA | NA | NA | NA | NA | NA | NA | NA |
| STAT6 | NA | NA | NA | 0.5964 | 0.619 | 0.6175 | NA | 0.0776 | NA | NA | NA | NA | 0.0063 | NA | 0.5761 |
| GRIA1 | NA | NA | NA | 0.21 | 0.216 | NA | NA | NA | 0.0735 | NA | NA | NA | NA | NA | NA |
| TBCCD1 | NA | NA | NA | 0.0589 | NA | 0.0246 | NA | 0.0749 | NA | NA | NA | NA | 0.2746 | NA | NA |
| CRYGS | NA | NA | NA | 0.0501 | NA | 0.0943 | NA | 0.0272 | NA | NA | NA | 0.0385 | 0.0297 | NA | NA |
| DNAJB11 | NA | NA | NA | 0.0424 | NA | 0.0037 | NA | 0.0802 | NA | NA | NA | NA | 0.1455 | NA | NA |
| EML2 | NA | NA | NA | 0.0301 | NA | NA | NA | NA | NA | NA | NA | NA | NA | NA | NA |
| EIF4A2 | NA | NA | NA | 0.0084 | NA | NA | NA | NA | NA | NA | NA | NA | NA | NA | NA |
| ESYT1 | NA | NA | NA | NA | NA | 0.1544 | NA | 0.0082 | 0.0446 | NA | NA | NA | 0.1475 | NA | NA |
| RFC4 | NA | NA | NA | NA | NA | 0.036 | NA | NA | NA | NA | NA | NA | NA | NA | NA |
| SPPL2C | NA | NA | NA | NA | NA | NA | 0.9618 | NA | NA | NA | NA | NA | NA | NA | NA |
| ISL2 | NA | NA | NA | NA | NA | NA | NA | 0.0863 | NA | NA | NA | NA | 0.1746 | NA | NA |
| MYL6B | NA | NA | NA | NA | NA | NA | NA | 0.0488 | NA | NA | NA | NA | 0.1294 | NA | NA |
| SPRYD4 | NA | NA | NA | NA | NA | NA | NA | 0.0377 | NA | NA | NA | NA | 0.0068 | NA | NA |
| ANKRD52 | NA | NA | NA | NA | NA | NA | NA | 0.0353 | NA | NA | NA | NA | 0.0235 | NA | NA |
| HAAO | NA | NA | NA | NA | NA | NA | NA | 0.0345 | NA | NA | NA | NA | 0.0024 | NA | NA |
| SPATA32 | NA | NA | NA | NA | NA | NA | NA | 0.0194 | NA | NA | NA | NA | 0.1911 | NA | NA |
| MIP | NA | NA | NA | NA | NA | NA | NA | 0.0156 | NA | NA | NA | NA | 0.1 | NA | NA |
| LRP1 | NA | NA | NA | NA | NA | NA | NA | 0.0136 | NA | NA | NA | NA | 0.2166 | NA | NA |
| MYL6 | NA | NA | NA | NA | NA | NA | NA | 0.009 | NA | NA | NA | NA | 0.0651 | NA | NA |
| POU3F2 | NA | NA | NA | NA | NA | NA | NA | 0.0023 | NA | NA | NA | NA | 0.0142 | NA | NA |
| ACMSD | NA | NA | NA | NA | NA | NA | NA | NA | 0.0714 | NA | NA | NA | NA | NA | NA |
| ETFA | NA | NA | NA | NA | NA | NA | NA | NA | NA | 0.2698 | NA | NA | NA | NA | NA |
| AS3MT | NA | NA | NA | NA | NA | NA | NA | NA | NA | 0.0861 | NA | NA | NA | NA | NA |
| TJP2 | NA | NA | NA | NA | NA | NA | NA | NA | NA | NA | 0.6452 | NA | NA | 0.6452 | NA |
| ECM2 | NA | NA | NA | NA | NA | NA | NA | NA | NA | NA | 0.2801 | NA | NA | 0.2801 | NA |
| GFI1 | NA | NA | NA | NA | NA | NA | NA | NA | NA | NA | 0.0093 | NA | NA | 0.0093 | NA |
| ASPN | NA | NA | NA | NA | NA | NA | NA | NA | NA | NA | NA | 0.7076 | NA | NA | NA |
| GLMN | NA | NA | NA | NA | NA | NA | NA | NA | NA | NA | NA | NA | 0.4426 | NA | NA |

| Cell | Chondrocytes | HFOB | HMSC_ Adipocyte | TFH | SGBS_DIFF | NAIVEB | HEPG2 | IPSC_NPC | IPSC_ Neurons | IPSC_ Hepatocytes | HESC_ Hypothalamic_ neuron | HMSC_ Osteoblasts | GCB | HESC_ Hypothalamic_ NPC | NAIVET |
|---|---|---|---|---|---|---|---|---|---|---|---|---|---|---|---|
| TMEM163 | NA | NA | NA | NA | NA | NA | NA | NA | NA | NA | NA | NA | 0.2294 | NA | NA |
| RPAP2 | NA | NA | NA | NA | NA | NA | NA | NA | NA | NA | NA | NA | 0.2145 | NA | NA |
| MGAT5 | NA | NA | NA | NA | NA | NA | NA | NA | NA | NA | NA | NA | 0.1899 | NA | NA |
| PPP6C | NA | NA | NA | NA | NA | NA | NA | NA | NA | NA | NA | NA | 0.0714 | NA | NA |
| RABEPK | NA | NA | NA | NA | NA | NA | NA | NA | NA | NA | NA | NA | 0.0466 | NA | NA |

**Supplementary Table 21. Significant eQTL colocalization results.** Only -log10 P values for significant gene-tissue colocalizations are provided. Full results for all gene-tissue pairs that met criteria for colocalization testing are included in Supplementary Table XX.

| Trait | Stratum | Lead SNP | Gene | N Genes in Region | N of eQTL datasets tested | Locus -log10 P-value threshold | Skeletal Muscle | Lung | Visceral Adipose | Subcutaneous Adipose |
|---|---|---|---|---|---|---|---|---|---|---|
| OSA | Multi-Population | rs12725199 | *PTBP2* | 2 | 5 | 2.00 | 7.23 | 7.97 | 7.03 | 8.00 |
| OSA | Multi-Population | rs9380814 | *SAYSD1* | 9 | 12 | 2.38 | - | - | 5.65 | - |
| OSA | Multi-Population | rs2958127 | *SPRYD4* | 34 | 5 | 2.00 | - | 4.50 | - | |
| OSA | Multi-Population | rs11642015 | *FTO* | 5 | 6 | 2.08 | 8.35 | - | - | - |
| OSA | Multi-Population | rs11642015 | *RBL2* | 5 | 6 | 2.08 | 3.29 | - | - | - |
| OSA | Multi-Population | rs11088984 | *FAM207A* | 10 | 15 | 2.48 | - | - | | 5.44 |
| OSAadjBMI | Multi-Population | rs2964006 | *GRIA1* | 4 | 12 | 2.38 | - | 7.44 | - | - |
| OSAadjBMI | Multi-Population | rs2964006 | *MFAP3* | 4 | 12 | 2.38 | - | - | 3.19 | 3.10 |
| OSAadjBMI | Multi-Population | rs4759254 | *SPRYD4* | 33 | 8 | 2.20 | - | 4.45 | - | - |
| OSAadjBMI | Multi-Population | rs10735928 | *LEMD3* | 6 | 7 | 2.15 | 2.72 | - | - | 3.56 |
| OSAadjBMI | Multi-Population | rs10735928 | *WIF1* | 6 | 7 | 2.15 | - | 7.38 | 4.19 | - |
| AHI | Multi-Population | rs13015934** | *DNAH7* | 2 | 6 | 2.08 | - | - | 2.89 | 2.52 |
| avgSAO2 | EUR-only | rs2194955 | *GFRA2* | 3 | 1 | 1.30 | - | - | 5.81 | - |

**Lead GWAS SNP at this locus was rs1200596362, which was not available in GTEx data. TOP GWAS SNP in overlapping set is provided.

**Supplementary Table 22.** eQTL colocaization results for all gene - tissue pairs that passed stage one significance criteria and were tested for colocalization.

| Trait | Strata | Lead SNP | Gene | N Genes | N of SNPs matching between GWAS and eQTL | N SNPs used in calculation | N of eQTL datasets tested | Locus -log10 P-value threshold | Whole Blood | Skeletal Muscle | Lung | Visceral Adipose | Subcutaneous Adipose | Brain - Hippocampus | Brain - Hypothalamus |
|---|---|---|---|---|---|---|---|---|---|---|---|---|---|---|---|
| OSA | multi-population | rs12725199 | *DPYD* | 2 | 2234 | 514 | 5 | 2.000 | - | - | 0.1020 | - | - | - | - |
| **OSA** | **multi-population** | **rs12725199** | ***PTBP2*** | 2 | 2234 | 1377 | 5 | 2.000 | - | **7.2300** | **7.9700** | **7.0300** | **8.0000** | - | - |
| OSA | multi-population | rs9831061 | *DGKG* | 10 | 2056 | 1255 | 2 | 1.602 | - | 0.0000 | - | - | - | - | - |
| OSA | multi-population | rs9831061 | *SENP2* | 10 | 2056 | 1219 | 2 | 1.602 | - | - | - | 0.1080 | - | - | - |
| OSA | multi-population | rs9380814 | *GLO1* | 9 | 2802 | 1824 | 12 | 2.380 | 0.0004 | 0.0000 | 0.0657 | - | 0.0031 | - | - |
| OSA | multi-population | rs9380814 | *GLP1R* | 9 | 2802 | 1824 | 12 | 2.380 | - | - | 0.2560 | - | - | - | - |
| OSA | multi-population | rs9380814 | *KCNK17* | 9 | 2802 | 1824 | 12 | 2.380 | 0.0803 | - | 0.2730 | - | 0.0026 | - | - |
| OSA | multi-population | rs9380814 | *KCNK5* | 9 | 2802 | 1824 | 12 | 2.380 | - | - | - | - | 1.4300 | - | - |
| **OSA** | **multi-population** | **rs9380814** | ***SAYSD1*** | 9 | 2802 | 1824 | 12 | 2.380 | 0.3240 | - | 0.0683 | **5.6500** | - | - | - |
| OSA | multi-population | rs12553324 | *ELAVL2* | 1 | 2810 | 1690 | 1 | 1.301 | - | - | - | 0.0038 | - | - | - |
| OSA | multi-population | rs2958127 | *RBMS2* | 34 | 1620 | 1003 | 5 | 2 | - | 0.6500 | - | - | - | - | - |
| **OSA** | **multi-population** | **rs2958127** | ***SPRYD4*** | 34 | 1620 | 1003 | 5 | 2 | - | - | **4.5000** | - | 0.2010 | - | - |
| OSA | multi-population | rs2958127 | *TMEM194A* | 34 | 1620 | 1003 | 5 | 2 | - | 0.0000 | 0.0012 | - | - | - | - |
| **OSA** | **multi-population** | **rs11642015** | ***FTO*** | 5 | 2503 | 1559 | 6 | 2.079181246 | - | **8.3500** | - | - | - | - | - |
| **OSA** | **multi-population** | **rs11642015** | ***RBL2*** | 5 | 2503 | 1559 | 6 | 2.079181246 | 0.7310 | **3.2900** | 0.2130 | 0.3410 | 0.0535 | - | - |
| OSA | multi-population | rs11642015 | *RPGRIP1L* | 5 | 2503 | 1559 | 6 | 2.079181246 | - | - | - | - | - | - | - |
| OSA | multi-population | rs1800437 | *CCDC61* | 35 | 2435 | 1536 | 20 | 2.602059991 | - | 0.1910 | - | - | - | - | - |
| OSA | multi-population | rs1800437 | *DMPK* | 35 | 2435 | 1536 | 20 | 2.602059991 | 0.0257 | - | 0.3410 | - | 0.4780 | - | - |
| OSA | multi-population | rs1800437 | *DMWD* | 35 | 2435 | 1536 | 20 | 2.602059991 | - | - | 0.2760 | 0.6620 | 0.8470 | - | - |
| OSA | multi-population | rs1800437 | *ERCC2* | 35 | 2435 | 1536 | 20 | 2.602059991 | - | 0.6150 | 0.2720 | 0.0176 | 0.7210 | - | - |
| OSA | multi-population | rs1800437 | *MARK4* | 35 | 2435 | 1536 | 20 | 2.602059991 | 0.0003 | 0.0010 | - | - | - | - | - |
| OSA | multi-population | rs1800437 | *OPA3* | 35 | 2435 | 1536 | 20 | 2.602059991 | - | 0.0373 | - | - | - | 0.0211 | - |
| OSA | multi-population | rs1800437 | *PPM1N* | 35 | 2435 | 1536 | 20 | 2.602059991 | - | - | 0.1290 | - | 0.0150 | - | - |
| OSA | multi-population | rs1800437 | *RTN2* | 35 | 2435 | 1536 | 20 | 2.602059991 | - | 1.0500 | - | - | - | - | - |
| OSA | multi-population | rs1800437 | *SNRPD2* | 35 | 2435 | 1536 | 20 | 2.602059991 | - | - | - | - | 0.9200 | - | - |
| OSA | multi-population | rs1800437 | *SYMPK* | 35 | 2435 | 1536 | 20 | 2.602059991 | - | - | 0.8460 | - | - | - | - |
| OSA | multi-population | rs11088984 | *COL18A1* | 10 | 3140 | 1939 | 15 | 2.477121255 | 0.0000 | 0.4260 | - | - | 0.0049 | - | - |
| **OSA** | **multi-population** | **rs11088984** | ***FAM207A*** | 10 | 3140 | 1939 | 15 | 2.477121255 | - | - | - | 0.8630 | **5.4400** | - | - |
| OSA | multi-population | rs11088984 | *PCBP3* | 10 | 3140 | 1939 | 15 | 2.477121255 | 1.0700 | 1.4800 | 1.9100 | 1.7600 | 1.4200 | - | - |
| OSA | multi-population | rs11088984 | *PTTG1IP* | 10 | 3140 | 1939 | 15 | 2.477121255 | 1.6300 | 0.0000 | 0.5410 | - | 0.9590 | - | - |
| OSA | multi-population | rs11088984 | *UBE2G2* | 10 | 3140 | 1939 | 15 | 2.477121255 | - | - | - | - | 0.0120 | - | - |
| OSA | EUR-only | rs34592089 | *BANK1* | 3 | 1698 | 1297 | 5 | 2 | - | 0.0168 | - | - | 0.0281 | - | - |
| OSA | EUR-only | rs34592089 | *SLC39A8* | 3 | 1698 | 1420 | 5 | 2 | 1.9900 | - | 0.1850 | - | 0.8980 | - | - |
| OSAadjBMI | multi-population | rs2964006 | *FAM114A2* | 4 | 2403 | 1682 | 12 | 2.380211242 | 0.7580 | 0.0000 | - | 0.0005 | 0.0794 | - | - |
| OSAadjBMI | multi-population | rs2964006 | *GALNT10* | 4 | 2403 | 1682 | 12 | 2.380211242 | - | - | 0.0002 | - | 0.0000 | 1.3600 | 0.7580 |
| OSAadjBMI | multi-population | rs2964006 | ***GRIA1*** | 4 | 2403 | 1682 | 12 | 2.380211242 | - | - | **7.4400** | - | - | - | - |
| OSAadjBMI | multi-population | rs2964006 | ***MFAP3*** | 4 | 2403 | 1682 | 12 | 2.380211242 | 0.0000 | - | - | **3.1900** | **3.1000** | - | - |
| OSAadjBMI | multi-population | rs4759254 | *RBMS2* | 33 | 1595 | 1000 | 8 | 2.204119983 | 0.0000 | 0.6890 | - | - | - | - | - |
| **OSAadjBMI** | **multi-population** | **rs4759254** | ***SPRYD4*** | 33 | 1595 | 1000 | 8 | 2.204119983 | - | - | **4.4500** | - | 0.7370 | - | - |
| OSAadjBMI | multi-population | rs4759254 | *STAT2* | 33 | 1595 | 1000 | 8 | 2.204119983 | - | - | - | 0.0000 | 0.0001 | - | - |
| OSAadjBMI | multi-population | rs4759254 | *TMEM194A* | 33 | 1595 | 1000 | 8 | 2.204119983 | - | 0.0003 | 0.0027 | - | - | - | - |
| **OSAadjBMI** | **multi-population** | **rs10735928** | ***LEMD3*** | 6 | 2017 | 1108 | 7 | 2.146128036 | - | **2.7200** | 2.1200 | 1.6300 | **3.5600** | - | - |
| **OSAadjBMI** | **multi-population** | **rs10735928** | ***WIF1*** | 6 | 2017 | 1108 | 7 | 2.146128036 | - | 0.1140 | **7.3800** | **4.1900** | - | - | - |
| OSAadjBMI | EUR-only | rs73649650* | *FAM189A2* | 8 | 1742 | 1493 | 12 | 2.380211242 | - | 0.0100 | 0.1540 | - | - | - | - |
| OSAadjBMI | EUR-only | rs73649650* | *FXN* | 8 | 1742 | 1493 | 12 | 2.380211242 | - | 0.3460 | - | - | - | 1.9400 | - |
| OSAadjBMI | EUR-only | rs73649650* | *PIP5K1B* | 8 | 1742 | 1363 | 12 | 2.380211242 | - | - | - | 0.1880 | 0.0081 | - | - |
| OSAadjBMI | EUR-only | rs73649650* | *PTAR1* | 8 | 1742 | 1493 | 12 | 2.380211242 | 0.0001 | - | - | - | - | - | - |
| OSAadjBMI | EUR-only | rs73649650* | *TJP2* | 8 | 1742 | 1493 | 12 | 2.380211242 | 0.0404 | 0.0171 | 0.0124 | 2.0600 | 0.9110 | - | - |
| **AHI** | **multi-population** | **rs13015934**** | ***DNAH7*** | 2 | 2511 | 1291 | 6 | 2.079181246 | - | - | - | **2.8900** | **2.5200** | - | - |
| AHI | multi-population | rs13015934** | *SLC39A10* | 2 | 2511 | 1400 | 6 | 2.079181246 | - | 1.8900 | 1.8000 | 2.0200 | 2.0100 | - | - |
| avgSAO$_2$ | multi-population | rs802729 | *KIAA0408* | 5 | 2404 | 1096 | 3 | 1.77815125 | 0.6730 | - | - | - | - | - | - |
| avgSAO$_2$ | multi-population | rs802729 | *SOGA3* | 5 | 2404 | 1096 | 3 | 1.77815125 | - | - | 0.7090 | - | 0.8760 | - | - |
| **avgSaO2** | **EUR-only** | **rs2194955** | ***GFRA2*** | 3 | 1961 | 1675 | 1 | 1.301029996 | - | - | - | **5.8100** | - | - | - |
| avgSaO2adjBMI | multi-population | rs79914264 | *EPB41* | 6 | 2370 | 1147 | 9 | 2.255272505 | - | 0.3720 | 0.7190 | - | 0.2810 | - | - |
| avgSaO2adjBMI | multi-population | rs79914264 | *MECR* | 6 | 2370 | 1147 | 9 | 2.255272505 | - | 0.2770 | 0.8810 | 0.6430 | - | - | - |
| avgSaO2adjBMI | multi-population | rs79914264 | *OPRD1* | 6 | 2370 | 1048 | 9 | 2.255272505 | - | 0.6740 | - | - | - | - | - |
| avgSaO2adjBMI | multi-population | rs79914264 | *PTPRU* | 6 | 2370 | 1147 | 9 | 2.255272505 | - | - | - | - | 0.0366 | - | - |
| avgSaO2adjBMI | multi-population | rs79914264 | *TMEM200B* | 6 | 2370 | 1147 | 9 | 2.255272505 | - | - | - | - | 0.3490 | - | - |

*Lead GWAS SNP at this locus was rs60791051, which was not available in GTEx data. TOP GWAS SNP in overlapping set is provided.

**Lead GWAS SNP at this locus was rs1200596362, which was not available in GTEx data. TOP GWAS SNP in overlapping set is provided.
